# Multi-omics Characterization of Neutrophil Extracellular Trap Formation in Severe and Mild COVID-19 Infections

**DOI:** 10.1101/2022.04.26.22274196

**Authors:** Lisa M. Bramer, Robert D. Hontz, Amie J. Eisfeld, Amy C. Sims, Young-Mo Kim, Kelly G. Stratton, Carrie D. Nicora, Marina A. Gritsenko, Athena A. Schepmoes, Osamu Akasaka, Michiko Koga, Takeya Tsutsumi, Morio Nakamura, Ichiro Nakachi, Rie Baba, Hiroki Tateno, Shoji Suzuki, Hideaki Nakajima, Hideaki Kato, Kazunari Ishida, Makoto Ishii, Yoshifumi Uwamino, Keiko Mitamura, Vanessa L. Paurus, Ernesto S. Nakayasu, Isaac K. Attah, Andrew G. Letizia, Katrina M. Waters, Thomas O. Metz, Karen Corson, Yoshihiro Kawaoka, Vincent R. Gerbasi

## Abstract

The detailed mechanisms of COVID-19 infection pathology remain poorly understood. To improve our understanding of SARS-CoV-2 pathology, we performed a multi-omics analysis of an immunologically naïve SARS-CoV-2 clinical cohort from the plasma of uninfected controls, mild, and severe infections. A comparison of healthy controls and patient samples showed activation of neutrophil degranulation pathways and formation of neutrophil extracellular trap (NET) complexes that were activated in a subset of the mild infections and more prevalent in severe infections (containing multiple NET proteins in individual patient samples). As a potential mechanism to suppress NET formation, multiple redox enzymes were elevated in the mild and severe symptom population. Analysis of metabolites from the same cohort showed a 24- and 60-fold elevation in plasma L-cystine, the oxidized form of cysteine, which is a substrate of the powerful antioxidant glutathione, in mild and severe patients, respectively. Unique to patients with mild infections, the carnosine dipeptidase modifying enzyme (CNDP1) was up-regulated. The strong protein and metabolite oxidation signatures suggest multiple compensatory pathways working to suppress oxidation and NET formation in SARS-CoV-2 infections.

## Introduction

To date the severe acute respiratory syndrome coronavirus-2 (SARS-CoV-2) pandemic has reached 500 million cases with 6.2 million deaths. Initial reports of the SARS-CoV-2 novel coronavirus-2019 (nCOV-2019) outbreak indicated the presence of a novel beta-coronavirus from bronchoalveolar lavage fluid causing fever and atypical pneumonia among infected patients (1). Severe acute respiratory syndrome coronavirus-1 (SARS-CoV-1) and Middle Eastern respiratory syndrome coronavirus (MERS-CoV) share similar transmission and symptom characteristics with SARS-CoV-2 but demonstrate considerably higher mortality rates (2). Despite the potential for SARS-CoV-2 to develop into acute respiratory distress syndrome (ARDS), the rate of asymptomatic infection is estimated to be 30-40% (3, 4). The penetrance of SARS-CoV-2 in the global population, and disparity in host response between individuals, underscores the need to characterize mechanisms and markers associated with immunopathology.

The measurement of biomolecules from blood cells and plasma using mass spectrometry has accelerated our understanding of coronavirus disease -2019 (COVID-19) infections (5, 6). While the sensitivity and reliability of mass spectrometry measurements has increased substantially in the past decade, mass analyzers lack an amplification mechanism similar to nucleic acid deep sequencing approaches that employ the logarithmic polymerase chain reaction. The lower sensitivity of “-omics” approaches employing mass spectrometry can still lead to discovery of prevailing differences between experimental and control populations. This is achievable by imposing appropriate quality control measures during sample preparation and analysis (7). We hypothesized that this might be the case for SARS-CoV-2 infections where infection severity varies greatly but is largely associated with extreme differences in inflammatory processes including neutrophil influx proximal to the alveolar-capillary barrier (8, 9).

In the study described herein, we characterized plasma proteins and metabolites from healthy controls (HC), and from both mild, and severe SARS-CoV-2 infections. All samples from this study originated from individuals infected with SARS-CoV-2 during March of 2020 (prior to vaccination and recurrent infections), thus the likelihood of any established adaptive immunity was low. As a result, our study likely demonstrates the natural course of infection with the ancestral strain of SARS-CoV-2 without the influence of poised adaptive immune responses against the virus.

Prior efforts towards identification of protective mechanisms have focused on adaptive immunity, and clearly showed that antibodies directed against SARS-CoV-2 Spike protein neutralize virus in natural infection and vaccine recipients (10-12). Few studies have worked to identify innate physiological mechanisms that compensate for stress imposed by the infection. Our results indicate that viral infection triggers antioxidant pathways in mild and severe patient cohorts. Patient populations with severe infection show elevation of antioxidant pathway enzymes including up-regulation of cellular superoxide dismutase, glutathione metabolism, and oxidized glutathione metabolite derivatives. Importantly, carnosine dipeptidase 1 (CNDP1), an enzyme required for controlling oxidative stress was elevated in the mild patient population, but not healthy controls or those with severe clinical outcomes. These results raise the possibility that one of the distinguishing features of severe and mild infections is the ability to suppress a threshold of oxidative stress, the ultimate trigger of neutrophil extracellular trap (NET) formation (13). NETs function as antiviral and antimicrobial DNA-protein complexes released by diverse infection types including bacterial (14), parasitic (15), and viral infections (5, 16, 17).

Importantly, NET levels were elevated in severe infection when compared to mild infection and healthy control populations. However, substantial NET formation was observed in a subpopulation of mild infections, suggesting NET formation might be a candidate for post-infection sequelae mechanisms in non-hospitalized patients. Additionally, NET levels were highly correlated with pulmonary surfactant protein leakage from the pulmonary microcapillaries. These results suggest that the extent of NET complex formation might drive the degree of pulmonary capillary permeability.

## Results

### Proteomics Analysis of Patient Plasma

We sought to characterize the host response among patients infected with SARS-CoV-2 through plasma proteomics analysis. A total of 151 patient samples were analyzed, including 40 healthy controls (HCs), 77 mild cases (a PCR-positive COVID 19 test but did not require hospitalization), and 34 severe cases that required oxygen, hospitalization, and/or other support (see methods). Prior to tandem-mass-tag (TMT) labeling (18) and mass spectrometry analysis, abundant plasma proteins were immunodepleted to improve sensitivity. Additionally, TMT-labeled peptides were subjected to high pH, reversed-phase chromatography to enhance the depth of protein identification and quantitation (Figure 1). A total of 2,766 proteins were quantified in our study. A comparison of HC to mild infections revealed a statistically significant difference (adjusted p<0.05) among 209 different proteins with 141 proteins elevated in mild cases and 68 proteins decreased relative to HCs. Comparing severe cases to HCs showed a statistically significant difference in 1,377 different proteins (adjusted p<0.05) with 1,129 proteins elevated and 248 proteins decreased in individuals with severe infection. When comparing mild to severe patients, there were 1,318 proteins statistically different (adjusted p<0.05), with an elevation in 1,097 proteins and a decrease in 221 proteins in severe relative to mild. All protein mean log2 fold changes between experimental groups and corresponding p values are located in Supplemental Table 1.

**Figure 1:**
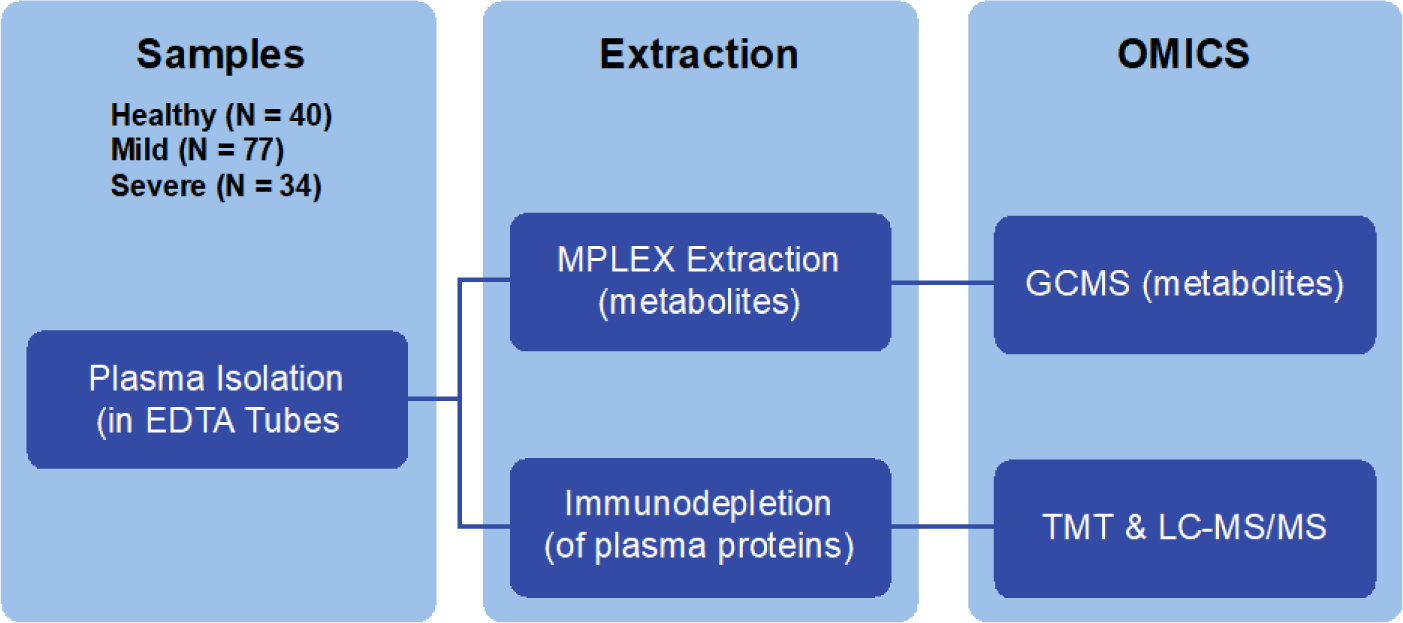
Multi-omics experimental design of COVID-19 plasma samples. (Left)-Plasma from Healthy (N=40), Mild (N=77), and Severe (N=34) disease patient samples were taken in EDTA-coated tubes. Plasma was subjected to MPLEx extraction to isolate metabolites or immunodepletion (to remove abundant plasma proteins). Following extraction metabolites were subjected to GC-MS analysis. Immunodepleted plasma proteins were TMT-labeled and analyzed by LC-MS/MS.

### Indicators of Pulmonary Distress and Neutrophil Activation in Severe Infections

Consistent with reported disruption of the alveolar-capillary barrier observed in ARDS patient samples and other COVID-19 plasma proteomics studies (5), we observed a statistically significant elevation in pulmonary surfactant B (PSPB) when comparing samples from severe infections to HC (adjusted p=3.92E-14) (Figure 2A&B). PSPB levels from mild patient samples were not statistically significant relative to HC (adjusted p=0.53). Consistent with previous reports (19), we also observed an increase in Surfactant Protein D in the severe relative to the mild infections (adjusted p=1.39E-05) and HC samples (adjusted p= 3.07E-05). The presence of pulmonary surfactant in the plasma of severe patient populations is consistent with an ARDS presentation with increased pulmonary capillary permeability (20).

**Figure 2:**
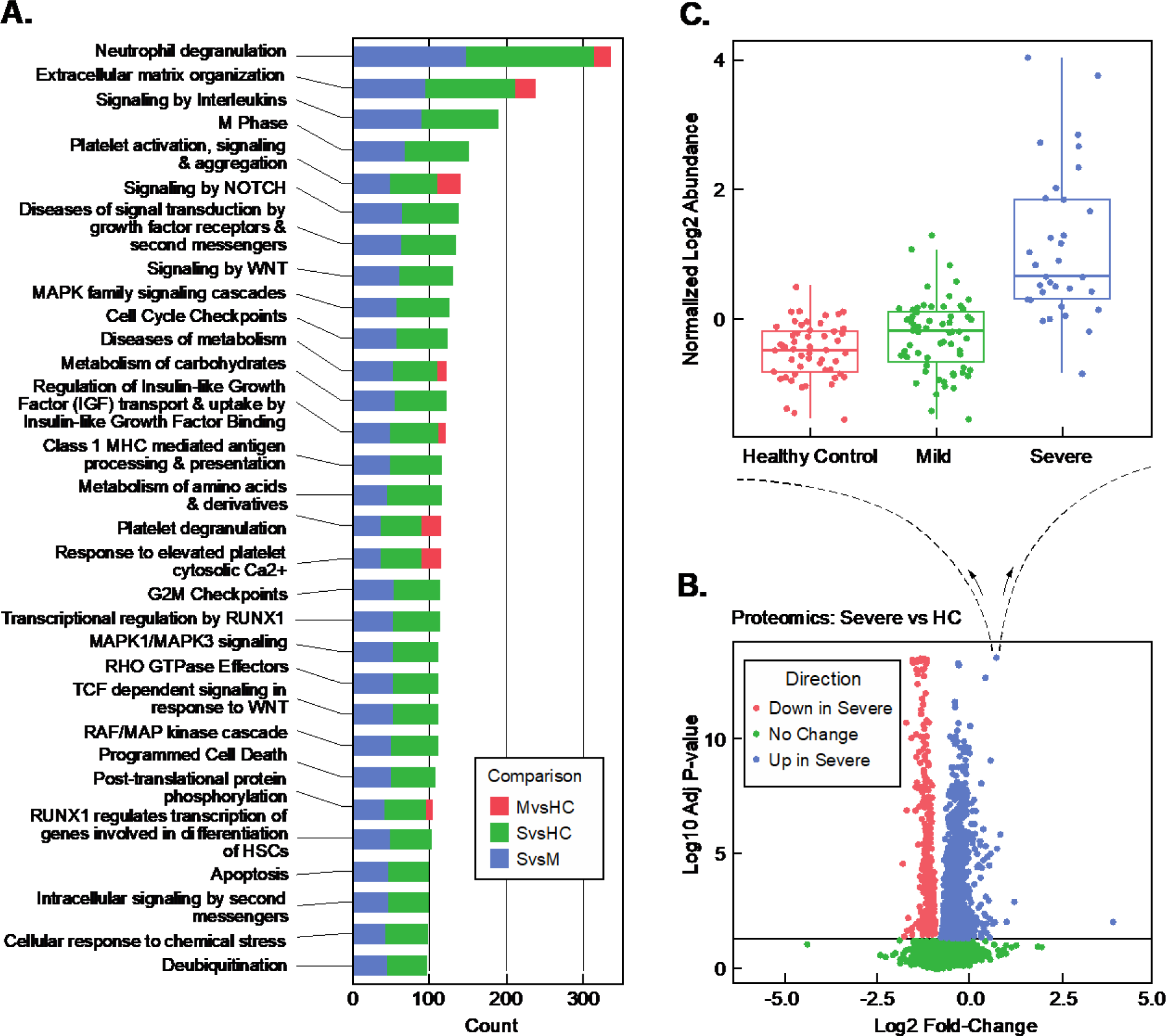
Indicators of Pulmonary Distress and Neutrophil Activation in Severe Infections. **(A)** Pathway analysis of plasma proteins changing between Mild (M), Severe (S) and Healthy controls (HC). **(B)** Volclano plot analysis of severe vs. healthy controls (HC). **(C)** Blow-up of individual, statistically-significant datapoint elevated in the Severe (S) samples showing a boxplot of the log2 abundance distribution of pulmonary surfactant B protein (PSPB) levels across all patients in the study.

A pathway enrichment analysis of proteins from samples derived from HC, mild, and severe cases strikingly showed the differential expression of proteins that mediate activation of neutrophil degranulation in mild and severe cases relative to HC samples (adjusted p<0.0001) (Figure 2C). In addition to differential regulation of neutrophil degranulation pathways, proteins involved in extracellular matrix organization (adjusted p<1.88E-09) were also elevated in COVID-19 patients relative to HCs. As expected, proteins involved in immune response pathways, platelet activation, and degranulation were also increased in COVID-19 patients, especially in severe cases. Other notable activated pathways included biological oxidation pathways that were significant in the severe cases when compared to HCs (adjusted p=0.0008) (Supplemental Table 2).

### Neutrophil Extracellular Traps in Severe and Mild Infections

Neutrophil extracellular traps (NET) are extracellular DNA-protein rich complexes released from neutrophils to capture pathogens during infection (14). Strong activation of both neutrophil degranulation and oxidation pathways is consistent with the reported physiological environment associated with triggering NET formation in viral infection and during free radical formation *in vitro* (13). NETs have been identified in COVID-19 lung tissue, sputum, and sera of infected patients (17, 21-25). Importantly, NETs occlude pulmonary vasculature in COVID-19 patients (26).

To characterize the proteome of NETs in our COVID-19 patients, we evaluated the mean relative levels and statistical significance of 18 NET protein components (27) including: DNA-binding proteins (Histones (H13, H4, H31T, H2B1C, and H2A2) and HMGNs (HMGN1, HMGN2, HMGN3, and HMGB1) Neutrophil collagenases (MMP8, MMP19, MMP14, and MMP9)), CAMP (Cathelicidin antimicrobial peptide), and Annexins (ANXA1, ANXA3, ANXA5, and ANXA6) in samples from HC, mild, and severe samples (Supplemental Figure 1). Only four NET proteins (MMP9 (adjusted p=0.002), CAMP (adjusted p=0.02), HMGN3 (adjusted p=0.04), and ANXA6 (adjusted p=0.04)) were elevated in the mild patient samples (Figure 3A) relative to HC. In contrast, all of the 18 NET complex proteins assessed were enriched in the severe relative to the mild patient samples (adjusted p<0.05) (Figure 3B) suggesting that NET complexes are more predominant in severe cases and/or are cleared more rapidly in mild cases.

**Figure 3:**
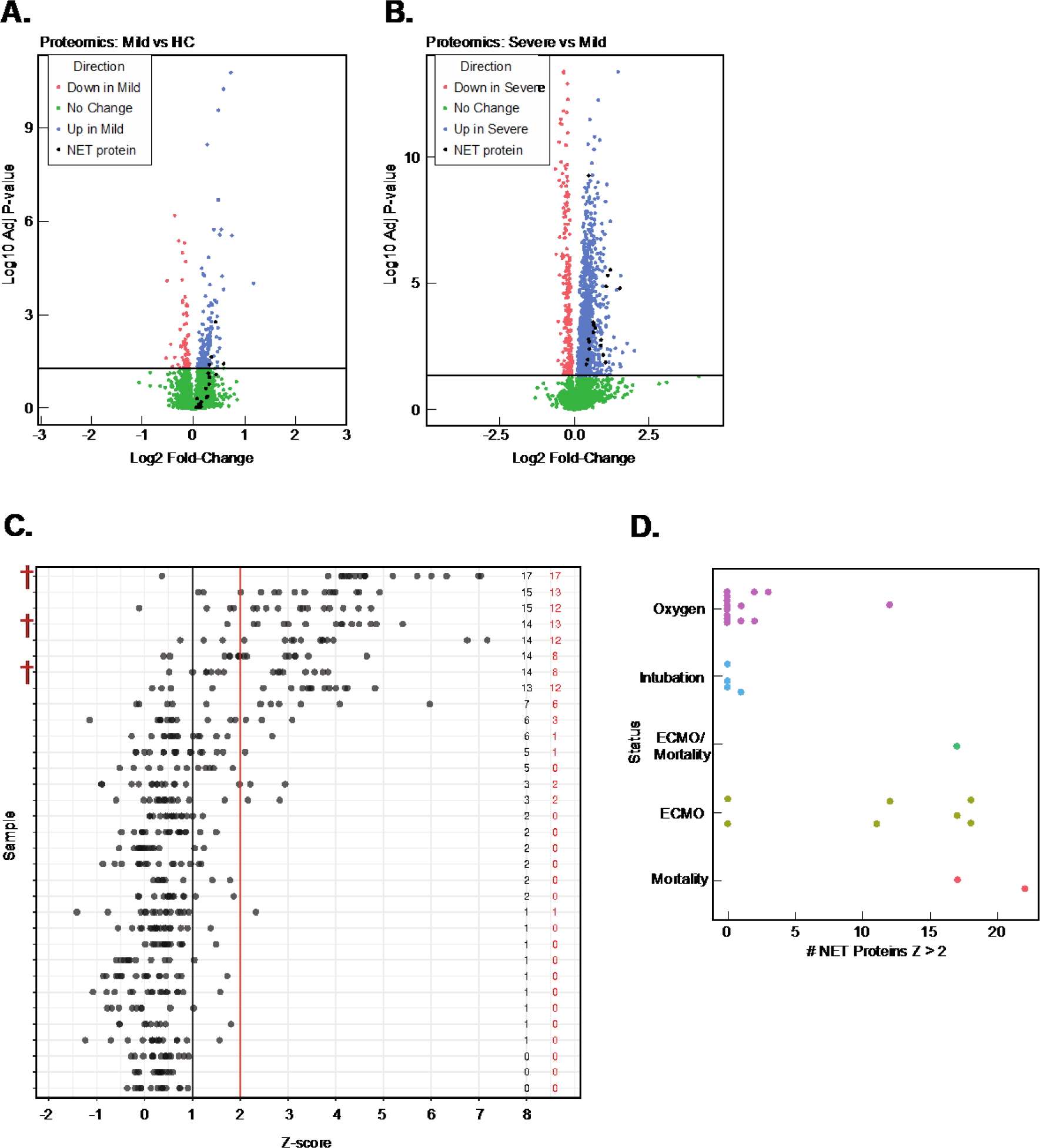
Neutrophil Extracellular Traps in Severe and Mild Infections. **(A)** Volcano plot analysis of mild (blue) vs. HC (red) plasma samples with the positions of (18) different NET protein components labeled in black. Proteins in green have a mean difference between mild and healthy controls (HC) with (P>0.05) **(B)** Volcano plot analysis of severe (blue) vs. mild (red) plasma samples with the positions of (18) different NET protein components labeled in black. Proteins in green have a mean difference between mild and severe with (P>0.05) **(C)** Analysis of NET protein z-score distributions among severe patient plasma samples. Numbers in red to the right indicate the number of NET proteins from the severe plasma sample with a z-score>2. Numbers to the right (in black) indicate the number of NET proteins from the individual severe plasma sample with a z-score >1. Individual NET protein z-score positions are shown as black dots. Severe patient mortalities are indicated with a red obelisk (ꝉ). **(D)** Distribution of NET protein complex z-scores by status of severe patients. Each dot on the string chart indicates an individual patient with the X position showing the number of proteins with a z-score > 2.

These results are consistent with recent reports of NETs in COVID-19 patient plasma (17, 28). We then investigated if NET proteins were both present and showed a statistically significant difference among individual patient samples. We addressed this question by evaluating the distribution of NET proteins from individual HC, mild, and severe patient samples by calculating z-scores relative to the HC mean abundance levels. NET proteins with a z-score >2 (see methods) were visualized in a string-plot from each patient to evaluate if: 1) Individual severe patient samples contributed more to the mean NET protein levels than others, 2) Multi-protein NET complexes could be observed from individual severe patent samples, 3) Randomly distributed NET proteins are observed across all severe patient samples. Importantly, an evaluation of NET protein z-score distributions showed: 1) That not all severe patients present detectable NET proteins in plasma (Figure 3C) and 2) A subset of the severe patients have detectable and significantly elevated levels of most of the NET proteins we monitored (Figure 3C). Importantly, these results suggest that NET formation is isolated to a subset of severe patients and several components of the NET complex are detectable among this subset of the severe patients. Additionally, our results indicate that a subset of mild patients have NET complexes with the number of detectable NET proteins and magnitude of NET protein z-scores considerably lower than that observed in severe cases (Supplemental Figure 2).

We next evaluated the individual characteristics of the severe patient population mean NET protein z-scores as a function of their respective clinical treatment or outcome (extracorporeal membrane oxygenation (ECMO), intubation, oxygen, and mortality). Patients that were either provided supplemental oxygen or intubated showed a clear trend towards lower mean NET protein z-scores compared to patients that eventually died in the hospital and/or were placed on ECMO treatment, which displayed higher mean NET protein z-scores (Figure 3C and Figure 3D). Only three patients died among the 151 patient cohort. Importantly, all three of the patients that died were among the severe group and were among those with the highest mean NET protein z-scores (Figure 3C). Consistent with age as a risk factor for severe disease (29, 30), nine of 33 total severe patients with the highest NET protein z-scores were 7.5 years older (mean value) than that of the 24 severe cases with lower mean NET protein z-scores (p=0.0124). Collectively, these results strongly suggest that formation of NETs is a marker of disease severity, age, and mortality during infection.

### Correlative Analysis of Mean NET z-scores, Pulmonary Surfactant, and other proteins

The precise role of NET complexes in severe COVID-19 infections is currently unknown but NETs have been reported in autopsies and suspected of contributing to increased pulmonary capillary permeability and pulmonary occlusions (26). Given the consistent observation of pulmonary surfactant B protein in COVID-19 patient plasma, we plotted the mean NET protein z-score in each patient against their relative log2 PSPB protein levels. Mean NET protein z-scores and PSPB levels were highly correlated with r= 0.70 (Supplemental Figure 3). We then performed a pan-correlative analysis of all proteins against each individual patient mean NET z-score. In addition to the previously mentioned correlations with PSPB and NET proteins, we observed strong correlations between mean NET z-scores and S100/calprotectin (S100A11) protein family members (27, 31, 32) and (r= 0.8-0.9) (Supplemental Figure 3). Since S100 proteins are established members of the NET complex (27, 32, 33), these results reinforce the validity of our correlative analysis but identify additional NET proteins beyond the original 18 NETs monitored in our survey. Importantly, our correlation analysis also identified the NET components myeloperoxidase (MPO) (r=0.823) and azurocidin (AZU1) (r=0.769). In addition to identifying other NET complex proteins, our correlative analysis also identified neutrophil cytosolic factor 1b (NCF-1B) (r=0.852), a subunit of NADPH oxidase, the primary source of reactive oxygen species in neutrophils (Supplemental Figure 3).

### Mild Infections Show Elevated Plasma Levels of Carnosine Dipeptidase Enzyme (CDNP1)

We next examined proteins specifically elevated in mild infections. We sought to determine if mild infections displayed signatures of neutrophil degranulation similar to severe infections. ISG-15, a ubiquitin-like protein, is a major component of neutrophil granules and is secreted in response to virus or IFN alpha-beta (34, 35). Consistent with the neutrophil degranulation pathway activation observed in severe COVID-19 samples (Figure 2C), we identified interferon stimulated gene 15 (ISG-15) elevated in mild samples relative to HC (adjusted p=1.86E-06) (Figure 4A&B). We then sought to identify candidate proteins among the mild infection samples that might highlight mechanistic differences between mild and severe disease. Analysis of a volcano plot between HCs and mild patient populations clearly showed multiple proteins elevated or decreased during a mild infection (Figure 4A). The carnosine dipeptidase 1 (CNDP1) was elevated in mild patients relative to HCs (adjusted p=6.12E-11) but was not statistically significant between HC and severe patient samples (Figure 4C) (adjusted p=0.621). This result was particularly interesting, as CNDP1 was among three proteins observed with increased differential expression in mild infection when comparing all three sample populations (HC, mild, and severe) among 2,766 total quantified proteins and might represent a compensatory pathway specific to mild patient populations. CNDP1 processes the antioxidant carnosine and is necessary for survival in animals upon oxidative stress challenge (36). The other two proteins with similar expression patterns (elevated in mild, but similar in HC and severe) were Trem-like transcript protein (TRML2) and Proto-oncogene tyrosine-protein kinase receptor (RET).

**Figure 4:**
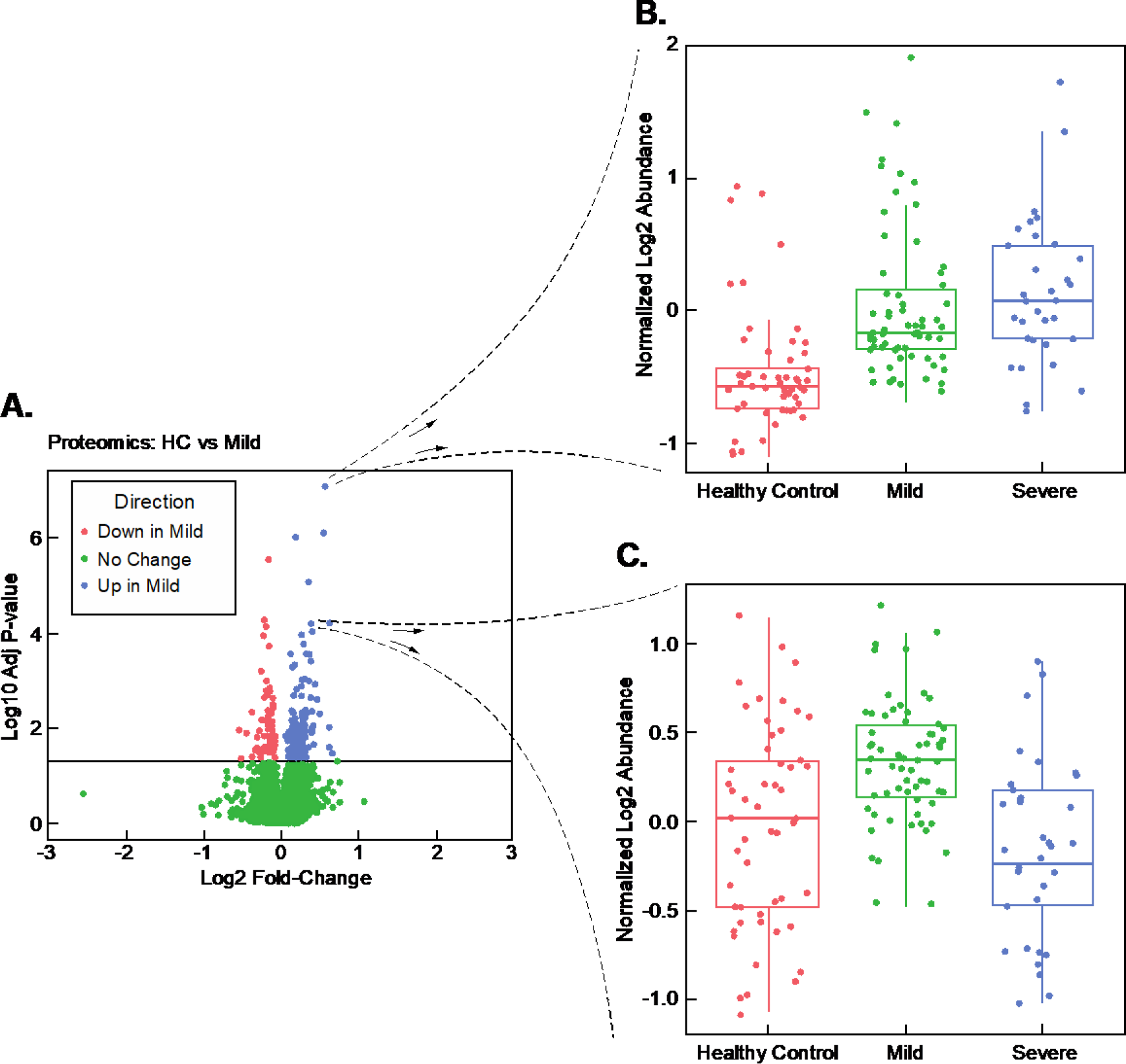
Mild Infections Show Elevated Plasma Levels of Carnosine Dipeptidase Enzyme (CDNP1). **(A)** Volcano plot analysis of Mild (M) vs. Healthy controls (HC) **(B)** Blow-up of individual, statistically-significant datapoint elevated in mild and severe samples showing a boxplot of the log2 abundance distribution of ISG-15 protein across all patients in the study. **(C)** Blow-up of individual, statistically-significant datapoint elevated in mild samples, but not severe samples, showing a boxplot of the log2 abundance distribution of the CNDP-1 protein across all patients in the study.

Our results suggested that surfactant protein levels might serve as a strong correlate of infection severity (Figure 2B). We reasoned that if CNDP1 was protective in the patient population with mild infection, then we might observe a negative correlation between lung surfactant levels (indicator of more severe disease phenotype) and CNDP1 (expression is potentially protective) in plasma. A comparison between CNDP1 and PSPB levels across all patient samples (HC, Mild, and Severe) showed a negative correlation (r= -0.335) (Supplemental Figure 3). When CNDP1 and PSPB levels were compared among the severe patient samples (alone) the negative correlation was much stronger with (r= -0.601) (Supplemental Figure 3). A comparison of CNDP1 and PSPB levels in HC and mild populations showed no significant correlation (HC r=-0.169, mild r= -0.151). Interestingly, a second, more promiscuous dipeptidase, cytosolic non-specific dipeptidase 2 (CNDP2, also known as carnosine dipeptidase 2), was also detected in our study and was elevated in the severe patient population relative to HC (adjusted p=0.001) but was not elevated in the mild infection relative to HC (adjusted p=0.659). CNDP2 and PSPB levels showed a positive correlation (r= 0.339) (Supplemental Figure 3). CNDP1 and CNDP2 share 54% amino acid sequence identity (Supplemental Figure 4). A manual investigation of peptide fragments quantified and identified from this study showed that the two proteins (CNDP1 and CNDP2) were unambiguously identified (Supplemental Figure 4).

While we observed a mean elevation in CDNP1 levels among mild infection samples, we found that some individual severe patients had CNDP1 levels matching or exceeding that of the mean CNDP1 levels in the mild infection group (Figure 4C). Given this result, we sought to evaluate a potential relationship between NET protein complex formation and CNDP1 levels among the patient sample groups. Therefore, we plotted mean NET protein z-scores observed in individual HC, Mild, and severe patients against their CNDP1 levels (assuming a linear model). In HC samples, the slope of CNDP1 levels vs. mean NET z-scores was positive (slope=0.463, p=0.036). In contrast, in mild and severe patient samples, the slope of CNDP1 levels vs. mean NET z-scores was negative (mild disease slope= -0.137, p=0.016, severe disease slope= -0.188, p=0.004) (Supplemental Figure 3). We also observed a positive correlation between mean NET protein z-scores and CNDP2 levels (r=0.59). There was no significant correlation between CNDP1 and CNDP2 levels (r=-0.06). Both CNDP1 and CNDP2 play essential roles in modifying potent antioxidants. These results highlight the intriguing possibility that each of the CNDP dipeptidases might have a different mechanistic role in suppression of NET protein complexes.

### L-cystine and Anti-oxidant Enzyme Elevation in COVID-19 Patient Plasma

In addition to analyzing protein levels between the previously mentioned patient populations, we quantified metabolite levels using gas chromatography-mass spectrometry (GC-MS)-based metabolomics analysis. In total, we quantified 205 metabolites across all patient samples (Supplemental Table 3). Comparing mild infection to HC showed a statistically significant difference in 22 metabolites (adjusted p<0.05) with 8 metabolites elevated in mild infection and 14 metabolites reduced. In contrast, a statistically significant difference among 71 total metabolites, with detection of higher quantities of 24 metabolites and decreased levels of 47 metabolites, was identified when severe infection plasma samples were compared to HC (adjusted p<0.05) (Figure 5A). Comparing severe vs. mild infections showed a difference in 56 metabolites with an elevation of 18 metabolites in severe relative to mild infection and an elevation of 38 metabolites in mild infection plasma relative to severe (adjusted p<0.05). A common elevated metabolite observed in both the mild and severe infection patient plasma with a mean fold-change of 23.75 (adjusted p<1E-15) between mild infection and HC and 60-fold mean increase (adjusted p<1E-15) in severe infection plasma relative to HC was L-cystine, the disulfide-bonded form of the amino acid cysteine (Figure 5A&5B). Cystine levels were 2.8-fold higher in severe vs. mild infection patient plasma (adjusted p=5.2E-07). L-cysteine (reduced L-cystine) levels were elevated in both mild (p=0.003) and severe (p=0.0004) relative to HC (Supplemental Table 3).

**Figure 5:**
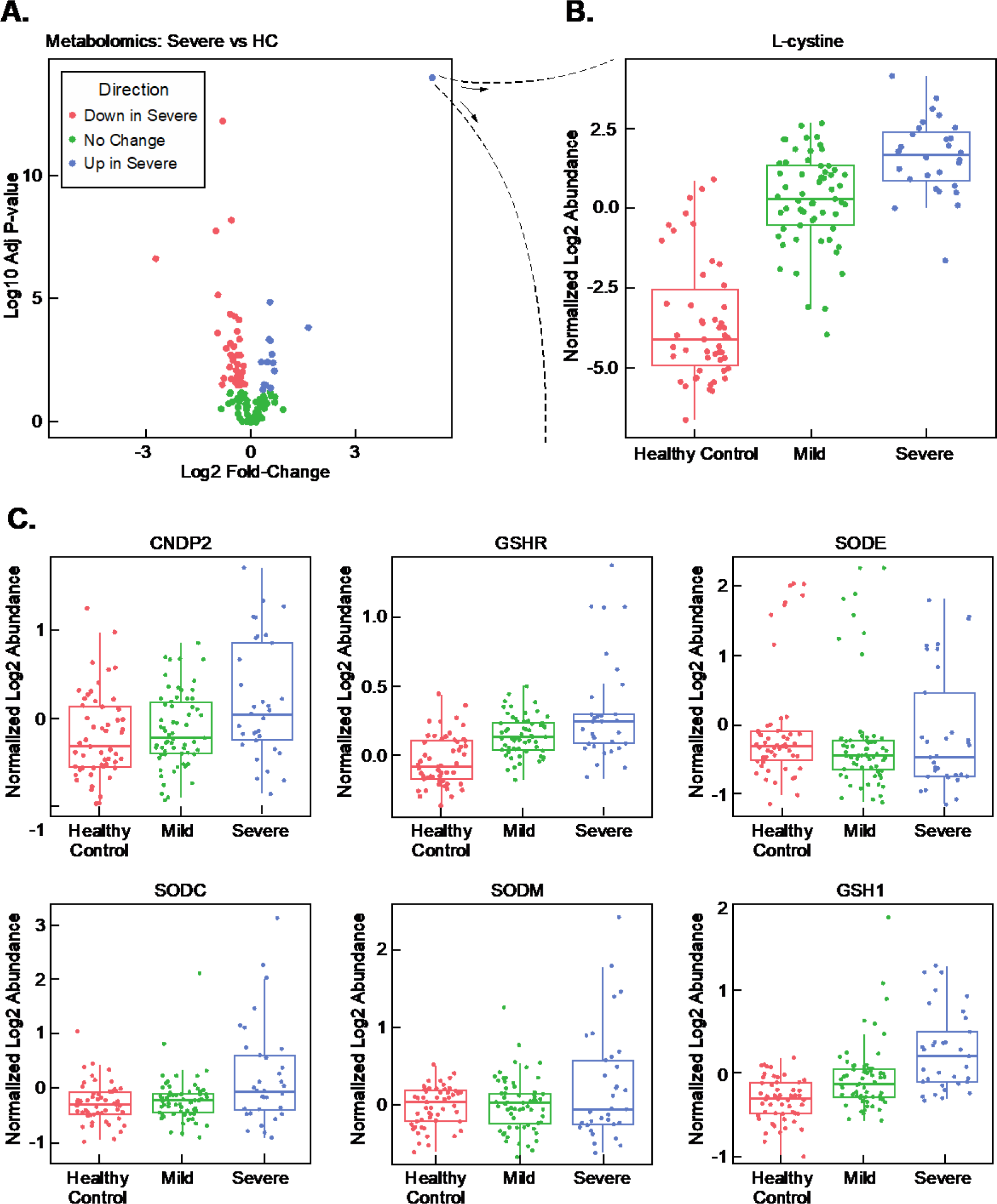
L-cystine and Anti-oxidant Enzyme Elevation in COVID-19 Patient Plasma. **(A)** Volcano plot analysis of severe vs. healthy controls (HC) metabolites **(B)** Blow-up of individual, statistically-significant mean datapoint from the volcano plot showing the boxplot distribution of normalized log2 abundance of L-cystine (disulfide bonded form of cysteine). **(C)** Boxplot analysis of protein abundance between HC, mild, and severe samples of antioxidation response proteins CNDP2, GSHR, SODE, SODC, SODM, and GSH1 levels.

NET formation is suppressed by antioxidants *in vitro* and *in vivo* (37, 38). Glutathione is the cell’s most potent antioxidant. L-cystine forms in the extracellular space from reduced glutathione. Upon transport into cells, L-cystine is an eventual substrate for glutathione synthesis. As increased levels of NET proteins were detected in a sub-population of our severe infection patient samples, we worked to evaluate relationships between mean NET protein z-scores observed in the severe samples and L-cystine log2-fold abundance relative to HCs. Interestingly, we observed that mean NET protein z-scores were correlated to L-cystine levels (r =0.40) (Supplemental Figure 3).

Reduced glutathione is eventually converted to L-cystine. Hence L-cystine might serve as an indicator of glutathione production and cycling during conditions of oxidative stress. Because we observed NET proteins in the severe infection population and L-cystine elevation among all COVID-19 patient populations, we analyzed the levels of proteins with key roles in the Cystine-Glutathione metabolic cycle and other enzymes with antioxidant activities-including those with roles in superoxide to peroxide conversion. Consistent with a cellular response to oxidative stress, we observed a statistically significant increase in enzymes critical for glutathione synthesis including glutathione S-reductase (GSHR) (Mild vs. HC adjusted p=3.73E-09, Severe vs. HC adjusted p=4.53E-14) and glutamate-cysteine ligase (GSH1) (Mild vs. HC adjusted p=1E-4, Severe vs. HC p=2E-11) (Figure 5C). Cellular SOD, which converts superoxide to peroxide, was elevated in severe infection samples relative to HC (Figure 5C) (adjusted p=0.0002). In contrast to cellular SOD (SODC) levels, mitochondrial SOD (SODM) and extracellular SOD (SODE) were unchanged relative to HCs (Figure 5C) (adjusted p>0.05).

## Discussion

Severe COVID-19 patients can develop acute respiratory distress syndrome (ARDS) which is characterized by hypoxemia and lung stiffness often related to increased permeability of the alveolar-capillary barrier, and an influx of neutrophils into the pulmonary space (1). Mouse models of ARDS recapitulate similar aspects of severe Coronavirus disease (38, 39). Multiple studies using bacterial-induced ARDS display evidence of increased microcapillary permeability (38, 39). Disruption of the alveolar-capillary barrier likely leads to both infiltration of leukocytes from the capillaries to the pulmonary space and exchange of alveolar contents to the blood. Consistent with this observation, a multi-omics analysis of plasma conducted prior to the study here found pulmonary surfactant B in SARS-CoV-2 patient plasma (5). In addition, ELISA-based analysis of SARS-CoV-2 patients has shown an increase in pulmonary surfactant D (40, 41). Taken together with results shown in our current study, these studies show that patients with severe SARS-CoV-2-induced ARDS exhibit an increase in surfactant B and surfactant D levels in severe infections. These results suggest that plasma surfactant B and D levels are a logical marker of ARDS indicating increased permeability of the alveolar-capillary barrier.

Bacteria-induced mouse models of ARDS show NET formation in the pulmonary capillaries (39). Similar to these ARDS models, general comparisons of SARS-CoV-2 patient populations vs. healthy controls have reported detection of NET components including DNA (17), myeloperoxidase (detected by ELISA), and histones (detected by immunofluorescence). In addition, plasma proteomics studies of SARS-CoV-2 and healthy control populations have reported elevated NET proteins in SARS-CoV-2 patients (5). Our study demonstrates that the most abundant NETs are found in a sub-population of severe SARS-CoV-2 infections who have the worst clinical outcomes. Importantly, our results also show that individuals with severe ARDS display multiple, detectable NET proteins. Patients with multiple elevated NETs not only had the worst infections but included all fatalities in the study and were older than the severe patients with reduced NET protein levels. Consistent with limited disease, few of the mild infection patients in our study showed elevated NET protein levels compared to healthy controls. However, a sub-population of the mild infection samples showed NET proteins with elevated z-scores. More than one NET protein was observed in these mild patient plasma samples, suggesting that significant NET formation can initiate in mild infections without an immediate requirement for hospitalization or displaying immediate indications of ARDS. That mild patient populations showed a clear initiation of NET formation is important and raises critical questions about NET formation during post-infection sequelae, breakthrough infections in vaccinated populations, and infection with SARS-CoV-2 variants of concern.

NETs are found in pulmonary capillary occlusions in mouse models of ARDS (39) and autopsies of coronavirus-induced ARDS (42). Occlusions and endothelial cell disruption within the pulmonary capillaries are hypothesized as a possible mechanism to explain the increased permeability observed in ARDS (43). In our study, comparison of mean NET protein z-scores to pulmonary surfactant B levels showed a strong correlation consistent with a direct relationship, suggesting that as NET formation increases, pulmonary capillary permeability increases. These results support the model of NETs driving permeability, potentially through the formation of capillary occlusions. In addition to significant correlations between the NET proteins and PSPB, we identified significant correlations between mean NET z-scores and 389 other proteins (r>0.6) (Supplemental Table 4). Among the other significant correlations were multiple established NET proteins characterized from other studies. We anticipate that within our correlative analysis to NETs there are other proteins that might stimulate NET formation. In addition, our correlative analysis likely reveals processes that occur after or during NET formation, including the activation of other leukocyte populations.

Recent studies from SARS-CoV-2 patient neutrophils show elevated release of reactive oxygen species (44). Free radicals and oxidative stress pathways trigger NET formation. One of the primary sources of ROS in neutrophils is the NADPH oxidase enzyme (45-49). We observed a clear correlation between mean NET z-protein scores and NAPDH protein levels in our study. In addition to activation of oxidation pathways driving NET formation, we observed a substantial elevation in antioxidant factors that might dampen NET formation. Specifically, we found that the glutathione substrate L-cystine was elevated in both the mild and severe infection patient samples. Activated neutrophils import L-cystine to produce glutathione (50). Importantly, our proteomics experiments showed a complementary elevation of enzymes critical for glutathione metabolism including GSHR (glutathione reductase), GSH1 (Glutamate-cysteine ligase), and CNDP2 (promiscuous dipeptidase) in the severe infection patients, with an elevation of GSHR and GSH1 in the mild infection patients. Taken together, the most logical interpretation of these results is that glutathione synthesis is elevated in the blood plasma of all SARS-COV-2 patient populations and this likely represents a mechanism to suppress over-activation of neutrophils (and NET formation), and other harmful processes triggered by oxidation (Figure 6).

**Figure 6:**
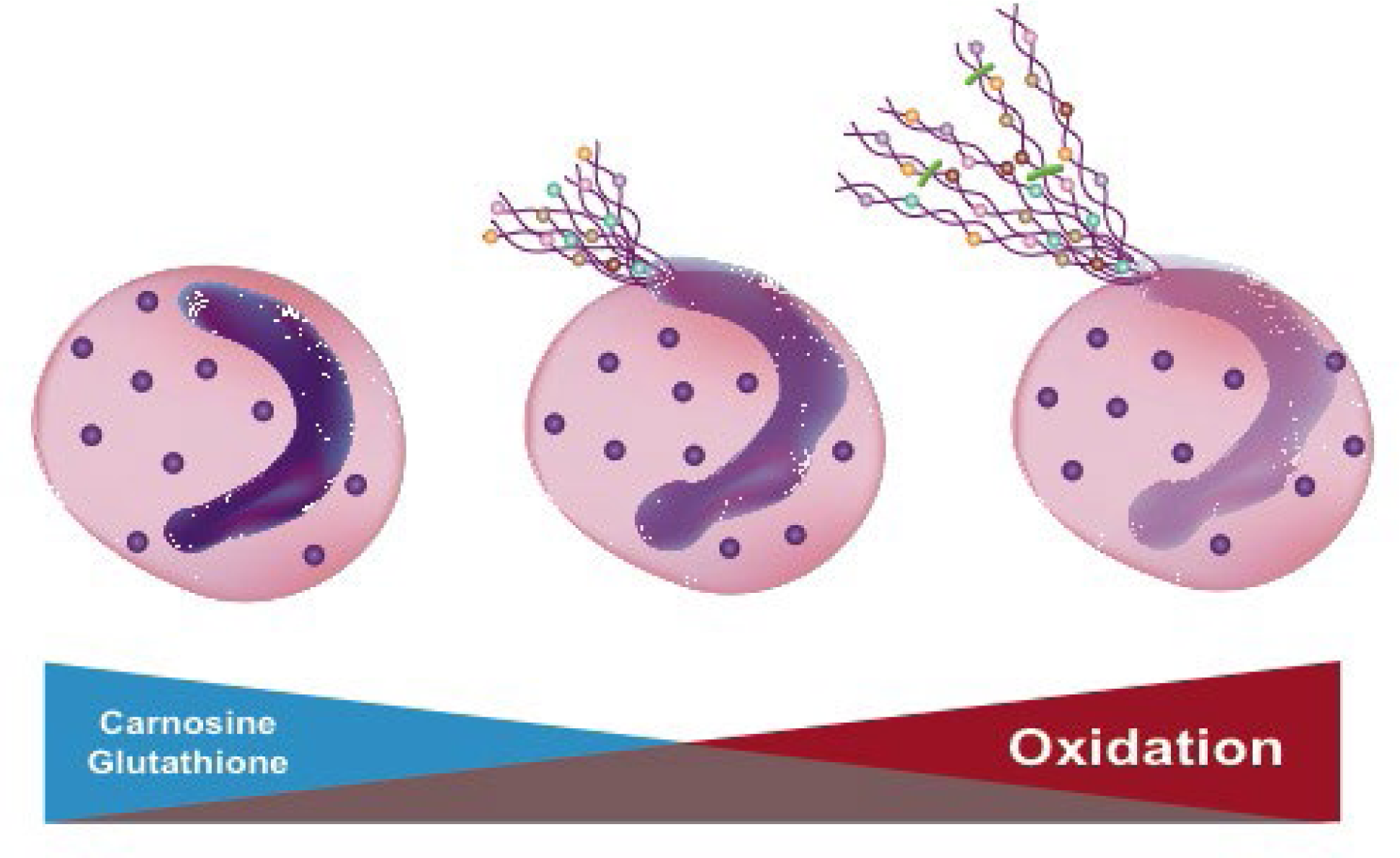
Hypothesis of NET formation in response to increased oxidation. (Left)-Healthy Neutrophil. (Center)-Neutrophil initiating NET formation. (Right)-Neutrophil with fully-formed NETs and elevated oxidative stress during COVID-19 infection.

In addition to observing multiple, elevated signatures of glutathione synthesis in our studies, we observed an elevation of the carnosinase CNDP1 in mild infection samples. Unlike CNDP2, CNDP1 lacks a reported activity against Cys-Gly dipeptides in glutathione metabolism and instead, specifically digests the antioxidant dipeptide carnosine (alanine-histidine) (51). Hence, CNDP1 activity is unlikely to accelerate glutathione synthesis. Instead, CNDP1’s narrow metabolic activity targeting carnosine in mild infection patients is likely to drive an independent antioxidant pathway working separately from the multiple enzymes observed driving glutathione synthesis. The substrate of CNDP1 (carnosine) has strong antioxidant activities (52-54). Reactivity of carnosine with harmful, chemical species typically occurs within Carnosine’s amine groups (distant from the peptide bond). Hence, it is possible that plasma CNDP1 carnosinase cleaves both modified carnosine derivative metabolites and non-oxidized forms. It is currently unclear how carnosinase activity would have a direct role in reducing oxidation and NET formation. One possibility is that cleavage of carnosine metabolites at the peptidyl bond facilitates excretion of adducted carnosine metabolites into the urine. Additionally, cleavage of carnosine into histidine and alanine metabolites could result in re-formation of carnosine through carnosine synthetase for subsequent rounds of antioxidant activity.

The unique upregulation of CNDP1 in mild, but not severe infection patients raises the possibility that CNDP1 has a protective role in patients with mild infections. In support of this possibility, our results clearly show that CNDP1 levels are negatively correlated with PSPB levels and, in both mild and severe infection populations, as CNDP1 levels increase, mean NET protein z-scores decrease. To our knowledge, CNDP1 would be one of the first candidate enzymes to regulate NET formation in the context of infection. Animals with targeted deletions in CNDP1 display a shortened lifespan and have increased sensitivity to oxidative stress (36). Importantly, administration of carnosine suppresses NET formation and capillary permeability in a mouse model of ARDS induced by bacteria (38) and suppresses capillary permeability, lung pathology, and neutrophil myeloperoxidase activity in mice infected with H9N2 influenza (55). Taken together, these studies and ours highlight the possibility of CNDP1 up-regulation as part of a protective mechanism distinct to individuals with mild infections to suppress oxidative stress and NET formation.

## METHODS

### Lead Contact

Further requests for resources and reagents should be directed to Vincent R. Gerbasi.

### Materials availability

This study did not generate any unique reagents.

### Data and Code Availability

All mass spectrometry datasets generated during this study have been deposited at the Mass Spectrometry Interactive Virtual Environment (MassIVE) under code XXXX.

### Experimental Model and Subject Details

Study design and participation were approved by the University of Wisconsin Health Sciences review board under IRB protocol # 2015-0802-CR006. Consent was obtained for all participants prior to enrolment. Plasma samples were collected from (n=40) HC patients within similar age ranges to patients with mild or severe SARS-CoV-2 infections. Study participants with infections were identified by a positive RT-PCR-based test (developed in-house) for SARS-CoV-2 collected by nasal swab. Mild infections (n=77) were characterized as patients with a positive test but not requiring supplemental oxygen or other significant support. Severe infections were characterized as those individuals requiring supplemental oxygen, ECMO, intubation, or mortality (n=34).

### Methods Details

#### Analyte Extraction

Plasma samples were isolated in EDTA-coated tubes and were subjected to both proteomic analysis preparation including immunodepletion, trypsinization, TMT-labeling, and basic-RP-fractionation and metabolomic analysis preparation using the <u>M</u>etabolite, <u>P</u>rotein, <u>L</u>ipid <u>Ex</u>traction method, essentially described previously (56, 57). Briefly, plasma samples were prepared and batched in a random order placing National Institute of Standards and Technology (NIST) Standard Reference Material (SRM) for Human Plasma (SRM 1950) (NIST Office of Reference Materials, Gaithersburg, MD) aliquots at the beginning and end of each batch (58). Plasma samples were thawed in a biosafety cabinet (BSC) and 110 µL of plasma was aliquoted into chloroform compatible microfuge tubes (Sorenson Biosciences, Salt Lake City, UT) along with 190 µl of a metabolite internal standard (in water) prepared as previously described (59-61) containing Leucine-13C6 (4 µg/mL), Creatine-D3 H2O (methyl-D3) (4 µg/mL), L-Leucine-D10 (4 µg/mL), L-Tryptophan-2,3,3-D3 (40 µg/mL), L-Tyrosine Ring-13C6 (4 µg/mL), L-Phenylalanine Ring-13C6 (4 µg/mL), N-BOC-L-tert-Leucine (4 µg/mL), N-BOC-L-Aspartic Acid (4 µg/mL), Propionic Acid 13C3 (8 µg/uL), Succinic Acid-2,2,3,3-d4 (4 µg/mL), Salicylic Acid D6 (4 ug/mL), Caffeine-d3 (1-methyl-d3) (4 µg/mL). 1190 µL of pre-made, cold chloroform/methanol mix (2:0.975 chloroform/methanol (v/v)) was added to each sample and NIST plasma along with 10 µl of SPLASH lipid standard in methanol (AVANTI polar lipids, Birmingham, AL) to make a final ratio of 3:4:8 H2O:methanol:chloroform (v/v). Viruses and other pathogens were inactivated at this point (62). Samples were vortexed and incubated -20°C for 10 mins and centrifuged at 10k x g for 10 mins to separate the analytes. The upper polar phase containing metabolites was collected into glass vials along with 200 µl of the lower non-polar phase (by carefully breaking through the protein interlayer). The remaining non-polar phase was removed and 500 µl of cold methanol was added to the tube containing the protein. The samples were vortexed and centrifuged again to pellet the protein and the remaining supernatant was added to the metabolites and dried *in vacuo* and stored at -20°C until derivatization and GC-MS analysis.

#### Metabolomic analysis

The extracted metabolites from MPLEx were chemically derivatized based on the method reported previously (62). To protect carbonyl groups and reduce the number of tautomeric isomers, methoxyamine (20 µL of a 30 mg/mL stock in anhydrous pyridine) was added to each sample, followed by incubation at 37°C with shaking for 90 min. To derivatize hydroxyl and amine groups to trimethylsilylated (TMS) forms, N-methyl-N-(trimethylsilyl)trifluoroacetamide with 1% trimethylchlorosilane (80 µL) was added to each vial, followed by incubation at 37°C with shaking for 30 min. The derivatized samples were analyzed by gas chromatography-mass spectrometry (GC-MS; Agilent 7890A GC with 5975 MSD single quadrupole; Agilent Technologies, Santa Clara, CA). Separation of metabolites were done by HP-5MS GC column (30 m × 0.25 mm × 0.25 µm; Agilent Technologies) and 1 µL of samples was injected in splitless mode. The helium flowrate was adjusted by retention time locking function, and the injection port temperature was held at 250°C throughout the analysis. The GC oven was held at 60°C for 1 min after injection, and the temperature was then increased to 325°C by 10°C/min, followed by a 10 min hold at 325°C. Data were collected over the mass range 50-600 m/z. A mixture of FAMEs (C8-C28) was analyzed together with the samples for retention index alignment purposes during subsequent data analysis. GC-MS data files were converted to CDF format and they are deconvoluted and aligned by Metabolite Detector (63). Identification of metabolites was done by matching with PNNL metabolomics databases – augmented version of Agilent Fiehn metabolomics database (64). The database contains mass spectra and retention index information of over 1,000 authentic chemical standards and they were cross-checked with commercial GC-MS databases such as NIST20 spectral library and Wiley 11th version GC-MS databases (65, 66). Three unique fragmented ions were selected and used to integrate peak area values, and a few metabolites were curated manually, when necessary.

#### Plasma Protein Processing, TMT-labeling, and Mass spectrometry

Major plasma proteins were immunodepleted using a Multiple Affinity Removal System (MARS) column (Hu-14 4.6 x 100 mm, Agilent Technologies, Santa Clara, CA) column in-line with an Agilent 1200 HPLC system and analyzed as described previously (67). Flow-through fractions were collected and concentrated from the depletion column as described previously (57). Depleted proteins were quantified using the BCA Protein Assay and denatured in 8M Urea, reduced in DTT, Alkylated in Iodoacetamide, and digested with trypsin. Following trypsinization, peptides were subjected to 16-plex TMT-labeling with an internal control sample and blank in each block of 14 samples analyzed. TMT-labeling reactions were quenched and desalted over C18 cartridges. Samples were fractionated into 96 fractions by basic reverse phase (68) and concatenated into 24 fractions by smart pooling to increase the depth of peptide identification. Fractions were concentrated and frozen prior to LC-MS analysis.

Immunodepleted, TMT-labeled peptide sample fractions were randomized and analyzed on a NanoAcuity UPLC system interfaced with a Thermofisher Q-Exactive HFX Orbital trap mass spectrometer (Thermofisher). Separation was carried out in a C18 revered-phase column (70 cm × 75 μm i.d., Phenomenex Jupiter, 3 μm particle size, 300 Å pore size) for 152 minutes. For detailed in gradient information and mass spectrometer settings please refer to previous work (57). Briefly, precursor ions were analyzed between 300-1800 Thomson, at 60K resolution, an automatic gain control (AGC) of 3E6, and an injection time of 20ms. The top-12 most abundant precursor ions were fragmented by HCD (NCE setting=30) with a quadrupole isolation width of 0.7 Thomson, employing 30K resolution, an AGC target of 1E5 ions. Dynamic exclusion was enabled for 30 seconds. Xcalibur spectral RAW files were acquired in centroid mode. Protein identifications were made by searching the SwissProt version of Uniprot Human Database (Release November 2019) using MS-GF+ combined to mzRefinery to recalibrate the mass spectra (69). The identification parameters included (a) parent ion mass tolerance of ±6 ppm, (b) at least partial tryptic digestion with 2 missed cleaved sites allowed, (c) static modification of cysteine carbamidomethylation (+57.0215 Da) and N-terminal/lysine TMT labeling (+229.1629 Da), and (d) variable modifications of methionine, cysteine, tyrosine and tryptophan oxidation (+15.9949 Da); cysteine dioxidation (+31.9898 Da); and asparagine, glutamine and arginine deamidation/deamination (+ 0.98402 Da). Peptide-spectrum matches, peptides and proteins were filtered based on MS-GF probabilities to a false-discovery rate of <1%.

Quantitative information of the TMT reporter ion intensities was extracted using MASIC (70). The median intensity of each peptide abundance for each sample was scaled to the combined internal standard TMT channel consisting of an equal concentration of plasma peptides from all patient samples as described previously (7). Quantitation and statistical analysis of TMT results to generate graphs and plots utilized an in-house R-studio package (described below).

### Quantification and Statistical Analysis

All data preprocessing was done in R version 4.1.0 with the *pmartR* package (71). Proteomics data was normalized to the reference sample by computing the ratio of each sample’s peptide abundances to the respective combined internal standard TMT channel calculated within each plex. Peptide-level data and metabolomics data were log2 transformed and filtered to remove peptides and metabolites without enough observations to conduct at least one statistical comparison of interest (72). One sample from the mild and one sample from the severe infection group was removed from the proteomics dataset and five samples from the metabolomics dataset (three from mild, one HC, one severe) as outliers based on a robust Mahalanobis distance with a p-value threshold of 0.0001 (73) and visual inspection of principal component analysis and correlation with other samples. For the proteomics data, samples were normalized by the median of rank-invariant peptides at a p-value of 0.15, the method selected by the statistical procedure for analysis of proteomics normalization strategies (SPANS) algorithm for determining the optimal normalization method (74). Data was then quantified to the protein-level using a median reference-based approach (75). The metabolomics data was normalized using the median of all metabolites for each sample.

Statistical comparisons between the HC, mild, and severe infection groups were conducted for each protein and metabolite by fitting an analysis of variance (ANOVA) model with post-hoc pairwise comparisons, and a Benjamini-Hochberg multiple test correction (76) to adjust ANOVA p-values. Correlation values were calculated using Pearson’s correlation. For each NET protein, the z-score was calculated as z-score = (log2 abundance – mean of log2 HC abundances)/(pooled standard deviation) for each sample. The interpretation of the z-score value is the number of standard deviations away from the mean of the HC group. Statistically, any protein or metabolite with a z-score > 1 is in the top 16% of values, and a z-score > 2 corresponds with approximately the top 2.5% of observations. Pathway enrichment analysis was conducted using the *ReactomePA* R package (77) and statistically significant proteins for each pairwise comparison (adjusted p<0.05). Significant pathways were identified and prioritized based on adjusted p-values and associated gene counts. Statistical significance of age differences between the 9-highest severe patient NET protein z-scores and those 24 severe patients with lower NET protein z-scores were evaluated using a non-parametric kruskal-wallis test. All visualizations were generated using the *ggplot2* (78) and *trelliscopejs* R packages.

## Data Availability

All data produced in the present work are contained in the manuscript.

## Acknowledgements

We thank Dr. Timothy J. Garrett at the University of Florida for sharing metabolite standard formulations. We thank Michael Perkins for assistance with generating figures. We thank Eisuke Adachi, Makoto Saito, Hiroyuki Nagai, and Hiroshi Yotsuyanagi at Institute of Medical Science of the University of Tokyo, Takayuki Ogura at Saiseikai Utsunomiya Hospital, Naoki Hasegawa, Tomoyasu Nishimura, and Katsunori Masaki at Keio University School of Medicine, Takahide Kikuchi, Daisuke Taniyama, and Kazuto Itoh at Saiseikai Central Hospital, and Shuichi Yoshida and Isano Hase at Saitama City Hospital for assistance with preparing clinical samples. This research was supported by Japan Program for Infectious diseases Research and Infrastructure (JP21wm0125002) and Research Program on Emerging and Re-emerging Infectious Diseases JP19fk0108113 from the Japan Agency for Medical Research and Development (AMED). VRG was supported by the Biomedical Resilience and Readiness in Adverse Operating Environment Initiative at PNNL. This work was supported by the PNNL Directorate Objective Biorisk Beyond the List. Sample testing and data analysis was funded by the U.S. Department of Defense’s Joint Program Executive Office (JPEO) COVID-19 FY20/21 CARES Act to U.S. Naval Medical Research Unit TWO (NAMRU-2) via Naval Medical Research Center (Appropriation: 9700130, JON: SW713). These funds were transferred via two Military Interagency Purchase Requests (MIPRs) from NAMRU-2 to PNNL (MIPRN6281420MPNX005, MIPRN6281421MPNX003) under a Memorandum of Agreement between the U.S. Departments of Defense and Energy (MOA; USA000707-13-DPAP dtd 01 May 2013).

The views expressed in this article reflect the results of research conducted by the authors and do not necessarily reflect the official policy or position of the Department of the Navy, Department of Defense, nor the United States Government. RDH, KC, and AGL are military Service member or employees of the United States Government. This work was prepared as part of their official duties. Title 17, U.S.C., §105 provides that ‘copyright protection under this title is not available for any work of the United States Government.’ Title 17, U.S.C., §101 defines a U.S. Government work as a work prepared by a military service member or employee of the U.S. Government as part of that person’s official duties. The study protocol was approved by the University of Wisconsin Health Sciences Institutional Review Board (IRB protocol # 2015-0802-CR006) in compliance with all applicable federal regulations governing the protection of human subjects.

## Author contributions

VRG: Designed experiments, analyzed results, wrote manuscript draft

LMB: Conducted statistical and pathway analyses, created visualizations, wrote manuscript draft

CDN: Prepared metabolite samples, designed experiments, contributed to manuscript draft

YMK: Analyzed results, designed experiments, contributed to manuscript

ACS: Designed experiments, analyzed results, contributed to manuscript draft TOM: Designed experiments, analyzed results, contributed to manuscript draft

RDH: Project conceptualization, administration and funding support, reviewed results, contributed to manuscript draft

KGS: Contributed to statistical analysis

MAG: Designed experiments, performed proteomics experiments

VLP: Analyzed metabolite samples

ESN: Designed experiments, analyzed results

AGL: Administrative oversight, reviewed manuscript

KMW: Designed experiments, Analyzed results

AAS: Sample Analysis

IKA: Sample Analysis

KC: Project conceptualization, administration and funding support, reviewed results, contributed to manuscript draft

YK: Conceptualization, funding acquisition, resources, supervision, writing

AJE: Conceptualization, project administration, writing

OA: Contributed clinical samples, provided clinical data

MK: Contributed clinical samples, provided clinical data

TT: Contributed clinical samples, provided clinical data

MK: Contributed clinical samples, provided clinical data

IN: Contributed clinical samples, provided clinical data

RB: Contributed clinical samples, provided clinical data

HT: Contributed clinical samples, provided clinical data

SS: Contributed clinical samples, provided clinical data

HN: Contributed clinical samples, provided clinical data

HK: Contributed clinical samples, provided clinical data

KI: Contributed clinical samples, provided clinical data

## Supplemental Information Legends

**Supplemental Figure 1:** Boxplot analysis of 18 different NET complex proteins in HC, Mild, and Severe plasma samples. (Attached in separate file)

**Supplemental Figure 2:** NET protein string plots of Severe (Top), Mild (Middle), and HC (Bottom). The vertical red line indicates the point where z-score >2. Each horizontal string line indicates an individual patient plasma sample. Uniprot identification numbers for each NET protein are labeled with different colors on the right. (Attached in separate file)

**Supplemental Figure 3:** Correlation analysis (A) mean NET z-score vs. PSPB (B) mean NET z-score vs. S100A11 protein (C) mean NET z-score vs. NCF-1B (D) mean NET z-score plotted against CNDP1 in HC (top), mild (center), and severe (bottom) (E) PSPB vs. CNDP1, (F) PSPB vs. CNDP2 (G) CNDP2 vs. CNDP1 (H) mean NET z-score vs. L-cystine. (Attached in separate file)

**Supplemental Figure 4:** Comparison of CNDP1 and CNDP2 mapping peptides identified from plasma. Bold (top, black) amino acid position indicate peptides used to identify CNDP1 (Top). Bold (bottom, red) peptides indicate positions within the protein used to identify CNDP2. (Attached in separate file)

**Supplemental Table I:** Quantitative Proteomics statistical analysis for all detected proteins from plasma. Column A: Uniprot Protein Identifier, Column B: Number of peptides detected from the protein in the first column (N_Peptides), Column C: number of Healthy control (N_HC) patient samples that detected the protein, Column D: number of Mild (N_Mild) patient samples that detected the protein, Column E: number of Severe (N_Severe) patient samples that detected the protein, Column F: Mean healthy control protein abundance (Mean_HC), Column G: Mean Mild abundance (Mean_Mild), Column H: Mean Severe abundance (Mean_Severe), Column I: Log base 2 fold difference between Mild and Healthy controls (Log2FC_MvsHC), Column J: Log base 2 fold difference between Severe and Healthy controls (Log2FC_SvsHC), Column K: Log base 2 fold difference between Severe and Mild (Log2FC_SvsM), Column L: p value for the comparison of means in the Mild vs. Healthy controls (Adj_Pvalue_MvsHC), Column M: p value for the comparison of means in the Severe vs. Healthy controls (Adj_Pvalue_SvsHC), Column N: p value for the comparison of means in the Severe vs. Mild (Adj_Pvalue_SvsM). (Attached in separate file)

**Supplemental Table II:** Plasma Pathway Analysis. Column A: Biological Pathway Description, Column B: p value of the pairwise comparison of Severe vs. Healthy control pathways (SvsHC_padjust), Column C: Number of pairwise protein differences in each pathway between Severe and Healthy Controls (SvsHC_Count), Column D: p value of the pairwise comparison of Severe vs. mild pathways (SvsM_padjust), Column E: Number of pairwise protein differences in each pathway between Severe and Mild (SvsM_Count), Column F: p value of the pairwise comparison of Mild vs. Healthy Control pathways (MvsHC_padjust) Column G: Number of pairwise protein differences in each pathway between Mild and Healthy Controls (MvsHC_Count). (Attached in separate file)

**Supplemental Table III:** Statistical Analysis of Plasma Metabolites. Column B: Metabolite name, Column C: Number of Healthy control samples where the metabolite was detected (N_HC), Column D: Number of Mild samples where the metabolite was detected (N_Mild), Column E: Number of Severe samples where the metabolite was detected (N_Severe), Column F: Mean healthy control protein abundance (Mean_HC), Column G: Mean Mild protein abundance (Mean_Mild), Column H: Mean Severe protein abundance (Mean_Severe), Column I: Log base 2 fold difference between Mild and Healthy controls (Log2FC_MvsHC), Column J: Log base 2 fold difference between Severe and Healthy controls (Log2FC_SvsHC), Column K: Log base 2 fold difference between Severe and Mild (Log2FC_SvsM) Column L: p value for the comparison of means in the Mild vs. Healthy controls (Adj_Pvalue_MvsHC), Column M: p value for the comparison of means in the Severe vs. Healthy controls (Adj_Pvalue_SvsHC), Column M: p value for the comparison of means in the Severe vs. Mild (Adj_Pvalue_SvsM). (Attached in separate file)

**Supplemental Table IV:** Correlative analysis of Neutrophil extracellular trap mean z-score with protein levels across the entire plasma study. Column A: Uniprot protein ID, Column B: uniprot_url link, Column C: Pearson r value indicated by “Cor_w_NET”. Significant r values were included between an r value range of -0.6 to -1.0 and 0.6 to 1.0. (Attached in separate file)

