## Supplemental Figures and Tables for "Multi-omics Characterization of Neutrophil Extracellular Trap Formation in Severe and Mild COVID-19 Infections"

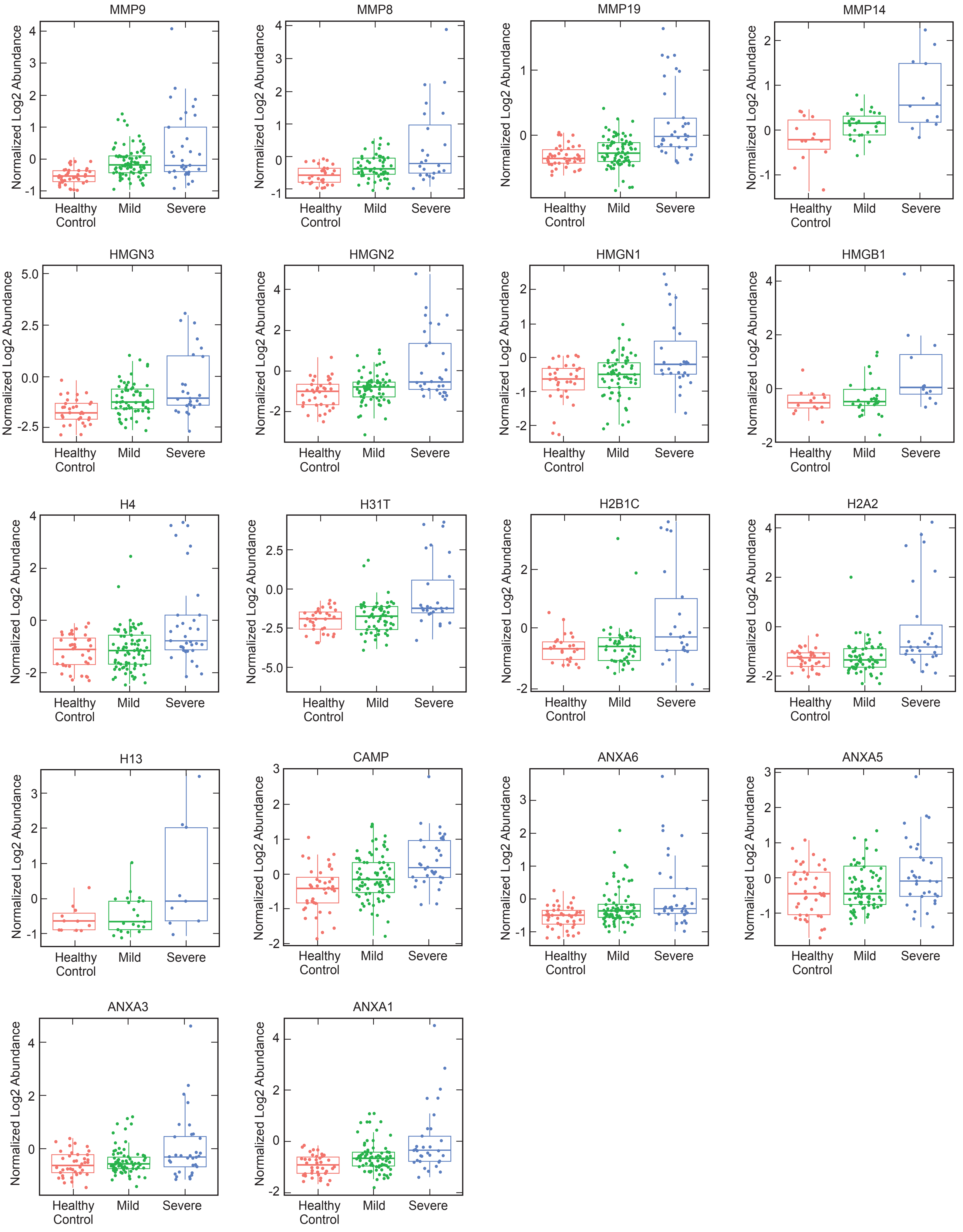


Supplemental Figure 1

Supplemental Figure 2

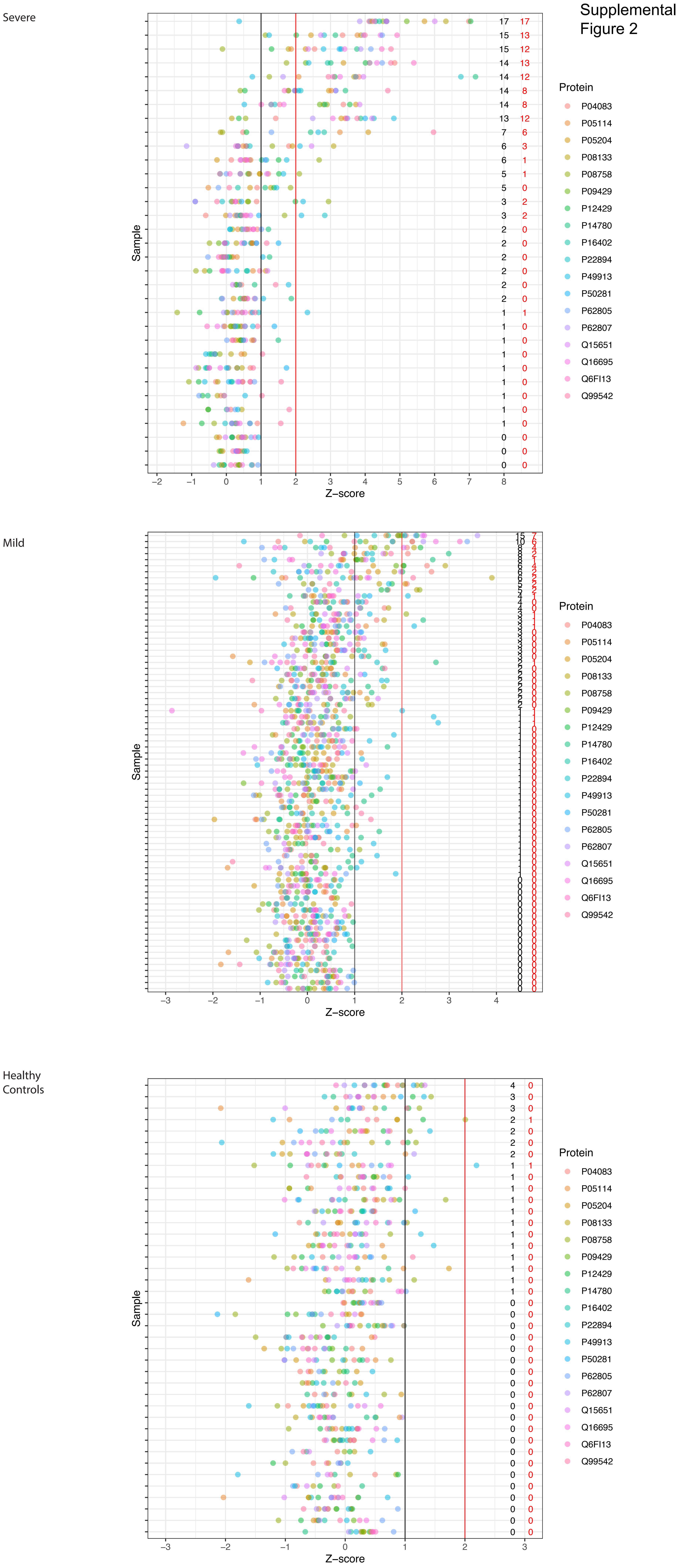

### Supplemental Figure 3

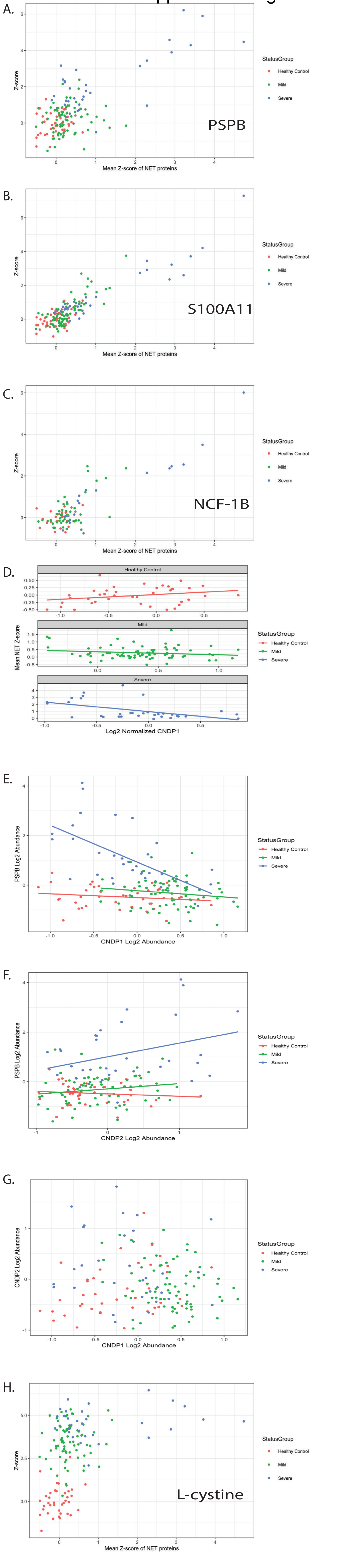

Supplemental Figure 4

>sp|Q96KN2|CNDP1_HUMAN Beta-Ala-His dipeptidase OS=Homo sapiens OX=9606 GN=CNDP1 PE=1 SV=4
MDPKLGRMAASLLAVLLLLLERGMFS**SPSPPPALLEKVFQYIDLHQDEFVQTLKEWVAIESDSVQPVPR**FRQELFR**MMAVAADTLQR**LGA
R**VASVDMGPQQLPDGQSLPIPPIILAELGSDPTKGTVCFYGHLDVQPADRGDGWLTDPYVLTEVDGK**LYGRGATDNK**GPVLAWINAVSAF**
**RALEQDLPVNIKFIIEGMEEAGSVALEELVEK**EKDRFFSGVDYIVISDNLWISQR**KPAITYGTRGNSYFMVEVK**CRDQDFHSGTFGGILH
EPMADLVALLGSLVDSSGH**ILVPGIYDEVVPLTEEEINTYKAIHLDLEEYR**NSSRVEKFLFDTK**EEILMHLWRYPSLSIHGIEGAFDEPG**
**TK**TVIPGRVIGKFSIRLVPHMNVSAVEKQVTR**HLEDVFSK**RNSSNK**MVVSMTLGLHPWIANIDDTQYLAAK**RAIR**TVFGTEPDMIRDGST**
**IPIAKMFQEIVHKSVVLIPLGAVDDGEHSQNEK**INR**WNYIEGTKLFAAFFLEMAQLH**

>sp|Q96KP4|CNDP2_HUMAN Cytosolic non-specific dipeptidase OS=Homo sapiens OX=9606 GN=CNDP2 PE=1 SV=2
MAALTTLFK**YIDENQDR**YIKKLAK**WVAIQSVSAWPEK**RGEIRR**MMEVAAADVKQLGGSVELVDIGKQKLPDGSEIPLPPILLGR**LGSDPQ
KKTVCIYGHLDVQPAALEDGWDSEPFTLVERDGKLYGRGSTDDKGPVAGWINALEAYQK**TGQEIPVNVR**FCLEGMEESGSEGLDELIFAR
KDTFFK**DVDYVCISDNYWLGK**KKPCITYGLRGICYFFIEVECSNKDLHSGVYGGSVHEAMTDLILLMGSLVDKRGNILIPGINEAVAAVT
EEEHKLYDDIDFDIEEFAK**DVGAQILLHSHK**KDILMHRWRYPSLSLHGIEGAFSGSGAKTVIPRKVVGKFSIRLVPNMTPEVVGEQVTSY
LTKKFAELRSPNEFK**VYMGHGGK**PWVSDFSHPHYLAGRRAMK**TVFGVEPDLTREGGSIPVTLTFQEATGK**NVMLLPVGSADDGAHSQNEK
LNRYNYIEGTK**MLAAYLYEVSQLK**D

### Supplemental Table I:

<table>
<tr><th>Protein</th><th>N_Peptides</th><th>N_HC</th><th>N_Mild</th><th>N_Severe</th><th>Mean_HC</th><th>Mean_Mild</th><th>Mean_Severe</th><th>Log2FC_MvsHC</th><th>Log2FC_SvsHC</th><th>Log2FC_SvsM</th><th>Adj_Pvalue_MvsHC</th><th>Adj_Pvalue_SvsHC</th><th>Adj_Pvalue_SvsM</th></tr>
<tr><td>sp|O15144|ARPC2_HUMAN</td><td>1</td><td>40</td><td>76</td><td>33</td><td>-0.02259</td><td>-0.06989545</td><td>0.20023523</td><td>-0.047303905</td><td>0.222826775</td><td>0.27013068</td><td>0.884546227</td><td>0.158120539</td><td>0.033549362</td></tr>
<tr><td>sp|Q99460|PSMD1_HUMAN</td><td>2</td><td>8</td><td>14</td><td>6</td><td>-0.11791</td><td>0.131884021</td><td>0.196822009</td><td>0.249796968</td><td>0.314734955</td><td>0.064937988</td><td>0.686891423</td><td>0.669587503</td><td>0.978954397</td></tr>
<tr><td>sp|O75882|ATRN_HUMAN</td><td>1</td><td>40</td><td>76</td><td>33</td><td>0.307815</td><td>0.192497743</td><td>0.02580914</td><td>-0.115317339</td><td>-0.282005942</td><td>-0.166688603</td><td>0.000526847</td><td>3.15E-12</td><td>1.92E-06</td></tr>
<tr><td>sp|P05976|MYL1_HUMAN</td><td>2</td><td>40</td><td>76</td><td>33</td><td>-2.04317</td><td>-2.246505281</td><td>-1.624210698</td><td>-0.203333813</td><td>0.418960771</td><td>0.622294583</td><td>0.480489602</td><td>0.120498268</td><td>0.003249631</td></tr>
<tr><td>sp|Q9NZP8|C1RL_HUMAN</td><td>2</td><td>40</td><td>76</td><td>33</td><td>0.183327</td><td>0.296293903</td><td>0.224965777</td><td>0.112967172</td><td>0.041639046</td><td>-0.071328125</td><td>0.003708868</td><td>0.575604385</td><td>0.131255072</td></tr>
<tr><td>sp|P05155|IC1_HUMAN</td><td>2</td><td>40</td><td>76</td><td>33</td><td>0.514166</td><td>0.480501436</td><td>0.604067741</td><td>-0.033664368</td><td>0.089901937</td><td>0.123566305</td><td>0.82524314</td><td>0.391527087</td><td>0.108174834</td></tr>
<tr><td>sp|P08697|A2AP_HUMAN</td><td>2</td><td>40</td><td>76</td><td>33</td><td>0.346401</td><td>0.310272943</td><td>0.093929984</td><td>-0.036128008</td><td>-0.252470968</td><td>-0.216342959</td><td>0.3167739</td><td>1.30E-13</td><td>4.87E-13</td></tr>
<tr><td>sp|Q8NI99|ANGL6_HUMAN</td><td>1</td><td>40</td><td>76</td><td>33</td><td>0.061497</td><td>0.142930088</td><td>-0.094752306</td><td>0.081433506</td><td>-0.156248889</td><td>-0.237682395</td><td>0.045414294</td><td>0.000541769</td><td>2.33E-09</td></tr>
<tr><td>sp|P02765|FETUA_HUMAN</td><td>36</td><td>40</td><td>76</td><td>33</td><td>0.522453</td><td>0.459110354</td><td>0.094100381</td><td>-0.063342935</td><td>-0.428352908</td><td>-0.365009973</td><td>0.30702339</td><td>2.65E-13</td><td>1.42E-12</td></tr>
<tr><td>sp|P0C0L5|CO4B_HUMAN</td><td>3</td><td>40</td><td>76</td><td>33</td><td>0.324434</td><td>0.375437291</td><td>0.244924753</td><td>0.051003073</td><td>-0.079509464</td><td>-0.130512538</td><td>0.565479717</td><td>0.385950352</td><td>0.04119814</td></tr>
<tr><td>sp|P19971|TYPH_HUMAN</td><td>2</td><td>40</td><td>76</td><td>33</td><td>-0.19728</td><td>-0.064090235</td><td>0.266718914</td><td>0.133187548</td><td>0.463996698</td><td>0.33080915</td><td>0.308394405</td><td>0.000110372</td><td>0.002322349</td></tr>
<tr><td>sp|P04217|A1BG_HUMAN</td><td>1</td><td>40</td><td>76</td><td>33</td><td>0.545594</td><td>0.513080348</td><td>0.407083746</td><td>-0.032513941</td><td>-0.138510543</td><td>-0.105996601</td><td>0.348069992</td><td>6.77E-06</td><td>0.000111899</td></tr>
<tr><td>sp|P01008|ANT3_HUMAN</td><td>4</td><td>40</td><td>76</td><td>33</td><td>0.483673</td><td>0.419441126</td><td>0.152338465</td><td>-0.064231988</td><td>-0.331334648</td><td>-0.26710266</td><td>0.107486331</td><td>5.64E-14</td><td>1.61E-12</td></tr>
<tr><td>sp|P05154|IPSP_HUMAN</td><td>6</td><td>40</td><td>76</td><td>33</td><td>0.266019</td><td>0.008342897</td><td>-0.073441734</td><td>-0.257675996</td><td>-0.339460627</td><td>-0.081784631</td><td>0.05317804</td><td>0.030390871</td><td>0.765790244</td></tr>
<tr><td>sp|P05543|THBG_HUMAN</td><td>1</td><td>40</td><td>76</td><td>33</td><td>0.279287</td><td>0.270144321</td><td>0.0098842</td><td>-0.009142869</td><td>-0.26940299</td><td>-0.260260121</td><td>0.969332834</td><td>1.08E-07</td><td>8.07E-09</td></tr>
<tr><td>sp|P36955|PEDF_HUMAN</td><td>2</td><td>40</td><td>76</td><td>33</td><td>0.318915</td><td>0.28045577</td><td>0.32161474</td><td>-0.038458994</td><td>0.002699976</td><td>0.04115897</td><td>0.467593663</td><td>0.997397407</td><td>0.465628761</td></tr>
<tr><td>sp|P00734|THRB_HUMAN</td><td>4</td><td>40</td><td>76</td><td>33</td><td>0.279823</td><td>0.248218247</td><td>0.007346056</td><td>-0.031604851</td><td>-0.272477042</td><td>-0.240872191</td><td>0.464349479</td><td>1.08E-13</td><td>1.18E-13</td></tr>
<tr><td>sp|P07357|CO8A_HUMAN</td><td>7</td><td>40</td><td>76</td><td>33</td><td>0.507877</td><td>0.480397854</td><td>0.404315724</td><td>-0.027479207</td><td>-0.103561336</td><td>-0.07608213</td><td>0.628664363</td><td>0.012659583</td><td>0.047778113</td></tr>
<tr><td>sp|Q9Y6R7|FCGBP_HUMAN</td><td>5</td><td>40</td><td>76</td><td>33</td><td>0.138323</td><td>0.220596563</td><td>0.447576704</td><td>0.082273767</td><td>0.309253908</td><td>0.226980141</td><td>0.274749238</td><td>1.10E-05</td><td>0.000312078</td></tr>
<tr><td>sp|Q2TV78|MST1L_HUMAN</td><td>11</td><td>40</td><td>76</td><td>33</td><td>0.15427</td><td>0.196031405</td><td>0.031882211</td><td>0.04176125</td><td>-0.122387944</td><td>-0.164149194</td><td>0.258062595</td><td>0.000527314</td><td>1.10E-07</td></tr>
<tr><td>sp|P00450|CERU_HUMAN</td><td>1</td><td>40</td><td>76</td><td>33</td><td>0.479359</td><td>0.518844654</td><td>0.384616701</td><td>0.039485218</td><td>-0.094742735</td><td>-0.134227954</td><td>0.562410496</td><td>0.105765613</td><td>0.003884347</td></tr>
<tr><td>sp|P09668|CATH_HUMAN</td><td>2</td><td>40</td><td>76</td><td>33</td><td>-0.09716</td><td>0.070885582</td><td>0.121576627</td><td>0.168040705</td><td>0.21873175</td><td>0.050691045</td><td>0.01079464</td><td>0.005214021</td><td>0.685491849</td></tr>
<tr><td>sp|P00918|CAH2_HUMAN</td><td>2</td><td>40</td><td>76</td><td>33</td><td>-0.23811</td><td>0.01430173</td><td>0.38264692</td><td>0.252412759</td><td>0.620757949</td><td>0.36834519</td><td>0.029359064</td><td>1.48E-06</td><td>0.001641871</td></tr>
<tr><td>sp|P62258|1433E_HUMAN</td><td>2</td><td>40</td><td>76</td><td>33</td><td>-0.08382</td><td>-0.138649778</td><td>0.347601151</td><td>-0.054832939</td><td>0.43141799</td><td>0.486250929</td><td>0.857868947</td><td>0.002112534</td><td>6.52E-05</td></tr>
<tr><td>sp|P02671|FIBA_HUMAN</td><td>2</td><td>40</td><td>76</td><td>33</td><td>0.135583</td><td>0.28042221</td><td>0.599116108</td><td>0.14483939</td><td>0.463533288</td><td>0.318693897</td><td>0.131792411</td><td>2.46E-06</td><td>0.000299489</td></tr>
<tr><td>sp|Q14624|ITIH4_HUMAN</td><td>4</td><td>40</td><td>76</td><td>33</td><td>0.65837</td><td>0.674030345</td><td>0.666827681</td><td>0.015660468</td><td>0.008457803</td><td>-0.007202665</td><td>0.884556113</td><td>0.975589431</td><td>0.977451359</td></tr>
<tr><td>sp|P08123|CO1A2_HUMAN</td><td>3</td><td>40</td><td>76</td><td>33</td><td>-0.01324</td><td>-0.038838481</td><td>0.310930369</td><td>-0.02559618</td><td>0.32417267</td><td>0.34976885</td><td>0.912115741</td><td>9.21E-05</td><td>1.70E-06</td></tr>
<tr><td>sp|P03952|KLKB1_HUMAN</td><td>6</td><td>40</td><td>76</td><td>33</td><td>0.503356</td><td>0.471927505</td><td>0.183779574</td><td>-0.031428425</td><td>-0.319576356</td><td>-0.288147931</td><td>0.748631082</td><td>2.33E-08</td><td>1.38E-08</td></tr>
<tr><td>sp|P00751|CFAB_HUMAN</td><td>4</td><td>40</td><td>76</td><td>33</td><td>0.236259</td><td>0.29120378</td><td>0.234919116</td><td>0.054944655</td><td>-0.001340009</td><td>-0.056284665</td><td>0.340552262</td><td>0.999552936</td><td>0.37034438</td></tr>
<tr><td>sp|P00748|FA12_HUMAN</td><td>2</td><td>40</td><td>76</td><td>33</td><td>0.231376</td><td>0.271177443</td><td>-0.004988407</td><td>0.039801727</td><td>-0.236364123</td><td>-0.27616585</td><td>0.759715504</td><td>0.001853044</td><td>2.73E-05</td></tr>
<tr><td>sp|P00742|FA10_HUMAN</td><td>3</td><td>40</td><td>76</td><td>33</td><td>0.218106</td><td>0.235792075</td><td>0.156493905</td><td>0.0176859</td><td>-0.06161227</td><td>-0.07929817</td><td>0.814997213</td><td>0.185187289</td><td>0.03051772</td></tr>
<tr><td>sp|P00740|FA9_HUMAN</td><td>22</td><td>40</td><td>76</td><td>33</td><td>0.249501</td><td>0.260068143</td><td>0.294564302</td><td>0.01056695</td><td>0.045063109</td><td>0.034496159</td><td>0.909627851</td><td>0.308821758</td><td>0.415238237</td></tr>
<tr><td>sp|P02751|FINC_HUMAN</td><td>3</td><td>40</td><td>76</td><td>33</td><td>0.625746</td><td>0.790995647</td><td>0.825774037</td><td>0.165249927</td><td>0.200028316</td><td>0.03477839</td><td>0.161416338</td><td>0.158280195</td><td>0.930299472</td></tr>
<tr><td>sp|P06396|GELS_HUMAN</td><td>5</td><td>40</td><td>76</td><td>33</td><td>0.522895</td><td>0.372026377</td><td>-0.003807242</td><td>-0.150868314</td><td>-0.526701934</td><td>-0.37583362</td><td>0.012312934</td><td>1.63E-13</td><td>1.01E-09</td></tr>
<tr><td>sp|P15291|B4GT1_HUMAN</td><td>1</td><td>40</td><td>76</td><td>33</td><td>0.009626</td><td>0.052458421</td><td>0.451220814</td><td>0.042831997</td><td>0.44159439</td><td>0.398762393</td><td>0.820356411</td><td>2.61E-06</td><td>1.70E-06</td></tr>
<tr><td>sp|P19827|ITIH1_HUMAN</td><td>57</td><td>40</td><td>76</td><td>33</td><td>0.625391</td><td>0.53613162</td><td>0.292288999</td><td>-0.089259218</td><td>-0.33310184</td><td>-0.243842621</td><td>0.01794115</td><td>8.62E-14</td><td>1.89E-10</td></tr>
<tr><td>sp|Q15063|POSTN_HUMAN</td><td>5</td><td>40</td><td>76</td><td>33</td><td>0.16417</td><td>0.117024309</td><td>0.122426851</td><td>-0.0471452</td><td>-0.041742658</td><td>0.005402542</td><td>0.811372998</td><td>0.892989916</td><td>0.997587612</td></tr>
<tr><td>sp|O00391|QSOX1_HUMAN</td><td>4</td><td>40</td><td>76</td><td>33</td><td>0.116624</td><td>0.090302507</td><td>0.135055549</td><td>-0.026321437</td><td>0.018431605</td><td>0.044753042</td><td>0.698653079</td><td>0.885472491</td><td>0.404581119</td></tr>
<tr><td>sp|P04114|APOB_HUMAN</td><td>2</td><td>40</td><td>76</td><td>33</td><td>0.764763</td><td>0.593495285</td><td>0.644518911</td><td>-0.171268201</td><td>-0.120244575</td><td>0.051023626</td><td>5.11E-06</td><td>0.011626374</td><td>0.34758575</td></tr>
<tr><td>sp|P22792|CPN2_HUMAN</td><td>5</td><td>40</td><td>76</td><td>33</td><td>0.275923</td><td>0.261397545</td><td>0.1685504</td><td>-0.014525886</td><td>-0.10737303</td><td>-0.092847145</td><td>0.884534912</td><td>0.011979902</td><td>0.014721229</td></tr>
<tr><td>sp|Q04756|HGFA_HUMAN</td><td>7</td><td>40</td><td>76</td><td>33</td><td>0.327763</td><td>0.24824213</td><td>-0.013154084</td><td>-0.079521164</td><td>-0.340917378</td><td>-0.261396214</td><td>0.174192042</td><td>5.99E-09</td><td>4.21E-07</td></tr>
<tr><td>sp|P02766|TTHY_HUMAN</td><td>4</td><td>40</td><td>76</td><td>33</td><td>0.285327</td><td>-0.078269826</td><td>-0.203788827</td><td>-0.363597196</td><td>-0.489116197</td><td>-0.125519001</td><td>6.68E-07</td><td>3.08E-08</td><td>0.187374208</td></tr>
<tr><td>sp|Q15582|BGH3_HUMAN</td><td>17</td><td>40</td><td>76</td><td>33</td><td>0.196206</td><td>0.237746952</td><td>0.155471904</td><td>0.041540528</td><td>-0.040734521</td><td>-0.082275049</td><td>0.707724296</td><td>0.794816346</td><td>0.30723364</td></tr>
<tr><td>sp|P15144|AMPN_HUMAN</td><td>15</td><td>40</td><td>76</td><td>33</td><td>0.097444</td><td>0.122107804</td><td>0.089799532</td><td>0.024663599</td><td>-0.007644673</td><td>-0.032308272</td><td>0.788024509</td><td>0.984295227</td><td>0.698756887</td></tr>
<tr><td>sp|Q96PD5|PGRP2_HUMAN</td><td>95</td><td>40</td><td>76</td><td>33</td><td>0.468645</td><td>0.336146306</td><td>0.032219974</td><td>-0.132499165</td><td>-0.436425497</td><td>-0.303926332</td><td>0.017971025</td><td>1.36E-11</td><td>6.43E-08</td></tr>
<tr><td>sp|P11047|LAMC1_HUMAN</td><td>4</td><td>40</td><td>76</td><td>33</td><td>-0.07942</td><td>-0.064538859</td><td>0.362251723</td><td>0.014885473</td><td>0.441676055</td><td>0.426790582</td><td>0.978990622</td><td>9.84E-06</td><td>1.42E-06</td></tr>
<tr><td>sp|P02655|APOC2_HUMAN</td><td>1</td><td>40</td><td>76</td><td>33</td><td>-0.10338</td><td>-0.255339291</td><td>0.181230468</td><td>-0.151955475</td><td>0.284614284</td><td>0.436569759</td><td>0.446702006</td><td>0.14546601</td><td>0.003831977</td></tr>
<tr><td>sp|P01019|ANGT_HUMAN</td><td>23</td><td>40</td><td>76</td><td>33</td><td>0.209328</td><td>0.220829991</td><td>0.123063076</td><td>0.011501634</td><td>-0.08626528</td><td>-0.097766914</td><td>0.97693301</td><td>0.407622709</td><td>0.232675296</td></tr>
<tr><td>sp|P10619|PPGB_HUMAN</td><td>2</td><td>40</td><td>76</td><td>33</td><td>0.003333</td><td>0.117193034</td><td>0.398990218</td><td>0.113859851</td><td>0.395657035</td><td>0.281797184</td><td>0.282591261</td><td>6.15E-05</td><td>0.001578439</td></tr>
<tr><td>sp|P41222|PTGDS_HUMAN</td><td>12</td><td>40</td><td>76</td><td>33</td><td>0.007307</td><td>-0.02519492</td><td>0.344469488</td><td>-0.032501893</td><td>0.337162515</td><td>0.369664407</td><td>0.91637922</td><td>0.002245293</td><td>0.000114912</td></tr>
<tr><td>sp|Q9UM47|NOTC3_HUMAN</td><td>12</td><td>40</td><td>76</td><td>33</td><td>0.116058</td><td>0.036273832</td><td>0.20083903</td><td>-0.079784309</td><td>0.084780889</td><td>0.164565198</td><td>0.174861681</td><td>0.255641092</td><td>0.001974784</td></tr>
<tr><td>sp|P02774|VTDB_HUMAN</td><td>32</td><td>40</td><td>76</td><td>33</td><td>0.7797</td><td>0.777676996</td><td>0.558558683</td><td>-0.002022528</td><td>-0.221140841</td><td>-0.219118312</td><td>0.997911497</td><td>3.19E-07</td><td>1.27E-08</td></tr>
<tr><td>sp|O94985|CSTN1_HUMAN</td><td>43</td><td>40</td><td>76</td><td>33</td><td>0.174248</td><td>0.123637023</td><td>-0.074112735</td><td>-0.050610882</td><td>-0.24836064</td><td>-0.197749758</td><td>0.178577228</td><td>5.99E-11</td><td>3.04E-09</td></tr>
<tr><td>sp|O43827|ANGL7_HUMAN</td><td>1</td><td>33</td><td>62</td><td>27</td><td>-0.00785</td><td>-0.042841294</td><td>-0.230260532</td><td>-0.034994054</td><td>-0.222413292</td><td>-0.187419238</td><td>0.844301465</td><td>0.011346637</td><td>0.017453082</td></tr>
<tr><td>sp|Q16610|ECM1_HUMAN</td><td>11</td><td>40</td><td>76</td><td>33</td><td>0.298802</td><td>0.258875681</td><td>0.083662517</td><td>-0.039926576</td><td>-0.21513974</td><td>-0.175213165</td><td>0.683638502</td><td>0.00080924</td><td>0.002312339</td></tr>
<tr><td>sp|P10643|CO7_HUMAN</td><td>1</td><td>40</td><td>76</td><td>33</td><td>0.220765</td><td>0.187521107</td><td>0.284261797</td><td>-0.033243508</td><td>0.063497181</td><td>0.09674069</td><td>0.752402755</td><td>0.490238103</td><td>0.125472078</td></tr>
<tr><td>sp|P0DOY3|IGLC3_HUMAN</td><td>1</td><td>40</td><td>76</td><td>33</td><td>-0.11017</td><td>-0.02962129</td><td>0.270139853</td><td>0.080549902</td><td>0.380311045</td><td>0.299761143</td><td>0.626220882</td><td>0.001163323</td><td>0.004466706</td></tr>
<tr><td>sp|A0A0C4DH67|KV108_HUMAN</td><td>79</td><td>40</td><td>76</td><td>33</td><td>0.072986</td><td>-0.068703187</td><td>-0.095495275</td><td>-0.141689306</td><td>-0.168481393</td><td>-0.026792088</td><td>0.451570564</td><td>0.460348447</td><td>0.975150532</td></tr>
<tr><td>sp|P35968|VGFR2_HUMAN</td><td>6</td><td>40</td><td>76</td><td>33</td><td>0.006248</td><td>0.052659445</td><td>-0.096338148</td><td>0.046411168</td><td>-0.102586425</td><td>-0.148997593</td><td>0.386705111</td><td>0.04366469</td><td>0.000330702</td></tr>
<tr><td>sp|P01042|KNG1_HUMAN</td><td>2</td><td>40</td><td>76</td><td>33</td><td>0.325089</td><td>0.337270891</td><td>0.195915881</td><td>0.012181988</td><td>-0.129173022</td><td>-0.141355011</td><td>0.899066337</td><td>0.000475246</td><td>1.26E-05</td></tr>
<tr><td>sp|P35443|TSP4_HUMAN</td><td>8</td><td>40</td><td>76</td><td>33</td><td>0.052356</td><td>0.163083237</td><td>0.126440982</td><td>0.110727623</td><td>0.074085369</td><td>-0.036642254</td><td>0.386859274</td><td>0.744243263</td><td>0.911970175</td></tr>
<tr><td>sp|P02649|APOE_HUMAN</td><td>2</td><td>40</td><td>76</td><td>33</td><td>-0.04666</td><td>0.062792736</td><td>0.380080261</td><td>0.109450463</td><td>0.426737988</td><td>0.317287525</td><td>0.14251754</td><td>2.10E-08</td><td>2.38E-06</td></tr>
<tr><td>sp|O75015|FCG3B_HUMAN</td><td>3</td><td>40</td><td>76</td><td>33</td><td>-0.10371</td><td>0.060623943</td><td>0.170453655</td><td>0.164338234</td><td>0.274167946</td><td>0.109829712</td><td>0.143176778</td><td>0.025543207</td><td>0.462255368</td></tr>
<tr><td>sp|Q9Y490|TLN1_HUMAN</td><td>5</td><td>40</td><td>76</td><td>33</td><td>-0.05974</td><td>-0.023726581</td><td>0.199546356</td><td>0.036009732</td><td>0.259282669</td><td>0.223272937</td><td>0.96868686</td><td>0.324687269</td><td>0.345722001</td></tr>
<tr><td>sp|P35858|ALS_HUMAN</td><td>10</td><td>40</td><td>76</td><td>33</td><td>0.3977</td><td>0.445965567</td><td>-0.030986087</td><td>0.048265556</td><td>-0.428686099</td><td>-0.476951655</td><td>0.681054832</td><td>1.90E-08</td><td>4.46E-12</td></tr>
<tr><td>sp|P35590|TIE1_HUMAN</td><td>2</td><td>40</td><td>76</td><td>33</td><td>0.093968</td><td>0.154126458</td><td>0.238795436</td><td>0.060158289</td><td>0.144827268</td><td>0.084668978</td><td>0.290972302</td><td>0.008545329</td><td>0.119176482</td></tr>
<tr><td>sp|P01031|CO5_HUMAN</td><td>14</td><td>40</td><td>76</td><td>33</td><td>0.302942</td><td>0.292839972</td><td>0.315227315</td><td>-0.010102285</td><td>0.012285057</td><td>0.022387343</td><td>0.931563648</td><td>0.930221537</td><td>0.737199253</td></tr>
<tr><td>sp|P06744|G6PI_HUMAN</td><td>23</td><td>40</td><td>76</td><td>33</td><td>-0.20999</td><td>-0.126265148</td><td>0.382625296</td><td>0.083729685</td><td>0.592620129</td><td>0.508890444</td><td>0.744725388</td><td>8.98E-05</td><td>0.000152915</td></tr>
<tr><td>sp|P01344|IGF2_HUMAN</td><td>4</td><td>40</td><td>76</td><td>33</td><td>0.096826</td><td>0.141707901</td><td>-0.142457832</td><td>0.044882348</td><td>-0.239283385</td><td>-0.284165733</td><td>0.498986958</td><td>5.02E-06</td><td>1.36E-09</td></tr>
<tr><td>sp|P48740|MASP1_HUMAN</td><td>1</td><td>40</td><td>76</td><td>33</td><td>0.20667</td><td>0.2373887</td><td>0.137660039</td><td>0.030719091</td><td>-0.069009569</td><td>-0.09972866</td><td>0.509916319</td><td>0.099711933</td><td>0.002712488</td></tr>
<tr><td>sp|P08294|SODE_HUMAN</td><td>11</td><td>40</td><td>76</td><td>33</td><td>-0.06067</td><td>-0.229638895</td><td>-0.094398501</td><td>-0.16896513</td><td>-0.033724736</td><td>0.135240393</td><td>0.546712468</td><td>0.983436333</td><td>0.71134692</td></tr>
<tr><td>sp|P05160|F13B_HUMAN</td><td>1</td><td>40</td><td>76</td><td>33</td><td>0.275354</td><td>0.197442907</td><td>0.074794826</td><td>-0.07791103</td><td>-0.200559111</td><td>-0.122648081</td><td>0.0791257</td><td>2.20E-05</td><td>0.004677605</td></tr>
<tr><td>sp|P15529|MCP_HUMAN</td><td>3</td><td>40</td><td>76</td><td>33</td><td>0.022636</td><td>-0.055177392</td><td>0.099139303</td><td>-0.07781321</td><td>0.076503484</td><td>0.154316695</td><td>0.217498087</td><td>0.359564794</td><td>0.006211887</td></tr>
<tr><td>sp|P02760|AMBP_HUMAN</td><td>1</td><td>40</td><td>76</td><td>33</td><td>0.301556</td><td>0.251846477</td><td>0.261650951</td><td>-0.049709873</td><td>-0.039905398</td><td>0.009804474</td><td>0.166619839</td><td>0.447031444</td><td>0.939567264</td></tr>
<tr><td>sp|P26927|HGFL_HUMAN</td><td>2</td><td>40</td><td>76</td><td>33</td><td>0.148903</td><td>0.187457604</td><td>0.028786606</td><td>0.038554646</td><td>-0.120116351</td><td>-0.158670997</td><td>0.259228242</td><td>0.000218014</td><td>3.03E-08</td></tr>
<tr><td>sp|P24821|TENA_HUMAN</td><td>13</td><td>40</td><td>76</td><td>33</td><td>-0.13175</td><td>-0.028445605</td><td>0.459526656</td><td>0.103305028</td><td>0.591277289</td><td>0.487972261</td><td>0.368014001</td><td>4.96E-09</td><td>4.64E-08</td></tr>
<tr><td>sp|P08603|CFAH_HUMAN</td><td>2</td><td>40</td><td>76</td><td>33</td><td>0.29547</td><td>0.315600982</td><td>0.298575461</td><td>0.020131062</td><td>0.00310554</td><td>-0.017025521</td><td>0.690407417</td><td>0.993911622</td><td>0.792157033</td></tr>
<tr><td>sp|P05156|CFAI_HUMAN</td><td>6</td><td>40</td><td>76</td><td>33</td><td>0.173447</td><td>0.176639496</td><td>0.162732118</td><td>0.003191998</td><td>-0.010715379</td><td>-0.013907377</td><td>0.993565329</td><td>0.951067199</td><td>0.898098973</td></tr>
<tr><td>sp|P00747|PLMN_HUMAN</td><td>3</td><td>40</td><td>76</td><td>33</td><td>0.365463</td><td>0.378233991</td><td>0.178147637</td><td>0.012770849</td><td>-0.187315505</td><td>-0.200086354</td><td>0.904377674</td><td>1.93E-06</td><td>1.15E-08</td></tr>
<tr><td>sp|Q16288|NTRK3_HUMAN</td><td>1</td><td>40</td><td>76</td><td>33</td><td>0.237847</td><td>0.074861321</td><td>-0.053768319</td><td>-0.162985549</td><td>-0.291615189</td><td>-0.12862964</td><td>0.000274199</td><td>5.10E-08</td><td>0.009646469</td></tr>
<tr><td>sp|O14498|ISLR_HUMAN</td><td>1</td><td>40</td><td>76</td><td>33</td><td>-0.04108</td><td>0.002942074</td><td>0.163535087</td><td>0.044022148</td><td>0.204615161</td><td>0.160593013</td><td>0.67629675</td><td>0.003957242</td><td>0.012455354</td></tr>
<tr><td>sp|P16035|TIMP2_HUMAN</td><td>6</td><td>40</td><td>76</td><td>33</td><td>0.227487</td><td>0.189866543</td><td>0.164991461</td><td>-0.03761998</td><td>-0.062495062</td><td>-0.024875083</td><td>0.64000817</td><td>0.429063968</td><td>0.842137804</td></tr>
<tr><td>sp|P01033|TIMP1_HUMAN</td><td>3</td><td>40</td><td>76</td><td>33</td><td>-0.13396</td><td>-0.026942589</td><td>0.488656665</td><td>0.10701433</td><td>0.622613585</td><td>0.515599254</td><td>0.399382936</td><td>1.18E-08</td><td>9.13E-08</td></tr>
<tr><td>sp|P01178|NEU1_HUMAN</td><td>10</td><td>33</td><td>62</td><td>27</td><td>-0.16851</td><td>-0.048725533</td><td>-0.027342676</td><td>0.119782337</td><td>0.141165194</td><td>0.021382857</td><td>0.560154358</td><td>0.573972601</td><td>0.983890232</td></tr>
<tr><td>sp|P07237|PDIA1_HUMAN</td><td>11</td><td>40</td><td>76</td><td>33</td><td>0.02636</td><td>0.073571361</td><td>0.313516422</td><td>0.047211228</td><td>0.287156289</td><td>0.239945061</td><td>0.763638492</td><td>0.00155774</td><td>0.003084264</td></tr>
<tr><td>sp|P19823|ITIH2_HUMAN</td><td>1</td><td>40</td><td>76</td><td>33</td><td>0.609194</td><td>0.524105858</td><td>0.338820419</td><td>-0.085088125</td><td>-0.270373564</td><td>-0.185285438</td><td>0.057453732</td><td>2.73E-08</td><td>1.66E-05</td></tr>
<tr><td>sp|Q9BWP8|COL11_HUMAN</td><td>60</td><td>40</td><td>76</td><td>33</td><td>0.023695</td><td>0.117462369</td><td>0.004304475</td><td>0.093767607</td><td>-0.019390287</td><td>-0.113157895</td><td>0.055992336</td><td>0.91614955</td><td>0.025812497</td></tr>
<tr><td>sp|P14625|ENPL_HUMAN</td><td>38</td><td>40</td><td>76</td><td>33</td><td>0.0737</td><td>0.079618312</td><td>0.442221621</td><td>0.00591826</td><td>0.368521568</td><td>0.362603308</td><td>0.993508484</td><td>2.78E-07</td><td>1.35E-08</td></tr>
<tr><td>sp|Q8IWV2|CNTN4_HUMAN</td><td>17</td><td>40</td><td>76</td><td>33</td><td>0.291558</td><td>0.179401088</td><td>-0.065423176</td><td>-0.112157019</td><td>-0.356981283</td><td>-0.244824264</td><td>0.029383595</td><td>7.04E-10</td><td>1.44E-06</td></tr>
<tr><td>sp|Q9HD45|TM9S3_HUMAN</td><td>6</td><td>26</td><td>48</td><td>20</td><td>-0.09869</td><td>-0.094890539</td><td>-0.036207433</td><td>0.00380286</td><td>0.062485966</td><td>0.058683106</td><td>0.999268044</td><td>0.876120673</td><td>0.864465605</td></tr>
<tr><td>sp|P02768|ALBU_HUMAN</td><td>15</td><td>40</td><td>76</td><td>33</td><td>-0.10454</td><td>0.058820025</td><td>0.435763937</td><td>0.163355136</td><td>0.540299048</td><td>0.376943912</td><td>0.48569911</td><td>0.005477203</td><td>0.03735157</td></tr>
<tr><td>sp|P05546|HEP2_HUMAN</td><td>1</td><td>40</td><td>76</td><td>33</td><td>0.359524</td><td>0.471067465</td><td>0.222903632</td><td>0.111543504</td><td>-0.136620329</td><td>-0.248163833</td><td>0.07446438</td><td>0.068144199</td><td>2.85E-05</td></tr>
<tr><td>sp|Q08380|LG3BP_HUMAN</td><td>6</td><td>40</td><td>76</td><td>33</td><td>-0.14116</td><td>0.070112323</td><td>0.274407307</td><td>0.211271496</td><td>0.41556648</td><td>0.204294984</td><td>0.034334016</td><td>0.000191711</td><td>0.061757539</td></tr>
<tr><td>sp|A1L4H1|SRCRL_HUMAN</td><td>1</td><td>40</td><td>76</td><td>33</td><td>0.209399</td><td>0.237918461</td><td>0.511397979</td><td>0.028519495</td><td>0.301999013</td><td>0.273479518</td><td>0.785841394</td><td>1.06E-07</td><td>5.74E-08</td></tr>
<tr><td>sp|P0C0L4|CO4A_HUMAN</td><td>7</td><td>40</td><td>76</td><td>33</td><td>0.191429</td><td>0.184627922</td><td>0.096938076</td><td>-0.006800634</td><td>-0.094490481</td><td>-0.087689847</td><td>0.988395858</td><td>0.216552904</td><td>0.187472908</td></tr>
<tr><td>sp|Q9UKX3|MYH13_HUMAN</td><td>9</td><td>40</td><td>76</td><td>33</td><td>-0.06614</td><td>-0.283020888</td><td>-0.075950718</td><td>-0.216884327</td><td>-0.009814156</td><td>0.207070171</td><td>0.022892902</td><td>0.994465886</td><td>0.047585705</td></tr>
<tr><td>sp|Q12907|LMAN2_HUMAN</td><td>2</td><td>40</td><td>76</td><td>33</td><td>-0.14019</td><td>-0.095541129</td><td>0.36384128</td><td>0.044646673</td><td>0.504029082</td><td>0.459382409</td><td>0.81371094</td><td>1.58E-07</td><td>7.21E-08</td></tr>
<tr><td>sp|P14209|CD99_HUMAN</td><td>40</td><td>40</td><td>76</td><td>33</td><td>0.052839</td><td>0.057374594</td><td>0.471837022</td><td>0.00453547</td><td>0.418997898</td><td>0.414462428</td><td>0.997820584</td><td>1.03E-05</td><td>8.40E-07</td></tr>
<tr><td>sp|Q9BWV1|BOC_HUMAN</td><td>7</td><td>40</td><td>76</td><td>33</td><td>0.077498</td><td>0.026060644</td><td>0.148844169</td><td>-0.05143694</td><td>0.071346584</td><td>0.122783525</td><td>0.393141181</td><td>0.290721332</td><td>0.011077619</td></tr>
<tr><td>sp|P43652|AFAM_HUMAN</td><td>5</td><td>40</td><td>76</td><td>33</td><td>0.385067</td><td>0.253051375</td><td>0.122744654</td><td>-0.132015748</td><td>-0.262322468</td><td>-0.13030672</td><td>0.036938472</td><td>0.000195323</td><td>0.058725572</td></tr>
<tr><td>sp|Q86WI1|PKHL1_HUMAN</td><td>7</td><td>40</td><td>76</td><td>33</td><td>0.261613</td><td>0.138643501</td><td>-0.016939501</td><td>-0.122969889</td><td>-0.278552891</td><td>-0.155583002</td><td>0.034926853</td><td>1.61E-05</td><td>0.009527171</td></tr>
<tr><td>sp|P0DJI8|SAA1_HUMAN</td><td>3</td><td>40</td><td>76</td><td>33</td><td>-1.49496</td><td>-1.407497905</td><td>-0.298253885</td><td>0.087458983</td><td>1.196703003</td><td>1.10924402</td><td>0.927021245</td><td>0.000128249</td><td>5.95E-05</td></tr>
<tr><td>sp|P06727|APOA4_HUMAN</td><td>15</td><td>40</td><td>76</td><td>33</td><td>0.476604</td><td>0.293984127</td><td>0.20275537</td><td>-0.182619551</td><td>-0.273848308</td><td>-0.091228758</td><td>0.009055007</td><td>0.000801025</td><td>0.343088035</td></tr>
<tr><td>sp|P10646|TFPI1_HUMAN</td><td>16</td><td>40</td><td>76</td><td>33</td><td>0.047449</td><td>0.049878758</td><td>0.307139095</td><td>0.002429759</td><td>0.259690097</td><td>0.257260338</td><td>0.998206756</td><td>3.61E-06</td><td>2.31E-07</td></tr>
</table>

<table>
<tr><th></th><th></th><th></th><th></th><th></th><th></th><th></th><th></th><th></th><th></th><th></th><th></th><th></th><th></th></tr>
<tr><td>sp|Q96GW7|PGCB_HUMAN</td><td>8</td><td>19</td><td>34</td><td>15</td><td>0.142379</td><td>0.045962672</td><td>0.297592569</td><td>-0.096416448</td><td>0.155213449</td><td>0.251629897</td><td>0.632457668</td><td>0.44441364</td><td>0.077305113</td></tr>
<tr><td>sp|P23142|FBLN1_HUMAN</td><td>2</td><td>40</td><td>76</td><td>33</td><td>0.171948</td><td>0.131944157</td><td>0.189187651</td><td>-0.040003496</td><td>0.017239998</td><td>0.057243494</td><td>0.560052504</td><td>0.928028662</td><td>0.354448728</td></tr>
<tr><td>sp|P80108|PHLD_HUMAN</td><td>11</td><td>40</td><td>76</td><td>33</td><td>0.224388</td><td>0.191833943</td><td>0.010829548</td><td>-0.032554367</td><td>-0.213558763</td><td>-0.181004396</td><td>0.873924571</td><td>0.021277925</td><td>0.029284466</td></tr>
<tr><td>sp|P10720|PF4V_HUMAN</td><td>3</td><td>40</td><td>76</td><td>33</td><td>-0.20378</td><td>0.094783359</td><td>0.113452321</td><td>0.298564675</td><td>0.317233636</td><td>0.018668962</td><td>0.008776218</td><td>0.024082417</td><td>0.983077694</td></tr>
<tr><td>sp|P04155|TFF1_HUMAN</td><td>16</td><td>18</td><td>34</td><td>16</td><td>-0.01114</td><td>-0.020211569</td><td>0.18852687</td><td>-0.009071029</td><td>0.19966741</td><td>0.208738439</td><td>0.99836213</td><td>0.56811451</td><td>0.453693795</td></tr>
<tr><td>sp|P02656|APOC3_HUMAN</td><td>5</td><td>40</td><td>76</td><td>33</td><td>0.221771</td><td>0.162782576</td><td>0.4740876</td><td>-0.058988509</td><td>0.252316515</td><td>0.311305025</td><td>0.807551893</td><td>0.07201966</td><td>0.006893779</td></tr>
<tr><td>sp|P55290|CAD13_HUMAN</td><td>1</td><td>40</td><td>76</td><td>33</td><td>0.333972</td><td>0.140759437</td><td>-0.207260745</td><td>-0.193212257</td><td>-0.541232438</td><td>-0.348020181</td><td>0.000938589</td><td>7.99E-14</td><td>1.51E-08</td></tr>
<tr><td>sp|P01009|A1AT_HUMAN</td><td>2</td><td>40</td><td>76</td><td>33</td><td>-1.91531</td><td>-1.925159911</td><td>-1.359917513</td><td>-0.009849232</td><td>0.555393166</td><td>0.565242397</td><td>0.999129388</td><td>0.153587816</td><td>0.085909041</td></tr>
<tr><td>sp|P13533|MYH6_HUMAN</td><td>15</td><td>40</td><td>76</td><td>33</td><td>0.112691</td><td>-0.09842701</td><td>0.277521771</td><td>-0.211117759</td><td>0.164831022</td><td>0.375948781</td><td>0.111642746</td><td>0.392842808</td><td>0.002787167</td></tr>
<tr><td>sp|Q9UKX2|MYH2_HUMAN</td><td>1</td><td>40</td><td>76</td><td>33</td><td>0.173455</td><td>-0.025016859</td><td>0.138616476</td><td>-0.198471738</td><td>-0.034838403</td><td>0.163633335</td><td>0.164007068</td><td>0.961597347</td><td>0.337190016</td></tr>
<tr><td>sp|P04004|VTNC_HUMAN</td><td>12</td><td>40</td><td>76</td><td>33</td><td>0.21222</td><td>0.288935156</td><td>0.05230798</td><td>0.076715254</td><td>-0.159911922</td><td>-0.236627177</td><td>0.066549873</td><td>0.000437951</td><td>3.58E-09</td></tr>
<tr><td>sp|Q07654|TFF3_HUMAN</td><td>32</td><td>40</td><td>76</td><td>33</td><td>-0.41037</td><td>-0.215905465</td><td>0.115910997</td><td>0.194461453</td><td>0.526277915</td><td>0.331816461</td><td>0.310879888</td><td>0.003542831</td><td>0.053262113</td></tr>
<tr><td>sp|P02753|RET4_HUMAN</td><td>3</td><td>40</td><td>76</td><td>33</td><td>0.196101</td><td>0.143449374</td><td>0.094783459</td><td>-0.052651599</td><td>-0.101317514</td><td>-0.048665915</td><td>0.482983321</td><td>0.15915836</td><td>0.578715956</td></tr>
<tr><td>sp|P69905|HBA_HUMAN</td><td>6</td><td>40</td><td>76</td><td>33</td><td>-1.04551</td><td>-0.791892941</td><td>0.079772118</td><td>0.253621312</td><td>1.125286372</td><td>0.871665059</td><td>0.439388525</td><td>3.75E-05</td><td>0.000354536</td></tr>
<tr><td>sp|P02790|HEMO_HUMAN</td><td>10</td><td>40</td><td>76</td><td>33</td><td>0.423362</td><td>0.385298989</td><td>0.085124151</td><td>-0.038062587</td><td>-0.338237426</td><td>-0.300174839</td><td>0.684449438</td><td>2.29E-08</td><td>2.22E-08</td></tr>
<tr><td>sp|P13667|PDIA4_HUMAN</td><td>29</td><td>40</td><td>76</td><td>33</td><td>-0.09626</td><td>-0.024321028</td><td>0.40313688</td><td>0.071943903</td><td>0.499401812</td><td>0.427457909</td><td>0.576811532</td><td>1.32E-07</td><td>3.42E-07</td></tr>
<tr><td>sp|P20851|C4BPB_HUMAN</td><td>5</td><td>40</td><td>76</td><td>33</td><td>0.053194</td><td>0.156823727</td><td>0.170295658</td><td>0.103629974</td><td>0.117101904</td><td>0.013471931</td><td>0.104588563</td><td>0.136073962</td><td>0.966287641</td></tr>
<tr><td>sp|P04278|SHBG_HUMAN</td><td>23</td><td>40</td><td>76</td><td>33</td><td>0.101983</td><td>0.191028486</td><td>-0.077847375</td><td>0.08904516</td><td>-0.179830702</td><td>-0.268875862</td><td>0.648594635</td><td>0.298674747</td><td>0.034670739</td></tr>
<tr><td>sp|Q01524|DEF6_HUMAN</td><td>13</td><td>12</td><td>20</td><td>9</td><td>-0.06602</td><td>0.246062121</td><td>0.667489404</td><td>0.312085555</td><td>0.733512838</td><td>0.421427283</td><td>0.909945022</td><td>0.701405933</td><td>0.867441954</td></tr>
<tr><td>sp|P09871|C1S_HUMAN</td><td>4</td><td>40</td><td>76</td><td>33</td><td>0.241875</td><td>0.221825844</td><td>0.19190511</td><td>-0.020049246</td><td>-0.049969981</td><td>-0.029920735</td><td>0.657011264</td><td>0.169509586</td><td>0.441355296</td></tr>
<tr><td>sp|P03951|FA11_HUMAN</td><td>13</td><td>40</td><td>76</td><td>33</td><td>0.221177</td><td>0.192625627</td><td>0.01123571</td><td>-0.02855118</td><td>-0.209941097</td><td>-0.181389917</td><td>0.693617638</td><td>5.16E-06</td><td>8.97E-06</td></tr>
<tr><td>sp|P17813|EGLN_HUMAN</td><td>1</td><td>40</td><td>76</td><td>33</td><td>0.214475</td><td>0.130168854</td><td>-0.128369124</td><td>-0.084306448</td><td>-0.342844426</td><td>-0.258537978</td><td>0.048102541</td><td>8.27E-13</td><td>5.77E-10</td></tr>
<tr><td>sp|Q13508|NAR3_HUMAN</td><td>3</td><td>40</td><td>76</td><td>33</td><td>0.132532</td><td>0.021468105</td><td>-0.01286212</td><td>-0.11106406</td><td>-0.145394285</td><td>-0.034330225</td><td>0.233063062</td><td>0.179517423</td><td>0.883456331</td></tr>
<tr><td>sp|A0A0C4DH38|HV551_HUMAN</td><td>3</td><td>31</td><td>55</td><td>23</td><td>-0.26932</td><td>-0.261574446</td><td>0.203326677</td><td>0.007740612</td><td>0.472641735</td><td>0.464901123</td><td>0.997544543</td><td>0.003426684</td><td>0.001275641</td></tr>
<tr><td>sp|P02743|SAMP_HUMAN</td><td>5</td><td>40</td><td>76</td><td>33</td><td>0.188159</td><td>0.242545292</td><td>0.166038375</td><td>0.054386396</td><td>-0.022120521</td><td>-0.076506917</td><td>0.691742133</td><td>0.958665302</td><td>0.528096233</td></tr>
<tr><td>sp|P02776|PLF4_HUMAN</td><td>3</td><td>40</td><td>76</td><td>33</td><td>-0.58204</td><td>-0.096470592</td><td>-0.010707919</td><td>0.48556528</td><td>0.571327954</td><td>0.085762674</td><td>0.002579096</td><td>0.003320988</td><td>0.840999745</td></tr>
<tr><td>sp|P01011|AACT_HUMAN</td><td>20</td><td>40</td><td>76</td><td>33</td><td>0.236646</td><td>0.280630269</td><td>0.506864728</td><td>0.043984666</td><td>0.270219125</td><td>0.226234459</td><td>0.736588288</td><td>0.000603493</td><td>0.001282076</td></tr>
<tr><td>sp|Q96KN2|CNDP1_HUMAN</td><td>19</td><td>40</td><td>76</td><td>33</td><td>-0.22405</td><td>0.367072691</td><td>-0.132794291</td><td>0.591119663</td><td>0.091252681</td><td>-0.499866982</td><td>6.12E-11</td><td>0.621036187</td><td>1.45E-07</td></tr>
<tr><td>sp|P02741|CRP_HUMAN</td><td>2</td><td>40</td><td>76</td><td>33</td><td>-1.59979</td><td>-1.206699544</td><td>-0.157644791</td><td>0.393095333</td><td>1.442150086</td><td>1.049054753</td><td>0.270229787</td><td>1.58E-05</td><td>0.000462012</td></tr>
<tr><td>sp|P12821|ACE_HUMAN</td><td>7</td><td>40</td><td>76</td><td>33</td><td>0.183914</td><td>0.084674064</td><td>-0.042393981</td><td>-0.099239882</td><td>-0.226307927</td><td>-0.127068045</td><td>0.176173141</td><td>0.002552868</td><td>0.083738505</td></tr>
<tr><td>sp|P19320|VCAM1_HUMAN</td><td>3</td><td>40</td><td>76</td><td>33</td><td>-0.01759</td><td>0.043719901</td><td>0.388243903</td><td>0.061308299</td><td>0.405832301</td><td>0.344524003</td><td>0.623898377</td><td>3.11E-06</td><td>8.07E-06</td></tr>
<tr><td>sp|Q9UIQ6|LCAP_HUMAN</td><td>2</td><td>30</td><td>56</td><td>24</td><td>-0.10896</td><td>0.073283857</td><td>0.079709963</td><td>0.182247515</td><td>0.18867362</td><td>0.006426106</td><td>0.025795297</td><td>0.066672697</td><td>0.995908679</td></tr>
<tr><td>sp|Q6P179|ERAP2_HUMAN</td><td>2</td><td>40</td><td>76</td><td>33</td><td>-0.09493</td><td>0.021851096</td><td>-0.040336698</td><td>0.116776789</td><td>0.054588996</td><td>-0.062187793</td><td>0.428242078</td><td>0.879104589</td><td>0.808484771</td></tr>
<tr><td>sp|P25311|ZA2G_HUMAN</td><td>3</td><td>40</td><td>76</td><td>33</td><td>0.343441</td><td>0.27863225</td><td>0.292038591</td><td>-0.064808752</td><td>-0.051402411</td><td>0.013406341</td><td>0.204325344</td><td>0.498679806</td><td>0.941166429</td></tr>
<tr><td>sp|P10909|CLUS_HUMAN</td><td>5</td><td>40</td><td>76</td><td>33</td><td>0.165864</td><td>0.196900202</td><td>0.155073487</td><td>0.031036237</td><td>-0.010790478</td><td>-0.041826715</td><td>0.455511158</td><td>0.936048703</td><td>0.287134249</td></tr>
<tr><td>sp|Q96IY4|CBPB2_HUMAN</td><td>5</td><td>40</td><td>76</td><td>33</td><td>0.270286</td><td>0.212007721</td><td>0.165369505</td><td>-0.058278214</td><td>-0.10491643</td><td>-0.046638216</td><td>0.104646263</td><td>0.007343349</td><td>0.277444623</td></tr>
<tr><td>sp|P02748|CO9_HUMAN</td><td>62</td><td>40</td><td>76</td><td>33</td><td>0.10426</td><td>0.223412757</td><td>0.37799522</td><td>0.119152462</td><td>0.273734924</td><td>0.154582463</td><td>0.249140859</td><td>0.007565817</td><td>0.130054584</td></tr>
<tr><td>sp|P10912|GHR_HUMAN</td><td>6</td><td>40</td><td>76</td><td>33</td><td>0.107297</td><td>0.138367675</td><td>-0.229596634</td><td>0.03107107</td><td>-0.33689324</td><td>-0.367964309</td><td>0.895504557</td><td>0.000260525</td><td>5.57E-06</td></tr>
<tr><td>sp|P02788|TRFL_HUMAN</td><td>1</td><td>40</td><td>76</td><td>33</td><td>-0.30832</td><td>-0.091070038</td><td>0.23328419</td><td>0.217250021</td><td>0.54160425</td><td>0.324354228</td><td>0.138094139</td><td>0.000335126</td><td>0.022300813</td></tr>
<tr><td>sp|Q01518|CAP1_HUMAN</td><td>19</td><td>40</td><td>76</td><td>33</td><td>-0.13564</td><td>-0.21508362</td><td>0.212788239</td><td>-0.079439276</td><td>0.348432583</td><td>0.427871858</td><td>0.84953234</td><td>0.119981886</td><td>0.018464195</td></tr>
<tr><td>sp|P17936|IBP3_HUMAN</td><td>2</td><td>40</td><td>76</td><td>33</td><td>0.141168</td><td>0.227785605</td><td>-0.23011564</td><td>0.086617658</td><td>-0.371283587</td><td>-0.457901245</td><td>0.258451498</td><td>2.83E-07</td><td>3.01E-12</td></tr>
<tr><td>sp|P60709|ACTB_HUMAN</td><td>2</td><td>40</td><td>76</td><td>33</td><td>0.237027</td><td>0.039583762</td><td>0.33664864</td><td>-0.197442777</td><td>0.099622102</td><td>0.297064878</td><td>0.107104581</td><td>0.670294183</td><td>0.012960373</td></tr>
<tr><td>sp|P02452|CO1A1_HUMAN</td><td>12</td><td>40</td><td>76</td><td>33</td><td>0.058498</td><td>0.071492318</td><td>0.513427475</td><td>0.012994686</td><td>0.454929843</td><td>0.441935157</td><td>0.983715655</td><td>4.45E-06</td><td>4.96E-07</td></tr>
<tr><td>sp|P55103|INHBC_HUMAN</td><td>59</td><td>40</td><td>76</td><td>33</td><td>0.011943</td><td>0.06850657</td><td>0.21263051</td><td>0.056563678</td><td>0.200687617</td><td>0.14412394</td><td>0.444630234</td><td>0.001308926</td><td>0.011671729</td></tr>
<tr><td>sp|Q8N149|LIRA2_HUMAN</td><td>18</td><td>40</td><td>76</td><td>33</td><td>-0.15294</td><td>-0.005983735</td><td>0.132762011</td><td>0.14695934</td><td>0.285705086</td><td>0.138745746</td><td>0.330116406</td><td>0.058495665</td><td>0.419146608</td></tr>
<tr><td>sp|O60493|SNX3_HUMAN</td><td>1</td><td>40</td><td>76</td><td>33</td><td>0.168679</td><td>0.158764672</td><td>0.444420389</td><td>-0.009914419</td><td>0.275741298</td><td>0.285655717</td><td>0.995160238</td><td>0.080323216</td><td>0.032957274</td></tr>
<tr><td>sp|Q6EMK4|VASN_HUMAN</td><td>6</td><td>40</td><td>76</td><td>33</td><td>0.201135</td><td>0.17227202</td><td>0.262538896</td><td>-0.028863394</td><td>0.061403483</td><td>0.090266876</td><td>0.549938871</td><td>0.158356952</td><td>0.007352571</td></tr>
<tr><td>sp|P08185|CBG_HUMAN</td><td>1</td><td>40</td><td>76</td><td>33</td><td>0.268934</td><td>0.35380291</td><td>0.051572744</td><td>0.084869127</td><td>-0.217361039</td><td>-0.302230166</td><td>0.282585922</td><td>0.004167405</td><td>3.30E-06</td></tr>
<tr><td>sp|P04180|LCAT_HUMAN</td><td>9</td><td>40</td><td>76</td><td>33</td><td>0.297369</td><td>0.187365324</td><td>0.132396678</td><td>-0.110003311</td><td>-0.164971958</td><td>-0.054968647</td><td>0.00485323</td><td>0.000317361</td><td>0.295867272</td></tr>
<tr><td>sp|P05019|IGF1_HUMAN</td><td>3</td><td>40</td><td>76</td><td>33</td><td>0.019512</td><td>0.18531131</td><td>-0.221431234</td><td>0.165798946</td><td>-0.240943598</td><td>-0.406742544</td><td>0.050695889</td><td>0.013729169</td><td>6.86E-07</td></tr>
<tr><td>sp|P20930|FILA_HUMAN</td><td>2</td><td>40</td><td>76</td><td>33</td><td>-0.27641</td><td>-0.441185323</td><td>-0.364450302</td><td>-0.164773203</td><td>-0.088038181</td><td>0.076735022</td><td>0.488588649</td><td>0.867669453</td><td>0.871773445</td></tr>
<tr><td>sp|Q8N6C8|LIRA3_HUMAN</td><td>2</td><td>40</td><td>76</td><td>33</td><td>0.15619</td><td>-0.156127833</td><td>-0.574352068</td><td>-0.312317567</td><td>-0.730541802</td><td>-0.418224235</td><td>0.352749162</td><td>0.021870188</td><td>0.195850565</td></tr>
<tr><td>sp|O14793|GDF8_HUMAN</td><td>1</td><td>40</td><td>76</td><td>33</td><td>0.237663</td><td>0.061276731</td><td>-0.339960324</td><td>-0.176386175</td><td>-0.577623229</td><td>-0.401237054</td><td>0.034889837</td><td>6.40E-10</td><td>9.89E-07</td></tr>
<tr><td>sp|O14960|LECT2_HUMAN</td><td>1</td><td>40</td><td>76</td><td>33</td><td>-0.06441</td><td>0.044816637</td><td>-0.059175919</td><td>0.109229584</td><td>0.005237029</td><td>-0.103992555</td><td>0.311851408</td><td>0.998130602</td><td>0.394708186</td></tr>
<tr><td>sp|P55854|SUMO3_HUMAN</td><td>2</td><td>40</td><td>76</td><td>33</td><td>-0.28427</td><td>-0.249271797</td><td>0.261246645</td><td>0.035002597</td><td>0.545521039</td><td>0.510518442</td><td>0.947932039</td><td>0.000259733</td><td>0.000108735</td></tr>
<tr><td>sp|P20160|CAP7_HUMAN</td><td>18</td><td>36</td><td>70</td><td>30</td><td>-0.57776</td><td>-0.447710242</td><td>0.019108455</td><td>0.130054229</td><td>0.596872927</td><td>0.466818697</td><td>0.609082819</td><td>0.001203984</td><td>0.00473791</td></tr>
<tr><td>sp|O75023|LIRB5_HUMAN</td><td>11</td><td>40</td><td>76</td><td>33</td><td>0.360687</td><td>0.391278458</td><td>0.306958696</td><td>0.030591692</td><td>-0.053728071</td><td>-0.084319763</td><td>0.759711187</td><td>0.558236778</td><td>0.164681596</td></tr>
<tr><td>sp|P02042|HBD_HUMAN</td><td>11</td><td>40</td><td>76</td><td>33</td><td>-1.30185</td><td>-1.121303744</td><td>-0.074072849</td><td>0.180545029</td><td>1.227775924</td><td>1.047230895</td><td>0.734887705</td><td>0.000121476</td><td>0.000226089</td></tr>
<tr><td>sp|P00746|CFAD_HUMAN</td><td>46</td><td>40</td><td>76</td><td>33</td><td>0.818716</td><td>0.803912318</td><td>0.905619649</td><td>-0.014803848</td><td>0.086903482</td><td>0.10170733</td><td>0.924500948</td><td>0.160050256</td><td>0.042830876</td></tr>
<tr><td>sp|P00738|HPT_HUMAN</td><td>4</td><td>40</td><td>76</td><td>33</td><td>-1.2273</td><td>-0.770042817</td><td>-0.442344916</td><td>0.457259866</td><td>0.784957767</td><td>0.327697902</td><td>0.173159885</td><td>0.030135194</td><td>0.449916211</td></tr>
<tr><td>sp|P07360|CO8G_HUMAN</td><td>3</td><td>40</td><td>76</td><td>33</td><td>0.216168</td><td>0.196732118</td><td>0.130749923</td><td>-0.019435722</td><td>-0.085417917</td><td>-0.065982195</td><td>0.775132099</td><td>0.037289824</td><td>0.080694446</td></tr>
<tr><td>sp|P07225|PROS_HUMAN</td><td>2</td><td>40</td><td>76</td><td>33</td><td>0.11409</td><td>0.181690018</td><td>0.163605324</td><td>0.067600115</td><td>0.049515421</td><td>-0.018084695</td><td>0.114147387</td><td>0.442689347</td><td>0.869949445</td></tr>
<tr><td>sp|P02675|FIBB_HUMAN</td><td>4</td><td>40</td><td>76</td><td>33</td><td>0.025417</td><td>0.158020323</td><td>0.308707445</td><td>0.132603476</td><td>0.283290598</td><td>0.150687122</td><td>0.130608293</td><td>0.002129923</td><td>0.100106426</td></tr>
<tr><td>sp|P04003|C4BPA_HUMAN</td><td>3</td><td>40</td><td>76</td><td>33</td><td>0.633682</td><td>0.757206224</td><td>0.757715697</td><td>0.123523792</td><td>0.124033265</td><td>0.000509473</td><td>0.056196178</td><td>0.132578508</td><td>0.999955686</td></tr>
<tr><td>sp|O00533|NCHL1_HUMAN</td><td>1</td><td>40</td><td>76</td><td>33</td><td>0.268685</td><td>0.17453486</td><td>-0.019263519</td><td>-0.094150124</td><td>-0.287948503</td><td>-0.19379838</td><td>0.129122816</td><td>6.07E-06</td><td>0.000722078</td></tr>
<tr><td>sp|Q92954|PRG4_HUMAN</td><td>3</td><td>40</td><td>76</td><td>33</td><td>0.08497</td><td>0.077784073</td><td>0.130829934</td><td>-0.007185867</td><td>0.045859994</td><td>0.053045861</td><td>0.990911406</td><td>0.774217312</td><td>0.647367757</td></tr>
<tr><td>sp|P00739|HPTR_HUMAN</td><td>9</td><td>40</td><td>76</td><td>33</td><td>-1.62603</td><td>-1.125860984</td><td>-0.718688342</td><td>0.500169478</td><td>0.90734212</td><td>0.407172642</td><td>0.185484346</td><td>0.02362249</td><td>0.372511091</td></tr>
<tr><td>sp|P02652|APOA2_HUMAN</td><td>14</td><td>40</td><td>76</td><td>33</td><td>-0.1989</td><td>-0.144776494</td><td>0.317291289</td><td>0.054123694</td><td>0.516191478</td><td>0.462067784</td><td>0.866158476</td><td>0.000245718</td><td>0.000211242</td></tr>
<tr><td>sp|Q9UK55|ZPI_HUMAN</td><td>4</td><td>40</td><td>76</td><td>33</td><td>0.268548</td><td>0.305477679</td><td>0.347612865</td><td>0.036930007</td><td>0.079065193</td><td>0.042135186</td><td>0.654359739</td><td>0.264877484</td><td>0.616067045</td></tr>
<tr><td>sp|Q02809|PLOD1_HUMAN</td><td>1</td><td>40</td><td>76</td><td>33</td><td>-0.05562</td><td>-0.017286833</td><td>0.180829571</td><td>0.038330804</td><td>0.236447207</td><td>0.198116403</td><td>0.760516436</td><td>0.001190867</td><td>0.002347623</td></tr>
<tr><td>sp|P29622|KAIN_HUMAN</td><td>18</td><td>40</td><td>76</td><td>33</td><td>0.434602</td><td>0.345553386</td><td>0.086601537</td><td>-0.089048728</td><td>-0.348000578</td><td>-0.258951849</td><td>0.135563766</td><td>1.31E-08</td><td>1.63E-06</td></tr>
<tr><td>sp|O15031|PLXB2_HUMAN</td><td>8</td><td>40</td><td>76</td><td>33</td><td>0.045856</td><td>0.080321344</td><td>0.15566624</td><td>0.034465635</td><td>0.10981053</td><td>0.075344896</td><td>0.45473795</td><td>0.00512898</td><td>0.039797634</td></tr>
<tr><td>sp|P40199|CEAM6_HUMAN</td><td>2</td><td>40</td><td>76</td><td>33</td><td>-0.03592</td><td>-0.090815561</td><td>0.295536417</td><td>-0.054896018</td><td>0.33145596</td><td>0.386351978</td><td>0.683522071</td><td>0.000150248</td><td>5.20E-07</td></tr>
<tr><td>sp|P04075|ALDOA_HUMAN</td><td>1</td><td>40</td><td>76</td><td>33</td><td>0.105665</td><td>0.006184584</td><td>0.282817789</td><td>-0.099479951</td><td>0.177153255</td><td>0.276633205</td><td>0.446964704</td><td>0.174698348</td><td>0.005373369</td></tr>
<tr><td>sp|P0DP58|LYNX1_HUMAN</td><td>1</td><td>40</td><td>76</td><td>33</td><td>0.145454</td><td>0.081168272</td><td>0.304881029</td><td>-0.064286025</td><td>0.159426732</td><td>0.223712757</td><td>0.710162315</td><td>0.237952146</td><td>0.029573822</td></tr>
<tr><td>sp|O95274|LYPD3_HUMAN</td><td>4</td><td>40</td><td>76</td><td>33</td><td>0.256415</td><td>0.19889304</td><td>0.013323547</td><td>-0.057521996</td><td>-0.243091488</td><td>-0.185569492</td><td>0.508378716</td><td>0.000423561</td><td>0.002837779</td></tr>
<tr><td>sp|Q08830|FGL1_HUMAN</td><td>1</td><td>40</td><td>76</td><td>33</td><td>-0.25487</td><td>-0.103852066</td><td>0.169516982</td><td>0.151018458</td><td>0.424387506</td><td>0.273369048</td><td>0.515374528</td><td>0.029989488</td><td>0.152180095</td></tr>
<tr><td>sp|Q6UXG3|CLM9_HUMAN</td><td>8</td><td>28</td><td>56</td><td>25</td><td>0.08453</td><td>0.086446592</td><td>0.176109581</td><td>0.001916283</td><td>0.091579272</td><td>0.089662989</td><td>0.999853786</td><td>0.790133384</td><td>0.744341458</td></tr>
<tr><td>sp|P00488|F13A_HUMAN</td><td>13</td><td>40</td><td>76</td><td>33</td><td>0.317704</td><td>0.24272329</td><td>0.013630564</td><td>-0.074981149</td><td>-0.304073875</td><td>-0.229092726</td><td>0.343033735</td><td>1.63E-05</td><td>0.000282261</td></tr>
<tr><td>sp|P05067|A4_HUMAN</td><td>4</td><td>40</td><td>76</td><td>33</td><td>0.016022</td><td>0.314741441</td><td>0.428218449</td><td>0.298719648</td><td>0.412196656</td><td>0.113477007</td><td>0.005028624</td><td>0.001060048</td><td>0.495245505</td></tr>
<tr><td>sp|P18428|LBP_HUMAN</td><td>2</td><td>40</td><td>76</td><td>33</td><td>-0.21929</td><td>-0.065429402</td><td>0.382062514</td><td>0.153863522</td><td>0.601355437</td><td>0.447491915</td><td>0.283544691</td><td>6.25E-06</td><td>0.000166843</td></tr>
<tr><td>sp|P06276|CHLE_HUMAN</td><td>26</td><td>40</td><td>76</td><td>33</td><td>0.225733</td><td>0.214939508</td><td>-0.131891973</td><td>-0.010793668</td><td>-0.357625149</td><td>-0.346831481</td><td>0.980177982</td><td>1.61E-06</td><td>1.60E-07</td></tr>
<tr><td>sp|Q9UHI8|ATS1_HUMAN</td><td>9</td><td>29</td><td>55</td><td>23</td><td>-0.35231</td><td>-0.324055165</td><td>0.290309574</td><td>0.028251171</td><td>0.642615911</td><td>0.614364739</td><td>0.968745574</td><td>5.51E-05</td><td>1.43E-05</td></tr>
<tr><td>sp|P15169|CBPN_HUMAN</td><td>3</td><td>40</td><td>76</td><td>33</td><td>0.234439</td><td>0.222964711</td><td>0.127038383</td><td>-0.011474384</td><td>-0.107400711</td><td>-0.095926327</td><td>0.933258193</td><td>0.018164174</td><td>0.017137539</td></tr>
<tr><td>sp|P06681|CO2_HUMAN</td><td>4</td><td>40</td><td>76</td><td>33</td><td>0.257948</td><td>0.269237325</td><td>0.232042223</td><td>0.011289163</td><td>-0.025905939</td><td>-0.037195102</td><td>0.905503455</td><td>0.697912676</td><td>0.391538956</td></tr>
<tr><td>sp|Q14019|COTL1_HUMAN</td><td>29</td><td>40</td><td>76</td><td>33</td><td>-0.12622</td><td>-0.108411016</td><td>0.279818169</td><td>0.017806708</td><td>0.406035893</td><td>0.388229185</td><td>0.988717309</td><td>0.020055713</td><td>0.010888462</td></tr>
<tr><td>sp|Q8N6Q3|CD177_HUMAN</td><td>1</td><td>36</td><td>69</td><td>30</td><td>-0.15533</td><td>-0.153645902</td><td>0.051699652</td><td>0.001688895</td><td>0.207034449</td><td>0.205345554</td><td>0.999893181</td><td>0.333999825</td><td>0.253050528</td></tr>
<tr><td>sp|P01023|A2MG_HUMAN</td><td>4</td><td>40</td><td>76</td><td>33</td><td>-0.07644</td><td>0.063439071</td><td>-0.074415787</td><td>0.139874833</td><td>0.002019974</td><td>-0.137854858</td><td>0.698392459</td><td>0.999948148</td><td>0.736160161</td></tr>
<tr><td>sp|O15394|NCAM2_HUMAN</td><td>6</td><td>40</td><td>76</td><td>33</td><td>0.256714</td><td>0.215811827</td><td>-0.041094738</td><td>-0.040902549</td><td>-0.297809114</td><td>-0.256906565</td><td>0.655963732</td><td>1.25E-06</td><td>2.38E-06</td></tr>
<tr><td>sp|P54108|CRIS3_HUMAN</td><td>9</td><td>40</td><td>76</td><td>33</td><td>0.18653</td><td>0.120707831</td><td>-0.152836664</td><td>-0.065822346</td><td>-0.339366841</td><td>-0.273544495</td><td>0.46072347</td><td>3.12E-06</td><td>2.32E-05</td></tr>
<tr><td>sp|P55899|FCGRN_HUMAN</td><td>12</td><td>40</td><td>76</td><td>33</td><td>0.009076</td><td>-0.0424877</td><td>-0.025923963</td><td>-0.051563984</td><td>-0.035000247</td><td>0.016563737</td><td>0.675356636</td><td>0.882377034</td><td>0.964933352</td></tr>
<tr><td>sp|P20333|TNR1B_HUMAN</td><td>9</td><td>40</td><td>76</td><td>33</td><td>-0.36902</td><td>-0.274144495</td><td>0.237347694</td><td>0.094875861</td><td>0.60636805</td><td>0.511492189</td><td>0.586700494</td><td>1.68E-06</td><td>5.23E-06</td></tr>
<tr><td>sp|Q15485|FCN2_HUMAN</td><td>10</td><td>40</td><td>76</td><td>33</td><td>-0.19451</td><td>-0.015030303</td><td>0.228479485</td><td>0.17947547</td><td>0.422985258</td><td>0.243509788</td><td>0.05694457</td><td>3.60E-05</td><td>0.01052918</td></tr>
<tr><td>sp|P35542|SAA4_HUMAN</td><td>6</td><td>40</td><td>76</td><td>33</td><td>-0.13964</td><td>-0.023872265</td><td>0.361113266</td><td>0.115762853</td><td>0.500748385</td><td>0.384985532</td><td>0.420096667</td><td>3.65E-05</td><td>0.000387728</td></tr>
<tr><td>sp|P55145|MANF_HUMAN</td><td>3</td><td>40</td><td>76</td><td>33</td><td>-0.47527</td><td>-0.319106945</td><td>0.032883602</td><td>0.156162951</td><td>0.508153498</td><td>0.351990547</td><td>0.446045571</td><td>0.003635656</td><td>0.03013312</td></tr>
<tr><td>sp|Q07954|LRP1_HUMAN</td><td>23</td><td>40</td><td>76</td><td>33</td><td>0.128293</td><td>0.193142263</td><td>0.358311952</td><td>0.064849537</td><td>0.230019226</td><td>0.165169688</td><td>0.129246258</td><td>1.60E-07</td><td>2.21E-05</td></tr>
<tr><td>sp|P13591|NCAM1_HUMAN</td><td>49</td><td>40</td><td>76</td><td>33</td><td>0.372043</td><td>0.241018327</td><td>-0.015935957</td><td>-0.131024737</td><td>-0.387979021</td><td>-0.256954284</td><td>0.001914771</td><td>8.40E-14</td><td>6.23E-09</td></tr>
<tr><td>sp|O00115|DNS2A_HUMAN</td><td>9</td><td>32</td><td>62</td><td>27</td><td>0.140401</td><td>0.044778938</td><td>0.180659758</td><td>-0.095622073</td><td>0.040258746</td><td>0.13588082</td><td>0.249031773</td><td>0.840688616</td><td>0.084637686</td></tr>
<tr><td>sp|P06865|HEXA_HUMAN</td><td>6</td><td>40</td><td>76</td><td>33</td><td>0.026034</td><td>-0.009466199</td><td>0.222623866</td><td>-0.035499969</td><td>0.196590096</td><td>0.232090066</td><td>0.704520101</td><td>0.000973447</td><td>7.77E-06</td></tr>
<tr><td>sp|P23528|COF1_HUMAN</td><td>2</td><td>40</td><td>76</td><td>33</td><td>-0.12173</td><td>-0.156557967</td><td>0.220751276</td><td>-0.034831769</td><td>0.342477474</td><td>0.377309244</td><td>0.973123653</td><td>0.167993506</td><td>0.06532846</td></tr>
<tr><td>sp|Q9Y287|ITM2B_HUMAN</td><td>2</td><td>40</td><td>76</td><td>33</td><td>-0.01076</td><td>0.101328262</td><td>0.336154917</td><td>0.112092871</td><td>0.346919527</td><td>0.234826655</td><td>0.080069071</td><td>3.48E-07</td><td>0.000108944</td></tr>
<tr><td>sp|P05062|ALDOB_HUMAN</td><td>19</td><td>40</td><td>76</td><td>33</td><td>-0.14971</td><td>0.064768901</td><td>0.198475965</td><td>0.214477543</td><td>0.348184606</td><td>0.133707063</td><td>0.148071743</td><td>0.032716014</td><td>0.516665838</td></tr>
<tr><td>sp|P06733|ENOA_HUMAN</td><td>6</td><td>40</td><td>76</td><td>33</td><td>-0.07376</td><td>0.026964738</td><td>0.270491384</td><td>0.10072595</td><td>0.344252596</td><td>0.243526646</td><td>0.598562448</td><td>0.018492999</td><td>0.075951998</td></tr>
<tr><td>sp|Q9UGM5|FETUB_HUMAN</td><td>7</td><td>40</td><td>76</td><td>33</td><td>0.132667</td><td>0.308372085</td><td>-0.043334702</td><td>0.175704616</td><td>-0.176002171</td><td>-0.351706787</td><td>0.00635172</td><td>0.028572533</td><td>1.02E-07</td></tr>
<tr><td>sp|P18206|VINC_HUMAN</td><td>1</td><td>40</td><td>76</td><td>33</td><td>0.13035</td><td>-0.086466733</td><td>0.280668598</td><td>-0.216816696</td><td>0.150318635</td><td>0.367135331</td><td>0.094420788</td><td>0.451314624</td><td>0.003204322</td></tr>
</table>

| | | | | | | | | | | | | |
|---|---|---|---|---|---|---|---|---|---|---|---|---|
| sp\|P10451\|OSTP_HUMAN | 1 | 40 | 76 | 33 | -0.46963 | -0.42325956 | 0.212183335 | 0.046374177 | 0.681817072 | 0.635442895 | 0.927180052 | 3.69E-05 | 1.39E-05 |
| sp\|P13671\|CO6_HUMAN | 1 | 40 | 76 | 33 | 0.268222 | 0.292828394 | 0.192802361 | 0.024606565 | -0.075419468 | -0.100026033 | 0.696417936 | 0.100461276 | 0.006704118 |
| sp\|P00736\|C1R_HUMAN | 7 | 40 | 76 | 33 | 0.263002 | 0.278652434 | 0.206691652 | 0.015650655 | -0.056310127 | -0.071960782 | 0.776728213 | 0.10961565 | 0.011227609 |
| sp\|P08493\|MGP_HUMAN | 6 | 40 | 76 | 33 | -0.09021 | -0.133145379 | 0.30744763 | -0.042930582 | 0.397662427 | 0.440593009 | 0.799246921 | 7.14E-06 | 2.24E-08 |
| sp\|P05164\|PERM_HUMAN | 4 | 40 | 76 | 33 | -0.02845 | 0.16398756 | 0.755639835 | 0.192440151 | 0.784092425 | 0.591652274 | 0.187003801 | 5.57E-08 | 3.57E-06 |
| sp\|P21333\|FLNA_HUMAN | 6 | 40 | 76 | 33 | 0.147181 | 0.423527811 | 0.675556589 | 0.276347163 | 0.52837594 | 0.252028777 | 0.088828977 | 0.002744037 | 0.168677358 |
| sp\|P13796\|PLSL_HUMAN | 4 | 40 | 76 | 33 | -0.01179 | 0.127697593 | 0.329261749 | 0.139487722 | 0.341051878 | 0.201564155 | 0.051847071 | 1.27E-05 | 0.004990522 |
| sp\|Q9H2G2\|SLK_HUMAN | 5 | 4 | 7 | 3 | 0.098117 | 0.24747819 | -0.114534379 | 0.149361201 | -0.212651368 | -0.362012569 | 0.818073492 | 0.761660575 | 0.402947891 |
| sp\|P04275\|VWF_HUMAN | 4 | 40 | 76 | 33 | -0.07417 | 0.040139918 | 0.739250711 | 0.114305854 | 0.813416647 | 0.699110793 | 0.476963056 | 4.98E-10 | 1.50E-09 |
| sp\|P43251\|BTD_HUMAN | 1 | 40 | 76 | 33 | 0.373401 | 0.23459162 | -0.028390258 | -0.13880981 | -0.401791688 | -0.262981878 | 0.000882448 | 4.55E-14 | 2.49E-09 |
| sp\|P02679\|FIBG_HUMAN | 1 | 40 | 76 | 33 | 0.036672 | 0.18714675 | 0.348800995 | 0.150475116 | 0.312129361 | 0.161654245 | 0.114617239 | 0.002074144 | 0.111437757 |
| sp\|P02786\|TFR1_HUMAN | 85 | 40 | 76 | 33 | 0.219743 | -0.047019844 | -0.065312901 | -0.266763267 | -0.285056324 | -0.018293057 | 0.055946135 | 0.101707369 | 0.987807052 |
| sp\|P26038\|MOES_HUMAN | 3 | 40 | 76 | 33 | 0.096381 | 0.078402148 | 0.292072805 | -0.017979244 | 0.195691413 | 0.213670657 | 0.961594421 | 0.045034603 | 0.009739573 |
| sp\|P25786\|PSA1_HUMAN | 6 | 40 | 76 | 33 | -0.2358 | -0.035786028 | 0.594118143 | 0.200014532 | 0.829918702 | 0.629904171 | 0.207745046 | 8.53E-08 | 4.35E-06 |
| sp\|Q9BYE9\|CDHR2_HUMAN | 2 | 40 | 76 | 33 | -0.81147 | -0.475053696 | 0.603392965 | 0.336420213 | 1.414866874 | 1.078446661 | 0.068436003 | 3.04E-12 | 1.15E-09 |
| sp\|O95810\|CAVN2_HUMAN | 1 | 40 | 76 | 33 | -0.19948 | -0.345327676 | -0.086786429 | -0.145850259 | 0.112690988 | 0.258541247 | 0.675789397 | 0.850600235 | 0.341864424 |
| sp\|P02750\|A2GL_HUMAN | 2 | 40 | 76 | 33 | -0.32463 | -0.077311582 | 0.427606841 | 0.247318626 | 0.752237048 | 0.504918423 | 0.165443353 | 2.61E-05 | 0.001847175 |
| sp\|P08118\|MSMB_HUMAN | 2 | 40 | 76 | 33 | -0.011 | -0.118385122 | 0.069963078 | -0.107385252 | 0.080962949 | 0.188348201 | 0.284999754 | 0.608780051 | 0.03619074 |
| sp\|P30101\|PDIA3_HUMAN | 2 | 40 | 76 | 33 | 0.084196 | 0.127754601 | 0.215235621 | 0.043558663 | 0.131039682 | 0.08748102 | 0.801015178 | 0.254425101 | 0.457917963 |
| sp\|Q06033\|ITIH3_HUMAN | 18 | 40 | 76 | 33 | 0.042838 | 0.110589409 | 0.421313948 | 0.067751879 | 0.378476418 | 0.310724539 | 0.631664305 | 0.00011367 | 0.000379249 |
| sp\|P46531\|NOTC1_HUMAN | 364 | 40 | 76 | 33 | 0.196245 | 0.202482781 | 0.093114619 | 0.006237937 | -0.103130225 | -0.109368162 | 0.978193302 | 0.018493055 | 0.003626976 |
| sp\|P07358\|CO8B_HUMAN | 2 | 40 | 76 | 33 | 0.614161 | 0.590249516 | 0.528524949 | -0.023911672 | -0.085636239 | -0.061724567 | 0.72154008 | 0.05995803 | 0.152837286 |
| sp\|P07359\|GP1BA_HUMAN | 4 | 40 | 76 | 33 | 0.16117 | 0.105910799 | 0.128622191 | -0.055259357 | -0.032547965 | 0.022711392 | 0.452727914 | 0.826205294 | 0.888362771 |
| sp\|Q9UNW1\|MINP1_HUMAN | 1 | 40 | 76 | 33 | 0.086723 | 0.136121968 | 0.082891529 | 0.049399103 | -0.003831337 | -0.053230439 | 0.231418684 | 0.993850036 | 0.225021468 |
| sp\|P02745\|C1QA_HUMAN | 12 | 40 | 76 | 33 | 0.154021 | 0.295186171 | 0.167036695 | 0.14116556 | 0.013016085 | -0.128149476 | 0.162742766 | 0.989203379 | 0.267011398 |
| sp\|O00187\|MASP2_HUMAN | 7 | 40 | 76 | 33 | 0.271055 | 0.384985026 | 0.387277349 | 0.113929943 | 0.116222266 | 0.002292324 | 0.032290116 | 0.083007146 | 0.99873613 |
| sp\|Q86VB7\|C163A_HUMAN | 2 | 40 | 76 | 33 | -0.1016 | 0.041289818 | 0.315158087 | 0.142894041 | 0.416762309 | 0.273868268 | 0.151870562 | 3.70E-05 | 0.002904192 |
| sp\|Q9UBV2\|SE1L1_HUMAN | 4 | 4 | 6 | 3 | 0.10389 | -0.002173201 | 1.141139113 | -0.106062726 | 1.037249588 | 1.143312314 | 0.979379051 | 0.28664279 | 0.184709389 |
| sp\|Q03591\|FHR1_HUMAN | 1 | 40 | 76 | 33 | 0.108033 | 0.141331317 | 0.107886347 | 0.03329873 | -0.00014624 | -0.03344497 | 0.353197122 | 0.999985996 | 0.397352279 |
| sp\|Q9UN70\|PCDGK_HUMAN | 7 | 40 | 76 | 33 | 0.00834 | 0.033538145 | 0.098508344 | 0.025197975 | 0.090168174 | 0.064970199 | 0.83180425 | 0.201328195 | 0.344762164 |
| sp\|Q6UX71\|PXDC2_HUMAN | 26 | 40 | 76 | 33 | 0.145115 | 0.086980075 | 0.177022739 | -0.058135241 | 0.031907423 | 0.090042664 | 0.211743178 | 0.72118351 | 0.0401906 |
| sp\|P11226\|MBL2_HUMAN | 4 | 40 | 76 | 33 | -0.06025 | -0.165551903 | -0.03387612 | -0.105304554 | 0.026371229 | 0.131675783 | 0.746356527 | 0.987383117 | 0.66955235 |
| sp\|P04196\|HRG_HUMAN | 3 | 40 | 76 | 33 | 0.235597 | 0.211578539 | -0.068086744 | -0.024018191 | -0.303683473 | -0.279665282 | 0.89774752 | 2.20E-05 | 1.02E-05 |
| sp\|Q6UWW8\|EST3_HUMAN | 21 | 33 | 62 | 27 | 0.024089 | -0.065266881 | -0.196851971 | -0.089356077 | -0.220941167 | -0.13158509 | 0.326991903 | 0.0108173 | 0.123394775 |
| sp\|Q8TDL5\|BPIB1_HUMAN | 1 | 36 | 69 | 30 | -0.09242 | 0.015488018 | 0.318556108 | 0.1079037 | 0.41097179 | 0.30306809 | 0.403625948 | 0.000223697 | 0.002511844 |
| sp\|P04070\|PROC_HUMAN | 1 | 40 | 76 | 33 | 0.197748 | 0.158685115 | -0.008921721 | -0.039062865 | -0.206669701 | -0.167606836 | 0.455331422 | 1.42E-06 | 1.05E-05 |
| sp\|P07711\|CATL1_HUMAN | 7 | 40 | 76 | 33 | 0.019604 | 0.067083537 | 0.182751159 | 0.047479851 | 0.163147473 | 0.115667621 | 0.490135204 | 0.0039612 | 0.027160839 |
| sp\|P61626\|LYSC_HUMAN | 6 | 40 | 76 | 33 | -0.0277 | 0.160130659 | 0.29527066 | 0.187826766 | 0.322966767 | 0.135140001 | 0.009006456 | 0.000100498 | 0.111273872 |
| sp\|P08571\|CD14_HUMAN | 1 | 40 | 76 | 33 | 0.057528 | 0.091927644 | 0.221708771 | 0.034399561 | 0.164180688 | 0.129781127 | 0.752027168 | 0.013604476 | 0.031866415 |
| sp\|Q71U36\|TBA1A_HUMAN | 1 | 40 | 76 | 33 | -0.33333 | -0.10018792 | 0.185740916 | 0.233140711 | 0.519069548 | 0.285928837 | 0.158319043 | 0.002372396 | 0.088962793 |
| sp\|P54289\|CA2D1_HUMAN | 13 | 40 | 76 | 33 | 0.293177 | 0.101622661 | -0.343494878 | -0.19155469 | -0.63667223 | -0.445117539 | 0.00716122 | 1.10E-13 | 1.37E-09 |
| sp\|P01024\|CO3_HUMAN | 9 | 40 | 76 | 33 | 0.43664 | 0.381683772 | 0.46079642 | -0.054956372 | 0.024156276 | 0.079112648 | 0.353820881 | 0.869502842 | 0.153526056 |
| sp\|P02747\|C1QC_HUMAN | 9 | 40 | 76 | 33 | -0.04302 | 0.130909731 | -0.039021824 | 0.173926879 | 0.003995324 | -0.169931555 | 0.089472039 | 0.999098964 | 0.131194997 |
| sp\|P02749\|APOH_HUMAN | 1 | 40 | 76 | 33 | 0.164332 | 0.123430789 | 0.130778943 | -0.040900738 | -0.033552584 | 0.007348154 | 0.645296304 | 0.815538845 | 0.987607159 |
| sp\|P01034\|CYTC_HUMAN | 29 | 40 | 76 | 33 | 0.017167 | 0.030203224 | 0.421950703 | 0.013035913 | 0.404783392 | 0.391747479 | 0.974370367 | 3.12E-07 | 2.56E-08 |
| sp\|P21589\|5NTD_HUMAN | 1 | 40 | 76 | 33 | -0.23397 | -0.060258289 | 0.001660226 | 0.173715419 | 0.235633934 | 0.061918515 | 0.083567385 | 0.043708342 | 0.753274355 |
| sp\|P16109\|LYAM3_HUMAN | 3 | 40 | 76 | 33 | -0.13254 | 0.007615598 | 0.236750321 | 0.14015271 | 0.369287434 | 0.229134724 | 0.044463159 | 1.33E-06 | 0.000885245 |
| sp\|P81605\|DCD_HUMAN | 1 | 40 | 76 | 33 | -0.3149 | -0.205176813 | 0.045615448 | 0.109723568 | 0.360515829 | 0.250792261 | 0.667177367 | 0.053082296 | 0.16063097 |
| sp\|Q96FE7\|P3IP1_HUMAN | 1 | 40 | 76 | 33 | -0.2316 | -0.223841487 | 0.121530272 | 0.007759444 | 0.353131203 | 0.345371759 | 0.995287676 | 0.001782797 | 0.000495074 |
| sp\|P17301\|ITA2_HUMAN | 5 | 40 | 76 | 33 | 0.042222 | 0.005627789 | -0.056083548 | -0.036594206 | -0.098305542 | -0.061711337 | 0.682152165 | 0.15337395 | 0.387005437 |
| sp\|P27487\|DPP4_HUMAN | 1 | 40 | 76 | 33 | 0.180163 | 0.07497484 | -0.148465078 | -0.105188053 | -0.328627971 | -0.223439918 | 0.088026318 | 4.54E-07 | 0.000119937 |
| sp\|Q12884\|SEPR_HUMAN | 11 | 40 | 76 | 33 | 0.141339 | 0.020234233 | -0.102345695 | -0.121105142 | -0.24368507 | -0.122579928 | 0.297594955 | 0.03639829 | 0.335681681 |
| sp\|Q9UHG3\|PCYOX_HUMAN | 1 | 40 | 76 | 33 | 0.102564 | 0.034863907 | 0.067183132 | -0.067699989 | -0.035380765 | 0.032319224 | 0.489598145 | 0.873351368 | 0.866085645 |
| sp\|P22891\|PROZ_HUMAN | 1 | 40 | 76 | 33 | 0.331102 | 0.264003263 | 0.02611638 | -0.067098424 | -0.304985308 | -0.237886884 | 0.554722539 | 0.000403518 | 0.002139633 |
| sp\|P23381\|SYWC_HUMAN | 10 | 40 | 76 | 33 | -0.22304 | 0.085538542 | 0.32735786 | 0.308581832 | 0.550401151 | 0.241819318 | 0.000515219 | 1.89E-07 | 0.014744249 |
| sp\|Q92484\|ASM3A_HUMAN | 3 | 40 | 76 | 33 | 0.127148 | 0.100277248 | 0.055475213 | -0.026870298 | -0.071672333 | -0.044802034 | 0.943764735 | 0.753188324 | 0.868363641 |
| sp\|P24593\|IBP5_HUMAN | 6 | 40 | 76 | 33 | 0.138379 | 0.124468836 | -0.026795904 | -0.013909876 | -0.165174616 | -0.15126474 | 0.936955962 | 0.002565408 | 0.00175853 |
| sp\|P22692\|IBP4_HUMAN | 1 | 40 | 76 | 33 | -0.00633 | 0.0247718 | 0.32296957 | 0.031097955 | 0.329295725 | 0.29819777 | 0.875726307 | 8.53E-05 | 5.87E-05 |
| sp\|P08833\|IBP1_HUMAN | 7 | 40 | 76 | 33 | -0.34916 | -0.264298895 | -0.12441712 | 0.084863499 | 0.224745274 | 0.139881775 | 0.800221624 | 0.343403008 | 0.58866805 |
| sp\|O00462\|MANBA_HUMAN | 5 | 40 | 76 | 33 | 0.114045 | 0.072253824 | 0.017346314 | -0.041791301 | -0.096698811 | -0.05490751 | 0.511291614 | 0.087791271 | 0.36313032 |
| sp\|P49747\|COMP_HUMAN | 1 | 40 | 76 | 33 | 0.281967 | 0.17946943 | -0.001261265 | -0.102497161 | -0.283227856 | -0.180730695 | 0.24655805 | 0.000933086 | 0.023953692 |
| sp\|P05452\|TETN_HUMAN | 34 | 40 | 76 | 33 | 0.262959 | 0.167538308 | -0.200176567 | -0.09542031 | -0.463135186 | -0.367714876 | 0.038308186 | 3.81E-14 | 4.47E-14 |
| sp\|Q9BQ66\|KR412_HUMAN | 2 | 4 | 7 | 3 | -0.03261 | 0.424495419 | -0.303638452 | 0.457103314 | -0.271030556 | -0.72813387 | 0.807766606 | 0.949959205 | 0.645717021 |
| sp\|Q16627\|CCL14_HUMAN | 55 | 40 | 76 | 33 | -0.07359 | 0.042166955 | 0.233119044 | 0.115759137 | 0.306711226 | 0.190952089 | 0.133006623 | 0.000109732 | 0.009179766 |
| sp\|Q8NBP7\|PCSK9_HUMAN | 1 | 40 | 76 | 33 | 0.025581 | 0.071592863 | 0.159083844 | 0.046011622 | 0.133502603 | 0.087490981 | 0.67085432 | 0.102933569 | 0.284839 |
| sp\|P28799\|GRN_HUMAN | 47 | 40 | 76 | 33 | 0.081921 | 0.142670847 | 0.246905849 | 0.060749914 | 0.164984916 | 0.104235002 | 0.420546446 | 0.0141627 | 0.109950933 |
| sp\|P27918\|PROP_HUMAN | 6 | 40 | 76 | 33 | 0.264207 | 0.185625664 | -0.087743494 | -0.078581222 | -0.35195038 | -0.273369158 | 0.127278396 | 5.96E-11 | 6.88E-09 |
| sp\|P60900\|PSA6_HUMAN | 28 | 40 | 76 | 33 | -0.37737 | -0.196698568 | 0.414974314 | 0.180673333 | 0.792346215 | 0.611672882 | 0.242407195 | 7.85E-08 | 2.78E-06 |
| sp\|P01597\|KV139_HUMAN | 8 | 33 | 62 | 27 | -0.08079 | -0.051727863 | 0.146442986 | 0.029060675 | 0.227231525 | 0.198170849 | 0.921879056 | 0.036909361 | 0.041473818 |
| sp\|P01602\|KV105_HUMAN | 9 | 15 | 27 | 13 | 0.233609 | 0.071514271 | 0.541900981 | -0.162095029 | 0.308291681 | 0.47038671 | 0.485313876 | 0.158690982 | 0.00658136 |
| sp\|P02746\|C1QB_HUMAN | 5 | 40 | 76 | 33 | 0.102249 | 0.224281676 | 0.111355924 | 0.12203228 | 0.009106529 | -0.112925752 | 0.119226415 | 0.991678655 | 0.200366623 |
| sp\|Q8IZF2\|AGRF5_HUMAN | 1 | 40 | 76 | 33 | 0.034451 | 0.057947183 | -0.008128922 | 0.02349629 | -0.042579815 | -0.066076104 | 0.860111027 | 0.711224807 | 0.35468399 |
| sp\|P07998\|RNAS1_HUMAN | 7 | 40 | 76 | 33 | -0.21894 | -0.225801972 | 0.33843286 | -0.006866442 | 0.55736839 | 0.564234832 | 0.996532611 | 9.86E-07 | 2.56E-08 |
| sp\|P20061\|TCO1_HUMAN | 3 | 40 | 76 | 33 | 0.081197 | 0.058705437 | 0.080363783 | -0.022491797 | -0.000833451 | 0.021658346 | 0.9426749 | 0.999944036 | 0.953070713 |
| sp\|Q14520\|HABP2_HUMAN | 3 | 40 | 76 | 33 | 0.344825 | 0.373591071 | 0.191400362 | 0.028766305 | -0.153424404 | -0.182190709 | 0.700920469 | 0.001428838 | 1.29E-05 |
| sp\|P03950\|ANGI_HUMAN | 1 | 40 | 76 | 33 | 0.059573 | 0.099438059 | 0.118266448 | 0.039865437 | 0.058693825 | 0.018828389 | 0.590350766 | 0.455620851 | 0.901507353 |
| sp\|A0A0B4J1V0\|HV315_HUMAN | 30 | 18 | 35 | 16 | -0.13195 | -0.114151981 | 0.093513322 | 0.017801955 | 0.225467259 | 0.207665304 | 0.988535998 | 0.276237608 | 0.243801688 |
| sp\|P01780\|HV307_HUMAN | 5 | 40 | 76 | 33 | -0.23515 | -0.102847561 | 0.078555412 | 0.13229961 | 0.313702582 | 0.181402973 | 0.293239759 | 0.010072465 | 0.134190096 |
| sp\|P01782\|HV309_HUMAN | 1 | 11 | 21 | 10 | 0.178613 | 0.022240117 | 0.158157508 | -0.156372619 | -0.020455228 | 0.135917391 | 0.188709486 | 0.978497181 | 0.301588547 |
| sp\|P36980\|FHR2_HUMAN | 6 | 40 | 76 | 33 | 0.045723 | 0.090525465 | 0.098000108 | 0.044802169 | 0.052276812 | 0.007474643 | 0.412932882 | 0.435461773 | 0.978421789 |
| sp\|P33908\|MA1A1_HUMAN | 1 | 40 | 76 | 33 | 0.134441 | 0.146438124 | 0.244298381 | 0.011996936 | 0.109857193 | 0.097860257 | 0.945112731 | 0.042349785 | 0.041106361 |
| sp\|Q8TDQ0\|HAVR2_HUMAN | 7 | 40 | 76 | 33 | -0.44603 | -0.30194773 | 0.353334152 | 0.144078346 | 0.799360228 | 0.655281882 | 0.329170936 | 2.34E-09 | 2.91E-08 |
| sp\|Q9Y6L7\|TLL2_HUMAN | 8 | 40 | 76 | 33 | 0.043821 | 0.071359603 | -0.020750971 | 0.027539055 | -0.06457152 | -0.092110575 | 0.830817895 | 0.496967219 | 0.166978049 |
| sp\|Q9BXR6\|FHR5_HUMAN | 2 | 40 | 76 | 33 | -0.08285 | 0.031326602 | 0.25947352 | 0.114173463 | 0.342320382 | 0.228146918 | 0.146879471 | 1.83E-05 | 0.001615099 |
| sp\|Q9P0K1\|ADA22_HUMAN | 2 | 16 | 27 | 12 | 0.12312 | 0.019448222 | 0.323606755 | -0.103671838 | 0.200486695 | 0.304158533 | 0.733444823 | 0.457168231 | 0.120429534 |
| sp\|Q24JP5\|T132A_HUMAN | 13 | 40 | 76 | 33 | 0.114078 | 0.101882807 | 0.097020514 | -0.012195124 | -0.017057418 | -0.004862293 | 0.931834291 | 0.909122463 | 0.990188276 |
| sp\|Q7Z7M0\|MEGF8_HUMAN | 3 | 40 | 76 | 33 | 0.215984 | 0.205832873 | 0.240832167 | -0.010151449 | 0.024847845 | 0.034999295 | 0.916774828 | 0.69895758 | 0.406960501 |
| sp\|Q12860\|CNTN1_HUMAN | 12 | 40 | 76 | 33 | 0.268409 | 0.116935509 | -0.226334612 | -0.151473153 | -0.494743274 | -0.343270121 | 0.011292911 | 1.71E-12 | 1.67E-08 |
| sp\|Q6WN34\|CRDL2_HUMAN | 46 | 40 | 76 | 33 | -0.00182 | -0.085963626 | -0.02648536 | -0.084142321 | -0.024664054 | 0.059478266 | 0.611464465 | 0.971119128 | 0.80538571 |
| sp\|P00338\|LDHA_HUMAN | 3 | 40 | 76 | 33 | 0.085971 | 0.191778806 | 0.491394197 | 0.105808121 | 0.405423513 | 0.299615392 | 0.362199307 | 7.73E-05 | 0.001171411 |
| sp\|P20062\|TCO2_HUMAN | 5 | 40 | 76 | 33 | 0.097667 | 0.104297099 | 0.132543198 | 0.006630525 | 0.034876625 | 0.028246099 | 0.988102097 | 0.79615121 | 0.826673256 |
| sp\|P07333\|CSF1R_HUMAN | 1 | 40 | 76 | 33 | 0.10416 | 0.115408268 | 0.168399204 | 0.011247855 | 0.06423879 | 0.052990935 | 0.975142591 | 0.569353307 | 0.61379054 |
| sp\|Q96BZ4\|PLD4_HUMAN | 3 | 40 | 76 | 33 | 0.083522 | 0.175415268 | -0.159824835 | 0.091893334 | -0.243346769 | -0.335240103 | 0.317724562 | 0.004904053 | 5.77E-06 |
| sp\|Q13740\|CD166_HUMAN | 20 | 40 | 76 | 33 | 0.146701 | 0.145581555 | 0.209696039 | -0.001119874 | 0.06299461 | 0.064114484 | 0.999485401 | 0.329362558 | 0.23257142 |
| sp\|Q12805\|FBLN3_HUMAN | 5 | 40 | 76 | 33 | 0.394228 | 0.342202985 | 0.568874388 | -0.052025508 | 0.174645895 | 0.226671403 | 0.557793258 | 0.012705551 | 0.000128998 |
| sp\|P51884\|LUM_HUMAN | 30 | 40 | 76 | 33 | 0.299395 | 0.171154296 | 0.174242487 | -0.128240775 | -0.125152584 | 0.003088191 | 0.025485391 | 0.086999934 | 0.998060805 |
| sp\|P07996\|TSP1_HUMAN | 27 | 40 | 76 | 33 | -0.46147 | 0.294608877 | 0.313293822 | 0.756078135 | 0.774763081 | 0.018684945 | 2.92E-06 | 7.45E-05 | 0.992302024 |
| sp\|P01762\|HV311_HUMAN | 19 | 21 | 41 | 19 | -0.20471 | -0.065753011 | 0.304878582 | 0.138958508 | 0.509590101 | 0.370631593 | 0.63302784 | 0.01564055 | 0.053910154 |
| sp\|P08519\|APOA_HUMAN | 4 | 40 | 76 | 33 | 0.252977 | 0.27721209 | 0.207015594 | 0.024234698 | -0.045961798 | -0.070196496 | 0.933527301 | 0.843226305 | 0.603910562 |
| sp\|P02787\|TRFE_HUMAN | 25 | 40 | 76 | 33 | -0.22504 | -0.54259436 | -0.786448147 | -0.317553246 | -0.561407032 | -0.243853787 | 0.167454761 | 0.02291042 | 0.393156849 |
| sp\|Q15113\|PCOC1_HUMAN | 10 | 40 | 76 | 33 | 0.156016 | 0.15873871 | 0.113395942 | 0.002722258 | -0.042620511 | -0.045342769 | 0.997405444 | 0.645750333 | 0.533344161 |
| sp\|P05362\|ICAM1_HUMAN | 1451 | 40 | 76 | 33 | -0.14869 | -0.04022144 | 0.464941221 | 0.108470285 | 0.613633346 | 0.50516306 | 0.176402795 | 1.40E-13 | 3.06E-12 |
| sp\|Q9H1U4\|MEGF9_HUMAN | 75 | 40 | 76 | 33 | 0.115668 | 0.16397621 | -0.000309358 | 0.048308503 | -0.115977066 | -0.164285568 | 0.358437539 | 0.019083888 | 6.91E-05 |
| sp\|P14543\|NID1_HUMAN | 13 | 40 | 76 | 33 | -0.08648 | 0.070955607 | 0.37537689 | 0.157438219 | 0.461859503 | 0.304421284 | 0.018499778 | 1.31E-09 | 5.37E-06 |
| sp\|Q9UBX1\|CATF_HUMAN | 152 | 40 | 76 | 33 | 0.095495 | 0.104295631 | 0.061377427 | 0.008800198 | -0.034118005 | -0.042918204 | 0.987505523 | 0.877892346 | 0.769618644 |

| | | | | | | | | | | | | | |
|---|---|---|---|---|---|---|---|---|---|---|---|---|---|
| sp\|P24387\|CRHBP_HUMAN | 29 | 40 | 76 | 33 | 0.058584 | 0.045870225 | 0.180745087 | -0.012713623 | 0.122161239 | 0.134874862 | 0.941443412 | 0.024695644 | 0.003580685 |
| sp\|O43278\|SPIT1_HUMAN | 12 | 40 | 76 | 33 | -0.10861 | -0.040181795 | 0.428777395 | 0.068429615 | 0.537388805 | 0.468959191 | 0.552807327 | 8.08E-10 | 1.41E-09 |
| sp\|P07093\|GDN_HUMAN | 2 | 40 | 76 | 33 | -0.39523 | -0.093272093 | 0.106135517 | 0.301961559 | 0.501369169 | 0.19940761 | 0.00774394 | 0.000136801 | 0.146983056 |
| sp\|Q9NY15\|STAB1_HUMAN | 35 | 40 | 76 | 33 | 0.241415 | 0.213948281 | 0.1580146 | -0.027466267 | -0.083399947 | -0.05593368 | 0.579960688 | 0.034537353 | 0.141639431 |
| sp\|Q14118\|DAG1_HUMAN | 29 | 40 | 76 | 33 | 0.052365 | 0.016311737 | 0.367380646 | -0.036052869 | 0.315016039 | 0.351068909 | 0.822055341 | 8.10E-05 | 6.39E-07 |
| sp\|Q9UBQ7\|GRHPR_HUMAN | 11 | 40 | 76 | 33 | -0.15371 | -0.099147765 | 0.180291889 | 0.054566785 | 0.334006439 | 0.279439654 | 0.824888114 | 0.008663327 | 0.014257821 |
| sp\|Q6UXB8\|PI16_HUMAN | 621 | 40 | 76 | 33 | 0.854951 | 0.773050443 | 0.493662266 | -0.081900532 | -0.36128871 | -0.279388178 | 0.570420983 | 0.000858229 | 0.004268681 |
| sp\|Q9UNN8\|EPCR_HUMAN | 340 | 40 | 76 | 33 | 0.107174 | 0.08388722 | 0.079238061 | -0.023287237 | -0.027936396 | -0.004649159 | 0.869506404 | 0.870359119 | 0.995111653 |
| sp\|P20933\|ASPG_HUMAN | 133 | 40 | 76 | 33 | 0.053938 | -0.027349533 | 0.0266917 | -0.081287418 | -0.027246186 | 0.054041233 | 0.348720015 | 0.920800931 | 0.662589563 |
| sp\|Q96CX2\|KCD12_HUMAN | 97 | 40 | 76 | 33 | 0.092534 | 0.014977797 | 0.027606074 | -0.077555824 | -0.064927547 | 0.012628277 | 0.14138827 | 0.384443201 | 0.954598041 |
| sp\|P12259\|FA5_HUMAN | 126 | 40 | 76 | 33 | 0.135399 | 0.200156794 | 0.25871773 | 0.064758095 | 0.123319031 | 0.058560935 | 0.18184469 | 0.015614506 | 0.292300457 |
| sp\|P25789\|PSA4_HUMAN | 140 | 40 | 76 | 33 | -0.35443 | -0.162137042 | 0.343397042 | 0.192288765 | 0.697822849 | 0.505534084 | 0.149765672 | 2.43E-07 | 2.51E-05 |
| sp\|P13727\|PRG2_HUMAN | 47 | 40 | 76 | 33 | 0.056493 | 0.015470379 | 0.119503124 | -0.041022863 | 0.063009881 | 0.104032744 | 0.804074712 | 0.701561991 | 0.295738334 |
| sp\|Q76LX8\|ATS13_HUMAN | 699 | 40 | 76 | 33 | 0.201634 | 0.204882409 | -0.115553754 | 0.003248829 | -0.317187334 | -0.320436163 | 0.996812752 | 1.90E-08 | 2.17E-10 |
| sp\|Q9Y5Y7\|LYVE1_HUMAN | 263 | 40 | 76 | 33 | 0.051896 | 0.104096326 | 0.142162671 | 0.052200632 | 0.090266978 | 0.038066345 | 0.710087573 | 0.494770633 | 0.852007419 |
| sp\|P10586\|PTPRF_HUMAN | 538 | 40 | 76 | 33 | 0.060004 | 0.131567008 | 0.263114763 | 0.071562788 | 0.203110544 | 0.131547755 | 0.257680711 | 0.000812075 | 0.01986941 |
| sp\|P14151\|LYAM1_HUMAN | 6 | 40 | 76 | 33 | 0.054749 | 0.164041472 | -0.206571303 | 0.10929236 | -0.261320416 | -0.370612775 | 0.120399002 | 0.000374831 | 1.01E-08 |
| sp\|Q02985\|FHR3_HUMAN | 44 | 40 | 76 | 33 | -0.01389 | -0.07095263 | -0.055590104 | -0.057065936 | -0.04170341 | 0.015362526 | 0.346654362 | 0.674923209 | 0.934143609 |
| sp\|P30043\|BLVRB_HUMAN | 32 | 40 | 76 | 33 | -0.36905 | -0.084450829 | 0.167175277 | 0.284596505 | 0.53622261 | 0.251626106 | 0.013913481 | 4.90E-05 | 0.051160093 |
| sp\|P23468\|PTPRD_HUMAN | 8 | 40 | 76 | 33 | 0.100514 | 0.091979236 | -0.000133862 | -0.008534674 | -0.100647772 | -0.092113098 | 0.959993357 | 0.023078616 | 0.018182086 |
| sp\|P98160\|PGBM_HUMAN | 6 | 40 | 76 | 33 | 0.388557 | 0.339729505 | 0.379861454 | -0.048827714 | -0.008695765 | 0.040131949 | 0.376389245 | 0.978629845 | 0.55879314 |
| sp\|P55268\|LAMB2_HUMAN | 335 | 40 | 76 | 33 | 0.121033 | 0.110980082 | 0.222593517 | -0.010052444 | 0.101560991 | 0.111613436 | 0.954117222 | 0.040732692 | 0.007915108 |
| sp\|P68871\|HBB_HUMAN | 149 | 40 | 76 | 33 | -1.49187 | -1.130994485 | -0.067617905 | 0.36087498 | 1.42425156 | 1.06337658 | 0.267043287 | 2.98E-06 | 9.08E-05 |
| sp\|Q9UHD8\|SEPT9_HUMAN | 453 | 4 | 7 | 3 | 0.245954 | 0.078410929 | -0.09573579 | -0.167543178 | -0.341689897 | -0.174146719 | 0.515087637 | 0.185933983 | 0.551440516 |
| sp\|P09493\|TPM1_HUMAN | 264 | 40 | 76 | 33 | 0.046251 | -0.318040714 | 0.031803931 | -0.364291873 | -0.014447229 | 0.349844645 | 0.022940224 | 0.995748836 | 0.046041229 |
| sp\|Q6E0U4\|DMKN_HUMAN | 207 | 40 | 76 | 33 | -0.2014 | -0.329819302 | -0.012565616 | -0.128422205 | 0.18883148 | 0.317253686 | 0.203158977 | 0.094539585 | 0.000330609 |
| sp\|P08670\|VIME_HUMAN | 273 | 40 | 76 | 33 | -1.00632 | -0.799528945 | -0.061507213 | 0.206794096 | 0.944815828 | 0.738021732 | 0.202748064 | 3.36E-09 | 1.60E-07 |
| sp\|P00915\|CAH1_HUMAN | 913 | 40 | 76 | 33 | -0.25181 | 0.072081735 | 0.351532431 | 0.32388705 | 0.603337745 | 0.279450695 | 0.008394358 | 2.01E-05 | 0.041795473 |
| sp\|P27169\|PON1_HUMAN | 15 | 40 | 76 | 33 | 0.40943 | 0.213537105 | 0.058614977 | -0.195893176 | -0.350815304 | -0.154922128 | 0.009087949 | 5.01E-05 | 0.071674696 |
| sp\|P68366\|TBA4A_HUMAN | 41 | 40 | 76 | 33 | -0.32623 | -0.067447938 | 0.048296921 | 0.258781264 | 0.374526123 | 0.115744859 | 0.063548424 | 0.019533334 | 0.60919296 |
| sp\|Q07960\|RHG01_HUMAN | 3 | 40 | 76 | 33 | -0.11493 | -0.046918472 | 0.143329737 | 0.068009613 | 0.258257823 | 0.19024821 | 0.666381884 | 0.020339564 | 0.065746932 |
| sp\|Q86UX2\|ITIH5_HUMAN | 2 | 40 | 76 | 33 | 0.003459 | -0.07717917 | -0.03079829 | -0.080637921 | -0.034257041 | 0.04638088 | 0.077394316 | 0.721673527 | 0.468907669 |
| sp\|Q8NHM4\|TRY6_HUMAN | 3 | 40 | 76 | 33 | 0.120517 | -0.005474735 | -0.128784244 | -0.125991548 | -0.249301057 | -0.123309509 | 0.0004668 | 6.91E-09 | 0.001472358 |
| sp\|O75636\|FCN3_HUMAN | 435 | 40 | 76 | 33 | 0.067236 | 0.193359997 | 0.031358961 | 0.126124308 | -0.035876729 | -0.162001037 | 0.10424066 | 0.878888259 | 0.039069151 |
| sp\|P07602\|SAP_HUMAN | 2 | 40 | 76 | 33 | -0.21912 | -0.116828449 | 0.41299935 | 0.102293754 | 0.632121553 | 0.529827799 | 0.345121921 | 1.03E-10 | 8.31E-10 |
| sp\|P14314\|GLU2B_HUMAN | 27 | 40 | 76 | 33 | -0.1373 | -0.13471165 | 0.313552341 | 0.002586103 | 0.450850094 | 0.448263991 | 0.998854056 | 2.08E-09 | 2.87E-11 |
| sp\|P22105\|TENX_HUMAN | 9 | 40 | 76 | 33 | 0.362 | 0.289604448 | 0.051228037 | -0.07239508 | -0.310771491 | -0.238376411 | 0.1885986 | 1.23E-08 | 7.38E-07 |
| sp\|P37840\|SYUA_HUMAN | 15 | 40 | 76 | 33 | -0.07674 | -0.19611775 | 0.203360604 | -0.119382182 | 0.280096172 | 0.399478354 | 0.385016716 | 0.029305124 | 0.000167878 |
| sp\|Q86UX7\|URP2_HUMAN | 2 | 40 | 76 | 33 | 0.035739 | -0.165911529 | -0.015045061 | -0.201650346 | -0.050783878 | 0.150866468 | 0.38486826 | 0.958684137 | 0.623960547 |
| sp\|P63098\|CANB1_HUMAN | 3 | 36 | 70 | 30 | 0.027884 | 0.14056918 | 0.440877701 | 0.112685129 | 0.41299365 | 0.300308521 | 0.382422795 | 0.000266906 | 0.003254415 |
| sp\|Q14766\|LTBP1_HUMAN | 1 | 40 | 76 | 33 | -0.32866 | -0.001228572 | 0.238439923 | 0.327435745 | 0.56710424 | 0.239668494 | 0.001257862 | 2.17E-06 | 0.038800996 |
| sp\|P49908\|SEPP1_HUMAN | 2 | 40 | 76 | 33 | 0.234509 | 0.211184037 | 0.002596856 | -0.023325353 | -0.231912534 | -0.208587181 | 0.82113298 | 6.21E-06 | 4.52E-06 |
| sp\|P20742\|PZP_HUMAN | 5 | 40 | 76 | 33 | -0.04039 | 0.148065212 | -0.175624183 | 0.18845853 | -0.135230865 | -0.323689395 | 0.559874709 | 0.813107424 | 0.225819133 |
| sp\|Q9NPY3\|C1QR1_HUMAN | 3 | 40 | 76 | 33 | 0.067231 | 3.95E-05 | 0.17631598 | -0.067191735 | 0.10908472 | 0.176276455 | 0.615539165 | 0.415282043 | 0.057238114 |
| sp\|P12955\|PEPD_HUMAN | 1 | 40 | 76 | 33 | 0.289395 | 0.221733471 | 0.062976468 | -0.067661746 | -0.226418749 | -0.158757003 | 0.090455598 | 8.15E-08 | 2.20E-05 |
| sp\|Q02487\|DSC2_HUMAN | 24 | 40 | 76 | 33 | 0.048635 | 0.024423056 | 0.390832463 | -0.024212095 | 0.342197312 | 0.366409407 | 0.929205475 | 9.71E-05 | 2.19E-06 |
| sp\|Q07075\|AMPE_HUMAN | 3 | 40 | 76 | 33 | 0.175578 | 0.26325343 | -0.014735199 | 0.087675652 | -0.190312977 | -0.277988629 | 0.037154144 | 4.38E-05 | 3.21E-11 |
| sp\|P05556\|ITB1_HUMAN | 6 | 40 | 76 | 33 | 0.230416 | 0.199440531 | 0.102158433 | -0.030975875 | -0.128257973 | -0.097282098 | 0.558657406 | 0.001521189 | 0.008009051 |
| sp\|P08709\|FA7_HUMAN | 3 | 40 | 76 | 33 | 0.162604 | 0.182994487 | 0.063210834 | 0.020390523 | -0.099393129 | -0.119783652 | 0.745388317 | 0.00997029 | 0.00026773 |
| sp\|P12111\|CO6A3_HUMAN | 2 | 40 | 76 | 33 | 0.259411 | 0.248207361 | 0.436117517 | -0.011203398 | 0.176706758 | 0.187910157 | 0.966518255 | 0.003982036 | 0.000417269 |
| sp\|P33241\|LSP1_HUMAN | 1 | 7 | 14 | 6 | 0.160112 | 0.089628174 | 0.148445503 | -0.070484254 | -0.011666925 | 0.058817329 | 0.836596497 | 0.99659437 | 0.893906491 |
| sp\|Q13683\|ITA7_HUMAN | 2 | 40 | 76 | 33 | 0.175567 | 0.116525728 | -0.141681897 | -0.059040987 | -0.317248612 | -0.258207625 | 0.454923808 | 9.62E-07 | 6.91E-06 |
| sp\|P22352\|GPX3_HUMAN | 3 | 40 | 76 | 33 | 0.186247 | 0.076020428 | -0.000133185 | -0.110226496 | -0.186380109 | -0.076153613 | 0.036800153 | 0.001794656 | 0.243861448 |
| sp\|Q6UWP8\|SBSN_HUMAN | 3 | 40 | 76 | 33 | 0.128392 | 0.032471539 | 0.192044829 | -0.095920271 | 0.06365302 | 0.15957329 | 0.184206814 | 0.594283244 | 0.018057812 |
| sp\|Q15942\|ZYX_HUMAN | 1 | 40 | 76 | 33 | 0.061643 | -0.061582917 | 0.184657447 | -0.123226381 | 0.123013982 | 0.246240364 | 0.472700885 | 0.596610487 | 0.075980886 |
| sp\|P16112\|PGCA_HUMAN | 2 | 40 | 76 | 33 | 0.120408 | 0.047411085 | -0.195512166 | -0.072996963 | -0.315920214 | -0.242923251 | 0.305892765 | 1.24E-06 | 2.70E-05 |
| sp\|P34096\|RNAS4_HUMAN | 1 | 40 | 76 | 33 | 0.031062 | 0.107663423 | 0.310186705 | 0.076601313 | 0.279124595 | 0.202523282 | 0.126452331 | 6.40E-08 | 9.30E-06 |
| sp\|Q13822\|ENPP2_HUMAN | 6 | 40 | 76 | 33 | 0.295282 | 0.286834707 | 0.146039511 | -0.008447639 | -0.149242836 | -0.140795195 | 0.987241725 | 0.067973209 | 0.048115253 |
| sp\|P09172\|DOPO_HUMAN | 1 | 40 | 76 | 33 | 0.290453 | 0.200814238 | -0.105175728 | -0.089639231 | -0.395629197 | -0.305989966 | 0.563874607 | 0.00074341 | 0.00380853 |
| sp\|Q6GTS8\|P20D1_HUMAN | 25 | 40 | 76 | 33 | -0.09134 | -0.258556455 | -0.090769695 | -0.167213667 | 0.000573094 | 0.167786761 | 0.568455506 | 0.999995394 | 0.60673456 |
| sp\|O00299\|CLIC1_HUMAN | 12 | 40 | 76 | 33 | -0.0897 | -0.03383157 | 0.19268642 | 0.055869176 | 0.282387165 | 0.22651799 | 0.905713895 | 0.179775514 | 0.244248481 |
| sp\|Q13093\|PAFA_HUMAN | 8 | 40 | 76 | 33 | 0.015804 | 0.173443181 | 0.327285946 | 0.157639409 | 0.311482174 | 0.153842765 | 0.141615454 | 0.006091946 | 0.194011659 |
| sp\|Q6MZM0\|HPHL1_HUMAN | 18 | 40 | 76 | 33 | 0.195469 | 0.136686713 | 0.009204251 | -0.058782485 | -0.186264948 | -0.127482463 | 0.592876829 | 0.029863111 | 0.119699429 |
| sp\|P62937\|PPIA_HUMAN | 14 | 40 | 76 | 33 | 0.047132 | -0.01032778 | 0.296061682 | -0.057459705 | 0.248929756 | 0.306389462 | 0.786867244 | 0.049030685 | 0.003477209 |
| sp\|P19961\|AMY2B_HUMAN | 9 | 40 | 76 | 33 | 0.112982 | 0.052298145 | -0.013164419 | -0.06068389 | -0.126146454 | -0.065462564 | 0.650019805 | 0.280017892 | 0.643999468 |
| sp\|P55056\|APOC4_HUMAN | 23 | 40 | 76 | 33 | 0.151037 | 0.163588342 | 0.321396634 | 0.012551147 | 0.170359439 | 0.157808292 | 0.991763055 | 0.353488527 | 0.321638583 |
| sp\|O95445\|APOM_HUMAN | 14 | 40 | 76 | 33 | 6.60E-05 | 0.157382891 | 0.269164552 | 0.157316881 | 0.269098542 | 0.111781661 | 0.095288475 | 0.009680759 | 0.347758579 |
| sp\|P04040\|CATA_HUMAN | 3 | 40 | 76 | 33 | -0.3043 | -0.143744511 | 0.415261567 | 0.160558665 | 0.719564743 | 0.559006078 | 0.286100499 | 2.48E-07 | 6.08E-06 |
| sp\|Q9UBP4\|DKK3_HUMAN | 9 | 40 | 76 | 33 | 0.161028 | 0.069602965 | 0.255097449 | -0.091425042 | 0.094069443 | 0.185494485 | 0.097747448 | 0.18095635 | 0.000351814 |
| sp\|P32119\|PRDX2_HUMAN | 8 | 40 | 76 | 33 | -0.58948 | -0.128517254 | 0.159956042 | 0.460961013 | 0.749434309 | 0.288473296 | 0.001117724 | 7.25E-06 | 0.087155125 |
| sp\|P08195\|4F2_HUMAN | 4 | 40 | 76 | 33 | 0.183861 | 0.170959865 | -0.123327291 | -0.012901402 | -0.307188558 | -0.294287155 | 0.942769619 | 4.20E-09 | 2.73E-10 |
| sp\|O95479\|G6PE_HUMAN | 6 | 40 | 76 | 33 | 0.06481 | 0.115900505 | 0.093070966 | 0.051090099 | 0.02826056 | -0.022829539 | 0.500465961 | 0.863302075 | 0.885066694 |
| sp\|Q9NQ79\|CRAC1_HUMAN | 1 | 40 | 76 | 33 | 0.336198 | 0.09688913 | -0.232195412 | -0.239308813 | -0.568393355 | -0.329084542 | 0.009921138 | 9.55E-08 | 0.000580454 |
| sp\|P00325\|ADH1B_HUMAN | 6 | 40 | 76 | 33 | -0.64119 | -0.185372139 | 0.108082933 | 0.455819687 | 0.749274759 | 0.293455072 | 0.005056872 | 7.68E-05 | 0.13683369 |
| sp\|P17927\|CR1_HUMAN | 3 | 40 | 76 | 33 | 0.0459 | 0.023309832 | 0.131353698 | -0.022589822 | 0.085454044 | 0.108043866 | 0.896896565 | 0.345231254 | 0.117745772 |
| sp\|Q05682\|CALD1_HUMAN | 35 | 40 | 76 | 33 | -0.00316 | -0.204837093 | 0.091389226 | -0.20167694 | 0.094549379 | 0.296226319 | 0.25311529 | 0.809825982 | 0.076512644 |
| sp\|P35527\|K1C9_HUMAN | 17 | 40 | 76 | 33 | 0.176197 | -0.23809524 | -0.096666351 | -0.41429183 | -0.272862941 | 0.141428889 | 0.044877985 | 0.386752946 | 0.72115239 |
| sp\|O75083\|WDR1_HUMAN | 71 | 40 | 76 | 33 | 0.060336 | -0.096188077 | 0.160208288 | -0.156524261 | 0.099872104 | 0.256396365 | 0.456555359 | 0.801263201 | 0.160901771 |
| sp\|Q15848\|ADIPO_HUMAN | 5 | 40 | 76 | 33 | 0.300206 | 0.110273796 | -0.12455343 | -0.189931849 | -0.424759075 | -0.234827226 | 0.136656748 | 0.001412907 | 0.070441972 |
| sp\|Q9P232\|CNTN3_HUMAN | 29 | 40 | 76 | 33 | 0.201044 | 0.144140838 | 0.050048263 | -0.056903518 | -0.150996093 | -0.094092575 | 0.358341803 | 0.008249638 | 0.088221724 |
| sp\|Q86SQ4\|AGRG6_HUMAN | 5 | 40 | 76 | 33 | 0.148435 | 0.142274496 | 0.230045076 | -0.006160929 | 0.081609651 | 0.08777058 | 0.985514228 | 0.176450578 | 0.079479931 |
| sp\|P33151\|CADH5_HUMAN | 2 | 40 | 76 | 33 | 0.090081 | 0.06494014 | 0.025264428 | -0.025140511 | -0.064816223 | -0.039675712 | 0.834302099 | 0.438164688 | 0.673522042 |
| sp\|Q93070\|NAR4_HUMAN | 11 | 40 | 76 | 33 | 0.032825 | 0.05717212 | 0.176530064 | 0.02434696 | 0.143704903 | 0.119357944 | 0.822990708 | 0.011330683 | 0.019185535 |
| sp\|P35442\|TSP2_HUMAN | 2 | 40 | 76 | 33 | -0.233 | -0.011957165 | 0.332073861 | 0.22104313 | 0.565074156 | 0.344031026 | 0.010074737 | 1.10E-08 | 8.73E-05 |
| sp\|Q8N0V5\|GNT2A_HUMAN | 15 | 40 | 76 | 33 | 0.162236 | 0.153063753 | 0.135955021 | -0.009172628 | -0.026281361 | -0.017108732 | 0.984616367 | 0.915993373 | 0.953765358 |
| sp\|Q4LDE5\|SVEP1_HUMAN | 10 | 40 | 76 | 33 | 0.137834 | 0.151828983 | 0.52186932 | 0.013995076 | 0.384035412 | 0.370040336 | 0.976513767 | 1.54E-05 | 2.54E-06 |
| sp\|P01833\|PIGR_HUMAN | 22 | 40 | 76 | 33 | 0.017303 | 0.022464695 | 0.246101514 | 0.005161803 | 0.228798622 | 0.223636819 | 0.996489646 | 0.01058296 | 0.004172384 |
| sp\|Q9H8L6\|MMRN2_HUMAN | 112 | 40 | 76 | 33 | 0.203427 | 0.187945729 | 0.09173017 | -0.015481737 | -0.111697296 | -0.096215559 | 0.902014162 | 0.028087017 | 0.034060433 |
| sp\|Q92859\|NEO1_HUMAN | 60 | 40 | 76 | 33 | 0.111223 | 0.061831967 | 0.132792414 | -0.049390585 | 0.021569862 | 0.070960447 | 0.508695023 | 0.914384022 | 0.295836777 |
| sp\|Q92820\|GGH_HUMAN | 20 | 40 | 76 | 33 | 0.085644 | 0.153018049 | 0.156866068 | 0.067374406 | 0.071222425 | 0.003848019 | 0.456316762 | 0.545483101 | 0.99773675 |
| sp\|Q16706\|MA2A1_HUMAN | 45 | 40 | 76 | 33 | 0.081935 | 0.080933999 | 0.146692621 | -0.001000737 | 0.064757885 | 0.065758622 | 0.999490774 | 0.234758156 | 0.150660634 |
| sp\|Q0VAF6\|SYCN_HUMAN | 76 | 40 | 76 | 33 | -0.0353 | 0.170438759 | 0.087870676 | 0.205741467 | 0.123173384 | -0.082568083 | 0.025523983 | 0.393328544 | 0.585200264 |
| sp\|P35555\|FBN1_HUMAN | 318 | 40 | 76 | 33 | 0.207734 | 0.179025223 | 0.213203324 | -0.02870883 | 0.005469271 | 0.034178102 | 0.686911793 | 0.990603124 | 0.626962558 |
| sp\|Q16651\|PRSS8_HUMAN | 340 | 36 | 69 | 30 | -0.04841 | -0.014901028 | 0.323886007 | 0.03350902 | 0.372296055 | 0.338787035 | 0.866210511 | 1.76E-05 | 9.94E-06 |
| sp\|P80370\|DLK1_HUMAN | 306 | 40 | 76 | 33 | -0.02948 | -0.171330001 | 0.156193703 | -0.141849109 | 0.185674595 | 0.327523704 | 0.209764962 | 0.158668785 | 0.000978893 |
| sp\|P0DJI9\|SAA2_HUMAN | 3 | 40 | 76 | 33 | -0.38527 | -0.243882966 | 0.691680725 | 0.141388073 | 1.076951763 | 0.93556369 | 0.671016748 | 8.13E-07 | 1.34E-06 |
| sp\|O60613\|SEP15_HUMAN | 25 | 36 | 69 | 31 | -0.07045 | 0.048812934 | 0.262461868 | 0.119266667 | 0.332915601 | 0.213648935 | 0.268196594 | 0.001104419 | 0.024253923 |
| sp\|Q9Y6N6\|LAMC3_HUMAN | 43 | 32 | 62 | 27 | -0.08799 | -0.038351018 | 0.357720183 | 0.049634525 | 0.445705726 | 0.396071201 | 0.886018189 | 0.001845245 | 0.001696888 |
| sp\|Q9HDC9\|APMAP_HUMAN | 64 | 40 | 76 | 33 | 0.059906 | -0.028625964 | 0.255373682 | -0.088531641 | 0.195468005 | 0.283999646 | 0.544367636 | 0.133263285 | 0.005297249 |
| sp\|Q07092\|COGA1_HUMAN | 62 | 25 | 48 | 21 | -0.04618 | -0.073542672 | 0.391115453 | -0.027357838 | 0.437300287 | 0.464658125 | 0.94213055 | 9.28E-05 | 2.74E-06 |
| sp\|Q6UWV6\|ENPP7_HUMAN | 107 | 40 | 76 | 33 | -0.25785 | -0.238858132 | -0.043384553 | 0.018992429 | 0.214466009 | 0.19547358 | 0.981933165 | 0.206429795 | 0.188941339 |
| sp\|Q8TER0\|SNED1_HUMAN | 51 | 40 | 76 | 33 | 0.091365 | 0.091052041 | 0.073581941 | -0.000312854 | -0.017782954 | -0.0174701 | 0.999957058 | 0.908797107 | 0.889211855 |
| sp\|P08174\|DAF_HUMAN | 256 | 40 | 76 | 33 | 0.172999 | 0.116370625 | 0.331494508 | -0.0566288 | 0.158495082 | 0.215123883 | 0.540714877 | 0.039396504 | 0.000684937 |
| sp\|Q8TCZ2\|C99L2_HUMAN | 323 | 40 | 76 | 33 | -0.24977 | -0.26842974 | 0.313390369 | -0.018657663 | 0.563162446 | 0.581820109 | 0.981698347 | 2.82E-05 | 1.00E-06 |
| sp\|P52566\|GDIR2_HUMAN | 51 | 40 | 76 | 33 | -0.24108 | -0.240339212 | 0.251484798 | 0.000736473 | 0.492560483 | 0.49182401 | 0.999986123 | 0.016499966 | 0.005780129 |

| | | | | | | | | | | | | | |
|---|---|---|---|---|---|---|---|---|---|---|---|---|---|
| sp\|P49257\|LMAN1_HUMAN | 1677 | 40 | 76 | 33 | -0.10139 | 0.013108577 | -0.010789061 | 0.114500545 | 0.090602907 | -0.023897638 | 0.291653348 | 0.584928698 | 0.953398409 |
| sp\|P06312\|KV401_HUMAN | 190 | 40 | 76 | 33 | -0.22804 | -0.037852202 | 0.227069582 | 0.190187765 | 0.455109549 | 0.264921784 | 0.107231539 | 0.000245748 | 0.023625917 |
| sp\|P01615\|KVD28_HUMAN | 292 | 10 | 21 | 9 | -0.07677 | 0.185804633 | 0.065648376 | 0.262577648 | 0.142421391 | -0.120156257 | 0.167997136 | 0.68166969 | 0.695588426 |
| sp\|P01860\|IGHG3_HUMAN | 7 | 40 | 76 | 33 | -0.40038 | -0.335915716 | -0.060101479 | 0.06446906 | 0.340283296 | 0.275814237 | 0.864286945 | 0.06529972 | 0.10111371 |
| sp\|P06310\|KV230_HUMAN | 252 | 32 | 62 | 27 | -0.29067 | -0.202347706 | -0.304109303 | 0.088326579 | -0.013435018 | -0.101761598 | 0.537887714 | 0.990020831 | 0.480708491 |
| sp\|O14791\|APOL1_HUMAN | 28 | 40 | 76 | 33 | -0.2087 | 0.037393493 | 0.143414573 | 0.246090777 | 0.352111857 | 0.10602108 | 0.008710675 | 0.001386032 | 0.447198991 |
| sp\|O00241\|SIRB1_HUMAN | 123 | 36 | 69 | 30 | -0.12119 | -0.143336145 | 0.250726036 | -0.022144709 | 0.371917472 | 0.394062181 | 0.962308553 | 0.000954283 | 6.10E-05 |
| sp\|P78324\|SHPS1_HUMAN | 1 | 40 | 76 | 33 | -0.07951 | 0.016233511 | 0.276750168 | 0.095746535 | 0.356263193 | 0.260516658 | 0.37506575 | 0.000169713 | 0.002344793 |
| sp\|Q9H4B7\|TBB1_HUMAN | 631 | 40 | 76 | 33 | -0.28316 | -0.09068047 | 0.03890709 | 0.192475176 | 0.322062737 | 0.129587561 | 0.178090913 | 0.037613461 | 0.499477496 |
| sp\|P04433\|KV311_HUMAN | 21 | 32 | 62 | 27 | -0.08358 | 0.01381066 | 0.042045839 | 0.0973916 | 0.125626779 | 0.028235179 | 0.358980834 | 0.307137719 | 0.925304518 |
| sp\|P01619\|KV320_HUMAN | 7 | 40 | 76 | 33 | 0.005591 | 0.038841551 | 0.357811861 | 0.033250529 | 0.352220839 | 0.31897031 | 0.920426958 | 0.002377726 | 0.001855212 |
| sp\|Q86YD3\|TMM25_HUMAN | 39 | 40 | 76 | 33 | 0.157941 | 0.065713533 | -0.150530981 | -0.092226986 | -0.3084715 | -0.216244513 | 0.101269879 | 1.65E-07 | 3.58E-05 |
| sp\|Q02325\|PLGB_HUMAN | 77 | 40 | 76 | 33 | 0.183319 | 0.217555616 | 0.045242553 | 0.034236655 | -0.138076408 | -0.172313063 | 0.56783938 | 0.002454732 | 1.19E-05 |
| sp\|Q15195\|PLGA_HUMAN | 62 | 29 | 56 | 24 | -2.97275 | -3.094563362 | -3.070406165 | -0.121816065 | -0.097658867 | 0.024157198 | 0.958336961 | 0.981370551 | 0.998528692 |
| sp\|O14594\|NCAN_HUMAN | 729 | 40 | 76 | 33 | 0.089593 | 0.02599 | -0.162109576 | -0.063602755 | -0.251702331 | -0.188099576 | 0.425036411 | 0.000190564 | 0.001982686 |
| sp\|Q14767\|LTBP2_HUMAN | 6 | 40 | 76 | 33 | -0.31773 | -0.405660703 | 0.388531702 | -0.087929074 | 0.706263332 | 0.794192406 | 0.60192488 | 5.40E-09 | 5.10E-13 |
| sp\|P16070\|CD44_HUMAN | 7 | 40 | 76 | 33 | 0.10818 | 0.094760603 | 0.206753098 | -0.013419067 | 0.098573429 | 0.111992495 | 0.938467685 | 0.099338186 | 0.023799429 |
| sp\|Q13103\|SPP24_HUMAN | 1 | 40 | 76 | 33 | 0.065319 | 0.165311551 | -0.032769947 | 0.0999921 | -0.098089398 | -0.198081498 | 0.687667519 | 0.779613652 | 0.278635192 |
| sp\|O95980\|RECK_HUMAN | 25 | 40 | 76 | 33 | 0.177645 | 0.148928799 | -0.080807647 | -0.028716078 | -0.258452524 | -0.229736446 | 0.703091814 | 4.74E-08 | 4.38E-08 |
| sp\|Q58FF3\|ENPLL_HUMAN | 140 | 7 | 14 | 6 | 0.313298 | 0.171936538 | 0.340892539 | -0.141361138 | 0.027594864 | 0.168956001 | 0.421674854 | 0.976602852 | 0.333437195 |
| sp\|Q8N3T6\|T132C_HUMAN | 326 | 40 | 76 | 33 | 0.181032 | 0.135252286 | 0.086119931 | -0.045779954 | -0.094912308 | -0.049132354 | 0.455170363 | 0.100492672 | 0.45118186 |
| sp\|P06702\|S10A9_HUMAN | 3 | 40 | 76 | 33 | -0.71468 | -0.409084213 | 0.370249465 | 0.305592403 | 1.084926081 | 0.779333678 | 0.165734223 | 8.84E-07 | 7.34E-05 |
| sp\|O43895\|XPP2_HUMAN | 8 | 40 | 76 | 33 | 0.012251 | -0.11402998 | -0.198241656 | -0.126281266 | -0.210492942 | -0.084211676 | 0.583613058 | 0.358068355 | 0.809779194 |
| sp\|P20023\|CR2_HUMAN | 372 | 40 | 76 | 33 | 0.017651 | 0.14266669 | -0.224836922 | 0.12501561 | -0.242488003 | -0.367503612 | 0.217497384 | 0.020935203 | 2.49E-05 |
| sp\|P48551\|INAR2_HUMAN | 379 | 37 | 69 | 29 | -0.21305 | -0.15747271 | 0.353901664 | 0.055579051 | 0.566953425 | 0.511374374 | 0.809918608 | 2.32E-06 | 1.81E-06 |
| sp\|Q86YW5\|TRML1_HUMAN | 55 | 40 | 76 | 33 | -0.29076 | -0.051606418 | 0.150221234 | 0.23915136 | 0.440979012 | 0.201827652 | 0.04162989 | 0.000768404 | 0.133745607 |
| sp\|P16562\|CRIS2_HUMAN | 4 | 40 | 76 | 33 | 0.060521 | -0.03674567 | 0.010118699 | -0.097266987 | -0.050402619 | 0.046864369 | 0.165928494 | 0.713235372 | 0.689605315 |
| sp\|Q9UIB8\|SLAF5_HUMAN | 11 | 40 | 76 | 33 | 0.063807 | 0.127551895 | 0.234294002 | 0.063744747 | 0.170486855 | 0.106742108 | 0.273903212 | 0.002265526 | 0.043794982 |
| sp\|P15085\|CBPA1_HUMAN | 30 | 40 | 76 | 33 | -0.13251 | -0.123465113 | 0.006449819 | 0.009043266 | 0.138958198 | 0.129914932 | 0.996088646 | 0.530373799 | 0.4941946 |
| sp\|P07737\|PROF1_HUMAN | 75 | 40 | 76 | 33 | 0.130939 | -0.003353463 | 0.350405248 | -0.134292294 | 0.219466416 | 0.353758711 | 0.5894748 | 0.379310393 | 0.043547763 |
| sp\|Q9Y6Z7\|COL10_HUMAN | 39 | 40 | 76 | 33 | 0.087338 | 0.176788009 | 0.116578377 | 0.089450285 | 0.029240654 | -0.060209632 | 0.085540467 | 0.830552049 | 0.370413406 |
| sp\|P39060\|COIA1_HUMAN | 7 | 40 | 76 | 33 | -0.02142 | 0.067809576 | 0.391590075 | 0.089231641 | 0.413012139 | 0.323780498 | 0.257701347 | 3.10E-08 | 9.05E-07 |
| sp\|Q15084\|PDIA6_HUMAN | 13 | 40 | 76 | 33 | -0.08837 | -0.026439989 | 0.379272558 | 0.061933696 | 0.467646243 | 0.405712547 | 0.825435815 | 0.000873733 | 0.001155566 |
| sp\|B2RNN3\|C1T9B_HUMAN | 2 | 32 | 63 | 27 | 0.117717 | 0.056655354 | -0.028265152 | -0.061061431 | -0.145981937 | -0.084920506 | 0.85462065 | 0.540016789 | 0.763194694 |
| sp\|Q9H2A7\|CXL16_HUMAN | 2674 | 40 | 76 | 33 | -0.06122 | -0.023393325 | 0.160473956 | 0.037822931 | 0.221690212 | 0.183867281 | 0.822639534 | 0.01191498 | 0.020277065 |
| sp\|Q9Y279\|VSIG4_HUMAN | 3 | 40 | 76 | 33 | -0.69033 | -0.622211852 | 0.294831233 | 0.06811592 | 0.985159005 | 0.917043085 | 0.882456386 | 1.70E-07 | 4.20E-08 |
| sp\|O00585\|CCL21_HUMAN | 2 | 40 | 76 | 33 | -0.09226 | 0.110073337 | 0.451612559 | 0.202332084 | 0.543871305 | 0.341539222 | 0.019994034 | 3.15E-08 | 9.19E-05 |
| sp\|P69849\|NOMO3_HUMAN | 7 | 40 | 76 | 33 | 0.059133 | 0.099883598 | 0.203753252 | 0.040750274 | 0.144619928 | 0.103869653 | 0.508601394 | 0.003739019 | 0.023837694 |
| sp\|P51693\|APLP1_HUMAN | 9 | 40 | 76 | 33 | 0.155975 | 0.004386748 | -0.174401974 | -0.151587796 | -0.330376519 | -0.178788722 | 0.019576832 | 6.43E-06 | 0.008527466 |
| sp\|P81172\|HEPC_HUMAN | 32 | 36 | 69 | 30 | 0.025926 | -0.032810453 | 0.120257848 | -0.05873678 | 0.094331521 | 0.153068301 | 0.716037665 | 0.552028317 | 0.139684832 |
| sp\|Q01459\|DIAC_HUMAN | 246 | 40 | 76 | 33 | 0.14634 | 0.138140988 | -0.081077499 | -0.008199079 | -0.227417566 | -0.219218487 | 0.98394502 | 0.000357816 | 9.44E-05 |
| sp\|O96019\|ACL6A_HUMAN | 3 | 8 | 14 | 5 | 0.013447 | -0.486654504 | -0.366623781 | -0.500101815 | -0.380071092 | 0.120030723 | 0.078971701 | 0.385469541 | 0.888439943 |
| sp\|P02654\|APOC1_HUMAN | 304 | 40 | 76 | 33 | 0.447107 | 0.200978877 | 0.561456897 | -0.246128541 | 0.114349479 | 0.36047802 | 0.077783237 | 0.677692435 | 0.009037536 |
| sp\|P40306\|PSB10_HUMAN | 2 | 40 | 76 | 33 | -0.36068 | -0.215363302 | 0.1967591 | 0.145313965 | 0.557436368 | 0.412122402 | 0.290800497 | 1.15E-05 | 0.000290107 |
| sp\|Q99436\|PSB7_HUMAN | 1 | 40 | 76 | 33 | -0.41317 | -0.231017016 | 0.417760503 | 0.182156932 | 0.830934451 | 0.648777519 | 0.299516637 | 2.56E-07 | 5.60E-06 |
| sp\|P43121\|MUC18_HUMAN | 40 | 40 | 76 | 33 | 0.336822 | 0.131489729 | -0.008136233 | -0.205332612 | -0.344958574 | -0.139625962 | 1.08E-05 | 1.14E-09 | 0.007198892 |
| sp\|Q969E1\|LEAP2_HUMAN | 1 | 40 | 76 | 33 | -0.53768 | -0.429433881 | 0.280206067 | 0.108248658 | 0.817888606 | 0.709639948 | 0.70093474 | 3.90E-06 | 6.27E-06 |
| sp\|Q02747\|GUC2A_HUMAN | 381 | 40 | 76 | 33 | -0.0344 | -0.131053044 | 0.205266188 | -0.096657833 | 0.239661399 | 0.336319232 | 0.41779866 | 0.027344055 | 0.000185176 |
| sp\|Q8WWA0\|ITLN1_HUMAN | 72 | 40 | 76 | 33 | 0.0891 | -0.195517499 | -0.099601003 | -0.284617341 | -0.188700845 | 0.095916496 | 0.066705509 | 0.432873405 | 0.758110403 |
| sp\|Q9Y4D7\|PLXD1_HUMAN | 2 | 40 | 76 | 33 | 0.096034 | 0.088971213 | 0.099455263 | -0.007063216 | 0.003420835 | 0.01048405 | 0.968589254 | 0.99484667 | 0.940150907 |
| sp\|P29401\|TKT_HUMAN | 7 | 40 | 76 | 33 | -0.23076 | -0.097539221 | 0.449098999 | 0.133223384 | 0.679861604 | 0.54663822 | 0.344189065 | 6.30E-08 | 8.74E-07 |
| sp\|P60174\|TPIS_HUMAN | 34 | 40 | 76 | 33 | -0.07552 | -0.015742368 | 0.279393454 | 0.059773455 | 0.354909277 | 0.295135822 | 0.792630668 | 0.004682691 | 0.00866387 |
| sp\|Q8IXL6\|FA20C_HUMAN | 26 | 40 | 76 | 33 | 0.124104 | 0.105862179 | 0.089258629 | -0.018241769 | -0.034845319 | -0.01660355 | 0.723962316 | 0.445285903 | 0.790435098 |
| sp\|Q9UJJ9\|GNPTG_HUMAN | 1 | 40 | 76 | 33 | 0.008504 | 0.045360364 | 0.223888238 | 0.036856087 | 0.215383961 | 0.178527875 | 0.555755428 | 4.30E-06 | 1.77E-05 |
| sp\|P08253\|MMP2_HUMAN | 8 | 40 | 76 | 33 | 0.206613 | 0.171547436 | 0.182229219 | -0.035065234 | -0.024383452 | 0.010681782 | 0.698529778 | 0.886914266 | 0.971091777 |
| sp\|O00151\|PDLI1_HUMAN | 2 | 40 | 76 | 33 | -0.05322 | -0.319696054 | 0.042158885 | -0.266477613 | 0.095377326 | 0.361854939 | 0.207086893 | 0.868221613 | 0.080118779 |
| sp\|P53004\|BIEA_HUMAN | 7 | 40 | 76 | 33 | -0.2009 | -0.055552402 | 0.207292025 | 0.145352138 | 0.408196565 | 0.262844427 | 0.145358996 | 5.96E-05 | 0.00475167 |
| sp\|P02775\|CXCL7_HUMAN | 13 | 40 | 76 | 33 | -0.38681 | 0.072136497 | 0.01250033 | 0.458950643 | 0.399314476 | -0.059636167 | 0.000399416 | 0.014524234 | 0.88221253 |
| sp\|P04066\|FUCO_HUMAN | 4 | 40 | 76 | 33 | 0.198694 | 0.132220063 | 0.315989101 | -0.06647394 | 0.117295099 | 0.183769039 | 0.431428157 | 0.167280075 | 0.004580844 |
| sp\|Q9BYH1\|SE6L1_HUMAN | 3 | 40 | 76 | 33 | 0.214811 | 0.162457878 | 0.041650306 | -0.052352781 | -0.173160352 | -0.120807572 | 0.380056245 | 0.001026333 | 0.012678206 |
| sp\|Q9HBB8\|CDHR5_HUMAN | 7 | 40 | 76 | 33 | 0.179966 | 0.218424572 | 0.137772142 | 0.03845872 | -0.042193709 | -0.08065243 | 0.712336716 | 0.754361969 | 0.273474114 |
| sp\|Q9BY67\|CADM1_HUMAN | 2 | 40 | 76 | 33 | 0.205371 | 0.189052077 | 0.204181075 | -0.016319255 | -0.001190258 | 0.015128997 | 0.934836197 | 0.999752524 | 0.950415926 |
| sp\|P54802\|ANAG_HUMAN | 1 | 40 | 76 | 33 | 0.183583 | 0.156264978 | 0.092865613 | -0.027318064 | -0.090717428 | -0.063399365 | 0.872879119 | 0.358886182 | 0.527565855 |
| sp\|Q9UBR2\|CATZ_HUMAN | 32 | 40 | 76 | 33 | -0.1116 | -0.092029363 | 0.152892575 | 0.01956672 | 0.264488658 | 0.244921938 | 0.927182402 | 0.00015953 | 7.67E-05 |
| sp\|P55058\|PLTP_HUMAN | 5 | 40 | 76 | 33 | 0.033958 | 0.067737381 | 0.247699168 | 0.033779156 | 0.213740944 | 0.179961788 | 0.723708233 | 0.000260305 | 0.000554064 |
| sp\|P49641\|MA2A2_HUMAN | 24 | 40 | 76 | 33 | 0.229657 | 0.136487844 | 0.006255797 | -0.093169136 | -0.223401183 | -0.130232047 | 0.004312322 | 4.93E-09 | 0.000119683 |
| sp\|P19021\|AMD_HUMAN | 52 | 40 | 76 | 33 | 0.122059 | 0.091245029 | 0.067717084 | -0.030813775 | -0.05434172 | -0.023527945 | 0.58553884 | 0.319436533 | 0.759777901 |
| sp\|P16233\|LIPP_HUMAN | 31 | 40 | 76 | 33 | -0.03807 | -0.039231146 | -0.012433026 | -0.001161393 | 0.025636727 | 0.02679812 | 0.999898437 | 0.966440904 | 0.953654328 |
| sp\|O14786\|NRP1_HUMAN | 5 | 40 | 76 | 33 | 0.132241 | 0.151239894 | 0.189070175 | 0.018999202 | 0.056829483 | 0.037830282 | 0.87221883 | 0.432706806 | 0.622394511 |
| sp\|P24043\|LAMA2_HUMAN | 28 | 40 | 76 | 33 | 0.084055 | 0.040992944 | 0.319790479 | -0.04306184 | 0.235735695 | 0.278797535 | 0.779825863 | 0.007430505 | 0.000217135 |
| sp\|Q9GZT8\|NIF3L_HUMAN | 12 | 40 | 76 | 33 | 0.019476 | 0.084843433 | 0.176357678 | 0.065367569 | 0.156881813 | 0.091514244 | 0.26432877 | 0.006158915 | 0.10357709 |
| sp\|P29322\|EPHA8_HUMAN | 22 | 7 | 13 | 7 | -0.00917 | 0.167092477 | 0.044197972 | 0.176259661 | 0.053365156 | -0.122894505 | 0.048379492 | 0.784452174 | 0.207115064 |
| sp\|P00533\|EGFR_HUMAN | 24 | 40 | 76 | 33 | 0.133033 | 0.092741898 | -0.265504985 | -0.040291562 | -0.398538446 | -0.358246883 | 0.462088984 | 3.97E-14 | 3.85E-14 |
| sp\|P08567\|PLEK_HUMAN | 2 | 40 | 76 | 33 | -0.12313 | -0.074909353 | 0.024248095 | 0.048221176 | 0.147378625 | 0.099157449 | 0.936623893 | 0.656794627 | 0.78458032 |
| sp\|P05109\|S10A8_HUMAN | 3 | 40 | 76 | 33 | -1.64433 | -1.198553667 | -0.239993561 | 0.445776803 | 1.404336908 | 0.958560105 | 0.079569605 | 2.09E-07 | 6.80E-05 |
| sp\|P14618\|KPYM_HUMAN | 23 | 40 | 76 | 33 | -0.02364 | -0.015859804 | 0.140065177 | 0.007777287 | 0.163702268 | 0.155924981 | 0.996205035 | 0.317191551 | 0.26629493 |
| sp\|P48304\|REG1B_HUMAN | 55 | 40 | 76 | 33 | -0.69405 | -0.573973476 | -0.036266062 | 0.120074137 | 0.657781551 | 0.537707415 | 0.800599931 | 0.012153332 | 0.022961451 |
| sp\|P12829\|MYL4_HUMAN | 262 | 28 | 55 | 25 | -1.62706 | -1.797625782 | -1.24207553 | -0.170566999 | 0.384983253 | 0.555550253 | 0.672567715 | 0.241946654 | 0.023946523 |
| sp\|P13497\|BMP1_HUMAN | 297 | 40 | 76 | 33 | 0.070552 | 0.144236187 | 0.068264082 | 0.073684144 | -0.002287961 | -0.075972105 | 0.274283387 | 0.99912792 | 0.298656905 |
| sp\|Q53GG5\|PDLI3_HUMAN | 243 | 4 | 6 | 3 | 0.192592 | 0.170532917 | 1.277432168 | -0.022059216 | 1.084840035 | 1.106899252 | 0.999335272 | 0.356939479 | 0.293655555 |
| sp\|O75106\|AOC2_HUMAN | 4 | 40 | 76 | 33 | 0.361579 | 0.277276374 | 0.26161398 | -0.084302504 | -0.099964898 | -0.015662394 | 0.242448274 | 0.252762468 | 0.957363661 |
| sp\|P31948\|STIP1_HUMAN | 47 | 40 | 76 | 33 | -0.23306 | -0.216422222 | 0.212838496 | 0.016635062 | 0.44589578 | 0.429260718 | 0.990577014 | 0.011315892 | 0.005271534 |
| sp\|P20810\|ICAL_HUMAN | 2 | 40 | 76 | 33 | -0.5702 | -0.321648471 | 0.224364351 | 0.248553667 | 0.794566489 | 0.546012822 | 0.028134151 | 4.78E-10 | 1.06E-06 |
| sp\|Q9NVA2\|SEP11_HUMAN | 4 | 29 | 55 | 23 | 0.004605 | -0.075392948 | 0.437237399 | -0.079997705 | 0.432632643 | 0.512630347 | 0.893830359 | 0.115561936 | 0.023463582 |
| sp\|Q99733\|NP1L4_HUMAN | 20 | 40 | 76 | 33 | -0.22323 | -0.235556183 | 0.169852464 | -0.012321358 | 0.393087289 | 0.405408647 | 0.995632728 | 0.050822941 | 0.018470508 |
| sp\|P14555\|PA2GA_HUMAN | 3 | 36 | 69 | 30 | -0.69869 | -0.629444372 | 0.299465217 | 0.069245873 | 0.998155463 | 0.92890959 | 0.886390621 | 3.39E-07 | 8.55E-08 |
| sp\|Q01813\|PFKAP_HUMAN | 3 | 40 | 76 | 33 | 0.095332 | -0.025858808 | 0.226809188 | -0.121190402 | 0.131477594 | 0.252667996 | 0.519670925 | 0.587281579 | 0.086162147 |
| sp\|P06753\|TPM3_HUMAN | 38 | 40 | 76 | 33 | -0.00734 | -0.207415191 | 0.31793214 | -0.200071219 | 0.325276111 | 0.525347331 | 0.482334442 | 0.266746458 | 0.014172686 |
| sp\|P07954\|FUMH_HUMAN | 1 | 40 | 76 | 33 | -0.49362 | -0.304354305 | 0.037638865 | 0.189267829 | 0.531260999 | 0.34199317 | 0.081803154 | 4.15E-06 | 0.001035777 |
| sp\|P27824\|CALX_HUMAN | 4 | 40 | 76 | 33 | -0.20648 | -0.253629497 | 0.337030801 | -0.047151814 | 0.543508483 | 0.590660297 | 0.827714605 | 2.90E-07 | 4.87E-10 |
| sp\|Q86VP6\|CAND1_HUMAN | 2 | 40 | 76 | 33 | -0.22863 | -0.048789506 | 0.267043164 | 0.179835956 | 0.495668726 | 0.31583267 | 0.065049061 | 2.26E-06 | 0.000834426 |
| sp\|Q15293\|RCN1_HUMAN | 10 | 40 | 76 | 33 | 0.001307 | -0.020955957 | -0.039302323 | -0.022262706 | -0.040609073 | -0.018346366 | 0.860605924 | 0.708984099 | 0.914327416 |
| sp\|Q9UPZ6\|THS7A_HUMAN | 5 | 40 | 76 | 33 | 0.125812 | -0.077984934 | 0.018810434 | -0.203797292 | -0.107001925 | 0.096795368 | 0.000350919 | 0.199683367 | 0.187043768 |
| sp\|P80723\|BASP1_HUMAN | 63 | 40 | 76 | 33 | -0.1464 | -0.167955149 | 0.390172519 | -0.02155475 | 0.536572918 | 0.558127668 | 0.971995847 | 1.82E-05 | 4.77E-07 |
| sp\|Q01995\|TAGL_HUMAN | 157 | 40 | 76 | 33 | -0.13773 | -0.198935341 | 0.369203109 | -0.06120706 | 0.50693139 | 0.56813845 | 0.694724781 | 3.05E-07 | 1.66E-10 |
| sp\|P29966\|MARCS_HUMAN | 364 | 40 | 76 | 33 | -0.31838 | -0.160840707 | 0.396898105 | 0.157540496 | 0.715279308 | 0.557738812 | 0.343636322 | 1.38E-06 | 2.25E-05 |
| sp\|P63316\|TNNC1_HUMAN | 37 | 14 | 27 | 13 | -1.60155 | -1.827927083 | -1.167331434 | -0.226380882 | 0.434214766 | 0.660595648 | 0.849153619 | 0.645964938 | 0.27509754 |
| sp\|P02585\|TNNC2_HUMAN | 12 | 10 | 20 | 9 | 0.07487 | 0.028474087 | 0.472774962 | -0.04639558 | 0.397905295 | 0.444300875 | 0.983142775 | 0.421099548 | 0.249155781 |
| sp\|P35908\|K22E_HUMAN | 5 | 40 | 76 | 33 | 0.681383 | 0.384533818 | 0.213165653 | -0.296848905 | -0.46821707 | -0.171368165 | 0.160895975 | 0.045253862 | 0.581665048 |
| sp\|Q12882\|DPYD_HUMAN | 28 | 40 | 76 | 33 | -0.29597 | -0.10486398 | 0.152873433 | 0.19110546 | 0.448842873 | 0.257737414 | 0.002014477 | 9.41E-10 | 6.76E-05 |
| sp\|P52758\|RIDA_HUMAN | 4 | 37 | 69 | 30 | -0.26391 | -0.120206083 | 0.354199816 | 0.143701002 | 0.618106901 | 0.474405899 | 0.370277453 | 1.17E-05 | 0.000171888 |
| sp\|P28838\|AMPL_HUMAN | 7 | 40 | 76 | 33 | -0.3537 | -0.08399013 | 0.279576916 | 0.269706784 | 0.63327383 | 0.363567046 | 0.078023271 | 0.00011117 | 0.018139065 |
| sp\|Q96RW7\|HMCN1_HUMAN | 6 | 40 | 76 | 33 | -0.11237 | 0.016240674 | 0.302969563 | 0.128612788 | 0.415341676 | 0.286728889 | 0.031985878 | 6.57E-10 | 1.17E-06 |
| sp\|Q5JRA6\|TGO1_HUMAN | 9 | 40 | 76 | 33 | -0.71869 | -0.617746316 | 0.166067747 | 0.100946278 | 0.884760341 | 0.783814063 | 0.647143994 | 3.89E-09 | 3.98E-09 |
| sp\|P05386\|RLA1_HUMAN | 104 | 40 | 76 | 33 | -0.40935 | -0.191317515 | 0.462402182 | 0.218035132 | 0.87175483 | 0.653719697 | 0.086116519 | 1.56E-10 | 4.42E-08 |

| | | | | | | | | | | | | |
|---|---|---|---|---|---|---|---|---|---|---|---|---|
| sp\|Q15833\|STXB2_HUMAN | 11 | 40 | 76 | 33 | 0.019239 | -0.149283798 | -0.006284893 | -0.168522833 | -0.025523929 | 0.142998904 | 0.314275532 | 0.981627547 | 0.479504913 |
| sp\|Q9UQ80\|PA2G4_HUMAN | 2 | 40 | 76 | 33 | -0.24792 | -0.229954693 | 0.071646855 | 0.017969167 | 0.319570715 | 0.301601547 | 0.978743111 | 0.011357274 | 0.00642988 |
| sp\|Q8WUJ3\|CEMIP_HUMAN | 5 | 40 | 76 | 33 | -0.07853 | -0.04207654 | 0.297640268 | 0.036450108 | 0.376166917 | 0.339716808 | 0.726560064 | 2.96E-09 | 1.57E-09 |
| sp\|P80511\|S10AC_HUMAN | 419 | 36 | 69 | 30 | -1.39417 | -1.11856984 | -0.245439346 | 0.275603294 | 1.148733789 | 0.873130494 | 0.416326916 | 6.75E-05 | 0.000704237 |
| sp\|P08319\|ADH4_HUMAN | 3 | 40 | 76 | 33 | -0.65984 | -0.339475607 | -0.091729393 | 0.320362015 | 0.568108229 | 0.247746214 | 0.057242696 | 0.002441923 | 0.218086519 |
| sp\|Q8N8Z6\|DCBD1_HUMAN | 2 | 40 | 76 | 33 | -0.01986 | 0.002329385 | 0.106028295 | 0.022191117 | 0.125890028 | 0.10369891 | 0.750121677 | 0.002422984 | 0.005300981 |
| sp\|O43747\|AP1G1_HUMAN | 269 | 29 | 55 | 24 | 0.049718 | -0.191307166 | -0.181454247 | -0.241024694 | -0.231171775 | 0.009852919 | 0.194226623 | 0.349698127 | 0.997540303 |
| sp\|P54920\|SNAA_HUMAN | 27 | 40 | 76 | 33 | -0.20573 | -0.225879349 | 0.177602422 | -0.020154247 | 0.383327523 | 0.40348177 | 0.976263741 | 0.003481912 | 0.00040284 |
| sp\|P09417\|DHPR_HUMAN | 282 | 40 | 76 | 33 | 0.058336 | 0.100535837 | 0.340296879 | 0.042199519 | 0.281960562 | 0.239761043 | 0.792050432 | 0.001200737 | 0.001993975 |
| sp\|P08727\|K1C19_HUMAN | 4 | 14 | 28 | 12 | -0.21435 | -0.631172412 | -0.675206059 | -0.416823995 | -0.460857642 | -0.044033647 | 0.598206194 | 0.646845012 | 0.994790638 |
| sp\|P17181\|INAR1_HUMAN | 1 | 36 | 69 | 30 | 0.149187 | 0.053800421 | 0.081971846 | -0.095386824 | -0.067215399 | 0.028171426 | 0.082172415 | 0.417279165 | 0.820623349 |
| sp\|Q8N4F0\|BPIB2_HUMAN | 29 | 15 | 28 | 12 | 0.01672 | -0.19040573 | 0.865449376 | -0.207125826 | 0.84872928 | 1.055855106 | 0.671010857 | 0.015104739 | 0.000511432 |
| sp\|P25787\|PSA2_HUMAN | 39 | 40 | 76 | 33 | -0.53475 | -0.397245882 | 0.177564758 | 0.137502617 | 0.712313257 | 0.57481064 | 0.434741215 | 1.18E-06 | 9.89E-06 |
| sp\|P42765\|THIM_HUMAN | 3 | 40 | 76 | 33 | -0.49653 | -0.356524431 | 0.179779995 | 0.140009689 | 0.676314115 | 0.536304425 | 0.558596954 | 0.000174473 | 0.000884359 |
| sp\|Q8N436\|CPXM2_HUMAN | 35 | 28 | 55 | 24 | -1.09592 | -1.076525019 | -0.5669982 | 0.019396656 | 0.528923475 | 0.509526819 | 0.987304867 | 0.00221221 | 0.00072111 |
| sp\|Q13449\|LSAMP_HUMAN | 32 | 40 | 76 | 33 | 0.142031 | 0.030251564 | -0.022141601 | -0.11177894 | -0.164172104 | -0.052393164 | 0.026414641 | 0.004898706 | 0.485598936 |
| sp\|Q9Y4L1\|HYOU1_HUMAN | 1 | 40 | 76 | 33 | 0.009288 | 0.057160359 | 0.25878506 | 0.047872111 | 0.249496812 | 0.201624701 | 0.434607229 | 1.01E-06 | 8.42E-06 |
| sp\|P08590\|MYL3_HUMAN | 12 | 36 | 69 | 30 | -2.09039 | -2.270948972 | -1.735967855 | -0.180555782 | 0.354425335 | 0.534981117 | 0.54858317 | 0.205427436 | 0.011506822 |
| sp\|Q9Y639\|NPTN_HUMAN | 14 | 4 | 6 | 3 | 0.557826 | 0.238293572 | 0.453853222 | -0.319532804 | -0.103973154 | 0.215559651 | 0.230305886 | 0.879784663 | 0.54312262 |
| sp\|O43508\|TNF12_HUMAN | 7 | 15 | 28 | 12 | 0.43171 | 0.392749971 | 0.405919376 | -0.038959642 | -0.025790237 | 0.013169404 | 0.949463663 | 0.984596329 | 0.994911607 |
| sp\|P11216\|PYGB_HUMAN | 20 | 40 | 76 | 33 | -0.04684 | -0.10683617 | 0.339929218 | -0.05999741 | 0.386767977 | 0.446765388 | 0.848454047 | 0.011045911 | 0.000589517 |
| sp\|Q92823\|NRCAM_HUMAN | 3 | 40 | 76 | 33 | 0.063941 | 0.049185724 | 0.071900354 | -0.014755587 | 0.007959043 | 0.02271463 | 0.955221635 | 0.990842084 | 0.909132326 |
| sp\|O94856\|NFASC_HUMAN | 13 | 29 | 55 | 24 | -0.11245 | -0.286263705 | -0.07848941 | -0.17380926 | 0.033965035 | 0.207774295 | 0.288563917 | 0.967193863 | 0.210891739 |
| sp\|Q15828\|CYTM_HUMAN | 7 | 40 | 76 | 33 | 0.044668 | -0.029915717 | 0.017499344 | -0.074583461 | -0.027168401 | 0.047415061 | 0.246708265 | 0.878208743 | 0.605710722 |
| sp\|P37837\|TALDO_HUMAN | 11 | 40 | 76 | 33 | -0.16175 | -0.063965472 | 0.501549889 | 0.097783757 | 0.663299118 | 0.565515361 | 0.525782325 | 2.64E-08 | 8.35E-08 |
| sp\|P0DN79\|CBSL_HUMAN | 26 | 26 | 48 | 20 | -0.67358 | -0.156934884 | 0.158366731 | 0.516649794 | 0.831951408 | 0.315301614 | 0.005785066 | 0.000194087 | 0.184908616 |
| sp\|P12318\|FCG2A_HUMAN | 11 | 40 | 76 | 33 | -0.01777 | -0.026969257 | 0.098989126 | -0.009192094 | 0.116761288 | 0.125953382 | 0.986096085 | 0.215704122 | 0.10495275 |
| sp\|O75071\|EFC14_HUMAN | 2 | 20 | 34 | 14 | -0.09069 | -0.044404964 | 0.437566819 | 0.046282939 | 0.528254722 | 0.481971783 | 0.96703706 | 0.066325079 | 0.065914744 |
| sp\|P07942\|LAMB1_HUMAN | 5 | 40 | 76 | 33 | 0.036503 | 0.029714388 | 0.21193838 | -0.006788594 | 0.175435398 | 0.182223992 | 0.985808081 | 0.002059315 | 0.000243221 |
| sp\|O75112\|LDB3_HUMAN | 238 | 10 | 20 | 9 | -0.74525 | -0.825486623 | -0.076699257 | -0.080238309 | 0.668549056 | 0.748787365 | 0.979747616 | 0.375696805 | 0.206204894 |
| sp\|Q8WWQ8\|STAB2_HUMAN | 39 | 40 | 76 | 33 | -0.03356 | -0.010701659 | -0.031147537 | 0.022856677 | 0.002410799 | -0.020445878 | 0.815513644 | 0.998432423 | 0.866436176 |
| sp\|P09525\|ANXA4_HUMAN | 4 | 40 | 76 | 33 | 0.425433 | 0.378029922 | 0.590869299 | -0.047403266 | 0.165436111 | 0.212839377 | 0.903717933 | 0.429950451 | 0.1719159 |
| sp\|Q9Y3F4\|STRAP_HUMAN | 14 | 40 | 76 | 33 | -0.28398 | -0.094416102 | 0.14044064 | 0.189559222 | 0.424415964 | 0.234856742 | 0.124219086 | 0.00102955 | 0.061382397 |
| sp\|P80303\|NUCB2_HUMAN | 2 | 40 | 76 | 33 | 0.152503 | 0.1243417 | 0.397221615 | -0.028161628 | 0.244718287 | 0.272879915 | 0.921504404 | 0.016804745 | 0.001791615 |
| sp\|P48594\|SPB4_HUMAN | 5 | 17 | 34 | 16 | -0.13451 | -0.387044132 | -0.656662688 | -0.252529594 | -0.52214815 | -0.269618556 | 0.591012768 | 0.201596531 | 0.562718896 |
| sp\|P11766\|ADHX_HUMAN | 24 | 40 | 76 | 33 | -0.0777 | -0.04336476 | 0.240916126 | 0.034338737 | 0.318619624 | 0.284280887 | 0.913113757 | 0.005932261 | 0.005568278 |
| sp\|Q8NDA2\|HMCN2_HUMAN | 35 | 40 | 76 | 33 | 0.109385 | 0.111681282 | -0.164877719 | 0.002296298 | -0.274262703 | -0.276559001 | 0.99921976 | 0.000786683 | 0.00011364 |
| sp\|Q9H2X3\|CLC4M_HUMAN | 3 | 40 | 76 | 33 | -0.1489 | -0.082870931 | 0.074736937 | 0.066031646 | 0.223639514 | 0.157607869 | 0.46081989 | 0.002951152 | 0.023338521 |
| sp\|P13716\|HEM2_HUMAN | 311 | 40 | 76 | 33 | -0.08511 | 0.036628935 | 0.35106773 | 0.121736565 | 0.43617536 | 0.314438794 | 0.335738177 | 0.000130089 | 0.002290035 |
| sp\|P07327\|ADH1A_HUMAN | 290 | 40 | 76 | 33 | -0.54792 | -0.249229489 | 0.011009361 | 0.298687993 | 0.558926842 | 0.26023885 | 0.094130608 | 0.003962561 | 0.204395991 |
| sp\|Q9Y2E5\|MA2B2_HUMAN | 36 | 40 | 76 | 33 | 0.002866 | -0.033141523 | 0.043689117 | -0.036007702 | 0.040822938 | 0.07683064 | 0.875846495 | 0.889065419 | 0.589980919 |
| sp\|Q9HBW9\|AGRL4_HUMAN | 91 | 40 | 76 | 33 | 0.051385 | -0.04905798 | -0.0505373 | -0.10044273 | -0.101922051 | -0.00147932 | 0.346221606 | 0.469544752 | 0.999795726 |
| sp\|P04083\|ANXA1_HUMAN | 29 | 40 | 76 | 33 | -0.93261 | -0.61537317 | 0.017196012 | 0.317241723 | 0.949810905 | 0.632569182 | 0.097516124 | 2.23E-06 | 0.00044989 |
| sp\|Q9Y623\|MYH4_HUMAN | 3 | 40 | 76 | 33 | 0.155193 | 0.023616799 | -0.003813989 | -0.131576205 | -0.159006993 | -0.027430788 | 0.000497478 | 0.000471172 | 0.731779221 |
| sp\|Q14563\|SEM3A_HUMAN | 1 | 40 | 76 | 33 | -0.01549 | -0.048066891 | 0.023679955 | -0.032577471 | 0.039169374 | 0.071746845 | 0.694897194 | 0.695538323 | 0.216307322 |
| sp\|P07911\|UROM_HUMAN | 20 | 40 | 76 | 33 | 0.17847 | 0.08228941 | -0.045005537 | -0.096180624 | -0.223475571 | -0.127294947 | 0.464988136 | 0.06125235 | 0.309561696 |
| sp\|P00558\|PGK1_HUMAN | 19 | 40 | 76 | 33 | -0.05701 | -0.010520251 | 0.379054069 | 0.046492596 | 0.436066916 | 0.38957432 | 0.890337486 | 0.001357911 | 0.001232618 |
| sp\|O95393\|BMP10_HUMAN | 9 | 40 | 76 | 33 | 0.120323 | -0.030459694 | -0.037028053 | -0.15078288 | -0.157351238 | -0.006568358 | 0.010236279 | 0.030778811 | 0.992029176 |
| sp\|Q9H939\|PPIP2_HUMAN | 9 | 40 | 76 | 33 | -0.49706 | -0.434273998 | -0.293924571 | 0.062781623 | 0.20313105 | 0.140349427 | 0.911715413 | 0.515016915 | 0.667511662 |
| sp\|P21860\|ERBB3_HUMAN | 7 | 40 | 76 | 33 | 0.302063 | 0.311005564 | 0.043626096 | 0.008942989 | -0.258436479 | -0.267379468 | 0.989311463 | 0.002925044 | 0.000410018 |
| sp\|Q02763\|TIE2_HUMAN | 28 | 40 | 76 | 33 | 0.030214 | 0.031708326 | 0.021568467 | 0.001494624 | -0.008645235 | -0.010139859 | 0.998927984 | 0.975549675 | 0.957601651 |
| sp\|Q14847\|LASP1_HUMAN | 22 | 40 | 76 | 33 | 0.005907 | -0.032363878 | 0.231794945 | -0.038270434 | 0.22588839 | 0.264158823 | 0.907338404 | 0.101849862 | 0.020044994 |
| sp\|P22234\|PUR6_HUMAN | 12 | 22 | 42 | 18 | -0.33633 | -0.135437843 | 0.382180622 | 0.200888131 | 0.718506596 | 0.517618465 | 0.333455596 | 0.000190838 | 0.002754873 |
| sp\|P49407\|ARRB1_HUMAN | 26 | 40 | 76 | 33 | -0.1131 | -0.157744711 | 0.038413053 | -0.044648681 | 0.151509083 | 0.196157764 | 0.943900214 | 0.633337149 | 0.379478908 |
| sp\|P22897\|MRC1_HUMAN | 15 | 40 | 76 | 33 | 0.040948 | 0.034831908 | 0.67623928 | -0.006116287 | 0.635291084 | 0.641407371 | 0.996951001 | 5.35E-09 | 4.78E-11 |
| sp\|Q96HD1\|CREL1_HUMAN | 7 | 40 | 76 | 33 | -0.03157 | 0.034645991 | 0.407541958 | 0.066212961 | 0.439108929 | 0.372895968 | 0.579822678 | 5.11E-07 | 1.49E-06 |
| sp\|P12814\|ACTN1_HUMAN | 3 | 40 | 76 | 33 | 0.121978 | -0.19590492 | 0.079079388 | -0.317882485 | -0.042898177 | 0.274984309 | 0.125548844 | 0.973708055 | 0.253260602 |
| sp\|O43707\|ACTN4_HUMAN | 19 | 40 | 76 | 33 | 0.034075 | -0.241070796 | 0.089800531 | -0.275145646 | 0.055725682 | 0.330871327 | 0.142457906 | 0.945287744 | 0.085343382 |
| sp\|P13929\|ENOB_HUMAN | 32 | 40 | 76 | 33 | 0.190285 | 0.224247186 | 0.281298561 | 0.03396246 | 0.091013834 | 0.057051375 | 0.866779614 | 0.494500899 | 0.702283557 |
| sp\|Q9H8J5\|MANS1_HUMAN | 4 | 40 | 76 | 33 | 0.035992 | 0.08162125 | 0.208510474 | 0.045629189 | 0.172518413 | 0.126889224 | 0.623880504 | 0.011492882 | 0.044319846 |
| sp\|O00451\|GFRA2_HUMAN | 4 | 40 | 76 | 33 | -0.02915 | -0.034417515 | 0.077938431 | -0.005267311 | 0.107088636 | 0.112355946 | 0.994539478 | 0.215142749 | 0.117806351 |
| sp\|Q8NHP8\|PLBL2_HUMAN | 31 | 36 | 69 | 30 | 0.160471 | 0.077875425 | 0.30129576 | -0.082595829 | 0.140824506 | 0.223420335 | 0.581522576 | 0.33845037 | 0.033495316 |
| sp\|Q8WWZ8\|OIT3_HUMAN | 69 | 40 | 76 | 33 | -0.09657 | 0.078302219 | 0.30558761 | 0.174870728 | 0.402156118 | 0.227285391 | 0.011335136 | 3.51E-07 | 0.001482724 |
| sp\|O15197\|EPHB6_HUMAN | 7 | 8 | 13 | 6 | -0.05621 | -0.119656468 | 0.291999016 | -0.063446867 | 0.348208617 | 0.411655484 | 0.926152782 | 0.223429128 | 0.090230368 |
| sp\|Q6YHK3\|CD109_HUMAN | 9 | 40 | 76 | 33 | 0.300341 | 0.22069223 | -0.008263013 | -0.079649046 | -0.308604289 | -0.228955243 | 0.18612152 | 2.21E-07 | 1.46E-05 |
| sp\|Q9NY47\|CA2D2_HUMAN | 10 | 40 | 76 | 33 | 0.164123 | 0.011973753 | -0.349971488 | -0.152149178 | -0.514094418 | -0.361945241 | 0.018303233 | 5.14E-12 | 2.33E-08 |
| sp\|Q8IUL8\|CILP2_HUMAN | 16 | 40 | 76 | 33 | 0.07398 | 0.118471015 | -0.52157901 | 0.044491357 | -0.595558667 | -0.640050024 | 0.862613593 | 1.43E-07 | 2.81E-10 |
| sp\|Q15262\|PTPRK_HUMAN | 159 | 40 | 76 | 33 | 0.009958 | 0.050432487 | 0.12310627 | 0.040474697 | 0.113148479 | 0.072673782 | 0.52019835 | 0.032574475 | 0.16086125 |
| sp\|Q99719\|SEPT5_HUMAN | 30 | 40 | 76 | 33 | -0.27137 | -0.181269452 | -0.102563725 | 0.090105034 | 0.16881076 | 0.078705726 | 0.75598484 | 0.509352459 | 0.828967562 |
| sp\|Q6NUS6\|TECT3_HUMAN | 2 | 40 | 76 | 33 | -0.06935 | -0.046805752 | -0.001201687 | 0.022543274 | 0.068147339 | 0.045604065 | 0.79491652 | 0.239573353 | 0.440652332 |
| sp\|P00441\|SODC_HUMAN | 2 | 40 | 76 | 33 | -0.36056 | -0.200358246 | 0.256677957 | 0.160203165 | 0.617239368 | 0.457036203 | 0.335688195 | 3.62E-05 | 0.000656213 |
| sp\|P48739\|PIPNB_HUMAN | 39 | 25 | 48 | 21 | -0.1428 | -0.090916424 | 0.119560259 | 0.051880558 | 0.262357241 | 0.210476683 | 0.889178649 | 0.131641539 | 0.186571616 |
| sp\|P61970\|NTF2_HUMAN | 17 | 40 | 76 | 33 | -0.18685 | -0.058105199 | 0.247892214 | 0.128749335 | 0.434746748 | 0.305997412 | 0.248599979 | 4.26E-05 | 0.001423172 |
| sp\|P35579\|MYH9_HUMAN | 6 | 40 | 76 | 33 | -0.58797 | -0.073992828 | 0.327624454 | 0.513978013 | 0.915595295 | 0.401617282 | 2.72E-06 | 1.05E-11 | 0.000725557 |
| sp\|P00390\|GSHR_HUMAN | 4 | 40 | 76 | 33 | -0.12592 | 0.149517036 | 0.329785421 | 0.27543895 | 0.455707335 | 0.180268385 | 3.73E-09 | 4.53E-14 | 0.000316139 |
| sp\|P35237\|SPB6_HUMAN | 15 | 40 | 76 | 33 | -0.13208 | -0.160262427 | 0.168018504 | -0.02818182 | 0.300099111 | 0.328280931 | 0.923787615 | 0.002886415 | 0.000171762 |
| sp\|P09622\|DLDH_HUMAN | 112 | 40 | 76 | 33 | 0.045724 | 0.037508694 | 0.123736235 | -0.008214951 | 0.07801259 | 0.086227541 | 0.991932334 | 0.605691926 | 0.459709684 |
| sp\|Q13231\|CHIT1_HUMAN | 24 | 40 | 76 | 33 | -0.44223 | -0.618768446 | 0.15301586 | -0.176536341 | 0.595247965 | 0.771784306 | 0.609277116 | 0.023337545 | 0.000437833 |
| sp\|P07307\|ASGR2_HUMAN | 3 | 40 | 76 | 33 | -0.19645 | -0.103679324 | 0.252585667 | 0.092773564 | 0.449038555 | 0.356264991 | 0.530618947 | 8.31E-05 | 0.000476496 |
| sp\|O00748\|EST2_HUMAN | 2 | 40 | 76 | 33 | 0.188686 | 0.14917813 | 0.167316555 | -0.039507599 | -0.021369173 | 0.018138426 | 0.593066946 | 0.899486346 | 0.907440866 |
| sp\|Q09666\|AHNK_HUMAN | 19 | 40 | 76 | 33 | -0.37453 | -0.359909737 | 0.201820575 | 0.014616801 | 0.576347114 | 0.561730313 | 0.985323021 | 9.32E-07 | 7.37E-08 |
| sp\|Q13418\|ILK_HUMAN | 4 | 40 | 76 | 33 | 0.042332 | -0.083490111 | -0.012888802 | -0.12582217 | -0.055220861 | 0.070601309 | 0.661287361 | 0.946320083 | 0.891641522 |
| sp\|O95747\|OXSR1_HUMAN | 59 | 27 | 48 | 21 | -0.22446 | -0.100452866 | 0.106557726 | 0.124011775 | 0.331022367 | 0.207010592 | 0.57147091 | 0.071014755 | 0.271447764 |
| sp\|Q99523\|SORT_HUMAN | 9 | 40 | 76 | 33 | -0.12348 | -0.135755352 | 0.168723625 | -0.012272531 | 0.292206446 | 0.304478977 | 0.978380596 | 0.000370915 | 2.39E-05 |
| sp\|P07900\|HS90A_HUMAN | 2 | 40 | 76 | 33 | -0.20665 | -0.137328577 | 0.265674007 | 0.069323062 | 0.472325645 | 0.403002584 | 0.801342597 | 0.001297917 | 0.002060238 |
| sp\|P20701\|ITAL_HUMAN | 28 | 40 | 76 | 33 | 0.212468 | 0.266908576 | 0.34031889 | 0.054440896 | 0.12785121 | 0.073410314 | 0.318944428 | 0.014791348 | 0.163398327 |
| sp\|P50453\|SPB9_HUMAN | 12 | 40 | 76 | 33 | -0.17553 | -0.048551572 | 0.162426329 | 0.126977854 | 0.337955755 | 0.210977901 | 0.324491997 | 0.005123334 | 0.068277874 |
| sp\|O75635\|SPB7_HUMAN | 19 | 4 | 6 | 3 | -0.13861 | 0.019646755 | -0.078373417 | 0.15825369 | 0.060233518 | -0.098020172 | 0.300837134 | 0.870118448 | 0.658732711 |
| sp\|P30740\|ILEU_HUMAN | 8 | 40 | 76 | 33 | -0.16577 | -0.093950031 | 0.398283623 | 0.071821327 | 0.564054981 | 0.492233654 | 0.781510497 | 6.99E-05 | 9.18E-05 |
| sp\|P36871\|PGM1_HUMAN | 14 | 40 | 76 | 33 | -0.10083 | -0.063341947 | 0.157596737 | 0.037484523 | 0.258423207 | 0.220938684 | 0.876815627 | 0.015987051 | 0.021118236 |
| sp\|Q86SR1\|GLT10_HUMAN | 4 | 40 | 76 | 33 | 0.092325 | 0.089279731 | 0.222047595 | -0.00304487 | 0.129722994 | 0.132767864 | 0.995920581 | 0.007679448 | 0.001670572 |
| sp\|Q9UNF0\|PACN2_HUMAN | 6 | 40 | 76 | 33 | -0.16043 | -0.398599102 | -0.102049236 | -0.238172208 | 0.058377658 | 0.296549866 | 0.121678679 | 0.914671801 | 0.058115682 |
| sp\|P26572\|MGAT1_HUMAN | 10 | 40 | 76 | 33 | -0.10428 | 0.01666867 | 0.199753947 | 0.120947066 | 0.304032343 | 0.183085277 | 0.066670376 | 1.82E-05 | 0.004951243 |
| sp\|P31146\|COR1A_HUMAN | 269 | 40 | 76 | 33 | -0.12015 | -0.17148238 | 0.290615618 | -0.051332141 | 0.410765856 | 0.462097997 | 0.90144065 | 0.012451079 | 0.001015054 |
| sp\|Q9H4M9\|EHD1_HUMAN | 30 | 40 | 76 | 33 | -0.06358 | -0.106563719 | 0.11604133 | -0.042978726 | 0.179626323 | 0.222605049 | 0.925900729 | 0.39868078 | 0.168479293 |
| sp\|P55285\|CADH6_HUMAN | 51 | 40 | 76 | 33 | 0.095758 | -0.022815486 | 0.073636027 | -0.118573199 | -0.022121686 | 0.096451513 | 0.199086011 | 0.961352764 | 0.388963853 |
| sp\|Q02952\|AKA12_HUMAN | 5 | 40 | 76 | 33 | -0.06091 | -0.005610404 | 0.040470126 | 0.05530307 | 0.101383601 | 0.04608053 | 0.500123038 | 0.203600707 | 0.654697543 |
| sp\|Q9Y4E8\|UBP15_HUMAN | 12 | 33 | 62 | 26 | -0.32586 | -0.163386554 | 0.260979896 | 0.16247421 | 0.58684066 | 0.42436645 | 0.351082973 | 0.000210762 | 0.003191925 |
| sp\|P48668\|K2C6C_HUMAN | 48 | 40 | 76 | 33 | -0.46462 | -0.408574507 | -0.404770412 | 0.056045151 | 0.059849247 | 0.003804096 | 0.931323841 | 0.945548477 | 0.999712032 |
| sp\|Q16181\|SEPT7_HUMAN | 9 | 40 | 76 | 33 | -0.15983 | -0.192410004 | 0.167021778 | -0.03258468 | 0.326847102 | 0.359431782 | 0.96996689 | 0.125957185 | 0.042723837 |
| sp\|Q9UQP3\|TENN_HUMAN | 16 | 40 | 76 | 33 | 0.046036 | 0.17744678 | 0.208209622 | 0.131410936 | 0.162173778 | 0.030762842 | 0.036598314 | 0.031111286 | 0.848358201 |
| sp\|O43493\|TGON2_HUMAN | 930 | 40 | 76 | 33 | -0.08002 | -0.106939609 | 0.251201405 | -0.026918421 | 0.331222593 | 0.358141013 | 0.940020082 | 0.00229366 | 0.000149051 |
| sp\|P08575\|PTPRC_HUMAN | 3 | 40 | 76 | 33 | -0.02292 | 0.071836936 | 0.082330376 | 0.094760015 | 0.105253455 | 0.01049344 | 0.074876313 | 0.10915777 | 0.971739924 |
| sp\|Q7RTS7\|K2C74_HUMAN | 15 | 4 | 7 | 2 | 0.138993 | 0.1367658 | 1.416073771 | -0.002227202 | 1.27708077 | 1.279307971 | 0.999967624 | 0.024327149 | 0.01587858 |
| sp\|P04264\|K2C1_HUMAN | 22 | 40 | 76 | 33 | 0.466468 | 0.240045832 | 0.186537758 | -0.226421918 | -0.279929992 | -0.053508074 | 0.343350057 | 0.324097188 | 0.948393372 |

| | | | | | | | | | | | | | |
|---|---|---|---|---|---|---|---|---|---|---|---|---|---|
| sp\|Q969T9\|WBP2_HUMAN | 271 | 33 | 62 | 27 | -0.04655 | -0.122013732 | 0.245844017 | -0.075467834 | 0.292389915 | 0.367857749 | 0.709538452 | 0.032610349 | 0.001336972 |
| sp\|Q8WZ42\|TITIN_HUMAN | 32 | 40 | 76 | 33 | -0.55802 | -0.750884428 | -0.272877896 | -0.192862631 | 0.285143901 | 0.478006531 | 0.320853112 | 0.181892072 | 0.002891602 |
| sp\|P10721\|KIT_HUMAN | 4 | 40 | 76 | 33 | 0.227579 | 0.153017148 | -0.2789374 | -0.074561373 | -0.506515921 | -0.431954548 | 0.331019549 | 8.91E-13 | 4.77E-12 |
| sp\|P48163\|MAOX_HUMAN | 14 | 40 | 76 | 33 | 0.09831 | 0.145706107 | 0.240464727 | 0.047395807 | 0.142154427 | 0.09475862 | 0.650682698 | 0.073738804 | 0.22507563 |
| sp\|P01133\|EGF_HUMAN | 7 | 37 | 69 | 29 | 0.068944 | 0.127317933 | 0.186167797 | 0.05837423 | 0.117224094 | 0.058849864 | 0.554769359 | 0.204754788 | 0.601609074 |
| sp\|Q03252\|LMNB2_HUMAN | 14 | 4 | 7 | 3 | 0.151853 | -0.042170483 | 1.400324825 | -0.194023381 | 1.248471928 | 1.442495309 | 0.949367796 | 0.276383236 | 0.139931971 |
| sp\|P15086\|CBPB1_HUMAN | 6 | 40 | 76 | 33 | -0.20066 | -0.180746662 | -0.011190155 | 0.019911474 | 0.189467981 | 0.169556507 | 0.976982339 | 0.238539303 | 0.232201113 |
| sp\|Q8TAB3\|PCD19_HUMAN | 2 | 7 | 14 | 6 | -0.05745 | 0.065943373 | 0.444724059 | 0.123391836 | 0.502172522 | 0.378780686 | 0.794581026 | 0.091842349 | 0.163106347 |
| sp\|Q92619\|HMHA1_HUMAN | 9 | 29 | 55 | 25 | 0.031913 | 0.08511729 | 0.478210129 | 0.053204705 | 0.446297545 | 0.39309284 | 0.91984715 | 0.01919265 | 0.019695153 |
| sp\|O95425\|SVIL_HUMAN | 1 | 4 | 6 | 3 | 0.243317 | -0.226095433 | 1.218756274 | -0.469412821 | 0.975438885 | 1.444851707 | 0.786055073 | 0.493217023 | 0.194515679 |
| sp\|P55822\|SH3BG_HUMAN | 6 | 8 | 13 | 6 | -0.72666 | -1.103676684 | 0.510068638 | -0.377017995 | 1.236727327 | 1.613745322 | 0.812051649 | 0.231299955 | 0.060461466 |
| sp\|P09110\|THIK_HUMAN | 15 | 40 | 76 | 33 | -0.83751 | -0.665883193 | -0.152312728 | 0.171630225 | 0.68520069 | 0.513570465 | 0.496853652 | 0.000742053 | 0.005251133 |
| sp\|P11234\|RALB_HUMAN | 10 | 24 | 49 | 21 | -0.01782 | 0.06399847 | 0.42918095 | 0.081820157 | 0.447002637 | 0.36518248 | 0.762044664 | 0.005253106 | 0.009680949 |
| sp\|Q9UHY7\|ENOPH_HUMAN | 67 | 14 | 27 | 11 | -0.00921 | -0.083043997 | 0.278227283 | -0.07383641 | 0.28743487 | 0.36127128 | 0.898392171 | 0.346426846 | 0.125750878 |
| sp\|P67936\|TPM4_HUMAN | 21 | 40 | 76 | 33 | -0.15415 | -0.336857936 | 0.002000705 | -0.18270918 | 0.156149461 | 0.338858641 | 0.595671209 | 0.769665516 | 0.212750521 |
| sp\|Q14315\|FLNC_HUMAN | 8 | 40 | 76 | 33 | -0.14132 | -0.12250901 | 0.088207162 | 0.018810572 | 0.229526744 | 0.210716172 | 0.953870361 | 0.009776602 | 0.007101676 |
| sp\|P29122\|PCSK6_HUMAN | 2 | 40 | 76 | 33 | -0.23033 | -0.025988119 | 0.181342163 | 0.204345809 | 0.411676091 | 0.207330282 | 0.014715123 | 1.55E-05 | 0.021752334 |
| sp\|Q9HCK4\|ROBO2_HUMAN | 12 | 40 | 76 | 33 | 0.029215 | 0.063280607 | 0.052478082 | 0.034065503 | 0.023262978 | -0.010802525 | 0.764267313 | 0.916978794 | 0.976483166 |
| sp\|Q16787\|LAMA3_HUMAN | 8 | 40 | 76 | 33 | 0.206064 | 0.141867202 | 0.070038279 | -0.064197247 | -0.13602617 | -0.071828923 | 0.4362144 | 0.080066578 | 0.402089397 |
| sp\|P53041\|PPP5_HUMAN | 3 | 36 | 69 | 30 | -0.102 | 0.251524849 | 0.43397875 | 0.353521289 | 0.53597519 | 0.182453901 | 0.035700989 | 0.005549314 | 0.446258752 |
| sp\|Q96QK1\|VPS35_HUMAN | 2 | 40 | 76 | 33 | 0.163227 | 0.144359015 | 0.517396178 | -0.018867881 | 0.354169282 | 0.373037163 | 0.979837771 | 0.008935615 | 0.001440755 |
| sp\|P04424\|ARLY_HUMAN | 16 | 40 | 76 | 33 | -0.29236 | -0.020819213 | 0.40133875 | 0.271537154 | 0.693695117 | 0.422157963 | 0.015804415 | 5.79E-08 | 0.000215021 |
| sp\|Q9UQV4\|LAMP3_HUMAN | 21 | 3 | 7 | 4 | -0.41429 | -0.071669337 | 0.688816876 | 0.342621705 | 1.103107918 | 0.760486213 | 0.849827861 | 0.289202172 | 0.40468055 |
| sp\|Q15404\|RSU1_HUMAN | 13 | 40 | 76 | 33 | 0.054102 | -0.295608353 | -0.129337092 | -0.349710452 | -0.183439191 | 0.166271261 | 0.101455773 | 0.642180073 | 0.629447057 |
| sp\|Q12864\|CAD17_HUMAN | 1 | 40 | 76 | 33 | 0.175346 | 0.007949473 | -0.054942274 | -0.167396267 | -0.230288014 | -0.062891747 | 0.0632174 | 0.027990518 | 0.704031935 |
| sp\|P07476\|INVO_HUMAN | 10 | 40 | 76 | 33 | -0.13827 | -0.042886094 | -0.108959783 | 0.095380699 | 0.02930701 | -0.066073689 | 0.560873989 | 0.962822031 | 0.783091241 |
| sp\|Q14515\|SPRL1_HUMAN | 2 | 40 | 76 | 33 | 0.152845 | 0.058465176 | 0.204251255 | -0.094380012 | 0.051406066 | 0.145786079 | 0.190564314 | 0.078876748 | 0.033015411 |
| sp\|P61088\|UBE2N_HUMAN | 1 | 40 | 76 | 33 | -0.21621 | -0.104952424 | 0.233114272 | 0.111256158 | 0.449322854 | 0.338066696 | 0.537731506 | 0.001395246 | 0.008086088 |
| sp\|P34932\|HSP74_HUMAN | 8 | 40 | 76 | 33 | -0.07247 | 0.031725686 | 0.429626471 | 0.104196594 | 0.50209738 | 0.397900785 | 0.342908913 | 2.96E-07 | 4.54E-06 |
| sp\|P13798\|ACPH_HUMAN | 13 | 40 | 76 | 33 | -0.30688 | -0.120009267 | 0.194618224 | 0.186872763 | 0.501500254 | 0.314627492 | 0.056589861 | 2.31E-06 | 0.001037365 |
| sp\|Q04721\|NOTC2_HUMAN | 16 | 40 | 76 | 33 | 0.203477 | 0.191898868 | 0.061188209 | -0.011577701 | -0.14228836 | -0.130710659 | 0.895358924 | 3.01E-05 | 1.50E-05 |
| sp\|Q9BWS9\|CHID1_HUMAN | 18 | 40 | 76 | 33 | 0.013346 | 0.014819768 | 0.088626403 | 0.001474092 | 0.075280728 | 0.073806635 | 0.999818703 | 0.722425474 | 0.672076086 |
| sp\|Q9HCB6\|SPON1_HUMAN | 2 | 40 | 76 | 33 | 0.052612 | 0.012048705 | 0.311158027 | -0.040563643 | 0.258545679 | 0.299109322 | 0.660186967 | 2.64E-05 | 4.25E-08 |
| sp\|P41226\|UBA7_HUMAN | 327 | 40 | 76 | 33 | 0.158238 | 0.065313936 | 0.180000888 | -0.092924325 | 0.021762627 | 0.114686952 | 0.506924616 | 0.974441249 | 0.403966643 |
| sp\|Q7L1Q6\|BZW1_HUMAN | 3 | 24 | 49 | 22 | -0.50633 | -0.339841256 | 0.311644147 | 0.166485403 | 0.817970805 | 0.651485402 | 0.81310533 | 0.033437326 | 0.056544734 |
| sp\|P35241\|RADI_HUMAN | 35 | 40 | 76 | 33 | -0.43462 | -0.430102053 | -0.145295017 | 0.004522718 | 0.289329753 | 0.284807036 | 0.998531173 | 0.018636873 | 0.007691953 |
| sp\|P31327\|CPSM_HUMAN | 17 | 36 | 69 | 31 | -0.61333 | -0.599322572 | -0.090528676 | 0.014006133 | 0.52280003 | 0.508793896 | 0.994333808 | 0.005180446 | 0.001780978 |
| sp\|Q15365\|PCBP1_HUMAN | 160 | 40 | 76 | 33 | -0.17608 | -0.074071722 | 0.280286387 | 0.102007618 | 0.456365726 | 0.354358109 | 0.610402388 | 0.001627995 | 0.006775224 |
| sp\|O95373\|IPO7_HUMAN | 1396 | 22 | 42 | 17 | -0.09793 | -0.009602119 | 0.320215247 | 0.088328989 | 0.418146355 | 0.329817366 | 0.816841433 | 0.056049132 | 0.101834523 |
| sp\|P54577\|SYYC_HUMAN | 1 | 40 | 76 | 33 | 0.027577 | -0.138012767 | 0.091208695 | -0.16558978 | 0.063631682 | 0.229221462 | 0.368774392 | 0.902477981 | 0.188895065 |
| sp\|Q96G03\|PGM2_HUMAN | 2 | 40 | 76 | 33 | -0.17965 | -0.088781556 | 0.147860337 | 0.090865898 | 0.327507791 | 0.236641893 | 0.686203609 | 0.037811015 | 0.110813005 |
| sp\|P53396\|ACLY_HUMAN | 1 | 40 | 76 | 33 | -0.32305 | -0.088494737 | 0.269100612 | 0.234550605 | 0.592145954 | 0.357595349 | 0.024743861 | 4.15E-07 | 0.000682818 |
| sp\|P32754\|HPPD_HUMAN | 1 | 40 | 76 | 33 | -0.34233 | -0.075399014 | 0.288466543 | 0.266929545 | 0.630795102 | 0.363865557 | 0.139270789 | 0.000738233 | 0.041728892 |
| sp\|P42892\|ECE1_HUMAN | 1 | 40 | 76 | 33 | 0.221646 | 0.225251725 | 0.292499476 | 0.00360552 | 0.070853271 | 0.067247751 | 0.995851456 | 0.334570231 | 0.285655041 |
| sp\|P49189\|AL9A1_HUMAN | 3 | 40 | 76 | 33 | -0.16833 | 0.001670913 | 0.22205474 | 0.170000122 | 0.390383949 | 0.220383827 | 0.050791922 | 4.01E-05 | 0.013070996 |
| sp\|Q14517\|FAT1_HUMAN | 43 | 40 | 76 | 33 | -0.15107 | -0.167552717 | 0.197348205 | -0.016480322 | 0.348420601 | 0.364900923 | 0.962248381 | 2.28E-05 | 5.38E-07 |
| sp\|P55786\|PSA_HUMAN | 12 | 40 | 76 | 33 | -0.14233 | -0.067522893 | 0.226838498 | 0.074806436 | 0.369167828 | 0.294361391 | 0.627075501 | 0.000661838 | 0.002483991 |
| sp\|P08581\|MET_HUMAN | 5 | 40 | 76 | 33 | 0.049651 | 0.085345856 | 0.035295617 | 0.035694981 | -0.014355259 | -0.050050239 | 0.605971722 | 0.945381699 | 0.422581146 |
| sp\|P32004\|L1CAM_HUMAN | 1 | 40 | 76 | 33 | 0.185866 | 0.089838635 | -0.127188316 | -0.096027419 | -0.313054371 | -0.217026952 | 0.069245071 | 3.59E-08 | 1.59E-05 |
| sp\|P05388\|RLA0_HUMAN | 27 | 26 | 48 | 20 | -0.13905 | -0.03123449 | 0.357237361 | 0.107813005 | 0.496284856 | 0.388471851 | 0.559278961 | 0.000562317 | 0.002862651 |
| sp\|P09211\|GSTP1_HUMAN | 4 | 40 | 76 | 33 | -0.05107 | -0.064108521 | 0.156521707 | -0.013042777 | 0.207587452 | 0.220630229 | 0.988727807 | 0.143505167 | 0.06285721 |
| sp\|Q99435\|NELL2_HUMAN | 12 | 40 | 76 | 33 | 0.062831 | -0.000506861 | -0.054734127 | -0.063338163 | -0.117565429 | -0.054227266 | 0.613960468 | 0.316153133 | 0.730134927 |
| sp\|P09603\|CSF1_HUMAN | 1 | 40 | 76 | 33 | -0.301 | -0.144118008 | 0.234903946 | 0.156880571 | 0.535902526 | 0.379021954 | 0.107855585 | 1.37E-07 | 2.66E-05 |
| sp\|Q00610\|CLH1_HUMAN | 1 | 40 | 76 | 33 | -0.29533 | -0.212656385 | 0.219687056 | 0.082668936 | 0.515012377 | 0.432343442 | 0.772169735 | 0.001504365 | 0.002839457 |
| sp\|Q9BUJ0\|ABHEA_HUMAN | 1 | 7 | 14 | 7 | -0.01682 | -0.067542797 | -0.014785491 | -0.050719623 | 0.002037683 | 0.052757306 | 0.943966961 | 0.99993002 | 0.939528084 |
| sp\|Q9UKU6\|TRHDE_HUMAN | 11 | 40 | 76 | 33 | 0.144368 | 0.207204101 | 0.037725701 | 0.06283584 | -0.10664256 | -0.1694784 | 0.716497675 | 0.5167209 | 0.123648227 |
| sp\|Q8N392\|RHG18_HUMAN | 2 | 40 | 76 | 33 | 0.003341 | -0.0781812 | 0.099175919 | -0.081521798 | 0.095835321 | 0.177357119 | 0.813000694 | 0.820841367 | 0.425537592 |
| sp\|Q9NZT1\|CALL5_HUMAN | 3 | 32 | 61 | 25 | -1.81332 | -1.842715555 | -2.375624479 | -0.029393843 | -0.562302766 | -0.532908923 | 0.995696997 | 0.352871973 | 0.307264809 |
| sp\|P10768\|ESTD_HUMAN | 2 | 40 | 76 | 33 | -0.03104 | 0.094380244 | 0.396245757 | 0.125422764 | 0.427288277 | 0.301865513 | 0.237626504 | 2.66E-05 | 0.000988723 |
| sp\|P11279\|LAMP1_HUMAN | 19 | 40 | 76 | 33 | 0.480463 | 0.47415684 | 0.304790057 | -0.006305723 | -0.175672506 | -0.169366784 | 0.978229724 | 2.44E-05 | 4.25E-06 |
| sp\|Q9UNZ2\|NSF1C_HUMAN | 5 | 40 | 76 | 33 | -0.05891 | -0.026784738 | 0.463720563 | 0.032125813 | 0.522631114 | 0.490505301 | 0.9372765 | 2.34E-05 | 7.32E-06 |
| sp\|P31939\|PUR9_HUMAN | 11 | 40 | 76 | 33 | -0.15128 | -0.118870347 | 0.196780142 | 0.032404702 | 0.348055191 | 0.315650489 | 0.931069548 | 0.004589544 | 0.003618706 |
| sp\|P48147\|PPCE_HUMAN | 13 | 40 | 76 | 33 | -0.18641 | -0.129781778 | 0.199814396 | 0.056632541 | 0.386228715 | 0.329596174 | 0.741128335 | 0.000145866 | 0.000266275 |
| sp\|O75116\|ROCK2_HUMAN | 17 | 37 | 69 | 29 | -0.41723 | -0.156790382 | 0.062776786 | 0.260442757 | 0.480009925 | 0.219567168 | 0.276433628 | 0.055243187 | 0.45878278 |
| sp\|Q96EG1\|ARSG_HUMAN | 73 | 29 | 55 | 24 | 0.354634 | 0.241949898 | 0.308564388 | -0.11268435 | -0.04606986 | 0.066614489 | 0.37519307 | 0.891687233 | 0.737717964 |
| sp\|P0DMV8\|HS71A_HUMAN | 8 | 40 | 76 | 33 | -0.16789 | -0.092692615 | 0.309835214 | 0.075200772 | 0.477728601 | 0.402527829 | 0.717898662 | 0.000201992 | 0.000434764 |
| sp\|Q16270\|IBP7_HUMAN | 16 | 40 | 76 | 33 | 0.192242 | 0.088750596 | 0.25673847 | -0.103491658 | 0.064496216 | 0.167987874 | 0.098565565 | 0.531548539 | 0.005481188 |
| sp\|O75019\|LIRA1_HUMAN | 1 | 40 | 76 | 33 | -0.15284 | -0.126984028 | -0.110902594 | 0.025851386 | 0.04193282 | 0.016081434 | 0.955594333 | 0.920878941 | 0.984681245 |
| sp\|O43280\|TREA_HUMAN | 384 | 40 | 76 | 33 | 0.058318 | -0.007116317 | 0.022090461 | -0.065434471 | -0.036227694 | 0.029206777 | 0.824773538 | 0.960011101 | 0.966808208 |
| sp\|P04406\|G3P_HUMAN | 9 | 40 | 76 | 33 | -0.3608 | -0.073297389 | 0.139312173 | 0.287499638 | 0.5001092 | 0.212609561 | 0.04264793 | 0.001695214 | 0.21433787 |
| sp\|Q93088\|BHMT1_HUMAN | 24 | 40 | 76 | 33 | -0.08339 | 0.034062278 | 0.397010355 | 0.117447684 | 0.48039576 | 0.362948076 | 0.568857094 | 0.002126061 | 0.010667043 |
| sp\|P08648\|ITA5_HUMAN | 16 | 40 | 76 | 33 | 0.073364 | 0.105279994 | 0.12250843 | 0.031916041 | 0.049144476 | 0.017228435 | 0.475045008 | 0.297688769 | 0.825674957 |
| sp\|P16278\|BGAL_HUMAN | 18 | 40 | 76 | 33 | 0.337721 | 0.320857767 | 0.465719458 | -0.016863511 | 0.12799818 | 0.144861691 | 0.954979172 | 0.165702979 | 0.055203214 |
| sp\|O60610\|DIAP1_HUMAN | 20 | 40 | 76 | 33 | -0.51719 | -0.274752136 | 0.186682371 | 0.24244123 | 0.703875737 | 0.461434507 | 0.082391833 | 1.98E-06 | 0.000524117 |
| sp\|Q9UP79\|ATS8_HUMAN | 1 | 40 | 76 | 33 | 0.091618 | -0.030340283 | -0.385618772 | -0.121958745 | -0.477237234 | -0.355278489 | 0.122089209 | 5.41E-09 | 8.25E-07 |
| sp\|O43396\|TXNL1_HUMAN | 1 | 40 | 76 | 33 | -0.23663 | -0.216724662 | 0.185082291 | 0.019906549 | 0.421713502 | 0.401806953 | 0.980753329 | 0.003415118 | 0.001490585 |
| sp\|P29017\|CD1C_HUMAN | 9 | 40 | 76 | 33 | 0.093286 | 0.147193406 | -0.366126494 | 0.05390775 | -0.45941215 | -0.513319899 | 0.72789937 | 8.58E-07 | 8.16E-10 |
| sp\|Q99519\|NEUR1_HUMAN | 201 | 32 | 62 | 28 | 0.010008 | -0.012996129 | 0.13729922 | -0.02300391 | 0.127291439 | 0.150295349 | 0.954405281 | 0.368136638 | 0.168209086 |
| sp\|Q9HC56\|PCDH9_HUMAN | 13 | 40 | 76 | 33 | 0.11206 | 0.126033029 | 0.026808226 | 0.013972952 | -0.085251851 | -0.099224803 | 0.948476588 | 0.261789739 | 0.101513546 |
| sp\|Q9BXX0\|EMIL2_HUMAN | 8 | 29 | 55 | 25 | -0.28051 | -0.254496208 | 0.295970338 | 0.026012357 | 0.576478903 | 0.550466546 | 0.962229861 | 9.24E-06 | 1.72E-06 |
| sp\|P42025\|ACTY_HUMAN | 43 | 22 | 42 | 18 | -0.11528 | 0.168490075 | 0.483774653 | 0.28376954 | 0.599054118 | 0.315284578 | 0.141347822 | 0.003557035 | 0.122107474 |
| sp\|Q9BY76\|ANGL4_HUMAN | 59 | 40 | 76 | 33 | -0.16339 | -0.15057275 | 0.251985786 | 0.012812482 | 0.415371018 | 0.402558536 | 0.989080888 | 0.000616688 | 0.000162583 |
| sp\|P29350\|PTN6_HUMAN | 3 | 40 | 76 | 33 | -0.15981 | -0.179895806 | 0.16420159 | -0.020085431 | 0.324011965 | 0.344097395 | 0.984546267 | 0.065989436 | 0.021146523 |
| sp\|Q9UBW5\|BIN2_HUMAN | 34 | 40 | 76 | 33 | -0.40978 | -0.103057665 | 0.049437503 | 0.306721219 | 0.459216387 | 0.152495168 | 0.027704283 | 0.004317658 | 0.448864921 |
| sp\|P02461\|CO3A1_HUMAN | 10 | 40 | 76 | 33 | -0.04054 | 0.035708122 | 0.296049134 | 0.076251676 | 0.336592688 | 0.260341012 | 0.48457503 | 0.00012623 | 0.000945185 |
| sp\|P09467\|F16P1_HUMAN | 4 | 40 | 76 | 33 | -0.13608 | 0.061394184 | 0.383799798 | 0.19747395 | 0.519879564 | 0.322405614 | 0.184336936 | 0.000488184 | 0.020855737 |
| sp\|P20618\|PSB1_HUMAN | 2 | 40 | 76 | 33 | -0.25628 | -0.117460826 | 0.416688656 | 0.138817697 | 0.672967178 | 0.534149482 | 0.356846789 | 4.19E-07 | 5.86E-06 |
| sp\|Q09327\|MGAT3_HUMAN | 3 | 33 | 62 | 26 | -0.08435 | 0.036710474 | 0.037335233 | 0.121065264 | 0.121690024 | 0.000624759 | 0.542523691 | 0.658240476 | 0.999986044 |
| sp\|P06732\|KCRM_HUMAN | 6 | 40 | 76 | 33 | 0.180023 | -0.010653858 | -0.206851889 | -0.190677139 | -0.38687517 | -0.196198031 | 0.102096176 | 0.001967406 | 0.119485525 |
| sp\|Q13018\|PLA2R_HUMAN | 28 | 15 | 28 | 11 | 0.116519 | 0.035925728 | 0.01624802 | -0.080593492 | -0.100271201 | -0.019677709 | 0.536288851 | 0.534404605 | 0.970004176 |
| sp\|P23526\|SAHH_HUMAN | 2 | 40 | 76 | 33 | -0.34157 | -0.115345668 | 0.306762613 | 0.226225113 | 0.648333394 | 0.422108281 | 0.021489994 | 5.69E-09 | 1.72E-05 |
| sp\|O75369\|FLNB_HUMAN | 63 | 4 | 7 | 3 | 0.243222 | 0.337315741 | 0.135645384 | 0.094093953 | -0.107576404 | -0.201670357 | 0.94037498 | 0.947284875 | 0.795112248 |
| sp\|P13987\|CD59_HUMAN | 2 | 40 | 76 | 33 | -0.18384 | -0.16364271 | 0.287937978 | 0.020197958 | 0.471778646 | 0.451580687 | 0.967552228 | 1.45E-05 | 2.77E-06 |
| sp\|Q6ZMI3\|GLDN_HUMAN | 6 | 40 | 76 | 33 | -0.05835 | -0.151138117 | 0.193218241 | -0.092783029 | 0.251573149 | 0.344356358 | 0.301219831 | 0.002998789 | 2.33E-06 |
| sp\|Q9H2K8\|TAOK3_HUMAN | 14 | 26 | 48 | 20 | 0.220982 | 0.113802958 | 0.226832761 | -0.107179216 | 0.005850587 | 0.113029803 | 0.614159566 | 0.999019737 | 0.635030585 |
| sp\|P49247\|RPIA_HUMAN | 19 | 40 | 76 | 33 | -0.11327 | -0.061257496 | 0.321700842 | 0.052016013 | 0.434974351 | 0.382958338 | 0.718212598 | 8.21E-07 | 9.74E-07 |
| sp\|P52209\|6PGD_HUMAN | 19 | 40 | 76 | 33 | -0.10629 | -0.083510472 | 0.367198996 | 0.022779106 | 0.473488574 | 0.450709468 | 0.970463133 | 0.000265556 | 8.34E-05 |
| sp\|Q99439\|CNN2_HUMAN | 5 | 40 | 76 | 33 | -0.10979 | -0.076031499 | 0.166896265 | 0.03376312 | 0.276690884 | 0.242927764 | 0.953252761 | 0.114245758 | 0.118978532 |
| sp\|Q86VZ4\|LRP11_HUMAN | 30 | 40 | 76 | 33 | 0.047119 | 0.014138657 | 0.129338078 | -0.032980259 | 0.082219163 | 0.115199421 | 0.698081729 | 0.218182049 | 0.02427381 |
| sp\|Q99704\|DOK1_HUMAN | 4 | 13 | 28 | 13 | -0.04967 | 0.144295519 | 0.278770258 | 0.193965402 | 0.328440141 | 0.134474739 | 0.444579765 | 0.188639839 | 0.674713354 |
| sp\|Q9UKY3\|CES1P_HUMAN | 110 | 40 | 76 | 33 | -0.04115 | -0.14250131 | -0.000903654 | -0.101352519 | 0.040245137 | 0.141597656 | 0.589297405 | 0.943818312 | 0.405495396 |
| sp\|P23141\|EST1_HUMAN | 13 | 40 | 76 | 33 | 0.009625 | -0.052385076 | 0.135541358 | -0.062010372 | 0.125916063 | 0.187926434 | 0.784786639 | 0.503489023 | 0.14661965 |
| sp\|Q9UBG0\|MRC2_HUMAN | 2 | 40 | 76 | 33 | 0.128336 | 0.086519123 | 0.086126825 | -0.041816996 | -0.042209295 | -0.000392298 | 0.792734123 | 0.849250706 | 0.999982008 |
| sp\|Q9Y696\|CLIC4_HUMAN | 4 | 40 | 76 | 33 | -0.03251 | -0.093029585 | 0.115542781 | -0.060519216 | 0.14805315 | 0.208572366 | 0.828830629 | 0.462805027 | 0.146269996 |
| sp\|Q9NQR4\|NIT2_HUMAN | 2 | 40 | 76 | 33 | -0.08688 | -0.079190033 | 0.150820248 | 0.007693557 | 0.237703839 | 0.230010281 | 0.993679924 | 0.01820326 | 0.008752256 |

<table>
<tr><td></td><td></td><td></td><td></td><td></td><td></td><td></td><td></td><td></td><td></td><td></td><td></td><td></td></tr>
<tr><td>sp|P35609|ACTN2_HUMAN</td><td>44</td><td>40</td><td>76</td><td>33</td><td>-0.09847</td><td>-0.426420381</td><td>-0.159514432</td><td>-0.32795233</td><td>-0.061046381</td><td>0.266905949</td><td>0.152812557</td><td>0.955243152</td><td>0.332413192</td></tr>
<tr><td>sp|P06756|ITAV_HUMAN</td><td>19</td><td>40</td><td>76</td><td>33</td><td>0.19413</td><td>-0.020816767</td><td>-0.087079336</td><td>-0.214947158</td><td>-0.281209726</td><td>-0.066262568</td><td>7.55E-05</td><td>1.59E-05</td><td>0.422295817</td></tr>
<tr><td>sp|Q10469|MGAT2_HUMAN</td><td>162</td><td>40</td><td>76</td><td>33</td><td>0.085913</td><td>0.107556461</td><td>0.05290921</td><td>0.021643923</td><td>-0.033003328</td><td>-0.054647251</td><td>0.73613862</td><td>0.6123297</td><td>0.184577795</td></tr>
<tr><td>sp|Q8WWY7|WFD12_HUMAN</td><td>266</td><td>10</td><td>21</td><td>10</td><td>0.289357</td><td>-0.052992503</td><td>-0.095465769</td><td>-0.34234992</td><td>-0.384823186</td><td>-0.042473267</td><td>0.254119728</td><td>0.277746586</td><td>0.978275374</td></tr>
<tr><td>sp|Q9UJ70|NAGK_HUMAN</td><td>3</td><td>14</td><td>28</td><td>12</td><td>0.000514</td><td>0.023857021</td><td>0.003010835</td><td>0.023342951</td><td>0.002496764</td><td>-0.020846186</td><td>0.969019129</td><td>0.999750588</td><td>0.977659761</td></tr>
<tr><td>sp|B9A064|IGLL5_HUMAN</td><td>5</td><td>40</td><td>76</td><td>33</td><td>-0.42722</td><td>-0.257397779</td><td>0.178249943</td><td>0.169824472</td><td>0.605472193</td><td>0.435647722</td><td>0.266256099</td><td>2.48E-05</td><td>0.000739441</td></tr>
<tr><td>sp|O75340|PDCD6_HUMAN</td><td>8</td><td>40</td><td>76</td><td>33</td><td>-0.02012</td><td>0.006282005</td><td>0.057175612</td><td>0.026398544</td><td>0.077292151</td><td>0.050893607</td><td>0.774753536</td><td>0.22538508</td><td>0.437027251</td></tr>
<tr><td>sp|O95965|ITGBL_HUMAN</td><td>11</td><td>33</td><td>62</td><td>27</td><td>-0.0327</td><td>-0.081225937</td><td>0.210973</td><td>-0.048526448</td><td>0.243672489</td><td>0.292198937</td><td>0.873887715</td><td>0.101913165</td><td>0.017028137</td></tr>
<tr><td>sp|Q96KG7|MEG10_HUMAN</td><td>1</td><td>40</td><td>76</td><td>33</td><td>0.124991</td><td>0.049293815</td><td>-0.123138812</td><td>-0.075697079</td><td>-0.248129705</td><td>-0.172432627</td><td>0.30892606</td><td>0.000295015</td><td>0.005857603</td></tr>
<tr><td>sp|Q15149|PLEC_HUMAN</td><td>2</td><td>40</td><td>76</td><td>33</td><td>-0.68312</td><td>-0.61066582</td><td>0.076894138</td><td>0.072453639</td><td>0.760013596</td><td>0.687559958</td><td>0.870706464</td><td>6.95E-05</td><td>4.83E-05</td></tr>
<tr><td>sp|Q9NPG4|PCD12_HUMAN</td><td>57</td><td>40</td><td>76</td><td>33</td><td>0.06355</td><td>0.062622721</td><td>0.072046344</td><td>-0.000927532</td><td>0.008496091</td><td>0.009423623</td><td>0.999711814</td><td>0.983455355</td><td>0.974223718</td></tr>
<tr><td>sp|P16930|FAAA_HUMAN</td><td>47</td><td>40</td><td>76</td><td>33</td><td>-0.11809</td><td>0.025671093</td><td>0.254566351</td><td>0.143762413</td><td>0.372657671</td><td>0.228895258</td><td>0.176374602</td><td>0.000504916</td><td>0.022780879</td></tr>
<tr><td>sp|P50990|TCPQ_HUMAN</td><td>3</td><td>40</td><td>76</td><td>33</td><td>-0.28918</td><td>-0.169614614</td><td>0.262959798</td><td>0.119563005</td><td>0.552137417</td><td>0.432574412</td><td>0.497213573</td><td>8.00E-05</td><td>0.000552122</td></tr>
<tr><td>sp|P08572|CO4A2_HUMAN</td><td>16</td><td>40</td><td>76</td><td>33</td><td>-0.11298</td><td>-0.061217719</td><td>0.296989766</td><td>0.051765268</td><td>0.409972753</td><td>0.358207485</td><td>0.754837785</td><td>1.73E-05</td><td>2.30E-05</td></tr>
<tr><td>sp|O15230|LAMA5_HUMAN</td><td>1</td><td>40</td><td>76</td><td>33</td><td>-0.03641</td><td>-0.106388281</td><td>0.177101654</td><td>-0.069983057</td><td>0.213506878</td><td>0.283489935</td><td>0.417450861</td><td>0.004701566</td><td>1.15E-05</td></tr>
<tr><td>sp|P0DMN0|ST1A4_HUMAN</td><td>8</td><td>40</td><td>76</td><td>33</td><td>-0.39284</td><td>-0.315369312</td><td>-0.034962438</td><td>0.077466446</td><td>0.35787332</td><td>0.280406874</td><td>0.860615395</td><td>0.114863534</td><td>0.182913392</td></tr>
<tr><td>sp|O00461|GOLI4_HUMAN</td><td>12</td><td>40</td><td>76</td><td>33</td><td>-0.33831</td><td>-0.289730713</td><td>0.075703403</td><td>0.04857445</td><td>0.414008565</td><td>0.365434116</td><td>0.776747416</td><td>1.16E-05</td><td>1.27E-05</td></tr>
<tr><td>sp|P61201|CSN2_HUMAN</td><td>7</td><td>37</td><td>69</td><td>30</td><td>-0.45366</td><td>-0.207125083</td><td>0.140929045</td><td>0.246530773</td><td>0.594584901</td><td>0.348054128</td><td>0.037174834</td><td>5.82E-06</td><td>0.003844503</td></tr>
<tr><td>sp|Q93091|RNAS6_HUMAN</td><td>34</td><td>40</td><td>76</td><td>33</td><td>-0.17069</td><td>-0.168186136</td><td>0.335030028</td><td>0.002507759</td><td>0.505723924</td><td>0.503216165</td><td>0.99960778</td><td>4.65E-05</td><td>4.51E-06</td></tr>
<tr><td>sp|O95861|BPNT1_HUMAN</td><td>1</td><td>40</td><td>76</td><td>33</td><td>-0.18316</td><td>-0.082619777</td><td>0.317884241</td><td>0.100537851</td><td>0.50104187</td><td>0.400504019</td><td>0.405155559</td><td>1.12E-06</td><td>1.15E-05</td></tr>
<tr><td>sp|Q9H115|SNAB_HUMAN</td><td>1</td><td>11</td><td>21</td><td>9</td><td>0.048851</td><td>-0.268757088</td><td>0.524567015</td><td>-0.317608517</td><td>0.475715587</td><td>0.793324103</td><td>0.163313521</td><td>0.066336341</td><td>0.000287042</td></tr>
<tr><td>sp|Q93063|EXT2_HUMAN</td><td>1</td><td>40</td><td>76</td><td>33</td><td>0.121302</td><td>0.062253339</td><td>0.104936451</td><td>-0.059048386</td><td>-0.016365274</td><td>0.042683113</td><td>0.08516321</td><td>0.874782784</td><td>0.318029246</td></tr>
<tr><td>sp|P55072|TERA_HUMAN</td><td>3</td><td>40</td><td>76</td><td>33</td><td>-0.1952</td><td>-0.157750125</td><td>0.155914264</td><td>0.03745201</td><td>0.351116399</td><td>0.31366439</td><td>0.952753136</td><td>0.05783132</td><td>0.055402919</td></tr>
<tr><td>sp|P08473|NEP_HUMAN</td><td>11</td><td>40</td><td>76</td><td>33</td><td>0.14022</td><td>0.220770701</td><td>0.099011956</td><td>0.080550636</td><td>-0.041208109</td><td>-0.121758745</td><td>0.583446768</td><td>0.906871297</td><td>0.341379907</td></tr>
<tr><td>sp|P50502|F10A1_HUMAN</td><td>19</td><td>40</td><td>76</td><td>33</td><td>-0.25296</td><td>-0.148154623</td><td>0.222838226</td><td>0.104808832</td><td>0.475801681</td><td>0.370992849</td><td>0.479902629</td><td>6.97E-05</td><td>0.000528908</td></tr>
<tr><td>sp|Q9UJM8|HAOX1_HUMAN</td><td>28</td><td>8</td><td>14</td><td>6</td><td>-0.20215</td><td>-0.329537047</td><td>0.258876492</td><td>-0.127388659</td><td>0.461024881</td><td>0.588413539</td><td>0.854901461</td><td>0.268770359</td><td>0.083001561</td></tr>
<tr><td>sp|Q9H4A4|AMPB_HUMAN</td><td>4</td><td>40</td><td>76</td><td>33</td><td>-0.17052</td><td>0.032206884</td><td>0.087375042</td><td>0.202727478</td><td>0.257895637</td><td>0.055168158</td><td>0.002081016</td><td>0.001059042</td><td>0.653380372</td></tr>
<tr><td>sp|O00339|MATN2_HUMAN</td><td>4</td><td>40</td><td>76</td><td>33</td><td>0.10791</td><td>0.059344755</td><td>0.099630265</td><td>-0.048564856</td><td>-0.008279347</td><td>0.040285509</td><td>0.459919945</td><td>0.984420331</td><td>0.624523279</td></tr>
<tr><td>sp|Q8TDQ7|GNPI2_HUMAN</td><td>19</td><td>40</td><td>76</td><td>33</td><td>0.018426</td><td>0.015852527</td><td>0.100826106</td><td>-0.002573857</td><td>0.082399721</td><td>0.084973579</td><td>0.998329636</td><td>0.311101146</td><td>0.207349331</td></tr>
<tr><td>sp|P16949|STMN1_HUMAN</td><td>1</td><td>33</td><td>62</td><td>26</td><td>-0.35333</td><td>-0.251464989</td><td>0.370839517</td><td>0.101868805</td><td>0.724173311</td><td>0.622304506</td><td>0.64227784</td><td>2.04E-06</td><td>4.59E-06</td></tr>
<tr><td>sp|P29144|TPP2_HUMAN</td><td>54</td><td>36</td><td>69</td><td>31</td><td>0.591173</td><td>0.5443854</td><td>0.800854724</td><td>-0.046788065</td><td>0.209681259</td><td>0.256469325</td><td>0.857525311</td><td>0.119312746</td><td>0.018276234</td></tr>
<tr><td>sp|P48637|GSHB_HUMAN</td><td>12</td><td>40</td><td>76</td><td>33</td><td>0.019691</td><td>0.087446548</td><td>0.294691394</td><td>0.067755901</td><td>0.275000747</td><td>0.207244846</td><td>0.417145812</td><td>0.000105328</td><td>0.001149375</td></tr>
<tr><td>sp|O75223|GGCT_HUMAN</td><td>28</td><td>40</td><td>76</td><td>33</td><td>-0.29014</td><td>-0.091838364</td><td>0.314999062</td><td>0.198305157</td><td>0.605142583</td><td>0.406837426</td><td>0.128844464</td><td>6.30E-06</td><td>0.000749736</td></tr>
<tr><td>sp|P60520|GBRL2_HUMAN</td><td>2</td><td>20</td><td>42</td><td>19</td><td>-0.17764</td><td>-0.068524771</td><td>0.238723404</td><td>0.109116738</td><td>0.416364913</td><td>0.307248176</td><td>0.707790872</td><td>0.032070434</td><td>0.077975059</td></tr>
<tr><td>sp|P07339|CATD_HUMAN</td><td>27</td><td>40</td><td>76</td><td>33</td><td>-0.00077</td><td>-0.001751449</td><td>0.523766741</td><td>-0.000980613</td><td>0.524537576</td><td>0.525518189</td><td>0.999921007</td><td>1.15E-06</td><td>4.22E-08</td></tr>
<tr><td>sp|P07148|FABPL_HUMAN</td><td>35</td><td>40</td><td>76</td><td>33</td><td>-0.32992</td><td>-0.212345684</td><td>0.282341809</td><td>0.117571602</td><td>0.612259095</td><td>0.494687492</td><td>0.535616656</td><td>2.50E-05</td><td>0.000131319</td></tr>
<tr><td>sp|P21926|CD9_HUMAN</td><td>8</td><td>40</td><td>76</td><td>33</td><td>-0.62429</td><td>-0.028947247</td><td>0.057669375</td><td>0.595347096</td><td>0.681963717</td><td>0.086616622</td><td>0.000156721</td><td>0.000339162</td><td>0.837500372</td></tr>
<tr><td>sp|P23471|PTPRZ_HUMAN</td><td>12</td><td>40</td><td>76</td><td>33</td><td>0.161103</td><td>0.12780302</td><td>0.042846395</td><td>-0.033299801</td><td>-0.118256425</td><td>-0.084956624</td><td>0.591249175</td><td>0.012347133</td><td>0.053457274</td></tr>
<tr><td>sp|Q9Y646|CBPQ_HUMAN</td><td>33</td><td>40</td><td>76</td><td>33</td><td>0.09597</td><td>0.127063991</td><td>-0.034178569</td><td>0.031093879</td><td>-0.130148681</td><td>-0.16124256</td><td>0.830383501</td><td>0.11083565</td><td>0.014854183</td></tr>
<tr><td>sp|Q9BTY2|FUCO2_HUMAN</td><td>450</td><td>40</td><td>76</td><td>33</td><td>0.200762</td><td>0.141108777</td><td>0.307139857</td><td>-0.059653557</td><td>0.106377523</td><td>0.166031079</td><td>0.507506965</td><td>0.228619576</td><td>0.011818278</td></tr>
<tr><td>sp|A8MU46|SMTL1_HUMAN</td><td>3</td><td>4</td><td>6</td><td>3</td><td>0.005489</td><td>0.202174101</td><td>1.882342508</td><td>0.196685121</td><td>1.876853529</td><td>1.680168407</td><td>0.975558479</td><td>0.248607165</td><td>0.269138602</td></tr>
<tr><td>sp|Q9Y2A7|NCKP1_HUMAN</td><td>10</td><td>11</td><td>21</td><td>10</td><td>-0.52624</td><td>-0.207158459</td><td>0.134178389</td><td>0.319079113</td><td>0.66041596</td><td>0.341336847</td><td>0.38069723</td><td>0.058380668</td><td>0.35522514</td></tr>
<tr><td>sp|Q92520|FAM3C_HUMAN</td><td>4</td><td>40</td><td>76</td><td>33</td><td>0.012301</td><td>-0.009011359</td><td>0.22772502</td><td>-0.021312564</td><td>0.215423814</td><td>0.236736379</td><td>0.912006532</td><td>0.002226412</td><td>0.000109191</td></tr>
<tr><td>sp|O43242|PSMD3_HUMAN</td><td>18</td><td>33</td><td>62</td><td>27</td><td>-0.35219</td><td>-0.091813365</td><td>0.21776263</td><td>0.260373915</td><td>0.569949909</td><td>0.309575995</td><td>0.091671895</td><td>0.000580702</td><td>0.053328618</td></tr>
<tr><td>sp|Q9BT78|CSN4_HUMAN</td><td>24</td><td>11</td><td>21</td><td>9</td><td>-0.07531</td><td>0.055462471</td><td>0.116949968</td><td>0.130773508</td><td>0.192261005</td><td>0.061487497</td><td>0.56594157</td><td>0.433043485</td><td>0.894611043</td></tr>
<tr><td>sp|P07951|TPM2_HUMAN</td><td>3</td><td>40</td><td>76</td><td>33</td><td>0.016833</td><td>-0.213330542</td><td>0.21036964</td><td>-0.230163603</td><td>0.193536579</td><td>0.423700182</td><td>0.246102036</td><td>0.502009023</td><td>0.017260162</td></tr>
<tr><td>sp|P10253|LYAG_HUMAN</td><td>86</td><td>40</td><td>76</td><td>33</td><td>0.020955</td><td>0.061961673</td><td>0.182870371</td><td>0.041007163</td><td>0.16191586</td><td>0.120908697</td><td>0.686165791</td><td>0.020117797</td><td>0.060640887</td></tr>
<tr><td>sp|Q13642|FHL1_HUMAN</td><td>44</td><td>40</td><td>76</td><td>33</td><td>-0.10269</td><td>-0.128245675</td><td>0.025586639</td><td>-0.025552928</td><td>0.128279386</td><td>0.153832314</td><td>0.9836418</td><td>0.751282075</td><td>0.593336705</td></tr>
<tr><td>sp|Q9NYU2|UGGG1_HUMAN</td><td>18</td><td>40</td><td>76</td><td>33</td><td>-0.05899</td><td>-0.015703162</td><td>0.149449474</td><td>0.043282069</td><td>0.208434705</td><td>0.165152636</td><td>0.70471156</td><td>0.004881364</td><td>0.013633411</td></tr>
<tr><td>sp|O95490|AGRL2_HUMAN</td><td>12</td><td>7</td><td>14</td><td>6</td><td>0.071898</td><td>-0.014092531</td><td>0.022744782</td><td>-0.085990472</td><td>-0.049153159</td><td>0.036837313</td><td>0.25991128</td><td>0.726501411</td><td>0.791195542</td></tr>
<tr><td>sp|Q15102|PA1B3_HUMAN</td><td>3</td><td>40</td><td>76</td><td>33</td><td>-0.06728</td><td>0.059956752</td><td>0.30899342</td><td>0.127232148</td><td>0.376268816</td><td>0.249036668</td><td>0.244071894</td><td>0.000344069</td><td>0.010128812</td></tr>
<tr><td>sp|P04179|SODM_HUMAN</td><td>14</td><td>40</td><td>76</td><td>33</td><td>-0.03583</td><td>0.007099039</td><td>0.249726046</td><td>0.042933179</td><td>0.285560186</td><td>0.242627007</td><td>0.875715026</td><td>0.020325094</td><td>0.027602125</td></tr>
<tr><td>sp|Q13442|HAP28_HUMAN</td><td>6</td><td>26</td><td>48</td><td>20</td><td>-0.04503</td><td>-0.105300241</td><td>0.855101522</td><td>-0.060271856</td><td>0.900129908</td><td>0.960401764</td><td>0.946956153</td><td>0.00064624</td><td>4.25E-05</td></tr>
<tr><td>sp|P04080|CYTB_HUMAN</td><td>12</td><td>40</td><td>76</td><td>33</td><td>-0.52071</td><td>-0.323983058</td><td>0.434465697</td><td>0.196724453</td><td>0.955173208</td><td>0.758448755</td><td>0.168133372</td><td>4.56E-11</td><td>2.65E-09</td></tr>
<tr><td>sp|P02647|APOA1_HUMAN</td><td>48</td><td>40</td><td>76</td><td>33</td><td>-0.29901</td><td>-0.012075004</td><td>0.356317907</td><td>0.28693495</td><td>0.65532786</td><td>0.368392911</td><td>0.015033347</td><td>9.96E-07</td><td>0.00255451</td></tr>
<tr><td>sp|P15907|SIAT1_HUMAN</td><td>4</td><td>40</td><td>76</td><td>33</td><td>-0.11412</td><td>-0.023935516</td><td>0.205838075</td><td>0.090188019</td><td>0.31996161</td><td>0.229773591</td><td>0.210909379</td><td>5.00E-06</td><td>0.000248869</td></tr>
<tr><td>sp|Q86SF2|GALT7_HUMAN</td><td>9</td><td>40</td><td>76</td><td>33</td><td>0.10894</td><td>0.145744092</td><td>-0.028825527</td><td>0.036804531</td><td>-0.137765088</td><td>-0.174569619</td><td>0.458929726</td><td>0.000851138</td><td>1.23E-06</td></tr>
<tr><td>sp|P00367|DHE3_HUMAN</td><td>27</td><td>40</td><td>76</td><td>33</td><td>-0.07067</td><td>-0.037644892</td><td>0.226572513</td><td>0.033024555</td><td>0.29724196</td><td>0.264217405</td><td>0.963976082</td><td>0.134186672</td><td>0.132760411</td></tr>
<tr><td>sp|P07108|ACBP_HUMAN</td><td>12</td><td>40</td><td>76</td><td>33</td><td>-0.41886</td><td>-0.332222241</td><td>0.317705236</td><td>0.086639971</td><td>0.736567448</td><td>0.649927477</td><td>0.747316545</td><td>2.58E-06</td><td>2.87E-06</td></tr>
<tr><td>sp|O15511|ARPC5_HUMAN</td><td>22</td><td>40</td><td>76</td><td>33</td><td>0.010229</td><td>-0.061010852</td><td>0.290381415</td><td>-0.071239864</td><td>0.280152402</td><td>0.351392266</td><td>0.848349403</td><td>0.177968989</td><td>0.033480808</td></tr>
<tr><td>sp|P52888|THOP1_HUMAN</td><td>9</td><td>40</td><td>76</td><td>33</td><td>-0.06418</td><td>0.086949042</td><td>0.304298709</td><td>0.151128368</td><td>0.368478035</td><td>0.217349667</td><td>0.064604364</td><td>2.93E-05</td><td>0.007677479</td></tr>
<tr><td>sp|Q15382|RHEB_HUMAN</td><td>15</td><td>14</td><td>27</td><td>12</td><td>0.16666</td><td>-0.048727528</td><td>0.340337528</td><td>-0.215387399</td><td>0.173677657</td><td>0.389065056</td><td>0.419482451</td><td>0.670097997</td><td>0.085463169</td></tr>
<tr><td>sp|P46060|RAGP1_HUMAN</td><td>74</td><td>4</td><td>6</td><td>3</td><td>-0.03804</td><td>0.319981703</td><td>0.549037978</td><td>0.358018963</td><td>0.587075238</td><td>0.229056275</td><td>0.273059761</td><td>0.105597839</td><td>0.616931936</td></tr>
<tr><td>sp|Q8NBS9|TXND5_HUMAN</td><td>11</td><td>40</td><td>76</td><td>33</td><td>-0.41805</td><td>-0.354398195</td><td>0.495637057</td><td>0.063653071</td><td>0.913688324</td><td>0.850035253</td><td>0.822251848</td><td>1.42E-10</td><td>2.11E-11</td></tr>
<tr><td>sp|O75594|PGRP1_HUMAN</td><td>3</td><td>40</td><td>76</td><td>33</td><td>-0.02609</td><td>0.040662056</td><td>0.252454534</td><td>0.066749813</td><td>0.278542291</td><td>0.211792478</td><td>0.646431884</td><td>0.006722481</td><td>0.024062919</td></tr>
<tr><td>sp|Q9NQW7|XPP1_HUMAN</td><td>46</td><td>40</td><td>76</td><td>33</td><td>-0.11921</td><td>-0.20465483</td><td>0.142337115</td><td>-0.085444964</td><td>0.26154698</td><td>0.346991944</td><td>0.736560991</td><td>0.14328445</td><td>0.014256487</td></tr>
<tr><td>sp|Q9GZX9|TWSG1_HUMAN</td><td>4</td><td>40</td><td>76</td><td>33</td><td>0.121155</td><td>0.031391639</td><td>0.175155552</td><td>-0.089763549</td><td>0.054000365</td><td>0.143763913</td><td>0.21241074</td><td>0.67596579</td><td>0.032633808</td></tr>
<tr><td>sp|P49368|TCPG_HUMAN</td><td>52</td><td>40</td><td>76</td><td>33</td><td>-0.29872</td><td>-0.118215893</td><td>0.199508248</td><td>0.180502105</td><td>0.498226246</td><td>0.317724141</td><td>0.204088756</td><td>0.000388969</td><td>0.014861281</td></tr>
<tr><td>sp|P50120|RET2_HUMAN</td><td>2</td><td>28</td><td>56</td><td>24</td><td>-0.00533</td><td>-0.016941607</td><td>0.052987418</td><td>-0.011612488</td><td>0.058316537</td><td>0.069929026</td><td>0.994213754</td><td>0.903762374</td><td>0.82784601</td></tr>
<tr><td>sp|Q9NZ08|ERAP1_HUMAN</td><td>8</td><td>40</td><td>76</td><td>33</td><td>-0.04029</td><td>0.026059144</td><td>0.275557806</td><td>0.066351925</td><td>0.315850587</td><td>0.249498663</td><td>0.710533912</td><td>0.006162506</td><td>0.016882599</td></tr>
<tr><td>sp|P22061|PIMT_HUMAN</td><td>9</td><td>40</td><td>76</td><td>33</td><td>-0.1485</td><td>-0.011697997</td><td>0.315955397</td><td>0.136803249</td><td>0.464456642</td><td>0.327653394</td><td>0.122925595</td><td>3.73E-07</td><td>5.56E-05</td></tr>
<tr><td>sp|P26447|S10A4_HUMAN</td><td>42</td><td>40</td><td>76</td><td>33</td><td>-0.06978</td><td>0.054155033</td><td>0.24223408</td><td>0.12393306</td><td>0.312012107</td><td>0.188079047</td><td>0.376135533</td><td>0.01593105</td><td>0.141282169</td></tr>
<tr><td>sp|Q08623|HDHD1_HUMAN</td><td>1</td><td>7</td><td>14</td><td>6</td><td>-0.08392</td><td>0.10165342</td><td>0.437205556</td><td>0.1855763</td><td>0.521128436</td><td>0.335552136</td><td>0.715063221</td><td>0.179365388</td><td>0.383261917</td></tr>
<tr><td>sp|Q00796|DHSO_HUMAN</td><td>13</td><td>40</td><td>76</td><td>33</td><td>-0.37611</td><td>-0.0847308</td><td>0.234741077</td><td>0.291379008</td><td>0.610850885</td><td>0.319471877</td><td>0.041754148</td><td>0.00011217</td><td>0.035216212</td></tr>
<tr><td>sp|Q93045|STMN2_HUMAN</td><td>21</td><td>13</td><td>28</td><td>12</td><td>-0.00208</td><td>-0.016819324</td><td>0.619493188</td><td>-0.014737981</td><td>0.621574531</td><td>0.636312512</td><td>0.997177212</td><td>0.037991039</td><td>0.011283658</td></tr>
<tr><td>sp|P49354|FNTA_HUMAN</td><td>17</td><td>11</td><td>20</td><td>9</td><td>-0.13625</td><td>-0.19752196</td><td>0.172266646</td><td>-0.061273717</td><td>0.30851489</td><td>0.369788606</td><td>0.901814378</td><td>0.1759962</td><td>0.049326369</td></tr>
<tr><td>sp|Q9Y617|SERC_HUMAN</td><td>7</td><td>40</td><td>76</td><td>33</td><td>-0.19524</td><td>0.007789921</td><td>0.270149565</td><td>0.203025681</td><td>0.465385326</td><td>0.262359644</td><td>0.067470705</td><td>0.000103195</td><td>0.020147578</td></tr>
<tr><td>sp|P13797|PLST_HUMAN</td><td>7</td><td>40</td><td>76</td><td>33</td><td>0.091354</td><td>0.118464415</td><td>0.107564645</td><td>0.027110496</td><td>0.016210726</td><td>-0.01089977</td><td>0.72536868</td><td>0.923657749</td><td>0.955326924</td></tr>
<tr><td>sp|Q9NZR1|TMOD2_HUMAN</td><td>93</td><td>32</td><td>62</td><td>27</td><td>-0.67433</td><td>-0.292789853</td><td>0.080548603</td><td>0.381536635</td><td>0.754875092</td><td>0.373338456</td><td>0.145993323</td><td>0.006483687</td><td>0.192562305</td></tr>
<tr><td>sp|Q9H008|LHPP_HUMAN</td><td>14</td><td>8</td><td>14</td><td>6</td><td>-0.03869</td><td>0.139624571</td><td>0.092903592</td><td>0.178311134</td><td>0.131590155</td><td>-0.046720979</td><td>0.480866148</td><td>0.760306037</td><td>0.958149372</td></tr>
<tr><td>sp|P52848|NDST1_HUMAN</td><td>4</td><td>40</td><td>76</td><td>33</td><td>0.062463</td><td>0.16376741</td><td>0.079085368</td><td>0.101304062</td><td>0.01662202</td><td>-0.084682041</td><td>0.025203963</td><td>0.931472337</td><td>0.10122715</td></tr>
<tr><td>sp|Q9NQ36|SCUB2_HUMAN</td><td>128</td><td>29</td><td>55</td><td>24</td><td>-0.01771</td><td>0.021503788</td><td>0.057959915</td><td>0.03920952</td><td>0.075665647</td><td>0.036456127</td><td>0.908512169</td><td>0.781440798</td><td>0.929590578</td></tr>
<tr><td>sp|Q99574|NEUS_HUMAN</td><td>4</td><td>40</td><td>76</td><td>33</td><td>-0.06075</td><td>-0.048063286</td><td>0.267984914</td><td>0.01269117</td><td>0.32873937</td><td>0.3160482</td><td>0.991431792</td><td>0.02173768</td><td>0.011400455</td></tr>
<tr><td>sp|P25940|CO5A3_HUMAN</td><td>4</td><td>26</td><td>48</td><td>20</td><td>0.113957</td><td>0.166400564</td><td>0.0463723</td><td>0.05244308</td><td>-0.067585184</td><td>-0.120028264</td><td>0.901677028</td><td>0.891188642</td><td>0.636670535</td></tr>
<tr><td>sp|P26641|EF1G_HUMAN</td><td>13</td><td>40</td><td>76</td><td>33</td><td>-0.50936</td><td>-0.289480914</td><td>0.251297429</td><td>0.219879109</td><td>0.760657453</td><td>0.540778343</td><td>0.229015887</td><td>1.52E-05</td><td>0.000622893</td></tr>
<tr><td>sp|O95372|LYPA2_HUMAN</td><td>23</td><td>30</td><td>55</td><td>23</td><td>-0.24436</td><td>-0.134683983</td><td>0.103869424</td><td>0.109678526</td><td>0.348231933</td><td>0.238553407</td><td>0.640062689</td><td>0.053918838</td><td>0.176625331</td></tr>
<tr><td>sp|P08133|ANXA6_HUMAN</td><td>24</td><td>40</td><td>76</td><td>33</td><td>-0.55289</td><td>-0.235239943</td><td>0.150436808</td><td>0.317648405</td><td>0.703325156</td><td>0.385676751</td><td>0.039802029</td><td>3.72E-05</td><td>0.016009863</td></tr>
<tr><td>sp|Q7LDG7|GRP2_HUMAN</td><td>3</td><td>26</td><td>48</td><td>20</td><td>0.024718</td><td>-0.154865141</td><td>0.082949338</td><td>-0.179583012</td><td>0.058231467</td><td>0.237814479</td><td>0.551046</td><td>0.958537578</td><td>0.418414107</td></tr>
<tr><td>sp|O14818|PSA7_HUMAN</td><td>4</td><td>40</td><td>76</td><td>33</td><td>-0.30796</td><td>-0.107193527</td><td>0.504533323</td><td>0.200766165</td><td>0.812493015</td><td>0.61172685</td><td>0.208231016</td><td>1.75E-07</td><td>9.05E-06</td></tr>
<tr><td>sp|Q96MK3|FA20A_HUMAN</td><td>7</td><td>40</td><td>76</td><td>33</td><td>-0.05417</td><td>0.009090109</td><td>0.149514645</td><td>0.063262963</td><td>0.203687499</td><td>0.140424536</td><td>0.281510238</td><td>0.000214772</td><td>0.005167454</td></tr>
<tr><td>sp|Q96QR1|SG3A1_HUMAN</td><td>4</td><td>33</td><td>62</td><td>28</td><td>-0.06526</td><td>-0.041371238</td><td>0.241983385</td><td>0.023893118</td><td>0.307247741</td><td>0.283354622</td><td>0.949211571</td><td>0.003428046</td><td>0.002205299</td></tr>
<tr><td>sp|Q70J99|UN13D_HUMAN</td><td>4</td><td>40</td><td>76</td><td>33</td><td>0.116965</td><td>-0.03565023</td><td>0.300164957</td><td>-0.152615667</td><td>0.18319952</td><td>0.335815187</td><td>0.450644697</td><td>0.452705493</td><td>0.036828241</td></tr>
<tr><td>sp|P08758|ANXA5_HUMAN</td><td>2</td><td>40</td><td>76</td><td>33</td><td>-0.3783</td><td>-0.323756095</td><td>0.109804245</td><td>0.054541303</td><td>0.488101643</td><td>0.43356034</td><td>0.917474474</td><td>0.010615531</td><td>0.010433019</td></tr>
<tr><td>sp|Q15043|S39AE_HUMAN</td><td>10</td><td>40</td><td>76</td><td>33</td><td>-0.03266</td><td>-0.000596582</td><td>-0.094779252</td><td>0.03206117</td><td>-0.062121499</td><td>-0.094182669</td><td>0.831800544</td><td>0.621533254</td><td>0.252140514</td></tr>
<tr><td>sp|Q8IV08|PLD3_HUMAN</td><td>1</td><td>37</td><td>69</td><td>30</td><td>0.014164</td><td>0.06167774</td><td>0.247198276</td><td>0.047514216</td><td>0.233034752</td><td>0.185520536</td><td>0.503555537</td><td>3.47E-05</td><td>0.000231855</td></tr>
<tr><td>sp|P12429|ANXA3_HUMAN</td><td>6</td><td>40</td><td>76</td><td>33</td><td>-0.56441</td><td>-0.461714334</td><td>0.028618823</td><td>0.10270031</td><td>0.593033468</td><td>0.490333158</td><td>0.746568353</td><td>0.001791085</td><td>0.0039113</td></tr>
<tr><td>sp|P21549|SPYA_HUMAN</td><td>20</td><td>40</td><td>76</td><td>33</td><td>-0.33953</td><td>-0.221831966</td><td>0.1355439</td><td>0.117702546</td><td>0.475078413</td><td>0.357375866</td><td>0.56070516</td><td>0.00214065</td><td>0.011131827</td></tr>
<tr><td>sp|O15467|CCL16_HUMAN</td><td>15</td><td>40</td><td>76</td><td>33</td><td>-0.12155</td><td>-0.078616964</td><td>0.293960148</td><td>0.042933646</td><td>0.415510758</td><td>0.372577113</td><td>0.724771237</td><td>2.08E-08</td><td>1.46E-08</td></tr>
<tr><td>sp|Q96RD9|FCRL5_HUMAN</td><td>11</td><td>40</td><td>76</td><td>33</td><td>-0.04775</td><td>-0.021407576</td><td>0.180873277</td><td>0.026342978</td><td>0.228623831</td><td>0.202280853</td><td>0.944269555</td><td>0.055413888</td><td>0.056019646</td></tr>
<tr><td>sp|Q12955|ANK3_HUMAN</td><td>28</td><td>4</td><td>7</td><td>3</td><td>-0.14271</td><td>-0.141115913</td><td>1.075314516</td><td>0.001595662</td><td>1.21802609</td><td>1.216430428</td><td>0.99999699</td><td>0.344398275</td><td>0.279164132</td></tr>
<tr><td>sp|Q08ET2|SIG14_HUMAN</td><td>21</td><td>33</td><td>61</td><td>27</td><td>-0.64189</td><td>-0.225670917</td><td>-0.696001677</td><td>0.416215918</td><td>-0.054114842</td><td>-0.47033076</td><td>0.517441228</td><td>0.99224484</td><td>0.479694635</td></tr>
<tr><td>sp|P30044|PRDX5_HUMAN</td><td>1</td><td>40</td><td>76</td><td>33</td><td>-0.31445</td><td>-0.236694704</td><td>0.063155242</td><td>0.077759916</td><td>0.377609862</td><td>0.299849946</td><td>0.717983945</td><td>0.005905062</td><td>0.015645018</td></tr>
<tr><td>sp|Q9NYQ8|FAT2_HUMAN</td><td>9</td><td>8</td><td>14</td><td>6</td><td>-0.12859</td><td>0.136530348</td><td>0.2724139</td><td>0.265125285</td><td>0.401008837</td><td>0.135883552</td><td>0.525237802</td><td>0.376229295</td><td>0.867043913</td></tr>
<tr><td>sp|P27816|MAP4_HUMAN</td><td>15</td><td>14</td><td>27</td><td>13</td><td>-0.11174</td><td>-0.010363011</td><td>0.470912272</td><td>0.101374114</td><td>0.582649397</td><td>0.481275284</td><td>0.799371438</td><td>0.007771963</td><td>0.012735775</td></tr>
<tr><td>sp|O43488|ARK72_HUMAN</td><td>17</td><td>36</td><td>70</td><td>30</td><td>-0.244</td><td>-0.205852239</td><td>0.025645776</td><td>0.038150936</td><td>0.269648951</td><td>0.231498015</td><td>0.93386591</td><td>0.1007823</td><td>0.11377865</td></tr>
</table>

| | | | | | | | | | | | | | |
|---|---|---|---|---|---|---|---|---|---|---|---|---|---|
| sp\|P40227\|TCPZ_HUMAN | 5 | 34 | 62 | 26 | -0.16859 | 0.038327204 | 0.483995585 | 0.206918081 | 0.652586463 | 0.445668381 | 0.428658538 | 0.00469298 | 0.041256659 |
| sp\|O00429\|DNM1L_HUMAN | 7 | 40 | 76 | 33 | -0.12049 | -0.172865603 | 0.080584933 | -0.052374711 | 0.201075825 | 0.253450535 | 0.905481915 | 0.367840205 | 0.135221217 |
| sp\|O00468\|AGRIN_HUMAN | 28 | 40 | 76 | 33 | -0.2517 | -0.122949499 | 0.3056017 | 0.128751115 | 0.557302314 | 0.428551199 | 0.116288379 | 9.62E-11 | 1.40E-08 |
| sp\|Q8IZP9\|AGRG2_HUMAN | 3 | 40 | 76 | 33 | 0.1653 | 0.101094477 | -0.12465334 | -0.064205183 | -0.289953001 | -0.225747817 | 0.311410855 | 5.17E-07 | 1.05E-05 |
| sp\|P54764\|EPHA4_HUMAN | 3 | 40 | 76 | 33 | 0.046674 | -0.032868684 | -0.179679924 | -0.079542691 | -0.226353931 | -0.146811241 | 0.393792713 | 0.006767675 | 0.064862415 |
| sp\|P98095\|FBLN2_HUMAN | 7 | 40 | 76 | 33 | 0.089558 | 0.056672562 | 0.396269519 | -0.032885797 | 0.306711159 | 0.339596957 | 0.829911274 | 3.93E-05 | 2.65E-07 |
| sp\|P30626\|SORCN_HUMAN | 9 | 36 | 69 | 30 | -0.11856 | -0.011692335 | 0.121110607 | 0.106864436 | 0.239667377 | 0.132802942 | 0.28018823 | 0.013804766 | 0.177674466 |
| sp\|Q13310\|PABP4_HUMAN | 18 | 23 | 41 | 18 | -0.07995 | -0.027345643 | 0.19214499 | 0.052605192 | 0.272095824 | 0.219490632 | 0.898968587 | 0.150127127 | 0.214920063 |
| sp\|P15259\|PGAM2_HUMAN | 2 | 40 | 76 | 33 | -0.06827 | 0.020353066 | 0.208435766 | 0.088623901 | 0.276706601 | 0.1880827 | 0.417881773 | 0.003719202 | 0.034644959 |
| sp\|O75326\|SEM7A_HUMAN | 4 | 40 | 76 | 33 | -0.0006 | 0.024316684 | 0.190044393 | 0.024912942 | 0.190640651 | 0.165727709 | 0.812470572 | 0.000419075 | 0.000553797 |
| sp\|P42785\|PCP_HUMAN | 6 | 40 | 76 | 33 | 0.012607 | 0.050361138 | 0.219722885 | 0.037753649 | 0.207115397 | 0.169361748 | 0.651980358 | 0.000269647 | 0.000851289 |
| sp\|P11597\|CETP_HUMAN | 3 | 40 | 76 | 33 | 0.291564 | 0.260833186 | -0.242505259 | -0.030730921 | -0.534069366 | -0.503338445 | 0.943022442 | 1.69E-05 | 4.67E-06 |
| sp\|Q68BL8\|OLM2B_HUMAN | 50 | 3 | 6 | 3 | -0.38182 | -0.544501866 | 0.691297093 | -0.162678344 | 1.073120614 | 1.235798958 | 0.935942624 | 0.170989376 | 0.063116169 |
| sp\|Q86U17\|SPA11_HUMAN | 4 | 40 | 76 | 33 | 0.094263 | 0.096942429 | 0.315637702 | 0.002679897 | 0.22137517 | 0.218695273 | 0.998813427 | 0.005019306 | 0.001505408 |
| sp\|O43390\|HNRPR_HUMAN | 1 | 29 | 55 | 24 | 0.100251 | 0.032631812 | 0.340007837 | -0.067619487 | 0.239756538 | 0.307376025 | 0.827774005 | 0.19935564 | 0.036901199 |
| sp\|Q9H9K5\|MER34_HUMAN | 11 | 40 | 76 | 33 | -0.07202 | -0.093766047 | -0.07294994 | -0.021746963 | -0.000930856 | 0.020816107 | 0.970173113 | 0.999961708 | 0.975934076 |
| sp\|O94923\|GLCE_HUMAN | 1 | 40 | 76 | 33 | -0.12255 | -0.007140733 | 0.036334079 | 0.11541122 | 0.158886033 | 0.043474812 | 0.206919327 | 0.128729507 | 0.81942481 |
| sp\|Q15369\|ELOC_HUMAN | 1 | 13 | 28 | 12 | -0.18299 | -0.069459759 | 0.496090685 | 0.113528224 | 0.679078668 | 0.565550444 | 0.784114514 | 0.004452609 | 0.006138884 |
| sp\|P48059\|LIMS1_HUMAN | 20 | 40 | 76 | 33 | -0.01136 | -0.148553371 | 0.013734528 | -0.137193103 | 0.025094797 | 0.1622879 | 0.685756216 | 0.991295105 | 0.629376073 |
| sp\|Q16851\|UGPA_HUMAN | 4 | 40 | 76 | 33 | 0.07271 | 0.212146026 | 0.66167452 | 0.139436519 | 0.588965012 | 0.449528494 | 0.416168118 | 5.12E-05 | 0.000559526 |
| sp\|P49006\|MRP_HUMAN | 2 | 10 | 21 | 10 | -0.66255 | -0.250249276 | 0.600870351 | 0.412305393 | 1.26342502 | 0.851119627 | 0.355806408 | 0.002149191 | 0.017873163 |
| sp\|P00491\|PNPH_HUMAN | 25 | 40 | 76 | 33 | -0.12577 | -0.099404057 | 0.193760586 | 0.026363285 | 0.319527928 | 0.293164643 | 0.963595598 | 0.026754363 | 0.020877645 |
| sp\|P40121\|CAPG_HUMAN | 7 | 40 | 76 | 33 | -0.19873 | -0.034773115 | 0.308611118 | 0.163960627 | 0.50734486 | 0.343384233 | 0.099291514 | 1.11E-06 | 0.000229151 |
| sp\|P21695\|GPDA_HUMAN | 3 | 40 | 76 | 33 | 0.093292 | 0.114783763 | 0.135897289 | 0.021491813 | 0.042605339 | 0.021113526 | 0.943617004 | 0.854535398 | 0.95200944 |
| sp\|Q9Y4C0\|NRX3A_HUMAN | 3 | 40 | 76 | 33 | 0.19888 | 0.064092504 | -0.108771128 | -0.134787977 | -0.30765161 | -0.172863633 | 0.008448457 | 1.76E-07 | 0.00115705 |
| sp\|Q6V0I7\|FAT4_HUMAN | 17 | 40 | 76 | 33 | 0.011107 | -0.005523844 | 0.02986644 | -0.016630996 | 0.018759288 | 0.035390285 | 0.837540587 | 0.855841388 | 0.496130247 |
| sp\|P05186\|PPBT_HUMAN | 2 | 40 | 76 | 33 | 0.009176 | 0.054222123 | 0.299258201 | 0.045046371 | 0.290082448 | 0.245036077 | 0.739982208 | 0.000342512 | 0.000681072 |
| sp\|P25445\|TNR6_HUMAN | 3 | 40 | 76 | 33 | 0.024861 | 0.036219295 | 0.406860959 | 0.011358459 | 0.382000123 | 0.370641664 | 0.980350717 | 1.25E-06 | 1.18E-07 |
| sp\|P58166\|INHBE_HUMAN | 29 | 40 | 76 | 33 | -0.12728 | -0.054671731 | 0.086906784 | 0.072603801 | 0.214182316 | 0.141578514 | 0.563171133 | 0.035094251 | 0.150987193 |
| sp\|P29323\|EPHB2_HUMAN | 1 | 40 | 76 | 33 | -0.15002 | -0.084396267 | -0.043966038 | 0.065626974 | 0.106057202 | 0.040430228 | 0.264397888 | 0.09335431 | 0.639259864 |
| sp\|Q02818\|NUCB1_HUMAN | 9 | 40 | 76 | 33 | -0.17067 | -0.066582577 | 0.490714227 | 0.104082612 | 0.661379416 | 0.557296804 | 0.546181296 | 3.98E-07 | 1.42E-06 |
| sp\|Q8WWV6\|FCAMR_HUMAN | 24 | 36 | 69 | 30 | -0.30305 | -0.1658975 | 0.185717709 | 0.137154115 | 0.488769323 | 0.351615208 | 0.322830388 | 0.000108675 | 0.001996284 |
| sp\|P40189\|IL6RB_HUMAN | 77 | 40 | 76 | 33 | 0.415713 | 0.413234574 | 0.450129958 | -0.002478001 | 0.034417383 | 0.036895384 | 0.99608293 | 0.594801979 | 0.468702674 |
| sp\|Q9UBQ6\|EXTL2_HUMAN | 3 | 40 | 76 | 33 | 0.219135 | 0.116564195 | -0.111923188 | -0.102570957 | -0.33105834 | -0.228487383 | 0.063441634 | 2.95E-08 | 1.52E-05 |
| sp\|P15311\|EZRI_HUMAN | 8 | 40 | 76 | 33 | 0.038952 | 0.071725209 | 0.270827295 | 0.032773632 | 0.231875719 | 0.199102086 | 0.871878125 | 0.010826891 | 0.014159942 |
| sp\|Q9Y5Z4\|HEBP2_HUMAN | 2 | 36 | 69 | 30 | -0.3635 | -0.343289674 | 0.185972901 | 0.02021245 | 0.549475024 | 0.529262575 | 0.985601143 | 0.001026424 | 0.000315628 |
| sp\|Q7Z7M9\|GALT5_HUMAN | 23 | 40 | 76 | 33 | 0.156736 | 0.064384129 | 0.038335725 | -0.092351483 | -0.118399887 | -0.026048404 | 0.263431977 | 0.220898851 | 0.909910339 |
| sp\|P16284\|PECA1_HUMAN | 2 | 40 | 76 | 33 | 0.064787 | 0.028594324 | 0.09911359 | -0.036192888 | 0.034326379 | 0.070519267 | 0.611502856 | 0.736487762 | 0.197905242 |
| sp\|Q06323\|PSME1_HUMAN | 4 | 40 | 76 | 33 | -0.24354 | -0.026656258 | 0.397959588 | 0.216884453 | 0.6415003 | 0.424615847 | 0.058898289 | 2.42E-07 | 0.00012607 |
| sp\|P00966\|ASSY_HUMAN | 143 | 32 | 62 | 28 | 0.167094 | 0.395494665 | 0.519292962 | 0.228400698 | 0.352198995 | 0.123798296 | 0.169757009 | 0.052739063 | 0.616545416 |
| sp\|P62191\|PRS4_HUMAN | 52 | 7 | 14 | 5 | 0.161759 | 0.164071119 | 0.537069751 | 0.002312027 | 0.375310659 | 0.372998633 | 0.99985174 | 0.110986077 | 0.06856326 |
| sp\|P07384\|CAN1_HUMAN | 1 | 40 | 76 | 33 | -0.04758 | -0.031381414 | 0.335256963 | 0.016198772 | 0.382837149 | 0.366638376 | 0.980282858 | 0.000791066 | 0.000261509 |
| sp\|Q11201\|SIA4A_HUMAN | 1 | 40 | 76 | 33 | 0.003834 | -0.031587704 | -0.022430451 | -0.035421322 | -0.026264069 | 0.009157254 | 0.59100741 | 0.818566719 | 0.96945156 |
| sp\|P49593\|PPM1F_HUMAN | 1 | 36 | 69 | 31 | 0.02806 | -0.007809086 | 0.126108097 | -0.035869555 | 0.098047629 | 0.133917184 | 0.83149905 | 0.382034517 | 0.103120414 |
| sp\|P47914\|RL29_HUMAN | 5 | 4 | 7 | 2 | -0.43034 | -0.109143119 | -0.251347951 | 0.321192958 | 0.178988126 | -0.142204832 | 0.479992483 | 0.880478467 | 0.910222532 |
| sp\|Q9HC84\|MUC5B_HUMAN | 3 | 40 | 76 | 33 | -0.15563 | -0.420972686 | -0.099118579 | -0.265344917 | 0.056509189 | 0.321854106 | 0.195232379 | 0.949345135 | 0.122461152 |
| sp\|P30049\|ATPD_HUMAN | 4 | 4 | 6 | 3 | 0.075098 | -0.175264566 | 0.70643548 | -0.250362276 | 0.63133777 | 0.881700046 | 0.861900524 | 0.52683393 | 0.258836782 |
| sp\|Q3KQU3\|MA7D1_HUMAN | 10 | 4 | 6 | 3 | -0.17913 | 0.043296157 | 1.245130275 | 0.222422095 | 1.424256213 | 1.201834118 | 0.949457095 | 0.265351138 | 0.324280735 |
| sp\|Q9Y2Q5\|LTOR2_HUMAN | 43 | 7 | 14 | 6 | 0.036403 | 0.251615787 | 0.56050311 | 0.215212898 | 0.524100221 | 0.308887323 | 0.308666334 | 0.015035713 | 0.123735689 |
| sp\|Q99426\|TBCB_HUMAN | 41 | 40 | 76 | 33 | -0.28789 | -0.255049674 | 0.256058937 | 0.032841904 | 0.543950515 | 0.511108611 | 0.971331616 | 0.005407908 | 0.002939302 |
| sp\|P98161\|PKD1_HUMAN | 8 | 40 | 76 | 33 | 0.007513 | -0.007896818 | -0.03889864 | -0.015409359 | -0.046411181 | -0.031001822 | 0.895061523 | 0.501761289 | 0.675166167 |
| sp\|Q12797\|ASPH_HUMAN | 8 | 40 | 76 | 33 | 0.06408 | 0.044529312 | 0.115735359 | -0.019550377 | 0.05165567 | 0.071206047 | 0.839870347 | 0.434010659 | 0.13616992 |
| sp\|P55084\|ECHB_HUMAN | 9 | 4 | 7 | 3 | 0.190986 | 0.083987679 | 1.136182537 | -0.1069984 | 0.945196458 | 1.052194858 | 0.96683354 | 0.215892713 | 0.112787241 |
| sp\|P08582\|TRFM_HUMAN | 74 | 40 | 76 | 33 | 0.186066 | 0.12559503 | -0.081956928 | -0.060470881 | -0.268022838 | -0.207551958 | 0.620546264 | 0.002223942 | 0.008874367 |
| sp\|P32019\|I5P2_HUMAN | 9 | 4 | 7 | 3 | 0.054315 | 0.115351947 | 0.495376334 | 0.061037435 | 0.441061821 | 0.380024386 | 0.826391819 | 0.012262772 | 0.016226255 |
| sp\|Q9NWV4\|CZIB_HUMAN | 5 | 40 | 76 | 33 | -0.35864 | -0.243416817 | 0.055844732 | 0.115222619 | 0.414484168 | 0.299261549 | 0.544528138 | 0.005631433 | 0.03038409 |
| sp\|Q8WWI1\|LMO7_HUMAN | 3 | 4 | 6 | 3 | -0.03253 | -0.00327693 | 0.789274667 | 0.029257982 | 0.82180958 | 0.792551598 | 0.998097412 | 0.379490593 | 0.352376624 |
| sp\|P21709\|EPHA1_HUMAN | 10 | 40 | 76 | 33 | -0.03018 | -0.009996048 | 0.279514879 | 0.02018717 | 0.309698097 | 0.289510926 | 0.879516409 | 2.08E-08 | 3.78E-09 |
| sp\|P62491\|RB11A_HUMAN | 497 | 40 | 76 | 33 | -0.43209 | -0.326782109 | 0.243005644 | 0.105307401 | 0.675095154 | 0.569787753 | 0.715706767 | 0.000162335 | 0.000347145 |
| sp\|P15289\|ARSA_HUMAN | 589 | 40 | 76 | 33 | 0.097949 | 0.098612848 | 0.242961098 | 0.000663628 | 0.145011879 | 0.144348251 | 0.999950026 | 0.198204785 | 0.131089321 |
| sp\|P54578\|UBP14_HUMAN | 6 | 40 | 76 | 33 | -0.12678 | -0.051051526 | 0.276607529 | 0.075723665 | 0.403382719 | 0.327659055 | 0.580304329 | 5.92E-05 | 0.000252865 |
| sp\|Q92734\|TFG_HUMAN | 5 | 4 | 7 | 3 | -0.08724 | 0.055128555 | 0.637784859 | 0.142369645 | 0.72502595 | 0.582656304 | 0.892856454 | 0.183193592 | 0.250411994 |
| sp\|Q9H3S1\|SEM4A_HUMAN | 41 | 40 | 76 | 33 | -0.07658 | -0.01603116 | 0.063473901 | 0.060548606 | 0.140053668 | 0.079505061 | 0.553031013 | 0.116342098 | 0.409090025 |
| sp\|Q6Q788\|APOA5_HUMAN | 18 | 40 | 76 | 33 | -0.00753 | -0.026630387 | 0.017907176 | -0.019104469 | 0.025433094 | 0.044537563 | 0.962416696 | 0.954240666 | 0.833177196 |
| sp\|P02545\|LMNA_HUMAN | 42 | 40 | 76 | 33 | -0.35142 | -0.264957001 | 0.403737699 | 0.08646102 | 0.75515572 | 0.6686947 | 0.646980226 | 4.33E-09 | 4.49E-09 |
| sp\|O75339\|CILP1_HUMAN | 5 | 40 | 76 | 33 | 0.030247 | 0.031004822 | 0.223262059 | 0.000758298 | 0.193015535 | 0.192257237 | 0.999915522 | 0.026349197 | 0.010576342 |
| sp\|P49773\|HINT1_HUMAN | 4 | 40 | 76 | 33 | -0.37109 | -0.155821992 | 0.222565956 | 0.215265439 | 0.593653387 | 0.378387949 | 0.085959488 | 7.64E-06 | 0.001660531 |
| sp\|P41250\|GARS_HUMAN | 12 | 40 | 76 | 33 | -0.35134 | -0.191809041 | 0.174967266 | 0.159531255 | 0.526307562 | 0.366776307 | 0.46554114 | 0.004198721 | 0.03171994 |
| sp\|Q7Z6M3\|MILR1_HUMAN | 23 | 22 | 41 | 18 | -0.12239 | 0.010494837 | 0.210402145 | 0.132882107 | 0.332789414 | 0.199907308 | 0.482865843 | 0.047843941 | 0.240987578 |
| sp\|P78386\|KRT85_HUMAN | 4 | 8 | 13 | 6 | -0.37485 | 0.435416434 | -0.803161042 | 0.810264281 | -0.428313195 | -1.238577476 | 0.346876125 | 0.80821564 | 0.13988317 |
| sp\|P13647\|K2C5_HUMAN | 1 | 37 | 69 | 30 | -0.10554 | -0.211995163 | 0.059407302 | -0.106456504 | 0.164945962 | 0.271402466 | 0.795500974 | 0.685801049 | 0.279217594 |
| sp\|P61163\|ACTZ_HUMAN | 4 | 29 | 55 | 24 | -0.0891 | 0.112858945 | 0.255451787 | 0.20195511 | 0.344547953 | 0.142592843 | 0.128477198 | 0.017828011 | 0.4010125 |
| sp\|Q9HBI1\|PARVB_HUMAN | 8 | 40 | 76 | 33 | 0.012548 | -0.180728307 | 0.001085328 | -0.193276367 | -0.011462732 | 0.181813634 | 0.491657972 | 0.998265392 | 0.575497304 |
| sp\|Q9Y6N7\|ROBO1_HUMAN | 1 | 40 | 76 | 33 | 0.027007 | 0.048615866 | 0.019474461 | 0.021609094 | -0.00753231 | -0.029141405 | 0.819303259 | 0.983398486 | 0.727727803 |
| sp\|P34059\|GALNS_HUMAN | 4 | 40 | 76 | 33 | 0.069049 | -0.030370743 | 0.127291302 | -0.099419952 | 0.058242094 | 0.157662045 | 0.16862705 | 0.652103027 | 0.021321147 |
| sp\|P12081\|SYHC_HUMAN | 2 | 15 | 28 | 11 | 0.18278 | 0.096971317 | 0.09830656 | -0.085808659 | -0.084473416 | 0.001335243 | 0.730965869 | 0.820479446 | 0.999938193 |
| sp\|O15061\|SYNEM_HUMAN | 15 | 4 | 6 | 3 | 0.11486 | 0.098117404 | 1.038768902 | -0.016742623 | 0.923908875 | 0.940651498 | 0.999562981 | 0.419962086 | 0.356632627 |
| sp\|P17050\|NAGAB_HUMAN | 8 | 40 | 76 | 33 | 0.227421 | 0.179452416 | 0.107944006 | -0.047968487 | -0.119476897 | -0.07150841 | 0.523767882 | 0.066586234 | 0.285434128 |
| sp\|Q9NR12\|PDLI7_HUMAN | 12 | 40 | 76 | 33 | 0.071441 | -0.073804319 | 0.349661095 | -0.145245129 | 0.278220286 | 0.423465415 | 0.4922195 | 0.169561739 | 0.006345442 |
| sp\|Q96CN7\|ISOC1_HUMAN | 166 | 40 | 76 | 33 | -0.126 | -0.158352177 | 0.013111569 | -0.032349318 | 0.139114428 | 0.171463746 | 0.874917764 | 0.18685109 | 0.041071731 |
| sp\|Q9H4G4\|GAPR1_HUMAN | 15 | 40 | 76 | 33 | 0.102094 | 0.064076594 | 0.007635722 | -0.038017018 | -0.09445789 | -0.056440872 | 0.661878747 | 0.176672908 | 0.451424978 |
| sp\|P52790\|HXK3_HUMAN | 15 | 40 | 76 | 33 | -0.4556 | -0.386865701 | 0.096672221 | 0.068737094 | 0.552275016 | 0.483537922 | 0.735740315 | 5.11E-06 | 6.70E-06 |
| sp\|P30613\|KPYR_HUMAN | 5 | 40 | 76 | 33 | -0.14506 | -0.015939458 | 0.239062987 | 0.129116086 | 0.384118531 | 0.255002445 | 0.216312468 | 0.000164586 | 0.006352511 |
| sp\|P00451\|FA8_HUMAN | 94 | 40 | 76 | 33 | -0.10071 | 0.037506493 | 0.438319966 | 0.138211567 | 0.539025039 | 0.400813472 | 0.041745594 | 1.84E-12 | 1.81E-09 |
| sp\|Q86UD1\|OAF_HUMAN | 10 | 40 | 76 | 33 | 0.157855 | 0.212153447 | 0.472405869 | 0.054298047 | 0.31455047 | 0.260252422 | 0.565441276 | 7.23E-06 | 2.92E-05 |
| sp\|Q9NR99\|MXRA5_HUMAN | 9 | 40 | 76 | 33 | 0.007784 | -0.004555788 | 0.281592709 | -0.012340274 | 0.273808222 | 0.286148497 | 0.974199715 | 0.000279381 | 1.55E-05 |
| sp\|Q9NNW7\|TRXR2_HUMAN | 17 | 4 | 7 | 3 | -0.11502 | -0.068510339 | 1.098196762 | 0.046512424 | 1.213219524 | 1.166707101 | 0.984448962 | 0.010470813 | 0.007053599 |
| sp\|Q92824\|PCSK5_HUMAN | 11 | 19 | 34 | 14 | -0.1695 | -0.148259847 | 0.472046855 | 0.02124072 | 0.641547423 | 0.620306703 | 0.986712491 | 0.000859439 | 0.000341569 |
| sp\|P23470\|PTPRG_HUMAN | 2 | 40 | 76 | 33 | 0.151802 | 0.047583604 | 0.094406068 | -0.104218204 | -0.057395741 | 0.046822464 | 0.028758629 | 0.465625048 | 0.523039195 |
| sp\|P16402\|H13_HUMAN | 1 | 11 | 21 | 9 | -0.59699 | -0.465027823 | 0.566445158 | 0.13195977 | 1.163432752 | 1.031472982 | 0.912898352 | 0.013824211 | 0.013803874 |
| sp\|Q14247\|SRC8_HUMAN | 9 | 40 | 76 | 33 | -0.0623 | -0.225813759 | 0.035404563 | -0.163510993 | 0.097707328 | 0.261218322 | 0.451478642 | 0.821046558 | 0.171378545 |
| sp\|P78504\|JAG1_HUMAN | 7 | 40 | 76 | 33 | 0.025911 | 0.063286311 | 0.102937244 | 0.03737559 | 0.077026523 | 0.039650933 | 0.557894441 | 0.184451936 | 0.561747245 |
| sp\|Q15019\|SEPT2_HUMAN | 144 | 40 | 76 | 33 | -0.01205 | -0.096925379 | 0.277008097 | -0.084874428 | 0.289059048 | 0.373933476 | 0.752533198 | 0.107604308 | 0.009724705 |
| sp\|P22748\|CAH4_HUMAN | 31 | 32 | 62 | 27 | -0.13582 | -0.158850451 | 0.105083001 | -0.023026386 | 0.240907066 | 0.263933452 | 0.970020505 | 0.105397585 | 0.032680304 |
| sp\|Q16394\|EXT1_HUMAN | 10 | 40 | 76 | 33 | 0.151433 | 0.074477438 | 0.13448841 | -0.076955743 | -0.016944772 | 0.060010972 | 0.08777317 | 0.919960609 | 0.268690014 |
| sp\|P02533\|K1C14_HUMAN | 2 | 36 | 70 | 30 | -0.01512 | -0.053497984 | -0.028845403 | -0.038375014 | -0.013722432 | 0.024652581 | 0.953226795 | 0.995790714 | 0.982682995 |
| sp\|Q8WZA1\|PMGT1_HUMAN | 13 | 40 | 76 | 33 | 0.150586 | 0.124991643 | -0.112569852 | -0.025594286 | -0.263155782 | -0.237561496 | 0.716160296 | 1.57E-09 | 8.31E-10 |
| sp\|P09960\|LKHA4_HUMAN | 16 | 40 | 76 | 33 | -0.22026 | -0.025233771 | 0.430040908 | 0.195030226 | 0.650304905 | 0.455274678 | 0.007030021 | 9.49E-14 | 1.09E-09 |
| sp\|O75509\|TNR21_HUMAN | 17 | 40 | 76 | 33 | 0.033807 | 0.006450347 | 0.179379338 | -0.027357007 | 0.145571984 | 0.172928992 | 0.876006406 | 0.080280189 | 0.011771372 |
| sp\|P34913\|HYES_HUMAN | 2 | 29 | 55 | 24 | -0.23947 | -0.06964499 | 0.052859595 | 0.169824653 | 0.292329238 | 0.122504585 | 0.393625923 | 0.151562032 | 0.650647366 |
| sp\|O15335\|CHAD_HUMAN | 2 | 40 | 76 | 33 | 0.289343 | 0.251055217 | -0.174470591 | -0.038287816 | -0.463813624 | -0.425525808 | 0.863171631 | 1.95E-06 | 8.50E-07 |
| sp\|O15143\|ARC1B_HUMAN | 5 | 40 | 76 | 33 | -0.21289 | -0.230643968 | 0.095333155 | -0.017754857 | 0.308222265 | 0.325977122 | 0.992096472 | 0.197505772 | 0.101008412 |
| sp\|O43665\|RGS10_HUMAN | 3 | 40 | 76 | 33 | -0.17396 | -0.204438005 | 0.083050841 | -0.030481082 | 0.257007764 | 0.287488846 | 0.96927526 | 0.221568282 | 0.092746585 |
| sp\|P02730\|B3AT_HUMAN | 8 | 40 | 76 | 33 | -1.17185 | -0.979120225 | -0.388328298 | 0.19272909 | 0.783521016 | 0.590791926 | 0.456663082 | 0.000249613 | 0.002184674 |
| sp\|P58215\|LOXL3_HUMAN | 5 | 40 | 76 | 33 | -0.27574 | 0.001089983 | -0.12735828 | 0.276832833 | 0.14838457 | -0.128448263 | 0.012312933 | 0.405163368 | 0.422380142 |

| | | | | | | | | | | | | | |
|---|---|---|---|---|---|---|---|---|---|---|---|---|---|
| sp\|P05771\|KPCB_HUMAN | 9 | 40 | 76 | 33 | -0.33718 | -0.186352856 | -0.025191144 | 0.150827121 | 0.311988833 | 0.161161712 | 0.543918821 | 0.169339282 | 0.543122572 |
| sp\|Q13561\|DCTN2_HUMAN | 1 | 33 | 63 | 27 | -0.22981 | -0.134230934 | 0.30926801 | 0.095578523 | 0.539077467 | 0.443498944 | 0.69807514 | 0.000716321 | 0.001830984 |
| sp\|Q9BZR6\|RTN4R_HUMAN | 12 | 36 | 68 | 30 | 0.085379 | 0.074627217 | 0.254402853 | -0.010751899 | 0.169023738 | 0.179775637 | 0.989031034 | 0.156357818 | 0.070862743 |
| sp\|P07585\|PGS2_HUMAN | 2 | 40 | 76 | 33 | -0.03701 | -0.006095973 | 0.219752173 | 0.030917684 | 0.256765831 | 0.225848147 | 0.910611037 | 0.01401282 | 0.014932328 |
| sp\|P62993\|GRB2_HUMAN | 159 | 40 | 76 | 33 | -0.17686 | -0.28047464 | 0.192277725 | -0.103612139 | 0.369140225 | 0.472752364 | 0.675709746 | 0.035804772 | 0.00120512 |
| sp\|Q9NQ38\|ISK5_HUMAN | 7 | 40 | 76 | 33 | 0.114116 | 0.018100474 | 0.186350223 | -0.09601547 | 0.072234279 | 0.168249748 | 0.230267932 | 0.560520885 | 0.021005146 |
| sp\|Q5VW32\|BROX_HUMAN | 6 | 18 | 35 | 16 | -0.01424 | -0.12915479 | 0.205633296 | -0.114916978 | 0.219871109 | 0.334788087 | 0.646764939 | 0.325600056 | 0.03911416 |
| sp\|P20929\|NEBU_HUMAN | 5 | 11 | 20 | 8 | 0.032741 | -0.152616468 | 0.339768481 | -0.18535774 | 0.307027209 | 0.492384949 | 0.68514487 | 0.511295314 | 0.130248855 |
| sp\|Q06830\|PRDX1_HUMAN | 15 | 40 | 76 | 33 | -0.0059 | 0.221103686 | 0.51550997 | 0.227005693 | 0.521411977 | 0.294406284 | 0.01941964 | 1.91E-06 | 0.003281959 |
| sp\|Q9H251\|CAD23_HUMAN | 2 | 40 | 76 | 33 | 0.084604 | 0.104236019 | -0.190857433 | 0.019632173 | -0.275461278 | -0.295093452 | 0.893394754 | 1.41E-06 | 6.87E-09 |
| sp\|P01036\|CYTS_HUMAN | 27 | 33 | 61 | 27 | -0.05515 | -0.250166058 | 0.019867454 | -0.195012163 | 0.075021349 | 0.270033511 | 0.503645841 | 0.931532786 | 0.31887211 |
| sp\|Q10472\|GALT1_HUMAN | 32 | 40 | 76 | 33 | 0.081366 | 0.10447713 | 0.142015582 | 0.023110967 | 0.060649418 | 0.037538452 | 0.731879127 | 0.231026151 | 0.486805826 |
| sp\|P00568\|KAD1_HUMAN | 1 | 40 | 76 | 33 | -0.24962 | -0.041415425 | 0.277244545 | 0.208204505 | 0.526864475 | 0.318659969 | 0.104381665 | 8.95E-05 | 0.010659548 |
| sp\|O00592\|PODXL_HUMAN | 17 | 40 | 76 | 33 | 0.158028 | 0.149865826 | 0.051220736 | -0.008162334 | -0.106807424 | -0.09864509 | 0.97570063 | 0.059470302 | 0.047072892 |
| sp\|P01880\|IGHD_HUMAN | 8 | 40 | 76 | 33 | -0.33563 | -0.144439631 | -0.320638842 | 0.19118721 | 0.014987998 | -0.176199211 | 0.350293375 | 0.995512282 | 0.456375359 |
| sp\|Q6UX73\|CP089_HUMAN | 5 | 29 | 55 | 25 | 0.040021 | 0.092594386 | 0.138977814 | 0.052572959 | 0.098956387 | 0.046383428 | 0.707594552 | 0.422910343 | 0.783461001 |
| sp\|P51149\|RAB7A_HUMAN | 6 | 40 | 76 | 33 | -0.19631 | -0.23634675 | 0.045128726 | -0.040034575 | 0.241440902 | 0.281475477 | 0.93888984 | 0.210648764 | 0.069734014 |
| sp\|P42229\|STA5A_HUMAN | 22 | 21 | 42 | 17 | -0.12341 | -0.137611224 | 0.38679079 | -0.014197596 | 0.510204418 | 0.524402015 | 0.994199529 | 0.009390031 | 0.002031621 |
| sp\|Q99832\|TCPH_HUMAN | 6 | 29 | 56 | 24 | 0.239755 | 0.217889496 | 0.545260015 | -0.021865384 | 0.305505135 | 0.327370519 | 0.972417266 | 0.027779812 | 0.005736574 |
| sp\|P69891\|HBG1_HUMAN | 4 | 40 | 76 | 33 | -1.34798 | -1.29786924 | -0.259552757 | 0.050107232 | 1.088423715 | 1.038316483 | 0.970149181 | 0.000120656 | 3.28E-05 |
| sp\|P04062\|GLCM_HUMAN | 189 | 40 | 76 | 33 | 0.132762 | 0.158286774 | 0.23984081 | 0.02552443 | 0.107078465 | 0.081554035 | 0.704881647 | 0.016809101 | 0.047469402 |
| sp\|A6NI73\|LIRA5_HUMAN | 72 | 29 | 55 | 24 | -0.43988 | -0.307286352 | 0.171978985 | 0.132595998 | 0.611861335 | 0.479265337 | 0.472126088 | 5.21E-05 | 0.000377084 |
| sp\|O00154\|BACH_HUMAN | 11 | 12 | 20 | 8 | -0.3169 | -0.232956491 | 0.091116285 | 0.083945444 | 0.40801822 | 0.324072776 | 0.945986065 | 0.440478942 | 0.538169281 |
| sp\|P60842\|IF4A1_HUMAN | 96 | 40 | 76 | 33 | -0.16454 | -0.134090281 | 0.286363142 | 0.030445206 | 0.45089863 | 0.420453423 | 0.966508031 | 0.00742058 | 0.004520629 |
| sp\|P51692\|STA5B_HUMAN | 41 | 4 | 7 | 3 | -0.32657 | -0.053245196 | 0.658991502 | 0.273325847 | 0.985562544 | 0.712236698 | 0.74101557 | 0.11268254 | 0.224221622 |
| sp\|P51161\|FABP6_HUMAN | 192 | 40 | 76 | 33 | 0.005392 | -0.083600226 | 0.151815706 | -0.088991835 | 0.146424097 | 0.235415932 | 0.653039206 | 0.452547952 | 0.077285119 |
| sp\|Q9H1E3\|NUCKS_HUMAN | 15 | 4 | 6 | 3 | -0.24323 | 0.290643712 | 1.909053721 | 0.533874303 | 2.152284312 | 1.618410009 | 0.853427779 | 0.205767346 | 0.33333906 |
| sp\|Q9H0R4\|HDHD2_HUMAN | 11 | 29 | 55 | 25 | -0.38148 | -0.105344769 | 0.152149096 | 0.276139358 | 0.533633223 | 0.257493865 | 0.125811237 | 0.005209323 | 0.193835042 |
| sp\|Q9NP79\|VTA1_HUMAN | 2 | 22 | 41 | 18 | 0.081402 | -0.071609832 | 0.156723741 | -0.153011832 | 0.075321742 | 0.228333573 | 0.513948751 | 0.893539983 | 0.277531466 |
| sp\|P98066\|TSG6_HUMAN | 1 | 40 | 76 | 33 | -0.07679 | -0.057004292 | 0.050763318 | 0.019789927 | 0.127557538 | 0.10776761 | 0.93668083 | 0.158778131 | 0.187433446 |
| sp\|Q06481\|APLP2_HUMAN | 24 | 40 | 76 | 33 | -0.33416 | -0.061016751 | 0.099758932 | 0.273145956 | 0.433921638 | 0.160775683 | 0.004598687 | 0.000117266 | 0.18283836 |
| sp\|P21399\|ACOC_HUMAN | 8 | 40 | 76 | 33 | -0.16954 | -0.097330072 | 0.108525329 | 0.072208503 | 0.278063903 | 0.205855401 | 0.615517287 | 0.008629648 | 0.034703102 |
| sp\|Q504Y2\|PKDCC_HUMAN | 43 | 40 | 76 | 33 | 0.046595 | -0.056143631 | -0.003646847 | -0.10273816 | -0.050241375 | 0.052496785 | 0.114941411 | 0.695047312 | 0.603728589 |
| sp\|P62906\|RL10A_HUMAN | 28 | 23 | 41 | 17 | -0.11861 | -0.050057588 | 0.297363194 | 0.06854939 | 0.415970172 | 0.347420782 | 0.872163021 | 0.041655696 | 0.064255814 |
| sp\|P01236\|PRL_HUMAN | 21 | 24 | 49 | 22 | -0.28144 | -0.127720008 | 0.190111353 | 0.153716534 | 0.471547895 | 0.317831361 | 0.440501836 | 0.005700967 | 0.041076729 |
| sp\|P25788\|PSA3_HUMAN | 22 | 40 | 76 | 33 | -0.44956 | -0.217854065 | 0.400699658 | 0.231706004 | 0.850259727 | 0.618553723 | 0.14138152 | 1.18E-07 | 1.37E-05 |
| sp\|O14737\|PDCD5_HUMAN | 14 | 13 | 27 | 12 | -0.67007 | -0.410843468 | 0.049206567 | 0.259224938 | 0.719274973 | 0.460050035 | 0.811576092 | 0.32691289 | 0.539882023 |
| sp\|O00194\|RB27B_HUMAN | 16 | 40 | 76 | 33 | 0.035361 | 0.109384775 | 0.276326329 | 0.074023752 | 0.240965306 | 0.166941553 | 0.751672594 | 0.128903438 | 0.283205613 |
| sp\|Q8WW52\|F151A_HUMAN | 5 | 30 | 56 | 24 | -0.26128 | 0.023936667 | -0.030377714 | 0.28521222 | 0.230897839 | -0.054314381 | 0.245775113 | 0.530412262 | 0.956423078 |
| sp\|P01130\|LDLR_HUMAN | 113 | 40 | 76 | 33 | -0.10144 | -0.03031294 | 0.270434734 | 0.071130592 | 0.371878267 | 0.300747675 | 0.620678075 | 0.000243824 | 0.000903357 |
| sp\|P60981\|DEST_HUMAN | 22 | 40 | 76 | 33 | 0.034237 | -0.169883879 | 0.092787272 | -0.204121183 | 0.058549967 | 0.262671151 | 0.353825615 | 0.942143045 | 0.222422411 |
| sp\|Q9Y281\|COF2_HUMAN | 27 | 7 | 13 | 6 | 0.413198 | 0.770429476 | 1.040904547 | 0.357231734 | 0.627706805 | 0.270475071 | 0.622939396 | 0.363732187 | 0.780782256 |
| sp\|Q99650\|OSMR_HUMAN | 4 | 40 | 76 | 33 | 0.155494 | 0.113459315 | 0.181370753 | -0.04203447 | 0.025876967 | 0.067911437 | 0.671105031 | 0.900668911 | 0.402901524 |
| sp\|Q9H361\|PABP3_HUMAN | 30 | 40 | 76 | 33 | -0.172 | -0.118554807 | 0.304786794 | 0.05344761 | 0.47678921 | 0.4233416 | 0.816375349 | 4.09E-05 | 3.97E-05 |
| sp\|Q99941\|ATF6B_HUMAN | 24 | 40 | 76 | 33 | 0.036678 | 0.036639231 | 0.027548085 | -3.86E-05 | -0.009129765 | -0.009091146 | 0.999999119 | 0.966604916 | 0.95805386 |
| sp\|Q702N8\|XIRP1_HUMAN | 5 | 4 | 6 | 3 | -0.06484 | -0.014239519 | 0.824595702 | 0.050603479 | 0.8894387 | 0.838835221 | 0.991002856 | 0.188108967 | 0.17839664 |
| sp\|Q8WVQ1\|CANT1_HUMAN | 1 | 40 | 76 | 33 | 0.021111 | 0.196580853 | 0.276370962 | 0.17546991 | 0.255260019 | 0.079790109 | 3.20E-05 | 4.74E-07 | 0.13049872 |
| sp\|Q8WUM4\|PDC6I_HUMAN | 119 | 40 | 76 | 33 | -0.11292 | -0.028997675 | 0.199589287 | 0.083919819 | 0.312506781 | 0.228586962 | 0.403988546 | 0.000311005 | 0.003595854 |
| sp\|P42126\|ECI1_HUMAN | 1 | 40 | 76 | 33 | -0.16136 | 0.045357055 | 0.142611229 | 0.206713168 | 0.303967342 | 0.097254174 | 0.045055493 | 0.0104873 | 0.538920349 |
| sp\|Q9UKV8\|AGO2_HUMAN | 3 | 37 | 69 | 30 | -0.15356 | -0.071916049 | 0.167408894 | 0.081640277 | 0.32096522 | 0.239324943 | 0.580544953 | 0.00417107 | 0.020054647 |
| sp\|Q12906\|ILF3_HUMAN | 147 | 40 | 76 | 33 | -0.27543 | -0.131706217 | 0.163708261 | 0.143719813 | 0.439134292 | 0.295414478 | 0.256768275 | 0.000278494 | 0.007699806 |
| sp\|Q12913\|PTPRJ_HUMAN | 7 | 40 | 76 | 33 | 0.263511 | 0.193336655 | -0.034453251 | -0.070174143 | -0.297964049 | -0.227789906 | 0.036296842 | 5.06E-14 | 1.05E-11 |
| sp\|O94933\|SLIK3_HUMAN | 1 | 40 | 76 | 33 | 0.104755 | 0.109955594 | -0.229708794 | 0.005200909 | -0.33446348 | -0.339664389 | 0.999030186 | 0.067773085 | 0.030098758 |
| sp\|P28827\|PTPRM_HUMAN | 5 | 40 | 76 | 33 | 0.113045 | 0.089021111 | 0.066582702 | -0.024024026 | -0.046462434 | -0.022438409 | 0.889662804 | 0.739981692 | 0.914323395 |
| sp\|Q04917\|1433F_HUMAN | 2 | 40 | 76 | 33 | -0.08393 | -0.152575511 | 0.317657712 | -0.068645264 | 0.401587959 | 0.470233224 | 0.868729955 | 0.040032702 | 0.004132006 |
| sp\|Q4VXU2\|PAP1L_HUMAN | 21 | 36 | 69 | 30 | -0.35828 | -0.301770973 | 0.075693842 | 0.056513177 | 0.433977992 | 0.377464816 | 0.841446751 | 0.001403103 | 0.001723266 |
| sp\|P27348\|1433T_HUMAN | 13 | 40 | 76 | 33 | -0.25805 | -0.149347771 | 0.251477453 | 0.108705229 | 0.509530453 | 0.400825224 | 0.60241251 | 0.000758496 | 0.003226708 |
| sp\|P31946\|1433B_HUMAN | 10 | 40 | 76 | 33 | -0.17466 | -0.233681362 | 0.30341794 | -0.059023725 | 0.478075577 | 0.537099302 | 0.896554367 | 0.008969775 | 0.000624615 |
| sp\|Q9NQS3\|NECT3_HUMAN | 13 | 40 | 76 | 33 | -0.00575 | -0.024263938 | 0.096108957 | -0.018517425 | 0.10185547 | 0.120372896 | 0.909693118 | 0.144023068 | 0.033613385 |
| sp\|Q9UKY7\|CDV3_HUMAN | 16 | 32 | 62 | 28 | -0.06272 | -0.119283203 | 0.421728055 | -0.056559337 | 0.484451921 | 0.541011258 | 0.890145631 | 0.003486455 | 0.000150078 |
| sp\|P08637\|FCG3A_HUMAN | 2 | 37 | 69 | 30 | -0.31379 | -0.07243856 | 0.164899075 | 0.241350914 | 0.478688549 | 0.237337635 | 0.060340471 | 0.000709589 | 0.093622031 |
| sp\|P61916\|NPC2_HUMAN | 4 | 40 | 76 | 33 | -0.04654 | 0.007275255 | 0.385785443 | 0.053814251 | 0.432324439 | 0.378510188 | 0.61749578 | 1.24E-08 | 1.83E-08 |
| sp\|Q86YT9\|JAML_HUMAN | 2 | 40 | 76 | 33 | -0.17935 | -0.113011228 | 0.189219759 | 0.06633686 | 0.368567847 | 0.302230987 | 0.614164364 | 7.48E-05 | 0.000267875 |
| sp\|P63010\|AP2B1_HUMAN | 3 | 40 | 76 | 33 | -0.15332 | -0.200031143 | 0.369853177 | -0.046710742 | 0.523173578 | 0.56988432 | 0.881696626 | 5.03E-05 | 5.78E-07 |
| sp\|O94769\|ECM2_HUMAN | 12 | 40 | 76 | 33 | 0.072605 | 0.056516112 | 0.153399867 | -0.01608887 | 0.080794886 | 0.096883755 | 0.873247579 | 0.099745501 | 0.015924693 |
| sp\|P49588\|SYAC_HUMAN | 3 | 40 | 76 | 33 | -0.11701 | -0.076581344 | 0.282269598 | 0.04043189 | 0.399282832 | 0.358850942 | 0.928963633 | 0.008860265 | 0.007815522 |
| sp\|P07306\|ASGR1_HUMAN | 8 | 29 | 55 | 25 | -0.38389 | -0.379165397 | 0.280273726 | 0.004722072 | 0.664161195 | 0.659439123 | 0.999352805 | 0.000283283 | 4.20E-05 |
| sp\|Q00722\|PLCB2_HUMAN | 13 | 4 | 7 | 3 | 0.198677 | 0.091203945 | 0.337457819 | -0.107473133 | 0.138780741 | 0.246253874 | 0.930154924 | 0.921971865 | 0.735909222 |
| sp\|Q9BYJ0\|FGFP2_HUMAN | 2 | 40 | 76 | 33 | -0.15178 | -0.006298747 | 0.06846106 | 0.145478391 | 0.220238199 | 0.074759807 | 0.033478806 | 0.005132404 | 0.445308049 |
| sp\|P17405\|ASM_HUMAN | 9 | 37 | 69 | 30 | 0.003549 | 0.132861663 | 0.448828496 | 0.129312427 | 0.44527926 | 0.315966833 | 0.336606715 | 0.000265468 | 0.00457549 |
| sp\|Q9NUQ9\|FA49B_HUMAN | 10 | 40 | 76 | 33 | -0.20585 | -0.160420243 | 0.322889653 | 0.045427507 | 0.528737403 | 0.483309896 | 0.825184285 | 1.80E-07 | 7.59E-08 |
| sp\|P11277\|SPTB1_HUMAN | 5 | 11 | 21 | 9 | 0.327362 | -0.011628958 | 0.448732151 | -0.33899078 | 0.12137033 | 0.460361109 | 0.255007834 | 0.882751261 | 0.116701274 |
| sp\|P06737\|PYGL_HUMAN | 3 | 40 | 76 | 33 | -0.14567 | -0.008880306 | 0.401445825 | 0.136787396 | 0.547113527 | 0.410326131 | 0.355123052 | 2.96E-05 | 0.000472703 |
| sp\|Q13576\|IQGA2_HUMAN | 127 | 36 | 70 | 30 | -0.07906 | -0.129148782 | 0.242835028 | -0.050092961 | 0.321890849 | 0.37198381 | 0.91612143 | 0.088420589 | 0.016852037 |
| sp\|P14780\|MMP9_HUMAN | 9 | 40 | 76 | 33 | -0.55379 | -0.118001263 | 0.34919934 | 0.43578706 | 0.902987663 | 0.467200603 | 0.001683035 | 3.45E-08 | 0.00159318 |
| sp\|P39059\|COFA1_HUMAN | 14 | 40 | 76 | 33 | 0.116499 | 0.130281771 | 0.280055419 | 0.013783118 | 0.163556767 | 0.149773649 | 0.9467242 | 0.00639133 | 0.004632723 |
| sp\|O75976\|CBPD_HUMAN | 128 | 40 | 76 | 33 | 0.034366 | 0.068904846 | 0.154830664 | 0.034538916 | 0.120464734 | 0.085925818 | 0.662438482 | 0.034995723 | 0.111214633 |
| sp\|Q12905\|ILF2_HUMAN | 10 | 40 | 76 | 33 | -0.14349 | 0.112838553 | 0.486245596 | 0.256330211 | 0.629737254 | 0.373407043 | 0.042988447 | 5.86E-06 | 0.003295312 |
| sp\|O14602\|IF1AY_HUMAN | 2 | 37 | 69 | 29 | -0.14951 | -0.088995234 | 0.278267986 | 0.060517409 | 0.427780629 | 0.36726322 | 0.797823546 | 0.000846562 | 0.00138506 |
| sp\|P05026\|AT1B1_HUMAN | 31 | 40 | 76 | 33 | 0.047828 | 0.052312889 | 0.083643651 | 0.004485004 | 0.035815767 | 0.031330762 | 0.989692692 | 0.635257857 | 0.642835961 |
| sp\|Q06141\|REG3A_HUMAN | 1 | 40 | 76 | 33 | -0.70058 | -0.73335799 | -0.094173879 | -0.032778569 | 0.606405542 | 0.639184111 | 0.976106477 | 0.004496317 | 0.000559057 |
| sp\|Q9BQE3\|TBA1C_HUMAN | 1 | 4 | 7 | 3 | 0.108225 | 0.258429718 | 0.02684863 | 0.150204826 | -0.081376263 | -0.231581088 | 0.780828958 | 0.951384191 | 0.622079923 |
| sp\|P13612\|ITA4_HUMAN | 44 | 7 | 13 | 6 | -0.0575 | 0.078249918 | 0.341809585 | 0.135745641 | 0.399305308 | 0.263559667 | 0.347659723 | 0.005036263 | 0.039301962 |
| sp\|P13639\|EF2_HUMAN | 4 | 40 | 76 | 33 | -0.73982 | -0.402636025 | 0.146990096 | 0.337186436 | 0.886812557 | 0.549626121 | 0.009523211 | 3.54E-09 | 3.39E-05 |
| sp\|P05166\|PCCB_HUMAN | 17 | 4 | 7 | 3 | 0.03017 | 0.189029052 | 1.330520947 | 0.158858725 | 1.30035062 | 1.141491895 | 0.926852711 | 0.069914571 | 0.078738006 |
| sp\|P61981\|1433G_HUMAN | 99 | 40 | 76 | 33 | -0.10204 | -0.147222514 | 0.368370248 | -0.045181508 | 0.470411255 | 0.515592762 | 0.925428238 | 0.00415119 | 0.000282604 |
| sp\|O76003\|GLRX3_HUMAN | 15 | 40 | 76 | 33 | -0.37048 | -0.210607346 | 0.149120069 | 0.15987033 | 0.519597746 | 0.359727415 | 0.310543764 | 0.000346651 | 0.006740651 |
| sp\|A6NMY6\|AXA2L_HUMAN | 10 | 33 | 62 | 26 | -0.26099 | -0.229940557 | 0.073789178 | 0.031054249 | 0.334783984 | 0.303729736 | 0.953350764 | 0.027532765 | 0.024257295 |
| sp\|Q9P1F3\|ABRAL_HUMAN | 5 | 37 | 69 | 29 | -0.76101 | -0.344132621 | 0.186823075 | 0.416879224 | 0.94783492 | 0.530955696 | 0.130960612 | 0.001176296 | 0.062480738 |
| sp\|O95967\|FBLN4_HUMAN | 16 | 40 | 76 | 33 | -0.15367 | -0.123780498 | 0.186061683 | 0.029889638 | 0.339731819 | 0.309842181 | 0.833132141 | 6.61E-07 | 3.16E-07 |
| sp\|P05090\|APOD_HUMAN | 10 | 40 | 76 | 33 | 0.230711 | 0.016963375 | 0.208200954 | -0.213747278 | -0.022509699 | 0.191237579 | 0.000393602 | 0.937197773 | 0.00359657 |
| sp\|Q4KMG0\|CDON_HUMAN | 10 | 40 | 76 | 33 | -0.00623 | 0.052039139 | -0.063597604 | 0.058270398 | -0.057366346 | -0.115636743 | 0.504772834 | 0.632373227 | 0.097922496 |
| sp\|Q16643\|DREB_HUMAN | 9 | 40 | 76 | 33 | -0.07542 | 0.018983494 | 0.463975501 | 0.09440846 | 0.539400467 | 0.444992007 | 0.762688281 | 0.003122394 | 0.006523477 |
| sp\|P49767\|VEGFC_HUMAN | 9 | 40 | 76 | 33 | 0.000121 | 0.255451509 | 0.285525049 | 0.255330892 | 0.285404432 | 0.03007354 | 0.002935846 | 0.006317928 | 0.927450821 |
| sp\|P23467\|PTPRB_HUMAN | 5 | 40 | 76 | 33 | 0.116048 | 0.118398265 | 0.024267706 | 0.002349943 | -0.091780616 | -0.094130559 | 0.996929968 | 0.043629351 | 0.015774495 |
| sp\|Q12841\|FSTL1_HUMAN | 2 | 40 | 76 | 33 | 0.056713 | 0.042239629 | 0.2509916 | -0.014473466 | 0.194278505 | 0.208751971 | 0.952156833 | 0.003182751 | 0.000261504 |
| sp\|P80294\|MT1H_HUMAN | 2 | 36 | 69 | 30 | -1.27888 | -1.084787199 | -0.157828232 | 0.194091791 | 1.121050758 | 0.926958968 | 0.49152635 | 6.38E-07 | 3.20E-06 |
| sp\|P07438\|MT1B_HUMAN | 2 | 21 | 41 | 17 | -1.02943 | -0.671455005 | 0.47443796 | 0.357976872 | 1.503869836 | 1.145892965 | 0.340750031 | 1.74E-05 | 0.000210898 |
| sp\|O75874\|IDHC_HUMAN | 31 | 40 | 76 | 33 | -0.2716 | 0.072158912 | 0.360794406 | 0.343754165 | 0.632389658 | 0.288635494 | 0.000441819 | 5.80E-08 | 0.007352951 |
| sp\|Q9BXJ4\|C1QT3_HUMAN | 26 | 40 | 76 | 33 | 0.563733 | 0.393978552 | 0.077391785 | -0.169754199 | -0.486340966 | -0.316586767 | 0.057555598 | 5.02E-07 | 0.000256112 |
| sp\|P29279\|CCN2_HUMAN | 24 | 40 | 76 | 33 | -0.18331 | -0.12562091 | 0.269438617 | 0.057684548 | 0.452744075 | 0.395059527 | 0.741717114 | 1.12E-05 | 1.55E-05 |
| sp\|P19957\|ELAF_HUMAN | 8 | 40 | 76 | 33 | -0.28215 | -0.202931119 | 0.221190476 | 0.079218624 | 0.503340218 | 0.424121595 | 0.700767096 | 0.000113447 | 0.00025625 |
| sp\|P03973\|SLPI_HUMAN | 4 | 40 | 76 | 33 | -0.14936 | -0.048391704 | 0.342911647 | 0.100969378 | 0.49227273 | 0.391303351 | 0.168918799 | 4.08E-11 | 2.32E-09 |
| sp\|Q9BX93\|PG12B_HUMAN | 8 | 40 | 76 | 33 | 0.047884 | 0.057815551 | 0.223263619 | 0.009932047 | 0.175380116 | 0.165448069 | 0.981646045 | 0.021697476 | 0.013342405 |
| sp\|Q8TEU8\|WFKN2_HUMAN | 6 | 40 | 76 | 33 | 0.170618 | 0.079797849 | -0.145553403 | -0.090820299 | -0.316171551 | -0.225351252 | 0.346178123 | 0.000258748 | 0.004158566 |

| | | | | | | | | | | | | |
|---|---|---|---|---|---|---|---|---|---|---|---|---|
| sp\|P04054\|PA21B_HUMAN | 9 | 40 | 76 | 33 | -0.19658 | -0.004269493 | -0.031707468 | 0.19231546 | 0.164877484 | -0.027437975 | 0.152742636 | 0.382080351 | 0.966338048 |
| sp\|Q14508\|WFDC2_HUMAN | 13 | 40 | 76 | 33 | -1.3826 | -1.302509458 | -0.325046331 | 0.080093057 | 1.057556183 | 0.977463127 | 0.878619298 | 1.16E-06 | 4.03E-07 |
| sp\|P01859\|IGHG2_HUMAN | 7 | 40 | 76 | 33 | -0.18204 | -0.120685319 | 0.013136108 | 0.061350846 | 0.195172273 | 0.133821428 | 0.77736841 | 0.177054574 | 0.352322295 |
| sp\|P11717\|MPRI_HUMAN | 16 | 40 | 76 | 33 | 0.071738 | 0.067760421 | 0.201935167 | -0.003977367 | 0.130197379 | 0.134174746 | 0.992109277 | 0.003946662 | 0.000642182 |
| sp\|P30086\|PEBP1_HUMAN | 1 | 40 | 76 | 33 | -0.14204 | -0.000800814 | 0.404649338 | 0.141241717 | 0.546691869 | 0.405450152 | 0.177511797 | 1.54E-07 | 1.12E-05 |
| sp\|P05161\|ISG15_HUMAN | 8 | 40 | 76 | 33 | -0.56885 | -0.022637958 | 0.150286693 | 0.54620845 | 0.7191331 | 0.172924651 | 1.86E-06 | 1.91E-07 | 0.272480933 |
| sp\|Q04760\|LGUL_HUMAN | 25 | 40 | 76 | 33 | -0.07746 | 0.041099664 | 0.282784825 | 0.118561573 | 0.360246735 | 0.241685162 | 0.194435693 | 6.45E-05 | 0.003236541 |
| sp\|P98155\|VLDLR_HUMAN | 36 | 40 | 76 | 33 | -0.07914 | -0.050531433 | 0.027894646 | 0.028612677 | 0.107038756 | 0.078426079 | 0.929541267 | 0.496186526 | 0.618876358 |
| sp\|P16581\|LYAM2_HUMAN | 2 | 40 | 76 | 33 | -0.10858 | -0.080787822 | 0.251369343 | 0.027791504 | 0.359948669 | 0.332157165 | 0.925301128 | 0.000253921 | 0.00013405 |
| sp\|Q96GP6\|SREC2_HUMAN | 157 | 40 | 76 | 33 | -0.00656 | -0.092745407 | 0.290013195 | -0.086183344 | 0.296575258 | 0.382758602 | 0.497994775 | 0.004390475 | 1.79E-05 |
| sp\|P17252\|KPCA_HUMAN | 1 | 10 | 21 | 9 | 0.000992 | 0.124239838 | 0.236062625 | 0.123247973 | 0.23507076 | 0.111822787 | 0.77887976 | 0.533521453 | 0.825587007 |
| sp\|P07477\|TRY1_HUMAN | 1 | 40 | 76 | 33 | 0.118633 | 0.106414072 | 0.148743315 | -0.012218598 | 0.030110644 | 0.042329243 | 0.979139732 | 0.915514357 | 0.801190766 |
| sp\|Q14314\|FGL2_HUMAN | 10 | 40 | 76 | 33 | -0.21237 | -0.073781698 | 0.098118569 | 0.138592875 | 0.310493142 | 0.171900267 | 0.054781052 | 7.93E-05 | 0.020563333 |
| sp\|Q9UK23\|NAGPA_HUMAN | 1 | 40 | 76 | 33 | 0.191227 | 0.168826488 | 0.043171273 | -0.022400583 | -0.148055798 | -0.125655215 | 0.737426085 | 0.000210901 | 0.000407396 |
| sp\|O95428\|PPN_HUMAN | 1 | 40 | 76 | 33 | 0.019309 | 0.039002354 | 0.205617777 | 0.019693434 | 0.186308857 | 0.166615422 | 0.929605063 | 0.013376537 | 0.012433545 |
| sp\|P78552\|I13R1_HUMAN | 8 | 40 | 76 | 33 | 0.006806 | 0.002666849 | 0.072811791 | -0.004138784 | 0.066006157 | 0.070144941 | 0.994664466 | 0.394690203 | 0.264388075 |
| sp\|Q86TH1\|ATL2_HUMAN | 17 | 40 | 76 | 33 | -0.05033 | 0.077220703 | 0.416597349 | 0.127546489 | 0.466923135 | 0.339376647 | 0.201518959 | 1.74E-06 | 9.71E-05 |
| sp\|Q9ULV4\|COR1C_HUMAN | 16 | 40 | 76 | 33 | -0.14697 | -0.149568157 | 0.086590542 | -0.002596184 | 0.233562515 | 0.236158699 | 0.999855337 | 0.448864027 | 0.353645123 |
| sp\|Q9BR76\|COR1B_HUMAN | 9 | 40 | 76 | 33 | -0.14703 | -0.201921878 | 0.15063118 | -0.054890274 | 0.297662784 | 0.352553058 | 0.896404511 | 0.114021507 | 0.022110506 |
| sp\|Q8IW52\|SLIK4_HUMAN | 7 | 25 | 48 | 21 | 0.344418 | 0.177491572 | 0.207318503 | -0.166926428 | -0.137099496 | 0.029826932 | 0.101347087 | 0.335865416 | 0.935093509 |
| sp\|P06731\|CEAM5_HUMAN | 1 | 4 | 6 | 3 | -0.13024 | 0.441305672 | 0.744486561 | 0.571546563 | 0.874727452 | 0.303180889 | 0.706735642 | 0.567017644 | 0.919730531 |
| sp\|P07203\|GPX1_HUMAN | 3 | 32 | 63 | 27 | -0.13805 | -0.125591183 | 0.184064708 | 0.012462706 | 0.322118597 | 0.309655891 | 0.993611779 | 0.057566514 | 0.03391693 |
| sp\|Q8NHW5\|RLA0L_HUMAN | 2 | 40 | 76 | 33 | -0.35629 | -0.166352771 | 0.465882703 | 0.189937532 | 0.822173006 | 0.632235474 | 0.163201539 | 2.33E-09 | 1.83E-07 |
| sp\|Q13867\|BLMH_HUMAN | 1 | 40 | 76 | 33 | -0.06557 | -0.019860967 | 0.250828891 | 0.045713853 | 0.316403711 | 0.270689858 | 0.737326053 | 9.89E-05 | 0.000177804 |
| sp\|Q9NYL9\|TMOD3_HUMAN | 9 | 17 | 35 | 15 | -0.36651 | -0.07500991 | 0.416681336 | 0.291501252 | 0.783192498 | 0.491691246 | 0.385827749 | 0.01137754 | 0.08916676 |
| sp\|P51858\|HDGF_HUMAN | 10 | 32 | 62 | 28 | -0.07412 | 0.05397265 | 0.468570942 | 0.128092628 | 0.542690921 | 0.414598292 | 0.68929475 | 0.011111759 | 0.032245995 |
| sp\|Q9UL46\|PSME2_HUMAN | 35 | 40 | 76 | 33 | -0.16594 | -0.017931937 | 0.332673701 | 0.14801158 | 0.498617218 | 0.350605637 | 0.139426001 | 1.02E-06 | 0.000116574 |
| sp\|P78509\|RELN_HUMAN | 14 | 40 | 76 | 33 | 0.19313 | 0.200121807 | 0.349912537 | 0.006991464 | 0.156782193 | 0.14979073 | 0.986920414 | 0.0126889 | 0.006481555 |
| sp\|P50053\|KHK_HUMAN | 1 | 8 | 14 | 6 | 0.0401 | 0.049079681 | 0.343576742 | 0.008979626 | 0.303476688 | 0.294497062 | 0.998598013 | 0.356575316 | 0.306758713 |
| sp\|Q13228\|SBP1_HUMAN | 5 | 40 | 76 | 33 | 0.201723 | 0.393090055 | 0.640658961 | 0.191367422 | 0.438936329 | 0.247568907 | 0.014366143 | 7.79E-07 | 0.002196868 |
| sp\|Q92496\|FHR4_HUMAN | 8 | 40 | 76 | 33 | -0.04066 | -0.059648774 | 0.004969077 | -0.018986138 | 0.045631713 | 0.064617851 | 0.955858583 | 0.835543585 | 0.633113242 |
| sp\|Q7LFX5\|CHSTF_HUMAN | 5 | 40 | 76 | 33 | 0.041682 | 0.050993152 | 0.279428874 | 0.009310718 | 0.237746439 | 0.228435721 | 0.980141925 | 0.000247004 | 6.52E-05 |
| sp\|P61160\|ARP2_HUMAN | 7 | 40 | 76 | 33 | -0.02665 | -0.083206882 | 0.171809641 | -0.056552686 | 0.198463837 | 0.255016523 | 0.834737422 | 0.220145138 | 0.043837557 |
| sp\|P48426\|PI42A_HUMAN | 7 | 40 | 76 | 33 | -0.08066 | -0.023082753 | 0.302345532 | 0.057579692 | 0.383007977 | 0.325428285 | 0.886128011 | 0.028410888 | 0.03760742 |
| sp\|Q5VY43\|PEAR1_HUMAN | 2 | 40 | 76 | 33 | 0.156371 | 0.124578936 | 0.078164841 | -0.031792164 | -0.078206259 | -0.046414095 | 0.625334154 | 0.145114502 | 0.416966845 |
| sp\|P52823\|STC1_HUMAN | 18 | 6 | 14 | 7 | 0.023067 | -0.259113836 | 0.578819627 | -0.282181246 | 0.555752218 | 0.837933464 | 0.594773018 | 0.226912962 | 0.013849447 |
| sp\|Q04446\|GLGB_HUMAN | 2 | 40 | 76 | 33 | -0.08099 | -0.068361392 | 0.121923719 | 0.012629141 | 0.202914252 | 0.190285111 | 0.970697991 | 0.006527498 | 0.003700174 |
| sp\|O95897\|NOE2_HUMAN | 2 | 40 | 76 | 33 | 0.083777 | 0.091578939 | 0.090752328 | 0.007801519 | 0.006974908 | -0.000826611 | 0.991848691 | 0.995496037 | 0.999919322 |
| sp\|Q15435\|PP1R7_HUMAN | 18 | 40 | 76 | 33 | -0.22515 | -0.143617446 | 0.124434805 | 0.081530668 | 0.349582919 | 0.268052251 | 0.406340274 | 3.01E-05 | 0.000346911 |
| sp\|P09958\|FURIN_HUMAN | 25 | 10 | 20 | 10 | 0.098563 | 0.220581031 | 0.541388348 | 0.122018406 | 0.442825723 | 0.320807317 | 0.617555325 | 0.014407403 | 0.046175067 |
| sp\|Q9UBW8\|CSN7A_HUMAN | 3 | 4 | 6 | 3 | 0.039803 | 0.168491914 | 0.001582195 | 0.1286893 | -0.038220419 | -0.166909719 | 0.390020543 | 0.936988546 | 0.279784202 |
| sp\|P11413\|G6PD_HUMAN | 2 | 40 | 76 | 33 | -0.34941 | -0.274410765 | 0.217794693 | 0.074998588 | 0.567204046 | 0.492205458 | 0.864069885 | 0.004250763 | 0.005281128 |
| sp\|P10645\|CMGA_HUMAN | 9 | 40 | 76 | 33 | -0.02011 | -0.185809101 | 0.262882237 | -0.165699942 | 0.282991396 | 0.448691338 | 0.162160068 | 0.027459018 | 2.16E-05 |
| sp\|P35030\|TRY3_HUMAN | 2 | 40 | 76 | 33 | 0.108814 | -0.059306606 | 0.076135193 | -0.168120213 | -0.032678414 | 0.135441799 | 0.009697086 | 0.88108392 | 0.067614984 |
| sp\|P21266\|GSTM3_HUMAN | 23 | 40 | 76 | 33 | -0.1396 | 0.005811232 | 0.359859991 | 0.145414988 | 0.499463747 | 0.354048759 | 0.171013867 | 2.43E-06 | 0.000184995 |
| sp\|P48060\|GLIP1_HUMAN | 4 | 33 | 62 | 28 | 0.298762 | 0.344817879 | 0.283040479 | 0.046055998 | -0.015721402 | -0.061777399 | 0.875223805 | 0.989120799 | 0.806902987 |
| sp\|P08238\|HS90B_HUMAN | 2 | 40 | 76 | 33 | -0.0372 | 0.009111358 | 0.426132695 | 0.046308935 | 0.463330272 | 0.417021337 | 0.889140279 | 0.000535356 | 0.000431186 |
| sp\|Q99466\|NOTC4_HUMAN | 5 | 40 | 76 | 33 | 0.013242 | -0.005224119 | -0.006303051 | -0.018466509 | -0.019545441 | -0.001078931 | 0.926964061 | 0.943053327 | 0.999772539 |
| sp\|O60704\|TPST2_HUMAN | 20 | 4 | 7 | 3 | 0.098164 | 0.119162914 | 0.305059781 | 0.020998507 | 0.206895374 | 0.185896867 | 0.967453064 | 0.162226394 | 0.16501523 |
| sp\|Q9NTU7\|CBLN4_HUMAN | 4 | 40 | 76 | 33 | 0.215563 | 0.131832302 | -0.383469298 | -0.083730267 | -0.599031867 | -0.5153016 | 0.4029073 | 6.82E-12 | 2.38E-11 |
| sp\|P23435\|CBLN1_HUMAN | 6 | 40 | 76 | 33 | 0.265362 | 0.009369555 | -0.314494551 | -0.255991988 | -0.579856095 | -0.323864106 | 0.002904233 | 9.44E-09 | 0.000322634 |
| sp\|Q8IUK8\|CBLN2_HUMAN | 3 | 37 | 69 | 30 | 0.497742 | 0.324586265 | 0.00424122 | -0.17315618 | -0.493501225 | -0.320345045 | 0.251920853 | 0.000719161 | 0.018656742 |
| sp\|Q9NP84\|TNR12_HUMAN | 26 | 40 | 76 | 33 | -0.88015 | -0.878124569 | 0.12707055 | 0.002025204 | 1.007220322 | 1.005195119 | 0.999913954 | 2.35E-06 | 1.13E-07 |
| sp\|P10599\|THIO_HUMAN | 4 | 40 | 76 | 33 | -0.16707 | -0.010094679 | 0.482406575 | 0.156973614 | 0.649474868 | 0.492501254 | 0.18671813 | 3.48E-08 | 2.21E-06 |
| sp\|P11142\|HSP7C_HUMAN | 2 | 40 | 76 | 33 | -0.06812 | -0.026920031 | 0.375715055 | 0.04119542 | 0.443830506 | 0.402635086 | 0.912479495 | 0.001058897 | 0.000782208 |
| sp\|Q05655\|KPCD_HUMAN | 38 | 15 | 28 | 11 | -0.18259 | -0.323322737 | 0.160827632 | -0.140733774 | 0.343416595 | 0.484150369 | 0.709169 | 0.272513823 | 0.04573299 |
| sp\|P48506\|GSH1_HUMAN | 1 | 40 | 76 | 33 | -0.39166 | -0.071624713 | 0.284937752 | 0.320036665 | 0.67659913 | 0.356562465 | 0.000110469 | 2.04E-11 | 5.22E-05 |
| sp\|O95185\|UNC5C_HUMAN | 20 | 30 | 55 | 23 | -0.0253 | -0.013648566 | 0.255259492 | 0.01164901 | 0.280557067 | 0.268908057 | 0.987932685 | 0.011592489 | 0.006329618 |
| sp\|P14207\|FOLR2_HUMAN | 3 | 40 | 76 | 33 | -0.14034 | -0.060280505 | 0.159666946 | 0.080057186 | 0.300004638 | 0.219947451 | 0.378310842 | 0.000162664 | 0.00220803 |
| sp\|Q9NPH3\|IL1AP_HUMAN | 1 | 40 | 76 | 33 | 0.141183 | 0.170276921 | -0.13460036 | 0.02909383 | -0.275783451 | -0.304877281 | 0.785491061 | 1.83E-06 | 3.46E-09 |
| sp\|O60667\|FAIM3_HUMAN | 27 | 40 | 76 | 33 | 0.007692 | 0.115997188 | 0.151320444 | 0.108305102 | 0.143628358 | 0.035323256 | 0.170434374 | 0.117791097 | 0.845296926 |
| sp\|Q9BRN9\|TM2D3_HUMAN | 1 | 13 | 27 | 13 | -0.10204 | -0.014182027 | -0.064550148 | 0.087853352 | 0.037485232 | -0.050368121 | 0.314282249 | 0.852268793 | 0.678746844 |
| sp\|P11362\|FGFR1_HUMAN | 10 | 40 | 76 | 33 | 0.204572 | 0.149975654 | 0.017493467 | -0.054596508 | -0.187078695 | -0.132482186 | 0.302452031 | 0.00012668 | 0.002751575 |
| sp\|P05106\|ITB3_HUMAN | 60 | 40 | 76 | 33 | -0.57087 | -0.289532118 | -0.24270006 | 0.281335465 | 0.328167522 | 0.046832057 | 0.042055937 | 0.050764182 | 0.923466709 |
| sp\|Q96TA1\|NIBA2_HUMAN | 3 | 40 | 76 | 33 | 0.124623 | 0.10460916 | 0.113989295 | -0.020013404 | -0.010633269 | 0.009380135 | 0.874468149 | 0.974182225 | 0.974431233 |
| sp\|Q9ULI3\|HEG1_HUMAN | 41 | 40 | 76 | 33 | 0.192811 | 0.145056861 | 0.183472614 | -0.047754042 | -0.00933829 | 0.038415753 | 0.236132972 | 0.962083549 | 0.438041992 |
| sp\|P54652\|HSP72_HUMAN | 11 | 11 | 20 | 9 | -0.18266 | -1.026036042 | -0.213692994 | -0.84337749 | -0.031034442 | 0.812343048 | 0.184660264 | 0.998320377 | 0.250651586 |
| sp\|Q14CN2\|CLCA4_HUMAN | 19 | 11 | 21 | 9 | 0.167327 | -0.289746537 | -0.285476955 | -0.457073996 | -0.452804413 | 0.004269582 | 0.072659067 | 0.163939274 | 0.999783905 |
| sp\|P13284\|GILT_HUMAN | 5 | 19 | 35 | 14 | -0.35422 | -0.298965468 | 0.22537581 | 0.055255336 | 0.579596614 | 0.524341278 | 0.873698535 | 0.000237948 | 0.000212968 |
| sp\|O14917\|PCD17_HUMAN | 6 | 36 | 69 | 30 | -0.19116 | -0.137870729 | 0.197284821 | 0.053288995 | 0.388444546 | 0.335155551 | 0.681485072 | 3.97E-06 | 6.85E-06 |
| sp\|O75051\|PLXA2_HUMAN | 45 | 40 | 76 | 33 | 0.013781 | -0.021197908 | -0.001979774 | -0.034978985 | -0.015760851 | 0.019218134 | 0.686187981 | 0.948438676 | 0.90474109 |
| sp\|P13501\|CCL5_HUMAN | 2 | 40 | 76 | 33 | -0.09394 | -0.057730983 | -0.176542136 | 0.036209817 | -0.082601336 | -0.118811153 | 0.958484506 | 0.858931737 | 0.67080477 |
| sp\|Q9P126\|CLC1B_HUMAN | 7 | 10 | 21 | 10 | -0.02646 | -0.156446936 | 0.670817902 | -0.129982195 | 0.697282643 | 0.827264838 | 0.843447188 | 0.037200316 | 0.002971447 |
| sp\|P14324\|FPPS_HUMAN | 13 | 40 | 76 | 33 | -0.0812 | -0.121247011 | 0.168695898 | -0.040043043 | 0.249899866 | 0.289942909 | 0.891185394 | 0.049602631 | 0.006465405 |
| sp\|Q9H2E6\|SEM6A_HUMAN | 14 | 40 | 76 | 33 | 0.03536 | 0.056740375 | 0.060394173 | 0.02138076 | 0.025034558 | 0.003653798 | 0.841869984 | 0.849724922 | 0.995588309 |
| sp\|P11586\|C1TC_HUMAN | 8 | 26 | 49 | 21 | -0.19821 | -0.098032252 | 0.213622266 | 0.10018184 | 0.411836358 | 0.311654518 | 0.484601895 | 0.000496905 | 0.003484185 |
| sp\|Q13201\|MMRN1_HUMAN | 6 | 40 | 76 | 33 | -0.33048 | 0.089761726 | 0.28252018 | 0.420241135 | 0.612999588 | 0.192758453 | 0.000349508 | 1.20E-05 | 0.208846464 |
| sp\|O75095\|MEGF6_HUMAN | 2 | 29 | 55 | 23 | -0.00734 | -0.002969365 | 0.559274744 | 0.00436935 | 0.56661346 | 0.562244109 | 0.999211522 | 0.000309653 | 5.29E-05 |
| sp\|O00548\|DLL1_HUMAN | 30 | 40 | 76 | 33 | -0.0603 | -0.026280845 | 0.291794654 | 0.034019808 | 0.352095307 | 0.318075499 | 0.876769987 | 0.000135348 | 9.87E-05 |
| sp\|Q03154\|ACY1_HUMAN | 76 | 40 | 76 | 33 | -0.43603 | -0.103209463 | 0.192779926 | 0.33282332 | 0.62881271 | 0.29598939 | 0.00740854 | 1.15E-05 | 0.03156321 |
| sp\|P15153\|RAC2_HUMAN | 2 | 32 | 62 | 28 | -0.4223 | -0.356901831 | 0.126993246 | 0.065395029 | 0.549290106 | 0.483895076 | 0.815168652 | 0.000100967 | 9.89E-05 |
| sp\|Q9Y274\|SIA10_HUMAN | 36 | 40 | 76 | 33 | 0.067329 | 0.071362207 | -0.052349599 | 0.004033264 | -0.119678542 | -0.123711806 | 0.991673358 | 0.007959377 | 0.001551986 |
| sp\|P07686\|HEXB_HUMAN | 22 | 40 | 76 | 33 | 0.052021 | 0.060846448 | 0.302417097 | 0.008825487 | 0.250396136 | 0.241570649 | 0.985465508 | 0.000532041 | 0.000147412 |
| sp\|O43866\|CD5L_HUMAN | 152 | 40 | 76 | 33 | -0.15356 | -0.056821836 | 0.415708213 | 0.096741855 | 0.569271905 | 0.47253005 | 0.471939981 | 1.68E-07 | 9.38E-07 |
| sp\|P00995\|ISK1_HUMAN | 17 | 40 | 76 | 33 | -0.24445 | -0.206174043 | 0.341394806 | 0.038278283 | 0.585847131 | 0.547568848 | 0.945154096 | 0.000227463 | 9.60E-05 |
| sp\|P01591\|IGJ_HUMAN | 1 | 40 | 76 | 33 | 0.16767 | 0.112705135 | 0.067138897 | -0.054965344 | -0.100531582 | -0.045566238 | 0.198218154 | 0.025781583 | 0.374168738 |
| sp\|Q5KU26\|COL12_HUMAN | 57 | 40 | 76 | 33 | -0.35396 | -0.373904568 | 0.002651595 | -0.019940723 | 0.35661544 | 0.376556163 | 0.952775396 | 6.11E-05 | 1.63E-06 |
| sp\|Q13564\|ULA1_HUMAN | 35 | 16 | 35 | 16 | -0.254 | -0.105924111 | 0.317897993 | 0.148072339 | 0.571894442 | 0.423822104 | 0.434115791 | 0.00035904 | 0.002077018 |
| sp\|Q5VU97\|CAHD1_HUMAN | 2 | 37 | 69 | 30 | 0.110529 | 0.182523255 | 0.243184113 | 0.071994222 | 0.13265508 | 0.060660859 | 0.239947627 | 0.038132876 | 0.412686456 |
| sp\|P21802\|FGFR2_HUMAN | 6 | 40 | 76 | 33 | 0.039804 | 0.037562806 | 0.115527555 | -0.002241309 | 0.07572344 | 0.077964749 | 0.99890738 | 0.426046847 | 0.317481164 |
| sp\|Q58EX2\|SDK2_HUMAN | 1 | 40 | 76 | 33 | 0.195973 | 0.216623581 | 0.050558112 | 0.020650526 | -0.145414942 | -0.166065468 | 0.884366736 | 0.017648681 | 0.00144781 |
| sp\|Q92626\|PXDN_HUMAN | 1 | 40 | 76 | 33 | -0.20099 | -0.057131534 | 0.225196867 | 0.143859227 | 0.426187628 | 0.282328401 | 0.035424081 | 1.97E-08 | 2.63E-05 |
| sp\|Q13630\|FCL_HUMAN | 1 | 40 | 76 | 33 | -0.26699 | -0.160413888 | 0.114827281 | 0.10657343 | 0.381814599 | 0.275241169 | 0.353202195 | 0.000192294 | 0.003004806 |
| sp\|P53621\|COPA_HUMAN | 5 | 14 | 27 | 11 | -0.07893 | 0.022602995 | 0.538906959 | 0.101532869 | 0.617836833 | 0.516303964 | 0.844540889 | 0.021762821 | 0.032575704 |
| sp\|P13645\|K1C10_HUMAN | 8 | 40 | 76 | 33 | 0.263668 | 0.056933835 | -0.101109936 | -0.206733844 | -0.364777615 | -0.158043771 | 0.380088511 | 0.128154071 | 0.607235943 |
| sp\|Q04695\|K1C17_HUMAN | 113 | 11 | 21 | 9 | 0.033091 | -0.062554612 | -0.203528975 | -0.095645487 | -0.23661985 | -0.140974363 | 0.945875777 | 0.792725582 | 0.900018392 |
| sp\|P01210\|PENK_HUMAN | 31 | 8 | 14 | 6 | -0.1476 | -0.115291744 | -0.055822795 | 0.032304851 | 0.0917738 | 0.059468949 | 0.989201786 | 0.942814961 | 0.970128003 |
| sp\|O75563\|SKAP2_HUMAN | 41 | 36 | 69 | 31 | 0.031184 | 0.022491202 | 0.234163194 | -0.008692722 | 0.20297927 | 0.211671992 | 0.996378492 | 0.253730255 | 0.148972916 |
| sp\|Q9Y266\|NUDC_HUMAN | 15 | 40 | 76 | 33 | -0.35116 | -0.232452349 | 0.331386433 | 0.11870619 | 0.682544972 | 0.563838782 | 0.532637949 | 2.80E-06 | 1.30E-05 |
| sp\|P22894\|MMP8_HUMAN | 3 | 28 | 55 | 24 | -0.55902 | -0.313844809 | 0.310740816 | 0.24517539 | 0.869761014 | 0.624585625 | 0.226736646 | 1.03E-05 | 0.000340072 |
| sp\|P11021\|BIP_HUMAN | 26 | 40 | 76 | 33 | 0.155221 | 0.201180766 | 0.174080132 | 0.045959931 | 0.018859297 | -0.027100634 | 0.365513991 | 0.888661684 | 0.733797093 |
| sp\|O60462\|NRP2_HUMAN | 12 | 40 | 76 | 33 | 0.016484 | 0.058944305 | 0.254246944 | 0.042460054 | 0.237762693 | 0.195302639 | 0.50738901 | 2.17E-06 | 1.16E-05 |
| sp\|Q92896\|GSLG1_HUMAN | 2 | 40 | 76 | 33 | -0.10753 | 0.044477988 | 0.191197298 | 0.152005156 | 0.298724466 | 0.14671931 | 0.011531461 | 1.42E-05 | 0.025245487 |
| sp\|Q05707\|COEA1_HUMAN | 5 | 37 | 69 | 29 | -0.02665 | 0.005219744 | 0.050427386 | 0.031872992 | 0.077080634 | 0.045207642 | 0.885426383 | 0.61974929 | 0.812748692 |
| sp\|Q13790\|APOF_HUMAN | 1 | 40 | 76 | 33 | -0.20939 | 0.190182297 | 0.343354898 | 0.399571707 | 0.552744309 | 0.153172601 | 1.87E-06 | 4.72E-08 | 0.150524632 |

| | | | | | | | | | | | | |
|---|---|---|---|---|---|---|---|---|---|---|---|---|
| sp\|Q14697\|GANAB_HUMAN | 31 | 40 | 76 | 33 | -0.06303 | 0.031931133 | 0.379093931 | 0.094959618 | 0.442122417 | 0.347162799 | 0.204910387 | 1.99E-09 | 8.95E-08 |
| sp\|P20908\|CO5A1_HUMAN | 6 | 40 | 76 | 33 | -0.241 | -0.195779948 | 0.575448015 | 0.045222163 | 0.816450126 | 0.771227963 | 0.906316967 | 8.19E-09 | 9.55E-10 |
| sp\|O00629\|IMA3_HUMAN | 2 | 4 | 7 | 3 | 0.050546 | 0.020761915 | 0.36609751 | -0.029784291 | 0.315551303 | 0.345335595 | 0.991784092 | 0.554447665 | 0.429871972 |
| sp\|P20042\|IF2B_HUMAN | 132 | 40 | 76 | 33 | -0.12475 | -0.026172971 | 0.326903781 | 0.098577161 | 0.451653913 | 0.353076752 | 0.606560404 | 0.001118326 | 0.004707319 |
| sp\|P09104\|ENOG_HUMAN | 4 | 40 | 76 | 33 | -0.02157 | -0.049409402 | -0.03433117 | -0.027841671 | -0.012763439 | 0.015078232 | 0.940223269 | 0.991097063 | 0.984245196 |
| sp\|P08254\|MMP3_HUMAN | 6 | 40 | 76 | 33 | 0.03444 | -0.068066899 | 0.056720865 | -0.10250669 | 0.022281074 | 0.124787764 | 0.445172038 | 0.97374753 | 0.349822996 |
| sp\|Q96Q06\|PLIN4_HUMAN | 139 | 4 | 6 | 3 | 0.071308 | 0.111021747 | 1.036226729 | 0.039714117 | 0.9649191 | 0.925204983 | 0.996620663 | 0.286352801 | 0.264499558 |
| sp\|Q6UWY5\|OLFL1_HUMAN | 3 | 40 | 76 | 33 | 0.069336 | 0.066637488 | 0.111651741 | -0.002698147 | 0.042316105 | 0.045014252 | 0.9970264 | 0.604884357 | 0.48568858 |
| sp\|Q10567\|AP1B1_HUMAN | 5 | 40 | 76 | 33 | -0.10173 | -0.138793332 | 0.333598051 | -0.037062377 | 0.435329005 | 0.472391383 | 0.948153775 | 0.008027196 | 0.000846999 |
| sp\|P35754\|GLRX1_HUMAN | 29 | 40 | 76 | 33 | -0.85434 | -0.568959001 | 0.115368385 | 0.285384954 | 0.96971234 | 0.684327386 | 0.062062301 | 5.10E-09 | 2.84E-06 |
| sp\|Q99471\|PFD5_HUMAN | 20 | 22 | 42 | 17 | -0.16234 | 0.085572314 | 0.228120697 | 0.247912205 | 0.390460588 | 0.142548383 | 0.108491935 | 0.027879796 | 0.531597781 |
| sp\|Q00169\|PIPNA_HUMAN | 5 | 7 | 14 | 7 | 0.003877 | 0.075812004 | 0.072705557 | 0.071934826 | 0.068828379 | -0.003106448 | 0.933853038 | 0.954054731 | 0.999871998 |
| sp\|Q14974\|IMB1_HUMAN | 1 | 40 | 76 | 33 | -0.22259 | -0.234879564 | 0.18178691 | -0.012286204 | 0.40438027 | 0.416666475 | 0.994610042 | 0.020645937 | 0.0056552 |
| sp\|Q9Y5Y6\|ST14_HUMAN | 2 | 40 | 76 | 33 | -0.37644 | -0.341136124 | 0.17296556 | 0.035306402 | 0.549408086 | 0.514101684 | 0.958287162 | 0.001284823 | 0.000631532 |
| sp\|Q14696\|MESD_HUMAN | 2 | 37 | 69 | 29 | -0.06169 | -0.020988538 | 0.339485515 | 0.040703441 | 0.401177494 | 0.360474053 | 0.897290205 | 0.001331967 | 0.001221305 |
| sp\|P27797\|CALR_HUMAN | 4 | 40 | 76 | 33 | -0.08299 | -0.028629574 | 0.531759992 | 0.054358416 | 0.614747982 | 0.560389566 | 0.797468226 | 3.92E-08 | 1.67E-08 |
| sp\|Q9UIW2\|PLXA1_HUMAN | 16 | 40 | 76 | 33 | 0.034166 | 0.014677663 | 0.111657278 | -0.01948821 | 0.077491405 | 0.096979615 | 0.81754039 | 0.116129192 | 0.014957522 |
| sp\|P54819\|KAD2_HUMAN | 28 | 37 | 69 | 30 | -0.01177 | -0.06262794 | 0.280539144 | -0.050861164 | 0.29230592 | 0.343167084 | 0.855164962 | 0.032447022 | 0.002983666 |
| sp\|Q6IBS0\|TWF2_HUMAN | 4 | 40 | 76 | 33 | 0.066394 | -0.002538292 | 0.374283949 | -0.068932617 | 0.307889624 | 0.37682224 | 0.821058005 | 0.07099324 | 0.007245035 |
| sp\|P06213\|INSR_HUMAN | 5 | 21 | 42 | 17 | 0.127372 | 0.144345289 | 0.036236103 | 0.016973334 | -0.091135851 | -0.108109186 | 0.987794498 | 0.789238597 | 0.65204256 |
| sp\|P09972\|ALDOC_HUMAN | 36 | 40 | 76 | 33 | 0.047322 | 0.040988708 | -0.032713493 | -0.006332792 | -0.080034993 | -0.073702201 | 0.986597257 | 0.231229804 | 0.206335549 |
| sp\|Q6UXI9\|NPNT_HUMAN | 23 | 40 | 76 | 33 | -0.08504 | -0.040867506 | 0.068042243 | 0.044170637 | 0.153080387 | 0.108909749 | 0.79375771 | 0.152419416 | 0.295226788 |
| sp\|Q15370\|ELOB_HUMAN | 5 | 40 | 76 | 33 | 0.289546 | 0.302913263 | 0.720167398 | 0.01336681 | 0.430620944 | 0.417254134 | 0.988152526 | 0.000375925 | 9.15E-05 |
| sp\|Q96NZ9\|PRAP1_HUMAN | 7 | 40 | 76 | 33 | -0.166 | -0.171767688 | 0.314076726 | -0.005771404 | 0.480073011 | 0.485844415 | 0.997581045 | 2.94E-05 | 1.74E-06 |
| sp\|Q00013\|EM55_HUMAN | 10 | 40 | 76 | 33 | -0.37381 | -0.265813922 | 0.104912113 | 0.107993105 | 0.47871914 | 0.370726035 | 0.674285103 | 0.006143907 | 0.019622255 |
| sp\|P55287\|CAD11_HUMAN | 5 | 40 | 76 | 33 | 0.134456 | 0.118424877 | 0.387918417 | -0.016030999 | 0.253462541 | 0.26949354 | 0.962871336 | 0.002169704 | 0.000180847 |
| sp\|P63279\|UBC9_HUMAN | 15 | 23 | 41 | 18 | -0.15026 | -0.142150031 | 0.224439791 | 0.00811324 | 0.374703062 | 0.366589822 | 0.997402914 | 0.02770691 | 0.014845485 |
| sp\|Q96RT1\|ERBIN_HUMAN | 10 | 19 | 34 | 14 | -0.83919 | -0.464221635 | -0.833783935 | 0.374968587 | 0.005406287 | -0.3695623 | 0.575343133 | 0.999923184 | 0.645421856 |
| sp\|Q8NCL4\|GALT6_HUMAN | 3 | 40 | 76 | 33 | 0.383465 | 0.323950626 | 0.398261781 | -0.059514321 | 0.014796833 | 0.074311154 | 0.398295065 | 0.961142003 | 0.284890083 |
| sp\|Q9Y316\|MEMO1_HUMAN | 8 | 8 | 13 | 6 | 0.263275 | -0.281233504 | 0.147225246 | -0.544508713 | -0.116049963 | 0.42845875 | 0.213613787 | 0.949347389 | 0.440842242 |
| sp\|Q8TAG5\|VTM2A_HUMAN | 1 | 40 | 76 | 33 | 0.034413 | -0.076670928 | -0.139057089 | -0.111084084 | -0.173470245 | -0.06238616 | 0.141456982 | 0.038874724 | 0.577146139 |
| sp\|P22303\|ACES_HUMAN | 5 | 36 | 69 | 30 | 0.181483 | 0.13645027 | 0.057506904 | -0.045032493 | -0.123975858 | -0.078943366 | 0.767666072 | 0.254387049 | 0.489446777 |
| sp\|P05121\|PAI1_HUMAN | 4 | 40 | 76 | 33 | -0.34254 | 0.032165671 | 0.208799663 | 0.374702746 | 0.551336738 | 0.176633992 | 0.000492989 | 1.63E-05 | 0.206513042 |
| sp\|P16157\|ANK1_HUMAN | 21 | 33 | 62 | 26 | -0.40746 | -0.090552314 | 0.510919122 | 0.316911709 | 0.918383145 | 0.601471437 | 0.082311619 | 3.21E-06 | 0.00071413 |
| sp\|Q16719\|KYNU_HUMAN | 15 | 29 | 55 | 25 | -0.2727 | -0.144326669 | 0.178135937 | 0.128375784 | 0.450838391 | 0.322462607 | 0.684056006 | 0.041077193 | 0.120067129 |
| sp\|Q9NS00\|C1GLT_HUMAN | 1 | 7 | 14 | 5 | 0.104182 | 0.070233551 | 0.055519616 | -0.033948352 | -0.048662287 | -0.014713935 | 0.936750388 | 0.919585065 | 0.990331266 |
| sp\|O00499\|BIN1_HUMAN | 8 | 22 | 41 | 18 | -0.76478 | -0.839207021 | -0.193810591 | -0.074431451 | 0.570964978 | 0.645396429 | 0.9316069 | 0.063441009 | 0.013038884 |
| sp\|Q01638\|ILRL1_HUMAN | 9 | 40 | 76 | 33 | -0.25226 | -0.238360902 | 0.463369246 | 0.013894505 | 0.715624652 | 0.701730147 | 0.994668347 | 0.000129558 | 2.10E-05 |
| sp\|P50552\|VASP_HUMAN | 3 | 40 | 76 | 33 | -0.34894 | -0.529260501 | -0.172265513 | -0.180318952 | 0.176676036 | 0.356994988 | 0.38168952 | 0.527209179 | 0.039268283 |
| sp\|P47756\|CAPZB_HUMAN | 7 | 40 | 76 | 33 | -0.22185 | -0.213229803 | 0.209877236 | 0.008625025 | 0.431732063 | 0.423107038 | 0.99695683 | 0.006705656 | 0.002347707 |
| sp\|P31749\|AKT1_HUMAN | 2 | 15 | 28 | 11 | 0.013476 | 0.093650182 | 0.110230819 | 0.080174138 | 0.096754775 | 0.016580637 | 0.882655425 | 0.888580583 | 0.995680645 |
| sp\|Q5JSH3\|WDR44_HUMAN | 9 | 40 | 76 | 33 | -0.13793 | -0.114977248 | 0.136803502 | 0.022957553 | 0.274738303 | 0.25178075 | 0.971000454 | 0.059366811 | 0.049181338 |
| sp\|P20774\|MIME_HUMAN | 1 | 40 | 76 | 33 | 0.26511 | 0.172931036 | 0.331909455 | -0.092178518 | 0.066799902 | 0.158978419 | 0.207782472 | 0.562503555 | 0.018267705 |
| sp\|O14980\|XPO1_HUMAN | 4 | 32 | 63 | 27 | -0.00735 | 0.140192734 | 0.574536294 | 0.14754508 | 0.581888639 | 0.434343559 | 0.322664046 | 1.89E-05 | 0.000315168 |
| sp\|P12883\|MYH7_HUMAN | 2 | 40 | 76 | 33 | -0.10036 | -0.305550988 | 0.097775136 | -0.205191722 | 0.198134402 | 0.403326124 | 0.125136361 | 0.259939741 | 0.001193061 |
| sp\|Q9BQB4\|SOST_HUMAN | 1 | 18 | 34 | 14 | 0.019923 | -0.170949623 | 0.389974329 | -0.190873115 | 0.370050837 | 0.560923951 | 0.328531866 | 0.066302209 | 0.00074088 |
| sp\|O95084\|PRS23_HUMAN | 4 | 37 | 69 | 30 | 0.063264 | 0.065847744 | 0.044914839 | 0.002583413 | -0.018349491 | -0.020932904 | 0.998609215 | 0.952874761 | 0.923802413 |
| sp\|Q02108\|GCYA1_HUMAN | 5 | 34 | 62 | 26 | -0.14175 | 0.077058133 | 0.257472199 | 0.218810516 | 0.399224582 | 0.180414066 | 0.078378686 | 0.004102018 | 0.231578329 |
| sp\|Q13332\|PTPRS_HUMAN | 13 | 40 | 76 | 33 | 0.161577 | 0.128591396 | -0.071360158 | -0.032985478 | -0.232937032 | -0.199951554 | 0.680653536 | 7.17E-06 | 1.41E-05 |
| sp\|P23284\|PPIB_HUMAN | 1 | 40 | 76 | 33 | -0.21479 | -0.04540357 | 0.151120439 | 0.169389103 | 0.365913112 | 0.196524009 | 0.124557841 | 0.001624481 | 0.086094585 |
| sp\|Q96HC4\|PDLI5_HUMAN | 16 | 22 | 41 | 17 | -0.41719 | -0.518964243 | 0.335764645 | -0.101776176 | 0.752952712 | 0.854728888 | 0.832574241 | 0.002248174 | 8.65E-05 |
| sp\|P48643\|TCPE_HUMAN | 9 | 36 | 69 | 30 | 0.143227 | 0.229696727 | 0.327382668 | 0.086470023 | 0.184155964 | 0.097685941 | 0.748034679 | 0.404765251 | 0.720849914 |
| sp\|P31937\|3HIDH_HUMAN | 10 | 29 | 55 | 24 | -0.28717 | 0.006289767 | 0.269111612 | 0.293460492 | 0.556282337 | 0.262821845 | 0.054863088 | 0.001044659 | 0.126042263 |
| sp\|Q15126\|PMVK_HUMAN | 46 | 21 | 41 | 18 | -0.2736 | -0.176520699 | 0.209726008 | 0.097080694 | 0.483327401 | 0.386246707 | 0.913469416 | 0.216883726 | 0.282179343 |
| sp\|Q6ZRP7\|QSOX2_HUMAN | 5 | 40 | 76 | 33 | 0.008693 | 0.013838157 | 0.02487588 | 0.005145274 | 0.016182997 | 0.011037723 | 0.997055788 | 0.980079347 | 0.988158388 |
| sp\|P0DP23\|CALM1_HUMAN | 126 | 40 | 76 | 33 | -0.23353 | -0.212735078 | 0.176535234 | 0.020792331 | 0.410062642 | 0.389270312 | 0.988812733 | 0.053484239 | 0.035301887 |
| sp\|P30041\|PRDX6_HUMAN | 4 | 40 | 76 | 33 | -0.39156 | -0.06183631 | 0.332256157 | 0.32972173 | 0.723814197 | 0.394092467 | 0.006045019 | 1.93E-07 | 0.001779029 |
| sp\|O75356\|ENTP5_HUMAN | 6 | 40 | 76 | 33 | 0.069634 | 0.076130332 | 0.09886606 | 0.00649655 | 0.029232277 | 0.022735727 | 0.993390361 | 0.911596731 | 0.931218575 |
| sp\|Q13177\|PAK2_HUMAN | 7 | 36 | 69 | 30 | -0.1328 | -0.137638969 | 0.136008899 | -0.004840294 | 0.268807574 | 0.273647868 | 0.997971443 | 0.01595919 | 0.004489604 |
| sp\|O43852\|CALU_HUMAN | 272 | 40 | 76 | 33 | -0.26332 | 0.050085396 | 0.088980914 | 0.313401223 | 0.352296742 | 0.038895518 | 0.004350935 | 0.008465441 | 0.925260428 |
| sp\|P09382\|LEG1_HUMAN | 1 | 40 | 76 | 33 | 0.033168 | 0.115787414 | 0.43411586 | 0.08261947 | 0.400947916 | 0.318328446 | 0.277581617 | 1.75E-08 | 4.06E-07 |
| sp\|O60883\|G37L1_HUMAN | 5 | 30 | 55 | 25 | -0.09879 | -0.239844654 | -0.10394466 | -0.141053629 | -0.005153635 | 0.135899994 | 0.662068311 | 0.999611244 | 0.712292509 |
| sp\|Q8WZ75\|ROBO4_HUMAN | 2 | 40 | 76 | 33 | 0.114322 | 0.089982902 | 0.149743429 | -0.024339386 | 0.035421141 | 0.059760527 | 0.875594934 | 0.823649691 | 0.497111319 |
| sp\|Q8NBF2\|NHLC2_HUMAN | 16 | 40 | 76 | 33 | -0.26775 | -0.363102033 | -0.074786617 | -0.095352167 | 0.192963249 | 0.288315417 | 0.698910088 | 0.365926123 | 0.060626571 |
| sp\|Q12765\|SCRN1_HUMAN | 1 | 19 | 34 | 15 | -0.15275 | -0.026176545 | 0.281412276 | 0.126576788 | 0.434165609 | 0.307588822 | 0.71659433 | 0.075539458 | 0.194193403 |
| sp\|Q9NZN3\|EHD3_HUMAN | 4 | 40 | 76 | 33 | -0.14924 | -0.243983974 | -0.081911274 | -0.094747143 | 0.067325557 | 0.1620727 | 0.731762665 | 0.896696262 | 0.450149216 |
| sp\|P68104\|EF1A1_HUMAN | 9 | 36 | 69 | 30 | -0.41077 | -0.29815505 | 0.215685382 | 0.112610318 | 0.62645075 | 0.513840432 | 0.632280755 | 0.000127922 | 0.000410919 |
| sp\|Q15303\|ERBB4_HUMAN | 42 | 36 | 69 | 30 | 0.226608 | 0.123980221 | 0.010520811 | -0.102627782 | -0.216087192 | -0.11345941 | 0.132531951 | 0.002628109 | 0.113122899 |
| sp\|Q9UDY2\|ZO2_HUMAN | 1 | 29 | 55 | 23 | 0.095324 | -0.079647291 | 0.219293224 | -0.174971302 | 0.123969214 | 0.298940516 | 0.611489525 | 0.845758395 | 0.296860559 |
| sp\|Q16658\|FSCN1_HUMAN | 5 | 40 | 76 | 33 | -0.09182 | 0.07040312 | 0.165614285 | 0.162223269 | 0.257434434 | 0.095211166 | 0.006122632 | 0.0001905 | 0.202548617 |
| sp\|P22455\|FGFR4_HUMAN | 4 | 40 | 76 | 33 | 0.173338 | 0.040807685 | -0.09964607 | -0.132530399 | -0.272984154 | -0.140453755 | 0.040658873 | 0.000139926 | 0.042450835 |
| sp\|Q9UJW2\|TINAG_HUMAN | 7 | 36 | 69 | 30 | 0.18841 | 0.070593426 | -0.1392192 | -0.117816318 | -0.327628944 | -0.209812626 | 0.208670002 | 0.000393238 | 0.014057348 |
| sp\|O15240\|VGF_HUMAN | 10 | 40 | 76 | 33 | 0.217822 | 0.122076911 | 0.015577737 | -0.095745421 | -0.202244595 | -0.106499174 | 0.391909993 | 0.059152162 | 0.361792417 |
| sp\|P42574\|CASP3_HUMAN | 1 | 40 | 76 | 33 | -0.35487 | -0.182987134 | 0.083972226 | 0.171882621 | 0.43884198 | 0.266959359 | 0.505208384 | 0.049894784 | 0.238194925 |
| sp\|P22607\|FGFR3_HUMAN | 4 | 33 | 62 | 27 | -0.03317 | -0.037052019 | 0.04215367 | -0.003881118 | 0.07532457 | 0.079205688 | 0.996036154 | 0.361016927 | 0.241723874 |
| sp\|P37802\|TAGL2_HUMAN | 3 | 40 | 76 | 33 | 0.037021 | -0.025179568 | 0.322011276 | -0.062200517 | 0.284990327 | 0.347190844 | 0.918952371 | 0.298283348 | 0.104380318 |
| sp\|Q92747\|ARC1A_HUMAN | 19 | 17 | 35 | 16 | -0.12193 | -0.051695981 | 0.386109847 | 0.070236991 | 0.508042819 | 0.437805828 | 0.843050419 | 0.003061762 | 0.003238313 |
| sp\|Q96LZ3\|CANB2_HUMAN | 3 | 21 | 42 | 19 | -0.00982 | -0.080187578 | 0.236609364 | -0.070369774 | 0.246427168 | 0.316796942 | 0.731043657 | 0.071679854 | 0.004245598 |
| sp\|P31949\|S10AB_HUMAN | 23 | 40 | 76 | 33 | -0.78064 | -0.489194716 | 0.215785408 | 0.291443399 | 0.996423523 | 0.704980124 | 0.073858895 | 1.22E-08 | 4.94E-06 |
| sp\|Q9NPR2\|SEM4B_HUMAN | 14 | 40 | 76 | 33 | 0.045841 | 0.11928455 | 0.244309402 | 0.073443728 | 0.198468581 | 0.125024853 | 0.198258645 | 0.000462834 | 0.018000831 |
| sp\|Q8IWY4\|SCUB1_HUMAN | 11 | 29 | 55 | 24 | -0.18078 | -0.13079434 | 0.262291678 | 0.049989563 | 0.443075581 | 0.393086018 | 0.90611271 | 0.006570009 | 0.006524861 |
| sp\|Q96MU8\|KREM1_HUMAN | 16 | 40 | 76 | 33 | 0.120595 | 0.176289928 | 0.136421354 | 0.055694798 | 0.015826224 | -0.039868574 | 0.35940011 | 0.944025961 | 0.629306956 |
| sp\|Q92854\|SEM4D_HUMAN | 11 | 40 | 76 | 33 | -0.13695 | 0.063450579 | 0.07437847 | 0.200396284 | 0.211324175 | 0.010927891 | 5.03E-05 | 0.000432339 | 0.971918578 |
| sp\|Q9Y570\|PPME1_HUMAN | 4 | 4 | 7 | 3 | 0.119982 | 0.105053345 | 0.352378333 | -0.014928999 | 0.232395989 | 0.247324988 | 0.995338278 | 0.49044995 | 0.381141435 |
| sp\|Q9H6X2\|ANTR1_HUMAN | 18 | 40 | 76 | 33 | 0.023907 | -0.058072385 | 0.07689033 | -0.081979524 | 0.052983191 | 0.134962715 | 0.328211293 | 0.723400988 | 0.073523117 |
| sp\|O75832\|PSD10_HUMAN | 8 | 30 | 55 | 23 | 0.238293 | 0.313810074 | 0.474011444 | 0.075516775 | 0.235718145 | 0.160201371 | 0.787647793 | 0.215836633 | 0.410758054 |
| sp\|P52907\|CAZA1_HUMAN | 2 | 40 | 76 | 33 | -0.22003 | -0.187112547 | 0.188290086 | 0.032922404 | 0.408325037 | 0.375402633 | 0.95191474 | 0.006928237 | 0.00484834 |
| sp\|O75368\|SH3L1_HUMAN | 2 | 40 | 76 | 33 | -0.19714 | -0.16777484 | 0.271903852 | 0.029366649 | 0.469045341 | 0.439678692 | 0.964136169 | 0.002365617 | 0.001209492 |
| sp\|Q13200\|PSMD2_HUMAN | 5 | 36 | 69 | 30 | -0.51712 | -0.323543221 | 0.147779601 | 0.193578593 | 0.664901414 | 0.471322822 | 0.215560156 | 1.22E-05 | 0.000530548 |
| sp\|P36969\|GPX4_HUMAN | 2 | 40 | 76 | 33 | 0.35453 | 0.284237736 | 0.386504808 | -0.070292192 | 0.03197488 | 0.102267071 | 0.696929139 | 0.949593434 | 0.512309533 |
| sp\|Q9H7Y0\|DIK2B_HUMAN | 32 | 40 | 76 | 33 | 0.132642 | 0.110853121 | 0.057388984 | -0.021788414 | -0.075252551 | -0.053464137 | 0.905344544 | 0.443859856 | 0.592560311 |
| sp\|P14384\|CBPM_HUMAN | 13 | 40 | 76 | 33 | 0.023882 | -0.015620769 | 0.097784273 | -0.039502462 | 0.07390258 | 0.113405042 | 0.48225057 | 0.175077327 | 0.006413585 |
| sp\|P00387\|NB5R3_HUMAN | 16 | 37 | 69 | 30 | -0.14001 | -0.068346989 | 0.190478421 | 0.071661876 | 0.330487286 | 0.258825411 | 0.704786302 | 0.007662701 | 0.022070462 |
| sp\|P07858\|CATB_HUMAN | 9 | 40 | 76 | 33 | -0.07811 | -0.156310742 | 0.365521175 | -0.078201192 | 0.443630725 | 0.521831916 | 0.564470671 | 1.09E-05 | 6.36E-09 |
| sp\|Q9NZK5\|ADA2_HUMAN | 13 | 40 | 76 | 33 | 0.094761 | 0.100107791 | 0.459232018 | 0.005346997 | 0.364471224 | 0.359124227 | 0.996528829 | 4.14E-05 | 4.88E-06 |
| sp\|Q9NT99\|LRC4B_HUMAN | 1 | 40 | 76 | 33 | 0.215465 | 0.121782958 | -0.142216475 | -0.093682183 | -0.357681615 | -0.263999433 | 0.188675773 | 3.65E-07 | 2.35E-05 |
| sp\|P19440\|GGT1_HUMAN | 6 | 40 | 76 | 33 | -0.15225 | -0.083059062 | 0.189835026 | 0.069185981 | 0.342080069 | 0.272894089 | 0.644260573 | 0.00096312 | 0.003370874 |
| sp\|P47755\|CAZA2_HUMAN | 110 | 40 | 76 | 33 | -0.10398 | -0.149310225 | 0.057949546 | -0.04532842 | 0.161931351 | 0.207259771 | 0.900989168 | 0.402427757 | 0.152850082 |
| sp\|P16083\|NQO2_HUMAN | 7 | 40 | 76 | 33 | -0.37861 | -0.353824073 | 0.066344597 | 0.024783823 | 0.444952492 | 0.420168669 | 0.981012577 | 0.016785268 | 0.009917409 |
| sp\|Q5TFE4\|NT5D1_HUMAN | 6 | 3 | 7 | 4 | -0.00443 | -0.043422583 | 0.156704125 | -0.038992182 | 0.161134526 | 0.200126708 | 0.97006895 | 0.664608777 | 0.409432905 |
| sp\|Q9UKX5\|ITA11_HUMAN | 12 | 25 | 49 | 21 | 0.331864 | 0.145703925 | -0.081867248 | -0.186160445 | -0.413731619 | -0.227571173 | 0.056291359 | 0.00012274 | 0.023050658 |
| sp\|Q92882\|OSTF1_HUMAN | 1 | 40 | 76 | 33 | 0.028412 | 0.007858416 | 0.307039138 | -0.020553556 | 0.278627166 | 0.299180721 | 0.980693894 | 0.089770953 | 0.030272617 |
| sp\|P19652\|A1AG2_HUMAN | 11 | 40 | 76 | 33 | -0.7884 | -0.842035044 | -0.329500929 | -0.053637398 | 0.458896717 | 0.512534115 | 0.941639666 | 0.052518948 | 0.010051293 |
| sp\|P61978\|HNRPK_HUMAN | 12 | 40 | 76 | 33 | -0.35291 | -0.199921782 | 0.424145992 | 0.152986076 | 0.77705385 | 0.624067774 | 0.466557865 | 5.24E-06 | 3.87E-05 |
| sp\|O95497\|VNN1_HUMAN | 13 | 40 | 76 | 33 | 0.152957 | -0.112997126 | -0.114251305 | -0.265954117 | -0.267208296 | -0.001254179 | 0.066554263 | 0.14910339 | 0.999945568 |

| | | | | | | | | | | | | | |
|---|---|---|---|---|---|---|---|---|---|---|---|---|---|
| sp\|O14656\|TOR1A_HUMAN | 8 | 4 | 7 | 3 | 0.088368 | 0.142624131 | 0.395967007 | 0.054255802 | 0.307598678 | 0.253342876 | 0.974736981 | 0.589879113 | 0.642591134 |
| sp\|P40925\|MDHC_HUMAN | 4 | 40 | 76 | 33 | 0.11526 | 0.17570081 | 0.338027203 | 0.06044124 | 0.222767633 | 0.162326393 | 0.594856111 | 0.009413207 | 0.040800607 |
| sp\|O75347\|TBCA_HUMAN | 13 | 40 | 76 | 33 | -0.24048 | -0.141014845 | 0.424380991 | 0.099469947 | 0.664865783 | 0.565395836 | 0.681165127 | 2.25E-05 | 4.94E-05 |
| sp\|P48681\|NEST_HUMAN | 11 | 7 | 13 | 5 | -0.72611 | -0.840250483 | 1.158487457 | -0.114144301 | 1.88459364 | 1.998737941 | 0.97170759 | 0.016711404 | 0.004769422 |
| sp\|Q15555\|MARE2_HUMAN | 12 | 40 | 76 | 33 | -0.16318 | -0.354016367 | -0.098259468 | -0.190839482 | 0.064917417 | 0.255756899 | 0.436238055 | 0.935407141 | 0.272083649 |
| sp\|P12277\|KCRB_HUMAN | 1 | 32 | 62 | 28 | -0.05305 | -0.242735247 | -0.334289547 | -0.189683068 | -0.281237369 | -0.0915543 | 0.354092189 | 0.20113142 | 0.799916485 |
| sp\|Q9NY97\|B3GN2_HUMAN | 7 | 40 | 76 | 33 | 0.173281 | 0.13387899 | 0.065271779 | -0.039402187 | -0.108009398 | -0.068607211 | 0.188843266 | 0.000300668 | 0.013319661 |
| sp\|Q9UHL4\|DPP2_HUMAN | 6 | 32 | 62 | 27 | -0.04003 | -0.039168729 | 0.149459467 | 0.00086266 | 0.189490856 | 0.188628196 | 0.999961327 | 0.279536632 | 0.19877525 |
| sp\|Q9UIV8\|SPB13_HUMAN | 44 | 7 | 14 | 5 | -0.00527 | 0.050683496 | -0.051052447 | 0.055955554 | -0.045780389 | -0.101735942 | 0.934574963 | 0.972037498 | 0.838887673 |
| sp\|Q9BRK5\|CAB45_HUMAN | 13 | 40 | 76 | 33 | -0.23021 | -0.165188008 | 0.220271897 | 0.065020128 | 0.450480033 | 0.385459905 | 0.602793186 | 4.31E-07 | 1.06E-06 |
| sp\|Q8WWU7\|ITLN2_HUMAN | 7 | 40 | 76 | 33 | 0.085793 | -0.079515904 | 0.019470517 | -0.165308907 | -0.066322485 | 0.098986422 | 0.254234778 | 0.857210921 | 0.647000962 |
| sp\|P50395\|GDIB_HUMAN | 9 | 40 | 76 | 33 | 0.083629 | 0.152282603 | 0.446143736 | 0.068654004 | 0.362515137 | 0.293861133 | 0.679959961 | 0.000962888 | 0.002828701 |
| sp\|Q8IW75\|SPA12_HUMAN | 108 | 18 | 35 | 14 | -0.02007 | 0.225622328 | -0.245482418 | 0.245689838 | -0.225414909 | -0.471104746 | 0.612297042 | 0.759790877 | 0.225776804 |
| sp\|Q10471\|GALT2_HUMAN | 6 | 40 | 76 | 33 | -0.07204 | -0.022247773 | 0.167477768 | 0.049794166 | 0.239519707 | 0.189725541 | 0.518546625 | 6.77E-05 | 0.000411851 |
| sp\|Q14C87\|T132D_HUMAN | 4 | 8 | 14 | 6 | 0.127904 | 0.273569283 | -0.76850829 | 0.145665311 | -0.896412263 | -1.042077573 | 0.938542653 | 0.219359093 | 0.08949218 |
| sp\|Q6UVK1\|CSPG4_HUMAN | 3 | 40 | 76 | 33 | 0.125091 | 0.029400593 | 0.291367378 | -0.095690097 | 0.166276688 | 0.261966785 | 0.121311224 | 0.013595737 | 3.48E-06 |
| sp\|Q16531\|DDB1_HUMAN | 1 | 40 | 76 | 33 | -0.25379 | -0.183749524 | 0.260174805 | 0.070038984 | 0.513963312 | 0.443924328 | 0.787685922 | 0.000277363 | 0.000408441 |
| sp\|Q7Z7M8\|B3GN8_HUMAN | 40 | 40 | 76 | 33 | 0.154531 | 0.085712374 | -0.022811796 | -0.068818472 | -0.177342642 | -0.10852417 | 0.22120066 | 0.001400299 | 0.039325346 |
| sp\|Q9Y5C1\|ANGL3_HUMAN | 10 | 40 | 76 | 33 | 0.018562 | 0.044960455 | 0.247354158 | 0.026398358 | 0.228792062 | 0.202393703 | 0.880162557 | 0.001992272 | 0.002042262 |
| sp\|P45378\|TNNT3_HUMAN | 2 | 11 | 20 | 9 | -0.80302 | -1.035245392 | 0.565286662 | -0.232228386 | 1.368303668 | 1.600532054 | 0.862989337 | 0.039058204 | 0.005336952 |
| sp\|Q9NZU0\|FLRT3_HUMAN | 2 | 21 | 42 | 19 | 0.090399 | 0.189111253 | 0.418346926 | 0.098712487 | 0.327948159 | 0.229235673 | 0.679075053 | 0.053901193 | 0.14919511 |
| sp\|P14735\|IDE_HUMAN | 2 | 32 | 62 | 27 | -0.1234 | 0.017363795 | 0.08959927 | 0.14076653 | 0.213002005 | 0.072235475 | 0.139278898 | 0.045495935 | 0.624311738 |
| sp\|Q96BJ3\|AIDA_HUMAN | 2 | 4 | 7 | 3 | 0.204489 | 0.170740902 | 0.349132953 | -0.033748347 | 0.144643705 | 0.178392051 | 0.929122758 | 0.429807339 | 0.227876981 |
| sp\|P08397\|HEM3_HUMAN | 2 | 40 | 76 | 33 | -0.30301 | -0.005561344 | 0.35639975 | 0.29744763 | 0.659408725 | 0.361961094 | 0.006734263 | 2.01E-07 | 0.00164051 |
| sp\|Q9UNM6\|PSD13_HUMAN | 2 | 4 | 7 | 3 | 0.307914 | 0.500701072 | 0.350281905 | 0.192786638 | 0.042367471 | -0.150419168 | 0.771360515 | 0.991388416 | 0.876355737 |
| sp\|P08887\|IL6RA_HUMAN | 32 | 40 | 76 | 33 | 0.036764 | 0.061304133 | -0.008197424 | 0.024540393 | -0.044961165 | -0.069501557 | 0.912105151 | 0.808317014 | 0.525144738 |
| sp\|P53618\|COPB_HUMAN | 2 | 7 | 14 | 6 | -0.12209 | 0.111871257 | 0.635599416 | 0.233958438 | 0.757686597 | 0.523728159 | 0.591209947 | 0.035062275 | 0.111328366 |
| sp\|Q5ZPR3\|CD276_HUMAN | 15 | 40 | 76 | 33 | -0.06203 | -0.026642474 | 0.298198138 | 0.035383252 | 0.360223864 | 0.324840612 | 0.785846448 | 3.11E-07 | 1.97E-07 |
| sp\|Q99715\|COCA1_HUMAN | 10 | 40 | 76 | 33 | 0.067394 | 0.03306853 | 0.040324122 | -0.03432526 | -0.027069669 | 0.007255592 | 0.685311462 | 0.84999192 | 0.985228689 |
| sp\|Q6ICL3\|TNG2_HUMAN | 22 | 40 | 76 | 33 | 0.622236 | 0.72615182 | 1.008805319 | 0.103916081 | 0.38656958 | 0.282653499 | 0.59646605 | 0.008868061 | 0.038307752 |
| sp\|Q08257\|QOR_HUMAN | 2 | 40 | 76 | 33 | -0.09703 | -0.01343668 | 0.077948683 | 0.083590971 | 0.174976334 | 0.091385363 | 0.494173678 | 0.122450482 | 0.477408832 |
| sp\|Q9NQ76\|MEPE_HUMAN | 2 | 40 | 76 | 33 | -0.13459 | 0.024660451 | 0.162047111 | 0.159255052 | 0.296641711 | 0.137386659 | 0.079183168 | 0.00281671 | 0.18788044 |
| sp\|P39687\|AN32A_HUMAN | 4 | 29 | 55 | 25 | -0.33859 | -0.210593079 | 0.391054201 | 0.128000182 | 0.729647462 | 0.60164728 | 0.610289847 | 4.22E-05 | 0.000137682 |
| sp\|Q13813\|SPTN1_HUMAN | 1 | 29 | 55 | 25 | -0.56171 | -0.315832639 | 0.231244287 | 0.245880683 | 0.792957608 | 0.547076926 | 0.225537267 | 5.15E-05 | 0.001870797 |
| sp\|Q14393\|GAS6_HUMAN | 15 | 40 | 76 | 33 | 0.011766 | 0.14643383 | 0.27028648 | 0.13466758 | 0.25852023 | 0.12385265 | 0.030140113 | 0.000206546 | 0.072515451 |
| sp\|P68133\|ACTS_HUMAN | 2 | 40 | 76 | 33 | 0.004768 | 0.035225495 | 0.044454468 | 0.03045757 | 0.039686544 | 0.009228973 | 0.8999168 | 0.883811758 | 0.991526448 |
| sp\|Q562R1\|ACTBL_HUMAN | 22 | 40 | 76 | 33 | -0.24186 | -0.402994982 | 0.019959262 | -0.16113061 | 0.261823635 | 0.422954245 | 0.530422243 | 0.316989414 | 0.024343473 |
| sp\|P02144\|MYG_HUMAN | 6 | 40 | 76 | 33 | -0.86123 | -0.977784507 | -0.397969153 | -0.116559278 | 0.463256076 | 0.579815354 | 0.708047016 | 0.026346093 | 0.000900014 |
| sp\|Q10588\|BST1_HUMAN | 1 | 40 | 76 | 33 | 0.103542 | 0.082387226 | -0.137681086 | -0.021155179 | -0.241223491 | -0.220068312 | 0.922770178 | 0.001182402 | 0.000814392 |
| sp\|Q08174\|PCDH1_HUMAN | 3 | 40 | 76 | 33 | -0.06347 | -0.042754483 | 0.324703189 | 0.020714445 | 0.388172117 | 0.367457672 | 0.922387645 | 5.39E-08 | 7.11E-09 |
| sp\|P78356\|PI42B_HUMAN | 2 | 40 | 76 | 33 | -0.15835 | -0.033506055 | 0.271845528 | 0.124841304 | 0.430192886 | 0.305351583 | 0.500036728 | 0.004463341 | 0.029164169 |
| sp\|Q6NW40\|RGMB_HUMAN | 7 | 40 | 76 | 33 | -0.03175 | -0.095770974 | 0.067552462 | -0.064025874 | 0.099297562 | 0.163323435 | 0.615429985 | 0.448022994 | 0.066562764 |
| sp\|Q15274\|NADC_HUMAN | 6 | 7 | 14 | 6 | -0.30538 | 0.282633174 | 0.259917317 | 0.588014327 | 0.56529847 | -0.022715856 | 0.052676461 | 0.139456721 | 0.995484284 |
| sp\|Q13637\|RAB32_HUMAN | 6 | 4 | 7 | 3 | -0.58172 | -0.279721263 | 0.225052878 | 0.301998788 | 0.806772929 | 0.504774141 | 0.613205536 | 0.132243802 | 0.344404575 |
| sp\|Q15121\|PEA15_HUMAN | 3 | 18 | 34 | 15 | 0.192503 | 0.20238187 | 0.473443642 | 0.009878851 | 0.280940624 | 0.271061772 | 0.996274983 | 0.132939297 | 0.093168223 |
| sp\|P22314\|UBA1_HUMAN | 1 | 40 | 76 | 33 | -0.13881 | -0.106269457 | 0.172415453 | 0.032543583 | 0.311228493 | 0.27868491 | 0.927873034 | 0.010970862 | 0.010051635 |
| sp\|P13611\|CSPG2_HUMAN | 2 | 40 | 76 | 33 | -0.11294 | -0.013628699 | 0.333961399 | 0.099308431 | 0.44689853 | 0.347590098 | 0.280837545 | 1.81E-07 | 4.60E-06 |
| sp\|P29508\|SPB3_HUMAN | 2 | 18 | 34 | 15 | -0.85642 | -0.977297911 | -1.393020142 | -0.120873306 | -0.536595537 | -0.415722231 | 0.951733182 | 0.511904389 | 0.59878759 |
| sp\|P30153\|2AAA_HUMAN | 15 | 40 | 76 | 33 | -0.28246 | -0.127310512 | 0.350180929 | 0.155150838 | 0.632642278 | 0.477491441 | 0.436066401 | 0.000150755 | 0.001477566 |
| sp\|Q15166\|PON3_HUMAN | 3 | 40 | 76 | 33 | 0.221849 | 0.158545197 | -0.289475651 | -0.06330357 | -0.511324417 | -0.448020848 | 0.533623547 | 9.56E-11 | 1.50E-10 |
| sp\|Q7Z5N4\|SDK1_HUMAN | 10 | 40 | 76 | 33 | 0.052193 | -0.017375044 | 0.04414293 | -0.069568379 | -0.008050406 | 0.061517973 | 0.16804111 | 0.98335031 | 0.29183371 |
| sp\|Q7Z7G0\|TARSH_HUMAN | 111 | 40 | 76 | 33 | 0.082764 | 0.070378893 | 0.082058077 | -0.012385181 | -0.000705998 | 0.011679183 | 0.97040185 | 0.999932619 | 0.976814062 |
| sp\|P98153\|IDD_HUMAN | 5 | 21 | 41 | 19 | -0.20981 | -0.069243772 | 0.495943637 | 0.140562501 | 0.70574991 | 0.565187409 | 0.598138207 | 0.000267679 | 0.000913235 |
| sp\|O95998\|I18BP_HUMAN | 26 | 40 | 76 | 33 | -0.2223 | -0.116711495 | 0.369545819 | 0.105592046 | 0.591849359 | 0.486257314 | 0.403846793 | 4.46E-08 | 3.72E-07 |
| sp\|O43529\|CHSTA_HUMAN | 5 | 40 | 76 | 33 | 0.114736 | 0.061161674 | -0.076865765 | -0.053573916 | -0.191601355 | -0.138027439 | 0.318327623 | 8.94E-05 | 0.001779141 |
| sp\|P49591\|SYSC_HUMAN | 8 | 40 | 76 | 33 | 0.461254 | 0.480357555 | 0.678091038 | 0.019103223 | 0.216836706 | 0.197733483 | 0.990656901 | 0.437257016 | 0.416907306 |
| sp\|Q9NRN5\|OLFL3_HUMAN | 18 | 40 | 76 | 33 | 0.041896 | 0.085085987 | 0.093491771 | 0.043189825 | 0.051595609 | 0.008405784 | 0.317942128 | 0.32350369 | 0.962174304 |
| sp\|Q9UJU6\|DBNL_HUMAN | 1 | 40 | 76 | 33 | -0.13376 | -0.10261703 | 0.170079545 | 0.031138697 | 0.303835241 | 0.272696574 | 0.958802642 | 0.06801756 | 0.063710795 |
| sp\|P32321\|DCTD_HUMAN | 5 | 10 | 21 | 9 | 0.021348 | -0.017716041 | 0.279993703 | -0.039064158 | 0.258645585 | 0.297709743 | 0.968557105 | 0.386051367 | 0.193645136 |
| sp\|O95260\|ATE1_HUMAN | 19 | 11 | 21 | 9 | -0.0489 | -0.188716721 | 0.003136427 | -0.139819503 | 0.052033644 | 0.191853147 | 0.799259023 | 0.978816916 | 0.693089016 |
| sp\|Q03167\|TGBR3_HUMAN | 6 | 40 | 76 | 33 | 0.13397 | 0.085115738 | 0.164584042 | -0.048854269 | 0.030614035 | 0.079468304 | 0.293424451 | 0.715081336 | 0.06067422 |
| sp\|Q96H15\|TIMD4_HUMAN | 3 | 40 | 76 | 33 | 0.00012 | 0.050823102 | 0.255888066 | 0.05070299 | 0.255767954 | 0.205064964 | 0.808162504 | 0.027055409 | 0.051203217 |
| sp\|O00602\|FCN1_HUMAN | 8 | 40 | 76 | 33 | -0.07251 | -0.052782035 | 0.148802349 | 0.019731037 | 0.221315421 | 0.201584384 | 0.977430287 | 0.143429064 | 0.129015031 |
| sp\|Q14162\|SREC_HUMAN | 2 | 36 | 69 | 30 | 0.022962 | 0.115053948 | -0.093354541 | 0.092092063 | -0.116316425 | -0.208408488 | 0.444360302 | 0.408936999 | 0.02839927 |
| sp\|P52306\|GDS1_HUMAN | 19 | 28 | 55 | 25 | 0.183925 | 0.109164614 | 0.420095027 | -0.074760304 | 0.236170108 | 0.310930412 | 0.821378969 | 0.252567265 | 0.047955638 |
| sp\|P60660\|MYL6_HUMAN | 11 | 40 | 76 | 33 | -0.49011 | -0.154119011 | 0.309039331 | 0.335991627 | 0.79914997 | 0.463158342 | 0.003506296 | 3.36E-09 | 0.000108719 |
| sp\|P20700\|LMNB1_HUMAN | 1 | 40 | 76 | 33 | -0.24998 | 0.014514748 | 0.320668412 | 0.264497117 | 0.570650782 | 0.306153665 | 0.008493666 | 7.83E-07 | 0.00382776 |
| sp\|Q9Y608\|LRRF2_HUMAN | 9 | 11 | 21 | 9 | 0.341048 | -0.097706222 | 0.139212294 | -0.43875428 | -0.201835764 | 0.236918516 | 0.137666258 | 0.738801338 | 0.590214368 |
| sp\|Q14240\|IF4A2_HUMAN | 3 | 40 | 76 | 33 | -0.19599 | -0.256649647 | 0.118739488 | -0.060662889 | 0.314726246 | 0.375389135 | 0.864485007 | 0.071735611 | 0.009346751 |
| sp\|P19022\|CADH2_HUMAN | 30 | 40 | 76 | 33 | 0.135251 | 0.138440502 | 0.455316661 | 0.003189092 | 0.320065251 | 0.316876159 | 0.998064863 | 6.25E-06 | 4.53E-07 |
| sp\|Q8TDY8\|IGDC4_HUMAN | 2 | 40 | 76 | 33 | 0.10024 | 0.070848833 | -0.22335083 | -0.029391172 | -0.323590835 | -0.294199663 | 0.855410109 | 8.49E-06 | 4.92E-06 |
| sp\|O60361\|NDK8_HUMAN | 4 | 40 | 76 | 33 | -0.29049 | -0.121598829 | 0.202617747 | 0.168894987 | 0.493111563 | 0.324216577 | 0.255789662 | 0.000530797 | 0.013823594 |
| sp\|O95633\|FSTL3_HUMAN | 13 | 40 | 76 | 33 | -0.19532 | -0.214993708 | 0.246275081 | -0.019677155 | 0.441591635 | 0.461268789 | 0.974477455 | 0.000255357 | 1.39E-05 |
| sp\|Q92876\|KLK6_HUMAN | 4 | 40 | 76 | 33 | 0.012115 | -0.069559526 | -0.136258849 | -0.081674626 | -0.148373949 | -0.066699323 | 0.329570451 | 0.082796604 | 0.520443191 |
| sp\|P35916\|VGFR3_HUMAN | 71 | 40 | 76 | 33 | 0.086589 | 0.148081343 | 0.102202239 | 0.061492807 | 0.015613702 | -0.045879105 | 0.375571351 | 0.956955019 | 0.618072313 |
| sp\|O76074\|PDE5A_HUMAN | 3 | 40 | 76 | 33 | -0.20392 | -0.013519189 | 0.201538094 | 0.190403068 | 0.405460352 | 0.215057284 | 0.162287645 | 0.00415635 | 0.13104673 |
| sp\|P16234\|PGFRA_HUMAN | 73 | 22 | 42 | 18 | -0.03378 | -0.051101527 | 0.275482748 | -0.017324231 | 0.309260045 | 0.326584276 | 0.976838729 | 0.00869848 | 0.001434182 |
| sp\|P51452\|DUS3_HUMAN | 2 | 40 | 76 | 33 | -0.29543 | -0.166255518 | 0.017336205 | 0.129172 | 0.312763724 | 0.183591723 | 0.515870378 | 0.07257142 | 0.311087443 |
| sp\|Q9HCL0\|PCD18_HUMAN | 2 | 40 | 76 | 33 | 0.124124 | 0.126901179 | 0.129932797 | 0.002777013 | 0.005808631 | 0.003031619 | 0.996690789 | 0.990044359 | 0.996537665 |
| sp\|Q96AP7\|ESAM_HUMAN | 4 | 40 | 76 | 33 | -0.08311 | -0.025615623 | 0.190387064 | 0.057493062 | 0.273495749 | 0.216002687 | 0.486010702 | 3.49E-05 | 0.000250125 |
| sp\|O60844\|ZG16_HUMAN | 16 | 40 | 76 | 33 | 0.022533 | 0.008262264 | 0.030817065 | -0.014271211 | 0.008283589 | 0.0225548 | 0.965450494 | 0.991857514 | 0.925822472 |
| sp\|P10644\|KAP0_HUMAN | 16 | 40 | 76 | 33 | -0.49778 | -0.349885652 | -0.201443696 | 0.14789567 | 0.296337626 | 0.148441956 | 0.347422522 | 0.056479544 | 0.39208412 |
| sp\|P56199\|ITA1_HUMAN | 4 | 40 | 76 | 33 | 0.157498 | 0.01144993 | -0.068757974 | -0.146048542 | -0.226256446 | -0.080207904 | 1.96E-05 | 4.08E-08 | 0.045453356 |
| sp\|P13489\|RINI_HUMAN | 1 | 40 | 76 | 33 | -0.14872 | 0.060031465 | 0.26606571 | 0.208752208 | 0.414786453 | 0.206034245 | 0.000801753 | 2.09E-08 | 0.0021248 |
| sp\|O43491\|E41L2_HUMAN | 1 | 11 | 20 | 8 | 0.026131 | 0.171719273 | 0.472508771 | 0.145588477 | 0.446377976 | 0.300789498 | 0.670508189 | 0.099420997 | 0.263500977 |
| sp\|P01834\|IGKC_HUMAN | 13 | 40 | 76 | 33 | -0.1687 | -0.054892413 | 0.146166285 | 0.113805563 | 0.314864261 | 0.201058698 | 0.320885985 | 0.00324515 | 0.047188438 |
| sp\|P13688\|CEAM1_HUMAN | 7 | 40 | 76 | 33 | 0.036614 | 0.052511496 | 0.021972487 | 0.015897929 | -0.01464108 | -0.030539009 | 0.90020086 | 0.94030159 | 0.711936106 |
| sp\|Q9BQS7\|HEPH_HUMAN | 4 | 40 | 76 | 33 | 0.031479 | 0.020668392 | 0.008717484 | -0.010810367 | -0.022761275 | -0.011950908 | 0.972518947 | 0.918344541 | 0.970542847 |
| sp\|Q8N126\|CADM3_HUMAN | 10 | 32 | 62 | 28 | 0.307966 | 0.125594217 | 0.406525397 | -0.182371574 | 0.098559606 | 0.28093118 | 0.04708479 | 0.52209359 | 0.001696211 |
| sp\|Q9UBV8\|PEF1_HUMAN | 19 | 36 | 69 | 30 | 0.127863 | 0.117696908 | 0.31024529 | -0.010165911 | 0.182382472 | 0.192548383 | 0.989410934 | 0.099311026 | 0.038571271 |
| sp\|A6NIZ1\|RP1BL_HUMAN | 30 | 40 | 76 | 33 | -0.36145 | -0.212704666 | 0.059888049 | 0.148750013 | 0.421342728 | 0.272592715 | 0.395307904 | 0.007211533 | 0.06809792 |
| sp\|O15145\|ARPC3_HUMAN | 7 | 40 | 76 | 33 | -0.12035 | -0.190632882 | 0.258049586 | -0.070278863 | 0.378403605 | 0.448682468 | 0.858608795 | 0.051764843 | 0.005622086 |
| sp\|P27482\|CALL3_HUMAN | 10 | 32 | 62 | 26 | -0.11652 | -0.009802256 | 0.045300155 | 0.106720659 | 0.16182307 | 0.055102411 | 0.910014548 | 0.863074791 | 0.978399783 |
| sp\|Q14126\|DSG2_HUMAN | 2 | 40 | 76 | 33 | 0.204375 | 0.103729348 | 0.039773277 | -0.100645303 | -0.164601374 | -0.063956071 | 0.08498793 | 0.011575942 | 0.411262552 |
| sp\|Q13421\|MSLN_HUMAN | 5 | 40 | 76 | 33 | -0.12244 | -0.23253913 | 0.42693469 | -0.110099922 | 0.549373898 | 0.65947382 | 0.375167854 | 3.80E-07 | 1.51E-11 |
| sp\|Q7KZF4\|SND1_HUMAN | 1 | 40 | 76 | 33 | -0.18924 | -0.375333111 | 0.045947273 | -0.186094166 | 0.235186218 | 0.421280384 | 0.297628707 | 0.263461032 | 0.005328666 |
| sp\|Q8IUX7\|AEBP1_HUMAN | 1 | 40 | 76 | 33 | 0.04153 | 0.024696108 | 0.134091923 | -0.016834093 | 0.092561721 | 0.109395814 | 0.952161216 | 0.363433524 | 0.167879001 |
| sp\|Q9BT09\|CNPY3_HUMAN | 17 | 21 | 42 | 19 | -0.28852 | -0.145399939 | 0.266024258 | 0.143117974 | 0.554542171 | 0.411424197 | 0.528742264 | 0.00196216 | 0.009897621 |
| sp\|P22413\|ENPP1_HUMAN | 30 | 40 | 76 | 33 | 0.057423 | 0.078061397 | 0.128737701 | 0.020638785 | 0.071315088 | 0.050676303 | 0.798333422 | 0.161311657 | 0.307221911 |
| sp\|P51665\|PSMD7_HUMAN | 1 | 23 | 42 | 17 | -0.43375 | -0.038193587 | 0.278569106 | 0.395554768 | 0.712317461 | 0.316762693 | 0.02125533 | 0.000438647 | 0.126261239 |
| sp\|O60664\|PLIN3_HUMAN | 39 | 40 | 76 | 33 | 0.064237 | 0.032217188 | 0.509741218 | -0.032019619 | 0.44550441 | 0.477524029 | 0.913049853 | 1.80E-05 | 2.12E-07 |
| sp\|Q9Y376\|CAB39_HUMAN | 11 | 40 | 76 | 32 | -0.10346 | -0.132407602 | 0.244913317 | -0.028951842 | 0.348369078 | 0.37732092 | 0.970240492 | 0.056155937 | 0.014681723 |
| sp\|O95678\|K2C75_HUMAN | 5 | 40 | 76 | 33 | -0.56484 | -0.739072292 | -0.728318561 | -0.174229307 | -0.163475576 | 0.010753731 | 0.697921937 | 0.803489359 | 0.998793178 |
| sp\|O43790\|KRT86_HUMAN | 16 | 8 | 13 | 6 | -0.31126 | 0.413582215 | -0.394629535 | 0.724844667 | -0.083367083 | -0.80821175 | 0.403353867 | 0.99137249 | 0.392752833 |
| sp\|Q9UKM7\|MA1B1_HUMAN | 7 | 34 | 62 | 27 | -0.00242 | 0.052613401 | -0.02542575 | 0.055036684 | -0.023002468 | -0.078039152 | 0.624424272 | 0.944912188 | 0.4458689 |

| | | | | | | | | | | | | | |
|---|---|---|---|---|---|---|---|---|---|---|---|---|---|
| sp\|Q96KP4\|CNDP2_HUMAN | 8 | 40 | 76 | 33 | -0.22969 | -0.138809183 | 0.218756825 | 0.090878401 | 0.448444409 | 0.357566008 | 0.659310351 | 0.001393374 | 0.004599584 |
| sp\|P08069\|IGF1R_HUMAN | 1 | 7 | 14 | 6 | 0.285919 | 0.061705322 | -0.104236424 | -0.224213936 | -0.390155682 | -0.165941746 | 0.425550539 | 0.178873269 | 0.650849089 |
| sp\|P12109\|CO6A1_HUMAN | 6 | 40 | 76 | 33 | 0.061964 | 0.114923749 | 0.268038152 | 0.052959621 | 0.206074024 | 0.153114403 | 0.533778577 | 0.00203208 | 0.011908725 |
| sp\|Q15181\|IPYR_HUMAN | 2 | 8 | 13 | 6 | -0.14418 | -0.160851385 | 0.152778967 | -0.016674415 | 0.296955937 | 0.313630352 | 0.993751702 | 0.273314467 | 0.182570367 |
| sp\|P06280\|AGAL_HUMAN | 3 | 40 | 76 | 33 | -0.03053 | 0.004813408 | 0.260779127 | 0.035341819 | 0.291307539 | 0.255965719 | 0.751590395 | 6.43E-06 | 7.78E-06 |
| sp\|P51114\|FXR1_HUMAN | 55 | 4 | 6 | 3 | -0.03556 | -0.055119396 | 0.111939283 | -0.019557437 | 0.147501242 | 0.167058679 | 0.999835579 | 0.993345938 | 0.990061708 |
| sp\|P01871\|IGHM_HUMAN | 19 | 40 | 76 | 33 | -0.20809 | 0.030150366 | 0.01682015 | 0.238239804 | 0.224909587 | -0.013330216 | 0.05685049 | 0.168127293 | 0.991911667 |
| sp\|P04745\|AMY1_HUMAN | 1 | 40 | 76 | 33 | 0.138923 | -1.88E-05 | -0.228560639 | -0.138942207 | -0.367484058 | -0.228541852 | 0.228280158 | 0.001156028 | 0.032101831 |
| sp\|Q13275\|SEM3F_HUMAN | 21 | 40 | 76 | 33 | 0.103232 | 0.087115814 | -0.07226834 | -0.016116233 | -0.175500387 | -0.159384155 | 0.855359701 | 1.06E-05 | 6.37E-06 |
| sp\|P55157\|MTP_HUMAN | 45 | 36 | 69 | 30 | -0.014 | -0.113998281 | -0.04727904 | -0.099999373 | -0.033280132 | 0.066719241 | 0.375901952 | 0.927064475 | 0.678722097 |
| sp\|Q15185\|TEBP_HUMAN | 3 | 40 | 76 | 33 | -0.17215 | -0.137377268 | 0.23496243 | 0.034774246 | 0.407113944 | 0.372339698 | 0.944293123 | 0.005830252 | 0.004257446 |
| sp\|Q6P988\|NOTUM_HUMAN | 1 | 36 | 69 | 30 | 0.038547 | 0.113288839 | 0.128192851 | 0.07474221 | 0.089646223 | 0.014904013 | 0.585602184 | 0.587142639 | 0.981260731 |
| sp\|P62136\|PP1A_HUMAN | 29 | 40 | 76 | 33 | -0.24966 | -0.125144348 | 0.238888386 | 0.124513442 | 0.488546176 | 0.364032734 | 0.296959056 | 8.62E-06 | 0.000207437 |
| sp\|Q16543\|CDC37_HUMAN | 2 | 37 | 69 | 29 | -0.48387 | -0.485325199 | 0.045551271 | -0.001458647 | 0.529417823 | 0.53087647 | 0.999931951 | 0.00339362 | 0.000843598 |
| sp\|Q14554\|PDIA5_HUMAN | 2 | 40 | 76 | 33 | 0.074756 | 0.069945296 | 0.160942702 | -0.004810238 | 0.086187169 | 0.090997407 | 0.999044386 | 0.809531403 | 0.741231859 |
| sp\|Q14956\|GPNMB_HUMAN | 4 | 40 | 76 | 33 | 0.096983 | 0.063303223 | 0.140967704 | -0.033679633 | 0.043984847 | 0.077664481 | 0.714332904 | 0.673251353 | 0.212130708 |
| sp\|Q6ZMJ2\|SCAR5_HUMAN | 1 | 40 | 76 | 33 | 0.078434 | 0.073818597 | 0.338054146 | -0.004615333 | 0.259620217 | 0.26423555 | 0.996745607 | 0.001290519 | 0.000182454 |
| sp\|P21757\|MSRE_HUMAN | 6 | 36 | 70 | 30 | -0.21243 | -0.092222846 | 0.468274661 | 0.12020725 | 0.680704756 | 0.560497507 | 0.513368814 | 2.33E-06 | 1.07E-05 |
| sp\|P50238\|CRIP1_HUMAN | 4 | 37 | 69 | 30 | -0.33527 | -0.117259064 | 0.36369664 | 0.218015742 | 0.698971446 | 0.480955704 | 0.035731606 | 1.99E-09 | 2.77E-06 |
| sp\|P08217\|CEL2A_HUMAN | 22 | 32 | 62 | 27 | -0.00298 | -0.006345959 | 0.156465579 | -0.003363438 | 0.1594481 | 0.162811538 | 0.999537813 | 0.489421744 | 0.38516639 |
| sp\|P61769\|B2MG_HUMAN | 3 | 40 | 76 | 33 | -0.1413 | -0.094636684 | 0.484946965 | 0.046662305 | 0.626245955 | 0.57958365 | 0.872500639 | 3.93E-07 | 1.23E-07 |
| sp\|Q9NZ53\|PDXL2_HUMAN | 1 | 8 | 14 | 5 | -0.02612 | 0.115828788 | 0.010711159 | 0.141946443 | 0.036828814 | -0.105117629 | 0.369747082 | 0.958593602 | 0.666521044 |
| sp\|Q00872\|MYPC1_HUMAN | 3 | 11 | 19 | 9 | -0.71091 | -0.856932509 | 0.238694321 | -0.146026795 | 0.949600035 | 1.09562683 | 0.937685842 | 0.160731444 | 0.054907314 |
| sp\|P07195\|LDHB_HUMAN | 1 | 40 | 76 | 33 | 0.077285 | 0.047021021 | 0.280201036 | -0.030264407 | 0.202915608 | 0.233180015 | 0.868744478 | 0.015215863 | 0.001056903 |
| sp\|Q15691\|MARE1_HUMAN | 7 | 40 | 76 | 33 | -0.12902 | -0.277771848 | 0.068094137 | -0.148748549 | 0.197117436 | 0.345865985 | 0.446811404 | 0.377421342 | 0.024453249 |
| sp\|P12110\|CO6A2_HUMAN | 10 | 40 | 76 | 33 | -0.03479 | -0.021198849 | 0.293710243 | 0.01359111 | 0.328500202 | 0.314909092 | 0.970115084 | 1.69E-05 | 3.18E-06 |
| sp\|P61158\|ARP3_HUMAN | 10 | 40 | 76 | 33 | -0.16197 | -0.218599064 | 0.137933558 | -0.056624822 | 0.299907799 | 0.356532622 | 0.889483199 | 0.108912997 | 0.019864827 |
| sp\|P05089\|ARGI1_HUMAN | 3 | 29 | 56 | 25 | -0.38958 | -0.087021405 | 0.163262082 | 0.302555511 | 0.552838999 | 0.250283488 | 0.116642936 | 0.007665498 | 0.260790608 |
| sp\|Q96PD2\|DCBD2_HUMAN | 6 | 40 | 76 | 33 | -0.02166 | -0.014235685 | 0.188815224 | 0.00742869 | 0.210479599 | 0.203050909 | 0.979324589 | 2.91E-05 | 5.14E-06 |
| sp\|P10321\|HLAC_HUMAN | 1 | 40 | 76 | 33 | -0.14716 | -0.017533823 | 0.085248092 | 0.129623883 | 0.232405798 | 0.102781915 | 0.351951029 | 0.101639094 | 0.560294668 |
| sp\|P17693\|HLAG_HUMAN | 9 | 36 | 69 | 30 | -0.1734 | 0.114994978 | 0.157807793 | 0.288391417 | 0.331204232 | 0.042812815 | 0.014047847 | 0.020114867 | 0.916662639 |
| sp\|Q9NS98\|SEM3G_HUMAN | 5 | 40 | 76 | 33 | 0.040029 | 0.002287706 | -0.05527108 | -0.037740875 | -0.095299661 | -0.057558786 | 0.553435584 | 0.078083144 | 0.301075819 |
| sp\|P15121\|ALDR_HUMAN | 4 | 40 | 76 | 33 | -0.02286 | -0.01534818 | 0.385379253 | 0.00750781 | 0.408235244 | 0.400727433 | 0.996151146 | 0.000670056 | 0.000145749 |
| sp\|Q99538\|LGMN_HUMAN | 2 | 36 | 69 | 30 | -0.1791 | -0.093305135 | 0.030269891 | 0.085795672 | 0.209370698 | 0.123575026 | 0.425543546 | 0.032687315 | 0.211334534 |
| sp\|Q16774\|KGUA_HUMAN | 21 | 4 | 7 | 3 | 0.198408 | 0.248641264 | 0.038798939 | 0.050233671 | -0.159608654 | -0.209842324 | 0.932173192 | 0.631275314 | 0.394273881 |
| sp\|O95881\|TXD12_HUMAN | 2 | 18 | 35 | 14 | -0.29215 | -0.114381231 | 0.354825583 | 0.177768452 | 0.646975266 | 0.469206814 | 0.640085434 | 0.025241553 | 0.081063813 |
| sp\|P19883\|FST_HUMAN | 7 | 18 | 34 | 14 | 0.106521 | 0.068844881 | 0.425945702 | -0.037675947 | 0.319424874 | 0.357100822 | 0.9570214 | 0.131080703 | 0.043691577 |
| sp\|Q16775\|GLO2_HUMAN | 89 | 37 | 69 | 30 | -0.08396 | -0.050387976 | 0.168723805 | 0.033573555 | 0.252685336 | 0.219111781 | 0.894527404 | 0.015706469 | 0.019248164 |
| sp\|P40967\|PMEL_HUMAN | 5 | 40 | 76 | 33 | 0.040215 | 0.032679788 | -0.258077848 | -0.007535021 | -0.298292657 | -0.290757636 | 0.992034051 | 0.000341051 | 7.37E-05 |
| sp\|P08138\|TNR16_HUMAN | 3 | 28 | 55 | 24 | -0.20965 | -0.3027074 | 0.51063215 | -0.093056964 | 0.720282586 | 0.81333955 | 0.804667487 | 0.000277312 | 2.80E-06 |
| sp\|Q9NRV9\|HEBP1_HUMAN | 18 | 22 | 41 | 19 | -0.45032 | -0.109412515 | 0.427862065 | 0.340904357 | 0.878178937 | 0.53727458 | 0.112873168 | 9.78E-05 | 0.008927888 |
| sp\|P07910\|HNRPC_HUMAN | 4 | 40 | 75 | 32 | -0.98933 | -0.586838085 | 0.246231869 | 0.402490012 | 1.235559966 | 0.833069955 | 0.096541044 | 1.37E-06 | 0.000294286 |
| sp\|O43505\|B4GA1_HUMAN | 7 | 40 | 76 | 33 | 0.182003 | 0.149764317 | -0.004435588 | -0.032238345 | -0.186438251 | -0.154199905 | 0.46482784 | 2.05E-07 | 1.22E-06 |
| sp\|P04350\|TBB4A_HUMAN | 3 | 40 | 76 | 33 | -0.28811 | -0.085002444 | 0.24397106 | 0.203104718 | 0.532078222 | 0.328973503 | 0.201801628 | 0.000769344 | 0.027020005 |
| sp\|P19105\|ML12A_HUMAN | 3 | 40 | 76 | 33 | -0.53211 | -0.175274517 | 0.327712707 | 0.356830647 | 0.859817871 | 0.502987224 | 0.002410817 | 6.56E-10 | 4.02E-05 |
| sp\|P10916\|MLRV_HUMAN | 2 | 7 | 13 | 6 | -0.96547 | -1.27577837 | -0.140130893 | -0.310311055 | 0.825336422 | 1.135647478 | 0.895375274 | 0.580565428 | 0.282540895 |
| sp\|Q02045\|MYL5_HUMAN | 1 | 4 | 6 | 2 | -0.38909 | -0.418351805 | 2.417389276 | -0.029264981 | 2.8064761 | 2.835741082 | 0.999464394 | 0.119376447 | 0.093552122 |
| sp\|Q96A32\|MLRS_HUMAN | 2 | 40 | 76 | 33 | -0.34127 | -0.484536014 | -0.397720874 | -0.143264267 | -0.056449128 | 0.086815139 | 0.145625425 | 0.810452826 | 0.532748119 |
| sp\|Q8NC51\|PAIRB_HUMAN | 5 | 11 | 20 | 9 | -0.04239 | -0.079127113 | 0.723601283 | -0.036737128 | 0.765991267 | 0.802728395 | 0.988639025 | 0.04306762 | 0.015094349 |
| sp\|P36222\|CH3L1_HUMAN | 8 | 40 | 76 | 33 | -0.17093 | -0.254403153 | 0.784653112 | -0.083474729 | 0.955581536 | 1.039056265 | 0.84676875 | 1.74E-06 | 5.61E-09 |
| sp\|Q96C86\|DCPS_HUMAN | 7 | 33 | 62 | 27 | -0.20252 | 0.012383461 | 0.320400936 | 0.214904958 | 0.522922433 | 0.308017475 | 0.075965551 | 6.05E-05 | 0.010881893 |
| sp\|P07988\|PSPB_HUMAN | 22 | 40 | 76 | 33 | -0.47014 | -0.319693324 | 1.125678494 | 0.150450673 | 1.595822491 | 1.445371819 | 0.538445244 | 3.92E-14 | 3.87E-14 |
| sp\|Q9P0L0\|VAPA_HUMAN | 25 | 32 | 63 | 28 | -0.14743 | -0.088737327 | 0.323365454 | 0.058689131 | 0.470791911 | 0.41210278 | 0.845331025 | 0.000887112 | 0.000918859 |
| sp\|O75131\|CPNE3_HUMAN | 2 | 32 | 63 | 27 | -0.16274 | -0.032107967 | 0.421159082 | 0.130630098 | 0.583897147 | 0.453267049 | 0.521248895 | 0.000265955 | 0.001467471 |
| sp\|Q8IZ83\|A16A1_HUMAN | 7 | 40 | 76 | 33 | -0.51388 | -0.025717117 | 0.378303426 | 0.488165804 | 0.892186347 | 0.404020542 | 2.12E-07 | 6.87E-14 | 6.04E-05 |
| sp\|Q15056\|IF4H_HUMAN | 1 | 14 | 28 | 12 | -0.24649 | -0.243835957 | 0.13977001 | 0.002657522 | 0.386263489 | 0.383605967 | 0.999941426 | 0.431732819 | 0.342617177 |
| sp\|Q96P70\|IPO9_HUMAN | 2 | 22 | 41 | 18 | 0.287471 | 0.284573063 | 0.39187543 | -0.002898113 | 0.104404254 | 0.107302367 | 0.999527024 | 0.656021986 | 0.570462865 |
| sp\|Q8WXD2\|SCG3_HUMAN | 3 | 40 | 76 | 33 | 0.002514 | -0.110773088 | -0.132451976 | -0.113287424 | -0.134965713 | -0.021678288 | 0.033083906 | 0.035419945 | 0.892779025 |
| sp\|Q9NSC7\|SIA7A_HUMAN | 56 | 40 | 76 | 33 | -0.02484 | -0.07628798 | -0.088990172 | -0.05144706 | -0.064149252 | -0.012702192 | 0.745610752 | 0.729909169 | 0.984365618 |
| sp\|Q9HBR0\|S38AA_HUMAN | 5 | 40 | 76 | 33 | 0.116567 | 0.126571117 | 0.277118233 | 0.00870449 | 0.160551607 | 0.151847117 | 0.985156755 | 0.033744017 | 0.02149495 |
| sp\|E9PAV3\|NACAM_HUMAN | 13 | 11 | 21 | 10 | -0.1475 | -0.236885983 | 0.449053046 | -0.08938958 | 0.596549449 | 0.685939028 | 0.900042172 | 0.044401323 | 0.00651159 |
| sp\|Q01973\|ROR1_HUMAN | 12 | 40 | 76 | 33 | 0.073323 | -0.059976579 | 0.088424742 | -0.133299585 | 0.015101736 | 0.148401321 | 0.025995845 | 0.966995133 | 0.019075546 |
| sp\|P18850\|ATF6A_HUMAN | 1 | 40 | 76 | 33 | 0.094232 | 0.056048176 | -0.05072649 | -0.038183401 | -0.144958067 | -0.106774666 | 0.559376227 | 0.00410819 | 0.021160094 |
| sp\|P27105\|STOM_HUMAN | 2 | 40 | 76 | 33 | -1.91668 | -1.578343645 | -1.116753805 | 0.338334633 | 0.799924473 | 0.46158984 | 0.05724129 | 3.45E-05 | 0.010114789 |
| sp\|Q9BYZ8\|REG4_HUMAN | 3 | 33 | 63 | 27 | -0.00254 | -0.054221327 | 0.057423138 | -0.051679585 | 0.05996488 | 0.111644465 | 0.594639507 | 0.618712714 | 0.125187083 |
| sp\|Q13308\|PTK7_HUMAN | 5 | 40 | 76 | 33 | -0.23527 | -0.209301045 | 0.105816068 | 0.025965399 | 0.341082512 | 0.315117113 | 0.893633543 | 6.55E-06 | 2.60E-06 |
| sp\|P78371\|TCPB_HUMAN | 9 | 36 | 70 | 30 | 0.091233 | 0.073254374 | 0.448830219 | -0.0179783 | 0.357597545 | 0.375575846 | 0.990700516 | 0.084158854 | 0.031236327 |
| sp\|Q01082\|SPTB2_HUMAN | 15 | 36 | 68 | 30 | 0.193872 | 0.308558679 | 0.521431806 | 0.114686758 | 0.327559885 | 0.212873127 | 0.811978141 | 0.310895498 | 0.53178239 |
| sp\|Q8N4A0\|GALT4_HUMAN | 12 | 18 | 35 | 16 | 0.071235 | 0.079379365 | 0.034101614 | 0.00814437 | -0.037133381 | -0.045277751 | 0.991520354 | 0.881731149 | 0.785019767 |
| sp\|Q6UXH1\|CREL2_HUMAN | 2 | 40 | 76 | 33 | -0.23183 | -0.141414741 | 0.41130974 | 0.090410547 | 0.643135028 | 0.552724481 | 0.583440742 | 8.92E-08 | 2.12E-07 |
| sp\|O15204\|ADEC1_HUMAN | 3 | 40 | 76 | 33 | -0.08493 | 0.042535848 | 0.021432253 | 0.127465042 | 0.106361448 | -0.021103594 | 0.049753172 | 0.231156994 | 0.928243433 |
| sp\|P19237\|TNNI1_HUMAN | 1 | 4 | 6 | 3 | 0.092164 | 0.081769679 | 1.416218122 | -0.010394099 | 1.324054344 | 1.334448444 | 0.999909314 | 0.38647283 | 0.33027147 |
| sp\|Q9NQC3\|RTN4_HUMAN | 1 | 7 | 14 | 6 | -0.82595 | -0.231590628 | 0.559954097 | 0.594363446 | 1.385908172 | 0.791544725 | 0.253980157 | 0.011450194 | 0.120751717 |
| sp\|Q92956\|TNR14_HUMAN | 24 | 36 | 69 | 30 | -0.38588 | -0.265533911 | 0.227777978 | 0.120345136 | 0.613657025 | 0.493311889 | 0.571987473 | 0.000104147 | 0.000459445 |
| sp\|Q9Y251\|HPSE_HUMAN | 4 | 40 | 76 | 33 | -0.24703 | -0.171095118 | 0.216799873 | 0.075939749 | 0.463834741 | 0.387894992 | 0.831739113 | 0.01073367 | 0.017342961 |
| sp\|Q9C0C9\|UBE2O_HUMAN | 6 | 21 | 42 | 19 | -0.48491 | -0.196613484 | 0.18681422 | 0.288294954 | 0.671722658 | 0.383427704 | 0.11331865 | 0.000444085 | 0.029532102 |
| sp\|P82987\|ATL3_HUMAN | 5 | 37 | 69 | 30 | 0.043084 | 0.03604677 | 0.297524406 | -0.00703693 | 0.254440705 | 0.261477636 | 0.995259605 | 0.016823103 | 0.004648189 |
| sp\|P78417\|GSTO1_HUMAN | 2 | 40 | 76 | 33 | -0.16819 | -0.055491529 | 0.398024503 | 0.112698792 | 0.566214824 | 0.453516032 | 0.298494486 | 1.49E-08 | 2.81E-07 |
| sp\|Q58FF6\|H90B4_HUMAN | 8 | 40 | 76 | 33 | -0.09403 | -0.110335329 | 0.271801544 | -0.016308535 | 0.365828338 | 0.382136873 | 0.992217272 | 0.07145385 | 0.026593591 |
| sp\|P23229\|ITA6_HUMAN | 1 | 40 | 76 | 33 | -0.17008 | 0.064780556 | -0.041385241 | 0.234857951 | 0.128692154 | -0.106165797 | 0.000250971 | 0.160378988 | 0.204139805 |
| sp\|P22626\|ROA2_HUMAN | 2 | 33 | 62 | 27 | -0.29832 | 0.005801421 | 0.529062623 | 0.304121338 | 0.82738254 | 0.523261202 | 0.393655295 | 0.010477352 | 0.093564313 |
| sp\|Q9BS26\|ERP44_HUMAN | 7 | 40 | 76 | 33 | 0.109996 | 0.097092172 | 0.079550304 | -0.012903568 | -0.030445436 | -0.017541868 | 0.960999921 | 0.858456255 | 0.937505796 |
| sp\|Q9UP52\|TFR2_HUMAN | 1 | 40 | 76 | 33 | -0.12758 | -0.296385855 | -0.173759175 | -0.168805094 | -0.046178414 | 0.12262668 | 0.360545353 | 0.948171987 | 0.621641398 |
| sp\|Q9BZD6\|TMG4_HUMAN | 2 | 8 | 14 | 6 | 0.144332 | 0.188924986 | 0.053338706 | 0.044592718 | -0.090993562 | -0.13558628 | 0.889722717 | 0.722767945 | 0.422179871 |
| sp\|P13010\|XRCC5_HUMAN | 2 | 14 | 27 | 12 | -0.63524 | -0.462883449 | 0.463749779 | 0.172353682 | 1.09898691 | 0.926633228 | 0.84367761 | 0.012468985 | 0.01759554 |
| sp\|P28845\|DHI1_HUMAN | 4 | 40 | 76 | 33 | 0.038297 | 0.086287847 | 0.086566086 | 0.04799096 | 0.048269199 | 0.000278239 | 0.65834263 | 0.746635688 | 0.999987608 |
| sp\|P28072\|PSB6_HUMAN | 3 | 40 | 76 | 33 | -0.11912 | 0.008246942 | 0.468511088 | 0.12737143 | 0.587635576 | 0.460264146 | 0.275715275 | 7.33E-08 | 1.87E-06 |
| sp\|Q9NS62\|THSD1_HUMAN | 19 | 40 | 76 | 33 | 0.141189 | 0.133797918 | -0.06826934 | -0.00739151 | -0.209458768 | -0.202067258 | 0.989808184 | 0.00464321 | 0.00182725 |
| sp\|Q6JBY9\|CPZIP_HUMAN | 4 | 24 | 48 | 21 | -0.14596 | 0.069579453 | 0.611494518 | 0.215535305 | 0.75745037 | 0.541915064 | 0.285666616 | 6.60E-05 | 0.001243103 |
| sp\|Q6UWB1\|I27RA_HUMAN | 6 | 26 | 48 | 21 | -0.28213 | -0.262645597 | -0.122983081 | 0.019482094 | 0.15914461 | 0.139662515 | 0.972696578 | 0.286551311 | 0.297860439 |
| sp\|P09681\|GIP_HUMAN | 11 | 7 | 14 | 6 | -0.07744 | -0.145250558 | 0.32228986 | -0.067812927 | 0.399727491 | 0.467540418 | 0.952417757 | 0.327374874 | 0.147426251 |
| sp\|P58335\|ANTR2_HUMAN | 5 | 40 | 76 | 33 | 0.153833 | -0.007171464 | -0.055278395 | -0.161004511 | -0.209111442 | -0.048106931 | 0.010626505 | 0.005234741 | 0.689464363 |
| sp\|Q9BXS4\|TMM59_HUMAN | 5 | 7 | 14 | 7 | -0.22837 | -0.048258065 | 0.356518962 | 0.180113138 | 0.584890165 | 0.404777027 | 0.465868653 | 0.006717758 | 0.032450446 |
| sp\|Q58FF8\|H90B2_HUMAN | 22 | 21 | 41 | 17 | -0.23441 | -0.377620772 | 0.502730232 | -0.143211766 | 0.737139239 | 0.880351004 | 0.78533852 | 0.017193761 | 0.000856455 |
| sp\|Q9H479\|FN3K_HUMAN | 1 | 40 | 76 | 33 | -0.30479 | -0.149940058 | 0.110761437 | 0.154854579 | 0.415556074 | 0.260701495 | 0.402578156 | 0.012734108 | 0.107203735 |
| sp\|Q7LGC8\|CHST3_HUMAN | 6 | 40 | 76 | 33 | 0.142589 | 0.101524931 | 0.074595398 | -0.041063948 | -0.06799348 | -0.026929533 | 0.358733723 | 0.146432272 | 0.677249244 |
| sp\|Q9UMS6\|SYNP2_HUMAN | 6 | 4 | 6 | 3 | 0.070832 | 0.157863916 | 1.022687643 | 0.087031581 | 0.951855308 | 0.864823726 | 0.984845154 | 0.314683029 | 0.326913269 |
| sp\|Q08345\|DDR1_HUMAN | 1 | 40 | 76 | 33 | -0.02788 | -0.042936385 | 0.080422842 | -0.015057791 | 0.108301436 | 0.123359227 | 0.92252317 | 0.06095754 | 0.010691857 |
| sp\|P80404\|GABT_HUMAN | 4 | 33 | 62 | 27 | -0.707 | -0.535257846 | -0.147132076 | 0.171747154 | 0.559872924 | 0.38812577 | 0.70190126 | 0.079927858 | 0.211003896 |
| sp\|P43034\|LIS1_HUMAN | 1 | 40 | 76 | 33 | -0.15827 | -0.176400725 | 0.164182425 | -0.018131521 | 0.322451628 | 0.34058315 | 0.984711488 | 0.03876068 | 0.010522496 |
| sp\|Q13126\|MTAP_HUMAN | 7 | 40 | 76 | 33 | -0.05922 | 0.011731498 | 0.229102443 | 0.070950766 | 0.28832171 | 0.217370944 | 0.530061548 | 0.001116882 | 0.006721581 |
| sp\|Q14165\|MLEC_HUMAN | 4 | 40 | 76 | 33 | -0.11526 | -0.137253954 | 0.150930162 | -0.021992606 | 0.26619151 | 0.288184116 | 0.94323856 | 0.003764041 | 0.00029575 |
| sp\|Q9NTK5\|OLA1_HUMAN | 1 | 40 | 76 | 33 | -0.27379 | -0.129032908 | 0.213293717 | 0.144760722 | 0.487087347 | 0.342326625 | 0.293146799 | 0.000137884 | 0.003166347 |
| sp\|P00352\|AL1A1_HUMAN | 11 | 40 | 76 | 33 | -0.36371 | -0.058949526 | 0.348630076 | 0.304758087 | 0.712337689 | 0.407579602 | 0.010276798 | 1.67E-07 | 0.000893401 |
| sp\|Q8IZA0\|K319L_HUMAN | 17 | 40 | 76 | 33 | 0.017313 | 0.064007174 | 0.236128393 | 0.046694147 | 0.218815366 | 0.172121219 | 0.53864319 | 0.000174048 | 0.000980966 |

| | | | | | | | | | | | | | |
|---|---|---|---|---|---|---|---|---|---|---|---|---|---|
| sp\|O15212\|PFD6_HUMAN | 7 | 15 | 27 | 11 | -0.06699 | 0.03833721 | 0.236498748 | 0.105329934 | 0.303491473 | 0.198161539 | 0.68570348 | 0.137488222 | 0.344791021 |
| sp\|Q9Y315\|DEOC_HUMAN | 5 | 20 | 42 | 19 | -0.12485 | -0.04611837 | 0.289204707 | 0.078726721 | 0.414049797 | 0.335323076 | 0.6320913 | 0.000310634 | 0.00073994 |
| sp\|P54687\|BCAT1_HUMAN | 2 | 4 | 7 | 2 | -0.2772 | 0.264303649 | 0.26273122 | 0.5414996 | 0.53992717 | -0.001572429 | 0.710250746 | 0.834254402 | 0.999998172 |
| sp\|P45974\|UBP5_HUMAN | 1 | 40 | 76 | 33 | -0.33605 | -0.18681597 | 0.146963602 | 0.149235087 | 0.483014659 | 0.333779572 | 0.237738684 | 6.85E-05 | 0.002418295 |
| sp\|Q13797\|ITA9_HUMAN | 5 | 40 | 76 | 33 | 0.136166 | 0.132441362 | 0.134442231 | -0.003724347 | -0.001723479 | 0.002000868 | 0.998380862 | 0.999760593 | 0.999589442 |
| sp\|O60240\|PLIN1_HUMAN | 2 | 22 | 42 | 17 | 0.042589 | 0.228680289 | 0.273357289 | 0.186091392 | 0.230768393 | 0.044677 | 0.333308609 | 0.325686965 | 0.947420162 |
| sp\|P21980\|TGM2_HUMAN | 39 | 29 | 56 | 24 | -0.45219 | -0.130048732 | 0.119388233 | 0.322142091 | 0.571579056 | 0.249436964 | 0.028649961 | 0.0006518 | 0.147674285 |
| sp\|P27930\|IL1R2_HUMAN | 36 | 40 | 76 | 33 | 0.060974 | 0.051165983 | 0.073007567 | -0.009808178 | 0.012033406 | 0.021841585 | 0.989715084 | 0.989320563 | 0.956002761 |
| sp\|Q8NBJ4\|GOLM1_HUMAN | 17 | 40 | 76 | 33 | -0.15074 | 0.014918011 | 0.493037598 | 0.165658391 | 0.643777977 | 0.478119587 | 0.239954826 | 1.70E-06 | 6.54E-05 |
| sp\|P09497\|CLCB_HUMAN | 1 | 12 | 20 | 9 | 0.228592 | 0.099497171 | 0.472928255 | -0.12909524 | 0.244335845 | 0.373431085 | 0.790182677 | 0.564068353 | 0.208790904 |
| sp\|Q13287\|NMI_HUMAN | 62 | 14 | 28 | 11 | 0.044521 | 0.196193332 | 0.364301984 | 0.151672292 | 0.319780944 | 0.168108653 | 0.520057385 | 0.155083726 | 0.507013298 |
| sp\|Q9H0U4\|RAB1B_HUMAN | 14 | 40 | 76 | 33 | -0.2034 | -0.211262644 | 0.330147985 | -0.007866449 | 0.53354418 | 0.541410629 | 0.997503996 | 0.000632995 | 7.76E-05 |
| sp\|Q9H173\|SIL1_HUMAN | 6 | 40 | 76 | 33 | -0.40544 | -0.34347171 | 0.163651291 | 0.061968537 | 0.569091538 | 0.507123001 | 0.827289975 | 4.47E-05 | 4.06E-05 |
| sp\|Q96FW1\|OTUB1_HUMAN | 12 | 40 | 76 | 33 | -0.23508 | -0.179648592 | -0.010190368 | 0.055435916 | 0.22489414 | 0.169458224 | 0.826146351 | 0.119432287 | 0.213407208 |
| sp\|Q9Y2T3\|GUAD_HUMAN | 6 | 40 | 76 | 33 | 0.00446 | 0.043389193 | 0.196049296 | 0.038928879 | 0.191588983 | 0.152660104 | 0.84349237 | 0.062779302 | 0.105570983 |
| sp\|P12882\|MYH1_HUMAN | 16 | 26 | 48 | 22 | -0.00323 | -0.025159004 | 0.025419898 | -0.02193098 | 0.028647922 | 0.050578902 | 0.987132463 | 0.984506713 | 0.940282905 |
| sp\|Q8N1Q1\|CAH13_HUMAN | 19 | 36 | 69 | 31 | -0.07284 | -0.169053384 | 0.084119058 | -0.096209658 | 0.156962785 | 0.253172443 | 0.780313021 | 0.628925254 | 0.216351567 |
| sp\|A6NMZ7\|CO6A6_HUMAN | 80 | 15 | 28 | 13 | 0.176156 | 0.123527063 | 0.252718359 | -0.052628462 | 0.076562834 | 0.129191296 | 0.921612842 | 0.884207402 | 0.641828317 |
| sp\|P38571\|LICH_HUMAN | 5 | 36 | 69 | 30 | -0.04576 | 0.031391707 | 0.297574318 | 0.077153876 | 0.343336488 | 0.266182611 | 0.497825079 | 0.000155561 | 0.001043274 |
| sp\|P58546\|MTPN_HUMAN | 10 | 40 | 76 | 33 | -0.20172 | -0.068257301 | 0.130465041 | 0.133459433 | 0.332181774 | 0.198722342 | 0.507223541 | 0.058599352 | 0.269131276 |
| sp\|O60506\|HNRPQ_HUMAN | 113 | 30 | 55 | 24 | -0.25623 | -0.252453972 | 0.240241591 | 0.003773531 | 0.496469095 | 0.492695563 | 0.999458059 | 0.002551562 | 0.000706624 |
| sp\|Q9BZQ8\|NIBA1_HUMAN | 22 | 14 | 28 | 11 | -0.32157 | -0.187242569 | 0.151883883 | 0.134327624 | 0.473454076 | 0.339126452 | 0.695444002 | 0.060205658 | 0.151228852 |
| sp\|P11215\|ITAM_HUMAN | 3 | 14 | 28 | 13 | 0.00194 | -0.089575649 | 0.478807924 | -0.091515591 | 0.476867982 | 0.568383573 | 0.779555169 | 0.01182027 | 0.000445721 |
| sp\|Q9NZL9\|MAT2B_HUMAN | 2 | 37 | 69 | 30 | -0.08915 | 0.139694605 | 0.432072963 | 0.228844794 | 0.521223151 | 0.292378358 | 0.029282793 | 9.08E-06 | 0.007230343 |
| sp\|P11940\|PABP1_HUMAN | 9 | 26 | 49 | 20 | 0.117483 | 0.122978417 | 0.34773759 | 0.005495079 | 0.230254251 | 0.224759173 | 0.997329779 | 0.050208556 | 0.028639369 |
| sp\|P16885\|PLCG2_HUMAN | 4 | 7 | 14 | 6 | -0.56219 | -0.466608725 | 0.672622371 | 0.09558466 | 1.234815756 | 1.139231096 | 0.959713325 | 0.018968128 | 0.013349851 |
| sp\|O00506\|STK25_HUMAN | 9 | 10 | 21 | 9 | -0.07104 | -0.027220683 | 0.716880196 | 0.043818885 | 0.787919764 | 0.744100879 | 0.986181109 | 0.056172001 | 0.034570397 |
| sp\|P17655\|CAN2_HUMAN | 4 | 40 | 76 | 33 | -0.14048 | -0.008432287 | 0.197218374 | 0.132044122 | 0.337694783 | 0.205650661 | 0.015999723 | 5.87E-08 | 0.000214288 |
| sp\|Q8NHL6\|LIRB1_HUMAN | 15 | 40 | 76 | 33 | 0.06448 | 0.015449345 | 0.097548003 | -0.049030801 | 0.033067857 | 0.082098658 | 0.697364208 | 0.892773464 | 0.413799287 |
| sp\|Q6PI73\|LIRA6_HUMAN | 1 | 40 | 76 | 33 | -0.21016 | -0.012801267 | 0.070622828 | 0.197360437 | 0.280784531 | 0.083424095 | 0.020516847 | 0.004749746 | 0.532301756 |
| sp\|Q16822\|PCKGM_HUMAN | 1 | 40 | 76 | 33 | -0.28061 | 0.035003968 | 0.086559509 | 0.31560959 | 0.36716513 | 0.05155554 | 0.008215445 | 0.011137525 | 0.88864976 |
| sp\|Q9HCN6\|GPVI_HUMAN | 5 | 40 | 76 | 33 | -0.20616 | 0.013315994 | 0.162575343 | 0.219477351 | 0.3687367 | 0.149259349 | 0.027704752 | 0.001148342 | 0.2258375 |
| sp\|Q14914\|PTGR1_HUMAN | 5 | 40 | 76 | 33 | -0.25433 | -0.104683861 | -0.046007175 | 0.149643804 | 0.208320489 | 0.058676686 | 0.218651167 | 0.132358237 | 0.812133199 |
| sp\|P18827\|SDC1_HUMAN | 2 | 29 | 55 | 24 | -0.77524 | -0.617425243 | 0.328042011 | 0.157816571 | 1.103283826 | 0.945467255 | 0.664038843 | 6.22E-06 | 1.25E-05 |
| sp\|Q6PCE3\|PGM2L_HUMAN | 5 | 7 | 14 | 6 | -0.53203 | -0.260304991 | 0.332223047 | 0.271723666 | 0.864251705 | 0.592528039 | 0.544622025 | 0.024927755 | 0.0911423 |
| sp\|P36551\|HEM6_HUMAN | 1 | 37 | 68 | 30 | -0.15748 | -0.072625518 | 0.105806492 | 0.084859069 | 0.263291079 | 0.17843201 | 0.483616959 | 0.009733353 | 0.065308766 |
| sp\|P20827\|EFNA1_HUMAN | 1 | 40 | 76 | 33 | -0.46422 | -0.35973953 | 0.089314798 | 0.104482685 | 0.553537013 | 0.449054328 | 0.528376376 | 1.43E-05 | 7.61E-05 |
| sp\|P62987\|RL40_HUMAN | 7 | 40 | 76 | 33 | -0.37093 | -0.180121859 | 0.339445315 | 0.190810642 | 0.710377817 | 0.519567175 | 0.188625499 | 6.97E-07 | 4.48E-05 |
| sp\|Q9NY33\|DPP3_HUMAN | 2 | 40 | 76 | 33 | -0.13073 | -0.106054348 | 0.246793573 | 0.024680406 | 0.377528327 | 0.352847921 | 0.939157503 | 9.60E-05 | 3.73E-05 |
| sp\|Q9H4D0\|CSTN2_HUMAN | 29 | 14 | 27 | 12 | 0.164509 | 0.077606954 | 0.388671145 | -0.086902058 | 0.224162133 | 0.311064191 | 0.764247141 | 0.293612078 | 0.054191639 |
| sp\|Q58FG0\|HS905_HUMAN | 2 | 40 | 76 | 33 | -0.34698 | -0.255075064 | 0.259979437 | 0.091902635 | 0.606957675 | 0.515055041 | 0.760011966 | 0.000467363 | 0.000853049 |
| sp\|Q29983\|MICA_HUMAN | 6 | 16 | 27 | 12 | -0.02831 | -0.013195373 | 0.294741956 | 0.015113397 | 0.323050726 | 0.30793733 | 0.992235194 | 0.099691846 | 0.080021791 |
| sp\|P11117\|PPAL_HUMAN | 212 | 21 | 42 | 18 | 0.095352 | 0.017774304 | 0.115192991 | -0.077577883 | 0.019840804 | 0.097418687 | 0.556266811 | 0.973547608 | 0.436469374 |
| sp\|Q9UKZ9\|PCOC2_HUMAN | 11 | 40 | 76 | 33 | 0.14236 | 0.112247693 | -0.067271069 | -0.030111843 | -0.209630604 | -0.179518762 | 0.714518584 | 3.71E-05 | 6.94E-05 |
| sp\|P05204\|HMGN2_HUMAN | 8 | 36 | 69 | 30 | -1.13318 | -0.874484969 | 0.23830951 | 0.258696238 | 1.371490717 | 1.112794478 | 0.43007925 | 6.19E-07 | 4.74E-06 |
| sp\|Q3LXA3\|TKFC_HUMAN | 2 | 40 | 76 | 33 | -0.15621 | 0.005442595 | 0.207736946 | 0.161652136 | 0.363946487 | 0.202294351 | 0.135871688 | 0.001276245 | 0.06561461 |
| sp\|Q99497\|PARK7_HUMAN | 8 | 40 | 76 | 33 | -0.10487 | -0.040654802 | 0.329448998 | 0.064218542 | 0.434322343 | 0.3701038 | 0.724127248 | 8.96E-05 | 0.000172728 |
| sp\|P08476\|INHBA_HUMAN | 3 | 30 | 55 | 23 | -0.09003 | 0.006718777 | 0.186828205 | 0.096753087 | 0.276862516 | 0.180109428 | 0.501575848 | 0.026093062 | 0.140242773 |
| sp\|P36269\|GGT5_HUMAN | 1 | 40 | 76 | 33 | 0.248686 | 0.197747369 | 0.119067763 | -0.050938795 | -0.129618401 | -0.078679606 | 0.434456809 | 0.026816475 | 0.177309409 |
| sp\|O15372\|EIF3H_HUMAN | 1 | 4 | 7 | 3 | -0.72543 | -0.424642871 | 0.407629848 | 0.300791489 | 1.133064208 | 0.832272719 | 0.83516151 | 0.220314703 | 0.351182789 |
| sp\|Q13443\|ADAM9_HUMAN | 31 | 36 | 69 | 30 | -0.21437 | -0.072437413 | 0.247630683 | 0.141932469 | 0.462000566 | 0.320068097 | 0.133948561 | 1.95E-06 | 0.000215379 |
| sp\|P01589\|IL2RA_HUMAN | 4 | 4 | 7 | 2 | 0.047202 | 0.603220202 | 0.00093951 | 0.556017979 | -0.046262712 | -0.602280692 | 0.711781608 | 0.998723322 | 0.781766729 |
| sp\|Q14112\|NID2_HUMAN | 1 | 40 | 76 | 33 | -0.14025 | -0.006054395 | 0.147044232 | 0.134196711 | 0.287295338 | 0.153098626 | 0.064932563 | 0.000275286 | 0.044482945 |
| sp\|P17900\|SAP3_HUMAN | 10 | 40 | 76 | 33 | -0.03985 | -0.029225198 | 0.540912108 | 0.010629433 | 0.580766738 | 0.570137306 | 0.992247275 | 8.17E-07 | 5.12E-08 |
| sp\|Q16143\|SYUB_HUMAN | 1 | 40 | 76 | 33 | -0.16094 | -0.194672933 | 0.257595297 | -0.033736974 | 0.418531256 | 0.45226823 | 0.958325293 | 0.01324135 | 0.001831903 |
| sp\|P16152\|CBR1_HUMAN | 17 | 40 | 76 | 33 | -0.16649 | -0.006089472 | 0.30163986 | 0.160397215 | 0.468126547 | 0.307729332 | 0.137537775 | 2.21E-05 | 0.002134637 |
| sp\|Q9H0P0\|5NT3A_HUMAN | 3 | 26 | 48 | 20 | -0.01183 | -0.021736644 | 0.078940508 | -0.009902196 | 0.090774956 | 0.100677152 | 0.995816478 | 0.790304703 | 0.697083575 |
| sp\|Q13361\|MFAP5_HUMAN | 10 | 40 | 76 | 33 | 0.061832 | -0.014185041 | 0.259995948 | -0.076016771 | 0.198164218 | 0.274180989 | 0.306852004 | 0.00493265 | 5.32E-06 |
| sp\|P43490\|NAMPT_HUMAN | 1 | 40 | 76 | 33 | -0.27993 | -0.128119909 | 0.264683327 | 0.15180954 | 0.544612776 | 0.392803236 | 0.293802555 | 4.61E-05 | 0.001104764 |
| sp\|P13521\|SCG2_HUMAN | 3 | 29 | 55 | 25 | 0.29165 | 0.286154492 | 0.237394042 | -0.005495587 | -0.054256038 | -0.048760451 | 0.997718583 | 0.854631175 | 0.850092698 |
| sp\|P15170\|ERF3A_HUMAN | 38 | 29 | 56 | 24 | -0.41856 | -0.295217729 | 0.248516933 | 0.123340411 | 0.667075073 | 0.543734662 | 0.72864694 | 0.002672764 | 0.006170037 |
| sp\|Q92187\|SIA8D_HUMAN | 7 | 40 | 76 | 33 | -0.01779 | 0.019952169 | 0.011954198 | 0.03774637 | 0.029748399 | -0.007997971 | 0.715377032 | 0.866027912 | 0.986838941 |
| sp\|Q15843\|NEDD8_HUMAN | 27 | 8 | 14 | 6 | 0.110791 | -0.007254823 | 0.355697638 | -0.118046219 | 0.244906242 | 0.362952461 | 0.620453883 | 0.263872917 | 0.037372726 |
| sp\|Q15257\|PTPA_HUMAN | 2 | 40 | 76 | 33 | -0.12014 | -0.087742218 | 0.311412058 | 0.032394119 | 0.431548394 | 0.399154275 | 0.911846038 | 3.62E-05 | 1.59E-05 |
| sp\|P55263\|ADK_HUMAN | 2 | 37 | 69 | 30 | -0.2524 | -0.107955162 | 0.180984753 | 0.144445062 | 0.433384977 | 0.288939915 | 0.207129656 | 0.000121252 | 0.005215717 |
| sp\|Q99983\|OMD_HUMAN | 3 | 40 | 76 | 33 | 0.122497 | 0.054527186 | -0.079528516 | -0.06796954 | -0.202025242 | -0.134055701 | 0.600385906 | 0.048348756 | 0.179005155 |
| sp\|P29218\|IMPA1_HUMAN | 7 | 36 | 69 | 30 | 0.048856 | -0.060507109 | 0.275579458 | -0.109362853 | 0.226723713 | 0.336086566 | 0.369218155 | 0.055021834 | 0.00043572 |
| sp\|P22392\|NDKB_HUMAN | 6 | 11 | 21 | 9 | -0.28559 | 0.012821661 | 0.619286665 | 0.298409723 | 0.904874727 | 0.606465004 | 0.415363935 | 0.007390846 | 0.051134402 |
| sp\|Q9BWD1\|THIC_HUMAN | 8 | 40 | 76 | 33 | -0.17356 | -0.05915392 | 0.140415767 | 0.11440767 | 0.313977357 | 0.199569687 | 0.361909184 | 0.00629467 | 0.069323573 |
| sp\|P48357\|LEPR_HUMAN | 5 | 40 | 76 | 33 | 0.077705 | 0.035055828 | 0.039575992 | -0.042648676 | -0.038128512 | 0.004520164 | 0.668352013 | 0.800403792 | 0.996016994 |
| sp\|P13473\|LAMP2_HUMAN | 7 | 40 | 76 | 33 | 0.058572 | 0.086522898 | -0.076353248 | 0.027950927 | -0.134925219 | -0.162876146 | 0.58912692 | 0.000369632 | 9.32E-07 |
| sp\|Q9H461\|FZD8_HUMAN | 11 | 19 | 34 | 14 | 0.285653 | 0.157824718 | 0.39371214 | -0.127828143 | 0.108059279 | 0.235887422 | 0.533256574 | 0.741517587 | 0.181586247 |
| sp\|Q6GMV3\|PTRD1_HUMAN | 1 | 8 | 14 | 6 | 0.053722 | 0.113645032 | 0.062924378 | 0.059923198 | 0.009202544 | -0.050720654 | 0.887569413 | 0.998097581 | 0.931791055 |
| sp\|Q86X29\|LSR_HUMAN | 5 | 40 | 76 | 33 | -0.09246 | -0.070812257 | 0.221934267 | 0.02164874 | 0.314395265 | 0.292746525 | 0.930316295 | 6.99E-05 | 2.84E-05 |
| sp\|Q9Y678\|COPG1_HUMAN | 2 | 27 | 48 | 20 | -0.38421 | -0.10696642 | 0.161979906 | 0.277242176 | 0.546188502 | 0.268946326 | 0.133964509 | 0.006912686 | 0.211046378 |
| sp\|O14672\|ADA10_HUMAN | 30 | 40 | 76 | 33 | -0.28191 | -0.166475693 | 0.211937206 | 0.115435169 | 0.493848068 | 0.378412899 | 0.321592221 | 2.73E-06 | 5.38E-05 |
| sp\|Q6P4E1\|CASC4_HUMAN | 2 | 40 | 76 | 33 | -0.02049 | 0.105389899 | 0.276337888 | 0.125884335 | 0.296832324 | 0.170947989 | 0.081449571 | 0.000119827 | 0.018328549 |
| sp\|P67775\|PP2AA_HUMAN | 23 | 40 | 76 | 33 | -0.48402 | -0.27397666 | 0.095216625 | 0.210038783 | 0.579232069 | 0.369193286 | 0.241588422 | 0.000870052 | 0.023241318 |
| sp\|Q9H4X1\|RGCC_HUMAN | 7 | 20 | 34 | 15 | -0.23293 | -0.17365589 | -0.275105581 | 0.059273196 | -0.042176495 | -0.101449691 | 0.964308024 | 0.987546819 | 0.915840204 |
| sp\|Q9UMX5\|NENF_HUMAN | 1 | 40 | 76 | 33 | 0.007757 | -0.085368441 | 0.126345122 | -0.093125649 | 0.118587914 | 0.211713563 | 0.217922092 | 0.182399477 | 0.001397556 |
| sp\|P48788\|TNNI2_HUMAN | 19 | 11 | 20 | 9 | -0.77774 | -1.008798709 | 0.202529215 | -0.23106231 | 0.980265614 | 1.211327924 | 0.847703389 | 0.140577169 | 0.028026453 |
| sp\|P05060\|SCG1_HUMAN | 3 | 40 | 76 | 33 | -0.04139 | -0.118718884 | 0.11196887 | -0.077325079 | 0.153362675 | 0.230687754 | 0.576556652 | 0.227628176 | 0.01581889 |
| sp\|P02794\|FRIH_HUMAN | 18 | 40 | 76 | 33 | -0.34192 | -0.206735573 | 0.372769274 | 0.135186306 | 0.714691153 | 0.579504847 | 0.565672597 | 4.48E-05 | 0.000204305 |
| sp\|Q6UXK5\|LRRN1_HUMAN | 24 | 40 | 76 | 33 | 0.214052 | 0.115936588 | -0.289501231 | -0.098115178 | -0.503552997 | -0.405437819 | 0.179014333 | 9.95E-12 | 4.36E-10 |
| sp\|P43405\|KSYK_HUMAN | 19 | 14 | 28 | 11 | -0.03273 | 0.029447956 | 0.556078129 | 0.062176066 | 0.58880624 | 0.526630174 | 0.946289894 | 0.047651725 | 0.044316664 |
| sp\|Q06187\|BTK_HUMAN | 9 | 40 | 76 | 33 | 0.018916 | -0.050504621 | 0.072699601 | -0.069420559 | 0.053783664 | 0.123204222 | 0.755408537 | 0.890143596 | 0.462251933 |
| sp\|Q01469\|FABP5_HUMAN | 3 | 40 | 76 | 33 | -0.23402 | -0.078529375 | 0.353050453 | 0.15549078 | 0.587070608 | 0.431579828 | 0.409615045 | 0.000283934 | 0.003192126 |
| sp\|P04271\|S100B_HUMAN | 1 | 8 | 14 | 5 | 0.27778 | 0.18394617 | -0.15902054 | -0.093833392 | -0.436800102 | -0.34296671 | 0.624361428 | 0.006719094 | 0.020451402 |
| sp\|O95817\|BAG3_HUMAN | 20 | 11 | 20 | 9 | 0.237214 | 0.122414771 | 0.53064989 | -0.11479944 | 0.293435679 | 0.408235119 | 0.872838028 | 0.542953413 | 0.235871039 |
| sp\|Q9BUL8\|PDC10_HUMAN | 14 | 36 | 69 | 30 | -0.36102 | -0.208985734 | 0.044217477 | 0.152031365 | 0.405234576 | 0.253203211 | 0.626566837 | 0.105199586 | 0.32063634 |
| sp\|P59998\|ARPC4_HUMAN | 1 | 40 | 76 | 33 | -0.08407 | -0.108455217 | 0.143423311 | -0.024385115 | 0.227493412 | 0.251878528 | 0.969988098 | 0.165959454 | 0.062446159 |
| sp\|O75914\|PAK3_HUMAN | 2 | 29 | 55 | 23 | -0.0593 | -0.037446033 | 0.309513134 | 0.021852482 | 0.368811649 | 0.346959167 | 0.97098134 | 0.00503794 | 0.002803392 |
| sp\|Q96C90\|PP14B_HUMAN | 152 | 4 | 6 | 3 | 0.035912 | 0.115297509 | 1.06768294 | 0.079385414 | 1.031770845 | 0.952385431 | 0.993204345 | 0.467699436 | 0.469662863 |
| sp\|P48960\|CD97_HUMAN | 15 | 40 | 76 | 33 | -0.00249 | -0.03061782 | 0.092649679 | -0.0281291 | 0.0951384 | 0.123267499 | 0.806561711 | 0.188084887 | 0.0301155 |
| sp\|Q9HC38\|GLOD4_HUMAN | 1 | 40 | 76 | 33 | -0.10151 | -0.058010749 | 0.327107318 | 0.043499389 | 0.428617456 | 0.385118067 | 0.836835811 | 2.13E-05 | 1.63E-05 |
| sp\|Q99674\|CGRE1_HUMAN | 2 | 40 | 76 | 33 | -0.14679 | -0.262241197 | 0.224538655 | -0.115446439 | 0.371333413 | 0.486779852 | 0.555456795 | 0.017425112 | 0.000206486 |
| sp\|P25815\|S100P_HUMAN | 61 | 40 | 76 | 33 | -0.7255 | -0.582685042 | 0.090479064 | 0.142819113 | 0.815983219 | 0.673164106 | 0.663564396 | 0.000199187 | 0.000579267 |
| sp\|O60749\|SNX2_HUMAN | 107 | 14 | 28 | 12 | -0.02175 | 0.022392839 | 0.355400151 | 0.044144746 | 0.377152058 | 0.333007311 | 0.95451503 | 0.106996138 | 0.103975698 |
| sp\|O00754\|MA2B1_HUMAN | 1 | 40 | 76 | 33 | 0.151673 | 0.184243714 | 0.440759612 | 0.032570987 | 0.289086884 | 0.256515898 | 0.877356506 | 0.001285189 | 0.001270048 |
| sp\|Q9NQ25\|SLAF7_HUMAN | 6 | 14 | 27 | 13 | 0.037432 | 0.098562772 | 0.150567301 | 0.061131132 | 0.113135662 | 0.05200453 | 0.808459656 | 0.589478216 | 0.86356626 |
| sp\|P28676\|GRAN_HUMAN | 14 | 7 | 13 | 6 | 0.380309 | 0.061499725 | 0.810382322 | -0.31880933 | 0.430073267 | 0.748882597 | 0.729044005 | 0.665916373 | 0.225897306 |
| sp\|O95831\|AIFM1_HUMAN | 2 | 4 | 7 | 3 | -0.1501 | 0.105246315 | 0.175336971 | 0.25534432 | 0.325434975 | 0.070090656 | 0.637932213 | 0.612720346 | 0.971326887 |
| sp\|P63104\|1433Z_HUMAN | 1 | 40 | 76 | 33 | -0.04385 | -0.243076568 | 0.129411075 | -0.199228505 | 0.173259138 | 0.372487643 | 0.289338554 | 0.521426893 | 0.024463335 |
| sp\|P48723\|HSP13_HUMAN | 4 | 40 | 76 | 33 | 0.02316 | -0.000497305 | 0.196377024 | -0.023657438 | 0.173216891 | 0.196874329 | 0.921120827 | 0.052372043 | 0.008558818 |
| sp\|P00492\|HPRT_HUMAN | 6 | 40 | 76 | 33 | -0.26965 | -0.0860166 | 0.238223793 | 0.183633931 | 0.507874324 | 0.324240392 | 0.039000567 | 2.23E-07 | 0.000215375 |

| | | | | | | | | | | | | | |
|---|---|---|---|---|---|---|---|---|---|---|---|---|---|
| sp\|Q86TY3\|ARMD4_HUMAN | 19 | 40 | 76 | 33 | 0.124548 | -0.005767869 | -0.198542523 | -0.130316146 | -0.3230908 | -0.192774654 | 0.037870584 | 3.03E-06 | 0.002209315 |
| sp\|Q9NR34\|MA1C1_HUMAN | 14 | 40 | 76 | 33 | -0.01629 | -0.001690063 | 0.196540649 | 0.014600104 | 0.212830816 | 0.198230712 | 0.937149872 | 0.000161609 | 7.02E-05 |
| sp\|P13686\|PPA5_HUMAN | 253 | 40 | 76 | 33 | -0.09247 | -0.024840512 | 0.223040994 | 0.067626142 | 0.315507648 | 0.247881505 | 0.499679033 | 7.12E-05 | 0.000482996 |
| sp\|Q9NQ30\|ESM1_HUMAN | 6 | 4 | 7 | 3 | 0.190104 | -0.007052533 | 0.181154736 | -0.197156425 | -0.008949156 | 0.188207269 | 0.458967925 | 0.998836184 | 0.551051049 |
| sp\|Q86SG5\|S1A7A_HUMAN | 8 | 40 | 76 | 33 | -1.77038 | -1.891262406 | -2.177428167 | -0.120882038 | -0.407047799 | -0.286165761 | 0.886307718 | 0.392165383 | 0.553844482 |
| sp\|P22304\|IDS_HUMAN | 16 | 40 | 76 | 33 | 0.293056 | 0.202264556 | 0.113690652 | -0.090791858 | -0.179365762 | -0.088573904 | 0.137303143 | 0.005642401 | 0.189246175 |
| sp\|Q92598\|HS105_HUMAN | 1 | 25 | 48 | 20 | -0.35481 | -0.045961505 | 0.425614652 | 0.308843515 | 0.780419672 | 0.471576157 | 0.100799441 | 0.000122016 | 0.011634551 |
| sp\|P16989\|YBOX3_HUMAN | 13 | 4 | 6 | 3 | 0.155497 | 0.145127887 | 0.825182739 | -0.01036951 | 0.669685342 | 0.680054852 | 0.99972131 | 0.464885534 | 0.403449686 |
| sp\|P63208\|SKP1_HUMAN | 3 | 40 | 76 | 33 | -0.38232 | -0.169453745 | 0.517504935 | 0.212864618 | 0.899823298 | 0.686958681 | 0.163299288 | 5.19E-09 | 4.20E-07 |
| sp\|P52799\|EFNB2_HUMAN | 13 | 40 | 76 | 33 | -0.11429 | -0.122487046 | 0.226896706 | -0.008197786 | 0.341185967 | 0.349383753 | 0.993535211 | 0.000740578 | 8.09E-05 |
| sp\|O00410\|IPO5_HUMAN | 4 | 28 | 56 | 24 | -0.39668 | -0.297189211 | 0.350088885 | 0.099495023 | 0.746773119 | 0.647278096 | 0.818910628 | 0.000799526 | 0.000932013 |
| sp\|P17174\|AATC_HUMAN | 11 | 40 | 76 | 33 | -0.04939 | -0.012906125 | 0.169724646 | 0.036479355 | 0.219110125 | 0.18263077 | 0.888458142 | 0.057232304 | 0.079078162 |
| sp\|Q8WY91\|THAP4_HUMAN | 49 | 4 | 6 | 3 | -0.34236 | -0.273675816 | 0.535740797 | 0.068679695 | 0.878096308 | 0.809416613 | 0.987613224 | 0.279947032 | 0.282689163 |
| sp\|P55000\|SLUR1_HUMAN | 4 | 30 | 55 | 25 | 0.206895 | 0.105836861 | 0.183223552 | -0.101057811 | -0.023671121 | 0.077386691 | 0.406151031 | 0.965484299 | 0.624707067 |
| sp\|P07451\|CAH3_HUMAN | 3 | 40 | 76 | 33 | -1.24722 | -1.204989977 | -0.634785177 | 0.04222852 | 0.61243332 | 0.5702048 | 0.933862384 | 0.000113911 | 4.83E-05 |
| sp\|P20036\|DPA1_HUMAN | 3 | 40 | 76 | 33 | -0.09792 | -0.206275714 | -0.204285752 | -0.108356715 | -0.106366753 | 0.001989962 | 0.441690625 | 0.579189973 | 0.999755942 |
| sp\|Q8IWL1\|SFPA2_HUMAN | 226 | 40 | 76 | 33 | -0.12242 | -0.059404939 | 0.301442432 | 0.063012338 | 0.423859709 | 0.360847371 | 0.525892431 | 3.42E-08 | 1.11E-07 |
| sp\|Q01433\|AMPD2_HUMAN | 1 | 7 | 14 | 6 | -0.08752 | 0.104942843 | 0.821032153 | 0.192466651 | 0.908555961 | 0.71608931 | 0.774154199 | 0.032891274 | 0.058852944 |
| sp\|P58400\|NRX1B_HUMAN | 16 | 29 | 56 | 24 | 0.264152 | 0.207784677 | 0.051786643 | -0.05636764 | -0.212365675 | -0.155998034 | 0.699196651 | 0.034823498 | 0.095571925 |
| sp\|Q9Y2J2\|E41L3_HUMAN | 16 | 10 | 21 | 9 | 0.031057 | -0.132578386 | 0.057755799 | -0.163635807 | 0.026698378 | 0.190334184 | 0.436267798 | 0.98428963 | 0.354510782 |
| sp\|P02763\|A1AG1_HUMAN | 1 | 40 | 76 | 33 | -0.98163 | -0.875308321 | -0.200417449 | 0.106323157 | 0.78121403 | 0.674890873 | 0.826567621 | 0.001299057 | 0.001780612 |
| sp\|Q9NRA1\|PDGFC_HUMAN | 5 | 40 | 76 | 33 | -0.00253 | 0.034922339 | 0.154930686 | 0.037451819 | 0.157460167 | 0.120008348 | 0.658485149 | 0.007686183 | 0.026182636 |
| sp\|O00231\|PSD11_HUMAN | 14 | 22 | 42 | 18 | -0.20446 | -0.10288312 | 0.208829449 | 0.101576574 | 0.413289143 | 0.311712568 | 0.741095053 | 0.039071497 | 0.092531012 |
| sp\|P08779\|K1C16_HUMAN | 2 | 17 | 35 | 16 | 0.055556 | 0.019821135 | -0.08210174 | -0.035734496 | -0.137657371 | -0.101922875 | 0.988745438 | 0.886269389 | 0.915551309 |
| sp\|Q9UKK9\|NUDT5_HUMAN | 10 | 40 | 76 | 33 | -0.07552 | -0.038357631 | 0.333326706 | 0.03716682 | 0.408851157 | 0.371684337 | 0.882377531 | 7.33E-05 | 4.67E-05 |
| sp\|Q9UJ68\|MSRA_HUMAN | 38 | 32 | 63 | 27 | 0.038727 | 0.024294817 | 0.271793038 | -0.014432637 | 0.233065584 | 0.247498221 | 0.988287201 | 0.126432932 | 0.050913403 |
| sp\|Q05209\|PTN12_HUMAN | 1 | 29 | 55 | 24 | -0.04877 | 0.03083437 | 0.264912067 | 0.079605491 | 0.313683188 | 0.234077697 | 0.885934367 | 0.277850184 | 0.401792763 |
| sp\|Q9BRT3\|MIEN1_HUMAN | 4 | 25 | 48 | 21 | -0.26966 | -0.310532607 | 0.376324174 | -0.040874436 | 0.645982346 | 0.686856782 | 0.966082591 | 0.003971509 | 0.000427043 |
| sp\|Q93074\|MED12_HUMAN | 4 | 4 | 6 | 3 | 0.030767 | -0.251086183 | 0.223481135 | -0.281853559 | 0.192713758 | 0.474567318 | 0.424149315 | 0.738077853 | 0.161135094 |
| sp\|O14618\|CCS_HUMAN | 5 | 34 | 62 | 26 | -0.20127 | -0.025498045 | 0.284961105 | 0.17577583 | 0.48623498 | 0.31045915 | 0.305821141 | 0.003131861 | 0.048966346 |
| sp\|Q8NFP4\|MDGA1_HUMAN | 3 | 22 | 42 | 18 | 0.134589 | 0.07183909 | -0.085002327 | -0.062749731 | -0.219591147 | -0.156841416 | 0.433145214 | 0.001617477 | 0.01339044 |
| sp\|P05451\|REG1A_HUMAN | 1 | 40 | 76 | 33 | -0.2005 | -0.145045922 | 0.246134396 | 0.055458504 | 0.446638821 | 0.391180317 | 0.913124411 | 0.020056649 | 0.021945672 |
| sp\|Q13510\|ASAH1_HUMAN | 22 | 40 | 76 | 33 | -0.08155 | -0.092691619 | 0.159318742 | -0.011146514 | 0.240863846 | 0.25201036 | 0.97449934 | 0.000450852 | 2.86E-05 |
| sp\|Q9UJC5\|SH3L2_HUMAN | 3 | 40 | 76 | 33 | -0.22248 | -0.25159108 | -0.206354313 | -0.029106693 | 0.016130074 | 0.045236767 | 0.981113168 | 0.995967185 | 0.960378843 |
| sp\|Q99972\|MYOC_HUMAN | 6 | 40 | 76 | 33 | 0.105383 | 0.097102472 | -0.007268605 | -0.008280929 | -0.112652005 | -0.104371077 | 0.989271157 | 0.257223906 | 0.227368199 |
| sp\|Q9BXD5\|NPL_HUMAN | 12 | 32 | 62 | 27 | -0.27801 | -0.177642435 | 0.323754667 | 0.100363146 | 0.601760248 | 0.501397102 | 0.632817209 | 3.69E-05 | 0.000100875 |
| sp\|O75695\|XRP2_HUMAN | 29 | 14 | 27 | 12 | -0.05946 | -0.093580327 | 0.303396568 | -0.034117018 | 0.362859877 | 0.396976895 | 0.951283567 | 0.026334781 | 0.00463503 |
| sp\|O60636\|TSN2_HUMAN | 4 | 14 | 28 | 13 | -0.0044 | -0.156276047 | 0.287132203 | -0.151875976 | 0.291532274 | 0.44340825 | 0.450528707 | 0.127314214 | 0.003108004 |
| sp\|P61758\|PFD3_HUMAN | 6 | 22 | 42 | 18 | -0.35171 | -0.099890016 | 0.343132309 | 0.251824953 | 0.694847278 | 0.443022326 | 0.56593135 | 0.057089101 | 0.220022385 |
| sp\|P30040\|ERP29_HUMAN | 1 | 40 | 76 | 33 | -0.11821 | -0.100145883 | 0.165006203 | 0.018065733 | 0.283217819 | 0.265152086 | 0.983638238 | 0.065920764 | 0.04863192 |
| sp\|Q8N257\|H2B3B_HUMAN | 5 | 40 | 76 | 33 | -0.44526 | -0.400679674 | 0.24314063 | 0.044579686 | 0.68839999 | 0.643820304 | 0.961266093 | 0.0022208 | 0.001161375 |
| sp\|P62807\|H2B1C_HUMAN | 12 | 25 | 48 | 21 | -0.653 | -0.548596655 | 0.330371625 | 0.104400963 | 0.983369243 | 0.87896828 | 0.903958698 | 0.003228626 | 0.002863285 |
| sp\|Q15631\|TSN_HUMAN | 34 | 40 | 76 | 33 | -0.31428 | -0.137594122 | 0.292287266 | 0.176685897 | 0.606567285 | 0.429881388 | 0.126999504 | 3.44E-07 | 4.84E-05 |
| sp\|O60234\|GMFG_HUMAN | 18 | 40 | 76 | 33 | -0.33829 | -0.181359876 | 0.198924791 | 0.156933765 | 0.537218432 | 0.380284667 | 0.444635269 | 0.002018318 | 0.017566721 |
| sp\|O95433\|AHSA1_HUMAN | 12 | 8 | 13 | 6 | -0.11479 | 0.016461502 | 0.294433064 | 0.131251119 | 0.409222682 | 0.277971562 | 0.786290772 | 0.217534907 | 0.419558338 |
| sp\|P17612\|KAPCA_HUMAN | 14 | 11 | 21 | 9 | 0.023313 | -0.179147898 | 0.568421404 | -0.202460967 | 0.545108335 | 0.747569303 | 0.669130067 | 0.148334114 | 0.014169793 |
| sp\|P22223\|CADH3_HUMAN | 1 | 28 | 55 | 24 | -0.10201 | -0.181275105 | 0.097835515 | -0.079267133 | 0.199843487 | 0.27911062 | 0.78577806 | 0.347951116 | 0.07364099 |
| sp\|O14745\|NHRF1_HUMAN | 8 | 40 | 76 | 33 | -0.11085 | -0.175088326 | 0.143720006 | -0.064235637 | 0.254572695 | 0.318808332 | 0.771922721 | 0.064983491 | 0.00491175 |
| sp\|P50897\|PPT1_HUMAN | 5 | 29 | 55 | 24 | 0.065309 | 0.077763118 | 0.151085651 | 0.012453787 | 0.085776319 | 0.073322533 | 0.981892225 | 0.551604354 | 0.574979592 |
| sp\|Q8WZA0\|LZIC_HUMAN | 2 | 36 | 69 | 30 | -0.11691 | -0.039380231 | 0.243776107 | 0.07753039 | 0.360686728 | 0.283156338 | 0.660920275 | 0.002899382 | 0.009473699 |
| sp\|Q15323\|K1H1_HUMAN | 9 | 8 | 13 | 6 | -0.20276 | 0.412831452 | -0.197189786 | 0.615595952 | 0.005574713 | -0.610021238 | 0.375467352 | 0.999941831 | 0.44760636 |
| sp\|P60983\|GMFB_HUMAN | 97 | 36 | 69 | 30 | -0.05131 | 0.098933408 | 0.412989298 | 0.150240831 | 0.464296721 | 0.31405589 | 0.407053435 | 0.003556099 | 0.03414401 |
| sp\|Q8TD46\|MO2R1_HUMAN | 20 | 16 | 27 | 12 | -0.318 | -0.173887882 | 0.099269688 | 0.144109747 | 0.417267317 | 0.27315757 | 0.673946863 | 0.114427192 | 0.315985704 |
| sp\|Q9NQZ7\|ENTP7_HUMAN | 80 | 19 | 34 | 14 | 0.041955 | 0.154485455 | 0.330671269 | 0.112530846 | 0.28871666 | 0.176185814 | 0.666204968 | 0.178436878 | 0.447630605 |
| sp\|P50135\|HNMT_HUMAN | 12 | 4 | 7 | 3 | -0.38845 | -0.066150653 | -0.188912636 | 0.322296821 | 0.199534839 | -0.122761982 | 0.09553984 | 0.490677924 | 0.710269246 |
| sp\|Q9Y2Q3\|GSTK1_HUMAN | 7 | 28 | 55 | 24 | -0.10035 | -0.299535639 | 0.271397876 | -0.199188031 | 0.371745483 | 0.570933514 | 0.4008797 | 0.112961596 | 0.001819114 |
| sp\|Q9Y2S2\|CRYL1_HUMAN | 32 | 31 | 55 | 23 | -0.27115 | -0.014369648 | 0.202324578 | 0.256782261 | 0.473476487 | 0.216694227 | 0.048285262 | 0.001407157 | 0.16631359 |
| sp\|P54317\|LIPR2_HUMAN | 44 | 15 | 28 | 11 | -0.04866 | 0.02014941 | 0.001689584 | 0.068813549 | 0.050353722 | -0.018459826 | 0.834773001 | 0.938941431 | 0.989505266 |
| sp\|Q9BVA1\|TBB2B_HUMAN | 11 | 40 | 76 | 33 | -0.39174 | -0.097067531 | 0.221001242 | 0.294676487 | 0.612745259 | 0.318068772 | 0.045679223 | 0.00016368 | 0.042655075 |
| sp\|P31150\|GDIA_HUMAN | 15 | 40 | 76 | 33 | 0.026653 | -0.046156709 | 0.180585831 | -0.073809291 | 0.15393325 | 0.227742541 | 0.637304311 | 0.262013185 | 0.026169762 |
| sp\|Q641Q3\|METRL_HUMAN | 25 | 40 | 76 | 33 | -0.00706 | 0.020886869 | 0.131194921 | 0.027948923 | 0.138256975 | 0.110308052 | 0.799641941 | 0.026163879 | 0.051234194 |
| sp\|P17987\|TCPA_HUMAN | 24 | 37 | 69 | 29 | -0.18748 | -0.112032053 | 0.287165958 | 0.07545047 | 0.474648481 | 0.399198011 | 0.715211986 | 0.000267037 | 0.000627841 |
| sp\|P15090\|FABP4_HUMAN | 1 | 40 | 76 | 33 | -0.35404 | -0.309634966 | 0.263891042 | 0.044403378 | 0.617929386 | 0.573526008 | 0.917022554 | 2.88E-05 | 1.16E-05 |
| sp\|P31997\|CEAM8_HUMAN | 2 | 32 | 62 | 27 | 0.092119 | 0.230126794 | 0.308698112 | 0.138008118 | 0.216579435 | 0.078571317 | 0.201105109 | 0.066885518 | 0.625165031 |
| sp\|Q13217\|DNJC3_HUMAN | 1 | 40 | 76 | 33 | -0.16827 | -0.063310643 | 0.14618093 | 0.104959644 | 0.314451217 | 0.209491573 | 0.127932805 | 8.98E-06 | 0.001052189 |
| sp\|Q9NS71\|GKN1_HUMAN | 6 | 40 | 76 | 33 | -0.10099 | -0.093985856 | 0.156270261 | 0.007001731 | 0.257257848 | 0.250256117 | 0.995609953 | 0.019559681 | 0.009082273 |
| sp\|Q9NS28\|RGS18_HUMAN | 4 | 19 | 34 | 15 | -0.0082 | 0.056604787 | 0.181208104 | 0.064803613 | 0.18940693 | 0.124603317 | 0.94528088 | 0.719739402 | 0.837577664 |
| sp\|Q9NX62\|IMPA3_HUMAN | 2 | 40 | 76 | 33 | -0.02308 | -0.070596892 | 0.178260436 | -0.047517458 | 0.20133987 | 0.248857328 | 0.530619095 | 0.000649222 | 1.38E-06 |
| sp\|Q16832\|DDR2_HUMAN | 57 | 37 | 69 | 30 | 0.187256 | 0.080085241 | -0.225713574 | -0.107171095 | -0.41296991 | -0.305798815 | 0.063436457 | 8.79E-11 | 4.38E-08 |
| sp\|O75558\|STX11_HUMAN | 4 | 8 | 14 | 6 | 0.177835 | -0.200094679 | -0.295861136 | -0.377929955 | -0.473696412 | -0.095766457 | 0.361495869 | 0.341685261 | 0.945313624 |
| sp\|Q14574\|DSC3_HUMAN | 3 | 40 | 76 | 33 | 0.10057 | 0.090483673 | 0.12584647 | -0.010086647 | 0.025276151 | 0.035362797 | 0.963564444 | 0.851631556 | 0.670996222 |
| sp\|P0DPI2\|GAL3A_HUMAN | 7 | 14 | 27 | 12 | -0.09999 | 0.186162949 | 0.877248266 | 0.28615288 | 0.977238197 | 0.691085317 | 0.351755255 | 0.000632638 | 0.006717942 |
| sp\|P06454\|PTMA_HUMAN | 7 | 40 | 76 | 33 | -0.74247 | -0.420720906 | 0.326347633 | 0.321751134 | 1.068819674 | 0.747068539 | 0.093607186 | 1.24E-07 | 3.11E-05 |
| sp\|Q96PX8\|SLIK1_HUMAN | 5 | 40 | 76 | 33 | 0.192498 | 0.03164121 | -0.054730116 | -0.160856293 | -0.247227619 | -0.086371326 | 0.006000066 | 0.000302555 | 0.260391175 |
| sp\|Q16881\|TRXR1_HUMAN | 8 | 40 | 76 | 33 | -0.08475 | 0.046621911 | 0.335324271 | 0.131369545 | 0.420071905 | 0.28870236 | 0.128546985 | 2.17E-06 | 0.000277557 |
| sp\|Q9NU53\|GINM1_HUMAN | 2 | 29 | 55 | 25 | -0.11114 | -0.046089277 | 0.144345258 | 0.065054576 | 0.25548911 | 0.190434535 | 0.745483489 | 0.045561553 | 0.108455572 |
| sp\|Q9C0C4\|SEM4C_HUMAN | 7 | 40 | 76 | 33 | 0.06489 | 0.059302324 | 0.093283401 | -0.005587239 | 0.028393838 | 0.033981077 | 0.986153168 | 0.780482916 | 0.637166358 |
| sp\|P26992\|CNTFR_HUMAN | 52 | 40 | 76 | 33 | 0.14903 | 0.125975439 | 0.008608679 | -0.023054071 | -0.140420831 | -0.11736676 | 0.864735067 | 0.027618125 | 0.040632496 |
| sp\|P00505\|AATM_HUMAN | 2 | 4 | 7 | 3 | 0.033082 | -0.204697867 | -0.138950978 | -0.237779536 | -0.172032647 | 0.065746889 | 0.64607559 | 0.853704233 | 0.971753764 |
| sp\|P36941\|TNR3_HUMAN | 8 | 27 | 48 | 20 | -0.03322 | -0.036661266 | 0.450860652 | -0.003445663 | 0.484076255 | 0.487521918 | 0.999438885 | 0.001233267 | 0.000279657 |
| sp\|P20138\|CD33_HUMAN | 27 | 40 | 76 | 33 | 0.091373 | 0.113742477 | 0.111952968 | 0.022369749 | 0.02058024 | -0.001789509 | 0.958967463 | 0.975823883 | 0.999764512 |
| sp\|Q7Z794\|K2C1B_HUMAN | 14 | 40 | 76 | 33 | -0.13065 | -0.396146987 | -0.495939341 | -0.265500581 | -0.365292936 | -0.099792354 | 0.203439242 | 0.126240819 | 0.818367626 |
| sp\|Q9BS40\|LXN_HUMAN | 5 | 30 | 55 | 23 | -0.01948 | 0.090793148 | 0.229763952 | 0.110270063 | 0.249240867 | 0.138970804 | 0.133264358 | 0.001469903 | 0.070484642 |
| sp\|O60476\|MA1A2_HUMAN | 1 | 36 | 69 | 30 | 0.051709 | 0.012619603 | 0.142743724 | -0.039088944 | 0.091035176 | 0.130124121 | 0.791981986 | 0.419513199 | 0.107160925 |
| sp\|Q9UBC2\|EP15R_HUMAN | 25 | 17 | 35 | 14 | -0.48678 | -0.338100414 | 0.546947258 | 0.148681196 | 1.033728868 | 0.885047672 | 0.776250602 | 0.000754338 | 0.001005993 |
| sp\|Q9Y624\|JAM1_HUMAN | 2 | 40 | 76 | 33 | -0.46828 | -0.351241175 | 0.309466319 | 0.117037572 | 0.777745066 | 0.660707494 | 0.533092895 | 6.73E-08 | 2.22E-07 |
| sp\|Q9UBQ0\|VPS29_HUMAN | 1 | 33 | 63 | 26 | -0.31122 | -0.23892293 | 0.239140766 | 0.072301796 | 0.550365491 | 0.478063696 | 0.810545789 | 0.000548225 | 0.000750306 |
| sp\|Q9NPF2\|CHSTB_HUMAN | 27 | 11 | 21 | 9 | -0.36228 | -0.080900286 | 0.274660832 | 0.281381671 | 0.636942789 | 0.355561118 | 0.144437935 | 0.002442364 | 0.071380078 |
| sp\|P28070\|PSB4_HUMAN | 4 | 40 | 76 | 33 | -0.25872 | -0.107753868 | 0.442695474 | 0.150966473 | 0.701415815 | 0.550449342 | 0.301098688 | 1.65E-07 | 3.49E-06 |
| sp\|P61020\|RAB5B_HUMAN | 48 | 26 | 48 | 21 | 0.005085 | 0.046379583 | 0.293780106 | 0.041294607 | 0.288695131 | 0.247400524 | 0.932058722 | 0.100931504 | 0.119663151 |
| sp\|Q9NRB3\|CHSTC_HUMAN | 2 | 40 | 76 | 33 | -0.13575 | -0.079775868 | -0.015884195 | 0.055969246 | 0.11986092 | 0.063891673 | 0.362825851 | 0.043356083 | 0.314053835 |
| sp\|Q16401\|PSMD5_HUMAN | 2 | 18 | 35 | 15 | -0.33324 | -0.18821322 | 0.346748072 | 0.145029315 | 0.679990607 | 0.534961292 | 0.633220542 | 0.00201577 | 0.006490977 |
| sp\|P50570\|DYN2_HUMAN | 7 | 15 | 28 | 12 | 0.197786 | -0.009994068 | 0.512913542 | -0.207780452 | 0.315127158 | 0.522907611 | 0.611994074 | 0.464858404 | 0.078003504 |
| sp\|P20472\|PRVA_HUMAN | 27 | 15 | 28 | 11 | 0.028368 | -0.102851438 | 0.032917531 | -0.131219408 | 0.00454956 | 0.135768969 | 0.831610897 | 0.999855367 | 0.852400494 |
| sp\|O95782\|AP2A1_HUMAN | 11 | 15 | 27 | 11 | 0.375981 | 0.272632536 | 0.28593216 | -0.1033482 | -0.090048576 | 0.013299624 | 0.730709075 | 0.854410477 | 0.995766406 |
| sp\|Q9BRF8\|CPPED_HUMAN | 6 | 4 | 7 | 3 | 0.407514 | 0.072416762 | 0.3639609 | -0.335097116 | -0.043552978 | 0.291544138 | 0.476918814 | 0.991004017 | 0.622258686 |
| sp\|O43847\|NRDC_HUMAN | 3 | 4 | 7 | 3 | -0.10921 | 0.077535583 | 0.74930486 | 0.186746171 | 0.858515448 | 0.671769277 | 0.824453183 | 0.105641793 | 0.170659169 |
| sp\|O94919\|ENDD1_HUMAN | 4 | 40 | 76 | 33 | -0.12233 | 0.005604214 | 0.235905479 | 0.127935807 | 0.358237071 | 0.230301264 | 0.153539538 | 8.12E-05 | 0.00575811 |
| sp\|Q14644\|RASA3_HUMAN | 14 | 37 | 69 | 30 | 0.039907 | -0.148071164 | -0.009799711 | -0.187977724 | -0.04970627 | 0.138271454 | 0.329667459 | 0.947420688 | 0.591741841 |
| sp\|Q14103\|HNRPD_HUMAN | 1 | 30 | 55 | 23 | -0.34951 | -0.096790414 | 0.304622161 | 0.252720229 | 0.654132803 | 0.401412574 | 0.186778518 | 0.000872406 | 0.031555388 |
| sp\|Q9UN19\|DAPP1_HUMAN | 3 | 22 | 41 | 17 | -0.03428 | -0.123313025 | 0.118520629 | -0.089028371 | 0.152805283 | 0.241833654 | 0.817698918 | 0.673129075 | 0.293696446 |
| sp\|Q00577\|PURA_HUMAN | 1 | 4 | 7 | 3 | 0.318005 | -0.05330921 | 0.314914994 | -0.371314593 | -0.00309039 | 0.368224203 | 0.653033192 | 0.999979293 | 0.705870982 |
| sp\|P00480\|OTC_HUMAN | 17 | 40 | 76 | 33 | -0.62398 | -0.427577134 | -0.136296615 | 0.196398331 | 0.48767885 | 0.291280519 | 0.354261354 | 0.013971362 | 0.137475087 |
| sp\|P24752\|THIL_HUMAN | 5 | 4 | 7 | 3 | -0.04723 | -0.175161228 | -0.205228972 | -0.127934634 | -0.158002378 | -0.030067744 | 0.833713937 | 0.829667226 | 0.991609243 |
| sp\|P62495\|ERF1_HUMAN | 3 | 32 | 62 | 28 | -0.15658 | -0.170235715 | 0.160626831 | -0.013654857 | 0.317207688 | 0.330862546 | 0.991193141 | 0.038825628 | 0.011114371 |

| | | | | | | | | | | | | | |
|---|---|---|---|---|---|---|---|---|---|---|---|---|---|
| sp\|P01903\|DRA_HUMAN | 4 | 40 | 76 | 33 | 0.032938 | 0.081040614 | 0.160757882 | 0.04810246 | 0.127819728 | 0.079717268 | 0.737265458 | 0.230653051 | 0.481111024 |
| sp\|Q9H4A9\|DPEP2_HUMAN | 6 | 40 | 76 | 33 | 0.09588 | 0.145783177 | -0.041640792 | 0.049903576 | -0.137520393 | -0.187423969 | 0.443636802 | 0.01630653 | 9.50E-05 |
| sp\|O95025\|SEM3D_HUMAN | 5 | 8 | 14 | 6 | -0.28087 | -0.312329713 | -0.288585088 | -0.0314619 | -0.007717275 | 0.023744625 | 0.957261913 | 0.998228426 | 0.979663815 |
| sp\|Q7L576\|CYFP1_HUMAN | 4 | 37 | 69 | 30 | -0.35028 | -0.119942453 | 0.156487067 | 0.23033445 | 0.50676397 | 0.27642952 | 0.218369158 | 0.007558786 | 0.150367874 |
| sp\|P00167\|CYB5_HUMAN | 23 | 30 | 55 | 24 | -0.07092 | 0.100064685 | 0.324278331 | 0.170984188 | 0.395197835 | 0.224213647 | 0.13776416 | 0.001066715 | 0.055055648 |
| sp\|P26639\|SYTC_HUMAN | 1 | 40 | 76 | 33 | -0.06252 | -0.094349723 | 0.244908558 | -0.031827873 | 0.307430408 | 0.339258281 | 0.946425324 | 0.032634116 | 0.005463051 |
| sp\|P30085\|KCY_HUMAN | 1 | 40 | 76 | 33 | -0.29152 | -0.141241513 | 0.127777461 | 0.150274716 | 0.419293689 | 0.269018973 | 0.290264234 | 0.001811473 | 0.033256459 |
| sp\|Q9NP98\|MYOZ1_HUMAN | 12 | 4 | 6 | 3 | 0.004958 | 0.010692812 | 1.586671638 | 0.005735218 | 1.581714043 | 1.575978825 | 0.999947928 | 0.107761144 | 0.081832946 |
| sp\|Q16772\|GSTA3_HUMAN | 3 | 36 | 69 | 30 | -0.36183 | -0.044941834 | 0.176669291 | 0.316885622 | 0.538496747 | 0.221611125 | 0.145382554 | 0.022942019 | 0.430081645 |
| sp\|Q9UIA9\|XPO7_HUMAN | 8 | 19 | 35 | 14 | -0.17237 | -0.031900101 | 0.31989531 | 0.140472171 | 0.492267582 | 0.351795411 | 0.546849705 | 0.011032261 | 0.052843887 |
| sp\|P02008\|HBAZ_HUMAN | 1 | 12 | 21 | 9 | -0.00516 | -0.15428431 | 0.653029089 | -0.149126561 | 0.658186837 | 0.807313398 | 0.804665198 | 0.070405955 | 0.009884215 |
| sp\|Q92743\|HTRA1_HUMAN | 11 | 40 | 76 | 33 | -0.23217 | -0.145512572 | 0.337879101 | 0.086660745 | 0.570052418 | 0.483391673 | 0.434253485 | 9.69E-10 | 4.44E-09 |
| sp\|Q6B0K9\|HBM_HUMAN | 8 | 4 | 7 | 3 | -0.40272 | -0.008090311 | 0.606140441 | 0.394634111 | 1.008864862 | 0.614230751 | 0.621083832 | 0.159473245 | 0.401481465 |
| sp\|Q9UGT4\|SUSD2_HUMAN | 7 | 40 | 76 | 33 | 0.081278 | 0.003640831 | 0.057746134 | -0.077637221 | -0.023531918 | 0.054105303 | 0.321853466 | 0.929902155 | 0.614635933 |
| sp\|P15531\|NDKA_HUMAN | 42 | 40 | 76 | 33 | -0.33588 | -0.105492124 | 0.165339494 | 0.230392181 | 0.501223799 | 0.270831618 | 0.018963615 | 6.10E-06 | 0.008460693 |
| sp\|P07814\|SYEP_HUMAN | 3 | 37 | 69 | 30 | -0.3997 | -0.164149395 | 0.267859191 | 0.235550269 | 0.667558855 | 0.432008586 | 0.071741955 | 2.05E-06 | 0.000659917 |
| sp\|P42224\|STAT1_HUMAN | 39 | 18 | 34 | 15 | -0.54305 | -0.223130075 | 0.242940656 | 0.319924791 | 0.785995522 | 0.46607073 | 0.227878284 | 0.003261471 | 0.066477687 |
| sp\|Q5JS37\|NHLC3_HUMAN | 10 | 33 | 62 | 27 | -0.08304 | -0.025850085 | 0.172017985 | 0.057191655 | 0.255059725 | 0.19786807 | 0.600036576 | 0.001459504 | 0.006342873 |
| sp\|P67870\|CSK2B_HUMAN | 10 | 23 | 41 | 18 | 0.058679 | -0.009287538 | 0.103138239 | -0.067966945 | 0.044458831 | 0.112425776 | 0.608346739 | 0.863653295 | 0.319094209 |
| sp\|P19367\|HXK1_HUMAN | 80 | 33 | 62 | 27 | 0.088627 | -0.422535215 | -0.136585828 | -0.511162585 | -0.225213198 | 0.285949387 | 8.25E-05 | 0.251927516 | 0.062744005 |
| sp\|P62826\|RAN_HUMAN | 21 | 40 | 76 | 33 | -0.297 | -0.071188584 | 0.288181641 | 0.225809511 | 0.585179737 | 0.359370225 | 0.02940403 | 4.00E-07 | 0.000530778 |
| sp\|P01857\|IGHG1_HUMAN | 2 | 40 | 76 | 33 | -0.4312 | -0.21572312 | 0.223048232 | 0.215473856 | 0.654245209 | 0.438771352 | 0.273795693 | 0.000439581 | 0.01044863 |
| sp\|Q96S96\|PEBP4_HUMAN | 1 | 40 | 76 | 33 | 0.127081 | 0.209943017 | 0.276355338 | 0.082861699 | 0.14927402 | 0.066412321 | 0.450391533 | 0.170679857 | 0.636658889 |
| sp\|Q8TAX7\|MUC7_HUMAN | 2 | 11 | 21 | 9 | 0.015322 | -0.165687424 | -0.171298826 | -0.181009427 | -0.186620829 | -0.005611402 | 0.384869789 | 0.495922008 | 0.999176004 |
| sp\|Q92542\|NICA_HUMAN | 3 | 15 | 27 | 12 | 0.021522 | 0.026244841 | 0.126217834 | 0.004723223 | 0.104696217 | 0.099972993 | 0.998672114 | 0.639758103 | 0.602415694 |
| sp\|Q9UBS4\|DJB11_HUMAN | 13 | 23 | 41 | 17 | -0.14516 | -0.076520817 | 0.098988255 | 0.06864194 | 0.244151011 | 0.175509071 | 0.755025099 | 0.102095878 | 0.23013643 |
| sp\|P98172\|EFNB1_HUMAN | 2 | 40 | 76 | 33 | -0.28156 | -0.138339178 | 0.327424692 | 0.143217326 | 0.608981196 | 0.46576387 | 0.261867052 | 3.99E-07 | 1.24E-05 |
| sp\|P41218\|MNDA_HUMAN | 1 | 14 | 28 | 12 | -0.46228 | -0.447059714 | 0.514861471 | 0.015219545 | 0.97714073 | 0.961921185 | 0.998453653 | 0.017920809 | 0.007020261 |
| sp\|P28062\|PSB8_HUMAN | 5 | 40 | 76 | 33 | -0.39403 | -0.239334468 | 0.273672313 | 0.15469756 | 0.667704341 | 0.513006781 | 0.292010193 | 8.18E-07 | 1.98E-05 |
| sp\|Q15818\|NPTX1_HUMAN | 21 | 36 | 69 | 31 | 0.190353 | 0.135726554 | 0.065250113 | -0.054626766 | -0.125103208 | -0.070476442 | 0.842588024 | 0.53301142 | 0.772971672 |
| sp\|P31944\|CASPE_HUMAN | 1 | 30 | 56 | 24 | -0.29526 | -0.283982978 | -0.552497572 | 0.011280201 | -0.257234393 | -0.268514594 | 0.99844969 | 0.578832799 | 0.473004689 |
| sp\|Q99784\|NOE1_HUMAN | 26 | 40 | 76 | 33 | 0.177181 | 0.152502415 | 0.113748594 | -0.024678471 | -0.063432293 | -0.038753821 | 0.735047761 | 0.249762111 | 0.514820136 |
| sp\|P17538\|CTRB1_HUMAN | 33 | 40 | 76 | 33 | -0.0701 | -0.022771653 | 0.062959961 | 0.0473281 | 0.133059713 | 0.085731614 | 0.765550044 | 0.237218991 | 0.465127684 |
| sp\|P00326\|ADH1G_HUMAN | 10 | 30 | 55 | 23 | -0.6153 | -0.26542586 | 0.073738383 | 0.349871886 | 0.68903613 | 0.339164243 | 0.144761377 | 0.007881176 | 0.217514799 |
| sp\|Q9P2T1\|GMPR2_HUMAN | 3 | 40 | 76 | 33 | -0.08769 | -0.066912386 | 0.281172952 | 0.020781021 | 0.368866359 | 0.348085338 | 0.984859307 | 0.040445107 | 0.026788774 |
| sp\|P55083\|MFAP4_HUMAN | 8 | 37 | 69 | 30 | 0.007503 | -0.04250214 | 0.263539282 | -0.050005435 | 0.256035988 | 0.306041422 | 0.82873403 | 0.037870349 | 0.003182575 |
| sp\|P02462\|CO4A1_HUMAN | 1 | 36 | 69 | 30 | -0.05744 | -0.048157983 | 0.190623327 | 0.009286888 | 0.248068198 | 0.23878131 | 0.99549375 | 0.113591109 | 0.07698687 |
| sp\|A6NGU5\|GGT3_HUMAN | 3 | 40 | 76 | 33 | -0.26162 | -0.205558179 | 0.150958776 | 0.056058179 | 0.412574752 | 0.356516574 | 0.847254422 | 0.002890931 | 0.003811102 |
| sp\|P14778\|IL1R1_HUMAN | 3 | 40 | 76 | 33 | 0.000684 | 0.056083913 | -0.003616071 | 0.055399805 | -0.004300179 | -0.059699985 | 0.357511775 | 0.995686498 | 0.350464111 |
| sp\|Q9GZP4\|PITH1_HUMAN | 3 | 40 | 76 | 33 | -0.29993 | -0.108672522 | 0.270153336 | 0.191256853 | 0.570082711 | 0.378825858 | 0.09776038 | 2.50E-06 | 0.00049873 |
| sp\|Q8IX30\|SCUB3_HUMAN | 1 | 37 | 69 | 30 | 0.001259 | -0.169032603 | 0.147625731 | -0.170291375 | 0.146366959 | 0.316658334 | 0.067743333 | 0.249615222 | 0.00046663 |
| sp\|Q9ULC4\|MCTS1_HUMAN | 1 | 26 | 48 | 20 | -0.22348 | 0.054931252 | 0.419521599 | 0.27841284 | 0.643003188 | 0.364590347 | 0.243280642 | 0.008120327 | 0.133781405 |
| sp\|P14222\|PERF_HUMAN | 8 | 40 | 76 | 33 | 0.091129 | 0.105622314 | 0.058832556 | 0.014493369 | -0.032296389 | -0.046789757 | 0.941138248 | 0.812596422 | 0.575673515 |
| sp\|Q495W5\|FUT11_HUMAN | 2 | 40 | 76 | 33 | 0.108765 | 0.086488866 | 0.138954473 | -0.022275827 | 0.03018978 | 0.052465607 | 0.863583823 | 0.830445182 | 0.491624175 |
| sp\|Q9UJA9\|ENPP5_HUMAN | 1 | 16 | 27 | 11 | 0.176317 | -0.007929412 | -0.105852776 | -0.184246362 | -0.282169726 | -0.097923364 | 0.02915102 | 0.005625011 | 0.43720768 |
| sp\|Q9Y644\|RFNG_HUMAN | 16 | 15 | 28 | 13 | 0.159421 | 0.129199002 | 0.151982507 | -0.030222233 | -0.007438728 | 0.022783504 | 0.921348444 | 0.996461479 | 0.958530727 |
| sp\|Q92485\|ASM3B_HUMAN | 16 | 12 | 20 | 8 | -0.03864 | -0.0625871 | 0.188787148 | -0.023946077 | 0.227428171 | 0.251374248 | 0.968131201 | 0.170043955 | 0.080616448 |
| sp\|P41439\|FOLR3_HUMAN | 57 | 40 | 76 | 33 | -0.32058 | -0.075712633 | 0.147568776 | 0.244870686 | 0.468152095 | 0.223281409 | 0.032665611 | 0.000265268 | 0.080405976 |
| sp\|P55209\|NP1L1_HUMAN | 1 | 40 | 76 | 33 | -0.255 | -0.319879806 | -0.102840044 | -0.06488267 | 0.152157092 | 0.217039763 | 0.923251092 | 0.739122803 | 0.459004066 |
| sp\|Q13555\|KCC2G_HUMAN | 17 | 37 | 69 | 30 | 0.014003 | 0.094399197 | -0.083040352 | 0.080395908 | -0.09704364 | -0.177439549 | 0.236327209 | 0.235578602 | 0.002968289 |
| sp\|P50281\|MMP14_HUMAN | 11 | 14 | 27 | 12 | -0.20374 | 0.097454623 | 0.788683657 | 0.301192763 | 0.992421797 | 0.691229034 | 0.166622672 | 1.66E-05 | 0.000580507 |
| sp\|O00264\|PGRC1_HUMAN | 67 | 40 | 76 | 33 | -0.03511 | -0.007208331 | 0.240999601 | 0.027904541 | 0.276112473 | 0.248207932 | 0.881225137 | 0.000370234 | 0.000300816 |
| sp\|P62256\|UBE2H_HUMAN | 52 | 4 | 7 | 2 | -0.0076 | -0.056069565 | -0.372923779 | -0.04847069 | -0.365324904 | -0.316854214 | 0.982023822 | 0.599046316 | 0.635997002 |
| sp\|Q5VVQ6\|OTU1_HUMAN | 153 | 37 | 69 | 30 | -0.02808 | 0.029818186 | 0.2020785 | 0.057895415 | 0.230155729 | 0.172260314 | 0.736433241 | 0.039955976 | 0.100311919 |
| sp\|Q9NR19\|ACSA_HUMAN | 2 | 14 | 28 | 12 | 0.061823 | 0.145390544 | 0.494910268 | 0.083567671 | 0.433087394 | 0.349519723 | 0.9023794 | 0.159763774 | 0.209554588 |
| sp\|Q13283\|G3BP1_HUMAN | 7 | 18 | 34 | 15 | -0.24801 | -0.116212096 | 0.210492719 | 0.131793292 | 0.458498106 | 0.326704814 | 0.678626045 | 0.045227368 | 0.129964915 |
| sp\|Q7Z5L0\|VMO1_HUMAN | 12 | 40 | 76 | 33 | -0.48779 | -0.346312225 | 0.302235796 | 0.141479445 | 0.790027466 | 0.648548021 | 0.389751659 | 2.79E-08 | 2.51E-07 |
| sp\|P54760\|EPHB4_HUMAN | 7 | 40 | 76 | 33 | 0.020349 | 0.043549568 | 0.263517066 | 0.023201049 | 0.243168548 | 0.219967498 | 0.906044711 | 0.000933773 | 0.000713406 |
| sp\|P05997\|CO5A2_HUMAN | 2 | 40 | 76 | 33 | 0.037691 | 0.058499135 | 0.289055753 | 0.020807701 | 0.251364319 | 0.230556618 | 0.942025891 | 0.003420499 | 0.002343335 |
| sp\|Q15008\|PSMD6_HUMAN | 619 | 15 | 28 | 11 | -0.06252 | -0.023407954 | 0.247308746 | 0.039112439 | 0.309829139 | 0.2707167 | 0.956671171 | 0.176269652 | 0.191607521 |
| sp\|O75503\|CLN5_HUMAN | 11 | 40 | 76 | 33 | 0.043991 | -0.017057598 | 0.148423176 | -0.061048272 | 0.104432502 | 0.165480774 | 0.56850297 | 0.321922065 | 0.02922191 |
| sp\|P16444\|DPEP1_HUMAN | 5 | 37 | 69 | 30 | 0.139027 | 0.064790629 | -0.004655915 | -0.074236567 | -0.143683111 | -0.069446544 | 0.567073475 | 0.235192677 | 0.64945087 |
| sp\|Q9UI15\|TAGL3_HUMAN | 7 | 40 | 76 | 33 | -0.0829 | -0.135715719 | 0.027849623 | -0.052819844 | 0.110745497 | 0.163565342 | 0.734849682 | 0.39520895 | 0.079336418 |
| sp\|Q86X76\|NIT1_HUMAN | 28 | 25 | 49 | 21 | -0.37041 | -0.073087753 | 0.342479941 | 0.297319183 | 0.712886876 | 0.415567694 | 0.071578166 | 7.41E-05 | 0.011581595 |
| sp\|P12931\|SRC_HUMAN | 62 | 40 | 76 | 33 | -0.07476 | -0.035906568 | 0.045051929 | 0.038854162 | 0.11981266 | 0.080958497 | 0.927276868 | 0.610690518 | 0.750395579 |
| sp\|O14894\|T4S5_HUMAN | 31 | 4 | 6 | 2 | -2.1392 | -1.44299171 | 2.694655742 | 0.696203907 | 4.833851359 | 4.137647452 | 0.824130384 | 0.030940855 | 0.048284016 |
| sp\|P51606\|RENBP_HUMAN | 12 | 18 | 35 | 16 | -0.17698 | -0.037077856 | 0.042047227 | 0.139903728 | 0.21902881 | 0.079125082 | 0.557821874 | 0.363750515 | 0.840444034 |
| sp\|P25774\|CATS_HUMAN | 5 | 40 | 76 | 33 | -0.10333 | -0.047667696 | 0.128174033 | 0.055663806 | 0.231505534 | 0.175841728 | 0.475070825 | 0.000262648 | 0.002085542 |
| sp\|P32320\|CDD_HUMAN | 19 | 36 | 69 | 30 | 0.11084 | 0.015629004 | 0.165220549 | -0.095210976 | 0.054380569 | 0.149591545 | 0.240514 | 0.722118239 | 0.047193724 |
| sp\|Q8TDF6\|GRP4_HUMAN | 1 | 25 | 48 | 22 | -0.40481 | -0.677193103 | -0.155136895 | -0.272380683 | 0.249673724 | 0.522054408 | 0.424684728 | 0.597608399 | 0.060390887 |
| sp\|Q96CG8\|CTHR1_HUMAN | 16 | 32 | 62 | 27 | -0.30181 | -0.208142301 | 0.246107618 | 0.093670966 | 0.547920885 | 0.454249919 | 0.586841248 | 1.36E-05 | 4.45E-05 |
| sp\|Q8NFL0\|B3GN7_HUMAN | 4 | 40 | 76 | 33 | -0.02187 | -0.014173325 | 0.064567211 | 0.007694633 | 0.086435169 | 0.078740536 | 0.97169524 | 0.087391374 | 0.076509283 |
| sp\|Q9H1R3\|MYLK2_HUMAN | 1 | 4 | 6 | 3 | -0.17246 | -0.219235197 | 0.972555048 | -0.046777278 | 1.145012968 | 1.191790246 | 0.995107815 | 0.174881463 | 0.120514641 |
| sp\|Q9H3U1\|UN45A_HUMAN | 4 | 29 | 56 | 24 | -0.73465 | -0.434990619 | 0.036316796 | 0.299664234 | 0.770971649 | 0.471307415 | 0.362817815 | 0.012088678 | 0.113930304 |
| sp\|P07355\|ANXA2_HUMAN | 2 | 17 | 35 | 15 | -0.07833 | 0.168711042 | 0.318798201 | 0.24704006 | 0.397127219 | 0.150087159 | 0.314229511 | 0.129002403 | 0.671706483 |
| sp\|P40763\|STAT3_HUMAN | 13 | 14 | 28 | 12 | -0.01819 | 0.047415804 | 0.129219358 | 0.065608137 | 0.147411691 | 0.081803554 | 0.801622273 | 0.466253763 | 0.734368483 |
| sp\|P08311\|CATG_HUMAN | 4 | 40 | 76 | 33 | -0.61053 | -0.505461915 | 0.057359062 | 0.105064436 | 0.667885413 | 0.562820977 | 0.624078996 | 7.65E-06 | 2.16E-05 |
| sp\|Q7RTY9\|PRS41_HUMAN | 3 | 40 | 76 | 33 | 0.06036 | 0.057634755 | 0.152716568 | -0.002725124 | 0.092356688 | 0.095081812 | 0.999234921 | 0.547348417 | 0.444408017 |
| sp\|Q9BQR3\|PRS27_HUMAN | 61 | 4 | 7 | 3 | -0.09488 | -0.026759395 | -0.179417772 | 0.068124751 | -0.084533626 | -0.152658376 | 0.78777819 | 0.781020603 | 0.396578781 |
| sp\|Q9H3G5\|CPVL_HUMAN | 15 | 40 | 76 | 33 | 0.056576 | -0.032582574 | -0.082632422 | -0.089158645 | -0.139208493 | -0.050049848 | 0.323241247 | 0.151877898 | 0.729664094 |
| sp\|P42773\|CDN2C_HUMAN | 6 | 15 | 27 | 11 | 0.022155 | 0.162193723 | 0.384696832 | 0.14003905 | 0.36254216 | 0.222503109 | 0.273706802 | 0.005447949 | 0.076360838 |
| sp\|Q15375\|EPHA7_HUMAN | 47 | 40 | 76 | 33 | -0.01111 | 0.019853158 | 0.182256575 | 0.030961321 | 0.193364738 | 0.162403417 | 0.875802584 | 0.031814418 | 0.044799293 |
| sp\|P01876\|IGHA1_HUMAN | 2 | 40 | 76 | 33 | -0.43794 | -0.376192359 | 0.223296853 | 0.061747113 | 0.661236325 | 0.599489211 | 0.88564708 | 0.000148826 | 0.000102487 |
| sp\|P54710\|ATNG_HUMAN | 2 | 4 | 6 | 3 | -0.29558 | -0.129104609 | 0.013977829 | 0.166471712 | 0.30955415 | 0.143082439 | 0.858521572 | 0.691909194 | 0.909844206 |
| sp\|Q16186\|ADRM1_HUMAN | 35 | 11 | 20 | 9 | -0.0378 | -0.155216405 | 0.603078117 | -0.117412997 | 0.640881526 | 0.758294523 | 0.78590973 | 0.012239329 | 0.000815503 |
| sp\|P50991\|TCPD_HUMAN | 28 | 30 | 55 | 25 | -0.26603 | -0.211915712 | 0.078792343 | 0.054118046 | 0.344826101 | 0.290708055 | 0.901973767 | 0.058132516 | 0.077457711 |
| sp\|Q99542\|MMP19_HUMAN | 2 | 40 | 76 | 33 | -0.34677 | -0.273336252 | 0.196054784 | 0.073429018 | 0.542820055 | 0.469391036 | 0.485959896 | 1.96E-10 | 4.92E-10 |
| sp\|Q15223\|NECT1_HUMAN | 6 | 40 | 76 | 33 | 0.11228 | 0.128504585 | -0.049449923 | 0.01622428 | -0.161730227 | -0.177954508 | 0.913955494 | 0.002972115 | 0.000163162 |
| sp\|Q3T906\|GNPTA_HUMAN | 2 | 4 | 7 | 3 | 0.144514 | 0.201437318 | -0.177453336 | 0.056923356 | -0.321967298 | -0.378890655 | 0.787209321 | 0.025528085 | 0.005173508 |
| sp\|O14798\|TR10C_HUMAN | 1 | 40 | 76 | 33 | -0.11044 | -0.175894711 | 0.321816665 | -0.065457333 | 0.432254042 | 0.497711375 | 0.825754441 | 0.004260005 | 0.000134777 |
| sp\|P27701\|CD82_HUMAN | 10 | 29 | 55 | 24 | -0.38176 | -0.141851361 | 0.372561538 | 0.239905844 | 0.754318744 | 0.514412899 | 0.133255916 | 5.19E-06 | 0.000499796 |
| sp\|Q14061\|COX17_HUMAN | 9 | 7 | 13 | 7 | -0.32402 | -0.29588121 | 0.593901935 | 0.028140254 | 0.917923399 | 0.889783145 | 0.997238392 | 0.127522045 | 0.084698843 |
| sp\|P31947\|1433S_HUMAN | 14 | 7 | 14 | 6 | -0.98284 | -0.815716758 | -0.454135171 | 0.167125488 | 0.528707075 | 0.361581586 | 0.592734372 | 0.040640684 | 0.12826348 |
| sp\|P09429\|HMGB1_HUMAN | 1 | 14 | 28 | 12 | -0.47853 | -0.331578184 | 0.61092487 | 0.146949607 | 1.08945266 | 0.942503053 | 0.859705883 | 0.005943258 | 0.006730612 |
| sp\|Q02790\|FKBP4_HUMAN | 1 | 22 | 41 | 17 | -0.34895 | -0.221588886 | 0.137548868 | 0.127363418 | 0.486501172 | 0.359137754 | 0.662477777 | 0.02226784 | 0.070813737 |
| sp\|P04156\|PRIO_HUMAN | 10 | 40 | 76 | 33 | -0.14898 | -0.193623945 | 0.211614635 | -0.044641752 | 0.360596828 | 0.40523858 | 0.821682368 | 0.000284474 | 3.36E-06 |
| sp\|Q8TAC1\|RFESD_HUMAN | 5 | 4 | 7 | 3 | -0.23068 | 0.490508985 | 0.185926449 | 0.721191401 | 0.416608864 | -0.304582536 | 0.229601569 | 0.692059509 | 0.783566162 |
| sp\|P21291\|CSRP1_HUMAN | 12 | 40 | 76 | 33 | -0.21184 | -0.235518009 | -0.000997138 | -0.02367327 | 0.2108476 | 0.234520871 | 0.984781158 | 0.435129161 | 0.271589766 |
| sp\|P04746\|AMYP_HUMAN | 1 | 40 | 76 | 33 | -0.00471 | 0.030860542 | 0.016209672 | 0.035568713 | 0.020917843 | -0.014650871 | 0.941116386 | 0.985614936 | 0.99099549 |
| sp\|P05107\|ITB2_HUMAN | 9 | 40 | 76 | 33 | -0.10672 | -0.018046674 | 0.107745021 | 0.088674903 | 0.214466598 | 0.125791695 | 0.094217886 | 0.000130819 | 0.016565576 |
| sp\|P52849\|NDST2_HUMAN | 3 | 12 | 20 | 9 | 0.058896 | 0.12117663 | 0.175257786 | 0.062281048 | 0.116362204 | 0.054081156 | 0.762575016 | 0.526470375 | 0.843949889 |
| sp\|P50851\|LRBA_HUMAN | 3 | 4 | 7 | 3 | -0.26951 | -0.030608003 | 0.674677814 | 0.238906536 | 0.944192353 | 0.705285817 | 0.676525719 | 0.043527638 | 0.097816647 |
| sp\|P14868\|SYDC_HUMAN | 4 | 40 | 76 | 33 | -0.37456 | -0.277898388 | 0.059468622 | 0.096666584 | 0.434033594 | 0.33736701 | 0.632036521 | 0.002403155 | 0.009118843 |
| sp\|Q9H3R1\|NDST4_HUMAN | 9 | 19 | 34 | 15 | -0.28799 | 0.014444225 | -0.150349268 | 0.302432797 | 0.137639304 | -0.164793493 | 0.002739912 | 0.39748484 | 0.197816381 |
| sp\|Q6UXH9\|PAMR1_HUMAN | 1 | 40 | 76 | 33 | -0.00027 | -0.016141911 | -0.006994618 | -0.01587588 | -0.006728587 | 0.009147293 | 0.90921999 | 0.98826382 | 0.972622927 |

| | | | | | | | | | | | | |
|---|---|---|---|---|---|---|---|---|---|---|---|---|
| sp\|Q9H2X0\|CHRD_HUMAN | 1 | 8 | 13 | 6 | 0.182855 | 0.085112697 | -0.038705507 | -0.097742005 | -0.221560209 | -0.123818205 | 0.658885002 | 0.242314743 | 0.576039218 |
| sp\|P01861\|IGHG4_HUMAN | 18 | 40 | 76 | 33 | -0.33895 | -0.141077692 | -0.098058106 | 0.197872395 | 0.240891981 | 0.043019587 | 0.105971624 | 0.100846328 | 0.909116626 |
| sp\|Q9UK05\|GDF2_HUMAN | 12 | 37 | 69 | 29 | -0.01669 | -0.015234448 | 0.103586624 | 0.001450623 | 0.120271695 | 0.118821072 | 0.999509997 | 0.108815415 | 0.066907094 |
| sp\|P61604\|CH10_HUMAN | 1 | 33 | 62 | 28 | -0.14643 | -0.003085354 | 0.443787702 | 0.143345481 | 0.590218536 | 0.446873056 | 0.547058852 | 0.001228041 | 0.006823874 |
| sp\|Q9GZM7\|TINAL_HUMAN | 16 | 29 | 55 | 24 | -0.20839 | -0.101380621 | 0.017524449 | 0.107005289 | 0.225910359 | 0.11890507 | 0.438349493 | 0.083122545 | 0.408487715 |
| sp\|Q9Y235\|ABEC2_HUMAN | 3 | 4 | 6 | 3 | 0.242025 | 0.007803717 | 1.895791247 | -0.234221647 | 1.653765883 | 1.887987531 | 0.963732484 | 0.311301933 | 0.186558221 |
| sp\|O60449\|LY75_HUMAN | 8 | 40 | 76 | 33 | -0.02348 | 0.037700989 | -0.041825469 | 0.061181441 | -0.018345017 | -0.079526458 | 0.525739084 | 0.960652524 | 0.386481259 |
| sp\|P01893\|HLAH_HUMAN | 90 | 22 | 41 | 19 | -0.24261 | -0.173590239 | 0.152786297 | 0.069018128 | 0.395394664 | 0.326376536 | 0.854238411 | 0.030566166 | 0.047463448 |
| sp\|Q9H7M9\|VISTA_HUMAN | 6 | 15 | 27 | 11 | -0.33166 | -0.115404578 | 0.05805084 | 0.216254246 | 0.389709665 | 0.173455418 | 0.222997285 | 0.04554505 | 0.451255098 |
| sp\|Q06124\|PTN11_HUMAN | 1 | 40 | 76 | 33 | -0.37647 | -0.011416875 | 0.346261433 | 0.365050437 | 0.722728746 | 0.357678309 | 0.00273989 | 4.02E-07 | 0.006631412 |
| sp\|P36959\|GMPR1_HUMAN | 4 | 31 | 55 | 23 | 0.028318 | 0.027789471 | 0.286400627 | -0.000528422 | 0.258082734 | 0.258611156 | 0.999993354 | 0.352960804 | 0.277937848 |
| sp\|Q14242\|SELPL_HUMAN | 11 | 28 | 56 | 25 | 0.148259 | 0.021884195 | 0.241575491 | -0.126374608 | 0.093316688 | 0.219691296 | 0.504884629 | 0.767108915 | 0.152199791 |
| sp\|P30520\|PURA2_HUMAN | 1 | 28 | 56 | 24 | -0.24482 | -0.114437384 | 0.310701804 | 0.130386215 | 0.555525402 | 0.425139188 | 0.50875674 | 0.000425156 | 0.002389361 |
| sp\|Q9BU40\|CRDL1_HUMAN | 12 | 36 | 69 | 30 | -0.0169 | -0.058748148 | 0.207767129 | -0.041846156 | 0.224669121 | 0.266515277 | 0.726195483 | 0.002487707 | 3.29E-05 |
| sp\|Q14982\|OPCM_HUMAN | 2 | 40 | 76 | 33 | 0.123098 | -0.032612387 | -0.058512219 | -0.155709964 | -0.181609796 | -0.025899832 | 0.028500907 | 0.035206729 | 0.914296645 |
| sp\|O60888\|CUTA_HUMAN | 4 | 40 | 76 | 33 | 0.16433 | 0.264993916 | 0.027209957 | 0.100663627 | -0.137120332 | -0.237783959 | 0.170071818 | 0.1047041 | 0.000288784 |
| sp\|P13693\|TCTP_HUMAN | 6 | 40 | 76 | 33 | -0.06581 | -0.053523713 | 0.26422203 | 0.012286383 | 0.330032126 | 0.317745743 | 0.989859795 | 0.007999429 | 0.003519508 |
| sp\|Q8N3J6\|CADM2_HUMAN | 1 | 32 | 62 | 28 | 0.051778 | -0.104393131 | -0.131568075 | -0.156171168 | -0.183346113 | -0.027174945 | 0.056605723 | 0.060649286 | 0.921111379 |
| sp\|O95450\|ATS2_HUMAN | 11 | 19 | 34 | 13 | -0.70765 | -0.580662711 | 0.832171499 | 0.12698542 | 1.539819629 | 1.41283421 | 0.869422204 | 2.34E-05 | 1.86E-05 |
| sp\|Q06418\|TYRO3_HUMAN | 9 | 40 | 76 | 33 | 0.083125 | 0.017367041 | 0.001555692 | -0.065757619 | -0.081568968 | -0.015811349 | 0.261010383 | 0.24055139 | 0.933223897 |
| sp\|P36888\|FLT3_HUMAN | 1 | 31 | 55 | 23 | 0.175336 | 0.272050175 | -0.044271096 | 0.09671467 | -0.219606602 | -0.316321271 | 0.465406365 | 0.076887391 | 0.001944089 |
| sp\|O60568\|PLOD3_HUMAN | 37 | 40 | 76 | 33 | -0.08285 | 0.032384678 | 0.271720659 | 0.11523962 | 0.354575601 | 0.239335981 | 0.130314053 | 5.60E-06 | 0.000659728 |
| sp\|Q9P289\|STK26_HUMAN | 3 | 40 | 76 | 33 | -0.09143 | -0.129272507 | 0.169489178 | -0.037840097 | 0.260921588 | 0.298761685 | 0.963041474 | 0.295228617 | 0.132860817 |
| sp\|P46109\|CRKL_HUMAN | 24 | 40 | 76 | 33 | 0.027208 | -0.010515511 | 0.268209234 | -0.037723226 | 0.24100152 | 0.278724745 | 0.911358465 | 0.078283512 | 0.014105917 |
| sp\|Q9UJ14\|GGT7_HUMAN | 2 | 40 | 76 | 33 | -0.07914 | -0.249036817 | -0.438043758 | -0.169895247 | -0.358902188 | -0.189006942 | 0.073883616 | 0.000483211 | 0.059333443 |
| sp\|O00233\|PSMD9_HUMAN | 2 | 32 | 62 | 27 | -0.21098 | -0.090593385 | 0.238470529 | 0.120390315 | 0.449454229 | 0.329063914 | 0.373195948 | 0.000163397 | 0.002081029 |
| sp\|Q9Y6Y9\|LY96_HUMAN | 15 | 33 | 62 | 27 | -0.23817 | -0.117661983 | 0.343416051 | 0.120509494 | 0.581587529 | 0.461078034 | 0.167827015 | 1.12E-10 | 5.79E-09 |
| sp\|Q8IYT4\|KATL2_HUMAN | 2 | 26 | 48 | 21 | -0.40884 | -0.407012088 | -0.147413072 | 0.001824908 | 0.261423924 | 0.259599016 | 0.99996663 | 0.625954463 | 0.559912078 |
| sp\|P46926\|GNPI1_HUMAN | 22 | 28 | 56 | 24 | -0.03629 | 0.051006322 | 0.274352523 | 0.087301207 | 0.310647408 | 0.223346201 | 0.536991503 | 0.005848324 | 0.0294107 |
| sp\|P21810\|PGS1_HUMAN | 1 | 25 | 49 | 22 | -0.20547 | -0.088656015 | 0.279986376 | 0.116816479 | 0.485458869 | 0.36864239 | 0.454663307 | 0.000178247 | 0.001315371 |
| sp\|Q86Z14\|KLOTB_HUMAN | 8 | 36 | 69 | 31 | -0.02803 | 0.048422217 | 0.045223113 | 0.076451837 | 0.073252733 | -0.003199104 | 0.649272544 | 0.756038452 | 0.999313097 |
| sp\|Q08209\|PP2BA_HUMAN | 5 | 8 | 14 | 6 | -0.17508 | 0.25556345 | -0.16362823 | 0.430645044 | 0.011453364 | -0.41919168 | 0.052972307 | 0.998407428 | 0.094733524 |
| sp\|Q8NCW5\|NNRE_HUMAN | 1 | 40 | 76 | 33 | 0.014373 | 0.086724741 | 0.331567667 | 0.07235222 | 0.317195147 | 0.244842926 | 0.429161794 | 3.59E-05 | 0.000361145 |
| sp\|O95394\|AGM1_HUMAN | 28 | 32 | 62 | 28 | -0.54778 | -0.204942061 | 0.322578634 | 0.342834157 | 0.870354852 | 0.527520695 | 0.008239257 | 6.43E-09 | 5.42E-05 |
| sp\|P14550\|AK1A1_HUMAN | 4 | 40 | 76 | 33 | -0.03364 | 0.036474016 | 0.386992757 | 0.070115287 | 0.420634028 | 0.350518741 | 0.567224357 | 3.56E-06 | 1.34E-05 |
| sp\|Q6UX06\|OLFM4_HUMAN | 13 | 25 | 48 | 21 | -0.24045 | -0.174527579 | 0.230457424 | 0.065924885 | 0.470909888 | 0.404985003 | 0.889475587 | 0.019727171 | 0.024064377 |
| sp\|Q8IX05\|CD302_HUMAN | 15 | 4 | 7 | 3 | 0.043217 | -0.044726091 | 0.815035297 | -0.087943161 | 0.771818228 | 0.859761388 | 0.979288917 | 0.371963041 | 0.23724257 |
| sp\|O60512\|B4GT3_HUMAN | 5 | 11 | 21 | 9 | 0.112409 | 0.29740452 | 0.490235625 | 0.18499541 | 0.377826514 | 0.192831105 | 0.591445058 | 0.23216581 | 0.607502815 |
| sp\|Q2TAA2\|IAH1_HUMAN | 3 | 29 | 55 | 24 | -0.2358 | -0.169052178 | 0.135837335 | 0.066745355 | 0.371634869 | 0.304889514 | 0.796551599 | 0.009978186 | 0.018737849 |
| sp\|Q13045\|FLII_HUMAN | 6 | 29 | 55 | 23 | -0.04968 | -0.161125844 | 0.211896459 | -0.111448424 | 0.261573879 | 0.373022303 | 0.769819509 | 0.38123858 | 0.087862519 |
| sp\|P24844\|MYL9_HUMAN | 161 | 10 | 21 | 9 | -0.37316 | -0.086718659 | 0.728294922 | 0.286446334 | 1.101459915 | 0.815013581 | 0.483456032 | 0.001792131 | 0.00804351 |
| sp\|Q9GZP0\|PDGFD_HUMAN | 3 | 36 | 69 | 30 | -0.1472 | 0.115682105 | 0.362801189 | 0.2628833 | 0.510002383 | 0.247119083 | 0.026355029 | 0.000128686 | 0.057051443 |
| sp\|Q15782\|CH3L2_HUMAN | 14 | 14 | 28 | 13 | 0.148082 | 0.115876526 | -0.035976594 | -0.032205096 | -0.184058216 | -0.15185312 | 0.951627586 | 0.319652662 | 0.358729182 |
| sp\|P26006\|ITA3_HUMAN | 3 | 25 | 48 | 22 | 0.063785 | -0.027806788 | 0.125544934 | -0.091592105 | 0.061759617 | 0.153351722 | 0.50182045 | 0.798753061 | 0.174343071 |
| sp\|Q9NR71\|ASAH2_HUMAN | 4 | 40 | 76 | 33 | -0.03685 | -0.06695104 | -0.306867687 | -0.030104086 | -0.270020733 | -0.239916647 | 0.960518065 | 0.112351584 | 0.111261433 |
| sp\|Q8N2Q7\|NLGN1_HUMAN | 12 | 11 | 21 | 10 | 0.159709 | 0.043791797 | 0.018372785 | -0.115916763 | -0.141335776 | -0.025419012 | 0.475958789 | 0.449534537 | 0.966366315 |
| sp\|Q92686\|NEUG_HUMAN | 2 | 40 | 76 | 33 | -0.35608 | -0.20345141 | 0.118201517 | 0.152632522 | 0.474285449 | 0.321652926 | 0.616731563 | 0.043612888 | 0.156068709 |
| sp\|P04626\|ERBB2_HUMAN | 3 | 40 | 76 | 33 | 0.166701 | 0.04311158 | -0.092623519 | -0.123589182 | -0.259324281 | -0.135735098 | 0.025863961 | 3.01E-05 | 0.021013667 |
| sp\|P10124\|SRGN_HUMAN | 3 | 40 | 76 | 33 | -0.26274 | 0.078045807 | 0.124099979 | 0.340781897 | 0.386836068 | 0.046054171 | 0.00484298 | 0.008520002 | 0.913760914 |
| sp\|P28066\|PSA5_HUMAN | 2 | 40 | 76 | 33 | -0.1923 | 0.005811276 | 0.547263422 | 0.198114162 | 0.739566308 | 0.541452146 | 0.143710559 | 8.16E-08 | 9.19E-06 |
| sp\|P50579\|MAP2_HUMAN | 61 | 8 | 13 | 6 | 0.324526 | 0.256840937 | 0.947388096 | -0.067684787 | 0.622862372 | 0.690547159 | 0.982243167 | 0.366429439 | 0.235103245 |
| sp\|P06132\|DCUP_HUMAN | 5 | 40 | 76 | 33 | -0.20517 | -0.107286474 | 0.215745765 | 0.097882211 | 0.42091445 | 0.323032239 | 0.551208602 | 0.000810631 | 0.004417019 |
| sp\|P35813\|PPM1A_HUMAN | 1 | 40 | 76 | 33 | -0.11144 | -0.141866174 | 0.120499451 | -0.030428141 | 0.231937483 | 0.262365624 | 0.922555779 | 0.043711825 | 0.006723818 |
| sp\|Q13616\|CUL1_HUMAN | 4 | 37 | 69 | 30 | -0.17642 | -0.096776994 | 0.191334752 | 0.07964018 | 0.367751926 | 0.288111746 | 0.524031985 | 0.000166045 | 0.001040921 |
| sp\|O43570\|CAH12_HUMAN | 2 | 4 | 7 | 3 | -0.3037 | -0.228098991 | 0.779589368 | 0.075604857 | 1.083293216 | 1.007688359 | 0.989153748 | 0.2650995 | 0.247175047 |
| sp\|P01266\|THYG_HUMAN | 7 | 29 | 55 | 24 | 0.358235 | 0.471453017 | 0.340072791 | 0.113217568 | -0.018162657 | -0.131380225 | 0.851819514 | 0.997143517 | 0.82698666 |
| sp\|O15067\|PUR4_HUMAN | 18 | 29 | 55 | 24 | -0.22361 | -0.036431833 | 0.465392684 | 0.187182719 | 0.689007237 | 0.501824518 | 0.267788196 | 1.74E-05 | 0.000457835 |
| sp\|Q9UQ52\|CNTN6_HUMAN | 1 | 18 | 35 | 15 | 0.204035 | 0.120331428 | 0.035233841 | -0.083703504 | -0.168801091 | -0.085097588 | 0.261921533 | 0.027492254 | 0.293516238 |
| sp\|O15117\|FYB1_HUMAN | 1 | 33 | 62 | 27 | -0.12514 | -0.100649507 | 0.122384711 | 0.024489813 | 0.247524031 | 0.223034218 | 0.978633496 | 0.224401551 | 0.215254224 |
| sp\|P68036\|UB2L3_HUMAN | 6 | 40 | 76 | 33 | -0.07117 | 0.130223237 | 0.426648097 | 0.201395764 | 0.497820625 | 0.296424861 | 0.105868755 | 0.000139999 | 0.015181779 |
| sp\|P12830\|CADH1_HUMAN | 14 | 40 | 76 | 33 | -0.03229 | -0.026784696 | 0.277443259 | 0.005505087 | 0.309733042 | 0.304227955 | 0.996158874 | 0.000420296 | 8.26E-05 |
| sp\|P32926\|DSG3_HUMAN | 5 | 8 | 14 | 6 | 0.068683 | 0.035822753 | -0.018120873 | -0.032860078 | -0.086803704 | -0.053943626 | 0.979193925 | 0.906219524 | 0.954380243 |
| sp\|P13861\|KAP2_HUMAN | 1 | 11 | 20 | 9 | -0.08651 | -0.059958663 | 0.495125969 | 0.026552931 | 0.581637563 | 0.555084632 | 0.986064973 | 0.015995798 | 0.009572349 |
| sp\|Q05315\|LEG10_HUMAN | 1 | 4 | 6 | 3 | 0.372435 | 0.222275542 | 0.223598758 | -0.150159235 | -0.14883602 | 0.001323215 | 0.922625785 | 0.944920251 | 0.999994745 |
| sp\|Q0ZGT2\|NEXN_HUMAN | 30 | 33 | 62 | 27 | -0.32762 | -0.332695008 | -0.013904212 | -0.005073094 | 0.313717702 | 0.318790797 | 0.999529794 | 0.294699021 | 0.203742827 |
| sp\|Q14203\|DCTN1_HUMAN | 24 | 11 | 21 | 9 | -0.14221 | 0.166154282 | 0.668519745 | 0.30836383 | 0.810729292 | 0.502365463 | 0.244620809 | 0.002903672 | 0.045071938 |
| sp\|P35611\|ADDA_HUMAN | 14 | 4 | 7 | 3 | -0.72214 | -1.12190512 | -0.758646192 | -0.399763296 | -0.036504368 | 0.363258928 | 0.889066329 | 0.999332496 | 0.922761621 |
| sp\|Q8IUK5\|PLDX1_HUMAN | 12 | 40 | 76 | 33 | 0.108717 | 0.06885278 | -0.066541023 | -0.039864461 | -0.175258264 | -0.135393803 | 0.463058787 | 8.04E-05 | 0.000673259 |
| sp\|Q16820\|MEP1B_HUMAN | 2 | 40 | 76 | 33 | -0.0459 | -0.054007058 | -0.203514096 | -0.008102569 | -0.157609608 | -0.149507039 | 0.99793505 | 0.584746029 | 0.5412468 |
| sp\|O60279\|SUSD5_HUMAN | 1 | 40 | 76 | 33 | -0.10143 | -0.238873002 | 0.062674858 | -0.137446364 | 0.164101496 | 0.30154786 | 0.026856804 | 0.028445199 | 9.25E-07 |
| sp\|Q16799\|RTN1_HUMAN | 4 | 4 | 7 | 3 | -0.02168 | 0.043119517 | 0.625568593 | 0.064799385 | 0.647248461 | 0.582449076 | 0.97712285 | 0.255204312 | 0.257678809 |
| sp\|O00232\|PSD12_HUMAN | 80 | 14 | 28 | 12 | -0.12468 | -0.036505771 | 0.582089491 | 0.088177622 | 0.706772883 | 0.618595262 | 0.81385769 | 0.00044972 | 0.000462118 |
| sp\|P41240\|CSK_HUMAN | 2 | 40 | 76 | 33 | -0.01294 | -0.186290873 | -0.09160977 | -0.173347315 | -0.078666211 | 0.094681103 | 0.223075739 | 0.805659679 | 0.671958838 |
| sp\|P54315\|LIPR1_HUMAN | 2 | 33 | 62 | 27 | -0.10534 | -0.135951751 | -0.05977855 | -0.030607828 | 0.045565373 | 0.076173201 | 0.97964084 | 0.969060018 | 0.894812414 |
| sp\|P28332\|ADH6_HUMAN | 2 | 16 | 27 | 11 | -0.90311 | -0.354525393 | 0.286371204 | 0.54858163 | 1.189478227 | 0.640896597 | 0.115295064 | 0.002408308 | 0.101564478 |
| sp\|Q9NYQ6\|CELR1_HUMAN | 19 | 18 | 34 | 15 | -0.13659 | -0.093917846 | -0.215411126 | 0.042676316 | -0.078816965 | -0.121493281 | 0.854631621 | 0.690094783 | 0.331114961 |
| sp\|P05120\|PAI2_HUMAN | 1 | 29 | 55 | 24 | -0.19388 | -0.127112589 | 0.059751664 | 0.066768509 | 0.253632762 | 0.186864253 | 0.76497861 | 0.074792756 | 0.163778126 |
| sp\|Q5VTT5\|MYOM3_HUMAN | 11 | 29 | 55 | 24 | 0.081978 | 0.006319462 | -0.037236663 | -0.075658903 | -0.119215029 | -0.043556126 | 0.699683911 | 0.542729105 | 0.900770764 |
| sp\|Q9Y275\|TN13B_HUMAN | 8 | 40 | 76 | 33 | -0.30633 | -0.209066726 | -0.093058088 | 0.097265363 | 0.213274002 | 0.116008638 | 0.347091224 | 0.032532751 | 0.267669309 |
| sp\|P30048\|PRDX3_HUMAN | 8 | 11 | 21 | 9 | -0.03329 | -0.025042596 | 0.191990004 | 0.008251078 | 0.225283678 | 0.2170326 | 0.996463644 | 0.179180119 | 0.133583593 |
| sp\|P04921\|GLPC_HUMAN | 12 | 3 | 7 | 4 | -0.24474 | -0.266212563 | 0.124731234 | -0.021475412 | 0.369468385 | 0.390943797 | 0.996410127 | 0.447293964 | 0.278772261 |
| sp\|Q9Y4G6\|TLN2_HUMAN | 101 | 26 | 48 | 21 | -0.14294 | -0.176675 | -0.016721784 | -0.033730261 | 0.126222955 | 0.159953216 | 0.980014445 | 0.823465048 | 0.676309993 |
| sp\|O00757\|F16P2_HUMAN | 5 | 40 | 76 | 33 | -0.1148 | 0.033058078 | 0.286048726 | 0.147854549 | 0.400845198 | 0.252990649 | 0.263480178 | 0.001617659 | 0.034815286 |
| sp\|Q9P121\|NTRI_HUMAN | 3 | 40 | 76 | 33 | 0.21461 | 0.102479298 | -0.073276308 | -0.112130715 | -0.28788632 | -0.175755605 | 0.014066348 | 3.26E-08 | 0.00014992 |
| sp\|Q8NFT8\|DNER_HUMAN | 3 | 40 | 76 | 33 | 0.236844 | 0.073399444 | -0.255630233 | -0.163444206 | -0.492473884 | -0.329029678 | 0.007712233 | 8.72E-12 | 1.54E-07 |
| sp\|Q9ULC0\|MUCEN_HUMAN | 2 | 37 | 69 | 30 | -0.09772 | -0.086262422 | 0.005451596 | 0.011458792 | 0.10317281 | 0.091714018 | 0.990855389 | 0.600865114 | 0.601685888 |
| sp\|P50895\|BCAM_HUMAN | 11 | 40 | 76 | 33 | 0.013681 | 0.023430703 | 0.209895876 | 0.009749974 | 0.196215147 | 0.186465173 | 0.977843841 | 0.00274851 | 0.001203388 |
| sp\|P31151\|S10A7_HUMAN | 2 | 37 | 69 | 30 | -1.68059 | -1.992991441 | -2.257660232 | -0.312400589 | -0.57706938 | -0.264668791 | 0.620946546 | 0.3294054 | 0.742652801 |
| sp\|P49746\|TSP3_HUMAN | 9 | 40 | 76 | 33 | 0.001888 | 0.212014652 | 0.17824857 | 0.210126297 | 0.176360216 | -0.033766082 | 0.038897881 | 0.200887458 | 0.926734376 |
| sp\|P30566\|PUR8_HUMAN | 2 | 40 | 76 | 33 | -0.6354 | 0.108032407 | 0.36597618 | 0.74343696 | 1.001380734 | 0.257943774 | 1.79E-11 | 1.59E-13 | 0.042221022 |
| sp\|P14621\|ACYP2_HUMAN | 2 | 18 | 34 | 15 | -0.28399 | -0.287815046 | 0.202076011 | -0.003820646 | 0.48607041 | 0.489891056 | 0.999576307 | 0.012480003 | 0.003916236 |
| sp\|P19438\|TNR1A_HUMAN | 29 | 37 | 69 | 30 | -0.23864 | -0.323303096 | 0.091646067 | -0.084660661 | 0.330288502 | 0.414949163 | 0.909676781 | 0.37485538 | 0.144578511 |
| sp\|Q99536\|VAT1_HUMAN | 2 | 40 | 76 | 33 | -0.08111 | -0.062827343 | 0.186661836 | 0.018280792 | 0.267769971 | 0.249489179 | 0.945724309 | 0.000474292 | 0.000225723 |
| sp\|P49327\|FAS_HUMAN | 3 | 22 | 41 | 18 | -0.29062 | 0.224961969 | 0.706437435 | 0.515584292 | 0.997059758 | 0.481475466 | 0.011660962 | 2.81E-05 | 0.031852475 |
| sp\|P42702\|LIFR_HUMAN | 2 | 32 | 62 | 27 | 0.085746 | 0.144455717 | 0.142746761 | 0.058709685 | 0.05700073 | -0.001708955 | 0.401368937 | 0.549179135 | 0.999303178 |
| sp\|A2VDF0\|FUCM_HUMAN | 4 | 25 | 48 | 22 | -0.38446 | -0.044865193 | 0.159846219 | 0.339591973 | 0.544303386 | 0.204711413 | 0.117190348 | 0.021752001 | 0.482000481 |
| sp\|P35998\|PRS7_HUMAN | 3 | 10 | 21 | 10 | -0.49071 | -0.109556582 | 0.156985397 | 0.381150142 | 0.647692121 | 0.266541979 | 0.123340634 | 0.015171584 | 0.347854313 |
| sp\|P48444\|COPD_HUMAN | 2 | 18 | 35 | 15 | -0.06113 | -0.074530274 | 0.733771976 | -0.013404913 | 0.794897337 | 0.80830225 | 0.998281179 | 0.020881342 | 0.006572625 |
| sp\|Q6QNK2\|AGRD1_HUMAN | 1 | 29 | 55 | 25 | 0.130587 | 0.090988347 | 0.363966674 | -0.039599121 | 0.233379206 | 0.272978327 | 0.798342164 | 0.005574721 | 0.000165078 |
| sp\|Q96BY9\|SARAF_HUMAN | 1 | 18 | 34 | 16 | 0.138896 | 0.061245739 | 0.212716605 | -0.077650699 | 0.073820167 | 0.151470866 | 0.742152004 | 0.823362451 | 0.355500927 |
| sp\|P26885\|FKBP2_HUMAN | 1 | 36 | 69 | 30 | -0.14415 | -0.152888245 | 0.269605419 | -0.008734915 | 0.413758749 | 0.422493664 | 0.996984395 | 0.011542569 | 0.002858338 |
| sp\|Q9UN36\|NDRG2_HUMAN | 1 | 14 | 27 | 12 | -0.39963 | -0.256772321 | 0.205171264 | 0.142857818 | 0.604801402 | 0.461943585 | 0.607540694 | 0.003859785 | 0.013665853 |
| sp\|O60547\|GMDS_HUMAN | 14 | 12 | 20 | 9 | -0.15569 | -0.13890428 | -0.049966111 | 0.016785553 | 0.105723722 | 0.088938169 | 0.986920439 | 0.701550224 | 0.738442332 |
| sp\|P01137\|TGFB1_HUMAN | 3 | 40 | 76 | 33 | -0.24918 | 0.047217375 | 0.154764372 | 0.296392413 | 0.40393941 | 0.107546997 | 0.001341859 | 0.000239049 | 0.444616017 |
| sp\|P40926\|MDHM_HUMAN | 24 | 37 | 69 | 29 | -0.12191 | -0.030462015 | 0.162068541 | 0.091448691 | 0.283979247 | 0.192530556 | 0.570871679 | 0.029284481 | 0.126048419 |

| | | | | | | | | | | | | |
|---|---|---|---|---|---|---|---|---|---|---|---|---|
| sp\|P24298\|ALAT1_HUMAN | 2 | 40 | 76 | 33 | -0.22069 | 0.004922728 | 0.086301778 | 0.225617688 | 0.306996739 | 0.08137905 | 0.065187415 | 0.03135812 | 0.726250917 |
| sp\|O75475\|PSIP1_HUMAN | 1 | 20 | 41 | 18 | -0.36977 | 0.15969219 | 1.310937882 | 0.529457366 | 1.680703057 | 1.151245692 | 0.401405811 | 0.002568678 | 0.021807454 |
| sp\|P43234\|CATO_HUMAN | 7 | 36 | 69 | 30 | 0.058253 | 0.17579594 | 0.304351003 | 0.117542675 | 0.246097737 | 0.128555062 | 0.161195767 | 0.004853083 | 0.145532943 |
| sp\|Q9Y5S9\|RBM8A_HUMAN | 7 | 7 | 14 | 7 | -0.13961 | 0.092546253 | 0.266089993 | 0.232158979 | 0.405702719 | 0.17354374 | 0.268736457 | 0.059857773 | 0.471089086 |
| sp\|Q9H8S9\|MOB1A_HUMAN | 2 | 40 | 76 | 33 | 0.162716 | 0.183810835 | 0.401354727 | 0.021094676 | 0.238638568 | 0.217543892 | 0.978260069 | 0.149169542 | 0.134021369 |
| sp\|Q96RP7\|G3ST4_HUMAN | 1 | 21 | 42 | 18 | 0.075034 | 0.068708069 | 0.16173594 | -0.006326043 | 0.086701828 | 0.093027871 | 0.995086049 | 0.530510405 | 0.389121467 |
| sp\|Q6UW63\|PLGT2_HUMAN | 1 | 4 | 7 | 3 | -0.01309 | -0.121271697 | 0.044090953 | -0.108186421 | 0.057176229 | 0.16536265 | 0.585407887 | 0.900351052 | 0.372386902 |
| sp\|Q9P1W8\|SIRPG_HUMAN | 3 | 25 | 48 | 22 | -0.60404 | -0.291472979 | -0.144479675 | 0.312568993 | 0.459562297 | 0.146993304 | 0.45905775 | 0.303934566 | 0.852599602 |
| sp\|O76061\|STC2_HUMAN | 1 | 40 | 76 | 33 | 0.052432 | 0.12979858 | 0.218512991 | 0.077366484 | 0.166080895 | 0.088714411 | 0.178626757 | 0.005076615 | 0.137645584 |
| sp\|P36575\|ARRC_HUMAN | 5 | 21 | 42 | 19 | 0.128884 | -0.006582434 | 0.21057501 | -0.135466822 | 0.081690622 | 0.217157444 | 0.420393285 | 0.796996348 | 0.129861383 |
| sp\|Q16539\|MK14_HUMAN | 2 | 40 | 76 | 33 | -0.06273 | -0.126872169 | 0.116823948 | -0.064141961 | 0.179554156 | 0.243696117 | 0.649846074 | 0.101720095 | 0.005503787 |
| sp\|P08134\|RHOC_HUMAN | 1 | 37 | 69 | 30 | 0.232162 | 0.23787826 | 0.513052361 | 0.005715872 | 0.280889973 | 0.2751741 | 0.998883996 | 0.162449887 | 0.111671811 |
| sp\|O14558\|HSPB6_HUMAN | 4 | 4 | 6 | 3 | 0.280811 | 0.292720133 | 1.45721134 | 0.011909172 | 1.176400378 | 1.164491207 | 0.999825301 | 0.337711319 | 0.294394821 |
| sp\|Q8IUZ5\|AT2L2_HUMAN | 7 | 40 | 76 | 33 | 0.046069 | 0.101838891 | 0.296409074 | 0.055769452 | 0.250339635 | 0.194570183 | 0.686961749 | 0.006877941 | 0.020858059 |
| sp\|Q9Y263\|PLAP_HUMAN | 3 | 29 | 55 | 24 | 0.115917 | 0.262442458 | 0.458112678 | 0.146525727 | 0.342195947 | 0.19567022 | 0.422097406 | 0.042537091 | 0.260569908 |
| sp\|Q9H993\|ARMT1_HUMAN | 5 | 30 | 55 | 23 | -0.01697 | 0.175451939 | 0.410839732 | 0.192422943 | 0.427810735 | 0.235387793 | 0.110557224 | 0.001025951 | 0.064938726 |
| sp\|P62333\|PRS10_HUMAN | 2 | 21 | 42 | 18 | -0.45219 | -0.157604876 | 0.184560324 | 0.29458413 | 0.636749329 | 0.3421652 | 0.054890582 | 0.000187279 | 0.030476958 |
| sp\|Q13162\|PRDX4_HUMAN | 2 | 36 | 69 | 30 | -0.36982 | -0.152910207 | 0.307768221 | 0.216905159 | 0.677583587 | 0.460678428 | 0.073164336 | 1.83E-07 | 6.21E-05 |
| sp\|Q8NFY4\|SEM6D_HUMAN | 23 | 40 | 76 | 33 | 0.005236 | -0.11288574 | -0.242259232 | -0.118122173 | -0.247495666 | -0.129373493 | 0.001092202 | 8.26E-09 | 0.000777655 |
| sp\|Q8IZP0\|ABI1_HUMAN | 1 | 39 | 76 | 33 | 0.059553 | 0.220400332 | 0.447801579 | 0.160847449 | 0.388248696 | 0.227401247 | 0.372992773 | 0.020902998 | 0.174526294 |
| sp\|Q9BQT9\|CSTN3_HUMAN | 2 | 36 | 69 | 30 | 0.00709 | 0.007990251 | 0.079781241 | 0.000900044 | 0.072691034 | 0.07179099 | 0.999849225 | 0.509103578 | 0.431782308 |
| sp\|P18065\|IBP2_HUMAN | 5 | 40 | 76 | 33 | 0.020491 | -0.004612951 | 0.602380258 | -0.0251042 | 0.581889008 | 0.606993208 | 0.964800005 | 7.19E-06 | 1.32E-07 |
| sp\|P31153\|METK2_HUMAN | 6 | 40 | 76 | 33 | -0.23302 | -0.054802912 | 0.247172094 | 0.178215554 | 0.48019056 | 0.301975006 | 0.05404848 | 1.74E-06 | 0.00085766 |
| sp\|Q9H1B5\|XYLT2_HUMAN | 1 | 40 | 76 | 33 | -0.28457 | -0.067928831 | 0.040873564 | 0.216645509 | 0.325447904 | 0.108802395 | 0.075910189 | 0.019091199 | 0.558243382 |
| sp\|Q08495\|DEMA_HUMAN | 1 | 28 | 55 | 24 | -0.25911 | -0.389182932 | -0.025443825 | -0.130077615 | 0.233661492 | 0.363739107 | 0.681486065 | 0.424685009 | 0.072694975 |
| sp\|Q13976\|KGP1_HUMAN | 6 | 6 | 14 | 7 | 0.239043 | -0.234895648 | 0.081841211 | -0.473938188 | -0.157201329 | 0.316736859 | 0.535218745 | 0.94714487 | 0.730043669 |
| sp\|Q14032\|BAAT_HUMAN | 1 | 11 | 20 | 9 | -0.22944 | -0.12664067 | 0.482436537 | 0.102797186 | 0.711874392 | 0.609077207 | 0.914105041 | 0.062939614 | 0.077683643 |
| sp\|Q9H741\|CL049_HUMAN | 1 | 4 | 7 | 3 | 0.231817 | 0.294387988 | 0.391859144 | 0.062570929 | 0.160042085 | 0.097471156 | 0.725969804 | 0.276633423 | 0.536478805 |
| sp\|Q8WWX9\|SELM_HUMAN | 1 | 4 | 6 | 3 | 0.137166 | -0.012428934 | 0.429091007 | -0.149594983 | 0.291924959 | 0.441519942 | 0.775549317 | 0.515700373 | 0.202205777 |
| sp\|Q9UBR1\|BUP1_HUMAN | 7 | 33 | 62 | 27 | 0.028697 | 0.216703407 | 0.288982576 | 0.188006818 | 0.260285987 | 0.072279169 | 0.223994957 | 0.139991455 | 0.821850931 |
| sp\|Q96C23\|GALM_HUMAN | 2 | 10 | 21 | 9 | -0.24404 | -0.023044637 | 0.179660018 | 0.22099985 | 0.423704505 | 0.202704655 | 0.421047918 | 0.117944011 | 0.506285385 |
| sp\|Q9Y240\|CLC11_HUMAN | 30 | 40 | 76 | 33 | -0.04901 | 0.00114192 | 0.246613364 | 0.05015039 | 0.295621834 | 0.245471444 | 0.694326861 | 0.000304609 | 0.000772365 |
| sp\|P28161\|GSTM2_HUMAN | 3 | 40 | 76 | 33 | -0.11142 | -0.174229337 | -0.008741912 | -0.062811477 | 0.102675947 | 0.165487425 | 0.73391634 | 0.566205338 | 0.156465959 |
| sp\|Q03013\|GSTM4_HUMAN | 2 | 14 | 28 | 13 | -0.80234 | -0.235094026 | -0.055229274 | 0.567250182 | 0.747114934 | 0.179864752 | 0.436964733 | 0.356167347 | 0.922600972 |
| sp\|Q99969\|RARR2_HUMAN | 2 | 40 | 76 | 33 | -0.0255 | 0.111585571 | 0.293267382 | 0.137082893 | 0.318764704 | 0.181681812 | 0.006545282 | 4.76E-08 | 0.00051647 |
| sp\|Q14210\|LY6D_HUMAN | 12 | 40 | 76 | 33 | -0.10494 | -0.184241268 | 0.140118478 | -0.07929921 | 0.245060536 | 0.324359747 | 0.68312555 | 0.085691426 | 0.004882964 |
| sp\|Q9HCK5\|AGO4_HUMAN | 1 | 4 | 7 | 3 | 0.013416 | 0.132972748 | 0.109029767 | 0.119557179 | 0.095614199 | -0.02394298 | 0.67686355 | 0.842254627 | 0.986701054 |
| sp\|P22307\|NLTP_HUMAN | 13 | 21 | 41 | 19 | 0.058689 | 0.058751366 | 0.338942892 | 6.28E-05 | 0.280254296 | 0.280191526 | 0.999999861 | 0.145275331 | 0.083157561 |
| sp\|P04216\|THY1_HUMAN | 7 | 40 | 76 | 33 | -0.06641 | -0.053586845 | 0.241981435 | 0.012825381 | 0.308393661 | 0.29556828 | 0.983992745 | 0.002349876 | 0.00090208 |
| sp\|P30405\|PPIF_HUMAN | 3 | 7 | 14 | 6 | -0.02023 | 0.15220586 | 0.158261015 | 0.172438463 | 0.178493618 | 0.006055155 | 0.873401716 | 0.904323994 | 0.999848863 |
| sp\|Q08752\|PPID_HUMAN | 1 | 7 | 14 | 6 | 0.241332 | 0.146491644 | 0.310160034 | -0.09484064 | 0.06882775 | 0.163668389 | 0.913722631 | 0.967552893 | 0.786484533 |
| sp\|P24855\|DNAS1_HUMAN | 4 | 22 | 42 | 17 | -0.04126 | -0.151651225 | -0.235034751 | -0.110393821 | -0.193777347 | -0.083383526 | 0.68562808 | 0.464206049 | 0.834367366 |
| sp\|Q14232\|EI2BA_HUMAN | 18 | 4 | 7 | 3 | -0.11417 | -0.063965437 | 0.37605825 | 0.050204428 | 0.490228115 | 0.440023687 | 0.972827306 | 0.216644991 | 0.220445207 |
| sp\|Q8TF64\|GIPC3_HUMAN | 1 | 4 | 7 | 3 | 0.114215 | 0.163210152 | -0.012509141 | 0.048994757 | -0.126724535 | -0.175719293 | 0.944643204 | 0.777372198 | 0.562700422 |
| sp\|Q9UMY4\|SNX12_HUMAN | 7 | 40 | 76 | 33 | 0.093435 | 0.034494576 | 0.398700906 | -0.058940181 | 0.305266149 | 0.364206329 | 0.75925359 | 0.007728124 | 0.000202171 |
| sp\|P32942\|ICAM3_HUMAN | 2 | 40 | 76 | 33 | 0.076718 | 0.187750998 | 0.123710186 | 0.111033104 | 0.046992291 | -0.064040812 | 0.049291722 | 0.682259464 | 0.407124358 |
| sp\|Q5JPE7\|NOMO2_HUMAN | 41 | 18 | 35 | 16 | 0.202384 | 0.172434703 | -0.08258028 | -0.029948927 | -0.28496391 | -0.255014983 | 0.896445649 | 0.001857207 | 0.001498576 |
| sp\|P11684\|UTER_HUMAN | 13 | 40 | 76 | 33 | -0.32314 | -0.244311999 | 0.258118634 | 0.078826976 | 0.581257609 | 0.502430633 | 0.728041175 | 2.14E-05 | 3.47E-05 |
| sp\|Q8N128\|F177A_HUMAN | 1 | 22 | 41 | 17 | 0.05478 | -0.055019418 | 0.257151708 | -0.109799905 | 0.202371221 | 0.312171126 | 0.551483995 | 0.26252905 | 0.021756227 |
| sp\|Q13557\|KCC2D_HUMAN | 7 | 40 | 76 | 33 | -0.11113 | 0.028483588 | -0.13666652 | 0.139611298 | -0.025538811 | -0.165150108 | 0.058531428 | 0.934765463 | 0.031279611 |
| sp\|Q13554\|KCC2B_HUMAN | 3 | 29 | 55 | 24 | -0.02179 | -0.02062228 | -0.272560382 | 0.001168306 | -0.250769796 | -0.251938102 | 0.999939503 | 0.152556571 | 0.090861178 |
| sp\|Q00266\|METK1_HUMAN | 3 | 4 | 7 | 3 | -0.54155 | -0.208530535 | 1.546266278 | 0.33302057 | 2.087817383 | 1.754796813 | 0.628211511 | 0.001399691 | 0.00240147 |
| sp\|Q8NCC3\|PAG15_HUMAN | 8 | 40 | 76 | 33 | 0.020723 | 0.030489969 | 0.103636272 | 0.009767341 | 0.082913644 | 0.073146303 | 0.976616436 | 0.312767776 | 0.31625804 |
| sp\|P62140\|PP1B_HUMAN | 11 | 34 | 62 | 26 | -0.04018 | -0.066069787 | 0.369521162 | -0.025892127 | 0.409698822 | 0.435590949 | 0.949148079 | 0.000336524 | 1.86E-05 |
| sp\|Q8NEU8\|DP13B_HUMAN | 6 | 18 | 34 | 14 | 0.008665 | -0.055424129 | 0.354647124 | -0.064089246 | 0.345982006 | 0.410071252 | 0.929856056 | 0.251061303 | 0.090993079 |
| sp\|P54727\|RD23B_HUMAN | 54 | 40 | 76 | 33 | -0.11863 | -0.169852183 | 0.264022466 | -0.051221444 | 0.382653205 | 0.433874649 | 0.845620458 | 0.002288085 | 6.63E-05 |
| sp\|Q9BY32\|ITPA_HUMAN | 1 | 7 | 14 | 6 | -0.13468 | -0.053870734 | 0.241586439 | 0.080811876 | 0.376269049 | 0.295457173 | 0.883365566 | 0.177443143 | 0.244954259 |
| sp\|P16871\|IL7RA_HUMAN | 6 | 40 | 76 | 33 | 0.257077 | 0.20904513 | 0.044830723 | -0.048032122 | -0.212246529 | -0.164214407 | 0.751868159 | 0.024521016 | 0.057883562 |
| sp\|Q5JZY3\|EPHAA_HUMAN | 3 | 7 | 14 | 6 | -0.05839 | -0.1263343 | -0.101797404 | -0.067943656 | -0.04340676 | 0.024536897 | 0.849343901 | 0.954671519 | 0.980891373 |
| sp\|P18510\|IL1RA_HUMAN | 3 | 10 | 20 | 10 | -0.96212 | -0.590016426 | 0.410720289 | 0.372105924 | 1.372842639 | 1.000736715 | 0.619171874 | 0.012986194 | 0.041216726 |
| sp\|Q92905\|CSN5_HUMAN | 1 | 15 | 28 | 12 | -0.53973 | -0.22454186 | 0.371997115 | 0.315189697 | 0.911728671 | 0.596538975 | 0.163843758 | 0.000147421 | 0.005718692 |
| sp\|P09326\|CD48_HUMAN | 33 | 40 | 76 | 33 | -0.05144 | 0.038894289 | 0.066907789 | 0.090332191 | 0.11834569 | 0.0280135 | 0.32678769 | 0.266674945 | 0.90894467 |
| sp\|Q15746\|MYLK_HUMAN | 6 | 40 | 76 | 33 | 0.044814 | -0.001335587 | 0.045720089 | -0.046149244 | 0.000906433 | 0.047055677 | 0.861924891 | 0.999960387 | 0.873147656 |
| sp\|P55212\|CASP6_HUMAN | 6 | 21 | 42 | 18 | -0.10829 | -0.184950066 | 0.184156958 | -0.076657957 | 0.292449067 | 0.369107024 | 0.898135644 | 0.344597677 | 0.114564301 |
| sp\|P24666\|PPAC_HUMAN | 9 | 40 | 76 | 33 | -0.14626 | -0.062666592 | 0.176557563 | 0.083597359 | 0.322821514 | 0.239224155 | 0.647129446 | 0.013530163 | 0.047594614 |
| sp\|Q14651\|PLSI_HUMAN | 4 | 40 | 76 | 33 | -0.19499 | 0.084921938 | 0.138101606 | 0.279916301 | 0.33309597 | 0.053179668 | 0.018127029 | 0.019805381 | 0.876078516 |
| sp\|Q08334\|I10R2_HUMAN | 29 | 3 | 7 | 4 | -0.11845 | 0.003958392 | 0.048734221 | 0.122407915 | 0.167183744 | 0.044775829 | 0.803703591 | 0.719465567 | 0.964576858 |
| sp\|Q09328\|MGT5A_HUMAN | 6 | 40 | 76 | 33 | 0.090994 | 0.056937572 | -0.026622114 | -0.034056071 | -0.117615758 | -0.083559686 | 0.6661006 | 0.038914357 | 0.121300359 |
| sp\|Q9H756\|LRC19_HUMAN | 1 | 37 | 69 | 30 | 0.000597 | 0.131432575 | -0.412282379 | 0.130835499 | -0.412879455 | -0.543714954 | 0.5592967 | 0.021490572 | 0.000320224 |
| sp\|Q12866\|MERTK_HUMAN | 42 | 40 | 76 | 33 | 0.017169 | 0.149230479 | 0.226365432 | 0.132061642 | 0.209196595 | 0.077134953 | 0.027382018 | 0.002291368 | 0.330974591 |
| sp\|Q05639\|EF1A2_HUMAN | 8 | 40 | 76 | 33 | -0.47845 | -0.330251769 | 0.255957199 | 0.148193366 | 0.734402333 | 0.586208967 | 0.34744325 | 1.64E-07 | 2.35E-06 |
| sp\|Q9BYG0\|B3GN5_HUMAN | 4 | 19 | 34 | 14 | -0.32097 | -0.241410722 | 0.124327445 | 0.079560468 | 0.445298634 | 0.365738167 | 0.847386389 | 0.039413002 | 0.066499528 |
| sp\|P10746\|HEM4_HUMAN | 6 | 4 | 7 | 2 | -0.46576 | -0.717232829 | -0.624152141 | -0.251471653 | -0.158390965 | 0.093080688 | 0.694739523 | 0.924966653 | 0.968914475 |
| sp\|Q96AT9\|RPE_HUMAN | 5 | 7 | 14 | 7 | -0.5335 | -0.289584358 | -0.679924273 | 0.243917698 | -0.146422216 | -0.390339915 | 0.761115365 | 0.928278378 | 0.503378954 |
| sp\|P30046\|DOPD_HUMAN | 21 | 40 | 76 | 33 | -0.51555 | -0.296782808 | 0.572224768 | 0.218763501 | 1.087771077 | 0.869007576 | 0.311061972 | 3.50E-08 | 6.23E-07 |
| sp\|Q01581\|HMCS1_HUMAN | 3 | 25 | 48 | 21 | -0.96699 | -0.655247835 | -0.428349135 | 0.311739371 | 0.538638071 | 0.226898699 | 0.290410799 | 0.080812427 | 0.555596201 |
| sp\|Q9ULB1\|NRX1A_HUMAN | 7 | 40 | 76 | 33 | 0.111935 | 0.003970341 | -0.225000329 | -0.107964469 | -0.336935138 | -0.22897067 | 0.04851259 | 1.91E-08 | 1.56E-05 |
| sp\|Q04637\|IF4G1_HUMAN | 14 | 8 | 13 | 6 | -0.41136 | -0.401110431 | 0.487204315 | 0.010253551 | 0.898568297 | 0.888314746 | 0.999385047 | 0.056884694 | 0.037267532 |
| sp\|P46108\|CRK_HUMAN | 32 | 25 | 48 | 21 | 0.18915 | 0.042766189 | 0.42055676 | -0.146383907 | 0.231406664 | 0.377790571 | 0.465338175 | 0.26805647 | 0.013532232 |
| sp\|P49862\|KLK7_HUMAN | 33 | 25 | 49 | 22 | 0.33991 | 0.249337277 | 0.171364301 | -0.090572265 | -0.168545842 | -0.077972977 | 0.509296055 | 0.196114182 | 0.631233383 |
| sp\|Q6UXI7\|VITRN_HUMAN | 2 | 10 | 21 | 9 | -0.18451 | -0.121479602 | 0.186608944 | 0.063026491 | 0.371115036 | 0.308088546 | 0.886362984 | 0.067183865 | 0.082814388 |
| sp\|Q94779\|CNTN5_HUMAN | 11 | 15 | 28 | 12 | 0.256265 | 0.060389235 | 0.06963881 | -0.195875422 | -0.186625846 | 0.009249575 | 0.092761029 | 0.222453925 | 0.995195883 |
| sp\|P05387\|RLA2_HUMAN | 9 | 40 | 76 | 33 | -0.51176 | -0.300078273 | 0.342532646 | 0.211684079 | 0.854294998 | 0.642610919 | 0.108589236 | 7.23E-10 | 1.33E-07 |
| sp\|P41271\|NBL1_HUMAN | 62 | 40 | 76 | 33 | -0.25321 | -0.293323665 | 0.315679085 | -0.040114337 | 0.568888413 | 0.60900275 | 0.91582354 | 1.72E-05 | 2.07E-07 |
| sp\|P53634\|CATC_HUMAN | 10 | 40 | 76 | 33 | -0.04166 | -0.025680037 | 0.203927473 | 0.015976834 | 0.245584343 | 0.229607509 | 0.96310324 | 0.003089472 | 0.00167663 |
| sp\|Q6UWH4\|GAK1B_HUMAN | 6 | 18 | 35 | 16 | 0.12446 | 0.062335141 | 0.247762612 | -0.062124986 | 0.123302484 | 0.18542747 | 0.884102062 | 0.70869242 | 0.368973144 |
| sp\|P13598\|ICAM2_HUMAN | 1 | 40 | 76 | 33 | 0.207718 | 0.2459264 | 0.154515813 | 0.038208381 | -0.053202206 | -0.091410588 | 0.691879312 | 0.611338153 | 0.161561647 |
| sp\|Q03113\|GNA12_HUMAN | 24 | 14 | 25 | 12 | -0.229 | -0.095763666 | 0.684597481 | 0.133238579 | 0.913599726 | 0.780361146 | 0.833597017 | 0.004422141 | 0.006655257 |
| sp\|P54725\|RD23A_HUMAN | 1 | 40 | 76 | 33 | -0.18612 | -0.176086007 | 0.212578844 | 0.010029121 | 0.398693972 | 0.388664851 | 0.990443097 | 7.27E-05 | 1.19E-05 |
| sp\|Q9BSL1\|UBAC1_HUMAN | 2 | 7 | 14 | 5 | -0.15775 | -0.084919162 | 0.405523809 | 0.072832021 | 0.563274993 | 0.490442971 | 0.929547502 | 0.087255328 | 0.095750692 |
| sp\|Q6UY14\|ATL4_HUMAN | 13 | 40 | 76 | 33 | 0.227612 | 0.061229267 | -0.089861343 | -0.16638232 | -0.31747293 | -0.15109061 | 0.00203212 | 5.31E-07 | 0.010458177 |
| sp\|P0C862\|C1T9A_HUMAN | 17 | 33 | 62 | 27 | 0.060508 | -0.011901315 | -0.184935671 | -0.07240887 | -0.245443227 | -0.173034356 | 0.819461872 | 0.211880292 | 0.373980241 |
| sp\|Q6UXD5\|SE6L2_HUMAN | 2 | 40 | 76 | 33 | 0.13201 | 0.117393929 | 0.251277306 | -0.014616152 | 0.119267225 | 0.133883377 | 0.973033569 | 0.289432562 | 0.139057785 |
| sp\|Q96IU4\|ABHEB_HUMAN | 89 | 40 | 76 | 33 | -0.13228 | 0.01823028 | 0.285662952 | 0.1505097 | 0.417942372 | 0.267432672 | 0.152322529 | 9.04E-05 | 0.006399181 |
| sp\|Q9NY65\|TBA8_HUMAN | 2 | 21 | 42 | 19 | -0.61458 | -0.208256822 | 0.270850234 | 0.406322912 | 0.885429969 | 0.479107056 | 0.275884972 | 0.015604093 | 0.189600923 |
| sp\|Q3ZCW2\|LEGL_HUMAN | 6 | 40 | 76 | 33 | -0.07968 | -0.181339757 | -0.100329896 | -0.10166296 | -0.020653099 | 0.081009861 | 0.688769595 | 0.989398099 | 0.812024653 |
| sp\|Q15819\|UB2V2_HUMAN | 15 | 4 | 7 | 3 | -0.06725 | -0.011514422 | 0.147403399 | 0.055733884 | 0.214651705 | 0.158917821 | 0.841941379 | 0.22100241 | 0.346713051 |
| sp\|Q13404\|UB2V1_HUMAN | 3 | 40 | 76 | 33 | -0.33855 | -0.133766486 | 0.212548313 | 0.20478727 | 0.551102069 | 0.346314799 | 0.12299045 | 5.98E-05 | 0.006069067 |
| sp\|Q13094\|LCP2_HUMAN | 11 | 37 | 69 | 29 | -0.08597 | -0.215903134 | 0.107864772 | -0.129931394 | 0.193836512 | 0.323767906 | 0.558182473 | 0.417669833 | 0.050383173 |
| sp\|P50591\|TNF10_HUMAN | 2 | 18 | 35 | 16 | 0.114234 | 0.179352545 | 0.121391404 | 0.065118234 | 0.007157094 | -0.05796114 | 0.439723062 | 0.992842638 | 0.546738453 |
| sp\|P13805\|TNNT1_HUMAN | 2 | 18 | 34 | 16 | -1.89163 | -2.07535678 | -1.122237023 | -0.183724476 | 0.769395281 | 0.953119757 | 0.892505706 | 0.246500645 | 0.067647076 |
| sp\|P61086\|UBE2K_HUMAN | 2 | 32 | 62 | 28 | -0.21892 | -0.120211866 | 0.266283547 | 0.098707604 | 0.485203016 | 0.386495412 | 0.607193741 | 0.000395329 | 0.001478961 |
| sp\|Q06828\|FMOD_HUMAN | 4 | 40 | 76 | 33 | 0.073155 | 0.04354437 | -0.104260785 | -0.029610533 | -0.177415688 | -0.147805155 | 0.845601322 | 0.018450151 | 0.02894859 |
| sp\|Q9BX67\|JAM3_HUMAN | 10 | 4 | 6 | 3 | 0.035853 | -0.015052604 | 0.533918496 | -0.05090529 | 0.49806581 | 0.5489711 | 0.960067504 | 0.145445553 | 0.077980191 |
| sp\|Q8TBC4\|UBA3_HUMAN | 10 | 12 | 21 | 9 | -0.28815 | -0.149385782 | 0.106337559 | 0.138767553 | 0.394490894 | 0.25572334 | 0.851363204 | 0.424036135 | 0.639524151 |

| | | | | | | | | | | | | | |
|---|---|---|---|---|---|---|---|---|---|---|---|---|---|
| sp\|P61019\|RAB2A_HUMAN | 1 | 33 | 62 | 27 | 0.019608 | 0.072924186 | 0.384679108 | 0.053315753 | 0.365070676 | 0.311754922 | 0.793115745 | 0.000977531 | 0.001601728 |
| sp\|P60022\|DEFB1_HUMAN | 1 | 36 | 69 | 30 | -0.17874 | -0.128414335 | 0.147492153 | 0.050323019 | 0.326229507 | 0.275906488 | 0.837069037 | 0.00739005 | 0.011057369 |
| sp\|Q9H082\|RB33B_HUMAN | 3 | 6 | 14 | 7 | -1.11648 | -0.85459409 | -0.770721971 | 0.261884352 | 0.345756471 | 0.083872119 | 0.841236267 | 0.793588627 | 0.980354202 |
| sp\|O00584\|RNT2_HUMAN | 13 | 40 | 76 | 33 | 0.206238 | 0.112552655 | 0.30685174 | -0.093685184 | 0.100613902 | 0.194299086 | 0.317776542 | 0.400693844 | 0.015054919 |
| sp\|Q9HAB8\|PPCS_HUMAN | 2 | 17 | 35 | 15 | 0.115327 | 0.132406285 | 0.332605124 | 0.017078795 | 0.217277634 | 0.200198838 | 0.988313232 | 0.274597125 | 0.236541979 |
| sp\|P12956\|XRCC6_HUMAN | 6 | 21 | 42 | 18 | -0.03628 | -0.042465921 | 0.506041172 | -0.006188219 | 0.542318874 | 0.548507093 | 0.99922254 | 0.020638745 | 0.006320244 |
| sp\|A8MVW5\|HECA2_HUMAN | 4 | 30 | 55 | 23 | 0.180464 | 0.055322393 | -0.073926125 | -0.125141492 | -0.254390011 | -0.129248518 | 0.120263061 | 0.003644132 | 0.150589502 |
| sp\|Q96C24\|SYTL4_HUMAN | 11 | 4 | 7 | 3 | 0.26423 | -0.0239036 | -0.30699022 | -0.288133534 | -0.571220154 | -0.28308662 | 0.584855454 | 0.268151418 | 0.649539013 |
| sp\|Q7Z5L7\|PODN_HUMAN | 24 | 36 | 69 | 30 | -0.04608 | -0.022314751 | 0.081605094 | 0.023767282 | 0.127687127 | 0.103919845 | 0.883608319 | 0.090014082 | 0.129245312 |
| sp\|Q02750\|MP2K1_HUMAN | 3 | 4 | 7 | 3 | -0.05741 | -0.080553087 | 0.107022439 | -0.023142678 | 0.164432849 | 0.187575526 | 0.990718411 | 0.734840788 | 0.616958977 |
| sp\|Q9HB71\|CYBP_HUMAN | 5 | 17 | 35 | 15 | -0.07436 | -0.188984275 | 0.396690923 | -0.114622849 | 0.471052349 | 0.585675198 | 0.803353196 | 0.084934454 | 0.008188888 |
| sp\|O15123\|ANGP2_HUMAN | 36 | 11 | 21 | 9 | -0.08855 | -0.107174049 | 0.239798187 | -0.018620648 | 0.328351588 | 0.346972236 | 0.989900671 | 0.130576893 | 0.059291642 |
| sp\|Q14116\|IL18_HUMAN | 1 | 24 | 48 | 21 | -0.19019 | -0.084680349 | 0.319089748 | 0.105506844 | 0.509276941 | 0.403770097 | 0.649104403 | 0.001561858 | 0.004612852 |
| sp\|P40197\|GPV_HUMAN | 88 | 40 | 76 | 33 | -0.12092 | 0.182341081 | 0.099982444 | 0.303260058 | 0.220901421 | -0.082358637 | 1.52E-05 | 0.013190008 | 0.452081994 |
| sp\|Q8TEM1\|PO210_HUMAN | 18 | 16 | 27 | 11 | 0.125305 | 0.122133614 | 0.102611276 | -0.00317106 | -0.022693398 | -0.019522338 | 0.999350846 | 0.978663479 | 0.981043305 |
| sp\|P07478\|TRY2_HUMAN | 14 | 40 | 76 | 33 | 0.033772 | 0.079990457 | 0.485737038 | 0.046218556 | 0.451965137 | 0.405746581 | 0.92371425 | 0.006884496 | 0.006099624 |
| sp\|O15400\|STX7_HUMAN | 32 | 22 | 41 | 18 | -0.11516 | -0.07652956 | 0.689638574 | 0.038626967 | 0.8047951 | 0.766168133 | 0.954468716 | 8.78E-06 | 2.15E-06 |
| sp\|Q6UWL2\|SUSD1_HUMAN | 11 | 25 | 49 | 21 | -0.05788 | 0.005975734 | -0.095981036 | 0.063857806 | -0.038098964 | -0.10195677 | 0.749785798 | 0.931557502 | 0.522868651 |
| sp\|P53990\|IST1_HUMAN | 2 | 18 | 35 | 16 | -0.42209 | -0.424178305 | 0.056443268 | -0.002091336 | 0.478530236 | 0.480621572 | 0.999957781 | 0.216499942 | 0.13752602 |
| sp\|P48507\|GSH0_HUMAN | 23 | 40 | 76 | 33 | -0.32794 | -0.071681768 | 0.277009143 | 0.256254467 | 0.604945378 | 0.348690911 | 0.006162451 | 2.51E-08 | 0.000321512 |
| sp\|O95336\|6PGL_HUMAN | 14 | 40 | 76 | 33 | -0.00835 | 0.036475215 | 0.587248793 | 0.044823441 | 0.595597019 | 0.550773578 | 0.892285662 | 4.40E-06 | 1.66E-06 |
| sp\|O94898\|LRIG2_HUMAN | 6 | 33 | 62 | 27 | 0.246463 | 0.081682514 | 0.085861561 | -0.164780326 | -0.160601279 | 0.004179047 | 0.095230856 | 0.211212474 | 0.998641984 |
| sp\|P63241\|IF5A1_HUMAN | 1 | 40 | 76 | 33 | -0.28743 | -0.125906846 | 0.285397961 | 0.16152809 | 0.572832897 | 0.411304807 | 0.352227627 | 0.000221006 | 0.003454 |
| sp\|P54136\|SYRC_HUMAN | 1 | 18 | 35 | 15 | -0.08075 | -0.177935288 | 0.245620155 | -0.097187235 | 0.326368208 | 0.423555443 | 0.711332335 | 0.079221968 | 0.00544486 |
| sp\|P06730\|IF4E_HUMAN | 37 | 40 | 76 | 33 | -0.11078 | -0.074417263 | 0.285913791 | 0.036363024 | 0.396694078 | 0.360331054 | 0.931208649 | 0.003948767 | 0.003040647 |
| sp\|P08514\|ITA2B_HUMAN | 2 | 36 | 70 | 30 | -0.38825 | -0.086720263 | 0.118107098 | 0.301527378 | 0.506354739 | 0.204827362 | 0.012781966 | 0.000291881 | 0.160961442 |
| sp\|Q99685\|MGLL_HUMAN | 5 | 33 | 62 | 27 | 0.050806 | 0.080304309 | 0.069195754 | 0.029497823 | 0.018389268 | -0.011108555 | 0.961694655 | 0.98958432 | 0.995172036 |
| sp\|P61601\|NCALD_HUMAN | 21 | 20 | 41 | 19 | -0.1067 | -0.042800498 | 0.240854205 | 0.063901344 | 0.347556048 | 0.283654703 | 0.882542461 | 0.076379061 | 0.10092793 |
| sp\|Q9Y2X8\|UB2D4_HUMAN | 23 | 11 | 20 | 9 | 0.068946 | 0.073220981 | 0.057426951 | 0.004275301 | -0.011518729 | -0.01579403 | 0.998814929 | 0.99401514 | 0.985951676 |
| sp\|P61106\|RAB14_HUMAN | 3 | 36 | 69 | 31 | -0.13784 | -0.154510354 | 0.272844759 | -0.016675226 | 0.410679887 | 0.427355113 | 0.98409974 | 0.001681048 | 0.000169948 |
| sp\|Q9UI42\|CBPA4_HUMAN | 10 | 10 | 21 | 10 | 0.003572 | 0.055684171 | -0.193024346 | 0.05211221 | -0.196596307 | -0.248708516 | 0.813406793 | 0.128500984 | 0.015389262 |
| sp\|P80188\|NGAL_HUMAN | 10 | 40 | 76 | 33 | -0.20375 | -0.040148718 | 0.490077091 | 0.163603093 | 0.693828901 | 0.530225808 | 0.287002959 | 1.06E-06 | 2.70E-05 |
| sp\|O75822\|EIF3J_HUMAN | 44 | 4 | 6 | 3 | -0.31883 | -0.166680457 | 0.36939182 | 0.152151882 | 0.68822416 | 0.536072277 | 0.965156708 | 0.610246812 | 0.701336858 |
| sp\|Q8N114\|SHSA5_HUMAN | 1 | 7 | 14 | 6 | -0.65669 | -0.515026662 | 0.568106758 | 0.141665852 | 1.224799272 | 1.08313342 | 0.933617823 | 0.045374665 | 0.043378191 |
| sp\|P63302\|SELW_HUMAN | 64 | 4 | 6 | 3 | 0.253349 | 0.095221251 | 1.59307659 | -0.158127991 | 1.339727347 | 1.497855338 | 0.958594133 | 0.165683034 | 0.086948616 |
| sp\|P09619\|PGFRB_HUMAN | 36 | 40 | 76 | 33 | 0.093555 | 0.009195013 | -0.061536077 | -0.084359611 | -0.155090701 | -0.070731089 | 0.390207173 | 0.114556571 | 0.558189531 |
| sp\|P31431\|SDC4_HUMAN | 6 | 40 | 76 | 33 | -0.34206 | 0.143470201 | 0.051915991 | 0.485531409 | 0.393977199 | -0.09155421 | 2.77E-10 | 1.70E-05 | 0.434829348 |
| sp\|P02792\|FRIL_HUMAN | 1 | 40 | 76 | 33 | -0.47744 | -0.187989291 | 0.759909989 | 0.289454376 | 1.237353656 | 0.94789928 | 0.183751482 | 1.10E-08 | 6.95E-07 |
| sp\|P30711\|GSTT1_HUMAN | 103 | 4 | 7 | 3 | 0.719449 | 0.454278197 | -0.091642988 | -0.265170503 | -0.811091687 | -0.545921184 | 0.448818232 | 0.023353916 | 0.091212975 |
| sp\|O43451\|MGA_HUMAN | 9 | 34 | 62 | 26 | -0.16281 | -0.115046812 | 0.136135037 | 0.047764375 | 0.298946224 | 0.251181849 | 0.902370989 | 0.072846971 | 0.09949115 |
| sp\|P14649\|MYL6B_HUMAN | 1 | 40 | 76 | 33 | -0.49445 | -0.256057406 | 0.404548643 | 0.23809548 | 0.898701529 | 0.660606049 | 0.150633012 | 8.51E-08 | 8.70E-06 |
| sp\|O00442\|RTCA_HUMAN | 2 | 11 | 21 | 9 | -0.10933 | 0.058270925 | 0.210837306 | 0.167599304 | 0.320165685 | 0.152566382 | 0.648940117 | 0.345885375 | 0.730633526 |
| sp\|Q92530\|PSMF1_HUMAN | 4 | 22 | 41 | 17 | -0.19804 | -0.005779436 | 0.375591419 | 0.192264431 | 0.573635287 | 0.381370856 | 0.251202629 | 0.000572302 | 0.012960691 |
| sp\|Q9H159\|CAD19_HUMAN | 6 | 40 | 76 | 33 | 0.077794 | 0.027653648 | 0.054696367 | -0.050139988 | -0.023097269 | 0.027042718 | 0.42923181 | 0.882684893 | 0.804564424 |
| sp\|Q16762\|THTR_HUMAN | 8 | 8 | 13 | 6 | -0.25624 | -0.003620026 | 0.570440111 | 0.252617371 | 0.826677508 | 0.574060137 | 0.520541546 | 0.01618121 | 0.077300325 |
| sp\|Q13185\|CBX3_HUMAN | 5 | 7 | 14 | 7 | -0.39835 | -0.417419065 | 0.408159322 | -0.019073955 | 0.806504431 | 0.825578387 | 0.998083915 | 0.098173097 | 0.043781748 |
| sp\|P35247\|SFTPD_HUMAN | 6 | 40 | 76 | 33 | -0.31402 | -0.264474921 | 0.329871435 | 0.049542636 | 0.643888992 | 0.594346356 | 0.9060551 | 3.07E-05 | 1.39E-05 |
| sp\|P11166\|GTR1_HUMAN | 7 | 15 | 28 | 11 | -0.85686 | -0.705924257 | -0.153694161 | 0.150933403 | 0.703163499 | 0.552230097 | 0.649575009 | 0.004425338 | 0.013962843 |
| sp\|Q01546\|K22O_HUMAN | 2 | 36 | 69 | 30 | 0.016083 | -0.462559727 | -0.651165557 | -0.478642751 | -0.667248582 | -0.188605831 | 0.122260625 | 0.060613029 | 0.745026802 |
| sp\|Q96FJ2\|DYL2_HUMAN | 8 | 37 | 69 | 30 | -0.25059 | -0.10826318 | 0.220801161 | 0.142331274 | 0.471395614 | 0.329064341 | 0.409053013 | 0.001729989 | 0.018155834 |
| sp\|Q9UBY9\|HSPB7_HUMAN | 8 | 4 | 6 | 3 | 0.033524 | 0.011014477 | 1.356688638 | -0.022509805 | 1.323164357 | 1.345674161 | 0.999398575 | 0.27407148 | 0.217900277 |
| sp\|O75608\|LYPA1_HUMAN | 3 | 40 | 76 | 33 | -0.11701 | -0.264543906 | -0.006371891 | -0.147534211 | 0.110637804 | 0.258172015 | 0.31195158 | 0.634181303 | 0.046388505 |
| sp\|P07741\|APT_HUMAN | 26 | 40 | 76 | 33 | -0.19666 | -0.197675619 | 0.147635992 | -0.001020255 | 0.344291356 | 0.345311611 | 0.999950975 | 0.024566257 | 0.009117289 |
| sp\|P04792\|HSPB1_HUMAN | 8 | 40 | 76 | 33 | -0.08222 | -0.237206141 | 0.317489116 | -0.154988113 | 0.399707144 | 0.554695258 | 0.482014462 | 0.038306758 | 0.00047214 |
| sp\|P26842\|CD27_HUMAN | 4 | 32 | 62 | 27 | -0.63438 | -0.633441341 | -0.159866804 | 0.000941815 | 0.474516351 | 0.473574537 | 0.9999736 | 0.012147186 | 0.003817384 |
| sp\|Q96P44\|COLA1_HUMAN | 4 | 14 | 28 | 12 | 0.08612 | 0.010009479 | -0.070485659 | -0.076110663 | -0.1566058 | -0.080495137 | 0.593646261 | 0.224464284 | 0.591607502 |
| sp\|P13942\|COBA2_HUMAN | 2 | 40 | 76 | 33 | 0.224592 | 0.009564559 | -0.602821423 | -0.215027343 | -0.827413326 | -0.612385983 | 0.107757159 | 3.55E-09 | 6.94E-07 |
| sp\|P68032\|ACTC_HUMAN | 4 | 40 | 76 | 33 | -0.14879 | -0.34040623 | -0.009222043 | -0.191620067 | 0.13956412 | 0.331184187 | 0.234988838 | 0.585547743 | 0.02445493 |
| sp\|A5A3E0\|POTEF_HUMAN | 10 | 24 | 48 | 22 | 0.144908 | -0.160965965 | 0.248516256 | -0.305874238 | 0.103607983 | 0.409482221 | 0.191182716 | 0.870000672 | 0.063821471 |
| sp\|P0CG38\|POTEI_HUMAN | 12 | 4 | 6 | 2 | 0.114757 | -0.349970452 | 0.573646559 | -0.464727365 | 0.458889646 | 0.923611011 | 0.322887148 | 0.52291406 | 0.091246671 |
| sp\|P0CG39\|POTEJ_HUMAN | 12 | 25 | 48 | 21 | -0.05971 | -0.261220455 | 0.112579538 | -0.2015146 | 0.172285393 | 0.373799993 | 0.405046477 | 0.630241586 | 0.067714715 |
| sp\|P48052\|CBPA2_HUMAN | 2 | 31 | 60 | 27 | 0.139744 | 0.00305196 | 0.157786068 | -0.136692493 | 0.018041615 | 0.154734108 | 0.27936954 | 0.984193275 | 0.22654723 |
| sp\|Q9Y4Y9\|LSM5_HUMAN | 38 | 4 | 6 | 3 | 0.485221 | 0.536160582 | 0.362727377 | 0.050939651 | -0.122493555 | -0.173433206 | 0.958573974 | 0.841657462 | 0.674440598 |
| sp\|Q96GG9\|DCNL1_HUMAN | 2 | 18 | 35 | 15 | -0.1495 | 0.019660009 | 0.252091315 | 0.169164237 | 0.401595543 | 0.232431306 | 0.300631907 | 0.012395327 | 0.138766618 |
| sp\|Q86TP1\|PRUN1_HUMAN | 1 | 22 | 42 | 18 | -0.04702 | 0.116560312 | 0.18708262 | 0.163579798 | 0.234102106 | 0.070522308 | 0.42701117 | 0.304683811 | 0.869654527 |
| sp\|Q8WVM8\|SCFD1_HUMAN | 3 | 28 | 55 | 24 | -0.27868 | -0.21037199 | 0.192852579 | 0.06830753 | 0.4715321 | 0.403224569 | 0.913560091 | 0.055504513 | 0.064630172 |
| sp\|Q9Y625\|GPC6_HUMAN | 27 | 17 | 34 | 16 | -0.15594 | -0.241972878 | 0.111020535 | -0.086029943 | 0.266963471 | 0.352993414 | 0.825535934 | 0.268818911 | 0.052933797 |
| sp\|Q9UPY8\|MARE3_HUMAN | 3 | 36 | 69 | 30 | -0.21845 | -0.461186476 | -0.161375999 | -0.242731992 | 0.057078486 | 0.299810477 | 0.365513606 | 0.961826963 | 0.258770313 |
| sp\|Q13444\|ADA15_HUMAN | 1 | 18 | 34 | 16 | -0.14122 | -0.038018722 | 0.447201885 | 0.103199138 | 0.588419745 | 0.485220607 | 0.698049337 | 0.000625632 | 0.001441639 |
| sp\|Q9Y2Z0\|SGT1_HUMAN | 5 | 7 | 14 | 6 | 0.139109 | 0.051207262 | 0.434030583 | -0.087901376 | 0.294921945 | 0.382823321 | 0.906035478 | 0.474017462 | 0.207179601 |
| sp\|P51888\|PRELP_HUMAN | 1 | 29 | 55 | 24 | 0.132474 | 0.042295349 | 0.428497856 | -0.090178911 | 0.296023596 | 0.386202507 | 0.724776922 | 0.096715032 | 0.007474531 |
| sp\|O00423\|EMAL1_HUMAN | 8 | 4 | 7 | 3 | 0.110214 | 0.185484964 | -0.263450385 | 0.075270944 | -0.373664405 | -0.448935349 | 0.964915045 | 0.569220985 | 0.383294902 |
| sp\|Q96E39\|RMXL1_HUMAN | 12 | 11 | 20 | 9 | -0.2201 | -0.11654231 | 0.816882137 | 0.103556616 | 1.036981063 | 0.933424447 | 0.952337257 | 0.044710273 | 0.042731451 |
| sp\|Q7Z3B1\|NEGR1_HUMAN | 37 | 40 | 76 | 33 | 0.195911 | 0.12168238 | 0.028324213 | -0.074228245 | -0.167586412 | -0.093358167 | 0.098730159 | 0.000443353 | 0.041291924 |
| sp\|Q9UDX3\|S14L4_HUMAN | 5 | 14 | 27 | 13 | -0.16775 | 0.171987146 | 0.452144014 | 0.339739093 | 0.619895961 | 0.280156869 | 0.099378268 | 0.005238371 | 0.218289207 |
| sp\|O76054\|S14L2_HUMAN | 1 | 19 | 34 | 15 | -0.3111 | -0.020253563 | 0.392581715 | 0.290842348 | 0.703677626 | 0.412835278 | 0.153452554 | 0.001033185 | 0.04292983 |
| sp\|Q9ULZ3\|ASC_HUMAN | 7 | 33 | 62 | 27 | -0.58693 | -0.482659836 | 0.358573003 | 0.104266921 | 0.94549976 | 0.841232839 | 0.754077902 | 1.05E-06 | 1.01E-06 |
| sp\|Q53FA7\|QORX_HUMAN | 8 | 21 | 42 | 18 | -0.23207 | -0.061325511 | 0.385618392 | 0.170745384 | 0.617689287 | 0.446943903 | 0.277106839 | 4.06E-05 | 0.000747473 |
| sp\|Q96DA0\|ZG16B_HUMAN | 5 | 37 | 69 | 30 | 0.050131 | 0.058119855 | 0.059856736 | 0.007989004 | 0.009725886 | 0.001736881 | 0.993723108 | 0.993601271 | 0.999741637 |
| sp\|Q9HA64\|KT3K_HUMAN | 4 | 40 | 76 | 33 | -0.3587 | -0.161640986 | 0.226422797 | 0.197063718 | 0.5851275 | 0.388063783 | 0.130261246 | 1.19E-05 | 0.001321087 |
| sp\|P06734\|FCER2_HUMAN | 5 | 11 | 20 | 9 | -0.15119 | -0.017998588 | 0.336371115 | 0.133195981 | 0.487565685 | 0.354369703 | 0.735549475 | 0.069357518 | 0.162879846 |
| sp\|P07205\|PGK2_HUMAN | 4 | 40 | 76 | 33 | -0.13936 | -0.046314046 | 0.232705642 | 0.093043313 | 0.372063001 | 0.279019688 | 0.547149904 | 0.001870096 | 0.010346122 |
| sp\|P18085\|ARF4_HUMAN | 15 | 36 | 69 | 30 | 0.041601 | -0.073411203 | 0.382705663 | -0.115012289 | 0.341104577 | 0.456116866 | 0.616032312 | 0.056695508 | 0.001798377 |
| sp\|Q9Y3I1\|FBX7_HUMAN | 3 | 32 | 62 | 28 | -0.181 | 0.082081147 | 0.415897957 | 0.263085884 | 0.596902655 | 0.333816771 | 0.088991616 | 0.000262076 | 0.029778405 |
| sp\|Q92995\|UBP13_HUMAN | 4 | 4 | 7 | 3 | 0.261128 | 0.134330761 | 0.058687077 | -0.126797128 | -0.202440812 | -0.075643684 | 0.54539984 | 0.367287099 | 0.830812507 |
| sp\|P51659\|DHB4_HUMAN | 4 | 4 | 7 | 2 | 0.232439 | -0.003208462 | 0.259795991 | -0.23564739 | 0.027357063 | 0.263004453 | 0.499575989 | 0.994745603 | 0.58440963 |
| sp\|Q5VUB5\|F1711_HUMAN | 2 | 26 | 48 | 20 | 0.493913 | 0.383716309 | 0.463779559 | -0.110196768 | -0.030133518 | 0.080063249 | 0.815838278 | 0.98982106 | 0.913846547 |
| sp\|P47972\|NPTX2_HUMAN | 19 | 40 | 76 | 33 | 0.056789 | -0.035648349 | 0.077788894 | -0.092437587 | 0.020999656 | 0.113437243 | 0.164429125 | 0.936620016 | 0.093205398 |
| sp\|P05413\|FABPH_HUMAN | 2 | 40 | 76 | 33 | -1.57037 | -1.636926905 | -0.793708097 | -0.066558922 | 0.776659885 | 0.843218807 | 0.920934692 | 0.000745189 | 2.81E-05 |
| sp\|Q86UN2\|R4RL1_HUMAN | 1 | 15 | 28 | 12 | 0.0882 | 0.125987118 | 0.181235514 | 0.037786909 | 0.093035305 | 0.055248396 | 0.859033363 | 0.536930833 | 0.756894248 |
| sp\|O14775\|GNB5_HUMAN | 14 | 8 | 14 | 6 | -0.04999 | -0.062163007 | -0.013057206 | -0.012177779 | 0.036928022 | 0.049105801 | 0.997563825 | 0.98501644 | 0.967851157 |
| sp\|P09486\|SPRC_HUMAN | 1 | 40 | 76 | 33 | -0.17129 | 0.18471035 | 0.168622142 | 0.35599766 | 0.339909452 | -0.016088208 | 0.001653772 | 0.016246151 | 0.98781971 |
| sp\|P08263\|GSTA1_HUMAN | 4 | 21 | 41 | 17 | -0.12246 | 0.177641527 | -0.038494414 | 0.300097727 | 0.083961785 | -0.216135941 | 0.133240584 | 0.895629085 | 0.397902783 |
| sp\|P41236\|IPP2_HUMAN | 27 | 7 | 13 | 6 | -0.03042 | 0.018712949 | 0.779929881 | 0.049131345 | 0.810348277 | 0.761216932 | 0.989235816 | 0.148193419 | 0.120115497 |
| sp\|Q9BYZ2\|LDH6B_HUMAN | 1 | 30 | 55 | 23 | -0.20423 | 0.032129896 | 0.280122304 | 0.236358212 | 0.484350619 | 0.247992408 | 0.146646949 | 0.00559662 | 0.17047974 |
| sp\|P62873\|GBB1_HUMAN | 5 | 40 | 76 | 33 | 0.023729 | 8.69E-05 | 0.16445001 | -0.023642217 | 0.140720889 | 0.164363106 | 0.906267422 | 0.095414307 | 0.018125568 |
| sp\|P17980\|PRS6A_HUMAN | 1 | 31 | 55 | 23 | -0.52097 | -0.324776314 | 0.057656496 | 0.196197348 | 0.578630159 | 0.38243281 | 0.277399315 | 0.00098905 | 0.021158922 |
| sp\|Q9H7Z3\|NRDE2_HUMAN | 3 | 7 | 14 | 6 | 0.060592 | -0.033236783 | 0.574172316 | -0.093829107 | 0.513579992 | 0.607409099 | 0.960937109 | 0.450126564 | 0.243849547 |
| sp\|P20936\|RASA1_HUMAN | 6 | 7 | 14 | 6 | -0.35671 | -0.18390223 | 0.669040828 | 0.172810135 | 1.025753193 | 0.852943058 | 0.848186436 | 0.031680754 | 0.042976986 |
| sp\|P28074\|PSB5_HUMAN | 12 | 40 | 76 | 33 | -0.13863 | -0.026236759 | 0.432775993 | 0.112390114 | 0.571402865 | 0.459012752 | 0.459338379 | 4.09E-06 | 3.12E-05 |
| sp\|O95834\|EMAL2_HUMAN | 1 | 40 | 76 | 33 | -0.17635 | -0.244777127 | 0.012451023 | -0.068423636 | 0.188804514 | 0.25722815 | 0.842507554 | 0.409256585 | 0.124429991 |
| sp\|P14174\|MIF_HUMAN | 11 | 40 | 76 | 33 | -0.3683 | -0.164121271 | 0.248835633 | 0.204176795 | 0.617133699 | 0.412956904 | 0.174437806 | 3.93E-05 | 0.002457087 |
| sp\|A6NI72\|NCF1B_HUMAN | 2 | 25 | 47 | 20 | -0.96373 | -0.689474092 | 0.176348076 | 0.274258575 | 1.140080742 | 0.865822168 | 0.495670405 | 0.000560187 | 0.003701556 |
| sp\|Q8TD30\|ALAT2_HUMAN | 14 | 40 | 76 | 33 | 0.062037 | 0.168219612 | 0.091877799 | 0.106182124 | 0.029840311 | -0.076341813 | 0.36702606 | 0.946321658 | 0.632841421 |
| sp\|Q16853\|AOC3_HUMAN | 9 | 40 | 76 | 33 | 0.196174 | 0.134597487 | 0.10864817 | -0.061576557 | -0.087525875 | -0.025949318 | 0.314185082 | 0.200606021 | 0.833266975 |

| | | | | | | | | | | | | |
|---|---|---|---|---|---|---|---|---|---|---|---|---|
| sp\|Q15286\|RAB35_HUMAN | 1 | 18 | 34 | 16 | -0.13916 | -0.048499787 | 0.293964115 | 0.090660944 | 0.433124846 | 0.342463902 | 0.853604491 | 0.083154985 | 0.133314969 |
| sp\|Q86TD4\|SRCA_HUMAN | 31 | 23 | 41 | 17 | 0.090936 | 0.062987447 | -0.007204807 | -0.027948759 | -0.098141013 | -0.070192254 | 0.921975741 | 0.517803234 | 0.659959601 |
| sp\|Q92930\|RAB8B_HUMAN | 2 | 18 | 35 | 14 | -0.40662 | -0.233565653 | 0.394880247 | 0.173054853 | 0.801500754 | 0.6284459 | 0.628163428 | 0.002600044 | 0.008631546 |
| sp\|P29992\|GNA11_HUMAN | 4 | 4 | 7 | 3 | -0.09034 | 0.325645524 | -0.090909245 | 0.415981093 | -0.000573675 | -0.416554769 | 0.38389427 | 0.999998655 | 0.447575741 |
| sp\|Q9BRR6\|ADPGK_HUMAN | 39 | 4 | 7 | 3 | -0.07251 | 0.078658462 | -0.077428256 | 0.151167272 | -0.004919446 | -0.156086718 | 0.575672048 | 0.999584814 | 0.613425926 |
| sp\|Q9GZN8\|CT027_HUMAN | 19 | 30 | 55 | 24 | -0.38382 | -0.249851045 | 0.11016999 | 0.133968505 | 0.493989539 | 0.360021034 | 0.595018695 | 0.010047835 | 0.044159714 |
| sp\|Q9UQC9\|CLCA2_HUMAN | 1 | 22 | 42 | 17 | 0.342175 | 0.107727461 | 0.016925687 | -0.234447161 | -0.325248935 | -0.090801773 | 0.010556393 | 0.003306762 | 0.543274864 |
| sp\|Q9UKU9\|ANGL2_HUMAN | 1 | 40 | 76 | 33 | -0.021 | 0.00148832 | 0.265042294 | 0.022491907 | 0.286045881 | 0.263553974 | 0.894538756 | 1.42E-05 | 6.31E-06 |
| sp\|O14561\|ACPM_HUMAN | 9 | 4 | 6 | 3 | 0.053716 | 0.094742797 | 1.231969003 | 0.041027229 | 1.178253435 | 1.137226206 | 0.998172428 | 0.378117747 | 0.350479246 |
| sp\|Q8NBZ7\|UXS1_HUMAN | 11 | 4 | 6 | 3 | -0.01299 | -0.082723904 | -0.50026263 | -0.069730726 | -0.487269452 | -0.417538726 | 0.807481502 | 0.010114496 | 0.015837482 |
| sp\|Q01344\|IL5RA_HUMAN | 8 | 4 | 7 | 3 | -0.37256 | -0.138157526 | -0.491612411 | 0.234399838 | -0.119055047 | -0.353454884 | 0.372767843 | 0.830910884 | 0.178634431 |
| sp\|Q15459\|SF3A1_HUMAN | 70 | 4 | 6 | 3 | -0.03816 | 0.274541136 | 0.753525381 | 0.312705929 | 0.791690174 | 0.478984245 | 0.859842288 | 0.519170796 | 0.747563458 |
| sp\|O15389\|SIGL5_HUMAN | 4 | 40 | 76 | 33 | -0.59687 | -0.218961074 | -0.340318677 | 0.377905018 | 0.256547415 | -0.121357603 | 0.198738375 | 0.594662342 | 0.861927042 |
| sp\|P07738\|PMGE_HUMAN | 6 | 40 | 76 | 33 | -0.34092 | -0.17039902 | 0.377781465 | 0.170523135 | 0.71870362 | 0.548180485 | 0.273930762 | 7.81E-07 | 2.21E-05 |
| sp\|P37173\|TGFR2_HUMAN | 11 | 40 | 76 | 33 | -0.08023 | -0.073690148 | 0.216442209 | 0.006542242 | 0.2966746 | 0.290132357 | 0.994899865 | 0.00114296 | 0.000292095 |
| sp\|Q9Y2V2\|CHSP1_HUMAN | 2 | 30 | 55 | 23 | -0.51584 | -0.211309047 | 0.25995396 | 0.304532792 | 0.775795799 | 0.471263007 | 0.066960672 | 2.32E-05 | 0.005311498 |
| sp\|Q10589\|BST2_HUMAN | 7 | 26 | 48 | 20 | -0.63504 | -0.165338103 | 0.377803533 | 0.469701465 | 1.0128431 | 0.543141636 | 0.044833921 | 0.000132863 | 0.031498638 |
| sp\|P08861\|CEL3B_HUMAN | 9 | 36 | 69 | 30 | -0.23754 | -0.230471201 | 0.136846922 | 0.007073541 | 0.374391664 | 0.367318123 | 0.998401912 | 0.049939514 | 0.025741482 |
| sp\|Q9NNX6\|CD209_HUMAN | 1 | 11 | 20 | 9 | -0.06518 | -0.05744733 | 0.354361634 | 0.007729319 | 0.419538283 | 0.411808963 | 0.998122118 | 0.031112185 | 0.016397054 |
| sp\|Q9NT22\|EMIL3_HUMAN | 3 | 36 | 69 | 30 | 0.193502 | 0.037307391 | -0.336515258 | -0.156194562 | -0.530017211 | -0.373822649 | 0.082520867 | 3.49E-08 | 1.00E-05 |
| sp\|P29692\|EF1D_HUMAN | 1 | 29 | 55 | 25 | -0.0604 | -0.048808395 | 0.26958106 | 0.011592194 | 0.329981649 | 0.318389455 | 0.996451218 | 0.137454367 | 0.095005338 |
| sp\|Q14964\|RB39A_HUMAN | 7 | 6 | 14 | 7 | -0.68491 | -0.298670244 | 0.286921122 | 0.386244379 | 0.971835745 | 0.585591366 | 0.390001131 | 0.018600331 | 0.104033003 |
| sp\|Q96N76\|HUTU_HUMAN | 20 | 30 | 55 | 24 | -0.0866 | 0.009628172 | 0.330226053 | 0.096226372 | 0.416824253 | 0.32059788 | 0.596619192 | 0.002044889 | 0.009301689 |
| sp\|P37235\|HPCL1_HUMAN | 3 | 37 | 69 | 30 | -0.18622 | -0.091397873 | 0.175578585 | 0.094825358 | 0.361801816 | 0.266976458 | 0.693117576 | 0.02890108 | 0.085082284 |
| sp\|Q16661\|GUC2B_HUMAN | 7 | 40 | 76 | 33 | 0.020956 | -0.029808753 | 0.182459616 | -0.050764285 | 0.161504083 | 0.212268369 | 0.778088784 | 0.178130063 | 0.024337373 |
| sp\|O95865\|DDAH2_HUMAN | 4 | 40 | 76 | 33 | 0.063922 | 0.006279683 | 0.294570315 | -0.057642321 | 0.230648311 | 0.288290632 | 0.656092513 | 0.011493539 | 0.000195882 |
| sp\|P05787\|K2C8_HUMAN | 1 | 26 | 48 | 22 | -0.46767 | -0.414624151 | 0.286428484 | 0.05304627 | 0.754098905 | 0.701052635 | 0.955364735 | 0.002491131 | 0.001483803 |
| sp\|Q15904\|VAS1_HUMAN | 23 | 40 | 76 | 33 | -0.17864 | -0.173884257 | 0.319730926 | 0.004752342 | 0.498367525 | 0.493615183 | 0.998157688 | 4.09E-06 | 2.69E-07 |
| sp\|P11171\|41_HUMAN | 1 | 4 | 7 | 3 | 0.240321 | 0.153907004 | 1.010893928 | -0.086413808 | 0.770573115 | 0.856986924 | 0.975397779 | 0.303655536 | 0.179893569 |
| sp\|P98088\|MUC5A_HUMAN | 2 | 14 | 27 | 13 | 0.439153 | 0.028635008 | 0.392929441 | -0.410517681 | -0.046223249 | 0.364294432 | 0.19968474 | 0.984616808 | 0.295644333 |
| sp\|A0A087WSY4\|HV432_HUMAN | 4 | 10 | 20 | 10 | 0.253084 | 0.426128387 | 0.247490202 | 0.173044473 | -0.005593712 | -0.178638185 | 0.566196697 | 0.999546395 | 0.545805039 |
| sp\|P16870\|CBPE_HUMAN | 1 | 26 | 48 | 21 | 0.187372 | 0.114615157 | 0.044441712 | -0.07275648 | -0.142929924 | -0.070173444 | 0.517109163 | 0.177964889 | 0.587180788 |
| sp\|Q9Y333\|LSM2_HUMAN | 1 | 4 | 7 | 3 | -0.0273 | -0.121016424 | -0.314292636 | -0.093712166 | -0.286988379 | -0.193276213 | 0.541131894 | 0.047439441 | 0.149518982 |
| sp\|Q5Y5X9\|LIPE_HUMAN | 4 | 40 | 76 | 33 | 0.038051 | 0.089028048 | 0.187108885 | 0.050976707 | 0.149057544 | 0.098080836 | 0.603161061 | 0.054547749 | 0.197120888 |
| sp\|Q9H074\|PAIP1_HUMAN | 2 | 7 | 14 | 6 | 0.00794 | 0.040751685 | 0.042811494 | 0.03281198 | 0.034871789 | 0.002059809 | 0.972774904 | 0.978641435 | 0.999901982 |
| sp\|P12104\|FABPI_HUMAN | 1 | 14 | 28 | 13 | -0.11999 | -0.078464368 | 0.245261618 | 0.041523463 | 0.365249449 | 0.323725987 | 0.978881561 | 0.312943704 | 0.30092847 |
| sp\|Q9P0V9\|SEP10_HUMAN | 3 | 7 | 14 | 6 | 0.193961 | -0.109119668 | 0.598734959 | -0.303080414 | 0.404774214 | 0.707854627 | 0.695962885 | 0.640267823 | 0.187251835 |
| sp\|O00148\|DX39A_HUMAN | 5 | 12 | 21 | 9 | -0.81573 | -0.326188301 | 0.156138888 | 0.489537491 | 0.971864679 | 0.482327189 | 0.09595356 | 0.003526441 | 0.14930763 |
| sp\|P28906\|CD34_HUMAN | 3 | 40 | 76 | 33 | -0.09972 | -0.111530639 | 0.034551482 | -0.011810671 | 0.13427145 | 0.146082122 | 0.98187378 | 0.201070056 | 0.091055712 |
| sp\|P54922\|ADPRH_HUMAN | 2 | 4 | 7 | 3 | 0.122534 | 0.099215275 | 0.284983314 | -0.023318572 | 0.162449467 | 0.185768038 | 0.983122496 | 0.588806934 | 0.438401028 |
| sp\|O43583\|DENR_HUMAN | 4 | 24 | 48 | 22 | -0.18226 | 0.015505176 | 0.314480627 | 0.197765666 | 0.496741117 | 0.298975451 | 0.202058244 | 0.001193694 | 0.034753179 |
| sp\|Q6XQN6\|PNCB_HUMAN | 1 | 40 | 76 | 33 | -0.35458 | -0.168040609 | 0.137722324 | 0.186536074 | 0.492299006 | 0.305762933 | 0.065782284 | 6.28E-06 | 0.002026157 |
| sp\|P25398\|RS12_HUMAN | 7 | 4 | 7 | 3 | -0.25928 | -0.096883918 | 0.267503284 | 0.162397376 | 0.526784578 | 0.364387202 | 0.716901936 | 0.134855744 | 0.282743816 |
| sp\|P20340\|RAB6A_HUMAN | 9 | 37 | 69 | 30 | -0.17152 | -0.148948565 | 0.331051771 | 0.022571911 | 0.502572247 | 0.480000336 | 0.971055831 | 0.000111964 | 3.21E-05 |
| sp\|P06858\|LIPL_HUMAN | 8 | 40 | 76 | 33 | 0.050351 | 0.032164392 | -0.157889718 | -0.018186337 | -0.208240447 | -0.19005411 | 0.965277951 | 0.045219621 | 0.037787495 |
| sp\|P55060\|XPO2_HUMAN | 21 | 7 | 14 | 6 | 0.148309 | 0.103369985 | 0.282151354 | -0.044939176 | 0.133842193 | 0.178781369 | 0.957985449 | 0.770307953 | 0.550962821 |
| sp\|P50452\|SPB8_HUMAN | 1 | 37 | 69 | 30 | -0.35134 | -0.211629672 | 0.293067358 | 0.139707708 | 0.644404738 | 0.50469703 | 0.600726575 | 0.000950159 | 0.004208762 |
| sp\|P21583\|SCF_HUMAN | 3 | 21 | 41 | 17 | 0.756751 | 0.542652916 | 0.927765788 | -0.21409769 | 0.171015182 | 0.385112872 | 0.617544576 | 0.811378973 | 0.264388696 |
| sp\|P24534\|EF1B_HUMAN | 4 | 40 | 76 | 33 | -0.47684 | -0.316703467 | 0.109197656 | 0.160135498 | 0.58603662 | 0.425901123 | 0.470320933 | 0.001396815 | 0.011022668 |
| sp\|Q13232\|NDK3_HUMAN | 30 | 40 | 76 | 33 | 0.121649 | 0.082513522 | -0.038095376 | -0.039135572 | -0.15974447 | -0.120608898 | 0.479721193 | 0.000384058 | 0.002975653 |
| sp\|P02689\|MYP2_HUMAN | 10 | 40 | 76 | 33 | -0.2832 | -0.232588896 | 0.485465039 | 0.050610691 | 0.768664626 | 0.718053935 | 0.930221584 | 3.04E-05 | 1.07E-05 |
| sp\|Q0Z7S8\|FABP9_HUMAN | 2 | 11 | 20 | 9 | -0.29734 | -0.234922547 | 0.96848306 | 0.062417607 | 1.265823213 | 1.203405607 | 0.980472441 | 0.007667031 | 0.004397898 |
| sp\|P62834\|RAP1A_HUMAN | 6 | 40 | 76 | 33 | -0.23503 | -0.164734832 | 0.045817668 | 0.070296526 | 0.280849027 | 0.210552501 | 0.727259153 | 0.033478392 | 0.086040197 |
| sp\|P63244\|RACK1_HUMAN | 2 | 4 | 7 | 3 | -0.42716 | -0.069932186 | 0.354885039 | 0.357230297 | 0.782047523 | 0.424817225 | 0.531489907 | 0.162505556 | 0.481686809 |
| sp\|Q9BSE5\|SPEB_HUMAN | 6 | 7 | 14 | 6 | -0.2777 | 0.17275587 | 0.769636249 | 0.450457794 | 1.047338713 | 0.596880379 | 0.695707411 | 0.272346859 | 0.566677109 |
| sp\|P78536\|ADA17_HUMAN | 1 | 40 | 76 | 33 | -0.01003 | 0.076760173 | 0.223541431 | 0.086785347 | 0.233566605 | 0.146781258 | 0.182707965 | 0.000340867 | 0.015557653 |
| sp\|P07949\|RET_HUMAN | 24 | 37 | 69 | 30 | 0.017881 | 0.185285239 | -0.073156426 | 0.167404024 | -0.091037641 | -0.258441666 | 0.023899623 | 0.456287403 | 0.000590968 |
| sp\|P35052\|GPC1_HUMAN | 27 | 40 | 76 | 33 | -0.05469 | -0.072020495 | 0.191410657 | -0.017328459 | 0.246102693 | 0.263431152 | 0.957070784 | 0.003150625 | 0.000273271 |
| sp\|P19256\|LFA3_HUMAN | 3 | 40 | 76 | 33 | 0.051751 | -0.013974755 | -0.182564458 | -0.065725301 | -0.234315004 | -0.168589703 | 0.185353916 | 1.77E-06 | 0.000116408 |
| sp\|Q4G148\|GXLT1_HUMAN | 1 | 40 | 76 | 33 | 0.153968 | 0.153438936 | 0.174100761 | -0.000529448 | 0.020132377 | 0.020661825 | 0.999952171 | 0.953425802 | 0.938087543 |
| sp\|Q7L7X3\|TAOK1_HUMAN | 4 | 7 | 14 | 6 | 0.330722 | 0.482525294 | 0.922729034 | 0.151803305 | 0.592007045 | 0.44020374 | 0.905686451 | 0.367594342 | 0.48268623 |
| sp\|P07437\|TBB5_HUMAN | 5 | 40 | 76 | 33 | -0.29108 | -0.095547128 | 0.155762854 | 0.195537682 | 0.446847664 | 0.251309982 | 0.136962406 | 0.00108436 | 0.057413859 |
| sp\|Q14194\|DPYL1_HUMAN | 8 | 36 | 69 | 30 | 0.059386 | 0.009300495 | 0.248542834 | -0.050085167 | 0.189157172 | 0.239242339 | 0.757668432 | 0.069553715 | 0.005007829 |
| sp\|P41159\|LEP_HUMAN | 5 | 40 | 76 | 33 | -0.17024 | -0.190483817 | -0.136163532 | -0.020242587 | 0.034077699 | 0.054320285 | 0.985811921 | 0.972448161 | 0.913683889 |
| sp\|Q99729\|ROAA_HUMAN | 9 | 7 | 13 | 6 | -0.26929 | -0.089471488 | 0.761459927 | 0.179817496 | 1.03074891 | 0.850931414 | 0.932459934 | 0.218225942 | 0.264370881 |
| sp\|Q92692\|NECT2_HUMAN | 1 | 40 | 76 | 33 | -0.3775 | -0.256240326 | 0.422788697 | 0.12126168 | 0.800290703 | 0.679029024 | 0.465314268 | 3.87E-09 | 1.59E-08 |
| sp\|Q15349\|KS6A2_HUMAN | 1 | 4 | 7 | 2 | -0.24002 | -0.095661429 | -0.148665144 | 0.144355214 | 0.0913515 | -0.053003714 | 0.959086596 | 0.991245407 | 0.996550435 |
| sp\|P30990\|NEUT_HUMAN | 23 | 16 | 28 | 11 | 0.216119 | 0.134795533 | -0.10793923 | -0.0813232 | -0.324057964 | -0.242734763 | 0.730560519 | 0.049750996 | 0.124567466 |
| sp\|P69892\|HBG2_HUMAN | 4 | 10 | 21 | 9 | -0.03591 | -0.321804362 | 0.547515882 | -0.285890189 | 0.583430055 | 0.869320244 | 0.641082737 | 0.282989 | 0.030777294 |
| sp\|P49721\|PSB2_HUMAN | 5 | 40 | 76 | 33 | -0.26105 | -0.085252753 | 0.416291052 | 0.175792333 | 0.677336139 | 0.501543806 | 0.179320642 | 1.90E-07 | 1.35E-05 |
| sp\|P63167\|DYL1_HUMAN | 1 | 7 | 14 | 6 | 0.000134 | 0.100355283 | 0.539235048 | 0.10022136 | 0.539101125 | 0.438879765 | 0.865678704 | 0.07554962 | 0.104461066 |
| sp\|Q9Y3C8\|UFC1_HUMAN | 1 | 40 | 76 | 33 | -0.3215 | -0.142202205 | 0.209631636 | 0.179298937 | 0.531132777 | 0.351833841 | 0.197230869 | 0.000108131 | 0.005049725 |
| sp\|Q7KYR7\|BT2A1_HUMAN | 26 | 40 | 76 | 33 | -0.08401 | -0.093906542 | 0.158487864 | -0.009896825 | 0.242497581 | 0.252394406 | 0.988648864 | 0.010953044 | 0.002186981 |
| sp\|P27695\|APEX1_HUMAN | 1 | 7 | 13 | 6 | -0.65697 | -0.419946713 | 0.383244529 | 0.237023471 | 1.040214714 | 0.803191242 | 0.887951492 | 0.218702641 | 0.310075364 |
| sp\|Q9NTX5\|ECHD1_HUMAN | 11 | 4 | 7 | 3 | 0.308702 | 0.408033924 | 0.346383274 | 0.099331716 | 0.037681066 | -0.061650651 | 0.798189981 | 0.977972269 | 0.929889603 |
| sp\|Q08431\|MFGM_HUMAN | 1 | 7 | 14 | 7 | -0.24572 | -0.113203412 | -0.146344466 | 0.132512848 | 0.099371793 | -0.033141055 | 0.389102148 | 0.665208637 | 0.940459155 |
| sp\|P35556\|FBN2_HUMAN | 78 | 40 | 76 | 33 | 0.025249 | -0.018175966 | 0.038270603 | -0.04342541 | 0.013021159 | 0.056446569 | 0.544285757 | 0.962745567 | 0.406744085 |
| sp\|P49720\|PSB3_HUMAN | 19 | 40 | 76 | 33 | -0.27391 | -0.122998947 | 0.400433838 | 0.150909705 | 0.67434249 | 0.523432784 | 0.2876818 | 2.85E-07 | 6.93E-06 |
| sp\|Q8TB96\|TIP_HUMAN | 7 | 15 | 27 | 13 | 0.069334 | -0.001122261 | 0.148042897 | -0.070456006 | 0.078709152 | 0.149165158 | 0.636961472 | 0.665672376 | 0.167939078 |
| sp\|Q9H6Z4\|RANB3_HUMAN | 23 | 4 | 6 | 3 | -0.18479 | 0.29288685 | 1.118012647 | 0.477675652 | 1.302801449 | 0.825125797 | 0.723558856 | 0.220011264 | 0.46410962 |
| sp\|Q9Y262\|EIF3L_HUMAN | 9 | 10 | 21 | 10 | -0.40518 | -0.235522063 | 0.547781218 | 0.169658617 | 0.952961898 | 0.783303281 | 0.807203322 | 0.012312099 | 0.017157788 |
| sp\|Q96B86\|RGMA_HUMAN | 14 | 40 | 76 | 33 | 0.134338 | 0.021306337 | -0.213794426 | -0.113031926 | -0.348132688 | -0.235100763 | 0.044728217 | 1.88E-08 | 1.75E-05 |
| sp\|O76011\|KRT34_HUMAN | 3 | 8 | 13 | 6 | -0.29216 | 0.346976165 | -0.547139593 | 0.639132894 | -0.254982864 | -0.894115758 | 0.42397004 | 0.906733409 | 0.256572682 |
| sp\|Q96P63\|SPB12_HUMAN | 15 | 19 | 35 | 15 | 0.231569 | 0.047626916 | -0.27018631 | -0.183941616 | -0.501754842 | -0.317813226 | 0.075312942 | 1.36E-05 | 0.002103952 |
| sp\|P84077\|ARF1_HUMAN | 14 | 40 | 76 | 33 | -0.20115 | -0.106230054 | 0.199513265 | 0.094921514 | 0.400664833 | 0.305743319 | 0.750730176 | 0.032980551 | 0.078051754 |
| sp\|Q86T26\|TM11B_HUMAN | 4 | 29 | 55 | 23 | -0.3654 | -0.382149285 | 0.054706473 | -0.016744831 | 0.420110927 | 0.436855758 | 0.994484213 | 0.102111676 | 0.04564331 |
| sp\|P31323\|KAP3_HUMAN | 3 | 10 | 21 | 10 | -0.11062 | 0.212627002 | 0.243078375 | 0.323246817 | 0.353698191 | 0.030451373 | 0.366788348 | 0.410908996 | 0.990869075 |
| sp\|Q9H0V9\|LMA2L_HUMAN | 5 | 8 | 13 | 5 | 0.503701 | 0.41338021 | 0.587499009 | -0.090320954 | 0.083797844 | 0.174118799 | 0.859698531 | 0.922084944 | 0.667156024 |
| sp\|P01160\|ANF_HUMAN | 6 | 31 | 55 | 23 | -0.60238 | -0.653148442 | 0.249907302 | -0.0507649 | 0.852290844 | 0.903055744 | 0.940496058 | 3.89E-05 | 1.40E-06 |
| sp\|Q8TD06\|AGR3_HUMAN | 33 | 29 | 55 | 24 | -0.02477 | 0.033489771 | 0.226870115 | 0.058257473 | 0.251637817 | 0.193380344 | 0.902615175 | 0.272219772 | 0.374617138 |
| sp\|P59665\|DEF1_HUMAN | 3 | 40 | 76 | 33 | -0.41199 | -0.344012535 | 0.402858165 | 0.067980001 | 0.814850702 | 0.746870701 | 0.857724716 | 1.52E-06 | 6.68E-07 |
| sp\|Q8NI22\|MCFD2_HUMAN | 1 | 18 | 35 | 15 | -0.08023 | 0.049709991 | 0.332263227 | 0.129943516 | 0.412496752 | 0.282553235 | 0.36965344 | 0.001920108 | 0.019603804 |
| sp\|P60033\|CD81_HUMAN | 7 | 40 | 76 | 33 | -0.10472 | -0.029235849 | 0.2457372 | 0.075485748 | 0.350458797 | 0.274973049 | 0.594137605 | 0.000718767 | 0.003178639 |
| sp\|P31025\|LCN1_HUMAN | 2 | 29 | 55 | 24 | -0.17422 | -0.088193097 | 0.077537698 | 0.086027109 | 0.251757904 | 0.165730795 | 0.809220934 | 0.290299922 | 0.50310228 |
| sp\|Q9Y6E0\|STK24_HUMAN | 15 | 11 | 21 | 10 | 0.045196 | 0.025293859 | 0.061807279 | -0.019902142 | 0.016611278 | 0.036513421 | 0.981497374 | 0.990601245 | 0.942781903 |
| sp\|A6NDG6\|PGP_HUMAN | 4 | 18 | 34 | 15 | -0.12613 | -0.027624509 | 0.388897386 | 0.098500954 | 0.515022849 | 0.416521895 | 0.707176473 | 0.00268112 | 0.006660829 |
| sp\|P01714\|LV319_HUMAN | 29 | 28 | 55 | 25 | -0.27476 | -0.136690891 | 0.287085965 | 0.138071937 | 0.561848794 | 0.423776856 | 0.490755151 | 0.000461707 | 0.002975049 |
| sp\|Q13618\|CUL3_HUMAN | 37 | 16 | 27 | 11 | -0.13715 | -0.10022022 | 0.018437008 | 0.036927764 | 0.155584993 | 0.118657228 | 0.977737639 | 0.772693782 | 0.835130214 |
| sp\|Q9BYF1\|ACE2_HUMAN | 4 | 28 | 56 | 24 | -0.71816 | -0.592788165 | 0.013163861 | 0.125371386 | 0.731323412 | 0.605952026 | 0.659776328 | 0.000148994 | 0.000353904 |
| sp\|Q92973\|TNPO1_HUMAN | 18 | 26 | 49 | 20 | -0.38383 | -0.16863342 | 0.345784941 | 0.215197488 | 0.729615849 | 0.514418361 | 0.606275387 | 0.025785339 | 0.097464176 |
| sp\|Q9BUD6\|SPON2_HUMAN | 47 | 40 | 76 | 33 | -0.29664 | -0.184519619 | 0.398906305 | 0.112118863 | 0.695544786 | 0.583425923 | 0.391211747 | 9.38E-10 | 6.17E-09 |
| sp\|Q15714\|T22D1_HUMAN | 10 | 4 | 6 | 3 | 0.149481 | -0.015876467 | 0.276147139 | -0.165357791 | 0.126665815 | 0.292023606 | 0.803067152 | 0.911060481 | 0.576695154 |
| sp\|O75923\|DYSF_HUMAN | 6 | 4 | 6 | 3 | -0.15708 | -0.154538406 | 0.31003581 | 0.002543754 | 0.467117971 | 0.464574216 | 0.999976875 | 0.590383804 | 0.547034045 |
| sp\|Q9UEW3\|MARCO_HUMAN | 11 | 40 | 76 | 33 | -0.10529 | 0.01188891 | 0.333476871 | 0.117181933 | 0.438769895 | 0.321587962 | 0.233652547 | 3.24E-06 | 0.000131958 |
| sp\|P04233\|HG2A_HUMAN | 2 | 11 | 20 | 9 | -0.04798 | -0.098357709 | 0.609930424 | -0.050373386 | 0.657914747 | 0.708288133 | 0.963579528 | 0.019920308 | 0.004374594 |

| | | | | | | | | | | | | | |
|---|---|---|---|---|---|---|---|---|---|---|---|---|---|
| sp\|P28908\|TNR8_HUMAN | 1 | 28 | 55 | 24 | -1.40294 | -1.280843284 | -1.033095939 | 0.122097474 | 0.369844819 | 0.247747345 | 0.810926794 | 0.26724214 | 0.462449593 |
| sp\|Q96EE4\|CC126_HUMAN | 144 | 40 | 76 | 33 | 0.136424 | 0.098313797 | -0.090093372 | -0.038109921 | -0.22651709 | -0.188407169 | 0.515207228 | 6.73E-07 | 3.12E-06 |
| sp\|A8K7I4\|CLCA1_HUMAN | 1 | 6 | 14 | 7 | -0.63532 | -0.531050159 | 0.974348601 | 0.104271821 | 1.609670582 | 1.505398761 | 0.978343166 | 0.032364525 | 0.015222171 |
| sp\|P60953\|CDC42_HUMAN | 2 | 15 | 27 | 12 | -0.18245 | -0.049059437 | 0.301947681 | 0.133389121 | 0.484396239 | 0.351007118 | 0.553164422 | 0.007544128 | 0.036320511 |
| sp\|Q96J42\|TXD15_HUMAN | 1 | 40 | 76 | 33 | -0.15576 | 0.018460151 | 0.126452743 | 0.174218947 | 0.282211539 | 0.107992592 | 0.004245218 | 7.36E-05 | 0.148120644 |
| sp\|Q13501\|SQSTM_HUMAN | 2 | 4 | 6 | 3 | 0.070055 | -0.037992803 | 0.911238278 | -0.108047348 | 0.841183732 | 0.94923108 | 0.988553433 | 0.622053197 | 0.501870458 |
| sp\|Q9NPH0\|PPA6_HUMAN | 1 | 16 | 27 | 11 | 0.330865 | 0.204190784 | 0.407567076 | -0.126674481 | 0.07670181 | 0.203376291 | 0.367727761 | 0.784853505 | 0.140631133 |
| sp\|O43157\|PLXB1_HUMAN | 38 | 40 | 76 | 33 | 0.03774 | 0.015772358 | 0.080998096 | -0.021967908 | 0.04325783 | 0.065225738 | 0.710792297 | 0.403398111 | 0.075697922 |
| sp\|Q15651\|HMGN3_HUMAN | 11 | 32 | 62 | 27 | -1.6746 | -1.0868435 | -0.210477739 | 0.58775848 | 1.46412424 | 0.876365761 | 0.036171068 | 2.59E-06 | 0.001744711 |
| sp\|Q9BXJ0\|C1QT5_HUMAN | 1 | 36 | 69 | 30 | 0.080929 | 0.158336885 | -0.038312601 | 0.077407983 | -0.119241503 | -0.196649486 | 0.230541195 | 0.092183865 | 0.00040061 |
| sp\|P01040\|CYTA_HUMAN | 10 | 36 | 69 | 30 | -0.46683 | -0.491230635 | 0.103229925 | -0.024401802 | 0.570058758 | 0.594460561 | 0.98831096 | 0.014471541 | 0.00305941 |
| sp\|Q9BUN1\|MENT_HUMAN | 4 | 40 | 76 | 33 | 0.120761 | 0.088740262 | 0.191085811 | -0.032020993 | 0.070324556 | 0.102345549 | 0.74522464 | 0.378463408 | 0.076134035 |
| sp\|P52943\|CRIP2_HUMAN | 5 | 4 | 7 | 3 | 0.524771 | 0.371857787 | 0.580871854 | -0.152912727 | 0.05610134 | 0.209014067 | 0.859773166 | 0.986218737 | 0.7936535 |
| sp\|Q56UQ5\|TPT1L_HUMAN | 4 | 36 | 69 | 30 | -0.25663 | -0.058015356 | 0.220930261 | 0.198611154 | 0.477556771 | 0.278945617 | 0.262819642 | 0.005928939 | 0.099785969 |
| sp\|Q14141\|SEPT6_HUMAN | 2 | 40 | 76 | 33 | -0.34321 | -0.288948985 | 0.053707734 | 0.054258038 | 0.396914757 | 0.342656719 | 0.929424734 | 0.072018598 | 0.082218338 |
| sp\|Q8IYD1\|ERF3B_HUMAN | 1 | 26 | 48 | 21 | -0.0981 | -0.014115959 | 0.392806748 | 0.083986606 | 0.490909313 | 0.406922707 | 0.793191441 | 0.006177283 | 0.011850612 |
| sp\|Q8IYS5\|OSCAR_HUMAN | 3 | 40 | 76 | 33 | -0.04516 | 0.027223103 | 0.116197968 | 0.072388011 | 0.161362876 | 0.088974865 | 0.482610028 | 0.08601559 | 0.381377437 |
| sp\|P01718\|LV327_HUMAN | 12 | 4 | 6 | 3 | -0.14266 | -0.064339321 | 0.415846864 | 0.078323356 | 0.558509541 | 0.480186185 | 0.960393426 | 0.276365179 | 0.323664087 |
| sp\|Q9GZP8\|IMUP_HUMAN | 1 | 40 | 76 | 33 | -0.18146 | -0.264193446 | -0.353706646 | -0.082733285 | -0.172246485 | -0.0895132 | 0.879252825 | 0.681263651 | 0.876119999 |
| sp\|Q16663\|CCL15_HUMAN | 1 | 40 | 76 | 33 | -0.10087 | -0.178758274 | 0.232576473 | -0.07789277 | 0.333441976 | 0.411334746 | 0.480499674 | 0.000188968 | 1.68E-07 |
| sp\|P02810\|PRPC_HUMAN | 2 | 28 | 55 | 25 | -0.44879 | -0.594497344 | 0.053387662 | -0.145709331 | 0.502175676 | 0.647885007 | 0.757032993 | 0.101088922 | 0.008171616 |
| sp\|P15151\|PVR_HUMAN | 8 | 40 | 76 | 33 | -0.00798 | 0.02325285 | 0.101496564 | 0.031229861 | 0.109473575 | 0.078243714 | 0.737117325 | 0.079942039 | 0.190687745 |
| sp\|P28482\|MK01_HUMAN | 1 | 15 | 28 | 12 | -0.07669 | -0.125897201 | -0.121519094 | -0.049208242 | -0.044830134 | 0.004378107 | 0.956076926 | 0.974864812 | 0.999693953 |
| sp\|Q9H9Q2\|CSN7B_HUMAN | 7 | 19 | 34 | 15 | -0.29338 | -0.099228455 | 0.286818651 | 0.194152366 | 0.580199472 | 0.386047106 | 0.455494413 | 0.011081588 | 0.076810214 |
| sp\|Q8NHJ6\|LIRB4_HUMAN | 1 | 7 | 14 | 6 | -0.65539 | -0.768523358 | 0.17477274 | -0.113130907 | 0.830165191 | 0.943296098 | 0.963945705 | 0.274645095 | 0.123589692 |
| sp\|Q9HAT2\|SIAE_HUMAN | 2 | 40 | 76 | 33 | -0.15295 | -0.170256283 | 0.185343697 | -0.017305889 | 0.338294091 | 0.35559998 | 0.963213198 | 0.00011899 | 4.40E-06 |
| sp\|P15813\|CD1D_HUMAN | 12 | 18 | 35 | 15 | 0.139167 | 0.092063481 | -0.091578348 | -0.047103763 | -0.230745592 | -0.183641829 | 0.90570533 | 0.203969336 | 0.272681936 |
| sp\|P49901\|MCSP_HUMAN | 1 | 4 | 6 | 3 | 0.311945 | 0.242975725 | 0.105745752 | -0.068969103 | -0.206199076 | -0.137229973 | 0.906280521 | 0.55091132 | 0.728126075 |
| sp\|Q9UFE4\|CCD39_HUMAN | 2 | 29 | 55 | 24 | 0.716525 | 0.290763536 | -0.065459297 | -0.425761314 | -0.781984147 | -0.356222833 | 0.163217562 | 0.016476283 | 0.324094603 |
| sp\|Q14012\|KCC1A_HUMAN | 1 | 21 | 42 | 19 | 0.015884 | -0.048406691 | 0.258961231 | -0.064291128 | 0.243076794 | 0.307367922 | 0.897797438 | 0.339387256 | 0.108153751 |
| sp\|P05114\|HMGN1_HUMAN | 5 | 32 | 62 | 27 | -0.70507 | -0.55342206 | 0.074229123 | 0.151650215 | 0.779301398 | 0.627651183 | 0.60687747 | 0.000235346 | 0.000858074 |
| sp\|Q96I15\|SCLY_HUMAN | 1 | 22 | 41 | 17 | -0.15616 | 0.056761868 | 0.230662711 | 0.212921998 | 0.386822841 | 0.173900843 | 0.262973121 | 0.056222007 | 0.469944193 |
| sp\|O60268\|K0513_HUMAN | 5 | 8 | 14 | 6 | -0.07693 | 0.08879441 | -0.01610933 | 0.165727132 | 0.060823392 | -0.10490374 | 0.718953644 | 0.970125104 | 0.895713804 |
| sp\|Q9Y3B3\|TMED7_HUMAN | 6 | 22 | 41 | 18 | -0.08389 | -0.318362588 | 0.088689747 | -0.234475064 | 0.17257727 | 0.407052334 | 0.172824828 | 0.511978428 | 0.011956379 |
| sp\|Q8NBK3\|SUMF1_HUMAN | 15 | 20 | 34 | 14 | -0.07795 | 0.075899907 | 0.55646583 | 0.15384786 | 0.634413784 | 0.480565923 | 0.312000451 | 1.97E-05 | 0.000372067 |
| sp\|Q5D862\|FILA2_HUMAN | 4 | 11 | 21 | 9 | -0.16787 | 0.20971719 | -0.330389769 | 0.377582782 | -0.162524178 | -0.54010696 | 0.07212482 | 0.699248625 | 0.011724782 |
| sp\|Q8IUI8\|CRLF3_HUMAN | 3 | 37 | 69 | 29 | -0.26255 | -0.235425799 | 0.240359578 | 0.027127739 | 0.502913117 | 0.475785377 | 0.982629869 | 0.020369663 | 0.012838848 |
| sp\|Q9UKJ1\|PILRA_HUMAN | 5 | 40 | 76 | 33 | -0.37394 | -0.253921807 | 0.426045751 | 0.120015129 | 0.799982687 | 0.679967558 | 0.459462758 | 2.09E-09 | 8.46E-09 |
| sp\|O43768\|ENSA_HUMAN | 2 | 22 | 41 | 18 | -0.32206 | -0.223877816 | 0.645821444 | 0.0981865 | 0.967885761 | 0.869699261 | 0.903439209 | 0.002063919 | 0.00184282 |
| sp\|P61586\|RHOA_HUMAN | 2 | 26 | 48 | 20 | -0.09993 | -0.088540054 | -0.15726956 | 0.011392601 | -0.057336906 | -0.068729507 | 0.995813964 | 0.931323581 | 0.880235118 |
| sp\|P04279\|SEMG1_HUMAN | 7 | 6 | 14 | 7 | -0.23925 | -0.195205927 | 0.868645472 | 0.044043007 | 1.107894407 | 1.0638514 | 0.993547275 | 0.062384655 | 0.028359458 |
| sp\|Q5T1J5\|CHCH9_HUMAN | 8 | 4 | 6 | 3 | -0.5975 | 0.03690491 | 0.348739294 | 0.634406826 | 0.94624121 | 0.311834384 | 0.338993969 | 0.197545014 | 0.788044813 |
| sp\|Q9NRX4\|PHP14_HUMAN | 1 | 40 | 76 | 33 | -0.4211 | -0.180946166 | 0.418703785 | 0.240149446 | 0.839799398 | 0.599649952 | 0.0461653 | 3.01E-10 | 2.87E-07 |
| sp\|Q9Y219\|JAG2_HUMAN | 1 | 32 | 62 | 27 | 0.033829 | 0.036752172 | -0.018432236 | 0.002922717 | -0.052261691 | -0.055184408 | 0.998533805 | 0.723100936 | 0.629079833 |
| sp\|Q96NU7\|HUTI_HUMAN | 10 | 26 | 49 | 21 | -0.23839 | -0.102813886 | 0.293379751 | 0.13557454 | 0.531768178 | 0.396193637 | 0.584459933 | 0.005072267 | 0.022614492 |
| sp\|P15941\|MUC1_HUMAN | 4 | 40 | 76 | 33 | -0.14284 | -0.066244195 | 0.430883373 | 0.076596034 | 0.573723601 | 0.497127568 | 0.535071588 | 1.65E-09 | 3.60E-09 |
| sp\|P22694\|KAPCB_HUMAN | 1 | 37 | 69 | 29 | -0.18444 | -0.181224796 | 0.171642573 | 0.003210571 | 0.35607794 | 0.352867369 | 0.999661343 | 0.065376483 | 0.035377089 |
| sp\|Q9UNN5\|FAF1_HUMAN | 2 | 4 | 7 | 3 | 0.241218 | 0.3214835 | 0.01139401 | 0.080265061 | -0.229824429 | -0.31008949 | 0.94275131 | 0.727933492 | 0.504790556 |
| sp\|Q14246\|AGRE1_HUMAN | 7 | 14 | 28 | 12 | 0.014539 | 0.000212575 | 0.01168524 | -0.014326317 | -0.002853653 | 0.011472664 | 0.99509974 | 0.999865061 | 0.997168653 |
| sp\|Q16134\|ETFD_HUMAN | 17 | 4 | 7 | 3 | 0.4642 | 0.466940077 | 1.596062405 | 0.002739682 | 1.13186201 | 1.129122328 | 0.999979418 | 0.141414064 | 0.099907348 |
| sp\|P01037\|CYTN_HUMAN | 3 | 14 | 27 | 13 | 0.562539 | 0.306884798 | 0.148744331 | -0.255654451 | -0.413794919 | -0.158140467 | 0.782852978 | 0.627209619 | 0.914404328 |
| sp\|O43915\|VEGFD_HUMAN | 6 | 36 | 69 | 30 | -0.08733 | -0.058279255 | 0.02208694 | 0.029055112 | 0.109421308 | 0.080366196 | 0.878460971 | 0.285044459 | 0.419411024 |
| sp\|Q9Y2Y8\|PRG3_HUMAN | 2 | 32 | 62 | 27 | -0.01522 | -0.002448586 | 0.098829087 | 0.012775708 | 0.114053381 | 0.101277673 | 0.981036038 | 0.351534512 | 0.346928229 |
| sp\|Q5DT21\|ISK9_HUMAN | 6 | 11 | 20 | 9 | -0.12733 | -0.058137703 | -0.017672629 | 0.069197129 | 0.109662203 | 0.040465074 | 0.865555802 | 0.777178517 | 0.957605859 |
| sp\|O76009\|KT33A_HUMAN | 7 | 4 | 6 | 3 | -0.19751 | 0.664105537 | -0.080742209 | 0.861612305 | 0.116764559 | -0.744847746 | 0.525675183 | 0.991022033 | 0.663341465 |
| sp\|Q9HCU0\|CD248_HUMAN | 18 | 40 | 76 | 33 | -0.0002 | -0.030364394 | 0.393737772 | -0.03016076 | 0.393941406 | 0.424102166 | 0.877850474 | 1.39E-06 | 5.69E-09 |
| sp\|P11150\|LIPC_HUMAN | 3 | 40 | 76 | 33 | 0.140667 | 0.027104502 | 0.009627512 | -0.113562413 | -0.131039403 | -0.01747699 | 0.210127469 | 0.238097765 | 0.967568088 |
| sp\|P17096\|HMGA1_HUMAN | 2 | 4 | 6 | 3 | 0.208939 | 0.310374722 | 0.800173869 | 0.101436 | 0.591235147 | 0.489799147 | 0.959255468 | 0.398541397 | 0.472348081 |
| sp\|Q27J81\|INF2_HUMAN | 6 | 14 | 28 | 12 | -0.48449 | -0.349250819 | 0.407247383 | 0.135241729 | 0.891739931 | 0.756498202 | 0.86823216 | 0.020302989 | 0.025592772 |
| sp\|Q01523\|DEF5_HUMAN | 81 | 4 | 6 | 3 | 1.322419 | 1.931226185 | 0.970586004 | 0.608806819 | -0.351833362 | -0.960640181 | 0.363942044 | 0.770223383 | 0.148950516 |
| sp\|Q6UXH0\|ANGL8_HUMAN | 4 | 40 | 76 | 33 | 0.144854 | 0.162025734 | 0.24073041 | 0.017171563 | 0.095876239 | 0.078704676 | 0.976110505 | 0.596090325 | 0.6414779 |
| sp\|Q93033\|IGSF2_HUMAN | 1 | 16 | 27 | 11 | 0.27692 | 0.274101443 | 0.041542808 | -0.002818559 | -0.235377194 | -0.232558635 | 0.999740142 | 0.318293436 | 0.263417243 |
| sp\|Q7L5N1\|CSN6_HUMAN | 1 | 33 | 62 | 27 | -0.24693 | -0.043298241 | 0.405174976 | 0.203629618 | 0.652102836 | 0.448473217 | 0.235599008 | 8.74E-05 | 0.00296798 |
| sp\|Q5VWZ2\|LYPL1_HUMAN | 12 | 10 | 21 | 9 | 0.000923 | -0.009496067 | 0.828541377 | -0.010419229 | 0.827618215 | 0.838037444 | 0.998441255 | 0.003144051 | 0.000576599 |
| sp\|Q9UFP1\|GAK1A_HUMAN | 1 | 36 | 69 | 30 | 0.060747 | 0.024583149 | -0.022958574 | -0.036164346 | -0.083706069 | -0.047541723 | 0.88516774 | 0.637422252 | 0.830127223 |
| sp\|P22102\|PUR2_HUMAN | 15 | 9 | 21 | 10 | -0.02196 | -0.057476229 | 0.298785774 | -0.035513557 | 0.320748445 | 0.356262002 | 0.961492556 | 0.105478956 | 0.022706868 |
| sp\|O43556\|SGCE_HUMAN | 77 | 19 | 34 | 14 | -0.03658 | 0.066587234 | 0.104604446 | 0.103167255 | 0.141184468 | 0.038017212 | 0.536466446 | 0.463432639 | 0.932814726 |
| sp\|P11169\|GTR3_HUMAN | 10 | 22 | 42 | 18 | -0.18121 | 0.120478806 | 0.241298974 | 0.301693187 | 0.422513355 | 0.120820168 | 0.114071678 | 0.055786467 | 0.731078642 |
| sp\|P16671\|CD36_HUMAN | 1 | 3 | 7 | 4 | 0.018616 | -0.182291319 | 0.396505643 | -0.200907461 | 0.377889502 | 0.578796962 | 0.827899208 | 0.589806132 | 0.192278588 |
| sp\|Q9H3T3\|SEM6B_HUMAN | 1 | 36 | 69 | 30 | -0.10144 | -0.073973332 | 0.157478042 | 0.027462377 | 0.258913751 | 0.231451374 | 0.896781405 | 0.001904135 | 0.001682217 |
| sp\|P00813\|ADA_HUMAN | 1 | 40 | 76 | 33 | -0.03209 | -0.0016598 | 0.016012958 | 0.030427073 | 0.048099831 | 0.017672757 | 0.804313624 | 0.687388877 | 0.937436269 |
| sp\|Q66PJ3\|AR6P4_HUMAN | 12 | 4 | 7 | 3 | 0.353841 | 0.406870892 | 0.905601396 | 0.053029426 | 0.551759931 | 0.498730504 | 0.977289972 | 0.232769063 | 0.232517846 |
| sp\|Q8IWK6\|AGRA3_HUMAN | 4 | 40 | 76 | 33 | 0.023089 | 0.078499818 | 0.059893717 | 0.05541061 | 0.036804509 | -0.018606101 | 0.450851123 | 0.783546166 | 0.923609606 |
| sp\|Q8NBJ7\|SUMF2_HUMAN | 3 | 23 | 41 | 18 | -0.00172 | 0.007214217 | 0.044386345 | 0.00893751 | 0.046109638 | 0.037172128 | 0.987759326 | 0.799421461 | 0.834968933 |
| sp\|P42330\|AK1C3_HUMAN | 5 | 15 | 27 | 13 | -0.11979 | 0.239701377 | 0.502882198 | 0.359492635 | 0.622673456 | 0.263180821 | 0.217371572 | 0.041420088 | 0.468783027 |
| sp\|Q9UNH7\|SNX6_HUMAN | 7 | 20 | 41 | 19 | 0.269985 | 0.026012325 | 0.311013719 | -0.243972646 | 0.041028749 | 0.285001394 | 0.267306751 | 0.972740002 | 0.177790286 |
| sp\|Q16695\|H31T_HUMAN | 1 | 32 | 61 | 27 | -2.00319 | -1.847611612 | -0.333600351 | 0.155576945 | 1.669588205 | 1.51401126 | 0.861697271 | 2.48E-05 | 1.53E-05 |
| sp\|Q8N339\|MT1M_HUMAN | 1 | 26 | 48 | 20 | -1.86906 | -1.522262895 | 0.019289087 | 0.346798611 | 1.888350593 | 1.541551983 | 0.420860187 | 6.15E-07 | 4.88E-06 |
| sp\|P16452\|EPB42_HUMAN | 4 | 4 | 7 | 3 | 0.154376 | 0.261702716 | 1.448530752 | 0.107327095 | 1.294155132 | 1.186828036 | 0.977473112 | 0.154853575 | 0.147683904 |
| sp\|P08865\|RSSA_HUMAN | 1 | 14 | 28 | 12 | -0.32196 | -0.10448897 | 0.342189282 | 0.217467649 | 0.6641459 | 0.446678251 | 0.592207 | 0.041588375 | 0.145857699 |
| sp\|Q99829\|CPNE1_HUMAN | 10 | 40 | 76 | 33 | 0.019518 | 0.100988057 | 0.252054775 | 0.081470345 | 0.232537062 | 0.151066718 | 0.616556456 | 0.070136994 | 0.235718417 |
| sp\|Q32MZ4\|LRRF1_HUMAN | 231 | 7 | 14 | 5 | 0.218915 | 0.387793847 | 0.464138054 | 0.168878721 | 0.245222928 | 0.076344207 | 0.88621575 | 0.853085346 | 0.980607336 |
| sp\|Q9NZU5\|LMCD1_HUMAN | 1 | 4 | 6 | 3 | -0.0109 | -0.083930733 | 0.50610888 | -0.073033837 | 0.517005776 | 0.590039613 | 0.95480102 | 0.239496884 | 0.130863391 |
| sp\|Q00688\|FKBP3_HUMAN | 5 | 37 | 69 | 30 | 0.04841 | 0.024502217 | 0.302936109 | -0.023907611 | 0.254526281 | 0.278433892 | 0.980897811 | 0.227831951 | 0.109064633 |
| sp\|Q96I99\|SUCB2_HUMAN | 5 | 4 | 7 | 3 | 0.186645 | 0.466749803 | 1.145929423 | 0.280105118 | 0.959284738 | 0.67917962 | 0.691949928 | 0.091947565 | 0.204622666 |
| sp\|P35613\|BASI_HUMAN | 1 | 4 | 7 | 3 | -0.21324 | -0.080536632 | 0.216049743 | 0.132698926 | 0.4292853 | 0.296586375 | 0.837645415 | 0.320475426 | 0.499249797 |
| sp\|Q9UHB6\|LIMA1_HUMAN | 15 | 7 | 14 | 6 | -0.29354 | -0.364197859 | 0.433963697 | -0.070653956 | 0.727507601 | 0.798161557 | 0.940508295 | 0.022569093 | 0.004160637 |
| sp\|P51805\|PLXA3_HUMAN | 6 | 7 | 14 | 5 | 0.089233 | 0.058673558 | 0.068210142 | -0.03055946 | -0.021022875 | 0.009536585 | 0.934960927 | 0.980260969 | 0.994827666 |
| sp\|O60242\|AGRB3_HUMAN | 1 | 11 | 20 | 10 | 0.116204 | 0.006827586 | 0.098349242 | -0.109376825 | -0.017855169 | 0.091521656 | 0.333950561 | 0.977960494 | 0.482185672 |
| sp\|O75063\|XYLK_HUMAN | 23 | 14 | 27 | 13 | 0.159683 | 0.363456023 | 0.508327341 | 0.203773178 | 0.348644497 | 0.144871319 | 0.170509521 | 0.026606125 | 0.419334795 |
| sp\|P18627\|LAG3_HUMAN | 3 | 8 | 13 | 5 | -0.08959 | 0.146024675 | 0.384942465 | 0.235615066 | 0.474532856 | 0.23891779 | 0.667054012 | 0.370771103 | 0.73711459 |
| sp\|Q08554\|DSC1_HUMAN | 1 | 40 | 76 | 33 | 0.178683 | 0.093975006 | 0.104448706 | -0.084708132 | -0.074234432 | 0.0104737 | 0.119620744 | 0.32082297 | 0.971282135 |
| sp\|Q9NZD4\|AHSP_HUMAN | 97 | 40 | 76 | 33 | -0.78445 | -0.469631613 | -0.027729385 | 0.314821496 | 0.756723724 | 0.441902228 | 0.033828406 | 4.11E-06 | 0.003267188 |
| sp\|O00560\|SDCB1_HUMAN | 14 | 15 | 28 | 13 | 0.192166 | 0.23537447 | 0.17764485 | 0.043208513 | -0.014521107 | -0.05772962 | 0.905553037 | 0.992025991 | 0.851508862 |
| sp\|Q15517\|CDSN_HUMAN | 11 | 25 | 48 | 21 | 0.062495 | 0.078314015 | 0.094239421 | 0.015819105 | 0.031744511 | 0.015925406 | 0.960858255 | 0.894477532 | 0.964674255 |
| sp\|Q6P5S2\|LEG1H_HUMAN | 4 | 4 | 7 | 3 | 1.049719 | 0.207831474 | 0.023086388 | -0.841887362 | -1.026632448 | -0.184745085 | 0.07208635 | 0.07186573 | 0.875062994 |
| sp\|P52565\|GDIR1_HUMAN | 15 | 40 | 76 | 33 | -0.13905 | -0.099685709 | 0.331630976 | 0.039364185 | 0.47068087 | 0.431316685 | 0.940778555 | 0.003393995 | 0.002348931 |
| sp\|Q16620\|NTRK2_HUMAN | 10 | 40 | 76 | 33 | 0.151119 | 0.086997221 | -0.195379815 | -0.064122047 | -0.346499083 | -0.282377035 | 0.330658513 | 6.06E-09 | 8.00E-08 |
| sp\|Q01970\|PLCB3_HUMAN | 12 | 26 | 48 | 21 | 0.023106 | 0.046770588 | 0.266898894 | 0.023664903 | 0.243793209 | 0.220128306 | 0.961827053 | 0.064832631 | 0.060667791 |
| sp\|P08236\|BGLR_HUMAN | 4 | 10 | 21 | 9 | 0.10676 | -0.001302585 | 0.09326753 | -0.108062609 | -0.013492495 | 0.094570115 | 0.684473125 | 0.995827728 | 0.762669416 |
| sp\|P24592\|IBP6_HUMAN | 11 | 40 | 76 | 33 | 0.098201 | 0.048089234 | 0.299694611 | -0.050111614 | 0.201493763 | 0.251605377 | 0.642006236 | 0.008821335 | 0.000122115 |
| sp\|P13747\|HLAE_HUMAN | 3 | 28 | 55 | 25 | -0.15121 | 0.023898771 | 0.349666023 | 0.175112217 | 0.50087947 | 0.325767253 | 0.227810705 | 0.000355618 | 0.010483009 |
| sp\|P23588\|IF4B_HUMAN | 11 | 11 | 21 | 9 | -0.50122 | -0.167415813 | 0.604573523 | 0.333801575 | 1.10579091 | 0.771989336 | 0.597299358 | 0.029009762 | 0.102211172 |
| sp\|P15374\|UCHL3_HUMAN | 5 | 7 | 14 | 6 | -0.12656 | -0.014647135 | 0.549972533 | 0.111908969 | 0.676528637 | 0.564619668 | 0.861958283 | 0.038490693 | 0.050455444 |
| sp\|Q12794\|HYAL1_HUMAN | 10 | 19 | 34 | 15 | -0.10666 | -0.023543122 | 0.060762501 | 0.0831131 | 0.167418723 | 0.084305624 | 0.706135624 | 0.383047714 | 0.736419835 |

<table>
<tr><td>sp|P25325|THTM_HUMAN</td><td>13</td><td>40</td><td>76</td><td>33</td><td>-0.1187</td><td>-0.081945974</td><td>0.148530866</td><td>0.036758473</td><td>0.267235313</td><td>0.23047684</td><td>0.887620141</td><td>0.015469346</td><td>0.019171997</td></tr>
<tr><td>sp|Q9H299|SH3L3_HUMAN</td><td>4</td><td>40</td><td>76</td><td>33</td><td>-0.03527</td><td>-0.153845103</td><td>0.276733707</td><td>-0.118571652</td><td>0.312007158</td><td>0.43057881</td><td>0.692050032</td><td>0.176675105</td><td>0.016564093</td></tr>
<tr><td>sp|Q16555|DPYL2_HUMAN</td><td>17</td><td>40</td><td>76</td><td>33</td><td>-0.20666</td><td>-0.155245816</td><td>0.109090895</td><td>0.051411814</td><td>0.315748526</td><td>0.264336711</td><td>0.83819436</td><td>0.012306134</td><td>0.019449571</td></tr>
<tr><td>sp|P55809|SCOT1_HUMAN</td><td>4</td><td>4</td><td>6</td><td>3</td><td>0.045647</td><td>0.416955692</td><td>0.286668257</td><td>0.371308713</td><td>0.241021278</td><td>-0.130287435</td><td>0.335819998</td><td>0.701088408</td><td>0.883342788</td></tr>
<tr><td>sp|Q13609|DNSL3_HUMAN</td><td>1</td><td>16</td><td>27</td><td>11</td><td>-0.22162</td><td>-0.219136391</td><td>0.139085379</td><td>0.002483629</td><td>0.360705399</td><td>0.358221769</td><td>0.999888772</td><td>0.22977162</td><td>0.17751223</td></tr>
<tr><td>sp|P80748|LV321_HUMAN</td><td>1</td><td>4</td><td>7</td><td>3</td><td>-0.01278</td><td>-0.048879932</td><td>0.171272339</td><td>-0.036099994</td><td>0.184052278</td><td>0.220152272</td><td>0.948317902</td><td>0.423341452</td><td>0.240359199</td></tr>
<tr><td>sp|Q9BRA2|TXD17_HUMAN</td><td>16</td><td>40</td><td>76</td><td>33</td><td>-0.36837</td><td>-0.175098772</td><td>0.259481933</td><td>0.193269976</td><td>0.62785068</td><td>0.434580705</td><td>0.142926135</td><td>2.82E-06</td><td>0.000295688</td></tr>
<tr><td>sp|Q969H8|MYDGF_HUMAN</td><td>6</td><td>40</td><td>76</td><td>33</td><td>-0.28814</td><td>-0.247213392</td><td>0.33946372</td><td>0.040923803</td><td>0.627600914</td><td>0.586677112</td><td>0.921692863</td><td>7.46E-06</td><td>2.28E-06</td></tr>
<tr><td>sp|Q13347|EIF3I_HUMAN</td><td>5</td><td>4</td><td>7</td><td>3</td><td>-0.27312</td><td>-0.139066389</td><td>-0.37814624</td><td>0.134049688</td><td>-0.105030164</td><td>-0.239079852</td><td>0.901574651</td><td>0.957828816</td><td>0.765161405</td></tr>
<tr><td>sp|Q5BLP8|CD048_HUMAN</td><td>2</td><td>14</td><td>28</td><td>13</td><td>-0.00959</td><td>-0.018077273</td><td>-0.002498604</td><td>-0.008489364</td><td>0.007089305</td><td>0.015578669</td><td>0.997877283</td><td>0.998930306</td><td>0.993217023</td></tr>
<tr><td>sp|P51148|RAB5C_HUMAN</td><td>49</td><td>4</td><td>7</td><td>3</td><td>0.358012</td><td>0.05685159</td><td>0.311064206</td><td>-0.301160153</td><td>-0.046947537</td><td>0.254212616</td><td>0.482055136</td><td>0.987287373</td><td>0.643358772</td></tr>
<tr><td>sp|P48595|SPB10_HUMAN</td><td>9</td><td>10</td><td>20</td><td>10</td><td>-0.30398</td><td>-0.259496144</td><td>0.529644748</td><td>0.044484318</td><td>0.833625211</td><td>0.789140893</td><td>0.990843667</td><td>0.10437489</td><td>0.069501631</td></tr>
<tr><td>sp|Q00839|HNRPU_HUMAN</td><td>2</td><td>10</td><td>21</td><td>9</td><td>0.182851</td><td>0.4357232</td><td>1.179890237</td><td>0.252872414</td><td>0.997039451</td><td>0.744167037</td><td>0.76728715</td><td>0.069044871</td><td>0.132563123</td></tr>
<tr><td>sp|O14933|UB2L6_HUMAN</td><td>6</td><td>10</td><td>21</td><td>10</td><td>-0.14757</td><td>0.103092736</td><td>0.505602146</td><td>0.25066726</td><td>0.653176671</td><td>0.40250941</td><td>0.47545766</td><td>0.032201345</td><td>0.156640197</td></tr>
<tr><td>sp|P04632|CPNS1_HUMAN</td><td>31</td><td>40</td><td>76</td><td>33</td><td>0.10255</td><td>0.142488859</td><td>0.438305</td><td>0.039938791</td><td>0.335754932</td><td>0.295816141</td><td>0.823809468</td><td>0.000169087</td><td>0.000186594</td></tr>
<tr><td>sp|P26599|PTBP1_HUMAN</td><td>4</td><td>17</td><td>35</td><td>16</td><td>-0.25893</td><td>-0.111179936</td><td>0.374083776</td><td>0.147752001</td><td>0.633015713</td><td>0.485263712</td><td>0.561144311</td><td>0.001114185</td><td>0.004269742</td></tr>
<tr><td>sp|O75752|B3GL1_HUMAN</td><td>5</td><td>40</td><td>76</td><td>33</td><td>0.073452</td><td>0.011457971</td><td>-0.03545984</td><td>-0.061993535</td><td>-0.108911346</td><td>-0.046917811</td><td>0.226230619</td><td>0.044500216</td><td>0.470865714</td></tr>
<tr><td>sp|P61956|SUMO2_HUMAN</td><td>9</td><td>22</td><td>42</td><td>17</td><td>-0.15649</td><td>-0.157442471</td><td>0.565817875</td><td>-0.000947484</td><td>0.722312861</td><td>0.723260346</td><td>0.999977062</td><td>0.000405688</td><td>6.70E-05</td></tr>
<tr><td>sp|Q6EEV6|SUMO4_HUMAN</td><td>2</td><td>22</td><td>41</td><td>19</td><td>0.023354</td><td>-0.095200643</td><td>0.314207986</td><td>-0.118555139</td><td>0.29085349</td><td>0.409408629</td><td>0.712351124</td><td>0.239807025</td><td>0.030643445</td></tr>
<tr><td>sp|A4D0S4|LAMB4_HUMAN</td><td>12</td><td>32</td><td>63</td><td>28</td><td>0.009813</td><td>0.018592422</td><td>0.129994932</td><td>0.008779429</td><td>0.120181938</td><td>0.11140251</td><td>0.981292483</td><td>0.088965984</td><td>0.067879467</td></tr>
<tr><td>sp|Q8NFZ3|NLGNY_HUMAN</td><td>26</td><td>14</td><td>28</td><td>13</td><td>0.038224</td><td>0.095512178</td><td>0.020805526</td><td>0.057288509</td><td>-0.017418143</td><td>-0.074706652</td><td>0.806083311</td><td>0.985644653</td><td>0.706358378</td></tr>
<tr><td>sp|P10747|CD28_HUMAN</td><td>13</td><td>8</td><td>13</td><td>5</td><td>-0.15554</td><td>0.028721999</td><td>-0.111763353</td><td>0.184259833</td><td>0.043774481</td><td>-0.140485353</td><td>0.922280153</td><td>0.99715644</td><td>0.966214465</td></tr>
<tr><td>sp|P99999|CYC_HUMAN</td><td>2</td><td>40</td><td>76</td><td>33</td><td>-0.08774</td><td>-0.033583976</td><td>0.063751641</td><td>0.05416066</td><td>0.151496277</td><td>0.097335616</td><td>0.696332691</td><td>0.146311944</td><td>0.36088883</td></tr>
<tr><td>sp|Q96FV2|SCRN2_HUMAN</td><td>1</td><td>36</td><td>70</td><td>30</td><td>0.060396</td><td>0.0155055</td><td>0.121877653</td><td>-0.044890734</td><td>0.061481419</td><td>0.106372153</td><td>0.834933177</td><td>0.79232775</td><td>0.411626117</td></tr>
<tr><td>sp|P50225|ST1A1_HUMAN</td><td>3</td><td>33</td><td>62</td><td>27</td><td>-0.08085</td><td>-0.16570238</td><td>0.164073907</td><td>-0.08484838</td><td>0.244927907</td><td>0.329776287</td><td>0.849028988</td><td>0.393937662</td><td>0.121366193</td></tr>
<tr><td>sp|P46782|RS5_HUMAN</td><td>1</td><td>4</td><td>6</td><td>3</td><td>-0.46447</td><td>-0.355664474</td><td>0.152327912</td><td>0.108805015</td><td>0.616797402</td><td>0.507992387</td><td>0.955922757</td><td>0.390160488</td><td>0.467947553</td></tr>
<tr><td>sp|P15586|GNS_HUMAN</td><td>1</td><td>19</td><td>35</td><td>15</td><td>-0.00226</td><td>0.006826454</td><td>0.314651557</td><td>0.009084257</td><td>0.316909361</td><td>0.307825103</td><td>0.993853893</td><td>0.00928011</td><td>0.004281957</td></tr>
<tr><td>sp|P26022|PTX3_HUMAN</td><td>11</td><td>40</td><td>76</td><td>33</td><td>-0.58731</td><td>-0.433677554</td><td>0.056334212</td><td>0.153636509</td><td>0.643648275</td><td>0.490011765</td><td>0.367598847</td><td>1.67E-05</td><td>0.000245288</td></tr>
<tr><td>sp|O00161|SNP23_HUMAN</td><td>6</td><td>36</td><td>69</td><td>30</td><td>-0.31989</td><td>-0.236309712</td><td>0.016291629</td><td>0.083582534</td><td>0.336183876</td><td>0.252601341</td><td>0.750812344</td><td>0.04470294</td><td>0.104060255</td></tr>
<tr><td>sp|Q9NTN9|SEM4G_HUMAN</td><td>8</td><td>40</td><td>76</td><td>33</td><td>0.036519</td><td>0.049291555</td><td>-0.02181018</td><td>0.012772324</td><td>-0.058329411</td><td>-0.071101735</td><td>0.971199161</td><td>0.657842084</td><td>0.454425074</td></tr>
<tr><td>sp|P43487|RANG_HUMAN</td><td>4</td><td>40</td><td>76</td><td>33</td><td>-0.25099</td><td>-0.105716858</td><td>0.221041294</td><td>0.145277696</td><td>0.472035848</td><td>0.326758152</td><td>0.339299218</td><td>0.000615753</td><td>0.009757017</td></tr>
<tr><td>sp|P46940|IQGA1_HUMAN</td><td>2</td><td>22</td><td>41</td><td>18</td><td>0.200983</td><td>0.234579474</td><td>0.54423006</td><td>0.033596471</td><td>0.343247057</td><td>0.309650585</td><td>0.974086373</td><td>0.158990123</td><td>0.151177209</td></tr>
<tr><td>sp|P40939|ECHA_HUMAN</td><td>4</td><td>4</td><td>7</td><td>3</td><td>0.055732</td><td>0.024483338</td><td>1.454645463</td><td>-0.031249079</td><td>1.398913046</td><td>1.430162125</td><td>0.998845773</td><td>0.255119743</td><td>0.183865829</td></tr>
<tr><td>sp|Q86WC4|OSTM1_HUMAN</td><td>9</td><td>19</td><td>35</td><td>14</td><td>0.017595</td><td>0.034551344</td><td>-0.233799677</td><td>0.016956141</td><td>-0.25139488</td><td>-0.268351021</td><td>0.952801252</td><td>0.002035831</td><td>0.000221997</td></tr>
<tr><td>sp|Q9H3K6|BOLA2_HUMAN</td><td>5</td><td>33</td><td>62</td><td>28</td><td>-0.37312</td><td>-0.084623399</td><td>0.306301076</td><td>0.288494011</td><td>0.679418486</td><td>0.390924475</td><td>0.011626586</td><td>1.94E-07</td><td>0.000816341</td></tr>
<tr><td>sp|P04118|COL_HUMAN</td><td>1</td><td>12</td><td>20</td><td>9</td><td>-0.22249</td><td>-0.264784514</td><td>0.449669433</td><td>-0.042294953</td><td>0.672158994</td><td>0.714453947</td><td>0.985966118</td><td>0.101740271</td><td>0.047561324</td></tr>
<tr><td>sp|Q16849|PTPRN_HUMAN</td><td>2</td><td>7</td><td>13</td><td>6</td><td>0.190037</td><td>0.057385284</td><td>0.541828534</td><td>-0.13265183</td><td>0.351791421</td><td>0.48444325</td><td>0.895347461</td><td>0.58226446</td><td>0.283868077</td></tr>
<tr><td>sp|P09093|CEL3A_HUMAN</td><td>2</td><td>40</td><td>76</td><td>33</td><td>-0.23224</td><td>-0.148989679</td><td>-0.032729791</td><td>0.083250296</td><td>0.199510184</td><td>0.116259888</td><td>0.707932208</td><td>0.258150195</td><td>0.554386395</td></tr>
<tr><td>sp|P45877|PPIC_HUMAN</td><td>12</td><td>40</td><td>76</td><td>33</td><td>0.171326</td><td>0.123114417</td><td>0.501590265</td><td>-0.048211439</td><td>0.330264409</td><td>0.378475848</td><td>0.71025184</td><td>4.24E-05</td><td>1.15E-07</td></tr>
<tr><td>sp|Q96L96|ALPK3_HUMAN</td><td>4</td><td>4</td><td>6</td><td>3</td><td>-0.18244</td><td>-0.08912599</td><td>0.520849006</td><td>0.093318515</td><td>0.703293511</td><td>0.609974996</td><td>0.983724854</td><td>0.535567926</td><td>0.575677566</td></tr>
<tr><td>sp|Q13451|FKBP5_HUMAN</td><td>3</td><td>15</td><td>27</td><td>12</td><td>-0.20583</td><td>-0.379568045</td><td>0.316475863</td><td>-0.173735891</td><td>0.522308017</td><td>0.696043908</td><td>0.725544303</td><td>0.145059292</td><td>0.01713505</td></tr>
<tr><td>sp|P62633|CNBP_HUMAN</td><td>2</td><td>7</td><td>14</td><td>6</td><td>-0.89258</td><td>-0.629965635</td><td>0.7354263</td><td>0.262612625</td><td>1.62800456</td><td>1.365391935</td><td>0.745014779</td><td>0.00245628</td><td>0.00369313</td></tr>
<tr><td>sp|P56537|IF6_HUMAN</td><td>2</td><td>7</td><td>14</td><td>6</td><td>-0.3092</td><td>-0.006061445</td><td>0.308480582</td><td>0.303138611</td><td>0.617680638</td><td>0.314542028</td><td>0.203235386</td><td>0.016870046</td><td>0.212832011</td></tr>
<tr><td>sp|Q53RD9|FBLN7_HUMAN</td><td>6</td><td>40</td><td>76</td><td>33</td><td>-0.03434</td><td>-0.037057739</td><td>0.210527746</td><td>-0.002722476</td><td>0.244863009</td><td>0.247585485</td><td>0.998170158</td><td>8.96E-05</td><td>7.13E-06</td></tr>
<tr><td>sp|P30530|UFO_HUMAN</td><td>27</td><td>40</td><td>76</td><td>33</td><td>-0.00158</td><td>-0.049391831</td><td>0.057477275</td><td>-0.047810747</td><td>0.05905836</td><td>0.106869106</td><td>0.433099704</td><td>0.414548948</td><td>0.0281981</td></tr>
<tr><td>sp|Q9P2E9|RRBP1_HUMAN</td><td>7</td><td>18</td><td>34</td><td>16</td><td>-0.44139</td><td>-0.281903602</td><td>0.036041142</td><td>0.159482912</td><td>0.477427657</td><td>0.317944744</td><td>0.866790165</td><td>0.403094919</td><td>0.59367015</td></tr>
<tr><td>sp|P06576|ATPB_HUMAN</td><td>9</td><td>23</td><td>41</td><td>18</td><td>0.095552</td><td>0.065411266</td><td>0.148517364</td><td>-0.030140688</td><td>0.05296541</td><td>0.083106098</td><td>0.948449843</td><td>0.894138104</td><td>0.711791549</td></tr>
<tr><td>sp|P26010|ITB7_HUMAN</td><td>2</td><td>11</td><td>20</td><td>9</td><td>0.157607</td><td>0.108662831</td><td>0.134946247</td><td>-0.048943878</td><td>-0.022660462</td><td>0.026283416</td><td>0.790114518</td><td>0.965178777</td><td>0.942006862</td></tr>
<tr><td>sp|P62805|H4_HUMAN</td><td>2</td><td>40</td><td>76</td><td>33</td><td>-1.20159</td><td>-1.109526427</td><td>-0.055256677</td><td>0.092062822</td><td>1.146332572</td><td>1.05426975</td><td>0.896917338</td><td>2.73E-05</td><td>1.32E-05</td></tr>
<tr><td>sp|P48509|CD151_HUMAN</td><td>9</td><td>4</td><td>7</td><td>3</td><td>-0.24169</td><td>-0.178336558</td><td>0.726804751</td><td>0.063350376</td><td>0.968491684</td><td>0.905141308</td><td>0.993952626</td><td>0.41564359</td><td>0.392882277</td></tr>
<tr><td>sp|Q8TCD5|NT5C_HUMAN</td><td>20</td><td>24</td><td>41</td><td>17</td><td>0.047837</td><td>0.014953118</td><td>0.253818473</td><td>-0.032883958</td><td>0.205981397</td><td>0.238865355</td><td>0.9470729</td><td>0.253486037</td><td>0.110906884</td></tr>
<tr><td>sp|P12273|PIP_HUMAN</td><td>8</td><td>33</td><td>62</td><td>27</td><td>0.071268</td><td>0.011478608</td><td>0.024463351</td><td>-0.059789527</td><td>-0.046804784</td><td>0.012984742</td><td>0.629936069</td><td>0.822084651</td><td>0.981042571</td></tr>
<tr><td>sp|Q8WUA8|TSK_HUMAN</td><td>22</td><td>40</td><td>76</td><td>33</td><td>-0.18443</td><td>-0.064286748</td><td>0.209268951</td><td>0.120143539</td><td>0.393699238</td><td>0.273555699</td><td>0.478118084</td><td>0.005392949</td><td>0.037947434</td></tr>
<tr><td>sp|O95502|NPTXR_HUMAN</td><td>6</td><td>40</td><td>76</td><td>33</td><td>0.091742</td><td>-0.00949521</td><td>-0.296706231</td><td>-0.101237273</td><td>-0.388448294</td><td>-0.287211021</td><td>0.208207822</td><td>7.13E-07</td><td>3.73E-05</td></tr>
<tr><td>sp|P55957|BID_HUMAN</td><td>10</td><td>11</td><td>21</td><td>10</td><td>0.133593</td><td>0.060789611</td><td>0.425385172</td><td>-0.072803001</td><td>0.29179256</td><td>0.364595561</td><td>0.870170587</td><td>0.21128101</td><td>0.049413645</td></tr>
<tr><td>sp|Q9C005|DPY30_HUMAN</td><td>1</td><td>6</td><td>14</td><td>7</td><td>0.127547</td><td>0.207820997</td><td>1.034147794</td><td>0.080274192</td><td>0.906600989</td><td>0.826326797</td><td>0.960293955</td><td>0.033212007</td><td>0.018715778</td></tr>
<tr><td>sp|P06748|NPM_HUMAN</td><td>1</td><td>7</td><td>14</td><td>5</td><td>-0.46682</td><td>-0.075993851</td><td>0.754941002</td><td>0.39082472</td><td>1.221759573</td><td>0.830934853</td><td>0.710717805</td><td>0.144724843</td><td>0.309963645</td></tr>
<tr><td>sp|Q13724|MOGS_HUMAN</td><td>5</td><td>33</td><td>62</td><td>27</td><td>0.193169</td><td>0.193198039</td><td>0.154601638</td><td>2.86E-05</td><td>-0.038567834</td><td>-0.038596401</td><td>0.999999807</td><td>0.784723745</td><td>0.735456304</td></tr>
<tr><td>sp|P55286|CADH8_HUMAN</td><td>7</td><td>4</td><td>7</td><td>3</td><td>0.009908</td><td>0.01672207</td><td>0.10685321</td><td>0.00681405</td><td>0.096945191</td><td>0.09013114</td><td>0.997739794</td><td>0.74096598</td><td>0.728387743</td></tr>
<tr><td>sp|P55327|TPD52_HUMAN</td><td>3</td><td>7</td><td>13</td><td>6</td><td>0.226879</td><td>0.180204597</td><td>0.475680872</td><td>-0.046674415</td><td>0.248801861</td><td>0.295476276</td><td>0.97933458</td><td>0.661324544</td><td>0.48192555</td></tr>
<tr><td>sp|O43399|TPD54_HUMAN</td><td>12</td><td>8</td><td>13</td><td>6</td><td>-0.08002</td><td>-0.123453465</td><td>-0.315054974</td><td>-0.043431613</td><td>-0.235033122</td><td>-0.191601509</td><td>0.965030352</td><td>0.49657308</td><td>0.570862605</td></tr>
<tr><td>sp|P01906|DQA2_HUMAN</td><td>1</td><td>28</td><td>56</td><td>25</td><td>-0.21534</td><td>-0.060811587</td><td>0.051161036</td><td>0.154533359</td><td>0.266505981</td><td>0.111972623</td><td>0.535293722</td><td>0.271601402</td><td>0.737053522</td></tr>
<tr><td>sp|P48745|CCN3_HUMAN</td><td>104</td><td>40</td><td>76</td><td>33</td><td>-0.31851</td><td>-0.3465985</td><td>0.113357256</td><td>-0.028085513</td><td>0.431870243</td><td>0.459955756</td><td>0.941384333</td><td>0.000121513</td><td>3.37E-06</td></tr>
<tr><td>sp|Q96JQ0|PCD16_HUMAN</td><td>1</td><td>40</td><td>76</td><td>33</td><td>0.099528</td><td>0.096455524</td><td>0.115814269</td><td>-0.003072044</td><td>0.016286701</td><td>0.019358745</td><td>0.995342836</td><td>0.913525048</td><td>0.850037814</td></tr>
<tr><td>sp|Q6NUJ1|SAPL1_HUMAN</td><td>8</td><td>4</td><td>7</td><td>3</td><td>0.025841</td><td>0.237705459</td><td>-0.145265944</td><td>0.211864514</td><td>-0.171106888</td><td>-0.382971402</td><td>0.251795611</td><td>0.521867843</td><td>0.044001908</td></tr>
<tr><td>sp|P23327|SRCH_HUMAN</td><td>1</td><td>6</td><td>14</td><td>7</td><td>0.103588</td><td>0.074237012</td><td>0.236471302</td><td>-0.029351259</td><td>0.132883032</td><td>0.16223429</td><td>0.955140004</td><td>0.495493432</td><td>0.232205576</td></tr>
<tr><td>sp|Q02413|DSG1_HUMAN</td><td>23</td><td>6</td><td>14</td><td>7</td><td>0.082254</td><td>0.131520713</td><td>-0.411726117</td><td>0.049266639</td><td>-0.49398019</td><td>-0.543246829</td><td>0.990115099</td><td>0.47575413</td><td>0.281712985</td></tr>
<tr><td>sp|P61081|UBC12_HUMAN</td><td>5</td><td>32</td><td>63</td><td>27</td><td>-0.00518</td><td>-0.026759832</td><td>0.318752831</td><td>-0.021578881</td><td>0.323933782</td><td>0.345512663</td><td>0.970325021</td><td>0.011738273</td><td>0.001687367</td></tr>
<tr><td>sp|P31152|MK04_HUMAN</td><td>2</td><td>22</td><td>41</td><td>17</td><td>0.229558</td><td>0.146720013</td><td>0.448029266</td><td>-0.082837803</td><td>0.21847145</td><td>0.301309253</td><td>0.894514991</td><td>0.597020562</td><td>0.296667246</td></tr>
<tr><td>sp|Q9H2H8|PPIL3_HUMAN</td><td>28</td><td>4</td><td>7</td><td>2</td><td>0.023781</td><td>-0.056253511</td><td>-0.137549009</td><td>-0.080034419</td><td>-0.161329917</td><td>-0.081295498</td><td>0.939000176</td><td>0.875482851</td><td>0.960999777</td></tr>
<tr><td>sp|Q8NBI6|XXLT1_HUMAN</td><td>9</td><td>14</td><td>27</td><td>12</td><td>0.06153</td><td>0.04177761</td><td>0.126285049</td><td>-0.019752496</td><td>0.064754943</td><td>0.08450744</td><td>0.962696079</td><td>0.752259805</td><td>0.538805293</td></tr>
<tr><td>sp|O95727|CRTAM_HUMAN</td><td>3</td><td>26</td><td>48</td><td>20</td><td>-0.0051</td><td>-0.00309484</td><td>0.241972998</td><td>0.002007953</td><td>0.24707579</td><td>0.245067837</td><td>0.999720898</td><td>0.065731233</td><td>0.036289559</td></tr>
<tr><td>sp|Q6FHJ7|SFRP4_HUMAN</td><td>9</td><td>40</td><td>76</td><td>33</td><td>-0.06736</td><td>-0.020126938</td><td>0.007945783</td><td>0.047234292</td><td>0.075307014</td><td>0.028072721</td><td>0.712429277</td><td>0.552623904</td><td>0.899947038</td></tr>
<tr><td>sp|O75581|LRP6_HUMAN</td><td>6</td><td>14</td><td>28</td><td>11</td><td>0.046156</td><td>0.10331158</td><td>0.03151501</td><td>0.05715574</td><td>-0.01464083</td><td>-0.07179657</td><td>0.67948476</td><td>0.983258332</td><td>0.597882112</td></tr>
<tr><td>sp|P02511|CRYAB_HUMAN</td><td>1</td><td>4</td><td>6</td><td>3</td><td>-0.19844</td><td>-0.360067962</td><td>1.570813483</td><td>-0.161623371</td><td>1.769258074</td><td>1.930881445</td><td>0.976922</td><td>0.187163393</td><td>0.110901496</td></tr>
<tr><td>sp|P62837|UB2D2_HUMAN</td><td>3</td><td>40</td><td>76</td><td>33</td><td>-0.11074</td><td>-0.076301085</td><td>0.1905693</td><td>0.034441171</td><td>0.301311556</td><td>0.266870385</td><td>0.891613402</td><td>0.003275484</td><td>0.003307533</td></tr>
<tr><td>sp|Q96Q42|ALS2_HUMAN</td><td>3</td><td>3</td><td>7</td><td>4</td><td>-0.37803</td><td>0.105178848</td><td>-0.111967914</td><td>0.483205694</td><td>0.266058933</td><td>-0.217146762</td><td>0.804648386</td><td>0.946830089</td><td>0.947392049</td></tr>
<tr><td>sp|Q9UH65|SWP70_HUMAN</td><td>1</td><td>6</td><td>14</td><td>7</td><td>-0.08262</td><td>-0.037628249</td><td>0.339908531</td><td>0.04498737</td><td>0.42252415</td><td>0.37753678</td><td>0.967904371</td><td>0.132599072</td><td>0.100241797</td></tr>
<tr><td>sp|P61960|UFM1_HUMAN</td><td>5</td><td>37</td><td>69</td><td>30</td><td>-0.31045</td><td>-0.198561796</td><td>0.250098243</td><td>0.111883457</td><td>0.560543496</td><td>0.448660039</td><td>0.507741678</td><td>2.60E-05</td><td>0.000167826</td></tr>
<tr><td>sp|Q7Z4W1|DCXR_HUMAN</td><td>10</td><td>40</td><td>76</td><td>33</td><td>-0.29581</td><td>-0.096956087</td><td>-0.061085708</td><td>0.198849165</td><td>0.234719544</td><td>0.035870379</td><td>0.175290308</td><td>0.187273693</td><td>0.950614297</td></tr>
<tr><td>sp|Q9H0Q0|FA49A_HUMAN</td><td>3</td><td>18</td><td>34</td><td>14</td><td>-0.49563</td><td>-0.435776327</td><td>0.075228826</td><td>0.059852181</td><td>0.570857335</td><td>0.511005153</td><td>0.916001519</td><td>0.007692077</td><td>0.007388501</td></tr>
<tr><td>sp|Q9UPP5|K1107_HUMAN</td><td>2</td><td>4</td><td>6</td><td>3</td><td>-0.01408</td><td>0.579812264</td><td>-0.27697043</td><td>0.59389318</td><td>-0.262889513</td><td>-0.856782694</td><td>0.502016576</td><td>0.902483255</td><td>0.320089768</td></tr>
<tr><td>sp|Q5TDH0|DDI2_HUMAN</td><td>8</td><td>25</td><td>48</td><td>21</td><td>-0.46862</td><td>-0.133974311</td><td>0.069574703</td><td>0.334647297</td><td>0.538196312</td><td>0.203549014</td><td>0.060093602</td><td>0.007433615</td><td>0.386777284</td></tr>
<tr><td>sp|P34896|GLYC_HUMAN</td><td>11</td><td>19</td><td>34</td><td>15</td><td>-0.49017</td><td>-0.317128461</td><td>0.207009868</td><td>0.173039131</td><td>0.697177461</td><td>0.524138329</td><td>0.743549281</td><td>0.043537149</td><td>0.106771298</td></tr>
<tr><td>sp|P30039|PBLD_HUMAN</td><td>1</td><td>4</td><td>7</td><td>3</td><td>0.157223</td><td>0.53357735</td><td>0.396939925</td><td>0.376354319</td><td>0.239716894</td><td>-0.136637425</td><td>0.512539544</td><td>0.825970446</td><td>0.925953108</td></tr>
<tr><td>sp|O14638|ENPP3_HUMAN</td><td>1</td><td>32</td><td>62</td><td>27</td><td>0.127019</td><td>0.07617882</td><td>-0.065660834</td><td>-0.050840613</td><td>-0.192680267</td><td>-0.141839654</td><td>0.752778473</td><td>0.064121115</td><td>0.14504659</td></tr>
<tr><td>sp|O60760|HPGDS_HUMAN</td><td>2</td><td>12</td><td>21</td><td>8</td><td>-0.00723</td><td>0.046174789</td><td>-0.287884993</td><td>0.053399867</td><td>-0.280659915</td><td>-0.334059782</td><td>0.921309722</td><td>0.254427503</td><td>0.10257888</td></tr>
<tr><td>sp|O43639|NCK2_HUMAN</td><td>5</td><td>32</td><td>62</td><td>27</td><td>0.374056</td><td>0.250056716</td><td>0.436902129</td><td>-0.123999227</td><td>0.062846187</td><td>0.186845413</td><td>0.735824183</td><td>0.946633607</td><td>0.538843723</td></tr>
<tr><td>sp|Q9Y6C2|EMIL1_HUMAN</td><td>1</td><td>40</td><td>76</td><td>33</td><td>0.039886</td><td>0.146898762</td><td>0.45417331</td><td>0.107013055</td><td>0.414287603</td><td>0.307274548</td><td>0.336781511</td><td>3.40E-05</td><td>0.000612879</td></tr>
<tr><td>sp|O75351|VPS4B_HUMAN</td><td>4</td><td>15</td><td>28</td><td>11</td><td>-0.12039</td><td>-0.187194627</td><td>0.224253285</td><td>-0.066803553</td><td>0.344644359</td><td>0.411447911</td><td>0.910173255</td><td>0.207591584</td><td>0.066181659</td></tr>
<tr><td>sp|O60256|KPRB_HUMAN</td><td>8</td><td>4</td><td>7</td><td>3</td><td>-0.44054</td><td>0.250818489</td><td>-0.343192533</td><td>0.691358116</td><td>0.097347093</td><td>-0.594011023</td><td>0.459055523</td><td>0.988870479</td><td>0.614043883</td></tr>
<tr><td>sp|Q8NES3|LFNG_HUMAN</td><td>3</td><td>23</td><td>41</td><td>17</td><td>-0.07068</td><td>0.034300351</td><td>0.153895351</td><td>0.104980096</td><td>0.224575096</td><td>0.119595</td><td>0.484535912</td><td>0.116636279</td><td>0.464679341</td></tr>
<tr><td>sp|Q9UBS9|SUCO_HUMAN</td><td>16</td><td>9</td><td>21</td><td>10</td><td>0.079608</td><td>0.193099972</td><td>0.141356325</td><td>0.11349224</td><td>0.061748593</td><td>-0.051743647</td><td>0.702276403</td><td>0.923730166</td><td>0.923421763</td></tr>
<tr><td>sp|Q5JXA9|SIRB2_HUMAN</td><td>5</td><td>11</td><td>20</td><td>9</td><td>-0.03456</td><td>0.101465029</td><td>0.351925923</td><td>0.136021858</td><td>0.386482752</td><td>0.250460894</td><td>0.357188158</td><td>0.006024033</td><td>0.055963531</td></tr>
<tr><td>sp|Q9H0X4|F234A_HUMAN</td><td>1</td><td>40</td><td>76</td><td>33</td><td>0.209418</td><td>0.15007222</td><td>-0.034054824</td><td>-0.059346069</td><td>-0.243473113</td><td>-0.184127044</td><td>0.308196229</td><td>4.61E-06</td><td>0.000101642</td></tr>
<tr><td>sp|Q6FI13|H2A2A_HUMAN</td><td>1</td><td>32</td><td>62</td><td>27</td><td>-1.30937</td><td>-1.243575407</td><td>-0.050768021</td><td>0.065798638</td><td>1.258606024</td><td>1.192807386</td><td>0.950968977</td><td>1.32E-05</td><td>2.89E-06</td></tr>
<tr><td>sp|Q15208|STK38_HUMAN</td><td>1</td><td>7</td><td>14</td><td>6</td><td>0.140602</td><td>0.118174983</td><td>0.084828134</td><td>-0.0224274</td><td>-0.055774249</td><td>-0.033346849</td><td>0.975859819</td><td>0.901004256</td><td>0.952592582</td></tr>
<tr><td>sp|Q9H6B4|CLMP_HUMAN</td><td>1</td><td>40</td><td>76</td><td>33</td><td>-0.09102</td><td>-0.128001188</td><td>0.144975489</td><td>-0.036979273</td><td>0.235997404</td><td>0.272976677</td><td>0.922671538</td><td>0.109537629</td><td>0.024526499</td></tr>
<tr><td>sp|Q8N3V7|SYNPO_HUMAN</td><td>1</td><td>4</td><td>6</td><td>3</td><td>0.129747</td><td>0.075548605</td><td>0.955976104</td><td>-0.054198882</td><td>0.826228618</td><td>0.880427499</td><td>0.994762759</td><td>0.448698548</td><td>0.355060606</td></tr>
<tr><td>sp|Q9NP55|BPIA1_HUMAN</td><td>6</td><td>3</td><td>7</td><td>4</td><td>-0.41521</td><td>0.431153105</td><td>0.129515359</td><td>0.846367758</td><td>0.544730012</td><td>-0.301637746</td><td>0.138417124</td><td>0.470540496</td><td>0.700027318</td></tr>
<tr><td>sp|Q9NS15|LTBP3_HUMAN</td><td>2</td><td>32</td><td>62</td><td>27</td><td>-0.13865</td><td>-0.097973978</td><td>0.333965516</td><td>0.040680624</td><td>0.472620117</td><td>0.431939493</td><td>0.865956212</td><td>7.46E-06</td><td>3.49E-06</td></tr>
<tr><td>sp|Q9UI17|M2GD_HUMAN</td><td>5</td><td>12</td><td>20</td><td>9</td><td>-0.08962</td><td>0.030474131</td><td>0.467099783</td><td>0.120091405</td><td>0.556717057</td><td>0.436625652</td><td>0.746519583</td><td>0.020902312</td><td>0.052260494</td></tr>
<tr><td>sp|O00764|PDXK_HUMAN</td><td>5</td><td>4</td><td>7</td><td>3</td><td>0.108758</td><td>0.278559732</td><td>0.586797084</td><td>0.169801587</td><td>0.478038939</td><td>0.308237352</td><td>0.83748809</td><td>0.413007277</td><td>0.625734556</td></tr>
<tr><td>sp|A0AVT1|UBA6_HUMAN</td><td>1</td><td>8</td><td>14</td><td>6</td><td>-0.22015</td><td>-0.098325077</td><td>0.208336787</td><td>0.121828196</td><td>0.42849006</td><td>0.306661863</td><td>0.948639507</td><td>0.649224644</td><td>0.761130486</td></tr>
<tr><td>sp|Q6IEE7|T132E_HUMAN</td><td>2</td><td>7</td><td>13</td><td>6</td><td>0.105399</td><td>0.210312352</td><td>0.156912704</td><td>0.104912997</td><td>0.051513349</td><td>-0.053399648</td><td>0.592273445</td><td>0.912530638</td><td>0.882686073</td></tr>
</table>

| | | | | | | | | | | | | | |
|---|---|---|---|---|---|---|---|---|---|---|---|---|---|
| sp\|P05198\|IF2A_HUMAN | 7 | 22 | 41 | 18 | -0.17859 | -0.009167412 | 0.357931344 | 0.169423474 | 0.53652223 | 0.367098756 | 0.610717495 | 0.038110174 | 0.138938898 |
| sp\|P34949\|MPI_HUMAN | 1 | 4 | 7 | 3 | -0.1131 | -0.250472743 | -0.040139379 | -0.13737361 | 0.072959754 | 0.210333364 | 0.795267782 | 0.956673489 | 0.647658235 |
| sp\|Q8NFZ8\|CADM4_HUMAN | 11 | 36 | 69 | 30 | 0.04587 | -0.051293499 | 0.23582418 | -0.097163701 | 0.189953977 | 0.287117678 | 0.403949863 | 0.094624458 | 0.001387606 |
| sp\|O95841\|ANGL1_HUMAN | 11 | 10 | 21 | 10 | 0.050047 | 0.067156536 | 0.144482815 | 0.017109097 | 0.094435377 | 0.07732628 | 0.992817069 | 0.850996119 | 0.863611646 |
| sp\|P09529\|INHBB_HUMAN | 6 | 22 | 41 | 19 | 0.228148 | 0.288235175 | 0.480697629 | 0.060087652 | 0.252550106 | 0.192462454 | 0.788159586 | 0.056765832 | 0.116880227 |
| sp\|Q9NWD8\|TM248_HUMAN | 2 | 18 | 34 | 14 | 0.093721 | 0.073397247 | 0.06525598 | -0.020323372 | -0.028464639 | -0.008141267 | 0.923183323 | 0.9004519 | 0.98923651 |
| sp\|Q5SRI9\|MANEA_HUMAN | 1 | 10 | 21 | 10 | -0.02344 | 0.060897981 | -0.024774389 | 0.084334404 | -0.001337965 | -0.08567237 | 0.629532422 | 0.999912816 | 0.620414583 |
| sp\|Q8N3F0\|MTURN_HUMAN | 17 | 12 | 21 | 9 | 0.230112 | 0.067780489 | 0.439453181 | -0.162331556 | 0.209341135 | 0.371672691 | 0.77281437 | 0.749475063 | 0.337040492 |
| sp\|Q14166\|TTL12_HUMAN | 16 | 7 | 14 | 6 | -0.01864 | -0.123630846 | 0.276634385 | -0.104992252 | 0.295272979 | 0.400265231 | 0.869633167 | 0.475232461 | 0.182543195 |
| sp\|Q9H2M3\|BHMT2_HUMAN | 7 | 40 | 76 | 33 | -0.28342 | -0.083205643 | 0.226764714 | 0.200212451 | 0.510182807 | 0.309970356 | 0.199546524 | 0.001056374 | 0.035712115 |
| sp\|O75791\|GRAP2_HUMAN | 7 | 29 | 56 | 24 | 0.013299 | -0.0701582 | -0.031560013 | -0.083457442 | -0.044859255 | 0.038598187 | 0.633018521 | 0.912793532 | 0.917203264 |
| sp\|Q9H0W9\|CK054_HUMAN | 10 | 12 | 21 | 9 | -0.45078 | -0.203385157 | 0.039945462 | 0.247397748 | 0.490728367 | 0.243330618 | 0.362375371 | 0.076585392 | 0.442668388 |
| sp\|O14773\|TPP1_HUMAN | 1 | 40 | 76 | 33 | -0.01553 | 0.011994608 | 0.155537007 | 0.027521674 | 0.171064074 | 0.143542399 | 0.806218693 | 0.004326992 | 0.007427484 |
| sp\|Q96DR8\|MUCL1_HUMAN | 1 | 10 | 21 | 9 | -0.02597 | 0.051085885 | -0.182140609 | 0.077050886 | -0.156175609 | -0.233226495 | 0.898374089 | 0.736402774 | 0.410148222 |
| sp\|P51124\|GRAM_HUMAN | 3 | 34 | 62 | 26 | 0.174636 | 0.198324812 | -0.01018267 | 0.023689281 | -0.184818202 | -0.208507482 | 0.887942425 | 0.010029589 | 0.000839865 |
| sp\|O15020\|SPTN2_HUMAN | 7 | 7 | 14 | 6 | -0.25174 | -0.163232132 | 0.224862229 | 0.088957414 | 0.476601575 | 0.387644161 | 0.811105041 | 0.028307776 | 0.043796745 |
| sp\|A0A075B6K4\|LV310_HUMAN | 4 | 11 | 20 | 9 | -0.08497 | -0.126924643 | -0.081619596 | -0.041957147 | 0.0033479 | 0.045305048 | 0.929758406 | 0.999675886 | 0.928424903 |
| sp\|P04899\|GNAI2_HUMAN | 8 | 11 | 20 | 9 | 0.116974 | -0.024753246 | 0.373930954 | -0.141727377 | 0.256956823 | 0.3986842 | 0.842143814 | 0.676223342 | 0.315555492 |
| sp\|P18577\|RHCE_HUMAN | 7 | 4 | 7 | 3 | -0.46239 | -0.38584723 | 1.357962172 | 0.076539258 | 1.820348661 | 1.743809403 | 0.994170348 | 0.155665005 | 0.128526785 |
| sp\|P56211\|ARP19_HUMAN | 8 | 8 | 14 | 6 | -0.02687 | -0.025071118 | 0.223746597 | 0.001794702 | 0.250612417 | 0.248817715 | 0.999890722 | 0.258738777 | 0.199151805 |
| sp\|P23378\|GCSP_HUMAN | 3 | 4 | 7 | 3 | -0.0988 | -0.000614304 | 0.897499587 | 0.0981826 | 0.996296491 | 0.89811389 | 0.97159635 | 0.182824796 | 0.184038727 |
| sp\|Q15661\|TRYB1_HUMAN | 1 | 40 | 76 | 33 | 0.077384 | 0.036895544 | 0.149862654 | -0.040488951 | 0.072478159 | 0.11296711 | 0.729930974 | 0.499965281 | 0.121080221 |
| sp\|Q9HBW1\|LRRC4_HUMAN | 9 | 16 | 27 | 12 | 0.167504 | 0.143061674 | -0.037881964 | -0.024442351 | -0.205385989 | -0.180943638 | 0.913519863 | 0.018551857 | 0.023099864 |
| sp\|Q9Y3B8\|ORN_HUMAN | 6 | 27 | 48 | 21 | -0.16453 | -0.015308032 | -0.082467707 | 0.149217567 | 0.082057892 | -0.067159674 | 0.208353728 | 0.718831401 | 0.760554325 |
| sp\|Q9UNS2\|CSN3_HUMAN | 3 | 17 | 35 | 15 | -0.08312 | 0.092227323 | 0.312502545 | 0.175350008 | 0.39562523 | 0.220275221 | 0.080430971 | 0.000317549 | 0.027959566 |
| sp\|O95166\|GBRAP_HUMAN | 1 | 18 | 34 | 14 | -0.10481 | -0.151076861 | 0.412593327 | -0.046264049 | 0.517406139 | 0.563670188 | 0.942333579 | 0.010512596 | 0.001423311 |
| sp\|O95390\|GDF11_HUMAN | 7 | 3 | 7 | 4 | 0.138893 | 0.254659351 | 0.00448885 | 0.115766258 | -0.134404242 | -0.2501705 | 0.782540643 | 0.763995565 | 0.284125116 |
| sp\|Q9H0B8\|CRLD2_HUMAN | 2 | 4 | 6 | 3 | 0.232332 | -0.048936811 | 0.346597416 | -0.28126864 | 0.114265587 | 0.395534227 | 0.76842984 | 0.96868525 | 0.652674933 |
| sp\|P55008\|AIF1_HUMAN | 1 | 18 | 34 | 14 | -0.13874 | -0.045247589 | 0.333641765 | 0.093491764 | 0.472381118 | 0.378889354 | 0.533135151 | 0.00010779 | 0.000493542 |
| sp\|Q9BXK5\|B2L13_HUMAN | 22 | 4 | 6 | 3 | -0.64224 | -0.256232967 | 0.040339692 | 0.386010846 | 0.682583505 | 0.296572659 | 0.831538098 | 0.66852894 | 0.912438999 |
| sp\|Q9UGN4\|CLM8_HUMAN | 22 | 36 | 69 | 30 | -0.25999 | -0.174822538 | 0.249528421 | 0.085165547 | 0.509516506 | 0.424350959 | 0.657774971 | 8.06E-05 | 0.000216664 |
| sp\|P61923\|COPZ1_HUMAN | 24 | 4 | 7 | 3 | -0.28791 | -0.289889651 | 0.02701929 | -0.001977988 | 0.314930953 | 0.316908941 | 0.999939923 | 0.391716132 | 0.319866397 |
| sp\|Q96AC1\|FERM2_HUMAN | 1 | 30 | 55 | 24 | -0.36961 | -0.674638485 | -0.262551123 | -0.305032586 | 0.107054776 | 0.412087362 | 0.378591853 | 0.920214659 | 0.219799773 |
| sp\|P28300\|LYOX_HUMAN | 8 | 40 | 76 | 33 | -0.21037 | -0.107971542 | 0.594466172 | 0.10240168 | 0.804839394 | 0.702437714 | 0.691939404 | 1.01E-06 | 1.48E-06 |
| sp\|P62942\|FKB1A_HUMAN | 18 | 40 | 76 | 33 | -0.20611 | -0.12056935 | 0.189967364 | 0.085535905 | 0.396072619 | 0.310536713 | 0.655812897 | 0.002731315 | 0.009341219 |
| sp\|O14556\|G3PT_HUMAN | 2 | 18 | 35 | 16 | 0.208371 | 0.175694263 | 0.07571847 | -0.032676299 | -0.132652093 | -0.099975794 | 0.855773684 | 0.169663708 | 0.267816827 |
| sp\|Q9UBX5\|FBLN5_HUMAN | 3 | 40 | 76 | 33 | -0.1267 | -0.090482731 | 0.18401435 | 0.036221307 | 0.310718389 | 0.274497082 | 0.811367655 | 6.33E-05 | 6.73E-05 |
| sp\|P01889\|HLAB_HUMAN | 12 | 40 | 76 | 33 | -0.02782 | 0.177500671 | 0.343392416 | 0.205325434 | 0.371217179 | 0.165891745 | 0.063541767 | 0.002436181 | 0.202180446 |
| sp\|Q9Y6U3\|ADSV_HUMAN | 1 | 17 | 34 | 16 | 0.387296 | 0.090303494 | -0.368901848 | -0.296992321 | -0.756197663 | -0.459205342 | 0.038855199 | 2.68E-06 | 0.00095501 |
| sp\|Q8TDZ2\|MICA1_HUMAN | 1 | 7 | 14 | 6 | -0.21997 | 0.107797838 | 0.253949341 | 0.327767659 | 0.473919162 | 0.146151503 | 0.763994589 | 0.678742919 | 0.952504862 |
| sp\|Q5SQ64\|LY66F_HUMAN | 3 | 40 | 76 | 33 | -0.34916 | 0.013312231 | 0.106334655 | 0.362467512 | 0.455489936 | 0.093022424 | 0.000848438 | 0.000470059 | 0.645843892 |
| sp\|Q8N2S1\|LTBP4_HUMAN | 1 | 40 | 76 | 33 | -0.1049 | -0.173331412 | 0.274499807 | -0.06843111 | 0.379400109 | 0.447831219 | 0.651858003 | 0.000228339 | 7.37E-07 |
| sp\|P35475\|IDUA_HUMAN | 2 | 8 | 13 | 6 | 0.088847 | 0.249875279 | 0.088461261 | 0.161028119 | -0.000385899 | -0.161414018 | 0.654227122 | 0.99999828 | 0.701494023 |
| sp\|P20962\|PTMS_HUMAN | 2 | 7 | 13 | 6 | -0.10876 | 0.073938374 | 0.456345973 | 0.18270038 | 0.56510798 | 0.382407599 | 0.664390427 | 0.081129228 | 0.216142058 |
| sp\|Q9Y5E6\|PCDB3_HUMAN | 9 | 7 | 14 | 6 | -0.09221 | 0.019627374 | 0.152163971 | 0.111838064 | 0.244374661 | 0.132536597 | 0.491666062 | 0.111677638 | 0.410665639 |
| sp\|O00139\|KIF2A_HUMAN | 1 | 21 | 42 | 18 | 0.237935 | 0.367967108 | 0.537548469 | 0.130031993 | 0.299613354 | 0.169581362 | 0.783924028 | 0.412600567 | 0.689459355 |
| sp\|P01911\|DRB_HUMAN | 1 | 40 | 76 | 33 | -0.18266 | -0.104069429 | -0.182752955 | 0.078590544 | -9.30E-05 | -0.078683526 | 0.88899585 | 0.999999886 | 0.901616572 |
| sp\|Q9H987\|SYP2L_HUMAN | 7 | 4 | 6 | 2 | -0.39022 | -0.596832695 | 2.477963797 | -0.206616234 | 2.868180258 | 3.074796492 | 0.975920189 | 0.128665492 | 0.081547453 |
| sp\|Q9UHG2\|PCS1N_HUMAN | 20 | 17 | 35 | 15 | -0.15796 | -0.231000476 | 0.061692244 | -0.073044623 | 0.219648097 | 0.29269272 | 0.886361383 | 0.472188468 | 0.178538294 |
| sp\|P55291\|CAD15_HUMAN | 4 | 29 | 55 | 24 | -0.19867 | -0.339616331 | 0.082185734 | -0.140949549 | 0.280852517 | 0.421802066 | 0.596104536 | 0.245623641 | 0.020218823 |
| sp\|P0DP04\|HV43D_HUMAN | 2 | 7 | 14 | 6 | 0.173395 | 0.283119619 | 0.250774924 | 0.109724367 | 0.077379672 | -0.032344695 | 0.764387044 | 0.91096051 | 0.978972024 |
| sp\|P0DP03\|HVC05_HUMAN | 8 | 14 | 28 | 13 | -0.03494 | -0.010094128 | 0.072755967 | 0.024844462 | 0.107694557 | 0.082850095 | 0.985451709 | 0.820340953 | 0.856825066 |
| sp\|Q13214\|SEM3B_HUMAN | 1 | 11 | 20 | 9 | 0.514553 | 0.307224402 | 0.3711916 | -0.207328197 | -0.143361 | 0.063967198 | 0.23885879 | 0.611629108 | 0.88325615 |
| sp\|Q16281\|CNGA3_HUMAN | 1 | 3 | 7 | 4 | 0.741788 | 0.084581371 | 0.059909623 | -0.657206456 | -0.681878205 | -0.024671749 | 0.189613701 | 0.226113108 | 0.996669306 |
| sp\|Q14623\|IHH_HUMAN | 8 | 21 | 41 | 18 | 0.020469 | -0.081641004 | -0.037837203 | -0.102109696 | -0.058305895 | 0.043803801 | 0.478350436 | 0.844152656 | 0.883817783 |
| sp\|Q68CJ9\|CR3L3_HUMAN | 2 | 33 | 62 | 27 | -0.07432 | -0.057093329 | 0.055302678 | 0.017228903 | 0.12962491 | 0.112396007 | 0.981187051 | 0.479594651 | 0.496563681 |
| sp\|Q6W4X9\|MUC6_HUMAN | 5 | 4 | 6 | 3 | -0.265 | -0.020117823 | 1.682362428 | 0.244886032 | 1.947366283 | 1.702480251 | 0.937585886 | 0.10109789 | 0.123758606 |
| sp\|Q86T13\|CLC14_HUMAN | 3 | 40 | 76 | 33 | -0.07922 | -0.078881093 | 0.132504338 | 0.000343718 | 0.211729149 | 0.211385431 | 0.999974092 | 0.001732012 | 0.000354674 |
| sp\|Q96DG6\|CMBL_HUMAN | 8 | 26 | 48 | 22 | 0.010506 | 0.082076791 | 0.211400106 | 0.0715711 | 0.200894414 | 0.129323315 | 0.626494036 | 0.079877933 | 0.259667048 |
| sp\|Q8WVN6\|SCTM1_HUMAN | 5 | 36 | 69 | 30 | 0.000786 | 0.038543529 | 0.31798891 | 0.037757107 | 0.317202488 | 0.279445381 | 0.82862672 | 0.000223849 | 0.00023875 |
| sp\|Q16363\|LAMA4_HUMAN | 12 | 17 | 34 | 16 | 0.22044 | 0.304731179 | 0.472598911 | 0.084291428 | 0.25215916 | 0.167867732 | 0.745646616 | 0.156707837 | 0.332832462 |
| sp\|P49913\|CAMP_HUMAN | 8 | 40 | 76 | 33 | -0.45545 | -0.107857896 | 0.373705653 | 0.347590138 | 0.829153686 | 0.481563549 | 0.02256585 | 1.25E-06 | 0.00195513 |
| sp\|Q96K76\|UBP47_HUMAN | 34 | 4 | 7 | 3 | 0.043246 | -0.25074724 | 0.343472528 | -0.293993216 | 0.300226552 | 0.594219768 | 0.766054012 | 0.828137614 | 0.427787393 |
| sp\|Q8N423\|LIRB2_HUMAN | 2 | 40 | 76 | 33 | -0.21849 | -0.128814056 | 0.085558606 | 0.08968042 | 0.304053083 | 0.214372663 | 0.48973286 | 0.004524716 | 0.030798192 |
| sp\|Q9HD89\|RETN_HUMAN | 19 | 21 | 41 | 19 | -0.48657 | -0.310869865 | 0.069057466 | 0.17569724 | 0.555624571 | 0.379927331 | 0.664083427 | 0.059254352 | 0.173730961 |
| sp\|Q15366\|PCBP2_HUMAN | 10 | 30 | 55 | 23 | -0.32152 | -0.214997017 | 0.328747685 | 0.106522277 | 0.65026698 | 0.543744703 | 0.784825683 | 0.003525032 | 0.00700089 |
| sp\|Q8WUT4\|LRRN4_HUMAN | 18 | 40 | 76 | 33 | -0.04481 | 0.056317406 | 0.019522361 | 0.10113146 | 0.064336415 | -0.036795045 | 0.41489505 | 0.780896059 | 0.902042314 |
| sp\|P13646\|K1C13_HUMAN | 1 | 19 | 34 | 14 | -0.03468 | -0.089375096 | 0.042059056 | -0.054694155 | 0.076739997 | 0.131434152 | 0.976321895 | 0.969288886 | 0.893694571 |
| sp\|Q86SJ2\|AMGO2_HUMAN | 24 | 22 | 42 | 17 | 0.093171 | 0.078893395 | -0.08756238 | -0.014277304 | -0.180733078 | -0.166455774 | 0.965753568 | 0.030089384 | 0.023841118 |
| sp\|P30047\|GFRP_HUMAN | 27 | 11 | 20 | 9 | -0.10086 | -0.030377099 | -0.074474756 | 0.070484179 | 0.026386522 | -0.044097657 | 0.510260501 | 0.935177844 | 0.791757867 |
| sp\|Q13753\|LAMC2_HUMAN | 8 | 32 | 62 | 27 | -0.11971 | -0.011441116 | 0.508115502 | 0.108267456 | 0.627824075 | 0.519556618 | 0.633948558 | 6.94E-05 | 0.000199452 |
| sp\|O75144\|ICOSL_HUMAN | 1 | 40 | 76 | 33 | 0.302181 | 0.218317797 | 0.011345192 | -0.083863693 | -0.290836298 | -0.206972605 | 0.186539465 | 3.51E-06 | 0.000223957 |
| sp\|P07902\|GALT_HUMAN | 16 | 37 | 69 | 29 | -0.06345 | -0.145397386 | 0.122766206 | -0.081952327 | 0.186211265 | 0.268163592 | 0.647311232 | 0.223659422 | 0.02229789 |
| sp\|Q8N0Y7\|PGAM4_HUMAN | 2 | 40 | 76 | 33 | -0.11231 | -0.035230577 | 0.339457469 | 0.077080957 | 0.451769003 | 0.374688046 | 0.653371193 | 9.66E-05 | 0.000282174 |
| sp\|Q13596\|SNX1_HUMAN | 12 | 13 | 27 | 12 | -0.02794 | 0.017094045 | 0.226879433 | 0.045037915 | 0.254823303 | 0.209785388 | 0.915289199 | 0.145555546 | 0.174371644 |
| sp\|P05981\|HEPS_HUMAN | 14 | 40 | 76 | 33 | -0.10087 | -0.066331376 | 0.194620373 | 0.034541281 | 0.295493031 | 0.26095175 | 0.870754108 | 0.001432041 | 0.001500959 |
| sp\|Q96PB7\|NOE3_HUMAN | 3 | 8 | 14 | 6 | 0.142775 | 0.070061189 | 0.185651271 | -0.072713546 | 0.042876536 | 0.115590082 | 0.568241653 | 0.873809947 | 0.317153837 |
| sp\|P0DJD7\|PEPA4_HUMAN | 3 | 40 | 76 | 33 | 0.064133 | -0.018051938 | 0.348830505 | -0.082185267 | 0.284697176 | 0.366882443 | 0.608487781 | 0.018907211 | 0.000315899 |
| sp\|Q12974\|TP4A2_HUMAN | 23 | 18 | 35 | 16 | 0.112216 | 0.18585786 | 0.220295002 | 0.073641779 | 0.108078921 | 0.034437142 | 0.781820902 | 0.686021016 | 0.951319356 |
| sp\|Q96LA6\|FCRL1_HUMAN | 1 | 3 | 7 | 4 | -0.0078 | 0.035617124 | -0.294845923 | 0.043417115 | -0.287045932 | -0.330463047 | 0.935644496 | 0.13920998 | 0.034373228 |
| sp\|P55884\|EIF3B_HUMAN | 3 | 7 | 14 | 6 | -0.58886 | -0.321018557 | 0.212797123 | 0.267836507 | 0.801652187 | 0.53381568 | 0.834416658 | 0.340456487 | 0.530534494 |
| sp\|Q9P2B2\|FPRP_HUMAN | 1 | 19 | 34 | 15 | 0.206736 | 0.225277014 | 0.323156847 | 0.018541345 | 0.116421178 | 0.097879833 | 0.98141846 | 0.604217822 | 0.642294753 |
| sp\|P56202\|CATW_HUMAN | 4 | 29 | 55 | 24 | -0.17102 | -0.156052158 | -0.13657637 | 0.014964022 | 0.03443981 | 0.019475788 | 0.990994204 | 0.967412788 | 0.986606524 |
| sp\|P05165\|PCCA_HUMAN | 3 | 4 | 7 | 3 | -0.27149 | -0.098101584 | 0.952237323 | 0.173393113 | 1.22373202 | 1.050338908 | 0.889821274 | 0.051807733 | 0.064997189 |
| sp\|Q99952\|PTN18_HUMAN | 1 | 30 | 55 | 23 | -0.1609 | -0.152759798 | -0.198225972 | 0.008141472 | -0.037324702 | -0.045466174 | 0.999265257 | 0.989694091 | 0.981035628 |
| sp\|Q86UN3\|R4RL2_HUMAN | 2 | 40 | 76 | 33 | 0.150976 | 0.047089144 | 0.08859854 | -0.103886574 | -0.062377178 | 0.041509396 | 0.079779539 | 0.526502948 | 0.695894366 |
| sp\|Q8IZP7\|H6ST3_HUMAN | 4 | 7 | 14 | 6 | 0.447771 | 0.072357117 | -0.47073659 | -0.375413913 | -0.91850762 | -0.543093707 | 0.073662291 | 0.000253234 | 0.011109203 |
| sp\|Q8TF66\|LRC15_HUMAN | 22 | 37 | 69 | 30 | 0.099407 | 0.012201304 | -0.08268967 | -0.087205934 | -0.182096908 | -0.094890975 | 0.675088973 | 0.310927743 | 0.667764094 |
| sp\|Q9NVG8\|TBC13_HUMAN | 2 | 7 | 14 | 6 | 0.114514 | 0.025281737 | 0.508617173 | -0.089232226 | 0.394103209 | 0.483335435 | 0.914913349 | 0.318495752 | 0.117975569 |
| sp\|Q9BQ51\|PD1L2_HUMAN | 32 | 40 | 76 | 33 | 0.19442 | 0.195803711 | 0.174348375 | 0.001384115 | -0.020071221 | -0.021455336 | 0.999534547 | 0.934727821 | 0.906545027 |
| sp\|P04440\|DPB1_HUMAN | 32 | 37 | 69 | 30 | -0.59456 | -0.292558535 | -0.139068491 | 0.30199886 | 0.455488904 | 0.153490044 | 0.208702758 | 0.088168419 | 0.700343543 |
| sp\|O00487\|PSDE_HUMAN | 40 | 11 | 21 | 8 | -0.00898 | 0.096878931 | 0.152502619 | 0.105861404 | 0.161485091 | 0.055623688 | 0.653806743 | 0.532374854 | 0.909264143 |
| sp\|P38606\|VATA_HUMAN | 2 | 40 | 76 | 33 | -0.31354 | -0.279230833 | 0.04575507 | 0.034304466 | 0.359290369 | 0.324985903 | 0.950117047 | 0.024048198 | 0.020751916 |
| sp\|Q9HCM3\|K1549_HUMAN | 3 | 7 | 14 | 7 | -0.0426 | 0.027579797 | -0.183874593 | 0.070181342 | -0.141273048 | -0.21145439 | 0.640453461 | 0.27151477 | 0.029354936 |
| sp\|P13224\|GP1BB_HUMAN | 5 | 18 | 34 | 15 | -0.06048 | -0.038656628 | 0.118792671 | 0.021825825 | 0.179275123 | 0.157449298 | 0.9788376 | 0.373596366 | 0.380439714 |
| sp\|Q16842\|SIA4B_HUMAN | 34 | 26 | 48 | 22 | -0.03477 | -0.104582569 | 0.037371839 | -0.069811761 | 0.072142626 | 0.141954408 | 0.703531013 | 0.766690316 | 0.277182674 |
| sp\|P54753\|EPHB3_HUMAN | 11 | 19 | 35 | 15 | -0.06715 | -0.112433046 | 0.124665441 | -0.045279072 | 0.191819415 | 0.237098487 | 0.865711064 | 0.180840859 | 0.041417255 |
| sp\|Q96DN2\|VWCE_HUMAN | 45 | 40 | 76 | 33 | -0.35382 | -0.291919128 | 0.336531075 | 0.061896124 | 0.690346327 | 0.628450203 | 0.823630772 | 4.91E-07 | 2.44E-07 |
| sp\|O76076\|WISP2_HUMAN | 2 | 36 | 69 | 30 | -0.0051 | -0.031155202 | 0.155723946 | -0.026059729 | 0.160819419 | 0.186879148 | 0.912312006 | 0.094730771 | 0.018416893 |
| sp\|Q9NZV1\|CRIM1_HUMAN | 9 | 22 | 41 | 19 | -0.04542 | -0.002686423 | 0.133902324 | 0.04272882 | 0.179317567 | 0.136588748 | 0.847470862 | 0.133507079 | 0.223064346 |
| sp\|Q0VD83\|APOBR_HUMAN | 5 | 28 | 55 | 25 | -1.18151 | -0.708136404 | 0.479938337 | 0.473368921 | 1.661443662 | 1.188074741 | 0.031152977 | 3.93E-11 | 3.40E-08 |
| sp\|P16144\|ITB4_HUMAN | 15 | 14 | 28 | 12 | 0.309701 | 0.182003873 | 0.115746655 | -0.127696922 | -0.193954141 | -0.066257218 | 0.474814227 | 0.308022494 | 0.832875744 |
| sp\|Q6GTX8\|LAIR1_HUMAN | 3 | 17 | 35 | 16 | -0.42721 | -0.576775561 | 0.336258894 | -0.149561987 | 0.763472468 | 0.913034455 | 0.799047988 | 0.019732461 | 0.000859702 |
| sp\|Q53H76\|PLA1A_HUMAN | 6 | 22 | 41 | 17 | 0.269572 | 0.189169201 | 0.317427094 | -0.080402512 | 0.04785538 | 0.128257892 | 0.701943015 | 0.919102649 | 0.47196542 |
| sp\|Q9HC57\|WFDC1_HUMAN | 2 | 18 | 34 | 15 | -0.43638 | -0.290805399 | 0.72809109 | 0.145573815 | 1.164470304 | 1.018896489 | 0.684117777 | 1.71E-06 | 2.26E-06 |

| | | | | | | | | | | | | | |
|---|---|---|---|---|---|---|---|---|---|---|---|---|---|
| sp\|P01185\|NEU2_HUMAN | 3 | 14 | 28 | 13 | 0.10219 | 0.318899877 | 0.444288698 | 0.216709834 | 0.342098656 | 0.125388822 | 0.546350158 | 0.340896125 | 0.823380078 |
| sp\|Q75N90\|FBN3_HUMAN | 9 | 8 | 14 | 6 | -0.10949 | -0.086872667 | -0.095657388 | 0.022620848 | 0.013836127 | -0.008784721 | 0.964710993 | 0.990978159 | 0.995533772 |
| sp\|P21741\|MK_HUMAN | 7 | 8 | 14 | 6 | -0.89474 | -1.001933813 | 0.533441799 | -0.10718975 | 1.428185862 | 1.535375612 | 0.972750953 | 0.054600703 | 0.019587262 |
| sp\|P30050\|RL12_HUMAN | 14 | 3 | 7 | 4 | -0.42921 | -0.267485471 | 0.349162239 | 0.161726591 | 0.778374301 | 0.61664771 | 0.831919844 | 0.066260178 | 0.076757062 |
| sp\|P0C7H8\|KRA23_HUMAN | 5 | 8 | 13 | 6 | -0.29859 | 0.111376211 | -1.20463398 | 0.409966725 | -0.906043466 | -1.316010191 | 0.876874053 | 0.645268068 | 0.341370033 |
| sp\|Q14008\|CKAP5_HUMAN | 8 | 4 | 7 | 3 | 0.008765 | -0.337702256 | 0.250885385 | -0.346467168 | 0.242120473 | 0.588587641 | 0.63573368 | 0.857848528 | 0.360289212 |
| sp\|Q08188\|TGM3_HUMAN | 15 | 21 | 42 | 19 | -0.40565 | -0.205091753 | -0.508327211 | 0.200556767 | -0.102678691 | -0.303235458 | 0.721965391 | 0.940721994 | 0.500713677 |
| sp\|Q9NVD7\|PARVA_HUMAN | 1 | 40 | 76 | 33 | -0.07466 | -0.420977077 | -0.196927477 | -0.346319474 | -0.122269874 | 0.2240496 | 0.109973518 | 0.823722476 | 0.439098269 |
| sp\|Q00973\|B4GN1_HUMAN | 3 | 20 | 34 | 14 | -0.12468 | -0.084988308 | -0.098852632 | 0.039688616 | 0.025824292 | -0.013864324 | 0.787664711 | 0.935844088 | 0.977230479 |
| sp\|Q15389\|ANGP1_HUMAN | 3 | 11 | 21 | 9 | 0.020675 | 0.191706938 | -0.053012084 | 0.171032192 | -0.073686831 | -0.244719023 | 0.415502842 | 0.89189764 | 0.214756783 |
| sp\|P01877\|IGHA2_HUMAN | 1 | 11 | 21 | 10 | -0.11885 | -0.084375008 | 0.305774736 | 0.03447641 | 0.424626154 | 0.390149744 | 0.976921653 | 0.091063586 | 0.074239964 |
| sp\|Q9NRR1\|CYTL1_HUMAN | 5 | 8 | 13 | 5 | 0.418674 | -0.020367669 | -0.063226638 | -0.439041407 | -0.481900376 | -0.042858969 | 0.008843904 | 0.024304233 | 0.959676052 |
| sp\|Q7L591\|DOK3_HUMAN | 2 | 11 | 21 | 9 | -0.22208 | -0.021767803 | 0.34590695 | 0.200308385 | 0.567983138 | 0.367674753 | 0.79304069 | 0.289375613 | 0.510288721 |
| sp\|Q13057\|COASY_HUMAN | 5 | 11 | 21 | 9 | 0.031699 | 0.03829523 | 0.431780175 | 0.006596295 | 0.40008124 | 0.393484945 | 0.999439844 | 0.257539708 | 0.191069509 |
| sp\|P18084\|ITB5_HUMAN | 7 | 40 | 76 | 33 | 0.029873 | -0.103540711 | 0.143483514 | -0.133413526 | 0.113610699 | 0.247024225 | 0.418372837 | 0.645320707 | 0.076030395 |
| sp\|P28065\|PSB9_HUMAN | 22 | 40 | 76 | 33 | -0.40578 | -0.179911869 | 0.430456715 | 0.225863713 | 0.836232297 | 0.610368584 | 0.154524464 | 1.77E-07 | 1.72E-05 |
| sp\|O75629\|CREG1_HUMAN | 14 | 32 | 62 | 27 | 0.20137 | 0.025422694 | -0.284496096 | -0.175947197 | -0.485865987 | -0.30991879 | 0.227621316 | 0.000663105 | 0.018818366 |
| sp\|Q8IW45\|NNRD_HUMAN | 7 | 4 | 7 | 3 | 0.02764 | -0.043297448 | 0.23848494 | -0.070937053 | 0.210845335 | 0.281782388 | 0.825147761 | 0.349111863 | 0.124819855 |
| sp\|Q9GZM5\|YIPF3_HUMAN | 8 | 32 | 62 | 28 | -0.04632 | -0.072143425 | 0.246923512 | -0.02582706 | 0.293239877 | 0.319066938 | 0.970505526 | 0.070944653 | 0.018624408 |
| sp\|Q9C0K3\|ARP3C_HUMAN | 12 | 25 | 48 | 20 | 0.007735 | -0.117330613 | 0.614319989 | -0.125065553 | 0.606585049 | 0.731650602 | 0.79880086 | 0.033085654 | 0.002316153 |
| sp\|Q16819\|MEP1A_HUMAN | 11 | 21 | 41 | 18 | -0.00023 | -0.135070018 | 0.16915014 | -0.13483615 | 0.169384008 | 0.304220158 | 0.47875164 | 0.444740314 | 0.039155678 |
| sp\|P10153\|RNAS2_HUMAN | 9 | 40 | 76 | 33 | -0.00716 | 0.008709582 | 0.093885138 | 0.015864845 | 0.101040401 | 0.085175556 | 0.953388942 | 0.267674093 | 0.303202565 |
| sp\|O14579\|COPE_HUMAN | 1 | 8 | 14 | 6 | -0.29112 | -0.478905861 | -0.247386427 | -0.187783387 | 0.043736046 | 0.231519433 | 0.737116312 | 0.988775202 | 0.683032994 |
| sp\|Q13098\|CSN1_HUMAN | 3 | 8 | 14 | 6 | 0.011288 | 0.315832865 | 0.32584834 | 0.304545173 | 0.314560648 | 0.010015475 | 0.110064126 | 0.196565661 | 0.997831786 |
| sp\|P0DP57\|SLUR2_HUMAN | 14 | 28 | 55 | 24 | 0.071256 | 0.007373791 | 0.327796373 | -0.0638826 | 0.256539982 | 0.320422582 | 0.741134257 | 0.039283283 | 0.001869457 |
| sp\|Q8IY63\|AMOL1_HUMAN | 3 | 29 | 56 | 23 | -0.15126 | -0.107659315 | -0.013485694 | 0.043598272 | 0.137771892 | 0.09417362 | 0.880968038 | 0.431034268 | 0.605283094 |
| sp\|Q7Z304\|MAMC2_HUMAN | 2 | 11 | 20 | 9 | -0.25373 | -0.228505958 | 0.382529668 | 0.025228438 | 0.636264064 | 0.611035626 | 0.992946622 | 0.056475175 | 0.037549737 |
| sp\|Q9UGM3\|DMBT1_HUMAN | 3 | 36 | 69 | 30 | -0.34827 | -0.320656662 | 0.45327968 | 0.027617519 | 0.80155386 | 0.773936341 | 0.976577541 | 5.20E-06 | 6.72E-07 |
| sp\|P0DMQ5\|INAM2_HUMAN | 2 | 6 | 14 | 7 | -0.13139 | -0.155000746 | 0.359297022 | -0.023608871 | 0.490688897 | 0.514297768 | 0.991629018 | 0.082743956 | 0.023909724 |
| sp\|P15509\|CSF2R_HUMAN | 7 | 25 | 49 | 22 | 0.02866 | 0.049745802 | 0.021785916 | 0.021086195 | -0.006873692 | -0.027959887 | 0.950071874 | 0.996157361 | 0.920759547 |
| sp\|Q8N729\|NPW_HUMAN | 9 | 23 | 41 | 18 | 0.225123 | 0.046234917 | 0.437939389 | -0.178887964 | 0.212816509 | 0.391704473 | 0.18157218 | 0.190878395 | 0.001593747 |
| sp\|Q9H773\|DCTP1_HUMAN | 50 | 14 | 28 | 12 | -0.4892 | -0.231797196 | 0.244303486 | 0.257405116 | 0.733505798 | 0.476100682 | 0.309136772 | 0.002696141 | 0.032383158 |
| sp\|O00505\|IMA4_HUMAN | 15 | 15 | 28 | 12 | 0.058754 | 0.002069102 | 0.220010451 | -0.056684887 | 0.161256462 | 0.217941348 | 0.925421457 | 0.653973922 | 0.38086773 |
| sp\|P01308\|INS_HUMAN | 9 | 4 | 7 | 3 | -0.13146 | 0.029723776 | -0.087373962 | 0.161180587 | 0.04408285 | -0.117097738 | 0.750498489 | 0.985268937 | 0.880813054 |
| sp\|Q8WTU2\|SRB4D_HUMAN | 3 | 12 | 20 | 8 | 0.148187 | 0.16260475 | -0.045313329 | 0.014417251 | -0.193500828 | -0.207918079 | 0.985972061 | 0.211637538 | 0.122610769 |
| sp\|Q68BL7\|OLM2A_HUMAN | 19 | 40 | 75 | 33 | 0.108881 | 0.032327065 | 0.411675007 | -0.076553884 | 0.302794059 | 0.379347943 | 0.824338452 | 0.128231058 | 0.01822628 |
| sp\|P29965\|CD40L_HUMAN | 2 | 12 | 21 | 9 | -0.01862 | 0.059349915 | 0.194016415 | 0.077965125 | 0.212631625 | 0.1346665 | 0.858914879 | 0.473076455 | 0.689420415 |
| sp\|P10636\|TAU_HUMAN | 4 | 4 | 6 | 3 | 0.514625 | 0.342990505 | 1.352312137 | -0.171634349 | 0.837687282 | 1.009321632 | 0.973544964 | 0.64601028 | 0.487568275 |
| sp\|P43686\|PRS6B_HUMAN | 4 | 4 | 7 | 3 | -0.18693 | -0.137227641 | 0.536682916 | 0.049706316 | 0.723616874 | 0.673910558 | 0.939541651 | 0.005160876 | 0.004196803 |
| sp\|Q16674\|MIA_HUMAN | 32 | 14 | 28 | 12 | -0.00024 | 0.041009159 | 0.100330488 | 0.04125088 | 0.10057221 | 0.059321329 | 0.854376915 | 0.527111952 | 0.746770853 |
| sp\|O15247\|CLIC2_HUMAN | 49 | 8 | 14 | 6 | -0.02805 | 0.048542645 | 0.203458855 | 0.07659703 | 0.23151324 | 0.15491621 | 0.819521746 | 0.30944646 | 0.517791902 |
| sp\|Q9Y2G1\|MYRF_HUMAN | 2 | 40 | 76 | 33 | -0.06723 | -0.050383413 | 0.313705535 | 0.016845485 | 0.380934432 | 0.364088947 | 0.956302586 | 1.00E-06 | 1.39E-07 |
| sp\|O75309\|CAD16_HUMAN | 3 | 7 | 14 | 6 | -0.11783 | -0.184471969 | 0.663761347 | -0.066645631 | 0.781587684 | 0.848233315 | 0.901348239 | 0.000798417 | 6.32E-05 |
| sp\|Q9Y336\|SIGL9_HUMAN | 29 | 30 | 56 | 24 | -0.06331 | 0.117173478 | 0.214005741 | 0.180483546 | 0.277315809 | 0.096832263 | 0.01342945 | 0.001180809 | 0.329082712 |
| sp\|P62857\|RS28_HUMAN | 5 | 14 | 27 | 12 | -0.22584 | -0.166556928 | 0.942769999 | 0.059287894 | 1.168614821 | 1.109326928 | 0.980217487 | 0.007799382 | 0.003946731 |
| sp\|Q969M7\|UBE2F_HUMAN | 13 | 17 | 35 | 15 | 0.049818 | 0.062506652 | 0.232396893 | 0.01268859 | 0.182578832 | 0.169890241 | 0.993665233 | 0.406361003 | 0.358835944 |
| sp\|Q8WUW1\|BRK1_HUMAN | 90 | 17 | 35 | 16 | -0.10985 | 0.045362463 | 0.229072748 | 0.15521105 | 0.338921336 | 0.183710285 | 0.291687178 | 0.018037315 | 0.193313195 |
| sp\|Q9UL54\|TAOK2_HUMAN | 1 | 8 | 14 | 6 | -0.70089 | -0.489286034 | 0.628287886 | 0.211600618 | 1.329174538 | 1.117573921 | 0.886845653 | 0.058994459 | 0.083028344 |
| sp\|Q9UHY1\|NRBP_HUMAN | 2 | 7 | 14 | 5 | -0.27122 | -0.239440698 | 0.457981725 | 0.031780283 | 0.729202706 | 0.697422423 | 0.993306666 | 0.134219709 | 0.101329368 |
| sp\|Q8N386\|LRC25_HUMAN | 6 | 4 | 7 | 3 | -0.24099 | -0.389319783 | -0.57913634 | -0.148326244 | -0.3381428 | -0.189816557 | 0.414639739 | 0.074273899 | 0.314191283 |
| sp\|Q9P2X0\|DPM3_HUMAN | 1 | 40 | 76 | 33 | -0.13276 | 0.073911377 | -0.049254789 | 0.206672807 | 0.08350664 | -0.123166167 | 0.019765489 | 0.63246645 | 0.284367984 |
| sp\|Q92688\|AN32B_HUMAN | 1 | 7 | 14 | 6 | -0.10537 | -0.097321185 | 0.446414696 | 0.00805082 | 0.551786702 | 0.543735882 | 0.999303181 | 0.127182223 | 0.078417159 |
| sp\|Q9ULA0\|DNPEP_HUMAN | 1 | 10 | 20 | 10 | -0.32503 | 0.025862798 | 0.352214135 | 0.350894316 | 0.677245653 | 0.326351337 | 0.214208238 | 0.018468949 | 0.261568881 |
| sp\|Q5XPI4\|RN123_HUMAN | 2 | 8 | 14 | 6 | 0.548053 | 0.71947609 | 0.818717173 | 0.171423211 | 0.270664294 | 0.099241083 | 0.811290488 | 0.70542241 | 0.943450793 |
| sp\|O00587\|MFNG_HUMAN | 2 | 23 | 41 | 17 | 0.053212 | 0.112763313 | 0.178892037 | 0.059551584 | 0.125680308 | 0.066128724 | 0.311124196 | 0.035786508 | 0.309101096 |
| sp\|Q9Y6N8\|CAD10_HUMAN | 2 | 36 | 69 | 30 | 0.054489 | -0.006070471 | 0.142114733 | -0.06055937 | 0.087625834 | 0.148185204 | 0.611401483 | 0.491284061 | 0.07850978 |
| sp\|Q6UWN8\|ISK6_HUMAN | 9 | 11 | 21 | 9 | 0.182248 | 0.126253636 | 0.106107853 | -0.055994328 | -0.076140112 | -0.020145784 | 0.945499878 | 0.931441135 | 0.993679555 |
| sp\|Q8TB24\|RIN3_HUMAN | 1 | 7 | 14 | 5 | -0.133 | -0.062234385 | 0.603942375 | 0.070769073 | 0.736945833 | 0.666176761 | 0.962804958 | 0.099755728 | 0.093325764 |
| sp\|Q8WWA1\|TMM40_HUMAN | 3 | 4 | 7 | 3 | 0.637924 | 0.206091392 | -0.195332979 | -0.43183266 | -0.833257031 | -0.401424371 | 0.50384779 | 0.206823394 | 0.607975345 |
| sp\|P15924\|DESP_HUMAN | 4 | 7 | 13 | 5 | -0.18266 | 0.333112734 | 0.008269933 | 0.515772792 | 0.190929991 | -0.324842801 | 0.68076379 | 0.966212266 | 0.884577507 |
| sp\|O95989\|NUDT3_HUMAN | 49 | 11 | 21 | 10 | 0.134219 | 0.139526159 | 0.251998945 | 0.005306741 | 0.117779527 | 0.112472786 | 0.999069721 | 0.719324061 | 0.678511109 |
| sp\|P17661\|DESM_HUMAN | 5 | 4 | 6 | 3 | -0.02009 | 0.056793736 | 0.821156991 | 0.076887981 | 0.841251236 | 0.764363255 | 0.986979081 | 0.36469423 | 0.377180795 |
| sp\|P04439\|HLAA_HUMAN | 3 | 40 | 76 | 33 | -0.23279 | -0.061325561 | 0.081552495 | 0.171468505 | 0.314346561 | 0.142878056 | 0.341005216 | 0.085510506 | 0.517498587 |
| sp\|Q30154\|DRB5_HUMAN | 3 | 14 | 27 | 13 | 0.046239 | 0.05920479 | 0.212494505 | 0.012965474 | 0.16625519 | 0.153289715 | 0.990995462 | 0.346154254 | 0.310055614 |
| sp\|O94766\|B3GA3_HUMAN | 3 | 36 | 69 | 30 | -0.173 | -0.091390263 | 0.263888169 | 0.081610416 | 0.436888848 | 0.355278433 | 0.509065363 | 6.73E-06 | 3.61E-05 |
| sp\|O94910\|AGRL1_HUMAN | 2 | 7 | 13 | 6 | 0.249384 | 0.13057617 | 0.534561983 | -0.118807853 | 0.28517796 | 0.403985813 | 0.427458224 | 0.044223965 | 0.001252578 |
| sp\|P38570\|ITAE_HUMAN | 2 | 4 | 7 | 3 | 0.608814 | 0.309740997 | 0.466667582 | -0.29907294 | -0.142146356 | 0.156926584 | 0.226431662 | 0.775201349 | 0.68676068 |
| sp\|Q9H488\|OFUT1_HUMAN | 33 | 7 | 14 | 6 | -0.04629 | 0.122493596 | 0.268314498 | 0.168782565 | 0.314603467 | 0.145820902 | 0.481321641 | 0.185718749 | 0.608635908 |
| sp\|O43598\|DNPH1_HUMAN | 16 | 23 | 41 | 18 | -0.28224 | -0.027755106 | 0.153395563 | 0.25448946 | 0.43564013 | 0.181150669 | 0.093919302 | 0.010207107 | 0.353591465 |
| sp\|Q9Y286\|SIGL7_HUMAN | 5 | 36 | 69 | 31 | 0.002241 | -0.052094483 | 0.155509279 | -0.054335725 | 0.153268036 | 0.207603761 | 0.706840717 | 0.148090213 | 0.012512743 |
| sp\|O75828\|CBR3_HUMAN | 21 | 30 | 55 | 24 | -0.02169 | 0.071187286 | 0.218010685 | 0.092880597 | 0.239703995 | 0.146823399 | 0.315153683 | 0.006554849 | 0.08695288 |
| sp\|Q8TCA0\|LRC20_HUMAN | 6 | 4 | 6 | 3 | -0.14387 | 0.086017865 | 0.692254532 | 0.229885803 | 0.83612247 | 0.606236667 | 0.965750984 | 0.726444637 | 0.819937967 |
| sp\|Q16378\|PROL4_HUMAN | 7 | 12 | 20 | 8 | 0.256967 | 0.24282339 | 0.202411758 | -0.014143433 | -0.054555064 | -0.040411631 | 0.984510547 | 0.862397601 | 0.907686144 |
| sp\|Q5VYK3\|ECM29_HUMAN | 11 | 8 | 13 | 6 | 0.093941 | 0.067718645 | -0.0473479 | -0.026222525 | -0.141289071 | -0.115066545 | 0.983640325 | 0.721195703 | 0.770774207 |
| sp\|A0A075B6R9\|KVD24_HUMAN | 2 | 8 | 14 | 5 | -0.01792 | 0.019342329 | -0.103519171 | 0.037259485 | -0.085602015 | -0.1228615 | 0.954097427 | 0.861392921 | 0.694606923 |
| sp\|A0A075B6H9\|LV469_HUMAN | 1 | 22 | 41 | 19 | -0.05403 | -0.008457314 | 0.108775802 | 0.045575127 | 0.162808243 | 0.117233116 | 0.897622596 | 0.379978691 | 0.526050049 |
| sp\|P18669\|PGAM1_HUMAN | 1 | 40 | 76 | 33 | -0.12824 | -0.067445481 | 0.264727395 | 0.060797254 | 0.392970129 | 0.332172875 | 0.783711082 | 0.001376449 | 0.002423084 |
| sp\|Q96J84\|KIRR1_HUMAN | 4 | 4 | 6 | 3 | 0.257123 | 0.201734535 | 0.582340557 | -0.055388446 | 0.325217576 | 0.380606022 | 0.837554206 | 0.042526164 | 0.012462302 |
| sp\|O95154\|ARK73_HUMAN | 16 | 22 | 41 | 19 | -0.26361 | -0.059707573 | 0.069897291 | 0.203899061 | 0.333503925 | 0.129604864 | 0.392446477 | 0.172376836 | 0.70748547 |
| sp\|Q8NHP1\|ARK74_HUMAN | 1 | 37 | 69 | 30 | -0.32882 | -0.165337218 | 0.050788907 | 0.163479334 | 0.379605459 | 0.216126125 | 0.441887135 | 0.051918123 | 0.291295351 |
| sp\|P01732\|CD8A_HUMAN | 22 | 26 | 48 | 21 | 0.036394 | 0.20390649 | 0.258552238 | 0.167512311 | 0.222158059 | 0.054645749 | 0.032751265 | 0.016533749 | 0.719261075 |
| sp\|Q9H4A3\|WNK1_HUMAN | 14 | 11 | 21 | 8 | -0.01147 | 0.169531286 | 0.508619222 | 0.181003217 | 0.520091153 | 0.339087936 | 0.602443628 | 0.080271119 | 0.249369486 |
| sp\|Q8WUD1\|RAB2B_HUMAN | 3 | 10 | 21 | 10 | 0.089838 | 0.106660666 | 0.352471897 | 0.016822528 | 0.262633759 | 0.245811231 | 0.994551556 | 0.384816219 | 0.32383565 |
| sp\|Q9Y303\|NAGA_HUMAN | 4 | 12 | 20 | 8 | 0.171923 | -0.055700922 | -0.242330983 | -0.227624201 | -0.414254262 | -0.186630061 | 0.149125107 | 0.022275008 | 0.367075012 |
| sp\|O95757\|HS74L_HUMAN | 8 | 40 | 76 | 33 | -0.34309 | -0.16469427 | 0.280388961 | 0.17839611 | 0.623479341 | 0.445083231 | 0.151136135 | 6.92E-07 | 6.90E-05 |
| sp\|P12838\|DEF4_HUMAN | 7 | 33 | 62 | 27 | -0.22795 | -0.213423045 | -0.045854974 | 0.014526633 | 0.182094704 | 0.167568071 | 0.995509134 | 0.615822511 | 0.594664594 |
| sp\|Q96JK4\|HIPL1_HUMAN | 2 | 7 | 13 | 6 | -0.24864 | -0.188946698 | 0.315045977 | 0.059689681 | 0.563682356 | 0.503992676 | 0.928620613 | 0.020193535 | 0.019174506 |
| sp\|P02771\|FETA_HUMAN | 7 | 4 | 6 | 3 | -0.22467 | -0.278486642 | 0.193039868 | -0.053816006 | 0.417710504 | 0.47152651 | 0.984715128 | 0.536965542 | 0.407504775 |
| sp\|Q14011\|CIRBP_HUMAN | 3 | 11 | 20 | 8 | 0.530509 | 0.413137385 | 1.225468837 | -0.117371839 | 0.694959614 | 0.812331452 | 0.915828789 | 0.149654729 | 0.045766648 |
| sp\|A8K2U0\|A2ML1_HUMAN | 98 | 4 | 7 | 3 | 0.328635 | -0.099442573 | -0.348226109 | -0.42807733 | -0.676860866 | -0.248783536 | 0.197016493 | 0.081618009 | 0.603620528 |
| sp\|Q9Y227\|ENTP4_HUMAN | 4 | 7 | 13 | 6 | 0.021926 | 0.091520607 | 0.0845179 | 0.069594324 | 0.062591617 | -0.007002707 | 0.869674863 | 0.922722299 | 0.998716798 |
| sp\|O75390\|CISY_HUMAN | 7 | 15 | 27 | 12 | -0.07758 | 0.177996888 | 0.468614585 | 0.255573386 | 0.546191083 | 0.290617697 | 0.162675748 | 0.00502917 | 0.133488443 |
| sp\|Q96QR8\|PURB_HUMAN | 1 | 3 | 7 | 4 | -0.321 | -0.157378155 | 0.141927313 | 0.163619872 | 0.462925341 | 0.299305468 | 0.701313553 | 0.138261692 | 0.269390741 |
| sp\|Q6ZVN8\|RGMC_HUMAN | 2 | 34 | 62 | 26 | -0.23364 | -0.103247388 | 0.121646429 | 0.130396917 | 0.355290733 | 0.224893817 | 0.220779721 | 0.00086025 | 0.02587274 |
| sp\|P17858\|PFKAL_HUMAN | 2 | 21 | 42 | 19 | -0.25825 | -0.280228638 | 0.235676262 | -0.021974279 | 0.493930622 | 0.5159049 | 0.993231654 | 0.095112115 | 0.0363512 |
| sp\|Q9ULF5\|S39AA_HUMAN | 2 | 12 | 21 | 8 | -0.04193 | -0.039532181 | -0.064631446 | 0.002400588 | -0.022698677 | -0.025099265 | 0.999690868 | 0.982783622 | 0.974704808 |
| sp\|P80217\|IN35_HUMAN | 71 | 29 | 55 | 24 | -0.08244 | 0.116237331 | 0.3181817 | 0.198681663 | 0.400626031 | 0.201944368 | 0.050687257 | 0.000375187 | 0.065861742 |
| sp\|Q9BRK3\|MXRA8_HUMAN | 4 | 40 | 76 | 33 | 0.037457 | 0.002280589 | 0.084600685 | -0.035176107 | 0.047143989 | 0.082320096 | 0.751508925 | 0.702022908 | 0.2571448 |
| sp\|P02458\|CO2A1_HUMAN | 8 | 40 | 76 | 33 | -1.22172 | -1.313490613 | -1.155888087 | -0.091771035 | 0.065831491 | 0.157602526 | 0.859152398 | 0.947473241 | 0.675645222 |
| sp\|P12532\|KCRU_HUMAN | 1 | 4 | 7 | 3 | 0.331502 | 0.219840086 | -0.240904668 | -0.111661491 | -0.572406245 | -0.460744754 | 0.916548812 | 0.255872421 | 0.329872891 |
| sp\|Q5IJ48\|CRUM2_HUMAN | 2 | 4 | 7 | 2 | 0.227397 | 0.206919345 | 0.02427279 | -0.020477288 | -0.203123843 | -0.182646555 | 0.988166619 | 0.560994736 | 0.578649659 |
| sp\|Q13477\|MADCA_HUMAN | 2 | 40 | 76 | 33 | -0.01343 | 0.019715659 | 0.074064814 | 0.033141091 | 0.087490246 | 0.054349155 | 0.876943018 | 0.533605407 | 0.73383588 |
| sp\|O75888\|TNF13_HUMAN | 6 | 36 | 69 | 30 | -0.18231 | -0.151132026 | -0.011439902 | 0.031180639 | 0.170872762 | 0.139692124 | 0.954052788 | 0.380260508 | 0.437346216 |
| sp\|O15273\|TELT_HUMAN | 4 | 4 | 6 | 3 | 0.001106 | -0.128932374 | 0.915607204 | -0.13003852 | 0.914501058 | 1.044539578 | 0.959453634 | 0.277526146 | 0.159274967 |
| sp\|Q9Y5X1\|SNX9_HUMAN | 5 | 4 | 6 | 3 | 0.032335 | 0.072261249 | 0.548123456 | 0.039925931 | 0.515788138 | 0.475862207 | 0.987419645 | 0.268623795 | 0.270787137 |

| | | | | | | | | | | | | | |
|---|---|---|---|---|---|---|---|---|---|---|---|---|---|
| sp\|P02538\|K2C6A_HUMAN | 7 | 18 | 34 | 13 | -1.79909 | -1.411949932 | -1.56589522 | 0.387135652 | 0.233190364 | -0.153945288 | 0.788048681 | 0.94588715 | 0.970229784 |
| sp\|O95292\|VAPB_HUMAN | 9 | 10 | 20 | 9 | -0.19236 | -0.136439759 | 0.341335952 | 0.055917537 | 0.533693248 | 0.477775712 | 0.93984357 | 0.027566904 | 0.023467936 |
| sp\|Q6ISS4\|LAIR2_HUMAN | 5 | 10 | 21 | 9 | -0.963 | -0.869393207 | 0.442060792 | 0.093606522 | 1.405060521 | 1.311453999 | 0.956602934 | 0.002893884 | 0.001340948 |
| sp\|Q9P0G3\|KLK14_HUMAN | 1 | 11 | 21 | 8 | 0.082968 | 0.051885391 | -0.190512981 | -0.031082788 | -0.27348116 | -0.242398372 | 0.948769193 | 0.088293937 | 0.091831701 |
| sp\|Q5T749\|KPRP_HUMAN | 15 | 26 | 48 | 21 | -0.53344 | -0.563311131 | -0.849578719 | -0.029875399 | -0.316142986 | -0.286267588 | 0.995250609 | 0.694009706 | 0.686217489 |
| sp\|O14657\|TOR1B_HUMAN | 9 | 11 | 20 | 9 | 0.095921 | 0.10496391 | 0.255774877 | 0.009042776 | 0.159853743 | 0.150810967 | 0.994185268 | 0.294049353 | 0.256577063 |
| sp\|Q86YZ3\|HORN_HUMAN | 3 | 40 | 76 | 33 | 0.925392 | 0.393523936 | 0.553420531 | -0.531868025 | -0.37197143 | 0.159896595 | 0.024115742 | 0.275555505 | 0.736204318 |
| sp\|Q8WX77\|IBPL1_HUMAN | 3 | 4 | 6 | 3 | 0.443231 | 0.750375662 | 1.704797438 | 0.307144829 | 1.261566605 | 0.954421776 | 0.886796952 | 0.27805259 | 0.410231801 |
| sp\|P07311\|ACYP1_HUMAN | 7 | 18 | 35 | 16 | -0.02005 | 0.154894713 | 0.167663444 | 0.174943427 | 0.187712159 | 0.012768731 | 0.180719197 | 0.243773144 | 0.991342908 |
| sp\|O43405\|COCH_HUMAN | 4 | 37 | 69 | 29 | -0.02037 | 0.024068154 | 0.153297965 | 0.044436424 | 0.173666236 | 0.129229812 | 0.823815202 | 0.141024311 | 0.253798899 |
| sp\|Q12931\|TRAP1_HUMAN | 10 | 21 | 41 | 19 | -0.2535 | -0.110941224 | 0.173045009 | 0.142559074 | 0.426545307 | 0.283986233 | 0.74395934 | 0.156685194 | 0.338561599 |
| sp\|O00170\|AIP_HUMAN | 1 | 3 | 7 | 4 | 0.350459 | 0.339443142 | 0.533492939 | -0.011015386 | 0.183034411 | 0.194049797 | 0.999389695 | 0.872990367 | 0.798698163 |
| sp\|O94788\|AL1A2_HUMAN | 21 | 4 | 6 | 3 | -0.49523 | -0.557538128 | -0.241082395 | -0.062310392 | 0.25414534 | 0.316455732 | 0.991551938 | 0.904971081 | 0.836001897 |
| sp\|P07204\|TRBM_HUMAN | 6 | 29 | 56 | 24 | 0.010542 | -0.013119032 | 0.317638067 | -0.023660578 | 0.307096521 | 0.330757099 | 0.944591148 | 0.002222654 | 0.00015398 |
| sp\|Q9Y2A9\|B3GN3_HUMAN | 7 | 4 | 6 | 3 | 0.292781 | 0.104528883 | 0.473019979 | -0.188252606 | 0.180238491 | 0.368491096 | 0.131118983 | 0.241347526 | 0.008553605 |
| sp\|P09466\|PAEP_HUMAN | 14 | 20 | 35 | 15 | 0.185691 | 0.037240627 | -0.436678275 | -0.148450038 | -0.622368939 | -0.473918902 | 0.89756797 | 0.285911782 | 0.408482785 |
| sp\|Q9Y2B0\|CNPY2_HUMAN | 111 | 12 | 21 | 9 | -0.15615 | -0.20579737 | 0.078669956 | -0.049644047 | 0.234823278 | 0.284467326 | 0.949431029 | 0.465064638 | 0.258559771 |
| sp\|P09327\|VILI_HUMAN | 3 | 23 | 42 | 17 | 0.219819 | 0.149380941 | -0.193618355 | -0.07043852 | -0.413437816 | -0.342999296 | 0.780059442 | 0.005531138 | 0.011344021 |
| sp\|Q66K79\|CBPZ_HUMAN | 5 | 4 | 7 | 3 | -0.02658 | 0.222491844 | -0.517699377 | 0.249067087 | -0.491124135 | -0.740191222 | 0.822702625 | 0.608676781 | 0.277968043 |
| sp\|Q6P9A2\|GLT18_HUMAN | 5 | 22 | 41 | 18 | 0.077897 | 0.10652073 | 0.363503102 | 0.028623718 | 0.28560609 | 0.256982372 | 0.96980252 | 0.129802396 | 0.124060165 |
| sp\|P60891\|PRPS1_HUMAN | 1 | 4 | 7 | 3 | 0.133493 | 0.18479391 | 0.191290117 | 0.05130111 | 0.057797318 | 0.006496207 | 0.923599548 | 0.934247844 | 0.998941276 |
| sp\|P68371\|TBB4B_HUMAN | 30 | 25 | 48 | 22 | -0.23247 | -0.126848032 | 0.374367388 | 0.105618198 | 0.606833619 | 0.501215421 | 0.776504327 | 0.004027198 | 0.007506991 |
| sp\|Q9BRP8\|PYM1_HUMAN | 11 | 11 | 20 | 8 | 0.038316 | -0.019258894 | 0.293930619 | -0.057574493 | 0.255615019 | 0.313189513 | 0.893188901 | 0.247796991 | 0.082447111 |
| sp\|P49589\|SYCC_HUMAN | 1 | 4 | 7 | 3 | 0.098454 | 0.147101513 | 0.472519507 | 0.048647079 | 0.374065073 | 0.325417994 | 0.913408151 | 0.062543002 | 0.073413033 |
| sp\|P59190\|RAB15_HUMAN | 15 | 4 | 7 | 3 | -0.38738 | -0.259253469 | 0.861663595 | 0.1281309 | 1.249047963 | 1.120917064 | 0.94368313 | 0.059214215 | 0.060994534 |
| sp\|P36507\|MP2K2_HUMAN | 17 | 15 | 27 | 12 | 0.025095 | 0.037333102 | 0.305482208 | 0.012237835 | 0.280386942 | 0.268149107 | 0.995365291 | 0.197279448 | 0.158720406 |
| sp\|Q9H8W4\|PKHF2_HUMAN | 5 | 13 | 28 | 13 | -0.23358 | -0.104134938 | 0.279351414 | 0.129442393 | 0.512928745 | 0.383486352 | 0.895898989 | 0.29202055 | 0.388267017 |
| sp\|O60218\|AK1BA_HUMAN | 4 | 4 | 7 | 3 | -0.40919 | -0.565739598 | 0.379165959 | -0.156549755 | 0.788355802 | 0.944905557 | 0.940991129 | 0.385511994 | 0.206386523 |
| sp\|O75022\|LIRB3_HUMAN | 5 | 7 | 14 | 7 | -0.1394 | 0.031407475 | -0.015831297 | 0.170803003 | 0.123564231 | -0.047238772 | 0.726205804 | 0.881041917 | 0.975512964 |
| sp\|Q99627\|CSN8_HUMAN | 10 | 3 | 7 | 4 | 0.185207 | -0.109957825 | 0.202703075 | -0.295164436 | 0.017496464 | 0.3126609 | 0.768898888 | 0.999227366 | 0.701665439 |
| sp\|Q9NPF0\|CD320_HUMAN | 10 | 25 | 49 | 22 | -0.13695 | -0.012671605 | 0.348183335 | 0.124279357 | 0.485134297 | 0.360854939 | 0.387636568 | 0.000109466 | 0.001163158 |
| sp\|O60245\|PCDH7_HUMAN | 2 | 12 | 21 | 8 | 0.111567 | 0.172781634 | 0.00776711 | 0.061214271 | -0.103800253 | -0.165014524 | 0.721710683 | 0.557196841 | 0.178440021 |
| sp\|Q8IUC8\|GLT13_HUMAN | 4 | 24 | 48 | 22 | -0.15441 | -0.084460956 | -0.047364653 | 0.069946548 | 0.107042852 | 0.037096303 | 0.573221519 | 0.394626989 | 0.862020667 |
| sp\|A6NH11\|GLTD2_HUMAN | 11 | 18 | 34 | 15 | 0.036455 | 0.180591505 | 0.20450766 | 0.144136801 | 0.168052957 | 0.023916155 | 0.297645242 | 0.31768971 | 0.970258826 |
| sp\|O75354\|ENTP6_HUMAN | 2 | 11 | 21 | 10 | 0.010696 | -0.053321292 | -0.137667546 | -0.064017025 | -0.148363279 | -0.084346254 | 0.689459447 | 0.245214549 | 0.547905086 |
| sp\|Q53FT3\|HIKES_HUMAN | 4 | 18 | 34 | 16 | 0.146219 | 0.066197172 | 0.083299418 | -0.080021826 | -0.062919579 | 0.017102246 | 0.727228164 | 0.867439862 | 0.986562368 |
| sp\|P62195\|PRS8_HUMAN | 1 | 16 | 27 | 11 | -0.1859 | -0.004579837 | 0.370851319 | 0.181316188 | 0.556747344 | 0.375431156 | 0.391433073 | 0.005519881 | 0.050641587 |
| sp\|O60259\|KLK8_HUMAN | 30 | 7 | 14 | 6 | 0.166203 | -0.025980711 | -0.120286863 | -0.19218418 | -0.286490332 | -0.094306151 | 0.363656796 | 0.218655081 | 0.796724373 |
| sp\|P18564\|ITB6_HUMAN | 25 | 30 | 55 | 23 | 0.113204 | 0.056663273 | -0.023332902 | -0.056540873 | -0.136537048 | -0.079996175 | 0.603668083 | 0.144135712 | 0.431555934 |
| sp\|P08246\|ELNE_HUMAN | 2 | 36 | 69 | 30 | -0.30093 | -0.135344957 | -0.09637545 | 0.165581673 | 0.20455118 | 0.038969507 | 0.401830155 | 0.382178603 | 0.955924715 |
| sp\|P55160\|NCKPL_HUMAN | 27 | 4 | 7 | 3 | -0.40625 | 0.026384922 | 0.478759681 | 0.432635325 | 0.885010085 | 0.45237476 | 0.488799663 | 0.163299253 | 0.522156719 |
| sp\|Q5T2D2\|TRML2_HUMAN | 4 | 40 | 76 | 33 | -0.12214 | 0.09088487 | -0.033773448 | 0.21302934 | 0.088371023 | -0.124658317 | 5.57E-05 | 0.282451125 | 0.043297404 |
| sp\|P04085\|PDGFA_HUMAN | 7 | 4 | 7 | 3 | -0.19346 | 0.19118705 | 0.272359513 | 0.384648136 | 0.465820599 | 0.081172463 | 0.566240298 | 0.570008222 | 0.978219566 |
| sp\|Q96AB3\|ISOC2_HUMAN | 4 | 8 | 13 | 6 | 0.031563 | 0.148288459 | 0.343266516 | 0.116725717 | 0.311703774 | 0.194978057 | 0.923254057 | 0.677957326 | 0.832091607 |
| sp\|P38405\|GNAL_HUMAN | 3 | 10 | 21 | 10 | -0.44318 | -0.207799047 | 0.430252477 | 0.235385382 | 0.873436906 | 0.638051524 | 0.571137287 | 0.006852175 | 0.023760631 |
| sp\|P84074\|HPCA_HUMAN | 6 | 17 | 35 | 14 | -0.10717 | -0.109026611 | 0.266435807 | -0.001861141 | 0.373601276 | 0.375462417 | 0.999864279 | 0.032324602 | 0.011922332 |
| sp\|P54296\|MYOM2_HUMAN | 8 | 37 | 69 | 30 | 0.08167 | -0.019037153 | -0.077416768 | -0.100707464 | -0.159087078 | -0.058379614 | 0.557472977 | 0.36848091 | 0.8425909 |
| sp\|Q86Y38\|XYLT1_HUMAN | 8 | 7 | 14 | 5 | 0.237067 | 0.326367498 | 0.056016052 | 0.089300469 | -0.181050977 | -0.270351446 | 0.886350437 | 0.735507228 | 0.430258999 |
| sp\|Q96C19\|EFHD2_HUMAN | 2 | 8 | 14 | 6 | 0.085231 | 0.185185781 | 0.052237043 | 0.099954374 | -0.032994363 | -0.132948737 | 0.810048151 | 0.984528322 | 0.736256246 |
| sp\|P06870\|KLK1_HUMAN | 3 | 15 | 28 | 12 | 0.149206 | -0.106708515 | -0.063023956 | -0.255914782 | -0.212230224 | 0.043684558 | 0.258658637 | 0.524566773 | 0.96567052 |
| sp\|Q9NR16\|C163B_HUMAN | 1 | 4 | 6 | 3 | -0.05901 | 0.032590401 | 0.6476174 | 0.091604558 | 0.706631558 | 0.615026999 | 0.952421581 | 0.177025406 | 0.21031667 |
| sp\|P57721\|PCBP3_HUMAN | 15 | 26 | 49 | 20 | -0.34798 | -0.274006569 | 0.169018161 | 0.073971825 | 0.516996555 | 0.44302473 | 0.929286749 | 0.099492203 | 0.118319601 |
| sp\|Q03405\|UPAR_HUMAN | 4 | 26 | 48 | 21 | -0.32279 | -0.235578572 | 0.161054179 | 0.087208485 | 0.483841237 | 0.396632752 | 0.721171933 | 0.001725457 | 0.004318885 |
| sp\|P62820\|RAB1A_HUMAN | 50 | 19 | 35 | 14 | -0.10816 | -0.032430715 | 0.148700958 | 0.075726149 | 0.256857821 | 0.181131673 | 0.792922214 | 0.182881289 | 0.346126328 |
| sp\|Q8IU80\|TMPS6_HUMAN | 11 | 40 | 76 | 33 | 0.168482 | 0.153926026 | -0.037344765 | -0.014555755 | -0.205826546 | -0.191270791 | 0.907342299 | 6.42E-06 | 2.23E-06 |
| sp\|Q9GZZ9\|UBA5_HUMAN | 3 | 4 | 7 | 3 | -1.83769 | -2.12764023 | -0.366193375 | -0.289948225 | 1.47149863 | 1.761446855 | 0.949158253 | 0.432101801 | 0.247795841 |
| sp\|O94973\|AP2A2_HUMAN | 10 | 15 | 27 | 11 | -0.16326 | -0.322078783 | 0.128553086 | -0.158818835 | 0.291813034 | 0.450631869 | 0.793100496 | 0.59953826 | 0.23023313 |
| sp\|P61026\|RAB10_HUMAN | 1 | 26 | 48 | 22 | -0.1584 | -0.147659455 | 0.266290647 | 0.010738237 | 0.424688339 | 0.413950102 | 0.993907178 | 0.002049472 | 0.000654773 |
| sp\|P48735\|IDHP_HUMAN | 1 | 40 | 76 | 33 | -0.34441 | -0.03660713 | 0.289186347 | 0.307805515 | 0.633598993 | 0.325793477 | 0.006038217 | 1.02E-06 | 0.006540239 |
| sp\|Q9BYE2\|TMPSD_HUMAN | 14 | 40 | 76 | 33 | 0.221018 | -0.058951166 | -0.237921936 | -0.279968853 | -0.458939623 | -0.178970771 | 4.30E-06 | 5.93E-10 | 0.008583336 |
| sp\|Q5SZK8\|FREM2_HUMAN | 8 | 8 | 13 | 5 | 0.199195 | 0.203875093 | 0.171607753 | 0.004680138 | -0.027587202 | -0.03226734 | 0.999075773 | 0.980254731 | 0.968505004 |
| sp\|Q96PE1\|AGRA2_HUMAN | 15 | 4 | 7 | 3 | 0.099983 | -0.063299483 | -0.471156732 | -0.163282412 | -0.571139661 | -0.407857249 | 0.74145192 | 0.125820307 | 0.25011881 |
| sp\|P11217\|PYGM_HUMAN | 4 | 11 | 21 | 8 | 0.238001 | -0.028375293 | 0.010503774 | -0.26637634 | -0.227497274 | 0.038879067 | 0.049615666 | 0.229128644 | 0.945309186 |
| sp\|Q14525\|KT33B_HUMAN | 39 | 4 | 7 | 3 | -0.23903 | 0.182980782 | -0.412042328 | 0.422005937 | -0.173017173 | -0.59502311 | 0.702137955 | 0.959591662 | 0.566574684 |
| sp\|Q9UHN6\|CEIP2_HUMAN | 2 | 3 | 7 | 4 | 0.138495 | 0.123169072 | -0.020942311 | -0.015326129 | -0.159437513 | -0.144111384 | 0.991133362 | 0.480050524 | 0.415649545 |
| sp\|Q13247\|SRSF6_HUMAN | 7 | 14 | 28 | 12 | -0.27745 | -0.166233651 | 0.536365645 | 0.111215346 | 0.813814642 | 0.702599296 | 0.834851825 | 0.002833585 | 0.003328984 |
| sp\|Q04759\|KPCT_HUMAN | 2 | 15 | 28 | 11 | 0.057223 | -0.065489586 | 0.384263789 | -0.122712673 | 0.327040702 | 0.449753375 | 0.845769548 | 0.466598719 | 0.173255568 |
| sp\|P41091\|IF2G_HUMAN | 4 | 7 | 14 | 6 | 0.195786 | -0.050424235 | 0.613993665 | -0.246210596 | 0.418207304 | 0.6644179 | 0.595785787 | 0.363761992 | 0.048591224 |
| sp\|Q5H9R7\|PP6R3_HUMAN | 9 | 7 | 14 | 6 | 0.047084 | 0.265251953 | 0.402450793 | 0.218167727 | 0.355366567 | 0.13719884 | 0.511404641 | 0.30056707 | 0.783953919 |
| sp\|P84157\|MXRA7_HUMAN | 11 | 14 | 27 | 13 | -0.26422 | -0.303549597 | 0.175072853 | -0.039326037 | 0.439296414 | 0.478622451 | 0.967599879 | 0.059856009 | 0.014797652 |
| sp\|O75084\|FZD7_HUMAN | 1 | 7 | 13 | 6 | -0.02871 | -0.120302467 | 0.280909605 | -0.091593089 | 0.309618982 | 0.401212072 | 0.782959485 | 0.158887645 | 0.027181447 |
| sp\|Q07507\|DERM_HUMAN | 2 | 7 | 14 | 6 | 0.274236 | 0.191859333 | 0.260982127 | -0.082377155 | -0.01325436 | 0.069122794 | 0.79731873 | 0.995907309 | 0.86582474 |
| sp\|O60496\|DOK2_HUMAN | 8 | 11 | 21 | 9 | -0.52942 | -0.055007285 | 0.48473374 | 0.474412271 | 1.014153296 | 0.539741025 | 0.508575387 | 0.130856514 | 0.466818416 |
| sp\|Q15762\|CD226_HUMAN | 13 | 33 | 62 | 28 | 0.037884 | -0.052069827 | 0.033898936 | -0.089953835 | -0.003985072 | 0.085968763 | 0.45378094 | 0.998899411 | 0.523366849 |
| sp\|Q9H853\|TBA4B_HUMAN | 4 | 14 | 28 | 13 | -0.12508 | 0.088602681 | 0.004884327 | 0.213681256 | 0.129962902 | -0.083718354 | 0.585180471 | 0.865490822 | 0.923974832 |
| sp\|Q06278\|AOXA_HUMAN | 21 | 4 | 7 | 3 | -0.30331 | 0.202802765 | 0.10633494 | 0.506109789 | 0.409641964 | -0.096467826 | 0.337770108 | 0.602600847 | 0.964642818 |
| sp\|P04259\|K2C6B_HUMAN | 1 | 4 | 7 | 3 | 0.660859 | -0.398713538 | -0.637947034 | -1.059572857 | -1.298806353 | -0.239233496 | 0.149115852 | 0.14630248 | 0.908986845 |
| sp\|P09564\|CD7_HUMAN | 1 | 4 | 6 | 3 | 0.030495 | 0.011498124 | 0.791422524 | -0.018997283 | 0.760927117 | 0.7799244 | 0.998815491 | 0.302217656 | 0.239264689 |
| sp\|Q7Z7H5\|TMED4_HUMAN | 7 | 8 | 13 | 6 | 0.250772 | 0.024392227 | 0.50336949 | -0.22637938 | 0.252597883 | 0.478977263 | 0.380882339 | 0.433037546 | 0.039660347 |
| sp\|P18054\|LOX12_HUMAN | 3 | 7 | 14 | 6 | -0.24481 | -0.104665786 | 0.343576333 | 0.140147335 | 0.588389454 | 0.448242119 | 0.868770486 | 0.200880167 | 0.291737412 |
| sp\|P08962\|CD63_HUMAN | 1 | 4 | 7 | 3 | 1.155806 | 1.069230794 | 0.843818172 | -0.086574942 | -0.311987564 | -0.225412622 | 0.93698646 | 0.580602977 | 0.701823886 |
| sp\|Q7Z3S9\|NT2NA_HUMAN | 3 | 26 | 48 | 21 | 0.048453 | -0.03815405 | 0.128627342 | -0.086607298 | 0.080174094 | 0.166781392 | 0.726508232 | 0.827814186 | 0.362070147 |
| sp\|Q9H772\|GREM2_HUMAN | 20 | 18 | 35 | 16 | -0.11092 | -0.160594565 | 0.241142216 | -0.049677493 | 0.352059288 | 0.401736781 | 0.919529604 | 0.057838734 | 0.009529611 |
| sp\|P42566\|EPS15_HUMAN | 4 | 12 | 21 | 8 | -0.33466 | -0.272852272 | 0.552152248 | 0.061805276 | 0.886809796 | 0.82500452 | 0.972200767 | 0.03669905 | 0.032116565 |
| sp\|O60518\|RNBP6_HUMAN | 2 | 4 | 7 | 2 | -0.25657 | -0.044563526 | 1.01782939 | 0.212002527 | 1.274395443 | 1.062392916 | 0.932113727 | 0.306483464 | 0.375355127 |
| sp\|Q9BZZ2\|SN_HUMAN | 2 | 32 | 62 | 27 | -0.51866 | -0.289385384 | 0.068241914 | 0.229270762 | 0.58689806 | 0.357627298 | 0.09861549 | 6.20E-05 | 0.007638417 |
| sp\|Q8TEA8\|DTD1_HUMAN | 2 | 25 | 49 | 20 | -0.05741 | -0.23164318 | 0.143626189 | -0.174228751 | 0.201040617 | 0.375269368 | 0.574992431 | 0.609578219 | 0.116284938 |
| sp\|Q76M96\|CCD80_HUMAN | 5 | 21 | 42 | 19 | -0.40747 | -0.323162841 | 0.277186638 | 0.084306599 | 0.684656078 | 0.600349479 | 0.809062887 | 0.000165935 | 0.000155597 |
| sp\|P35558\|PCKGC_HUMAN | 14 | 7 | 14 | 5 | 0.18947 | 0.357118635 | 0.598719224 | 0.167648354 | 0.409248943 | 0.241600588 | 0.870590604 | 0.602209822 | 0.797500987 |
| sp\|P0DP01\|HV108_HUMAN | 2 | 7 | 14 | 6 | -0.5333 | -0.188576827 | 0.338371326 | 0.344727661 | 0.871675814 | 0.526948153 | 0.512252966 | 0.067395607 | 0.255986439 |
| sp\|Q07955\|SRSF1_HUMAN | 1 | 4 | 6 | 3 | -0.23547 | -0.06035606 | -0.002036418 | 0.175115273 | 0.233434914 | 0.058319642 | 0.643418936 | 0.57516306 | 0.958390084 |
| sp\|P01127\|PDGFB_HUMAN | 3 | 7 | 14 | 6 | -0.01336 | -0.231791852 | 0.72971255 | -0.218429082 | 0.74307532 | 0.961504402 | 0.699688187 | 0.075554696 | 0.00660365 |
| sp\|Q96FN4\|CPNE2_HUMAN | 2 | 10 | 21 | 9 | 0.111212 | 0.098976911 | 0.705559165 | -0.012235239 | 0.594347016 | 0.606582254 | 0.997537763 | 0.026360085 | 0.007812473 |
| sp\|P68363\|TBA1B_HUMAN | 2 | 36 | 69 | 30 | -0.63967 | -0.168901978 | 0.068773079 | 0.470771403 | 0.70844646 | 0.237675057 | 0.015242129 | 0.001657748 | 0.376686577 |
| sp\|Q9Y613\|FHOD1_HUMAN | 5 | 4 | 7 | 3 | 0.098943 | 0.236042741 | -0.386908521 | 0.137100127 | -0.485851136 | -0.622951263 | 0.941084443 | 0.610961991 | 0.386971026 |
| sp\|Q86V81\|THOC4_HUMAN | 1 | 4 | 6 | 3 | -0.14692 | 0.257369884 | 0.628506688 | 0.404290349 | 0.775427152 | 0.371136803 | 0.696526055 | 0.407475906 | 0.773528213 |
| sp\|Q99598\|TSNAX_HUMAN | 1 | 4 | 7 | 3 | 0.482411 | 0.650097671 | 1.355874137 | 0.167686356 | 0.873462822 | 0.705776466 | 0.809047589 | 0.052061701 | 0.083762717 |
| sp\|P35318\|ADML_HUMAN | 1 | 40 | 76 | 33 | -0.50141 | -0.580594135 | 0.292292864 | -0.07918314 | 0.793703859 | 0.872886999 | 0.84457393 | 2.56E-05 | 1.72E-07 |
| sp\|P13535\|MYH8_HUMAN | 9 | 4 | 7 | 3 | 0.104229 | -0.019147401 | 0.003783924 | -0.123376056 | -0.100444731 | 0.022931325 | 0.518975483 | 0.738668128 | 0.980325351 |
| sp\|Q96EM0\|T3HPD_HUMAN | 2 | 10 | 21 | 10 | -0.24417 | 0.160954343 | 0.435041112 | 0.405120256 | 0.679207025 | 0.274086769 | 0.182918288 | 0.034974275 | 0.449792276 |
| sp\|O43897\|TLL1_HUMAN | 3 | 40 | 76 | 33 | -0.07546 | 0.039609497 | -0.151091577 | 0.115073857 | -0.075627218 | -0.190701074 | 0.19495919 | 0.610700912 | 0.021201213 |
| sp\|P22004\|BMP6_HUMAN | 2 | 22 | 41 | 18 | 0.115789 | -0.037876225 | -0.155618439 | -0.153664898 | -0.271407112 | -0.117742214 | 0.416267787 | 0.155661242 | 0.635896748 |
| sp\|Q8IWU5\|SULF2_HUMAN | 8 | 33 | 62 | 26 | -0.08322 | -0.191066059 | 0.319366714 | -0.107844003 | 0.40258877 | 0.510432773 | 0.600916583 | 0.01039545 | 0.000148283 |
| sp\|P20142\|PEPC_HUMAN | 3 | 14 | 28 | 12 | -0.00735 | -0.081255752 | 0.188145792 | -0.073908359 | 0.195493184 | 0.269401543 | 0.929579909 | 0.703712081 | 0.424329832 |
| sp\|Q9BX10\|GTPB2_HUMAN | 1 | 3 | 7 | 4 | 0.292547 | 0.070685181 | 0.364532803 | -0.221862279 | 0.071985342 | 0.293847622 | 0.787925405 | 0.979303166 | 0.609884444 |

| | | | | | | | | | | | | | |
|---|---|---|---|---|---|---|---|---|---|---|---|---|---|
| sp\|Q5TFQ8\|SIRBL_HUMAN | 229 | 30 | 55 | 24 | -0.34539 | -0.069523328 | -0.121868815 | 0.275870012 | 0.223524525 | -0.052345487 | 0.340556606 | 0.612921608 | 0.966719751 |
| sp\|O75487\|GPC4_HUMAN | 16 | 4 | 7 | 3 | -0.02067 | 0.017984564 | -0.127974765 | 0.038652987 | -0.107306342 | -0.145959329 | 0.918753865 | 0.653328665 | 0.398603874 |
| sp\|P0C7U1\|ASA2B_HUMAN | 30 | 23 | 41 | 17 | 0.203415 | 0.092183289 | -0.097155334 | -0.111231709 | -0.300570332 | -0.189338623 | 0.440678971 | 0.022776266 | 0.149242511 |
| sp\|O94903\|PLPHP_HUMAN | 6 | 11 | 20 | 9 | -0.14922 | -0.12929562 | 0.28183041 | 0.019922219 | 0.431048249 | 0.41112603 | 0.992828966 | 0.111238885 | 0.083494157 |
| sp\|O95302\|FKBP9_HUMAN | 3 | 11 | 20 | 9 | 0.114261 | 0.150392806 | 0.246434448 | 0.036131487 | 0.132173129 | 0.096041643 | 0.950102247 | 0.624025094 | 0.730709176 |
| sp\|P20337\|RAB3B_HUMAN | 41 | 24 | 48 | 22 | -0.19569 | -0.083824729 | 0.23894058 | 0.111868968 | 0.434634277 | 0.322765309 | 0.419855306 | 0.000214764 | 0.001829913 |
| sp\|P12724\|ECP_HUMAN | 1 | 24 | 49 | 21 | -0.05481 | -0.070090477 | 0.141470896 | -0.01527773 | 0.196283642 | 0.211561372 | 0.992219378 | 0.412794823 | 0.261813699 |
| sp\|Q8N1N4\|K2C78_HUMAN | 2 | 14 | 28 | 13 | 0.109004 | -0.056593359 | -0.364940975 | -0.165597752 | -0.473945369 | -0.308347616 | 0.891156387 | 0.510188883 | 0.685406027 |
| sp\|A8MUU1\|FB5L3_HUMAN | 28 | 40 | 76 | 33 | -0.52415 | -0.373750618 | 0.091972651 | 0.150403505 | 0.616126774 | 0.465723268 | 0.523288523 | 0.000882698 | 0.005494206 |
| sp\|Q9C0H2\|TTYH3_HUMAN | 1 | 15 | 28 | 12 | -1.09134 | 0.094287384 | 0.150328271 | 1.185623958 | 1.241664845 | 0.056040887 | 0.000100929 | 0.00074551 | 0.978480657 |
| sp\|O95866\|G6B_HUMAN | 2 | 33 | 62 | 27 | -0.42435 | 0.137710678 | 0.059612075 | 0.562059654 | 0.483961052 | -0.078098602 | 5.87E-05 | 0.005331295 | 0.832556361 |
| sp\|Q9BYX7\|ACTBM_HUMAN | 1 | 15 | 27 | 11 | -0.01456 | -0.191253411 | 0.055619203 | -0.176690788 | 0.070181826 | 0.246872614 | 0.461665051 | 0.921649503 | 0.29810833 |
| sp\|Q9Y3Q8\|T22D4_HUMAN | 9 | 7 | 14 | 6 | -0.69278 | -0.112799324 | 0.78922918 | 0.579982511 | 1.482011015 | 0.902028504 | 0.163422999 | 0.001412357 | 0.026616062 |
| sp\|Q9UKR3\|KLK13_HUMAN | 12 | 7 | 14 | 6 | -0.4191 | -0.113066021 | -0.41877019 | 0.306037611 | 0.000333442 | -0.305704169 | 0.733542801 | 0.999999742 | 0.756787138 |
| sp\|P17066\|HSP76_HUMAN | 15 | 40 | 76 | 33 | 0.053013 | 0.087365127 | 0.125023723 | 0.034351831 | 0.072010427 | 0.037658596 | 0.726715521 | 0.382399175 | 0.714078862 |
| sp\|P35606\|COPB2_HUMAN | 6 | 7 | 14 | 6 | -0.12641 | 0.057965745 | 0.285366268 | 0.184372484 | 0.411773007 | 0.227400523 | 0.795870014 | 0.463420335 | 0.732419047 |
| sp\|Q96A72\|MGN2_HUMAN | 2 | 4 | 6 | 3 | -0.22958 | 0.154200522 | 0.145421695 | 0.383775543 | 0.374996716 | -0.008778827 | 0.387573855 | 0.513709491 | 0.999543271 |
| sp\|Q14213\|IL27B_HUMAN | 2 | 15 | 28 | 12 | -0.13393 | -0.009977554 | 0.164186439 | 0.123952175 | 0.298116167 | 0.174163993 | 0.547280731 | 0.101320583 | 0.362833705 |
| sp\|P52597\|HNRPF_HUMAN | 16 | 7 | 14 | 6 | -0.13317 | -0.148799432 | 0.417620619 | -0.01562567 | 0.550794381 | 0.566420051 | 0.998143243 | 0.224710333 | 0.134696174 |
| sp\|Q6ZMR3\|LDH6A_HUMAN | 27 | 40 | 76 | 33 | -0.09341 | -0.028550867 | 0.242722796 | 0.06485695 | 0.336130612 | 0.271273663 | 0.653489759 | 0.000639817 | 0.002101959 |
| sp\|O95803\|NDST3_HUMAN | 7 | 4 | 7 | 3 | 0.102677 | 0.049329383 | 0.089655466 | -0.053348002 | -0.013021919 | 0.040326084 | 0.728633786 | 0.987007237 | 0.859318 |
| sp\|Q08397\|LOXL1_HUMAN | 3 | 7 | 13 | 6 | 0.3771 | 0.290344449 | 0.243998255 | -0.086755092 | -0.133101286 | -0.046346193 | 0.83028477 | 0.734157437 | 0.952937475 |
| sp\|Q969P0\|IGSF8_HUMAN | 1 | 10 | 21 | 10 | -0.11057 | 0.022930109 | -0.074410952 | 0.133498648 | 0.036157588 | -0.09734106 | 0.801886683 | 0.988041589 | 0.888915836 |
| sp\|P25705\|ATPA_HUMAN | 3 | 8 | 13 | 6 | 0.001067 | -0.265465586 | 0.712665497 | -0.266532495 | 0.711598588 | 0.978131083 | 0.856756996 | 0.475862154 | 0.199065417 |
| sp\|P55010\|IF5_HUMAN | 22 | 4 | 7 | 3 | -0.59973 | -0.32072814 | 0.869485553 | 0.279004826 | 1.469218519 | 1.190213693 | 0.753177515 | 0.023505569 | 0.041065902 |
| sp\|P84085\|ARF5_HUMAN | 5 | 11 | 21 | 9 | 0.241137 | 0.259086088 | 0.274888544 | 0.017949429 | 0.033751885 | 0.015802456 | 0.996389743 | 0.99127047 | 0.997556378 |
| sp\|O75821\|EIF3G_HUMAN | 15 | 8 | 13 | 6 | -0.40528 | -0.447617256 | 0.219675868 | -0.042341014 | 0.62495211 | 0.667293124 | 0.996442881 | 0.591693555 | 0.491500028 |
| sp\|Q6UXB4\|CLC4G_HUMAN | 6 | 12 | 21 | 9 | -0.30135 | -0.149380349 | -0.054536803 | 0.151967967 | 0.246811513 | 0.094843546 | 0.187451429 | 0.056627679 | 0.573463908 |
| sp\|Q8N6G6\|ATL1_HUMAN | 3 | 4 | 7 | 3 | -0.18692 | -0.407801993 | -0.5536664 | -0.220880315 | -0.366744722 | -0.145864407 | 0.128829086 | 0.035418823 | 0.435640356 |
| sp\|P03971\|MIS_HUMAN | 9 | 4 | 7 | 3 | 0.340621 | 0.259253028 | -0.012323823 | -0.081368018 | -0.352944869 | -0.271576851 | 0.899663564 | 0.299748209 | 0.406511867 |
| sp\|Q96F85\|CNRP1_HUMAN | 8 | 11 | 21 | 9 | -0.0877 | -0.135857114 | 0.411997537 | -0.04815756 | 0.499697092 | 0.547854652 | 0.96666576 | 0.096820296 | 0.031621468 |
| sp\|Q02223\|TNR17_HUMAN | 28 | 19 | 35 | 15 | 0.005062 | -0.077278082 | 0.347746762 | -0.082339755 | 0.34268509 | 0.425024844 | 0.62237323 | 0.005933509 | 0.000102744 |
| sp\|P35625\|TIMP3_HUMAN | 2 | 7 | 14 | 7 | -0.03721 | 0.056059792 | 0.143654655 | 0.093269179 | 0.180864043 | 0.087594863 | 0.873128227 | 0.684852533 | 0.887141301 |
| sp\|Q14353\|GAMT_HUMAN | 446 | 4 | 7 | 3 | -0.16289 | -0.178496346 | 0.356494451 | -0.01560923 | 0.519381567 | 0.534990797 | 0.998980402 | 0.491388599 | 0.40407771 |
| sp\|P14866\|HNRPL_HUMAN | 1 | 7 | 14 | 5 | -0.41804 | -0.430417606 | 0.736405412 | -0.012379897 | 1.15444312 | 1.166823017 | 0.999467886 | 0.077345743 | 0.04067815 |
| sp\|Q16553\|LY6E_HUMAN | 2 | 25 | 48 | 21 | 0.019135 | 0.073633991 | 0.125110693 | 0.054499411 | 0.105976113 | 0.051476702 | 0.751827762 | 0.475310562 | 0.797433522 |
| sp\|P01611\|KVD12_HUMAN | 15 | 15 | 27 | 12 | 0.005928 | -0.006959527 | -0.017744453 | -0.012887912 | -0.023672838 | -0.010784926 | 0.994385089 | 0.986953973 | 0.996608481 |

### Supplemental Table II:

| Description | SvsHC_padjust | SvsHC_Count | SvsM_padjust | SvsM_Count | MvsHC_padjust | MvsHC_Count |
|---|---|---|---|---|---|---|
| Neutrophil degranulation | 3.92E-59 | 183 | 7.76E-59 | 177 | 0.000172546 | 23 |
| Extracellular matrix organization | 5.50E-40 | 120 | 4.05E-34 | 109 | 1.88E-09 | 25 |
| Regulation of Insulin-like Growth Factor (IGF) transport and uptake by Insulin-like Growth Factor Binding Proteins (IGFBPs) | 3.40E-27 | 63 | 5.85E-24 | 58 | 0.005455229 | 9 |
| Post-translational protein phosphorylation | 5.76E-24 | 55 | 1.61E-20 | 50 | 0.002464872 | 9 |
| Platelet degranulation | 1.21E-23 | 60 | 7.65E-15 | 47 | 1.89E-16 | 24 |
| Binding and Uptake of Ligands by Scavenger Receptors | 6.84E-23 | 33 | 4.35E-18 | 29 | NA | NA |
| Response to elevated platelet cytosolic Ca2+ | 9.84E-23 | 60 | 3.52E-14 | 47 | 2.41E-16 | 24 |
| AUF1 (hnRNP D0) binds and destabilizes mRNA | 9.84E-23 | 38 | 8.94E-20 | 35 | NA | NA |
| ER-Phagosome pathway | 2.67E-21 | 47 | 9.21E-14 | 37 | NA | NA |
| Antigen processing-Cross presentation | 2.67E-21 | 51 | 1.40E-13 | 40 | NA | NA |
| The role of GTSE1 in G2/M progression after G2 checkpoint | 5.29E-21 | 43 | 9.21E-14 | 34 | NA | NA |
| Regulation of activated PAK-2p34 by proteasome mediated degradation | 1.85E-20 | 34 | 1.89E-15 | 29 | NA | NA |
| RUNX1 regulates transcription of genes involved in differentiation of HSCs | 2.31E-20 | 56 | 2.89E-17 | 51 | NA | NA |
| Vif-mediated degradation of APOBEC3G | 3.82E-20 | 35 | 2.49E-15 | 30 | NA | NA |
| FBXL7 down-regulates AURKA during mitotic entry and in early mitosis | 3.82E-20 | 35 | 2.49E-15 | 30 | NA | NA |
| Negative regulation of NOTCH4 signaling | 3.82E-20 | 35 | 3.17E-16 | 31 | NA | NA |
| Cross-presentation of soluble exogenous antigens (endosomes) | 8.25E-20 | 33 | 8.21E-15 | 28 | NA | NA |
| SCF-beta-TrCP mediated degradation of Emi1 | 8.25E-20 | 35 | 4.73E-15 | 30 | NA | NA |
| Vpu mediated degradation of CD4 | 8.25E-20 | 34 | 6.31E-15 | 29 | NA | NA |
| NIK-->noncanonical NF-kB signaling | 1.60E-19 | 36 | 5.71E-15 | 31 | NA | NA |
| Regulation of Apoptosis | 1.88E-19 | 34 | 1.07E-14 | 29 | NA | NA |
| Dectin-1 mediated noncanonical NF-kB signaling | 3.27E-19 | 36 | 9.02E-15 | 31 | NA | NA |
| Regulation of RUNX3 expression and activity | 9.00E-19 | 34 | 2.42E-13 | 28 | NA | NA |
| Autodegradation of the E3 ubiquitin ligase COP1 | 9.00E-19 | 33 | 4.75E-14 | 28 | NA | NA |
| Ubiquitin Mediated Degradation of Phosphorylated Cdc25A | 9.00E-19 | 33 | 4.75E-14 | 28 | NA | NA |
| p53-Independent DNA Damage Response | 9.00E-19 | 33 | 4.75E-14 | 28 | NA | NA |
| p53-Independent G1/S DNA damage checkpoint | 9.00E-19 | 33 | 4.75E-14 | 28 | NA | NA |
| Ubiquitin-dependent degradation of Cyclin D | 9.00E-19 | 33 | 4.75E-14 | 28 | NA | NA |
| G2/M Checkpoints | 9.33E-19 | 62 | 3.55E-16 | 57 | NA | NA |
| SCF(Skp2)-mediated degradation of p27/p21 | 2.83E-18 | 35 | 6.24E-14 | 30 | NA | NA |
| Degradation of GLI1 by the proteasome | 2.83E-18 | 35 | 1.32E-15 | 32 | NA | NA |
| Degradation of GLI2 by the proteasome | 2.83E-18 | 35 | 1.32E-15 | 32 | NA | NA |
| GLI3 is processed to GLI3R by the proteasome | 2.83E-18 | 35 | 1.32E-15 | 32 | NA | NA |
| Regulation of ornithine decarboxylase (ODC) | 4.68E-18 | 32 | 2.14E-13 | 27 | NA | NA |
| Degradation of AXIN | 9.02E-18 | 33 | 2.42E-13 | 28 | NA | NA |
| Regulation of mRNA stability by proteins that bind AU-rich elements | 1.40E-17 | 42 | 4.45E-17 | 41 | NA | NA |
| Oxygen-dependent proline hydroxylation of Hypoxia-inducible Factor Alpha | 1.61E-17 | 36 | 1.69E-13 | 31 | NA | NA |
| Hh mutants are degraded by ERAD | 1.83E-17 | 33 | 4.12E-13 | 28 | NA | NA |
| Signaling by NOTCH | 3.49E-17 | 73 | 2.86E-16 | 70 | NA | NA |
| Degradation of DVL | 3.54E-17 | 33 | 6.44E-13 | 28 | NA | NA |
| Stabilization of p53 | 3.54E-17 | 33 | 6.44E-13 | 28 | NA | NA |
| ROS sensing by NFE2L2 | 3.54E-17 | 33 | 9.40E-14 | 29 | NA | NA |
| Interleukin-1 signaling | 4.97E-17 | 45 | 3.41E-13 | 39 | NA | NA |
| Hedgehog ligand biogenesis | 7.20E-17 | 35 | 6.44E-13 | 30 | NA | NA |
| Regulation of HMOX1 expression and activity | 7.20E-17 | 35 | 1.04E-13 | 31 | NA | NA |
| CLEC7A (Dectin-1) signaling | 1.28E-16 | 44 | 3.17E-16 | 43 | NA | NA |
| Hh mutants abrogate ligand secretion | 1.33E-16 | 33 | 1.76E-12 | 28 | NA | NA |
| CDT1 association with the CDC6:ORC:origin complex | 1.33E-16 | 33 | 1.76E-12 | 28 | NA | NA |
| FCERI mediated NF-kB activation | 2.25E-16 | 39 | 6.44E-13 | 34 | NA | NA |
| Activation of NF-kappaB in B cells | 2.35E-16 | 35 | 1.65E-12 | 30 | NA | NA |
| Defective CFTR causes cystic fibrosis | 4.93E-16 | 33 | 4.73E-12 | 28 | NA | NA |
| ECM proteoglycans | 5.55E-16 | 37 | 6.33E-11 | 30 | 3.58E-05 | 10 |
| Cellular response to chemical stress | 6.16E-16 | 56 | 4.80E-13 | 50 | NA | NA |
| Regulation of RUNX2 expression and activity | 8.51E-16 | 36 | 2.09E-11 | 30 | NA | NA |
| TNFR2 non-canonical NF-kB pathway | 1.15E-15 | 43 | 5.56E-12 | 37 | NA | NA |
| Metabolism of polyamines | 1.25E-15 | 32 | 1.29E-11 | 27 | NA | NA |
| Platelet activation, signaling and aggregation | 1.32E-15 | 75 | 3.10E-11 | 64 | 2.33E-13 | 28 |
| Signaling by NOTCH4 | 1.52E-15 | 38 | 6.44E-13 | 34 | NA | NA |
| Orc1 removal from chromatin | 2.15E-15 | 35 | 9.35E-12 | 30 | NA | NA |
| Degradation of the extracellular matrix | 2.15E-15 | 51 | 3.26E-17 | 53 | NA | NA |
| Cellular response to hypoxia | 2.30E-15 | 36 | 7.95E-12 | 31 | NA | NA |
| Downstream signaling events of B Cell Receptor (BCR) | 2.36E-15 | 38 | 3.66E-14 | 36 | NA | NA |
| Autodegradation of Cdh1 by Cdh1:APC/C | 2.68E-15 | 33 | 1.87E-11 | 28 | NA | NA |
| Asymmetric localization of PCP proteins | 2.68E-15 | 33 | 2.80E-12 | 29 | NA | NA |
| Regulation of RAS by GAPs | 3.12E-15 | 34 | 1.65E-11 | 29 | NA | NA |
| MAPK6/MAPK4 signaling | 5.61E-15 | 39 | 6.24E-14 | 37 | NA | NA |
| p53-Dependent G1 DNA Damage Response | 8.34E-15 | 33 | 4.30E-11 | 28 | NA | NA |
| p53-Dependent G1/S DNA damage checkpoint | 8.34E-15 | 33 | 4.30E-11 | 28 | NA | NA |
| Signaling by Interleukins | 1.02E-14 | 106 | 6.42E-13 | 98 | NA | NA |
| Interleukin-1 family signaling | 1.33E-14 | 50 | 3.32E-12 | 45 | NA | NA |
| Degradation of beta-catenin by the destruction complex | 1.54E-14 | 37 | 2.91E-11 | 32 | NA | NA |
| APC/C:Cdc20 mediated degradation of Securin | 2.39E-14 | 33 | 9.42E-11 | 28 | NA | NA |
| Assembly of the pre-replicative complex | 2.39E-14 | 33 | 9.42E-11 | 28 | NA | NA |
| G1/S DNA Damage Checkpoints | 2.39E-14 | 33 | 9.42E-11 | 28 | NA | NA |
| Amyloid fiber formation | 3.43E-14 | 43 | 4.29E-12 | 39 | NA | NA |
| Apoptosis | 3.82E-14 | 57 | 3.11E-13 | 54 | NA | NA |
| Host Interactions of HIV factors | 3.82E-14 | 47 | 4.90E-11 | 41 | NA | NA |
| Regulation of PTEN stability and activity | 3.92E-14 | 33 | 1.39E-10 | 28 | NA | NA |
| Hedgehog 'off' state | 6.96E-14 | 43 | 6.24E-10 | 36 | NA | NA |
| Transcriptional regulation by RUNX3 | 1.02E-13 | 39 | 1.79E-09 | 32 | NA | NA |
| Formation of Fibrin Clot (Clotting Cascade) | 1.44E-13 | 24 | 4.05E-08 | 18 | 0.009288971 | 5 |
| UCH proteinases | 1.86E-13 | 40 | 1.16E-10 | 35 | NA | NA |
| Downstream TCR signaling | 2.22E-13 | 39 | 1.58E-10 | 34 | NA | NA |
| Integrin cell surface interactions | 2.28E-13 | 36 | 2.88E-10 | 31 | 0.002532341 | 8 |
| Regulation of APC/C activators between G1/S and early anaphase | 2.50E-13 | 35 | 3.79E-10 | 30 | NA | NA |
| ABC transporter disorders | 2.68E-13 | 34 | 4.97E-10 | 29 | NA | NA |
| Cdc20:Phospho-APC/C mediated degradation of Cyclin A | 2.75E-13 | 33 | 6.31E-10 | 28 | NA | NA |
| CDK-mediated phosphorylation and removal of Cdc6 | 2.75E-13 | 33 | 6.31E-10 | 28 | NA | NA |
| Programmed Cell Death | 2.96E-13 | 61 | 1.43E-12 | 58 | NA | NA |
| C-type lectin receptors (CLRs) | 3.32E-13 | 48 | 1.87E-14 | 49 | NA | NA |
| APC/C:Cdh1 mediated degradation of Cdc20 and other APC/C:Cdh1 targeted proteins in late mitosis/early G1 | 4.28E-13 | 33 | 8.96E-10 | 28 | NA | NA |
| APC:Cdc20 mediated degradation of cell cycle proteins prior to satisfation of the cell cycle checkpoint | 4.28E-13 | 33 | 8.96E-10 | 28 | NA | NA |

| | | | | | | |
|---|---|---|---|---|---|---|
| Cyclin E associated events during G1/S transition | 5.62E-13 | 35 | 7.34E-10 | 30 | NA | NA |
| TCR signaling | 7.06E-13 | 43 | 5.67E-11 | 39 | NA | NA |
| APC/C:Cdc20 mediated degradation of mitotic proteins | 1.07E-12 | 33 | 1.78E-09 | 28 | NA | NA |
| Signaling by the B Cell Receptor (BCR) | 1.17E-12 | 41 | 1.26E-12 | 40 | NA | NA |
| Cyclin A:Cdk2-associated events at S phase entry | 1.28E-12 | 35 | 1.37E-09 | 30 | NA | NA |
| Activation of APC/C and APC/C:Cdc20 mediated degradation of mitotic proteins | 1.63E-12 | 33 | 2.44E-09 | 28 | NA | NA |
| PCP/CE pathway | 3.56E-12 | 36 | 2.48E-11 | 34 | NA | NA |
| Transcriptional regulation by RUNX1 | 3.87E-12 | 64 | 2.52E-09 | 56 | NA | NA |
| APC/C-mediated degradation of cell cycle proteins | 4.19E-12 | 35 | 3.33E-09 | 30 | NA | NA |
| Regulation of mitotic cell cycle | 4.19E-12 | 35 | 3.33E-09 | 30 | NA | NA |
| Metabolism of carbohydrates | 5.95E-12 | 73 | 1.12E-11 | 70 | NA | NA |
| G2/M Transition | 6.48E-12 | 56 | 7.83E-08 | 46 | NA | NA |
| Mitotic G2-G2/M phases | 1.02E-11 | 56 | 1.08E-07 | 46 | NA | NA |
| Fc epsilon receptor (FCERI) signaling | 1.09E-11 | 44 | 3.48E-11 | 42 | NA | NA |
| Cytoprotection by HMOX1 | 1.10E-11 | 42 | 2.44E-09 | 37 | NA | NA |
| Switching of origins to a post-replicative state | 1.28E-11 | 35 | 7.89E-09 | 30 | NA | NA |
| Non-integrin membrane-ECM interactions | 3.73E-11 | 27 | 3.13E-09 | 24 | 0.000241742 | 8 |
| Hedgehog 'on' state | 4.06E-11 | 33 | 6.33E-09 | 29 | NA | NA |
| DNA Replication Pre-Initiation | 4.06E-11 | 33 | 2.83E-08 | 28 | NA | NA |
| Scavenging by Class A Receptors | 4.31E-11 | 15 | 9.05E-13 | 16 | NA | NA |
| Gene and protein expression by JAK-STAT signaling after Interleukin-12 stimulation | 9.32E-11 | 21 | 3.09E-09 | 19 | 0.048698799 | 4 |
| Cell surface interactions at the vascular wall | 1.02E-10 | 43 | 1.46E-06 | 34 | 4.85E-06 | 14 |
| TCF dependent signaling in response to WNT | 1.14E-10 | 60 | 3.37E-10 | 57 | NA | NA |
| Signaling by Hedgehog | 1.44E-10 | 45 | 4.40E-08 | 39 | NA | NA |
| ABC-family proteins mediated transport | 1.51E-10 | 36 | 4.61E-08 | 31 | NA | NA |
| Scavenging of heme from plasma | 1.75E-10 | 12 | 4.05E-05 | 8 | NA | NA |
| RHO GTPases activate PKNs | 2.59E-10 | 34 | 6.33E-11 | 34 | NA | NA |
| Collagen degradation | 3.56E-10 | 27 | 1.12E-10 | 27 | NA | NA |
| Packaging Of Telomere Ends | 4.35E-10 | 24 | 1.52E-10 | 24 | NA | NA |
| M Phase | 7.61E-10 | 87 | 3.06E-07 | 76 | NA | NA |
| Cell Cycle Checkpoints | 8.52E-10 | 68 | 3.36E-09 | 64 | NA | NA |
| Common Pathway of Fibrin Clot Formation | 1.27E-09 | 15 | 9.39E-05 | 10 | 0.009288971 | 4 |
| Interleukin-12 signaling | 1.95E-09 | 22 | 3.26E-08 | 20 | NA | NA |
| Diseases of programmed cell death | 2.28E-09 | 34 | 8.90E-09 | 32 | NA | NA |
| Recognition and association of DNA glycosylase with site containing an affected purine | 2.76E-09 | 24 | 9.19E-10 | 24 | NA | NA |
| Cleavage of the damaged purine | 2.76E-09 | 24 | 9.19E-10 | 24 | NA | NA |
| Depurination | 2.76E-09 | 24 | 9.19E-10 | 24 | NA | NA |
| Diseases of metabolism | 2.76E-09 | 59 | 2.39E-09 | 57 | 0.005049397 | 13 |
| Interleukin-12 family signaling | 4.26E-09 | 24 | 2.55E-07 | 21 | NA | NA |
| Transcriptional regulation by RUNX2 | 5.72E-09 | 37 | 7.07E-07 | 32 | NA | NA |
| Regulation of expression of SLITs and ROBOs | 1.87E-08 | 45 | 7.26E-07 | 40 | NA | NA |
| Recognition and association of DNA glycosylase with site containing an affected pyrimidine | 2.16E-08 | 24 | 6.71E-09 | 24 | NA | NA |
| Cleavage of the damaged pyrimidine | 2.16E-08 | 24 | 6.71E-09 | 24 | NA | NA |
| Depyrimidination | 2.16E-08 | 24 | 6.71E-09 | 24 | NA | NA |
| Elastic fibre formation | 3.72E-08 | 20 | 3.00E-06 | 17 | 0.002763594 | 6 |
| Base-Excision Repair, AP Site Formation | 4.57E-08 | 24 | 1.39E-08 | 24 | NA | NA |
| G1/S Transition | 6.22E-08 | 37 | 4.50E-06 | 32 | NA | NA |
| Molecules associated with elastic fibres | 6.25E-08 | 18 | 7.26E-06 | 15 | 0.008831027 | 5 |
| Synthesis of DNA | 6.25E-08 | 35 | 5.86E-06 | 30 | NA | NA |
| HSP90 chaperone cycle for steroid hormone receptors (SHR) | 6.68E-08 | 22 | 0.000239457 | 16 | NA | NA |
| Signaling by WNT | 6.88E-08 | 69 | 5.65E-11 | 74 | NA | NA |
| Nonhomologous End-Joining (NHEJ) | 7.02E-08 | 25 | 2.15E-08 | 25 | NA | NA |
| Meiotic synapsis | 7.66E-08 | 27 | 2.17E-08 | 27 | NA | NA |
| Metabolism of amino acids and derivatives | 7.66E-08 | 75 | 0.000962166 | 58 | NA | NA |
| Separation of Sister Chromatids | 7.79E-08 | 47 | 7.64E-05 | 38 | NA | NA |
| Plasma lipoprotein remodeling | 8.53E-08 | 16 | 2.39E-06 | 14 | 0.000444061 | 6 |
| Plasma lipoprotein assembly, remodeling, and clearance | 9.54E-08 | 25 | 2.97E-08 | 25 | 0.004215306 | 7 |
| Recruitment and ATM-mediated phosphorylation of repair and signaling proteins at DNA double strand breaks | 1.86E-07 | 26 | 2.43E-07 | 25 | NA | NA |
| Diseases of glycosylation | 2.23E-07 | 38 | 4.40E-08 | 38 | NA | NA |
| RNA Polymerase I Promoter Opening | 2.23E-07 | 23 | 7.31E-08 | 23 | NA | NA |
| Inhibition of DNA recombination at telomere | 2.38E-07 | 24 | 7.50E-08 | 24 | NA | NA |
| DNA Double Strand Break Response | 2.45E-07 | 26 | 3.17E-07 | 25 | NA | NA |
| Mitotic Anaphase | 2.47E-07 | 53 | 0.00019331 | 43 | NA | NA |
| Mitotic Metaphase and Anaphase | 2.85E-07 | 53 | 0.000211739 | 43 | NA | NA |
| Signaling by ROBO receptors | 2.94E-07 | 50 | 4.27E-08 | 50 | NA | NA |
| MAPK family signaling cascades | 3.42E-07 | 66 | 2.33E-09 | 69 | NA | NA |
| DNA Replication | 3.43E-07 | 35 | 2.38E-05 | 30 | NA | NA |
| G2/M DNA damage checkpoint | 3.54E-07 | 29 | 9.66E-08 | 29 | NA | NA |
| Beta-catenin independent WNT signaling | 3.83E-07 | 38 | 7.12E-09 | 40 | NA | NA |
| Laminin interactions | 3.97E-07 | 15 | 1.49E-06 | 14 | 0.023394658 | 4 |
| MAPK1/MAPK3 signaling | 4.08E-07 | 60 | 4.64E-08 | 60 | NA | NA |
| DNA methylation | 4.08E-07 | 23 | 1.40E-07 | 23 | NA | NA |
| DNA Damage/Telomere Stress Induced Senescence | 4.13E-07 | 26 | 1.28E-07 | 26 | NA | NA |
| RAF/MAP kinase cascade | 4.41E-07 | 59 | 5.22E-08 | 59 | NA | NA |
| Intrinsic Pathway of Fibrin Clot Formation | 4.53E-07 | 13 | 2.36E-06 | 12 | NA | NA |
| L1CAM interactions | 5.45E-07 | 33 | 0.000314061 | 26 | 0.013689406 | 8 |
| Mitotic G1 phase and G1/S transition | 6.54E-07 | 38 | 7.39E-05 | 32 | NA | NA |
| Pre-NOTCH Expression and Processing | 7.11E-07 | 31 | 1.91E-07 | 31 | NA | NA |
| Activated PKN1 stimulates transcription of AR (androgen receptor) regulated genes KLK2 and KLK3 | 7.47E-07 | 23 | 2.64E-07 | 23 | NA | NA |
| SIRT1 negatively regulates rRNA expression | 1.01E-06 | 23 | 3.55E-07 | 23 | NA | NA |
| HIV Infection | 1.11E-06 | 51 | 3.39E-05 | 45 | NA | NA |
| Regulation of TLR by endogenous ligand | 1.21E-06 | 12 | 0.000402886 | 9 | NA | NA |
| Condensation of Prophase Chromosomes | 1.32E-06 | 24 | 4.50E-07 | 24 | NA | NA |
| Collagen formation | 1.39E-06 | 27 | 4.32E-07 | 27 | NA | NA |
| Formation of tubulin folding intermediates by CCT/TriC | 1.59E-06 | 13 | 0.001629824 | 9 | NA | NA |
| RUNX1 regulates genes involved in megakaryocyte differentiation and platelet function | 2.01E-06 | 28 | 2.50E-05 | 25 | NA | NA |
| Class I MHC mediated antigen processing & presentation | 2.12E-06 | 71 | 0.000397383 | 60 | NA | NA |
| Ub-specific processing proteases | 2.40E-06 | 48 | 3.54E-05 | 43 | NA | NA |
| Neddylation | 2.67E-06 | 50 | 0.00015853 | 43 | NA | NA |
| MET activates PTK2 signaling | 2.96E-06 | 14 | 1.49E-06 | 14 | NA | NA |
| PTEN Regulation | 3.26E-06 | 35 | 0.000336898 | 29 | NA | NA |
| MHC class II antigen presentation | 3.84E-06 | 32 | 0.023821381 | 21 | NA | NA |
| Metabolism of nucleotides | 3.86E-06 | 28 | 4.29E-05 | 25 | NA | NA |

| | | | | | | |
|---|---|---|---|---|---|---|
| HCMV Early Events | 3.88E-06 | 34 | 0.00017554 | 29 | NA | NA |
| PRC2 methylates histones and DNA | 3.96E-06 | 23 | 1.46E-06 | 23 | NA | NA |
| Defective pyroptosis | 3.96E-06 | 23 | 1.46E-06 | 23 | NA | NA |
| RHO GTPase Effectors | 4.17E-06 | 63 | 7.12E-09 | 68 | NA | NA |
| Signaling by PDGF | 4.20E-06 | 20 | 1.72E-06 | 20 | 0.039349738 | 5 |
| Syndecan interactions | 4.68E-06 | 13 | 0.002930495 | 9 | 0.000225937 | 6 |
| Deposition of new CENPA-containing nucleosomes at the centromere | 5.06E-06 | 23 | 1.86E-06 | 23 | NA | NA |
| Nucleosome assembly | 5.06E-06 | 23 | 1.86E-06 | 23 | NA | NA |
| Immunoregulatory interactions between a Lymphoid and a non-Lymphoid cell | 6.51E-06 | 33 | 0.0002921 | 28 | NA | NA |
| Cooperation of Prefoldin and TriC/CCT in actin and tubulin folding | 7.34E-06 | 14 | 0.002675233 | 10 | NA | NA |
| Diseases associated with glycosaminoglycan metabolism | 8.08E-06 | 16 | 3.89E-06 | 16 | NA | NA |
| MET promotes cell motility | 8.08E-06 | 16 | 6.68E-07 | 17 | NA | NA |
| ERCC6 (CSB) and EHMT2 (G9a) positively regulate rRNA expression | 8.33E-06 | 23 | 7.80E-07 | 24 | NA | NA |
| Pre-NOTCH Transcription and Translation | 9.33E-06 | 26 | 8.87E-07 | 27 | NA | NA |
| COPI-independent Golgi-to-ER retrograde traffic | 9.83E-06 | 18 | 0.010670328 | 12 | NA | NA |
| Assembly of collagen fibrils and other multimeric structures | 9.92E-06 | 20 | 9.23E-07 | 21 | NA | NA |
| Transport to the Golgi and subsequent modification | 9.96E-06 | 41 | 0.000436547 | 35 | NA | NA |
| Diseases of signal transduction by growth factor receptors and second messengers | 1.14E-05 | 71 | 5.37E-06 | 69 | NA | NA |
| Asparagine N-linked glycosylation | 1.49E-05 | 58 | 0.000647254 | 50 | NA | NA |
| Aggrephagy | 1.64E-05 | 16 | 0.007962825 | 11 | NA | NA |
| Gluconeogenesis | 1.68E-05 | 14 | 0.004327317 | 10 | NA | NA |
| Signaling by MET | 1.68E-05 | 23 | 1.68E-06 | 24 | NA | NA |
| Deregulated CDK5 triggers multiple neurodegenerative pathways in Alzheimer's disease models | 1.95E-05 | 11 | 0.000600248 | 9 | 7.61E-05 | 6 |
| Neurodegenerative Diseases | 1.95E-05 | 11 | 0.000600248 | 9 | 7.61E-05 | 6 |
| Chaperone Mediated Autophagy | 1.95E-05 | 11 | 1.21E-05 | 11 | NA | NA |
| Meiotic recombination | 2.29E-05 | 24 | 8.80E-06 | 24 | NA | NA |
| Glutathione synthesis and recycling | 2.54E-05 | 8 | 0.002007169 | 6 | NA | NA |
| Processing of DNA double-strand break ends | 2.54E-05 | 26 | 9.23E-06 | 26 | NA | NA |
| PIP3 activates AKT signaling | 2.54E-05 | 52 | 0.000208567 | 47 | NA | NA |
| Chondroitin sulfate/dermatan sulfate metabolism | 3.20E-05 | 17 | 0.009060506 | 12 | NA | NA |
| Glutathione conjugation | 3.59E-05 | 14 | 0.001845358 | 11 | NA | NA |
| COPI-mediated anterograde transport | 4.55E-05 | 26 | 0.002513656 | 21 | NA | NA |
| Toll-like Receptor Cascades | 4.60E-05 | 35 | 0.000208567 | 32 | NA | NA |
| Prefoldin mediated transfer of substrate to CCT/TriC | 5.02E-05 | 12 | 0.000908139 | 10 | NA | NA |
| Detoxification of Reactive Oxygen Species | 5.11E-05 | 14 | 0.000143072 | 13 | 0.008475125 | 5 |
| Defective B3GALTL causes Peters-plus syndrome (PpS) | 5.11E-05 | 14 | 0.000635663 | 12 | 0.046379153 | 4 |
| Defective B3GAT3 causes JDSSDHD | 5.26E-05 | 10 | 0.000258445 | 9 | NA | NA |
| Dissolution of Fibrin Clot | 5.71E-05 | 8 | 0.003254419 | 6 | NA | NA |
| Diseases associated with O-glycosylation of proteins | 5.79E-05 | 20 | 9.49E-05 | 19 | NA | NA |
| Formation of the beta-catenin:TCF transactivating complex | 6.24E-05 | 24 | 2.50E-05 | 24 | NA | NA |
| HCMV Infection | 6.89E-05 | 35 | 0.001410284 | 30 | NA | NA |
| O-glycosylation of TSR domain-containing proteins | 7.13E-05 | 14 | 0.000817677 | 12 | 0.048698799 | 4 |
| Base Excision Repair | 7.53E-05 | 24 | 3.05E-05 | 24 | NA | NA |
| Disorders of transmembrane transporters | 8.74E-05 | 37 | 0.000698581 | 33 | NA | NA |
| S Phase | 9.00E-05 | 35 | 0.001685437 | 30 | NA | NA |
| Retinoid metabolism and transport | 0.000102323 | 15 | 0.003098587 | 12 | 0.013689406 | 5 |
| Interconversion of nucleotide di- and triphosphates | 0.000110685 | 12 | 0.001587191 | 10 | NA | NA |
| CS/DS degradation | 0.000115908 | 8 | 0.000755221 | 7 | NA | NA |
| Deubiquitination | 0.000122016 | 54 | 0.001241469 | 48 | NA | NA |
| A tetrasaccharide linker sequence is required for GAG synthesis | 0.000129489 | 11 | 0.00219405 | 9 | NA | NA |
| Post-chaperonin tubulin folding pathway | 0.000142712 | 10 | NA | NA | NA | NA |
| Transcriptional regulation of granulopoiesis | 0.000156027 | 23 | 6.60E-05 | 23 | NA | NA |
| Collagen biosynthesis and modifying enzymes | 0.000159742 | 19 | 7.62E-05 | 19 | NA | NA |
| ER to Golgi Anterograde Transport | 0.00017853 | 33 | 0.00655574 | 27 | NA | NA |
| B-WICH complex positively regulates rRNA expression | 0.00018563 | 23 | 2.50E-05 | 24 | NA | NA |
| RNA Polymerase I Promoter Escape | 0.00018563 | 23 | 2.50E-05 | 24 | NA | NA |
| Mitotic Prophase | 0.000207357 | 31 | 7.16E-05 | 31 | NA | NA |
| Pentose phosphate pathway | 0.000216613 | 8 | 0.001219647 | 7 | NA | NA |
| Recycling pathway of L1 | 0.000235207 | 15 | 0.015736693 | 11 | NA | NA |
| Glycosaminoglycan metabolism | 0.000252833 | 28 | 0.000246678 | 27 | NA | NA |
| Complement cascade | 0.000252833 | 17 | 1.22E-08 | 23 | NA | NA |
| Meiosis | 0.000266728 | 27 | 0.000103247 | 27 | NA | NA |
| Transcriptional regulation by small RNAs | 0.000293402 | 25 | 0.000120101 | 25 | NA | NA |
| Metabolism of fat-soluble vitamins | 0.000303638 | 15 | 0.006534901 | 12 | 0.019256708 | 5 |
| Ethanol oxidation | 0.000307735 | 7 | NA | NA | NA | NA |
| HDACs deacetylate histones | 0.000308893 | 23 | 0.000135438 | 23 | NA | NA |
| Defective B4GALT7 causes EDS, progeroid type | 0.000376111 | 9 | 0.001548229 | 8 | NA | NA |
| Defective B3GALT6 causes EDSP2 and SEMDJL1 | 0.000376111 | 9 | 0.001548229 | 8 | NA | NA |
| Translocation of SLC2A4 (GLUT4) to the plasma membrane | 0.00044238 | 19 | 0.001837812 | 17 | NA | NA |
| Glucose metabolism | 0.000528749 | 22 | 0.001648163 | 20 | NA | NA |
| Estrogen-dependent gene expression | 0.000582822 | 31 | 0.002213128 | 28 | NA | NA |
| Advanced glycosylation endproduct receptor signaling | 0.000588928 | 7 | 0.003254419 | 6 | NA | NA |
| RHOBTB GTPase Cycle | 0.000588928 | 12 | 0.001485548 | 11 | NA | NA |
| O-linked glycosylation | 0.000623949 | 25 | 0.001573624 | 23 | NA | NA |
| MyD88 deficiency (TLR2/4) | 0.000632962 | 8 | 0.014323589 | 6 | NA | NA |
| Intracellular signaling by second messengers | 0.000643979 | 53 | 1.82E-05 | 56 | NA | NA |
| Senescence-Associated Secretory Phenotype (SASP) | 0.000717819 | 25 | 0.000300099 | 25 | NA | NA |
| Removal of aminoterminal propeptides from gamma-carboxylated proteins | 0.000826653 | 6 | 0.000638339 | 6 | NA | NA |
| Chylomicron assembly | 0.000826653 | 6 | 0.000638339 | 6 | 0.010004681 | 3 |
| Chylomicron remodeling | 0.000826653 | 6 | 0.000638339 | 6 | 0.010004681 | 3 |
| HDL remodeling | 0.000826653 | 6 | 4.64E-05 | 7 | NA | NA |
| Regulation of Complement cascade | 0.000881679 | 14 | 2.97E-05 | 16 | NA | NA |
| Microtubule-dependent trafficking of connexons from Golgi to the plasma membrane | 0.001002007 | 8 | NA | NA | NA | NA |
| IRAK4 deficiency (TLR2/4) | 0.001002007 | 8 | 0.018949236 | 6 | NA | NA |
| Interleukin-4 and Interleukin-13 signaling | 0.00100795 | 24 | 0.005644107 | 21 | NA | NA |
| Attenuation phase | 0.001020949 | 7 | 0.005000985 | 6 | NA | NA |
| TGF-beta receptor signaling activates SMADs | 0.001043282 | 11 | 0.002675233 | 10 | NA | NA |
| Toll Like Receptor 4 (TLR4) Cascade | 0.00107709 | 28 | 0.000982359 | 27 | NA | NA |
| Reproduction | 0.001253369 | 29 | 0.001092839 | 28 | NA | NA |
| RHOBTB2 GTPase cycle | 0.001264327 | 9 | 0.000862559 | 9 | NA | NA |
| Plasma lipoprotein clearance | 0.001399953 | 11 | 0.003372905 | 10 | NA | NA |
| Transport of connexons to the plasma membrane | 0.001524045 | 8 | NA | NA | NA | NA |
| LDL clearance | 0.001524045 | 8 | 0.024470873 | 6 | NA | NA |

| | | | | | | |
|---|---|---|---|---|---|---|
| Collagen chain trimerization | 0.001556626 | 13 | 0.000920395 | 13 | NA | NA |
| Endosomal/Vacuolar pathway | 0.001571254 | 6 | 0.049623749 | 4 | NA | NA |
| Gamma-carboxylation, transport, and amino-terminal cleavage of proteins | 0.001571254 | 6 | 0.001196477 | 6 | NA | NA |
| Oxidative Stress Induced Senescence | 0.001640441 | 26 | 0.000691473 | 26 | NA | NA |
| Activation of BAD and translocation to mitochondria | 0.001658641 | 7 | 0.000156212 | 8 | NA | NA |
| GRB2:SOS provides linkage to MAPK signaling for Integrins | 0.001658641 | 7 | NA | NA | NA | NA |
| Platelet Adhesion to exposed collagen | 0.001658641 | 7 | 0.035312991 | 5 | 0.028806283 | 3 |
| HDMs demethylate histones | 0.001680419 | 14 | 0.00097685 | 14 | NA | NA |
| Platelet Aggregation (Plug Formation) | 0.001680419 | 12 | NA | NA | NA | NA |
| RHO GTPases activate PAKs | 0.001756173 | 9 | 3.37E-05 | 11 | NA | NA |
| Positive epigenetic regulation of rRNA expression | 0.0018227 | 23 | 0.000120101 | 25 | NA | NA |
| Telomere Maintenance | 0.0019187 | 24 | 0.000863046 | 24 | NA | NA |
| Antigen processing: Ubiquitination & Proteasome degradation | 0.001988697 | 51 | 0.007767537 | 46 | NA | NA |
| NoRC negatively regulates rRNA expression | 0.002078402 | 23 | 0.000959129 | 23 | NA | NA |
| Chondroitin sulfate biosynthesis | 0.002193527 | 8 | 0.031175551 | 6 | NA | NA |
| Antimicrobial peptides | 0.002420953 | 21 | 0.001168376 | 21 | NA | NA |
| Defects of contact activation system (CAS) and kallikrein/kinin system (KKS) | 0.002587482 | 7 | 0.001841868 | 7 | NA | NA |
| Diseases of hemostasis | 0.002587482 | 7 | 0.001841868 | 7 | NA | NA |
| HCMV Late Events | 0.002796947 | 24 | 0.001244581 | 24 | NA | NA |
| SUMOylation of chromatin organization proteins | 0.002997977 | 17 | 0.001573624 | 17 | NA | NA |
| Glycolysis | 0.002997977 | 17 | 0.001573624 | 17 | NA | NA |
| Negative epigenetic regulation of rRNA expression | 0.003038265 | 23 | 0.001402057 | 23 | NA | NA |
| RNA Polymerase I Promoter Clearance | 0.003038265 | 23 | 0.000587807 | 24 | NA | NA |
| Gene Silencing by RNA | 0.003272849 | 27 | 0.001381685 | 27 | NA | NA |
| Toll Like Receptor TLR1:TLR2 Cascade | 0.003300637 | 22 | 0.007962825 | 20 | NA | NA |
| Toll Like Receptor 2 (TLR2) Cascade | 0.003300637 | 22 | 0.007962825 | 20 | NA | NA |
| Gamma carboxylation, hypusine formation and arylsulfatase activation | 0.003340833 | 12 | 0.006679624 | 11 | NA | NA |
| HDR through Homologous Recombination (HRR) or Single Strand Annealing (SSA) | 0.003611063 | 26 | 0.001557407 | 26 | NA | NA |
| RNA Polymerase I Transcription | 0.003869433 | 23 | 0.000755818 | 24 | NA | NA |
| EPH-Ephrin signaling | 0.003872328 | 20 | 2.53E-06 | 26 | NA | NA |
| RHO GTPases activate IQGAPs | 0.004156637 | 10 | 0.002675233 | 10 | NA | NA |
| Infection with Mycobacterium tuberculosis | 0.004316415 | 9 | 0.011557951 | 8 | NA | NA |
| Erythrocytes take up carbon dioxide and release oxygen | 0.004316415 | 6 | 0.000436547 | 7 | NA | NA |
| O2/CO2 exchange in erythrocytes | 0.004316415 | 6 | 0.000436547 | 7 | NA | NA |
| Trafficking and processing of endosomal TLR | 0.004316415 | 6 | 0.019354134 | 5 | NA | NA |
| Keratan sulfate degradation | 0.004316415 | 6 | 4.05E-05 | 8 | NA | NA |
| Purine salvage | 0.004316415 | 6 | 0.019354134 | 5 | NA | NA |
| Chk1/Chk2(Cds1) mediated inactivation of Cyclin B:Cdk1 complex | 0.004316415 | 6 | 0.003254419 | 6 | NA | NA |
| Heparan sulfate/heparin (HS-GAG) metabolism | 0.004316415 | 14 | 0.045848681 | 11 | NA | NA |
| Post-translational modification: synthesis of GPI-anchored proteins | 0.004324987 | 20 | 0.011705452 | 18 | NA | NA |
| Selective autophagy | 0.005057682 | 18 | NA | NA | NA | NA |
| Activation of Matrix Metalloproteinases | 0.00523388 | 10 | 0.003372905 | 10 | NA | NA |
| Pre-NOTCH Processing in Golgi | 0.005498592 | 7 | 0.003974798 | 7 | NA | NA |
| Metabolism of Angiotensinogen to Angiotensins | 0.005498592 | 7 | NA | NA | NA | NA |
| Sulfur amino acid metabolism | 0.005658755 | 9 | NA | NA | NA | NA |
| Phase II - Conjugation of compounds | 0.005924181 | 22 | 0.026648914 | 19 | NA | NA |
| NCAM signaling for neurite out-growth | 0.005924181 | 15 | 0.00334773 | 15 | NA | NA |
| Keratan sulfate/keratin metabolism | 0.006615454 | 10 | 5.20E-05 | 13 | NA | NA |
| Homology Directed Repair | 0.00663756 | 26 | 0.002930495 | 26 | NA | NA |
| Gamma-carboxylation of protein precursors | 0.007221244 | 5 | 0.005603758 | 5 | NA | NA |
| Folding of actin by CCT/TriC | 0.007221244 | 5 | 4.64E-05 | 7 | NA | NA |
| Metabolism of vitamins and cofactors | 0.007226646 | 33 | 0.009694596 | 31 | NA | NA |
| Glycogen metabolism | 0.007284199 | 9 | 0.00496138 | 9 | NA | NA |
| Apoptotic execution phase | 0.007335592 | 13 | 0.00443201 | 13 | NA | NA |
| Assembly of active LPL and LIPC lipase complexes | 0.007676325 | 7 | 0.005557148 | 7 | NA | NA |
| Triglyceride catabolism | 0.00772085 | 8 | 0.000221681 | 10 | NA | NA |
| Semaphorin interactions | 0.008021306 | 15 | 0.001648163 | 16 | 0.002763594 | 7 |
| Signaling by Nuclear Receptors | 0.008040292 | 47 | 0.002470854 | 47 | NA | NA |
| Golgi-to-ER retrograde transport | 0.008087523 | 25 | NA | NA | NA | NA |
| GPVI-mediated activation cascade | 0.008120336 | 10 | 0.017683844 | 9 | NA | NA |
| Biological oxidations | 0.008241539 | 37 | NA | NA | NA | NA |
| trans-Golgi Network Vesicle Budding | 0.008646441 | 16 | NA | NA | NA | NA |
| Signaling by CSF3 (G-CSF) | 0.009190361 | 9 | 0.021558731 | 8 | NA | NA |
| Prolactin receptor signaling | 0.009696333 | 6 | 0.007312466 | 6 | NA | NA |
| p130Cas linkage to MAPK signaling for integrins | 0.009696333 | 6 | NA | NA | NA | NA |
| Activation of anterior HOX genes in hindbrain development during early embryogenesis | 0.010045639 | 23 | 0.004880256 | 23 | NA | NA |
| Activation of HOX genes during differentiation | 0.010045639 | 23 | 0.004880256 | 23 | NA | NA |
| Receptor-type tyrosine-protein phosphatases | 0.010350013 | 7 | 0.007560886 | 7 | NA | NA |
| NCAM1 interactions | 0.010350013 | 11 | 0.006679624 | 11 | NA | NA |
| MyD88:MAL(TIRAP) cascade initiated on plasma membrane | 0.011176935 | 20 | 0.025834797 | 18 | NA | NA |
| Toll Like Receptor TLR6:TLR2 Cascade | 0.011176935 | 20 | 0.025834797 | 18 | NA | NA |
| Dermatan sulfate biosynthesis | 0.011393367 | 5 | 0.008959336 | 5 | NA | NA |
| DNA Double-Strand Break Repair | 0.012445536 | 29 | 0.010202569 | 28 | NA | NA |
| Downregulation of TGF-beta receptor signaling | 0.012885256 | 8 | NA | NA | NA | NA |
| Nuclear Envelope (NE) Reassembly | 0.012885256 | 16 | NA | NA | NA | NA |
| Glycogen synthesis | 0.013695241 | 6 | NA | NA | NA | NA |
| Signal regulatory protein family interactions | 0.013695241 | 6 | 0.001841868 | 7 | NA | NA |
| Signal transduction by L1 | 0.013784594 | 7 | NA | NA | 0.008699543 | 4 |
| ESR-mediated signaling | 0.015324819 | 36 | 0.003254419 | 37 | NA | NA |
| Metabolism of porphyrins | 0.016392045 | 8 | NA | NA | NA | NA |
| Integrin signaling | 0.016392045 | 8 | NA | NA | NA | NA |
| Caspase-mediated cleavage of cytoskeletal proteins | 0.017245803 | 5 | NA | NA | NA | NA |
| HSF1 activation | 0.017245803 | 5 | 0.013662988 | 5 | NA | NA |
| GP1b-IX-V activation signalling | 0.017245803 | 5 | NA | NA | NA | NA |
| Purine ribonucleoside monophosphate biosynthesis | 0.017245803 | 5 | 0.002007169 | 6 | NA | NA |
| TAK1 activates NFkB by phosphorylation and activation of IKKs complex | 0.017495455 | 9 | 0.03665047 | 8 | NA | NA |
| Intra-Golgi and retrograde Golgi-to-ER traffic | 0.0176637 | 33 | NA | NA | NA | NA |
| Signaling by ERBB2 TMD/JMD mutants | 0.017941042 | 7 | 0.047323229 | 6 | NA | NA |
| PKMTs methylate histone lysines | 0.017966218 | 15 | 0.010645707 | 15 | NA | NA |
| Glycosphingolipid metabolism | 0.020745945 | 11 | 0.035510466 | 10 | NA | NA |
| Autophagy | 0.020898154 | 26 | NA | NA | NA | NA |
| RMTs methylate histone arginines | 0.020957759 | 16 | 0.012080994 | 16 | NA | NA |
| Nucleotide salvage | 0.023154118 | 7 | 0.016859694 | 7 | NA | NA |

| | | | | | | |
|---|---|---|---|---|---|---|
| RHOBTB1 GTPase cycle | 0.023154118 | 7 | 0.016859694 | 7 | NA | NA |
| Activation of AMPK downstream of NMDARs | 0.025332545 | 8 | NA | NA | NA | NA |
| Signaling by TGFB family members | 0.02595621 | 19 | NA | NA | NA | NA |
| Regulation of PLK1 Activity at G2/M Transition | 0.026510185 | 17 | NA | NA | NA | NA |
| HSF1-dependent transactivation | 0.029516368 | 7 | 0.00542735 | 8 | NA | NA |
| Activation of BH3-only proteins | 0.031072616 | 8 | 0.00628775 | 9 | NA | NA |
| EPHA-mediated growth cone collapse | 0.031072616 | 8 | NA | NA | 0.023394658 | 4 |
| Chromosome Maintenance | 0.03153092 | 24 | 0.015672058 | 24 | NA | NA |
| Ephrin signaling | 0.033017058 | 6 | 0.005557148 | 7 | NA | NA |
| Plasma lipoprotein assembly | 0.033017058 | 6 | 0.000161907 | 9 | 0.048999353 | 3 |
| Methylation | 0.034678478 | 5 | 0.026460847 | 5 | NA | NA |
| Heme biosynthesis | 0.034678478 | 5 | NA | NA | NA | NA |
| Golgi Associated Vesicle Biogenesis | 0.034768198 | 12 | NA | NA | NA | NA |
| IKK complex recruitment mediated by RIP1 | 0.036265016 | 7 | 0.00706967 | 8 | NA | NA |
| Signaling by ERBB2 KD Mutants | 0.036265016 | 7 | NA | NA | NA | NA |
| Inactivation of CSF3 (G-CSF) signaling | 0.036265016 | 7 | NA | NA | NA | NA |
| Antigen Presentation: Folding, assembly and peptide loading of class I MHC | 0.036265016 | 7 | NA | NA | NA | NA |
| Diseases of Immune System | 0.036819132 | 8 | NA | NA | NA | NA |
| Uptake and actions of bacterial toxins | 0.036819132 | 8 | 0.007844962 | 9 | NA | NA |
| Diseases associated with the TLR signaling cascade | 0.036819132 | 8 | NA | NA | NA | NA |
| Sealing of the nuclear envelope (NE) by ESCRT-III | 0.036819132 | 8 | NA | NA | NA | NA |
| Cell-Cell communication | 0.039665264 | 22 | 0.001117136 | 26 | NA | NA |
| Apoptotic cleavage of cellular proteins | 0.042669498 | 9 | 0.02915956 | 9 | NA | NA |
| DNA Damage Recognition in GG-NER | 0.042669498 | 9 | 0.02915956 | 9 | NA | NA |
| EPH-ephrin mediated repulsion of cells | 0.042769695 | 11 | 0.010670328 | 12 | NA | NA |
| Signaling by ERBB2 in Cancer | 0.044033774 | 7 | NA | NA | NA | NA |
| Purinergic signaling in leishmaniasis infection | 0.044033774 | 7 | NA | NA | NA | NA |
| Cell recruitment (pro-inflammatory response) | 0.044033774 | 7 | NA | NA | NA | NA |
| Iron uptake and transport | 0.044033774 | 12 | 0.011449431 | 13 | NA | NA |
| Urea cycle | 0.044990414 | 4 | NA | NA | NA | NA |
| Signaling by NOTCH1 HD Domain Mutants in Cancer | 0.045086085 | 5 | 0.035312991 | 5 | NA | NA |
| Constitutive Signaling by NOTCH1 HD Domain Mutants | 0.045086085 | 5 | 0.035312991 | 5 | NA | NA |
| ERBB2 Regulates Cell Motility | 0.045086085 | 5 | NA | NA | NA | NA |
| Nucleobase biosynthesis | 0.045086085 | 5 | 0.007312466 | 6 | NA | NA |
| Recruitment of NuMA to mitotic centrosomes | 0.046770436 | 17 | NA | NA | NA | NA |
| TP53 Regulates Metabolic Genes | 0.047968599 | 16 | NA | NA | NA | NA |
| Signaling by TGF-beta Receptor Complex | 0.049572079 | 14 | NA | NA | NA | NA |
| Smooth Muscle Contraction | NA | NA | 1.53E-05 | 15 | 0.010004681 | 5 |
| Initial triggering of complement | NA | NA | 2.09E-05 | 11 | NA | NA |
| DARPP-32 events | NA | NA | 0.000473668 | 10 | NA | NA |
| Striated Muscle Contraction | NA | NA | 0.000480462 | 12 | NA | NA |
| EPHB-mediated forward signaling | NA | NA | 0.000582218 | 13 | NA | NA |
| Calcineurin activates NFAT | NA | NA | 0.001196477 | 6 | NA | NA |
| eNOS activation | NA | NA | 0.003254419 | 6 | NA | NA |
| Adherens junctions interactions | NA | NA | 0.003372905 | 10 | NA | NA |
| Keratan sulfate biosynthesis | NA | NA | 0.003811283 | 9 | NA | NA |
| Creation of C4 and C2 activators | NA | NA | 0.005000985 | 6 | NA | NA |
| CREB1 phosphorylation through the activation of Adenylate Cyclase | NA | NA | 0.005000985 | 6 | NA | NA |
| Fcgamma receptor (FCGR) dependent phagocytosis | NA | NA | 0.005139349 | 18 | NA | NA |
| Regulation of actin dynamics for phagocytic cup formation | NA | NA | 0.006691711 | 14 | NA | NA |
| Triglyceride metabolism | NA | NA | 0.008044861 | 10 | NA | NA |
| CaMK IV-mediated phosphorylation of CREB | NA | NA | 0.013662988 | 5 | NA | NA |
| Metabolism of nitric oxide: NOS3 activation and regulation | NA | NA | 0.014323589 | 6 | NA | NA |
| Signaling by BRAF and RAF fusions | NA | NA | 0.015611743 | 14 | NA | NA |
| Epigenetic regulation of gene expression | NA | NA | 0.015736693 | 25 | NA | NA |
| Signaling by moderate kinase activity BRAF mutants | NA | NA | 0.015736693 | 11 | NA | NA |
| Signaling by RAS mutants | NA | NA | 0.015736693 | 11 | NA | NA |
| Paradoxical activation of RAF signaling by kinase inactive BRAF | NA | NA | 0.015736693 | 11 | NA | NA |
| Signaling downstream of RAS mutants | NA | NA | 0.015736693 | 11 | NA | NA |
| HATs acetylate histones | NA | NA | 0.01684925 | 24 | NA | NA |
| Downstream signal transduction | NA | NA | 0.017806081 | 8 | NA | NA |
| Crosslinking of collagen fibrils | NA | NA | 0.018949236 | 6 | NA | NA |
| Uptake and function of anthrax toxins | NA | NA | 0.019354134 | 5 | NA | NA |
| Cell junction organization | NA | NA | 0.020432009 | 17 | NA | NA |
| RHO GTPases Activate WASPs and WAVEs | NA | NA | 0.021079591 | 9 | NA | NA |
| RAF activation | NA | NA | 0.021079591 | 9 | NA | NA |
| Cytosolic tRNA aminoacylation | NA | NA | 0.02121053 | 7 | NA | NA |
| MyD88-independent TLR4 cascade | NA | NA | 0.021474363 | 18 | NA | NA |
| TRIF(TICAM1)-mediated TLR4 signaling | NA | NA | 0.021474363 | 18 | NA | NA |
| Clathrin-mediated endocytosis | NA | NA | 0.022928951 | 24 | NA | NA |
| Signaling by RAF1 mutants | NA | NA | 0.023135898 | 10 | NA | NA |
| Protein methylation | NA | NA | 0.024470873 | 6 | NA | NA |
| Calmodulin induced events | NA | NA | 0.02479162 | 9 | NA | NA |
| CaM pathway | NA | NA | 0.02479162 | 9 | NA | NA |
| Activated NOTCH1 Transmits Signal to the Nucleus | NA | NA | 0.025961493 | 8 | NA | NA |
| Cellular Senescence | NA | NA | 0.027033861 | 30 | NA | NA |
| PKA activation | NA | NA | 0.031175551 | 6 | NA | NA |
| Parasite infection | NA | NA | 0.031816869 | 12 | NA | NA |
| Leishmania phagocytosis | NA | NA | 0.031816869 | 12 | NA | NA |
| FCGR3A-mediated phagocytosis | NA | NA | 0.031816869 | 12 | NA | NA |
| Ca-dependent events | NA | NA | 0.034214398 | 9 | NA | NA |
| Anchoring fibril formation | NA | NA | 0.035312991 | 5 | NA | NA |
| CREB1 phosphorylation through the activation of CaMKII/CaMKK/CaMKIV cascasde | NA | NA | 0.036049905 | 4 | NA | NA |
| STAT5 activation downstream of FLT3 ITD mutants | NA | NA | 0.036049905 | 4 | NA | NA |
| VEGFR2 mediated cell proliferation | NA | NA | 0.038871267 | 6 | NA | NA |
| PI5P, PP2A and IER3 Regulate PI3K/AKT Signaling | NA | NA | 0.039986865 | 18 | NA | NA |
| TRAF6 mediated induction of NFkB and MAP kinases upon TLR7/8 or 9 activation | NA | NA | 0.041540161 | 16 | NA | NA |
| VEGFA-VEGFR2 Pathway | NA | NA | 0.042747921 | 17 | NA | NA |
| Oncogenic MAPK signaling | NA | NA | 0.043998945 | 15 | NA | NA |
| Muscle contraction | NA | NA | 0.045104075 | 29 | NA | NA |
| MyD88 dependent cascade initiated on endosome | NA | NA | 0.045473832 | 16 | NA | NA |
| Spry regulation of FGF signaling | NA | NA | 0.045764395 | 5 | NA | NA |
| Sema3A PAK dependent Axon repulsion | NA | NA | 0.045764395 | 5 | NA | NA |

| | | | | | | |
|---|---|---|---|---|---|---|
| Signaling by ERBB2 ECD mutants | NA | NA | 0.045764395 | 5 | NA | NA |
| Interleukin-3, Interleukin-5 and GM-CSF signaling | NA | NA | 0.046079633 | 10 | NA | NA |
| Translation of structural proteins | NA | NA | 0.046096413 | 7 | NA | NA |
| Signaling by VEGF | NA | NA | 0.04667717 | 18 | NA | NA |
| PKA-mediated phosphorylation of CREB | NA | NA | 0.047323229 | 6 | NA | NA |
| NOTCH2 Activation and Transmission of Signal to the Nucleus | NA | NA | 0.047323229 | 6 | NA | NA |
| Toll Like Receptor 7/8 (TLR7/8) Cascade | NA | NA | 0.048861225 | 16 | NA | NA |
| Regulation of localization of FOXO transcription factors | NA | NA | 0.049623749 | 4 | NA | NA |
| Constitutive Signaling by Overexpressed ERBB2 | NA | NA | 0.049623749 | 4 | NA | NA |
| N-glycan trimming in the ER and Calnexin/Calreticulin cycle | NA | NA | 0.049623749 | 8 | NA | NA |
| Sema4D induced cell migration and growth-cone collapse | NA | NA | NA | NA | 0.000823938 | 5 |
| Sema4D in semaphorin signaling | NA | NA | NA | NA | 0.001896423 | 5 |
| PECAM1 interactions | NA | NA | NA | NA | 0.016233959 | 3 |
| Metabolic disorders of biological oxidation enzymes | NA | NA | NA | NA | 0.03563081 | 4 |
| Other semaphorin interactions | NA | NA | NA | NA | 0.048999353 | 3 |

### Supplemental Table III:

| | Metabolite | N_HC | N_Mild | N_Severe | Mean_HC | Mean_Mild | Mean_Severe | Log2FC_MvsHC | Log2FC_SvsHC | Log2FC_SvsM | Adj_Pvalue_MvsHC | Adj_Pvalue_SvsHC | Adj_Pvalue_SvsM |
|---|---|---|---|---|---|---|---|---|---|---|---|---|---|
| 1 | 1,5-anhydroglucitol* | 39 | 74 | 33 | 2.258717 | 2.232249644 | 1.100972136 | -0.026467215 | -1.157744723 | -1.131277508 | 0.990340108 | 9.24E-06 | 9.67E-07 |
| 2 | 1H-pyrrole-2,5-dione* | 39 | 74 | 33 | 3.569952 | 3.157935613 | 2.818393171 | -0.412016615 | -0.751559057 | -0.339542442 | 0.342165897 | 0.085338469 | 0.520074093 |
| 3 | 2,4-dihydroxybutanoic acid* | 39 | 74 | 33 | -3.53612 | -3.514166238 | -3.62395165 | 0.021949489 | -0.087835923 | -0.109785412 | 0.976275601 | 0.764607687 | 0.586367931 |
| 4 | 2-aminobutanoic acid | 39 | 74 | 33 | 0.865915 | 0.684656532 | 0.495757932 | -0.181258372 | -0.370156972 | -0.1888986 | 0.707386211 | 0.366665795 | 0.714615413 |
| 5 | 2-aminomalonic acid* | 39 | 74 | 33 | -2.74367 | -2.835537866 | -2.786744639 | -0.091869762 | -0.043076535 | 0.048793227 | 0.883322549 | 0.981069368 | 0.969180576 |
| 6 | 2-hydroxybutyric acid | 39 | 74 | 33 | -0.98074 | -0.899189477 | -0.699341749 | 0.081554667 | 0.281402395 | 0.199847728 | 0.842062382 | 0.243307625 | 0.400498211 |
| 7 | 2-hydroxyisopentanoic acid* | 39 | 74 | 33 | 0.965316 | 1.014410903 | 0.952162693 | 0.04909462 | -0.01315359 | -0.06224821 | 0.88026827 | 0.993605071 | 0.832689392 |
| 8 | 2-ketoisocaproic acid | 39 | 74 | 33 | -2.3118 | -2.197012565 | -2.542510069 | 0.114787908 | -0.230709595 | -0.345497504 | 0.448443292 | 0.107182405 | 0.002141952 |
| 9 | 3,4-dihydroxybutanoic acid* | 39 | 74 | 33 | -3.22469 | -3.309905283 | -2.764042942 | -0.085211429 | 0.460650913 | 0.545862341 | 0.854058771 | 0.043854951 | 0.004192373 |
| 10 | 3-aminopiperidine-2,6-dione* | 39 | 74 | 33 | 4.379419 | 4.443755751 | 3.933498653 | 0.06433663 | -0.445920469 | -0.510257098 | 0.798289449 | 0.000856348 | 1.18E-05 |
| 11 | 5-methylhydantoin* | 39 | 74 | 33 | -0.5709 | -0.562861429 | -0.873622542 | 0.008039145 | -0.302721967 | -0.310761112 | 0.996610301 | 0.038555346 | 0.013132951 |
| 12 | aconitic acid | 39 | 74 | 33 | -0.91178 | -0.76602275 | -1.023482957 | 0.145760602 | -0.111699605 | -0.257460208 | 0.547399069 | 0.779881244 | 0.189936174 |
| 13 | adipic acid (contaminated?) | 39 | 74 | 33 | -1.68085 | -1.523894823 | -1.630778038 | 0.156950418 | 0.050067202 | -0.106883216 | 0.603326527 | 0.964475122 | 0.810524572 |
| 14 | alpha-tocopherol | 39 | 74 | 33 | 2.330629 | 1.411284926 | 1.469777338 | -0.919343904 | -0.860851492 | 0.058492412 | 0.000166724 | 0.004065037 | 0.966234413 |
| 15 | benzoic acid | 39 | 74 | 33 | 0.707619 | 0.634729448 | 0.504074364 | -0.072889592 | -0.203544676 | -0.130655084 | 0.699129537 | 0.146444137 | 0.360463572 |
| 16 | beta-alanine | 39 | 74 | 33 | -2.17937 | -1.959309062 | -1.726042243 | 0.220063197 | 0.453330015 | 0.233266819 | 0.554920641 | 0.177638227 | 0.553607183 |
| 17 | beta-sitosterol | 39 | 72 | 33 | -2.44424 | -2.58426056 | -1.939475214 | -0.140023089 | 0.504762257 | 0.644785347 | 0.785113406 | 0.113533508 | 0.012376256 |
| 18 | capric acid | 39 | 74 | 33 | -2.14626 | -2.024212206 | -1.287509461 | 0.122048343 | 0.858751089 | 0.736702745 | 0.871368622 | 0.010509258 | 0.01367807 |
| 19 | carbonate ion | 39 | 74 | 33 | 3.871691 | 3.787083072 | 3.690215955 | -0.084608048 | -0.181475165 | -0.096867117 | 0.505836486 | 0.115294643 | 0.450540763 |
| 20 | cholest-7-en-3-ol* | 39 | 74 | 33 | -1.99293 | -1.971140897 | -1.818431591 | 0.021785553 | 0.174494859 | 0.152709306 | 0.993898161 | 0.760316807 | 0.764927791 |
| 21 | cholestan-3-ol* | 39 | 74 | 33 | -1.90192 | -1.826180612 | -0.955989235 | 0.075738499 | 0.945929876 | 0.870191377 | 0.948514721 | 0.004271694 | 0.002820436 |
| 22 | cholesterol | 39 | 74 | 33 | 6.376818 | 6.368245588 | 6.408369167 | -0.008572351 | 0.031551229 | 0.040123579 | 0.989830727 | 0.907720495 | 0.818984244 |
| 23 | citric acid | 39 | 74 | 33 | 2.332407 | 2.34428339 | 2.093860763 | 0.01187674 | -0.238545886 | -0.250422626 | 0.994800887 | 0.23478325 | 0.131785127 |
| 24 | creatinine | 39 | 74 | 33 | 0.065332 | -0.085225044 | 0.131593897 | -0.150557543 | 0.066261398 | 0.216818941 | 0.621071115 | 0.937199709 | 0.415237897 |
| 25 | dehydroalanine | 39 | 74 | 33 | -2.27595 | -2.005150548 | -1.64357635 | 0.270798909 | 0.632373107 | 0.361574198 | 0.154143071 | 0.001124478 | 0.052569536 |
| 26 | D-fructose | 39 | 74 | 33 | 2.364601 | 2.314893584 | 2.433092914 | -0.049707507 | 0.068491823 | 0.11819933 | 0.946752416 | 0.929884437 | 0.758954403 |
| 27 | D-gluconic acid | 39 | 74 | 33 | -2.94591 | -2.784881561 | -2.013864814 | 0.161031343 | 0.93204809 | 0.771016746 | 0.682544335 | 0.000256498 | 0.000681116 |
| 28 | D-glucose | 39 | 74 | 33 | 7.982834 | 8.035088871 | 7.885109589 | 0.052255208 | -0.097724074 | -0.149979282 | 0.766441079 | 0.522871662 | 0.146069103 |
| 29 | D-glucuronic acid | 39 | 74 | 33 | -2.31403 | -2.222406887 | -1.282496169 | 0.091623856 | 1.031534573 | 0.939910718 | 0.904665079 | 0.00027803 | 0.000176501 |
| 30 | D-mannose | 39 | 74 | 33 | 3.170393 | 3.289055416 | 3.65188024 | 0.118662801 | 0.481487625 | 0.362824824 | 0.644781408 | 0.007979228 | 0.028776106 |
| 31 | D-ribitol | 39 | 74 | 33 | -3.2467 | -3.090229308 | -2.899399801 | 0.156465887 | 0.347295394 | 0.190829507 | 0.362822049 | 0.033067777 | 0.261089302 |
| 32 | D-sorbitol | 39 | 74 | 33 | 0.427151 | 0.401638501 | 1.326221849 | -0.025512863 | 0.899070486 | 0.924583349 | 0.992762203 | 0.002637665 | 0.000382294 |
| 33 | D-xylitol | 39 | 74 | 33 | -1.51925 | -1.417680569 | -1.627852353 | 0.101568206 | -0.108603578 | -0.210171784 | 0.833218283 | 0.864103563 | 0.499427579 |
| 34 | dopamine | 39 | 74 | 33 | -1.45695 | -1.468079798 | -1.684221496 | -0.011125806 | -0.227267504 | -0.216141698 | 0.994127744 | 0.184668805 | 0.14292145 |
| 35 | EDTA [major] | 39 | 74 | 33 | 8.606051 | 8.530070058 | 8.278870439 | -0.075981044 | -0.327180664 | -0.251199619 | 0.769372578 | 0.037061021 | 0.082040816 |
| 36 | EDTA [minor] | 39 | 74 | 33 | 2.670986 | 2.704961756 | 2.666024342 | 0.033976177 | -0.004961237 | -0.038937413 | 0.944168539 | 0.999142499 | 0.93480426 |
| 37 | erythritol | 39 | 74 | 33 | 0.100895 | -0.04185635 | 0.188420365 | -0.142751661 | 0.087525054 | 0.230276715 | 0.841537381 | 0.955562607 | 0.67019497 |
| 38 | erythronic acid | 39 | 74 | 33 | -0.28 | -0.394577605 | 0.228529106 | -0.114579522 | 0.508527188 | 0.623106711 | 0.681796624 | 0.006529867 | 9.52E-05 |
| 39 | fumaric acid | 39 | 74 | 33 | 0.11458 | 0.052985812 | 0.433868765 | -0.061594024 | 0.319288929 | 0.380882953 | 0.900354536 | 0.144307213 | 0.031372377 |
| 40 | glyceric acid | 39 | 74 | 33 | -0.15249 | -1.269282945 | -1.3023105 | -1.116790668 | -1.149818223 | -0.033027555 | 7.23E-12 | 2.15E-09 | 0.974967897 |
| 41 | glycerol | 39 | 74 | 33 | 5.019067 | 4.96825257 | 4.956532895 | -0.050814363 | -0.062534039 | -0.011719676 | 0.896692304 | 0.890856301 | 0.994824385 |
| 42 | glycerol 3-phosphate | 39 | 74 | 33 | 0.49184 | 0.554680649 | 0.416532567 | 0.062840279 | -0.075307803 | -0.138148082 | 0.575491326 | 0.573865036 | 0.096199881 |
| 43 | glycine | 39 | 74 | 33 | 4.897205 | 4.953353077 | 4.715128227 | 0.056148079 | -0.182076772 | -0.238224851 | 0.834632213 | 0.268667265 | 0.05917539 |
| 44 | glycolic acid | 39 | 74 | 33 | -0.95133 | -0.992351747 | -1.067793768 | -0.041022605 | -0.116464627 | -0.075442021 | 0.702293075 | 0.140897359 | 0.346464459 |
| 45 | heptadecanoic acid | 39 | 74 | 33 | 0.8576 | 0.818743437 | 0.599534064 | -0.03885698 | -0.258066353 | -0.219209373 | 0.917211445 | 0.074462376 | 0.090911918 |
| 46 | hypoxanthine | 39 | 74 | 33 | 0.167798 | 1.337003646 | 1.450483345 | 1.169205636 | 1.282685335 | 0.1134797 | 7.12E-05 | 0.000288464 | 0.915392784 |
| 47 | inosine | 39 | 74 | 33 | -4.57783 | -3.459710848 | -3.836039772 | 1.118120577 | 0.741791652 | -0.376328924 | 0.007655366 | 0.211588054 | 0.596861828 |
| 48 | inositol phosphate | 39 | 74 | 33 | -2.02445 | -2.047156842 | -2.540455572 | -0.02270192 | -0.51600065 | -0.49329873 | 0.981037796 | 0.001538748 | 0.000561681 |
| 52 | internal standard - creatinine-d3 | 39 | 74 | 33 | 0.127152 | -0.397612959 | -0.380825668 | -0.524765166 | -0.507977876 | 0.016787291 | 0.061550425 | 0.157792718 | 0.997377342 |
| 58 | internal standard - tryptophan -d3 | 39 | 74 | 33 | 10.31714 | 10.23395924 | 10.03918339 | -0.083177384 | -0.277953238 | -0.194775855 | 0.277944154 | 9.67E-05 | 0.002530497 |
| 59 | internal standard - tyrosine 13C6 | 39 | 74 | 33 | 1.644392 | 1.544808034 | 1.593218101 | -0.099584258 | -0.051174191 | 0.048410067 | 0.297608929 | 0.797412004 | 0.772154555 |
| 60 | lactic acid | 39 | 74 | 33 | 6.856348 | 7.227829257 | 7.434542889 | 0.371481431 | 0.578195064 | 0.206713632 | 0.000510894 | 4.55E-06 | 0.1093796 |
| 61 | L-alanine | 39 | 74 | 33 | 4.861541 | 4.89860976 | 4.677545629 | 0.037068676 | -0.183995456 | -0.221064131 | 0.95755244 | 0.476014184 | 0.256887118 |
| 62 | L-aspartic acid | 39 | 74 | 33 | 2.541342 | 2.293359106 | 2.279315588 | -0.247983098 | -0.262026617 | -0.014043518 | 0.064038606 | 0.115242533 | 0.991923814 |
| 63 | lauric acid | 39 | 74 | 33 | -0.35774 | -0.559024109 | -0.914942366 | -0.201284246 | -0.557202502 | -0.355918257 | 0.42901726 | 0.012587542 | 0.097528556 |
| 64 | L-cysteine | 39 | 74 | 33 | 1.687302 | 1.969798743 | 2.088693118 | 0.282496498 | 0.401390873 | 0.118894375 | 0.003649661 | 0.000423418 | 0.393637326 |
| 65 | L-cystine | 39 | 74 | 33 | -4.4104 | 0.159927892 | 1.655743949 | 4.570332793 | 6.06614885 | 1.495816057 | 0 | 0 | 5.20E-07 |
| 66 | L-glutamic acid | 39 | 74 | 33 | 2.098891 | 1.631444472 | 2.119978613 | -0.467446555 | 0.021087586 | 0.488534141 | 0.007784742 | 0.992751129 | 0.00869898 |
| 67 | L-glutamine | 39 | 74 | 33 | 0.749781 | 1.310597489 | 1.39495025 | 0.560816793 | 0.64516974 | 0.084352761 | 0.177360852 | 0.201058071 | 0.965020092 |
| 68 | linoleic acid | 39 | 74 | 33 | 0.888911 | 0.713000799 | 0.788522456 | -0.175909777 | -0.100388119 | 0.075521658 | 0.639497229 | 0.902771942 | 0.928733451 |
| 69 | L-isoleucine | 39 | 74 | 33 | 3.56852 | 3.565730027 | 3.520874776 | -0.002789801 | -0.047645053 | -0.044855251 | 0.999707815 | 0.942126865 | 0.934767804 |
| 70 | L-leucine | 39 | 74 | 33 | 4.165779 | 4.143353326 | 3.812494364 | -0.022425493 | -0.353284455 | -0.330858963 | 0.991871843 | 0.247438094 | 0.209879982 |
| 71 | L-lysine | 39 | 74 | 33 | 3.778293 | 3.826268014 | 4.00763541 | 0.047975184 | 0.22934258 | 0.181367396 | 0.894823592 | 0.174375533 | 0.246666135 |
| 72 | L-malic acid | 39 | 74 | 33 | -1.5561 | -1.369814995 | -0.995851565 | 0.186284725 | 0.560248155 | 0.37396343 | 0.299113523 | 0.000753351 | 0.014805825 |
| 73 | L-methionine | 39 | 74 | 33 | 0.798596 | 0.740628887 | 0.725925127 | -0.057967251 | -0.072671011 | -0.01470376 | 0.906570916 | 0.897734172 | 0.994370709 |
| 74 | L-ornithine | 39 | 74 | 33 | 3.778267 | 3.69627731 | 3.917014117 | -0.081990082 | 0.138746725 | 0.220736807 | 0.697658513 | 0.487340168 | 0.101816506 |
| 75 | L-phenylalanine | 39 | 74 | 33 | 2.027119 | 2.047518848 | 1.925752636 | 0.020400227 | -0.101365984 | -0.121766211 | 0.994617356 | 0.911034853 | 0.842384723 |
| 76 | L-proline | 39 | 74 | 33 | 4.207605 | 4.069263648 | 4.09310557 | -0.138340898 | -0.114498976 | 0.023841922 | 0.66939975 | 0.824590945 | 0.989358734 |
| 77 | L-pyroglutamic acid | 39 | 74 | 33 | 4.330487 | 4.349493422 | 4.41396617 | 0.019006298 | 0.083479046 | 0.064472748 | 0.96159856 | 0.591010224 | 0.669603234 |
| 78 | L-serine | 39 | 74 | 33 | 3.031949 | 2.935482762 | 2.648483913 | -0.096466356 | -0.383465205 | -0.286998849 | 0.679170038 | 0.016329276 | 0.050927226 |
| 79 | L-threonine | 39 | 74 | 33 | 3.179093 | 3.185276024 | 3.03687539 | 0.006183342 | -0.142217291 | -0.148400634 | 0.997223957 | 0.361307796 | 0.244294519 |
| 80 | L-tryptophan | 39 | 74 | 33 | 6.442334 | 6.41538639 | 6.104576801 | -0.026947181 | -0.33775677 | -0.310809589 | 0.924820287 | 0.000358454 | 0.000197561 |
| 81 | L-tyrosine | 39 | 74 | 33 | 1.787962 | 1.863698363 | 2.068091137 | 0.075736737 | 0.280129512 | 0.204392775 | 0.817423802 | 0.150225837 | 0.273435916 |
| 82 | L-valine | 39 | 74 | 33 | 5.215604 | 5.228520006 | 5.111846762 | 0.012916258 | -0.103756986 | -0.116673244 | 0.989848077 | 0.632163564 | 0.477987971 |
| 83 | maltose | 39 | 74 | 33 | -1.56724 | -1.448002051 | -0.724807312 | 0.119242567 | 0.842437306 | 0.723194739 | 0.820646982 | 0.001543856 | 0.002214983 |
| 84 | myristic acid | 39 | 74 | 33 | 4.856915 | 4.329056297 | 4.20424188 | -0.527858369 | -0.652672787 | -0.124814417 | 2.76E-11 | 6.81E-12 | 0.224238287 |
| 85 | nonanoic acid | 39 | 74 | 33 | 1.414495 | 1.350085583 | 1.131794578 | -0.06440976 | -0.282700765 | -0.218291005 | 0.812385093 | 0.065382565 | 0.123661281 |
| 86 | oleic acid | 39 | 74 | 33 | 2.258491 | 2.129187466 | 2.228825087 | -0.129303043 | -0.029665423 | 0.099637621 | 0.84117488 | 0.993637215 | 0.912239791 |
| 87 | O-phosphocolamine | 39 | 74 | 33 | -0.20918 | -0.093950485 | -0.0943086 | 0.115227404 | 0.114869289 | -0.000358115 | 0.590689068 | 0.692939993 | 0.99999543 |
| 88 | oxalic acid | 39 | 74 | 33 | -0.60866 | -0.732956593 | -0.719635345 | -0.12429293 | -0.110971681 | 0.013321249 | 0.633294896 | 0.77459328 | 0.995298994 |
| 89 | palmitic acid | 39 | 74 | 33 | 1.207671 | 0.911010128 | 0.863469426 | -0.296660466 | -0.344201168 | -0.047540702 | 0.003260413 | 0.004464429 | 0.870031237 |
| 90 | phosphate ion | 39 | 74 | 33 | 7.254326 | 7.281284712 | 7.044272547 | 0.026958492 | -0.210053674 | -0.237012165 | 0.884710771 | 0.007086985 | 0.000400354 |
| 91 | pipecolic acid | 39 | 74 | 33 | -2.48554 | -2.096760896 | -1.818360921 | 0.388777751 | 0.667177725 | 0.278399975 | 0.224420714 | 0.048283555 | 0.501274326 |
| 92 | pseudouridine | 39 | 74 | 33 | -2.51293 | -2.37070842 | -2.202968657 | 0.142220193 | 0.309959956 | 0.167739763 | 0.162243985 | 0.003011915 | 0.105350594 |
| 93 | pyrophosphate | 39 | 74 | 33 | -1.09599 | -1.141913963 | -1.399887678 | -0.045920216 | -0.303893931 | -0.257973715 | 0.944112278 | 0.177024376 | 0.202832548 |
| 94 | pyrrole -2-carboxylic acid | 39 | 74 | 33 | -4.38165 | -4.075317296 | -3.898889197 | 0.306331254 | 0.482759352 | 0.176428099 | 0.30207451 | 0.127220431 | 0.699042914 |
| 95 | pyruvic acid | 39 | 74 | 33 | 1.378261 | 1.665836574 | 1.649717559 | 0.287575473 | 0.271456458 | -0.016119015 | 0.360795269 | 0.528158394 | 0.997110541 |
| 96 | sedoheptulose | 39 | 74 | 33 | 4.418601 | 4.412133951 | 4.237585947 | -0.00646749 | -0.181015495 | -0.174548004 | 0.999220415 | 0.653365638 | 0.603595743 |
| 97 | S-methylcysteine* | 39 | 74 | 33 | -0.64542 | -0.64668391 | -0.709962399 | -0.001268864 | -0.064547353 | -0.063278489 | 0.999957993 | 0.926797004 | 0.910951467 |
| 98 | stearic acid | 39 | 74 | 33 | 5.117182 | 4.898514975 | 4.827230748 | -0.218667066 | -0.289951294 | -0.071284228 | 0.016537897 | 0.006721384 | 0.667017033 |
| 99 | succinic acid | 39 | 74 | 33 | -0.9171 | -1.025636135 | -0.727779533 | -0.108538352 | 0.18931825 | 0.297856602 | 0.816627439 | 0.650323717 | 0.260126714 |
| 100 | tagatose | 39 | 74 | 33 | 0.946803 | 0.904709086 | 1.138568874 | -0.042094077 | 0.191765712 | 0.233859789 | 0.979218895 | 0.73778799 | 0.562289506 |
| 101 | taurine | 39 | 74 | 33 | 1.203558 | 1.391116324 | 1.189744995 | 0.187558291 | -0.013813038 | -0.201371329 | 0.451109911 | 0.996956786 | 0.440531903 |
| 102 | urea | 39 | 74 | 33 | 9.027862 | 9.084553305 | 9.213236604 | 0.056691265 | 0.185374565 | 0.1286833 | 0.898140249 | 0.450248468 | 0.61105484 |
| 103 | uric acid | 39 | 74 | 33 | 4.240503 | 4.364625656 | 4.191925775 | 0.124122198 | -0.048577683 | -0.172699881 | 0.483284211 | 0.924501635 | 0.286250556 |
| 104 | Unknown 001 | 39 | 74 | 33 | 0.55874 | 0.454994924 | 0.119123277 | -0.103745198 | -0.439616845 | -0.335871647 | 0.687324964 | 0.010967387 | 0.033323481 |
| 105 | Unknown 002 | 39 | 74 | 33 | -0.44623 | 0.084338764 | 0.140607234 | 0.530573247 | 0.586841717 | 0.05626847 | 0.283372347 | 0.339004925 | 0.987237168 |

| | | | | | | | | | | | | | |
|---|---|---|---|---|---|---|---|---|---|---|---|---|---|
| 106 | Unknown 003 | 39 | 74 | 33 | -1.77166 | -1.884480017 | -2.219016821 | -0.112817767 | -0.447354571 | -0.334536804 | 0.258866383 | 1.75E-06 | 5.72E-05 |
| 107 | Unknown 004 | 39 | 74 | 33 | -0.88967 | -0.947458161 | -0.925476297 | -0.057786523 | -0.035804659 | 0.021981865 | 0.909018522 | 0.974676286 | 0.987730245 |
| 108 | Unknown 005 | 39 | 74 | 33 | 1.323075 | 1.56873333 | 1.59330191 | 0.245658483 | 0.270227063 | 0.02456858 | 0.599745359 | 0.648344974 | 0.995415829 |
| 109 | Unknown 006 | 39 | 74 | 33 | -2.12565 | -2.025838115 | -2.062469315 | 0.099815353 | 0.063184153 | -0.0366312 | 0.542147573 | 0.841548022 | 0.928568149 |
| 110 | Unknown 007 | 39 | 74 | 33 | 0.143794 | -0.238663866 | -0.025425348 | -0.382457675 | -0.169219156 | 0.213238519 | 0.268021684 | 0.833088716 | 0.691084892 |
| 111 | Unknown 008 | 39 | 74 | 33 | -0.51502 | -0.537098751 | -0.954542847 | -0.022077463 | -0.43952156 | -0.417444097 | 0.981852811 | 0.00797828 | 0.003982978 |
| 112 | Unknown 009 | 39 | 74 | 33 | -0.96472 | -1.018678959 | -1.594254595 | -0.053957436 | -0.629533073 | -0.575575636 | 0.931902194 | 0.001834094 | 0.001231867 |
| 113 | Unknown 010 | 39 | 73 | 33 | -0.90913 | -0.903513611 | -0.838531303 | 0.005618907 | 0.070601214 | 0.064982307 | 0.999481254 | 0.944056066 | 0.939878564 |
| 114 | Unknown 011 | 39 | 74 | 33 | 1.645347 | 1.48689251 | 1.322538874 | -0.158454028 | -0.322807664 | -0.164353636 | 0.557082238 | 0.18643854 | 0.569734106 |
| 115 | Unknown 012 | 39 | 74 | 33 | 1.484225 | 1.274154182 | 1.2218215 | -0.210070678 | -0.26240336 | -0.052332682 | 0.332545485 | 0.300886441 | 0.9401225 |
| 116 | Unknown 013 | 39 | 74 | 33 | 0.136522 | 0.227572977 | 0.642180643 | 0.091051421 | 0.505659087 | 0.414607667 | 0.829958155 | 0.020852078 | 0.035429288 |
| 117 | Unknown 014 | 39 | 74 | 33 | 1.707932 | 1.889165011 | 1.590197041 | 0.181233244 | -0.117734726 | -0.29896797 | 0.643985982 | 0.877725691 | 0.345412448 |
| 118 | Unknown 015 | 39 | 74 | 33 | 2.561528 | 2.65037149 | 2.034134673 | 0.088843437 | -0.527393379 | -0.616236817 | 0.923523325 | 0.14604756 | 0.03667455 |
| 119 | Unknown 016 | 39 | 74 | 33 | -3.15119 | -2.943529694 | -3.126726518 | 0.207663869 | 0.024467045 | -0.183196824 | 0.413598901 | 0.991378503 | 0.540241961 |
| 120 | Unknown 017 | 39 | 74 | 33 | -1.38626 | -1.357706955 | -1.789058233 | 0.028549076 | -0.402802202 | -0.431351278 | 0.959862641 | 0.004523914 | 0.000440689 |
| 121 | Unknown 018 | 39 | 74 | 33 | -0.40465 | -0.561085497 | -0.134719815 | -0.156432874 | 0.269932808 | 0.426365682 | 0.422333891 | 0.168837847 | 0.004266051 |
| 122 | Unknown 019 | 39 | 74 | 33 | -4.10777 | -3.517471502 | -3.521761028 | 0.590300908 | 0.586011381 | -0.004289527 | 1.14E-06 | 5.68E-05 | 0.999262654 |
| 123 | Unknown 020 | 39 | 74 | 33 | -3.24372 | -3.082603378 | -2.811841744 | 0.161116948 | 0.431878582 | 0.270761634 | 0.765993357 | 0.265685507 | 0.511787272 |
| 124 | Unknown 021 | 39 | 74 | 33 | -1.42938 | -1.526442269 | -2.049434253 | -0.097059027 | -0.62005101 | -0.522991983 | 0.735054897 | 0.000296723 | 0.000594771 |
| 125 | Unknown 022 | 39 | 74 | 33 | -3.0661 | -3.203002409 | -3.430466018 | -0.136906058 | -0.364369667 | -0.227463609 | 0.49285671 | 0.033059849 | 0.177915936 |
| 126 | Unknown 023 | 39 | 73 | 33 | -2.96581 | -2.988169204 | -3.293331461 | -0.022357524 | -0.327519781 | -0.305162257 | 0.978734386 | 0.043376096 | 0.031676768 |
| 127 | Unknown 024 | 39 | 74 | 32 | -0.34443 | -0.248643154 | -0.42431828 | 0.095783411 | -0.079891716 | -0.175675126 | 0.91910639 | 0.960397083 | 0.780549162 |
| 128 | Unknown 025 | 39 | 74 | 33 | -1.87751 | -1.720503607 | -1.983623363 | 0.157006757 | -0.106112998 | -0.263119755 | 0.193402153 | 0.5877101 | 0.017902582 |
| 129 | Unknown 026 | 39 | 74 | 33 | 0.425196 | 0.476457069 | 0.427128788 | 0.051260936 | 0.001932655 | -0.049328281 | 0.839758937 | 0.999825942 | 0.865402622 |
| 130 | Unknown 027 | 39 | 74 | 33 | -1.02035 | -1.056280425 | -1.095584969 | -0.035931138 | -0.075235683 | -0.039304544 | 0.87725301 | 0.669861089 | 0.869342837 |
| 131 | Unknown 028 | 39 | 74 | 33 | 0.073129 | 0.266293724 | -0.365731861 | 0.193165017 | -0.438860568 | -0.632025585 | 0.521017644 | 0.098851198 | 0.002715798 |
| 132 | Unknown 029 | 12 | 23 | 12 | -4.7284 | -4.915376484 | -3.996834245 | -0.186976407 | 0.731565831 | 0.918542238 | 0.973358034 | 0.731877283 | 0.526713209 |
| 133 | Unknown 030 | 39 | 74 | 33 | -1.50808 | -1.418581572 | -1.66932242 | 0.089495222 | -0.161245626 | -0.250740848 | 0.612183714 | 0.330367078 | 0.035424934 |
| 134 | Unknown 031 | 39 | 74 | 33 | -1.64108 | -1.630846031 | -1.106482047 | 0.010231157 | 0.534595141 | 0.524363984 | 0.997329077 | 0.007766687 | 0.002724 |
| 135 | Unknown 032 | 4 | 8 | 10 | 0.648028 | 0.419621978 | 0.799908784 | -0.228406229 | 0.151880576 | 0.380286806 | 0.965521794 | 0.98349785 | 0.851336019 |
| 136 | Unknown 033 | 39 | 74 | 33 | 0.090941 | -0.625701617 | -0.336216366 | -0.716642891 | -0.42715764 | 0.289485251 | 2.16E-06 | 0.02871913 | 0.120864867 |
| 137 | Unknown 034 | 39 | 74 | 33 | -3.73172 | -3.446270094 | -3.651881591 | 0.285445027 | 0.07983353 | -0.205611497 | 0.21694533 | 0.918544746 | 0.489340474 |
| 138 | Unknown 035 | 39 | 74 | 33 | -1.28787 | -1.837411179 | -1.932508354 | -0.549544841 | -0.644642016 | -0.095097175 | 0.000205736 | 0.000275212 | 0.781223938 |
| 139 | Unknown 036 | 39 | 74 | 33 | -0.96805 | -1.181906433 | -1.143067263 | -0.213855182 | -0.175016013 | 0.03883917 | 0.088345895 | 0.315742129 | 0.929288814 |
| 140 | Unknown 037 | 39 | 74 | 33 | -3.91437 | -3.603787366 | -3.504900194 | 0.310584462 | 0.409471634 | 0.098887172 | 0.024011066 | 0.011094624 | 0.704661332 |
| 141 | Unknown 038 | 39 | 74 | 33 | -1.79303 | -1.626575372 | -1.297895018 | 0.166452871 | 0.495133224 | 0.328680354 | 0.734527142 | 0.152762548 | 0.344293619 |
| 142 | Unknown 039 | 39 | 73 | 33 | -3.51713 | -4.40957706 | -4.204023185 | -0.892447091 | -0.686893216 | 0.205553875 | 0.144630516 | 0.44257029 | 0.910701302 |
| 143 | Unknown 040 | 39 | 74 | 33 | 2.990792 | 2.906559887 | 2.522003652 | -0.084232573 | -0.468788808 | -0.384556235 | 0.600010602 | 4.24E-05 | 0.00015746 |
| 144 | Unknown 041 | 39 | 74 | 33 | 3.848689 | 3.841320365 | 3.729823029 | -0.007368763 | -0.1188661 | -0.111497336 | 0.99677782 | 0.557617635 | 0.51910796 |
| 145 | Unknown 042 | 39 | 74 | 33 | 7.598106 | 7.593278517 | 7.504044217 | -0.00482748 | -0.094061781 | -0.0892343 | 0.99658204 | 0.406172729 | 0.355642543 |
| 146 | Unknown 043 | 39 | 74 | 33 | 4.17127 | 4.081114192 | 3.850854731 | -0.090155389 | -0.320414851 | -0.230259461 | 0.510942344 | 0.00355367 | 0.022631998 |
| 147 | Unknown 044 | 39 | 74 | 33 | 1.630782 | 1.475907002 | 1.637211408 | -0.154874859 | 0.006429547 | 0.161304407 | 0.806433742 | 0.999739929 | 0.811759067 |
| 148 | Unknown 045 | 39 | 74 | 33 | -0.00816 | -0.031086683 | -0.585295705 | -0.022924659 | -0.577133681 | -0.554209022 | 0.983039887 | 0.000859292 | 0.000269272 |
| 149 | Unknown 046 | 39 | 74 | 33 | 3.774661 | 3.704232701 | 3.57756929 | -0.070428306 | -0.197091717 | -0.126663411 | 0.717221932 | 0.166372654 | 0.385097948 |
| 150 | Unknown 047 | 39 | 74 | 33 | -2.10572 | -1.728737763 | -1.260526133 | 0.376978296 | 0.845189926 | 0.46821163 | 0.753434283 | 0.372143617 | 0.677184418 |
| 151 | Unknown 048 | 39 | 74 | 33 | -1.3509 | -1.394541577 | -1.102394819 | -0.043644997 | 0.248501762 | 0.292146758 | 0.963030992 | 0.428591572 | 0.226511251 |
| 152 | Unknown 049 | 39 | 74 | 33 | -0.18613 | 0.156855614 | 0.132637338 | 0.342983505 | 0.318765229 | -0.024218276 | 0.615124675 | 0.744999023 | 0.997825986 |
| 153 | Unknown 050 | 39 | 74 | 33 | 2.79875 | 2.76177089 | 2.736962375 | -0.036978984 | -0.061787499 | -0.024808516 | 0.95797332 | 0.919562389 | 0.982875164 |
| 154 | Unknown 051 | 39 | 74 | 33 | 3.171386 | 3.032831389 | 2.935917142 | -0.13855421 | -0.235468458 | -0.096914247 | 0.619957997 | 0.382344709 | 0.810880345 |
| 155 | Unknown 052 | 39 | 74 | 33 | 0.392426 | 0.028125463 | -0.083727622 | -0.364300801 | -0.476153886 | -0.111853085 | 0.130880534 | 0.088793257 | 0.839678735 |
| 156 | Unknown 053 | 39 | 74 | 33 | 1.344447 | 1.475568631 | 1.28524087 | 0.131121975 | -0.059205786 | -0.190327761 | 0.622030298 | 0.934218026 | 0.410700017 |
| 157 | Unknown 054 | 39 | 74 | 33 | 2.490648 | 2.504647417 | 2.647005078 | 0.013999408 | 0.156357069 | 0.142357661 | 0.994076659 | 0.597008957 | 0.579400284 |
| 158 | Unknown 055 | 39 | 74 | 33 | 1.42603 | 1.486535709 | 0.97805283 | 0.060505366 | -0.447977513 | -0.508482879 | 0.86037962 | 0.004252164 | 0.00016572 |
| 159 | Unknown 056 | 39 | 74 | 33 | -3.21598 | -3.04872778 | -3.126605462 | 0.167249293 | 0.089371612 | -0.077877682 | 0.339949158 | 0.804418057 | 0.809765211 |
| 160 | Unknown 057 | 39 | 74 | 33 | -3.34147 | -3.138676588 | -3.173066741 | 0.202788932 | 0.168398778 | -0.034390154 | 0.185051485 | 0.439637231 | 0.956830507 |
| 161 | Unknown 058 | 39 | 74 | 33 | -0.69092 | -0.290669697 | -0.505171684 | 0.400253641 | 0.185751654 | -0.214501987 | 0.14432838 | 0.743095371 | 0.603854279 |
| 162 | Unknown 059 | 39 | 74 | 33 | -0.10186 | 0.223860219 | 0.433884059 | 0.325724439 | 0.535748278 | 0.210023839 | 0.084657807 | 0.010302821 | 0.39362277 |
| 163 | Unknown 060 | 39 | 74 | 33 | 0.478817 | 0.50638845 | 0.313291332 | 0.027571292 | -0.165525827 | -0.193097119 | 0.915971507 | 0.114694547 | 0.024644409 |
| 164 | Unknown 061 | 39 | 74 | 33 | -1.24489 | -1.167345735 | -1.946946297 | 0.077545418 | -0.702055144 | -0.779600562 | 0.894592548 | 0.00244922 | 0.000102827 |
| 165 | Unknown 062 | 39 | 74 | 33 | -0.08492 | -0.099892493 | -0.491353954 | -0.014970445 | -0.406431906 | -0.391461461 | 0.987992616 | 0.002836333 | 0.001029415 |
| 166 | Unknown 063 | 39 | 74 | 33 | 0.000616 | -0.099984169 | -0.485928118 | -0.100600181 | -0.486544129 | -0.385943948 | 0.537940716 | 9.10E-05 | 0.00050199 |
| 167 | Unknown 064 | 39 | 74 | 33 | -0.15406 | -0.139241139 | -0.491370583 | 0.014818711 | -0.337310732 | -0.352129444 | 0.987622231 | 0.013337701 | 0.002705988 |
| 168 | Unknown 065 | 39 | 74 | 33 | -1.15987 | -1.225547126 | -1.45891011 | -0.065681113 | -0.299044097 | -0.233362984 | 0.944756998 | 0.441435545 | 0.528861157 |
| 169 | Unknown 066 | 39 | 74 | 33 | 0.108702 | 0.031816078 | 0.062944427 | -0.076886238 | -0.045757889 | 0.031128349 | 0.781243886 | 0.94052223 | 0.964406525 |
| 170 | Unknown 067 | 39 | 74 | 33 | 0.518708 | 0.643390886 | 0.816140436 | 0.124682482 | 0.297432032 | 0.17274955 | 0.711181778 | 0.261077084 | 0.558149051 |
| 171 | Unknown 068 | 39 | 74 | 33 | -1.12346 | -1.123810739 | -1.844464924 | -0.000354632 | -0.721008817 | -0.720654185 | 0.999997903 | 0.003289472 | 0.000755285 |
| 172 | Unknown 069 | 39 | 74 | 33 | -1.42071 | -1.104633301 | -1.474647564 | 0.316072478 | -0.053941784 | -0.370014262 | 0.350363965 | 0.978614625 | 0.277704842 |
| 173 | Unknown 070 | 39 | 74 | 33 | -1.81281 | -1.984478327 | -2.030271087 | -0.171664517 | -0.217457278 | -0.045792761 | 0.035174869 | 0.023654144 | 0.80251545 |
| 174 | Unknown 071 | 39 | 74 | 33 | -0.17137 | -0.080892017 | -0.526234745 | 0.090476392 | -0.354866336 | -0.445342728 | 0.868563902 | 0.224182384 | 0.051769733 |
| 175 | Unknown 072 | 39 | 74 | 33 | 1.086682 | 1.089513572 | 0.957282546 | 0.002831371 | -0.129399655 | -0.132231026 | 0.999269245 | 0.347657378 | 0.245749189 |
| 176 | Unknown 073 | 39 | 74 | 33 | 6.944682 | 6.929978755 | 6.602363403 | -0.014703343 | -0.342318695 | -0.327615352 | 0.9897188 | 0.023092481 | 0.012531041 |
| 177 | Unknown 074 | 39 | 74 | 33 | -2.14625 | -2.134793238 | -2.296320623 | 0.011451824 | -0.150075561 | -0.161527385 | 0.997516328 | 0.742340111 | 0.644030952 |
| 178 | Unknown 075 | 39 | 72 | 33 | -3.40806 | -3.683535025 | -4.057526643 | -0.275474272 | -0.64946589 | -0.373991619 | 0.531277612 | 0.087462372 | 0.354095508 |
| 179 | Unknown 076 | 39 | 74 | 33 | -3.34872 | -3.563960159 | -3.492178682 | -0.215240707 | -0.14345923 | 0.071781477 | 0.687327045 | 0.889673211 | 0.963290273 |
| 180 | Unknown 077 | 39 | 74 | 33 | -2.73741 | -2.596929336 | -2.792540901 | 0.140478938 | -0.055132627 | -0.195611565 | 0.420759182 | 0.910188598 | 0.225375942 |
| 181 | Unknown 078 | 39 | 74 | 33 | -1.5669 | -1.381661352 | -2.03161464 | 0.185241535 | -0.464711753 | -0.649953288 | 0.558342838 | 0.080868059 | 0.00233437 |
| 182 | Unknown 079 | 39 | 74 | 33 | -4.2185 | -4.318584358 | -3.654997017 | -0.100085253 | 0.563502088 | 0.663587341 | 0.881166664 | 0.065210387 | 0.008773501 |
| 183 | Unknown 080 | 39 | 74 | 33 | -0.47814 | -0.285719424 | -1.492036461 | 0.192420914 | -1.013896123 | -1.206317037 | 0.793570918 | 0.013498334 | 0.000532597 |
| 184 | Unknown 081 | 39 | 74 | 33 | -2.42938 | -2.36962419 | -2.244059002 | 0.059758749 | 0.185323937 | 0.125565188 | 0.855054149 | 0.352043243 | 0.540779981 |
| 185 | Unknown 082 | 39 | 74 | 33 | -3.60667 | -3.456276173 | -3.940233212 | 0.150391771 | -0.333565268 | -0.483957039 | 0.397990459 | 0.045036676 | 0.000355395 |
| 186 | Unknown 083 | 39 | 74 | 33 | -1.72511 | -2.310133941 | -2.448621559 | -0.585019139 | -0.723506757 | -0.138487618 | 1.30E-07 | 4.90E-08 | 0.400272505 |
| 187 | Unknown 084 | 39 | 74 | 33 | -0.89243 | -0.93818929 | -1.599986696 | -0.045756429 | -0.707553835 | -0.661797406 | 0.958631917 | 0.001348534 | 0.000655208 |
| 188 | Unknown 085 | 39 | 74 | 33 | -0.9422 | -0.773080888 | -0.791887042 | 0.169120915 | 0.150314761 | -0.018806154 | 0.74052616 | 0.84677031 | 0.996675561 |
| 189 | Unknown 086 | 39 | 74 | 33 | -2.33279 | -2.446507932 | -2.895437011 | -0.113718257 | -0.562647335 | -0.448929078 | 0.563951937 | 0.000121422 | 0.000587876 |
| 190 | Unknown 087 | 39 | 74 | 33 | 0.030176 | 0.073068249 | 0.132026309 | 0.042892435 | 0.101850496 | 0.058958061 | 0.947924648 | 0.81002458 | 0.913694808 |
| 191 | Unknown 088 | 39 | 72 | 33 | 0.004447 | -0.288637088 | 0.011933471 | -0.293084083 | 0.007486476 | 0.300570559 | 0.633610174 | 0.999788586 | 0.650881791 |
| 192 | Unknown 089 | 39 | 74 | 33 | -3.333 | -3.317049519 | -3.212752646 | 0.015952541 | 0.120249414 | 0.104296873 | 0.989245934 | 0.65178285 | 0.662850192 |
| 193 | Unknown 090 | 39 | 74 | 33 | -1.5538 | -1.547602386 | -1.390202693 | 0.006202605 | 0.163602298 | 0.157399693 | 0.999317383 | 0.717987264 | 0.676189982 |
| 194 | Unknown 091 | 39 | 74 | 33 | -2.87333 | -2.721607155 | -3.266681882 | 0.151719873 | -0.393354853 | -0.545074727 | 0.89262816 | 0.587460813 | 0.274297283 |
| 195 | Unknown 092 | 39 | 73 | 32 | -3.35162 | -3.678600331 | -3.878867451 | -0.326980077 | -0.527247197 | -0.200267121 | 0.012076219 | 0.000458601 | 0.22426002 |
| 196 | Unknown 093 | 39 | 74 | 32 | 1.688379 | 0.714967779 | 0.580605135 | -0.97341171 | -1.107774353 | -0.134362644 | 4.04E-12 | 4.43E-11 | 0.575444817 |
| 197 | Unknown 094 | 39 | 74 | 33 | 1.299567 | 1.157334048 | 0.889025593 | -0.142232843 | -0.410541298 | -0.268308455 | 0.80820727 | 0.292833727 | 0.509791036 |
| 198 | Unknown 095 | 39 | 74 | 33 | -0.18717 | -1.113023304 | -1.158390971 | -0.925849151 | -0.971216818 | -0.045367667 | 0.001654201 | 0.006729967 | 0.98543024 |
| 199 | Unknown 096 | 39 | 74 | 33 | -1.72277 | -1.681506985 | -1.453590123 | 0.041263185 | 0.269180047 | 0.227916862 | 0.978601538 | 0.527313098 | 0.556480956 |
| 200 | Unknown 097 | 39 | 74 | 33 | 2.761275 | 2.908066387 | 2.433086007 | 0.14679137 | -0.32818901 | -0.47498038 | 0.643768143 | 0.218052841 | 0.018857427 |
| 201 | Unknown 098 | 39 | 74 | 33 | -0.72195 | -0.843674559 | -0.912619293 | -0.121729474 | -0.190674208 | -0.068944734 | 0.802833012 | 0.68627865 | 0.938888771 |
| 202 | Unknown 099 | 39 | 74 | 33 | 2.900109 | 2.749293819 | 2.490485769 | -0.150815187 | -0.409623238 | -0.25880805 | 0.052987706 | 1.13E-06 | 0.000614356 |
| 203 | Unknown 100 | 39 | 74 | 33 | 1.628652 | 1.574013628 | 1.346401505 | -0.054638644 | -0.282250767 | -0.227612123 | 0.762020078 | 0.007907047 | 0.017409097 |

| | | | | | | | | | | | | | |
|---|---|---|---|---|---|---|---|---|---|---|---|---|---|
| 204 | Unknown 101 | 39 | 74 | 33 | -2.22748 | -2.261232904 | -2.358659818 | -0.033749888 | -0.131176802 | -0.097426914 | 0.993087642 | 0.929335817 | 0.949682941 |
| 205 | Unknown 102 | 39 | 73 | 33 | -3.51935 | -1.981837844 | -1.641701816 | 1.537507857 | 1.877643885 | 0.340136028 | 1.69E-05 | 1.04E-05 | 0.586642877 |
| 206 | Unknown 103 | 39 | 74 | 33 | 0.011495 | 0.172496474 | 0.154895125 | 0.161001151 | 0.143399801 | -0.01760135 | 0.591706571 | 0.74670605 | 0.994379888 |
| 207 | Unknown 104 | 39 | 70 | 32 | -1.64607 | -4.286801006 | -5.077334368 | -2.640731845 | -3.431265207 | -0.790533362 | 8.66E-08 | 6.60E-09 | 0.228642052 |
| 208 | Unknown 105 | 39 | 74 | 33 | 0.44489 | 0.336857909 | 0.281883988 | -0.108032012 | -0.163005933 | -0.054973921 | 0.811256064 | 0.716876499 | 0.952671057 |
| 209 | Unknown 106 | 39 | 74 | 33 | -0.65443 | -0.650681906 | -0.677986907 | 0.003748679 | -0.023556323 | -0.027305002 | 0.999834414 | 0.995434048 | 0.992180286 |
| 210 | Unknown 107 | 39 | 74 | 33 | 4.079474 | 4.187140091 | 4.252216362 | 0.10766614 | 0.172742411 | 0.065076271 | 0.833517076 | 0.720677336 | 0.942209108 |
| 211 | Unknown 108 | 39 | 74 | 33 | -1.60148 | -1.531654425 | -1.474809211 | 0.06982402 | 0.126669234 | 0.056845214 | 0.950927434 | 0.890638518 | 0.970631518 |
| 212 | Unknown 109 | 39 | 74 | 33 | -2.04133 | -1.727937458 | -1.950211685 | 0.31339412 | 0.091119892 | -0.222274227 | 0.510154406 | 0.960698314 | 0.738046604 |
| 213 | Unknown 110 | 38 | 74 | 33 | -1.07312 | -0.557021525 | -0.37689771 | 0.516100639 | 0.696224454 | 0.180123815 | 0.264552258 | 0.183479702 | 0.861466226 |

### Supplemental Table IV:

| Protein | uniprot_url | Cor_w_NET |
|---|---|---|
| P62258 | https://www.uniprot.org/uniprot/P62258 | 0.691252814 |
| P02671 | https://www.uniprot.org/uniprot/P02671 | 0.608444092 |
| P15291 | https://www.uniprot.org/uniprot/P15291 | 0.791351511 |
| P11047 | https://www.uniprot.org/uniprot/P11047 | 0.762476326 |
| P0DOY3 | https://www.uniprot.org/uniprot/P0DOY3 | 0.684550547 |
| P06744 | https://www.uniprot.org/uniprot/P06744 | 0.808941074 |
| P24821 | https://www.uniprot.org/uniprot/P24821 | 0.682467661 |
| P01033 | https://www.uniprot.org/uniprot/P01033 | 0.827003164 |
| P14625 | https://www.uniprot.org/uniprot/P14625 | 0.739693826 |
| Q12907 | https://www.uniprot.org/uniprot/Q12907 | 0.686606335 |
| P0DJI8 | https://www.uniprot.org/uniprot/P0DJI8 | 0.743892057 |
| P69905 | https://www.uniprot.org/uniprot/P69905 | 0.619340603 |
| P13667 | https://www.uniprot.org/uniprot/P13667 | 0.761316849 |
| P01011 | https://www.uniprot.org/uniprot/P01011 | 0.678784994 |
| P02741 | https://www.uniprot.org/uniprot/P02741 | 0.74196483 |
| P02788 | https://www.uniprot.org/uniprot/P02788 | 0.740338601 |
| P55854 | https://www.uniprot.org/uniprot/P55854 | 0.661225392 |
| P20160 | https://www.uniprot.org/uniprot/P20160 | 0.769067958 |
| P02042 | https://www.uniprot.org/uniprot/P02042 | 0.612421726 |
| Q02809 | https://www.uniprot.org/uniprot/Q02809 | 0.635092928 |
| P18428 | https://www.uniprot.org/uniprot/P18428 | 0.769097603 |
| Q9UHI8 | https://www.uniprot.org/uniprot/Q9UHI8 | 0.655870394 |
| P20333 | https://www.uniprot.org/uniprot/P20333 | 0.641866 |
| Q15485 | https://www.uniprot.org/uniprot/Q15485 | 0.606446189 |
| P10451 | https://www.uniprot.org/uniprot/P10451 | 0.667184233 |
| P05164 | https://www.uniprot.org/uniprot/P05164 | 0.82327699 |
| P13796 | https://www.uniprot.org/uniprot/P13796 | 0.666850137 |
| P04275 | https://www.uniprot.org/uniprot/P04275 | 0.628584018 |
| P25786 | https://www.uniprot.org/uniprot/P25786 | 0.797779734 |
| Q9BYE9 | https://www.uniprot.org/uniprot/Q9BYE9 | 0.605212344 |
| P02750 | https://www.uniprot.org/uniprot/P02750 | 0.667660953 |
| Q06033 | https://www.uniprot.org/uniprot/Q06033 | 0.635130951 |
| P01034 | https://www.uniprot.org/uniprot/P01034 | 0.652683816 |
| P23381 | https://www.uniprot.org/uniprot/P23381 | 0.701027123 |
| P60900 | https://www.uniprot.org/uniprot/P60900 | 0.805532798 |
| P07998 | https://www.uniprot.org/uniprot/P07998 | 0.601512371 |
| P33908 | https://www.uniprot.org/uniprot/P33908 | 0.604425892 |
| Q8TDQ0 | https://www.uniprot.org/uniprot/Q8TDQ0 | 0.722261494 |
| P00338 | https://www.uniprot.org/uniprot/P00338 | 0.732384091 |
| P01762 | https://www.uniprot.org/uniprot/P01762 | 0.703321131 |
| P05362 | https://www.uniprot.org/uniprot/P05362 | 0.713910191 |
| P14543 | https://www.uniprot.org/uniprot/P14543 | 0.791857117 |
| O43278 | https://www.uniprot.org/uniprot/O43278 | 0.646364245 |
| Q14118 | https://www.uniprot.org/uniprot/Q14118 | 0.626801502 |
| P25789 | https://www.uniprot.org/uniprot/P25789 | 0.802991068 |
| P68871 | https://www.uniprot.org/uniprot/P68871 | 0.666536499 |

| | | |
|---|---|---|
| P08670 | https://www.uniprot.org/uniprot/P08670 | 0.884880768 |
| P07602 | https://www.uniprot.org/uniprot/P07602 | 0.796856263 |
| P14314 | https://www.uniprot.org/uniprot/P14314 | 0.816199799 |
| P34096 | https://www.uniprot.org/uniprot/P34096 | 0.634115535 |
| P04040 | https://www.uniprot.org/uniprot/P04040 | 0.760608695 |
| P35442 | https://www.uniprot.org/uniprot/P35442 | 0.719742337 |
| P0DJI9 | https://www.uniprot.org/uniprot/P0DJI9 | 0.807233734 |
| Q8TCZ2 | https://www.uniprot.org/uniprot/Q8TCZ2 | 0.653251288 |
| P06312 | https://www.uniprot.org/uniprot/P06312 | 0.654293225 |
| P01619 | https://www.uniprot.org/uniprot/P01619 | 0.617770057 |
| Q14767 | https://www.uniprot.org/uniprot/Q14767 | 0.699846897 |
| P06702 | https://www.uniprot.org/uniprot/P06702 | 0.879841835 |
| P39060 | https://www.uniprot.org/uniprot/P39060 | 0.689603103 |
| Q9Y279 | https://www.uniprot.org/uniprot/Q9Y279 | 0.794195668 |
| P40306 | https://www.uniprot.org/uniprot/P40306 | 0.720995502 |
| Q99436 | https://www.uniprot.org/uniprot/Q99436 | 0.776661454 |
| Q969E1 | https://www.uniprot.org/uniprot/Q969E1 | 0.677559028 |
| P29401 | https://www.uniprot.org/uniprot/P29401 | 0.855001398 |
| P53004 | https://www.uniprot.org/uniprot/P53004 | 0.665754034 |
| P24043 | https://www.uniprot.org/uniprot/P24043 | 0.629258452 |
| P05109 | https://www.uniprot.org/uniprot/P05109 | 0.90658628 |
| P20810 | https://www.uniprot.org/uniprot/P20810 | 0.771421498 |
| P14555 | https://www.uniprot.org/uniprot/P14555 | 0.809310297 |
| P07954 | https://www.uniprot.org/uniprot/P07954 | 0.725504356 |
| P27824 | https://www.uniprot.org/uniprot/P27824 | 0.724100564 |
| Q86VP6 | https://www.uniprot.org/uniprot/Q86VP6 | 0.709622404 |
| P80723 | https://www.uniprot.org/uniprot/P80723 | 0.709838428 |
| P29966 | https://www.uniprot.org/uniprot/P29966 | 0.689532859 |
| Q12882 | https://www.uniprot.org/uniprot/Q12882 | 0.659915578 |
| Q5JRA6 | https://www.uniprot.org/uniprot/Q5JRA6 | 0.708539419 |
| P05386 | https://www.uniprot.org/uniprot/P05386 | 0.799088732 |
| P80511 | https://www.uniprot.org/uniprot/P80511 | 0.800440275 |
| P25787 | https://www.uniprot.org/uniprot/P25787 | 0.760424836 |
| Q9Y4L1 | https://www.uniprot.org/uniprot/Q9Y4L1 | 0.694284962 |
| P37837 | https://www.uniprot.org/uniprot/P37837 | 0.896216698 |
| P22234 | https://www.uniprot.org/uniprot/P22234 | 0.774540359 |
| P22897 | https://www.uniprot.org/uniprot/P22897 | 0.782048936 |
| P00441 | https://www.uniprot.org/uniprot/P00441 | 0.690698022 |
| P61970 | https://www.uniprot.org/uniprot/P61970 | 0.665618755 |
| P00390 | https://www.uniprot.org/uniprot/P00390 | 0.703576077 |
| Q09666 | https://www.uniprot.org/uniprot/Q09666 | 0.685486192 |
| Q99523 | https://www.uniprot.org/uniprot/Q99523 | 0.617867479 |
| P07900 | https://www.uniprot.org/uniprot/P07900 | 0.702408536 |
| P30740 | https://www.uniprot.org/uniprot/P30740 | 0.733393687 |
| P11234 | https://www.uniprot.org/uniprot/P11234 | 0.674426981 |
| P34932 | https://www.uniprot.org/uniprot/P34932 | 0.820761479 |
| P13798 | https://www.uniprot.org/uniprot/P13798 | 0.644591851 |

| | | |
|---|---|---|
| P53396 | https://www.uniprot.org/uniprot/P53396 | 0.675525827 |
| P55786 | https://www.uniprot.org/uniprot/P55786 | 0.686136776 |
| P05388 | https://www.uniprot.org/uniprot/P05388 | 0.785111322 |
| P09603 | https://www.uniprot.org/uniprot/P09603 | 0.632987154 |
| Q9UNZ2 | https://www.uniprot.org/uniprot/Q9UNZ2 | 0.747321332 |
| P48147 | https://www.uniprot.org/uniprot/P48147 | 0.714580535 |
| P0DMV8 | https://www.uniprot.org/uniprot/P0DMV8 | 0.756017785 |
| P02461 | https://www.uniprot.org/uniprot/P02461 | 0.681093691 |
| P20618 | https://www.uniprot.org/uniprot/P20618 | 0.798548973 |
| P23526 | https://www.uniprot.org/uniprot/P23526 | 0.788609142 |
| P13987 | https://www.uniprot.org/uniprot/P13987 | 0.63943807 |
| P49247 | https://www.uniprot.org/uniprot/P49247 | 0.776232211 |
| P52209 | https://www.uniprot.org/uniprot/P52209 | 0.786552322 |
| B9A064 | https://www.uniprot.org/uniprot/B9A064 | 0.71264617 |
| P08572 | https://www.uniprot.org/uniprot/P08572 | 0.623118125 |
| O00461 | https://www.uniprot.org/uniprot/O00461 | 0.661835631 |
| P61201 | https://www.uniprot.org/uniprot/P61201 | 0.691035947 |
| Q93091 | https://www.uniprot.org/uniprot/Q93091 | 0.638613142 |
| O95861 | https://www.uniprot.org/uniprot/O95861 | 0.647008222 |
| P50502 | https://www.uniprot.org/uniprot/P50502 | 0.74662288 |
| P16949 | https://www.uniprot.org/uniprot/P16949 | 0.801325287 |
| P48637 | https://www.uniprot.org/uniprot/P48637 | 0.648133704 |
| O75223 | https://www.uniprot.org/uniprot/O75223 | 0.756694041 |
| O43242 | https://www.uniprot.org/uniprot/O43242 | 0.615271567 |
| Q13442 | https://www.uniprot.org/uniprot/Q13442 | 0.623034728 |
| P04080 | https://www.uniprot.org/uniprot/P04080 | 0.770977664 |
| P15907 | https://www.uniprot.org/uniprot/P15907 | 0.733997889 |
| P07108 | https://www.uniprot.org/uniprot/P07108 | 0.727733443 |
| P52888 | https://www.uniprot.org/uniprot/P52888 | 0.622283895 |
| Q8NBS9 | https://www.uniprot.org/uniprot/Q8NBS9 | 0.703060157 |
| O75594 | https://www.uniprot.org/uniprot/O75594 | 0.635250222 |
| P22061 | https://www.uniprot.org/uniprot/P22061 | 0.723423623 |
| O14818 | https://www.uniprot.org/uniprot/O14818 | 0.793963594 |
| O00468 | https://www.uniprot.org/uniprot/O00468 | 0.788048156 |
| P98095 | https://www.uniprot.org/uniprot/P98095 | 0.64833475 |
| P15259 | https://www.uniprot.org/uniprot/P15259 | 0.695824103 |
| Q16851 | https://www.uniprot.org/uniprot/Q16851 | 0.645059979 |
| P40121 | https://www.uniprot.org/uniprot/P40121 | 0.719552129 |
| Q02818 | https://www.uniprot.org/uniprot/Q02818 | 0.781380357 |
| Q8WWV6 | https://www.uniprot.org/uniprot/Q8WWV6 | 0.618795762 |
| Q9Y5Z4 | https://www.uniprot.org/uniprot/Q9Y5Z4 | 0.711916306 |
| Q06323 | https://www.uniprot.org/uniprot/Q06323 | 0.783788055 |
| P02545 | https://www.uniprot.org/uniprot/P02545 | 0.750270025 |
| P49773 | https://www.uniprot.org/uniprot/P49773 | 0.686902169 |
| P52790 | https://www.uniprot.org/uniprot/P52790 | 0.845000604 |
| P00451 | https://www.uniprot.org/uniprot/P00451 | 0.624939349 |
| Q9NR99 | https://www.uniprot.org/uniprot/Q9NR99 | 0.650671598 |

| | | |
|---|---|---|
| P09960 | https://www.uniprot.org/uniprot/P09960 | 0.814630169 |
| P02730 | https://www.uniprot.org/uniprot/P02730 | 0.717541184 |
| Q06830 | https://www.uniprot.org/uniprot/Q06830 | 0.663615455 |
| P69891 | https://www.uniprot.org/uniprot/P69891 | 0.605236387 |
| A6NI73 | https://www.uniprot.org/uniprot/A6NI73 | 0.651110764 |
| P25788 | https://www.uniprot.org/uniprot/P25788 | 0.785862302 |
| Q9H361 | https://www.uniprot.org/uniprot/Q9H361 | 0.760458296 |
| Q12906 | https://www.uniprot.org/uniprot/Q12906 | 0.618754621 |
| Q4VXU2 | https://www.uniprot.org/uniprot/Q4VXU2 | 0.715610352 |
| P61916 | https://www.uniprot.org/uniprot/P61916 | 0.645563702 |
| P63010 | https://www.uniprot.org/uniprot/P63010 | 0.659390987 |
| P07306 | https://www.uniprot.org/uniprot/P07306 | 0.665685674 |
| Q9NUQ9 | https://www.uniprot.org/uniprot/Q9NUQ9 | 0.829239619 |
| P06737 | https://www.uniprot.org/uniprot/P06737 | 0.751087571 |
| Q12905 | https://www.uniprot.org/uniprot/Q12905 | 0.628902881 |
| O14602 | https://www.uniprot.org/uniprot/O14602 | 0.683273548 |
| P13639 | https://www.uniprot.org/uniprot/P13639 | 0.843786004 |
| P61981 | https://www.uniprot.org/uniprot/P61981 | 0.674721076 |
| P80294 | https://www.uniprot.org/uniprot/P80294 | 0.67762907 |
| P03973 | https://www.uniprot.org/uniprot/P03973 | 0.746195038 |
| Q14508 | https://www.uniprot.org/uniprot/Q14508 | 0.653018746 |
| P30086 | https://www.uniprot.org/uniprot/P30086 | 0.758130738 |
| Q8NHW5 | https://www.uniprot.org/uniprot/Q8NHW5 | 0.790055642 |
| Q13867 | https://www.uniprot.org/uniprot/Q13867 | 0.717831344 |
| Q9UL46 | https://www.uniprot.org/uniprot/Q9UL46 | 0.75539559 |
| Q13228 | https://www.uniprot.org/uniprot/Q13228 | 0.603222157 |
| Q15435 | https://www.uniprot.org/uniprot/Q15435 | 0.742304035 |
| P08238 | https://www.uniprot.org/uniprot/P08238 | 0.686148961 |
| Q9NP84 | https://www.uniprot.org/uniprot/Q9NP84 | 0.681656528 |
| P10599 | https://www.uniprot.org/uniprot/P10599 | 0.786085967 |
| P48506 | https://www.uniprot.org/uniprot/P48506 | 0.670152681 |
| P11586 | https://www.uniprot.org/uniprot/P11586 | 0.612034072 |
| P15153 | https://www.uniprot.org/uniprot/P15153 | 0.833603382 |
| P00995 | https://www.uniprot.org/uniprot/P00995 | 0.72332499 |
| Q92626 | https://www.uniprot.org/uniprot/Q92626 | 0.743751689 |
| Q13630 | https://www.uniprot.org/uniprot/Q13630 | 0.635029067 |
| Q9Y266 | https://www.uniprot.org/uniprot/Q9Y266 | 0.798887125 |
| O60462 | https://www.uniprot.org/uniprot/O60462 | 0.640631045 |
| Q14697 | https://www.uniprot.org/uniprot/Q14697 | 0.78318059 |
| P20042 | https://www.uniprot.org/uniprot/P20042 | 0.649747686 |
| P35754 | https://www.uniprot.org/uniprot/P35754 | 0.800126516 |
| Q9Y5Y6 | https://www.uniprot.org/uniprot/Q9Y5Y6 | 0.65505868 |
| P27797 | https://www.uniprot.org/uniprot/P27797 | 0.771731828 |
| Q96NZ9 | https://www.uniprot.org/uniprot/Q96NZ9 | 0.601253962 |
| P16157 | https://www.uniprot.org/uniprot/P16157 | 0.633971499 |
| Q01638 | https://www.uniprot.org/uniprot/Q01638 | 0.713302436 |
| Q13177 | https://www.uniprot.org/uniprot/Q13177 | 0.624772918 |

| | | |
|---|---|---|
| P09382 | https://www.uniprot.org/uniprot/P09382 | 0.667246177 |
| P68104 | https://www.uniprot.org/uniprot/P68104 | 0.790391821 |
| Q16658 | https://www.uniprot.org/uniprot/Q16658 | 0.665200763 |
| P31949 | https://www.uniprot.org/uniprot/P31949 | 0.88475895 |
| Q13200 | https://www.uniprot.org/uniprot/Q13200 | 0.629455712 |
| P07858 | https://www.uniprot.org/uniprot/P07858 | 0.618477795 |
| P61978 | https://www.uniprot.org/uniprot/P61978 | 0.825352539 |
| O75347 | https://www.uniprot.org/uniprot/O75347 | 0.697386778 |
| Q9BRK5 | https://www.uniprot.org/uniprot/Q9BRK5 | 0.717790543 |
| Q10471 | https://www.uniprot.org/uniprot/Q10471 | 0.738567938 |
| Q16531 | https://www.uniprot.org/uniprot/Q16531 | 0.650909323 |
| P39687 | https://www.uniprot.org/uniprot/P39687 | 0.850021715 |
| Q13813 | https://www.uniprot.org/uniprot/Q13813 | 0.619384321 |
| Q08174 | https://www.uniprot.org/uniprot/Q08174 | 0.636897162 |
| P13611 | https://www.uniprot.org/uniprot/P13611 | 0.724051615 |
| P98153 | https://www.uniprot.org/uniprot/P98153 | 0.613935648 |
| O95998 | https://www.uniprot.org/uniprot/O95998 | 0.623194939 |
| O95633 | https://www.uniprot.org/uniprot/O95633 | 0.624199458 |
| P13489 | https://www.uniprot.org/uniprot/P13489 | 0.789004913 |
| P01834 | https://www.uniprot.org/uniprot/P01834 | 0.669639026 |
| P51665 | https://www.uniprot.org/uniprot/P51665 | 0.707004596 |
| O60664 | https://www.uniprot.org/uniprot/O60664 | 0.791108729 |
| P06280 | https://www.uniprot.org/uniprot/P06280 | 0.674895749 |
| P50238 | https://www.uniprot.org/uniprot/P50238 | 0.663914728 |
| P61769 | https://www.uniprot.org/uniprot/P61769 | 0.651460958 |
| P07195 | https://www.uniprot.org/uniprot/P07195 | 0.695176465 |
| P05089 | https://www.uniprot.org/uniprot/P05089 | 0.708832877 |
| P15121 | https://www.uniprot.org/uniprot/P15121 | 0.616598513 |
| P08138 | https://www.uniprot.org/uniprot/P08138 | 0.664875656 |
| Q9NRV9 | https://www.uniprot.org/uniprot/Q9NRV9 | 0.788998135 |
| P07910 | https://www.uniprot.org/uniprot/P07910 | 0.806804824 |
| P36222 | https://www.uniprot.org/uniprot/P36222 | 0.655045005 |
| Q96C86 | https://www.uniprot.org/uniprot/Q96C86 | 0.687900474 |
| P07988 | https://www.uniprot.org/uniprot/P07988 | 0.699334504 |
| O75131 | https://www.uniprot.org/uniprot/O75131 | 0.727058108 |
| Q8IZ83 | https://www.uniprot.org/uniprot/Q8IZ83 | 0.788303888 |
| P27105 | https://www.uniprot.org/uniprot/P27105 | 0.720468946 |
| Q13308 | https://www.uniprot.org/uniprot/Q13308 | 0.628866313 |
| Q6UXH1 | https://www.uniprot.org/uniprot/Q6UXH1 | 0.752349756 |
| P78417 | https://www.uniprot.org/uniprot/P78417 | 0.729390151 |
| P28072 | https://www.uniprot.org/uniprot/P28072 | 0.821925636 |
| Q6JBY9 | https://www.uniprot.org/uniprot/Q6JBY9 | 0.785798788 |
| Q9Y315 | https://www.uniprot.org/uniprot/Q9Y315 | 0.689412694 |
| Q8NBJ4 | https://www.uniprot.org/uniprot/Q8NBJ4 | 0.699382091 |
| Q9H173 | https://www.uniprot.org/uniprot/Q9H173 | 0.722431588 |
| O60506 | https://www.uniprot.org/uniprot/O60506 | 0.726807869 |
| Q9NZL9 | https://www.uniprot.org/uniprot/Q9NZL9 | 0.69667276 |

| | | |
|---|---|---|
| P18827 | https://www.uniprot.org/uniprot/P18827 | 0.730199956 |
| P20827 | https://www.uniprot.org/uniprot/P20827 | 0.656947227 |
| P62987 | https://www.uniprot.org/uniprot/P62987 | 0.781970623 |
| Q9NY33 | https://www.uniprot.org/uniprot/Q9NY33 | 0.762818836 |
| Q13443 | https://www.uniprot.org/uniprot/Q13443 | 0.612125461 |
| P17900 | https://www.uniprot.org/uniprot/P17900 | 0.640704294 |
| P43490 | https://www.uniprot.org/uniprot/P43490 | 0.820392279 |
| Q15257 | https://www.uniprot.org/uniprot/Q15257 | 0.601434859 |
| Q86X29 | https://www.uniprot.org/uniprot/Q86X29 | 0.668829564 |
| O14672 | https://www.uniprot.org/uniprot/O14672 | 0.664301678 |
| Q01469 | https://www.uniprot.org/uniprot/Q01469 | 0.695924109 |
| O75914 | https://www.uniprot.org/uniprot/O75914 | 0.731455374 |
| Q9HC38 | https://www.uniprot.org/uniprot/Q9HC38 | 0.727357995 |
| Q99674 | https://www.uniprot.org/uniprot/Q99674 | 0.601708954 |
| P25815 | https://www.uniprot.org/uniprot/P25815 | 0.798284445 |
| P00492 | https://www.uniprot.org/uniprot/P00492 | 0.75955418 |
| Q9NR34 | https://www.uniprot.org/uniprot/Q9NR34 | 0.619223159 |
| Q92598 | https://www.uniprot.org/uniprot/Q92598 | 0.653606542 |
| P63208 | https://www.uniprot.org/uniprot/P63208 | 0.782839697 |
| P07451 | https://www.uniprot.org/uniprot/P07451 | 0.603100052 |
| P02763 | https://www.uniprot.org/uniprot/P02763 | 0.749420828 |
| Q9UKK9 | https://www.uniprot.org/uniprot/Q9UKK9 | 0.721754684 |
| Q9BRT3 | https://www.uniprot.org/uniprot/Q9BRT3 | 0.702019537 |
| Q9BXD5 | https://www.uniprot.org/uniprot/Q9BXD5 | 0.621504387 |
| Q8N257 | https://www.uniprot.org/uniprot/Q8N257 | 0.763069683 |
| Q15631 | https://www.uniprot.org/uniprot/Q15631 | 0.819628314 |
| Q13217 | https://www.uniprot.org/uniprot/Q13217 | 0.719962899 |
| P06454 | https://www.uniprot.org/uniprot/P06454 | 0.865976042 |
| Q16881 | https://www.uniprot.org/uniprot/Q16881 | 0.701881099 |
| Q9Y624 | https://www.uniprot.org/uniprot/Q9Y624 | 0.803186487 |
| P28070 | https://www.uniprot.org/uniprot/P28070 | 0.801440986 |
| Q14103 | https://www.uniprot.org/uniprot/Q14103 | 0.769266925 |
| Q92743 | https://www.uniprot.org/uniprot/Q92743 | 0.733124935 |
| P62826 | https://www.uniprot.org/uniprot/P62826 | 0.688077584 |
| P01857 | https://www.uniprot.org/uniprot/P01857 | 0.642443755 |
| P28062 | https://www.uniprot.org/uniprot/P28062 | 0.770206188 |
| Q9GZP4 | https://www.uniprot.org/uniprot/Q9GZP4 | 0.631781469 |
| Q9ULC4 | https://www.uniprot.org/uniprot/Q9ULC4 | 0.673500964 |
| O00264 | https://www.uniprot.org/uniprot/O00264 | 0.607970054 |
| Q7Z5L0 | https://www.uniprot.org/uniprot/Q7Z5L0 | 0.681220682 |
| Q86X76 | https://www.uniprot.org/uniprot/Q86X76 | 0.632589941 |
| P08311 | https://www.uniprot.org/uniprot/P08311 | 0.858982831 |
| P01876 | https://www.uniprot.org/uniprot/P01876 | 0.771883138 |
| P30520 | https://www.uniprot.org/uniprot/P30520 | 0.745140226 |
| O60568 | https://www.uniprot.org/uniprot/O60568 | 0.632292258 |
| O00233 | https://www.uniprot.org/uniprot/O00233 | 0.661014934 |
| Q9Y6Y9 | https://www.uniprot.org/uniprot/Q9Y6Y9 | 0.613219522 |

| | | |
|---|---|---|
| P46926 | https://www.uniprot.org/uniprot/P46926 | 0.735025546 |
| P21810 | https://www.uniprot.org/uniprot/P21810 | 0.672676195 |
| Q8NCW5 | https://www.uniprot.org/uniprot/Q8NCW5 | 0.687564085 |
| O95394 | https://www.uniprot.org/uniprot/O95394 | 0.600350877 |
| P14550 | https://www.uniprot.org/uniprot/P14550 | 0.638386732 |
| P28066 | https://www.uniprot.org/uniprot/P28066 | 0.795397992 |
| Q13616 | https://www.uniprot.org/uniprot/Q13616 | 0.671298538 |
| O15067 | https://www.uniprot.org/uniprot/O15067 | 0.71596756 |
| Q99536 | https://www.uniprot.org/uniprot/Q99536 | 0.786074887 |
| P49327 | https://www.uniprot.org/uniprot/P49327 | 0.673887109 |
| P62333 | https://www.uniprot.org/uniprot/P62333 | 0.615959516 |
| Q13162 | https://www.uniprot.org/uniprot/Q13162 | 0.686741801 |
| P31153 | https://www.uniprot.org/uniprot/P31153 | 0.712044925 |
| P54727 | https://www.uniprot.org/uniprot/P54727 | 0.645380013 |
| Q05639 | https://www.uniprot.org/uniprot/Q05639 | 0.843328698 |
| P30046 | https://www.uniprot.org/uniprot/P30046 | 0.853496731 |
| P05387 | https://www.uniprot.org/uniprot/P05387 | 0.862579409 |
| P54725 | https://www.uniprot.org/uniprot/P54725 | 0.750410491 |
| P61019 | https://www.uniprot.org/uniprot/P61019 | 0.624661109 |
| P12956 | https://www.uniprot.org/uniprot/P12956 | 0.761338566 |
| Q14116 | https://www.uniprot.org/uniprot/Q14116 | 0.683504008 |
| O15400 | https://www.uniprot.org/uniprot/O15400 | 0.707219605 |
| P48507 | https://www.uniprot.org/uniprot/P48507 | 0.638370407 |
| O95336 | https://www.uniprot.org/uniprot/O95336 | 0.701424653 |
| P80188 | https://www.uniprot.org/uniprot/P80188 | 0.760372384 |
| P14649 | https://www.uniprot.org/uniprot/P14649 | 0.816281201 |
| Q9ULZ3 | https://www.uniprot.org/uniprot/Q9ULZ3 | 0.865894831 |
| Q53FA7 | https://www.uniprot.org/uniprot/Q53FA7 | 0.803909843 |
| Q9HA64 | https://www.uniprot.org/uniprot/Q9HA64 | 0.609071828 |
| Q9Y3I1 | https://www.uniprot.org/uniprot/Q9Y3I1 | 0.606062589 |
| P28074 | https://www.uniprot.org/uniprot/P28074 | 0.720016396 |
| A6NI72 | https://www.uniprot.org/uniprot/A6NI72 | 0.852281366 |
| Q9GZN8 | https://www.uniprot.org/uniprot/Q9GZN8 | 0.705905765 |
| P07738 | https://www.uniprot.org/uniprot/P07738 | 0.643240803 |
| Q9Y2V2 | https://www.uniprot.org/uniprot/Q9Y2V2 | 0.759232581 |
| Q10589 | https://www.uniprot.org/uniprot/Q10589 | 0.644233232 |
| P05787 | https://www.uniprot.org/uniprot/P05787 | 0.665309038 |
| Q15904 | https://www.uniprot.org/uniprot/Q15904 | 0.696472942 |
| O43583 | https://www.uniprot.org/uniprot/O43583 | 0.638075556 |
| Q6XQN6 | https://www.uniprot.org/uniprot/Q6XQN6 | 0.673602181 |
| P20340 | https://www.uniprot.org/uniprot/P20340 | 0.63845793 |
| P35052 | https://www.uniprot.org/uniprot/P35052 | 0.617890378 |
| Q92692 | https://www.uniprot.org/uniprot/Q92692 | 0.702121439 |
| P49721 | https://www.uniprot.org/uniprot/P49721 | 0.808445279 |
| P49720 | https://www.uniprot.org/uniprot/P49720 | 0.794014591 |
| P59665 | https://www.uniprot.org/uniprot/P59665 | 0.79513905 |
| P01714 | https://www.uniprot.org/uniprot/P01714 | 0.71805358 |

| | | |
|---|---|---|
| Q9BYF1 | https://www.uniprot.org/uniprot/Q9BYF1 | 0.720579407 |
| Q9BUD6 | https://www.uniprot.org/uniprot/Q9BUD6 | 0.721758049 |
| P01040 | https://www.uniprot.org/uniprot/P01040 | 0.663948357 |
| O43768 | https://www.uniprot.org/uniprot/O43768 | 0.660633154 |
| Q9NRX4 | https://www.uniprot.org/uniprot/Q9NRX4 | 0.762781059 |
| P15941 | https://www.uniprot.org/uniprot/P15941 | 0.647359555 |
| Q7L5N1 | https://www.uniprot.org/uniprot/Q7L5N1 | 0.668374737 |
| Q8N339 | https://www.uniprot.org/uniprot/Q8N339 | 0.697872099 |
| Q9NZD4 | https://www.uniprot.org/uniprot/Q9NZD4 | 0.613851719 |
| Q9BRA2 | https://www.uniprot.org/uniprot/Q9BRA2 | 0.711480914 |
| Q969H8 | https://www.uniprot.org/uniprot/Q969H8 | 0.71654806 |
| P61956 | https://www.uniprot.org/uniprot/P61956 | 0.644932756 |
| P26022 | https://www.uniprot.org/uniprot/P26022 | 0.69311028 |
| Q9H3K6 | https://www.uniprot.org/uniprot/Q9H3K6 | 0.703815073 |
| P61960 | https://www.uniprot.org/uniprot/P61960 | 0.695981137 |
| Q9Y6C2 | https://www.uniprot.org/uniprot/Q9Y6C2 | 0.627662486 |
| Q9UGN4 | https://www.uniprot.org/uniprot/Q9UGN4 | 0.636194445 |
| Q15366 | https://www.uniprot.org/uniprot/Q15366 | 0.657711599 |
| Q8N0Y7 | https://www.uniprot.org/uniprot/Q8N0Y7 | 0.783859006 |
| Q0VD83 | https://www.uniprot.org/uniprot/Q0VD83 | 0.859209897 |
| P28065 | https://www.uniprot.org/uniprot/P28065 | 0.720210275 |
| Q9UGM3 | https://www.uniprot.org/uniprot/Q9UGM3 | 0.669636509 |
| O94766 | https://www.uniprot.org/uniprot/O94766 | 0.668513746 |
| P18669 | https://www.uniprot.org/uniprot/P18669 | 0.711038914 |
| O95757 | https://www.uniprot.org/uniprot/O95757 | 0.799694829 |
| P61026 | https://www.uniprot.org/uniprot/P61026 | 0.719862103 |
| P35318 | https://www.uniprot.org/uniprot/P35318 | 0.715675821 |
| A8MUU1 | https://www.uniprot.org/uniprot/A8MUU1 | 0.678150647 |
| Q6ZMR3 | https://www.uniprot.org/uniprot/Q6ZMR3 | 0.645773022 |
| O75882 | https://www.uniprot.org/uniprot/O75882 | -0.659858961 |
| P02765 | https://www.uniprot.org/uniprot/P02765 | -0.617346155 |
| P01008 | https://www.uniprot.org/uniprot/P01008 | -0.666697339 |
| P00734 | https://www.uniprot.org/uniprot/P00734 | -0.708108656 |
| P03952 | https://www.uniprot.org/uniprot/P03952 | -0.693444911 |
| P06396 | https://www.uniprot.org/uniprot/P06396 | -0.71627292 |
| P19827 | https://www.uniprot.org/uniprot/P19827 | -0.722990599 |
| Q96PD5 | https://www.uniprot.org/uniprot/Q96PD5 | -0.722746601 |
| P35858 | https://www.uniprot.org/uniprot/P35858 | -0.648599681 |
| P19823 | https://www.uniprot.org/uniprot/P19823 | -0.711987021 |
| P43652 | https://www.uniprot.org/uniprot/P43652 | -0.628269694 |
| P55290 | https://www.uniprot.org/uniprot/P55290 | -0.676283516 |
| P02790 | https://www.uniprot.org/uniprot/P02790 | -0.678288707 |
| P17936 | https://www.uniprot.org/uniprot/P17936 | -0.692204038 |
| O14793 | https://www.uniprot.org/uniprot/O14793 | -0.610419332 |
| P29622 | https://www.uniprot.org/uniprot/P29622 | -0.759938045 |
| P06276 | https://www.uniprot.org/uniprot/P06276 | -0.665540769 |
| P43251 | https://www.uniprot.org/uniprot/P43251 | -0.679348744 |

| | | |
|---|---|---|
| P54289 | https://www.uniprot.org/uniprot/P54289 | -0.637604299 |
| P05452 | https://www.uniprot.org/uniprot/P05452 | -0.702117454 |
| P27918 | https://www.uniprot.org/uniprot/P27918 | -0.614091681 |
| Q12860 | https://www.uniprot.org/uniprot/Q12860 | -0.63492054 |
| Q76LX8 | https://www.uniprot.org/uniprot/Q76LX8 | -0.643813892 |
| P49908 | https://www.uniprot.org/uniprot/P49908 | -0.645291133 |
| P00533 | https://www.uniprot.org/uniprot/P00533 | -0.643473616 |
| Q9UBQ6 | https://www.uniprot.org/uniprot/Q9UBQ6 | -0.605808746 |
| O15335 | https://www.uniprot.org/uniprot/O15335 | -0.668053383 |
| Q12913 | https://www.uniprot.org/uniprot/Q12913 | -0.641679392 |
| Q16832 | https://www.uniprot.org/uniprot/Q16832 | -0.635131472 |
| Q8NFT8 | https://www.uniprot.org/uniprot/Q8NFT8 | -0.625062185 |
| Q16620 | https://www.uniprot.org/uniprot/Q16620 | -0.703619993 |
| O75144 | https://www.uniprot.org/uniprot/O75144 | -0.617410449 |
